# Genotypic infection endpoint analysis to understand efficacy and escape potential of the malaria monoclonal antibody CIS43LS

**DOI:** 10.64898/2026.11.04.26367264

**Authors:** Philipp Schwabl, Amadou Niangaly, William Nickols, Viviane Callier, Allyson Mateja, Kassoum Kayentao, Aissata Ongoiba, Jeff Skinner, Anne C. Preston, Safiatou Doumbo, Didier Doumtabe, Jorge E. Amaya-Romero, Katrina A. Kelley, Shanping Li, Margaret Laws, Mary E. Peterson, Tuan M. Tran, Sean C. Murphy, Boubacar Traore, Robert A. Seder, Peter D. Crompton, Gail E. Potter, Daniel E. Neafsey

**Affiliations:** Department of Immunology and Infectious Diseases, Harvard TH Chan School of Public Health; Boston, MA, USA; Infectious Disease and Microbiome Program, Broad Institute; Cambridge, MA, USA; Parasites and Microbes Research and Training Centre, Mali International Center of Excellence in Research, University of Sciences, Techniques and Technologies of Bamako; Bamako, Mali; Department of Biostatistics, Harvard TH Chan School of Public Health; Boston, MA, USA; Clinical Monitoring Research Program Directorate, Frederick National Laboratory for Cancer Research; Rockville, MD, USA; Malaria Infection Biology and Immunity Section, Laboratory of Immunogenetics, Division of Intramural Research, National Institute of Allergy and Infectious Diseases, National Institutes of Health; Rockville, MD, USA; Division of Infectious Diseases, Department of Medicine, Indiana University School of Medicine; Indianapolis, IN, USA; Department of Laboratory Medicine and Pathology and Center for Emerging and Re-emerging Infectious Diseases, University of Washington; Seattle, WA, USA; Cellular Immunology Section, Vaccine Research Center, National Institute of Allergy and Infectious Diseases, National Institutes of Health; Bethesda, MD, USA; Biostatistics Research Branch, National Institute of Allergy and Infectious Diseases, National Institutes of Health, Rockville; MD, USA

## Abstract

Early-phase trials of candidate malaria interventions in endemic areas often use “time-to-first” infection as a primary efficacy endpoint. This approach does not assess infection risk after the initial infection, and its censoring of trial participants after their first infection can introduce bias by removing more high-risk participants from control than treatment arms over time. Efficacy estimate precision may also decline as risk set size decreases. Here we assessed the utility of genotyping all infections in an intervention trial to determine the total infection incidence in each participant for the full follow-up period. Specifically, we genotyped all *Plasmodium falciparum* infections detected in a previously-reported phase 2 trial of the antimalarial monoclonal antibody CIS43LS in 330 Malian adults. We determined the molecular force of infection (molFOI; rate of new *P. falciparum* strains) for each participant, expressing efficacy as the percent molFOI reduction in CIS43LS versus placebo arms (Eff_molFOI_). Six-month Eff_molFOI_ was 86.5% (95% CI: 80.1-90.8) and 70.7% (61.0-78.0) for the 40 mg/kg and 10 mg/kg doses, respectively. These estimates are similar to those from time-to-first infection analysis, but more precise when analyzing efficacy by trial-month and when stratifying by *P. falciparum* infection-status at enrollment. Participants infected at enrollment had lower Eff_molFOI_ and acquired infections earlier in the trial, creating systematic differences in risk composition between arms over time. Genotyping showed no evidence for escape-polymorphisms in the CIS43LS epitope. These results demonstrate how genotypic endpoints can provide a more comprehensive understanding of candidate malaria interventions.

**One sentence summary:** Malaria parasite genotyping and molecular force of infection inference improves standard malaria intervention efficacy analysis.

## INTRODUCTION

Malaria remains a significant public health threat, causing over 600,000 deaths and 250 million clinical cases each year (1). The disease burden is greatest in Sub-Saharan Africa, where *Plasmodium falciparum* is the dominant species of malaria parasite. Following a decade of stagnating progress in malaria control, new momentum is expected from the recent implementation of the malaria vaccines RTS,S/AS01 and R21/Matrix-M in Africa, primarily aimed at reducing disease burden among young children, and from a recent surge in the development of new countermeasures suitable for wider age groups as well as during pregnancy. New biomedical interventions in development include next-generation vaccines, long-acting monoclonal antibodies (mAbs) (2), and long-acting injectable small-molecule drugs (LAIs) (3). These strategies are intended to complement mosquito vector interventions, which continue to represent the mainstay of malaria control (4).

Traditionally, early-phase clinical trials that are designed to assess the efficacy of candidate malaria interventions in endemic regions have often relied on “time-to-first event” analysis using Cox proportional hazards models. These may use time-to-first malaria infection (5–10) based on thick blood smear microscopy (TBS) or PCR-based methods performed on serially sampled blood as primary endpoints (8,9,11).

The use of time-to-first-event endpoints in malaria intervention trials has several important limitations. First, TBS and PCR-based methods cannot distinguish new vs. persisting infections. The approach assumes that time to first infection adequately reflects the overall longitudinal risk of infection during the entire trial period. Second, censoring participants after their first infection can introduce bias by decreasing the size of the study population and changing its risk profile over time. For example, individuals in the control arm who are at high risk of infective mosquito bites, such as those living near mosquito breeding sites, are more likely to be infected and censored early in the trial, while those in the intervention arm(s) who are at equally high risk of infective mosquito bites may remain in the at-risk population longer if the intervention is protective. The control arm is thus expected to become enriched for individuals at lower risk, resulting in an artefactual decline in intervention efficacy over time. Beyond distorting time-specific efficacy estimates, the time-to-first event approach can bias the composite efficacy metric by violating the proportional hazards and constant risk composition assumptions that underlie Cox models and similar survival regression methods (12,13).

The above limitations of time-to-event analysis can be addressed by including parasite genotypic endpoints to distinguish new from persistent infection states and thereby resolve total infection incidence. This approach enables all participants to remain in the at-risk population throughout the trial follow-up period, allowing for a more complete picture of the infection burden prevented by the intervention. In addition, strain-and allele-level analysis of breakthrough infections can identify parasite polymorphisms linked to intervention ‘escape’, providing insight into the mechanisms underlying intervention failure.

Here, we explored the utility of including parasite genotypic endpoints in clinical trials of malaria interventions by applying this approach to a previously described phase 2 trial of the long-acting mAb CIS43LS (6,14). CIS43LS targets a conserved ‘junctional’ epitope spanning the N-terminal and central repeat regions of the *P. falciparum* circumsporozoite protein (PfCSP) (15). As previously reported, a single intravenous dose of 40 mg/kg of CIS43LS provided 88.2% protective efficacy against the first *P. falciparum* infection detected by TBS (6) and 87.4% efficacy against the first infection detected by qRT-PCR (14) during a six-month malaria season. We re-analyzed this trial using genotypic endpoints generated via PCR amplicon sequencing (AmpSeq) at seven polymorphic *P. falciparum* targets, estimating the number of distinct strains newly acquired per follow-up visit per participant. We used the genotypic data to calculate three CIS43LS efficacy estimates. The first is efficacy against the molecular force of infection (molFOI), the number of newly acquired parasite strains over time (16), which we denote as Eff_molFOI_. Eff_molFOI_ is defined as one minus the ratio of incidence of new strains in the treated vs. control arms and incorporates the number of events over follow-up, but does not consider time-to-first event.

The second genotyping-informed estimate is the proportion reduction in the incidence of new malaria infections (where one “infection” may include multiple strains) over the course of follow-up. We denote this Eff_IRR_ for the incidence rate ratio.

The third estimate, Eff_FH_, integrates time to first infection together with the number of strains at first infection, but it does not consider subsequent infection events. Proposed by Follmann and Huang, this efficacy estimate is defined as the proportional reduction in mean number of strains that develop into a blood-stage infection from an exposure (17). Eff_FH_ is estimated as one minus the product of the Cox-estimated hazard ratio (HR) multiplied by the ratio of the mean number of strains at first infection in treated vs. control arms. We compare these genotyping-informed efficacy estimates to the time-to-first event-based efficacy estimates previously used to evaluate CIS43LS efficacy in this trial (6,14). These include efficacy calculated as 1 - HR based on thick blood smear microscopy, efficacy as 1 - HR based on qRT-PCR, and efficacy as 1 - risk ratio for the risk of at least one infection detected by qRT-PCR during the follow-up period.

## RESULTS

### Trial enrollment and follow-up

As previously reported, between May 6 and August 6, 2021, 330 healthy adult participants who met eligibility criteria were randomized and received a single IV infusion of normal saline placebo (n=110), or CIS43LS at a dose of 10 mg/kg (n=110) or 40 mg/kg (n=110) (6). Thereafter, dried blood spot (DBS) and TBS samples were collected at 3 to 14-day intervals for 24 weeks and during unscheduled illness visits when malaria was suspected (Fig. 1A). Follow-up through the last trial visit was completed by 102 (92.7%) participants who received placebo, 105 (95.5%) who received 10 mg/kg of CIS43LS, and 104 (94.5%) who received 40 mg/kg of CIS43LS.

**Fig. 1.**
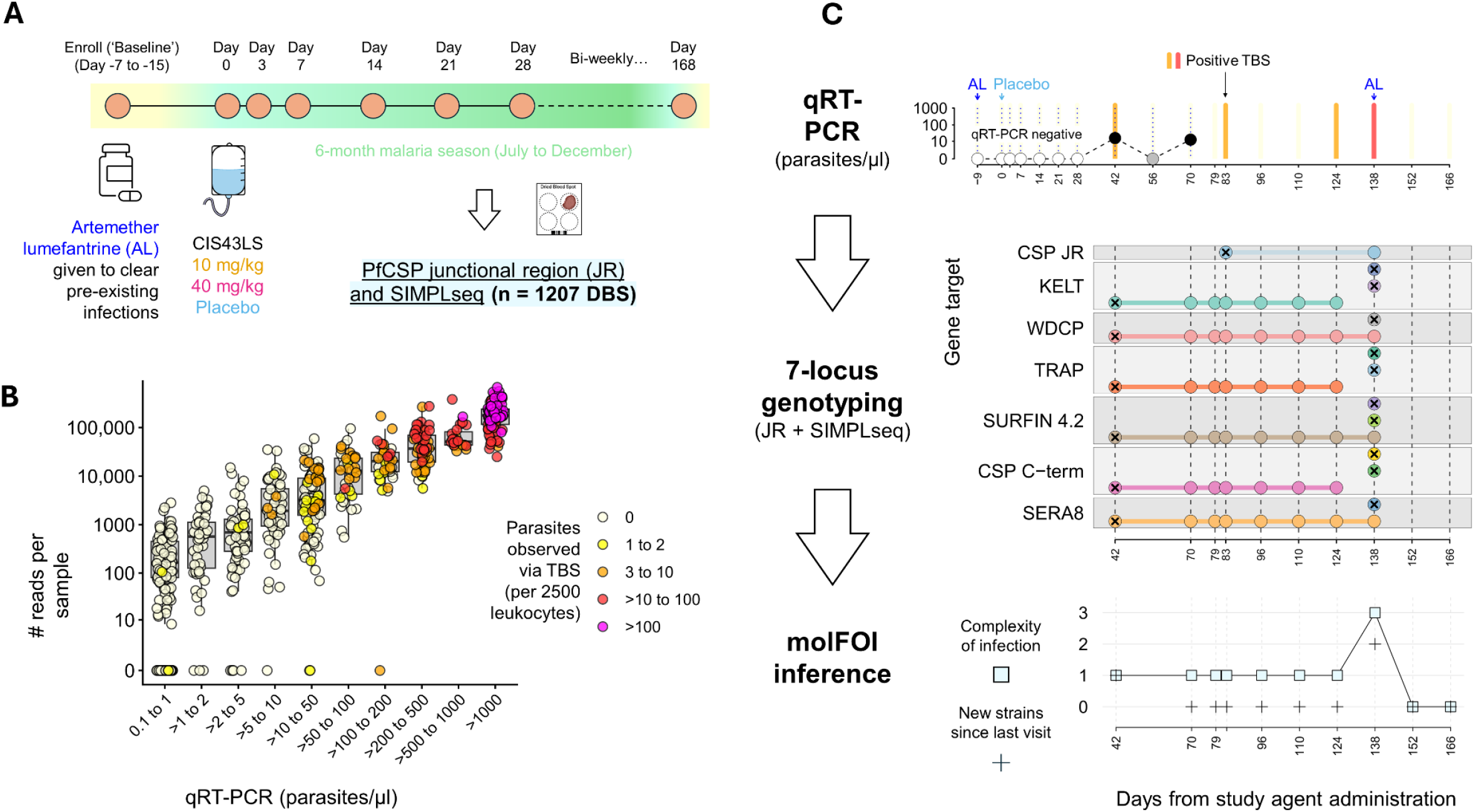
Study design leading to molFOI inference. (A) Time course of scheduled visits (tan circles) collecting thick blood smears (TBS) for microscopic examination and dried blood spots (DBS) for qRT-PCR and AmpSeq analysis targeting the PfCSP junctional region (JR) and 6 SIMPLseq loci. (B) AmpSeq sensitivity per DBS sample (circle) shown as total reads (y-axis, all 7 loci combined) per parasitemia bin, as determined by qRT-PCR (x-axis). Plot excludes samples below the qRT-PCR limit of detection (LOD) of 0.1 parasites/µl. Circle fill color indicates parasitemia by TBS microscopy. (C) Schematic of the quantitative parasitemia and genotypic assay results (example participant 0175G). In the top panel, dotted vertical lines indicate timepoints at which qRT-PCR testing was performed. Black fill circles indicate qRT-PCR results ≥0.1 parasites/µl (y-axis in log10). Grey and white fill circles indicate sub-LOD and negative qRT-PCR results, respectively. Shaded vertical line colors indicate TBS microscopy results as in B. The middle panel shows alleles (circles) detected per AmpSeq locus (boxes). Matching circle colors represent repeated detection of the same allele. X-symbols mark newly acquired alleles. The bottom panel translates the genotypic data into complexity of infection values and into the molecular force of infection endpoint (same y-axis).

### AmpSeq sensitivity and longitudinal infection dynamics

Seven-locus *P. falciparum* AmpSeq genotyping (SIMPLseq + Nterm assay, see Materials and Methods) was performed using DNA obtained from DBS collected from 232 participants (70.3% of total study population) during 1,207 study visits, from a total of 5,657 visits with available DBS. Of these 1,207 visits, 865 (71.7%) showed AmpSeq signal at one or more loci (Fig. 1B). Of note, 753 of the 1,207 samples subjected to AmpSeq had also been analyzed by qRT-PCR; sensitivity comparisons are described in Fig. S1. Together with the qRT-PCR and TBS data, the AmpSeq data provided longitudinal infection profiles for each study participant. Fig. 1C provides an example infection profile of a single participant in the placebo arm whose first infection was detected on day 42. For this infection, 1 allele was observed at each of the 7 genotyped loci, suggesting the presence of a single parasite strain (i.e. complexity of infection (COI) = 1). This participant carried the same strain at fluctuating parasitemias until day 138, at which point a symptomatic superinfection consisting of 2 new strains was observed, prompting successful treatment with artemether-lumefantrine (AL). Data files S1 and S2 provide infection trajectories for each AmpSeq-positive participant, with and without imputation for missing genotype data at AmpSeq-negative timepoints.

### CIS43LS efficacy based on molFOI

The combination of qRT-PCR and AmpSeq data enabled us to track new parasite strain acquisition over the complete follow-up period for each participant (Fig. 2A). Ten participants already showed new strain acquisition within 2 weeks after study agent administration (see triangles in Fig. 2B-C), possibly representing infections that were in the liver stage at enrollment and during initial AL treatment. The mean molFOI over the study period (i.e., infection rate expressed as new strains per year) was 10.5 in the placebo arm, 3.1 in the 10 mg/kg CIS43LS arm, and 1.4 in the 40 mg/kg CIS43LS arm, yielding efficacy estimates (Eff_molFOI_) of 70.7% [95% CI: 61.0, 78.0] for 10 mg/kg of CIS43LS and 86.5% [95% CI: 80.1, 90.8] for 40 mg/kg of CIS43LS.

**Fig. 2.**
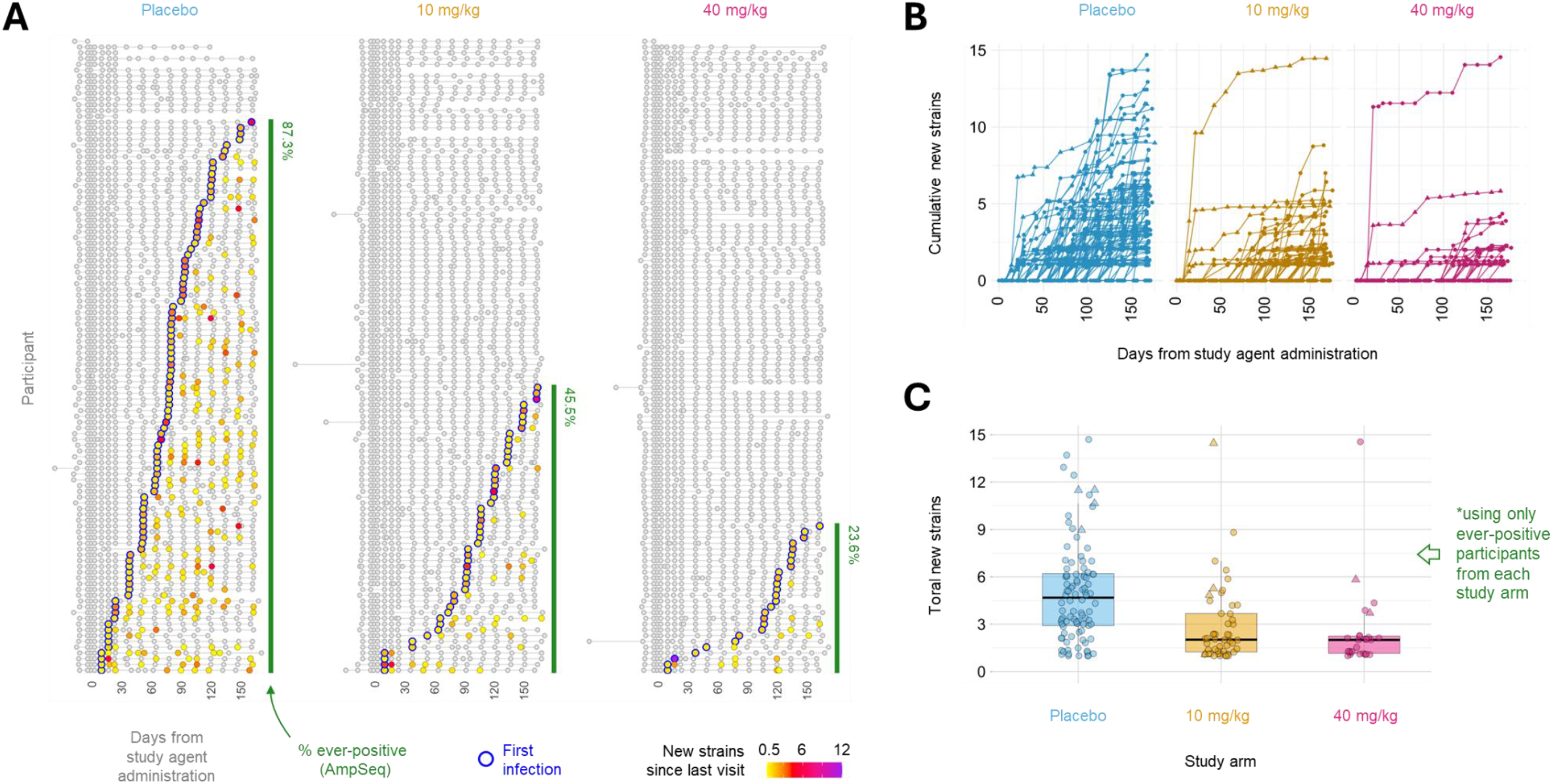
Infection profiles of individual study participants over the follow-up period. (A) Each row represents the visit series (circles) of one participant over the 6-month study (x-axis), ordered by time of first infection (green border) and grouped by study arm. Circle fill color indicates the number of new strains since the previous visit (grey if zero, otherwise see color scale). The fraction of participants in each study arm who had at least one new strain detected (>0.5 probability) after day 7 is indicated by blue bars at right. (B) As in A, each line represents the visit series of one participant over the 6-month study (x-axis), grouped by study arm. Point position on the y-axis indicates cumulative new strain acquisition by the time of each visit. Triangles are used for visit series in which new strain acquisition occurred by day 14. (C) Boxplots show median and inter-quartile ranges for the total number of new strains accumulated by the end of the study for each study arm. Only participants who had at least one new strain detected are included here (as opposed to yearly molFOI means reported in the main text, which also incorporate never-infected participants). Triangles are used as in B.

Eff_molFOI_ estimates were similar to efficacy estimates generated by the HR and FH methods: Eff_HR_ for qRT-PCR positivity was 76.0% [95% CI: 65.7, 83.2] and 85.0% [95% CI: 77.2, 90.1] for 10 mg/kg and 40 mg/kg of CIS43LS, respectively; and Eff_FH_ was 75.9% [95% CI: 62.3, 84.6] and 88.2% [95% CI: 74.2, 94.6] for 10 mg/kg and 40 mg/kg of CIS43LS, respectively (Fig. 3). Eff_FH_ and Eff_HR_ were similar since the mean number of incident strains at the first follow-up infection was nearly identical among study arms (1.66 for placebo, 1.66 for 10 mg/kg of CIS43LS, and 1.62 for 40 mg/kg of CIS43LS). Efficacy against the incidence of distinct infection events (Eff_IRR_, with incidence rate ratio distinguished by genotyping) was slightly lower than Eff_molFOI_ and Eff_HR_: 68% [95%CI: 56, 76] and 82% [95%CI: 74, 95] for 10 mg/kg and 40 mg/kg of CIS43LS, respectively. Efficacy estimates by HR for TBS positivity were similar to those for qRT-PCR positivity and had similar precision. Estimates were lower using risk ratio efficacy, which does not factor in the timing at which infections occur and does not account for multiple infections: 49.4% [95% CI: 37.0, 59.4] for 10 mg/kg of CIS43LS, and 66.4% [95% CI: 54.6, 75.0] for 40 mg/kg of CIS43LS (Fig. 3). See Materials and Methods for further definition of efficacy estimates.

**Fig. 3.**
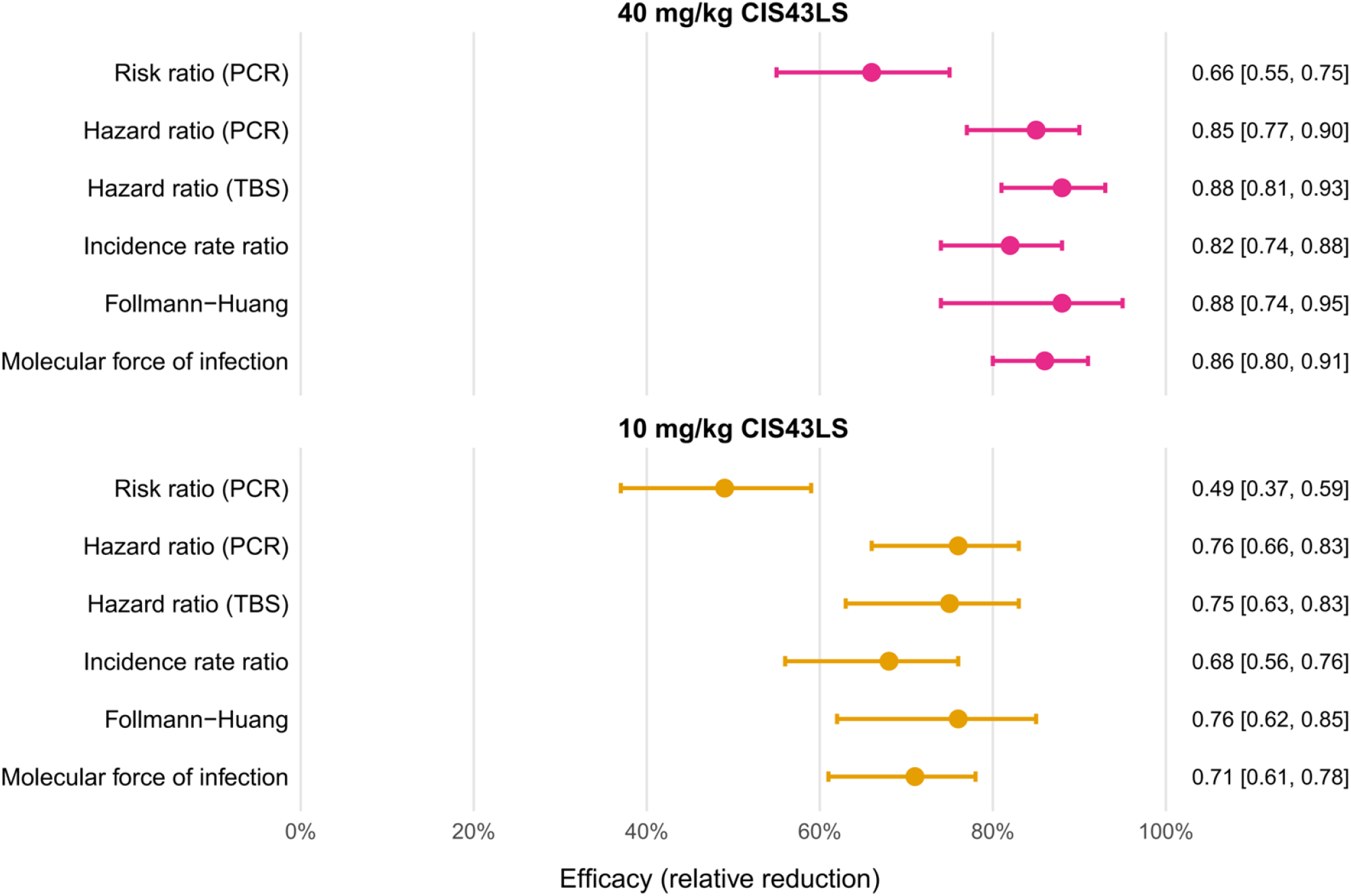
CIS43LS efficacy estimates with 95% confidence intervals using time to first infection and genotypic data. Risk ratio efficacy is the CIS43LS-induced proportion reduction in probability of qRT-PCR-detected malaria infection during follow-up. Hazard ratio efficacy is the CIS43LS-induced proportion reduction in hazard of qRT-PCR-detected or TBS-detected malaria infection during follow-up. Incidence rate ratio efficacy is the proportion reduction in the incidence of new malaria infections (where one “infection” may include multiple strains) over the course of follow-up. Follmann-Huang efficacy is the CIS43LS-induced proportion reduction in mean number of infecting strains per exposure. Molecular force of infection efficacy is the proportion reduction in the incidence of strains per unit time over the course of follow-up.

### molFOI trajectories

Next, we plotted the predicted time-varying molFOI trajectories by study arm over the 6-month trial period (Fig. 4A). molFOI was relatively low for all 3 study arms during the first 28 days of follow-up but then rapidly increased in the placebo arm, peaking at 16.0 strains/year at day 96 before decreasing to 8.8 strains/year by day 168. In the 10 mg/kg CIS43LS arm, molFOI gradually increased from approximately day 40 through day 168, peaking at approximately 5.7 strains/year. In the 40 mg/kg CIS43LS arm, molFOI remained near zero throughout the 6-month trial with the exception of a modestly elevated initial level and a small peak (3.1 strains/year) around day 122. The monthly discretized molFOI estimates (Fig. 4B) were similar to the continuous predictions shown in Fig. 4A.

**Fig. 4.**
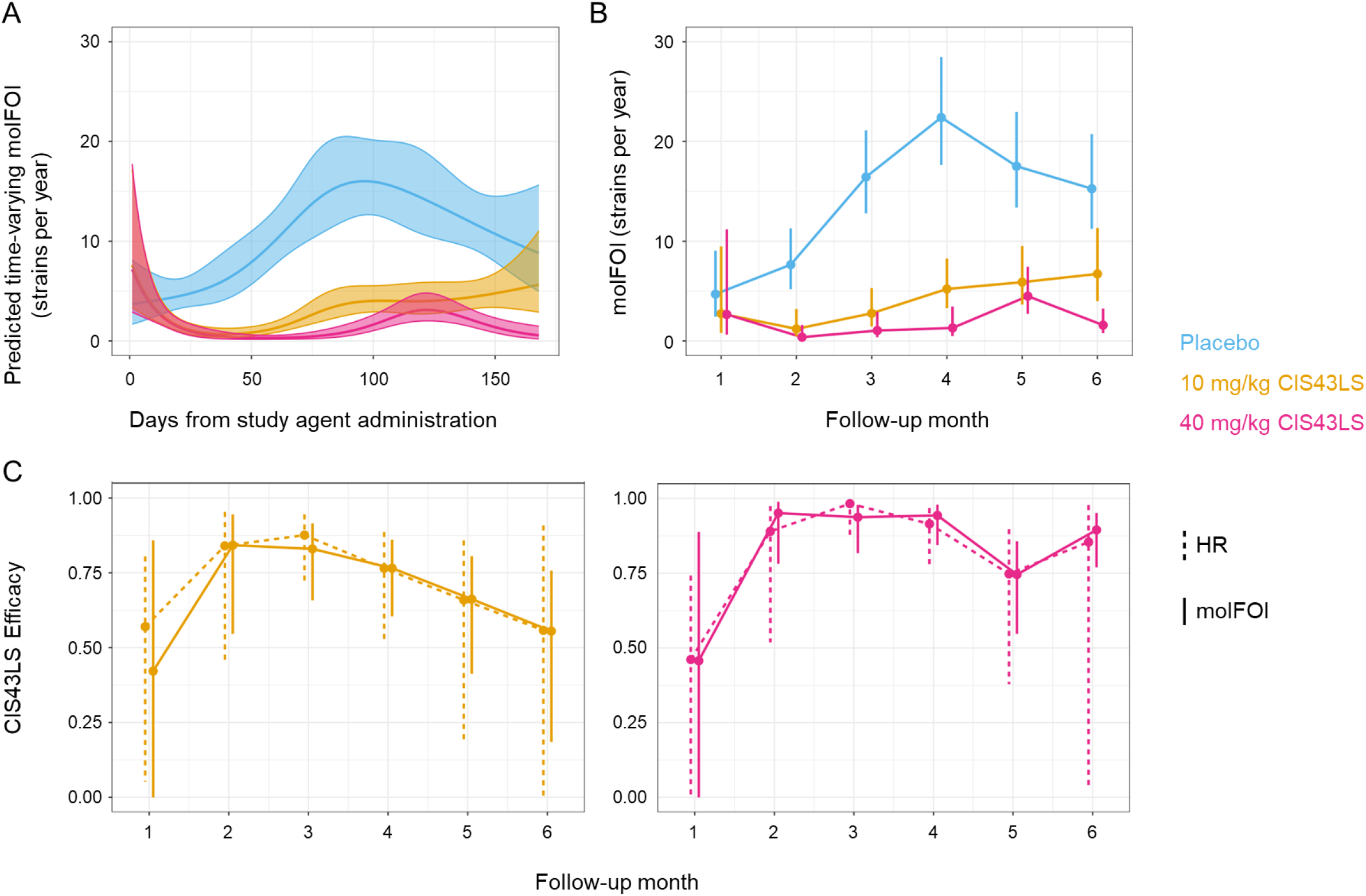
Time-varying molFOI trajectories by study arm and CIS43LS efficacy by the molFOI and HR methods over the 6-month trial. (A) Continuous predicted time-varying molFOI trajectories by study arm over the 6-month trial. Shaded areas indicate 95% confidence intervals. (B) Average monthly discretized molFOI estimates by study arm over the 6-month trial. Circles represent sample means, and vertical whiskers represent 95% confidence intervals. (C) CIS43LS efficacy versus placebo for the 10 mg/kg arm (left panel) and 40 mg/kg arm (right panel) estimated per month by the molFOI method (solid lines) and HR method (dashed lines). Circles represent sample means and vertical whiskers represent 95% confidence intervals.

Monthly CIS43LS Eff_molFOI_ and Eff_HR_ estimates compared to placebo are visualized in Fig. 4C. 40 mg/kg of CIS43LS showed higher Eff_molFOI_ than 10 mg/kg each month of the 6-month follow-up period, and Eff_molFOI_ more clearly waned in the 10 mg/kg arm than the 40 mg/kg arm over the follow-up period. Temporal patterns of Eff_HR_ and Eff_molFOI_ were highly congruent over the follow-up period, but 95% confidence intervals for monthly efficacy estimates were wider by the Eff_HR_ method compared to Eff_molFOI_ (with the exception of month 1), especially during months 4-6 (Table S1). The wider 95% confidence intervals for Eff_HR_ at month 6 fail to detect statistically significant Eff_HR_ for the 10 mg/kg dose during month 6 (Eff_HR_= 55.8%; 95% CI: −0.01, 90.8; p = 0.31), whereas Eff_molFOI_ for 10 mg/kg was statistically significant at month 6 (55.5%; 95% CI: 18.5, 75.7; p = 0.009). For 40 mg/kg of CIS43LS both Eff_HR_ and Eff_molFO_ were statistically significant at all timepoints, but the higher precision of the Eff_molFOI_ method resulted in narrower 95% confidence intervals; for example, the lower bound of the 95% confidence interval at 6 months was 0.77 for Eff_molFOI_ compared to 0.032 for Eff_HR_.

### Relationship between baseline *P. falciparum* infection-status and subsequent molFOI, clinical malaria risk, and CIS43LS efficacy

To evaluate differential changes between arms in risk set composition, we explored the relationship between *P. falciparum* infection-status at study enrollment, as determined by qRT-PCR, and the subsequent molFOI, risk of clinical malaria, and CIS43LS efficacy. After clearance of baseline infection with AL, we found that monthly estimates of molFOI were higher among participants who were enrollment-positive (EP, n = 133) versus enrollment-negative (EN, n = 194) (Fig. 5A). EP participants also acquired their first new *P. falciparum* infection significantly earlier than EN participants (Fig. 5B, p = 0.026), resulting in more rapid depletion (i.e. censoring) of EP participants from the study population included in time-to-first infection analysis. This accelerated depletion effect was apparent across all three study arms during early follow-up (Fig. S2A), suggesting it may result from infections that were at the hepatic stage during enrollment. For example, from day 0 to 21, the proportion of EP participants declined by 18.5% in the placebo arm, 7.4% in the 10 mg/kg arm, and 10.9% in the 40 mg/kg arm. The proportion of EN participants increased, respectively, by 14.2%, 4.8%, and 7.8%. In contrast, the initial decline in EP proportion in the risk set for TBS positivity was similar between treatment groups, but the EP proportion declined more steeply in controls starting around Day 120. (Fig. S2B).

**Fig. 5.**
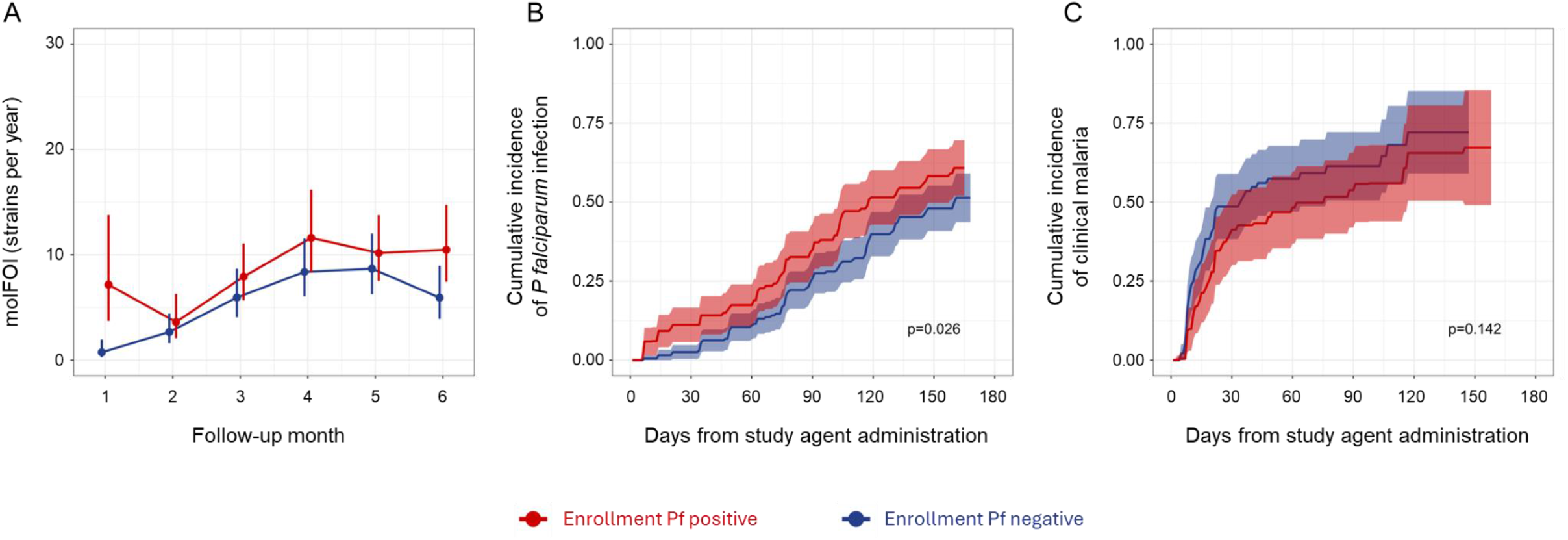
Relationship between baseline *P. falciparum* infection and the subsequent risk of *P. falciparum* infection and clinical malaria. (A) Monthly estimates of molFOI among participants stratified by the presence (n=133) or absence (n=194) of qRT-PCR-detected *P. falciparum* infection at enrollment. Vertical whiskers indicate 95% confidence intervals. (B) Cumulative incidence of qRT-PCR-detected *P. falciparum* infection (irrespective of symptoms being present) among participants stratified by the presence (n=133) or absence (n=194) of qRT-PCR-detected *P. falciparum* infection at enrollment. Shaded areas indicate 95% confidence intervals. (C) Cumulative incidence of symptomatic malaria after first infection stratified by the presence (n=133) or absence (n=194) of qRT-PCR-detected *P. falciparum* infection at enrollment. Shaded areas indicate 95% confidence intervals.

*P. falciparum* infection at enrollment may indicate a higher overall risk of infection, and as a result, a greater degree of acquired immunity to symptomatic malaria caused by blood stage parasites (18). Accordingly, despite the higher risk of infection among EP participants (Fig. 5B, p = 0.026), we observed a trend toward lower risk of clinical malaria among those who became infected (Fig. 5C) and lower median parasitemia when infected compared to EN participants (9.9 parasites/μl vs. 78.9 parasites/μl, p < 0.001, Fig. S3A). In addition, the average duration of asymptomatic blood-stage infections was longer in EP participants compared to EN participants, based on the length of allele persistence detected by genotyping over time in the absence of AL treatment (29 days vs. 15 days, p = 0.043, Fig. S3B), consistent with enhanced clinical immunity to blood-stage parasites.

Through interaction tests we also found that infection at enrollment was associated with reduced CIS43LS Eff_molFOI_ in the 40 mg/kg study arm (EP Eff_molFOI_ = 81.0% [69.1, 88.3], EN Eff_molFOI_ = 91.5% [84.2, 95.5], p = 0.043) (Fig. S4). We observed a similar trend in the 10 mg/kg arm, although this interaction was not statistically significant (EP Eff_molFOI_ = 64.0% [46.5, 75.8], EN Eff_molFOI_ = 75.5% [63.3, 83.6], p *= 0*.*184*). We applied the same interaction testing to the Eff_HR_ method and found that estimates were similar to Eff_molFOI_ but less precise with no significant interactions detected (Table S2).

### Polymorphisms in the region targeted by CIS43LS were not detected in breakthrough infections

Finally, for all qRT-PCR-detected infections that occurred after study agent administration, we sequenced the PfCSP junctional region (JR) epitope targeted by CIS43LS (PfCSP amino acid positions 101 to 115 covered by the Nterm assay) to determine whether breakthrough infections in CIS43LS recipients were associated with polymorphisms in this epitope, and no polymorphisms were detected. In addition, we investigated two polymorphisms proximal to the JR that are known to occur in West Africa (19): a nonsynonymous single nucleotide substitution from C to G causing an Alanine to Glycine amino acid change at codon position 98 (A98G), and a 57 bp insertion between codon positions 80 and 81 (with respect to the Pf3D7 reference genome). In this trial, we found that the A98G polymorphism and the 57 bp insertion were present in 27.9% and 33.0% of all newly acquired strains, respectively. However, there were no statistically significant differences among trial arms in the proportion of the A98G polymorphism (placebo: 30.1%; 10 mg/kg: 22.5%; 40 mg/kg: 23.1%; *p = 0*.*59*) or the 57 bp insertion in newly acquired strains (placebo: 33.3%; 10 mg/kg: 35.0%; 40 mg/kg: 23.1%; p = 0.72) (Fig. S5). Moreover, the presence of one polymorphism did not affect the probability of observing the other in the same haplotype (p = 0.16).

## DISCUSSION

Here we explored the use of *P. falciparum* genotypic analysis in a clinical trial of a candidate malaria intervention to estimate efficacy and to detect parasite polymorphisms that might escape the intervention.

Specifically, we genotyped *P. falciparum* infections that were detected in a phase 2 trial of the anti-malaria mAb CIS43LS and compared efficacy estimates based on molFOI to traditional methods that use PCR- or TBS-based infection endpoints and time-to-first infection (HR) analysis. Overall, we found that parasite genotypic analysis provided a more comprehensive picture of intervention efficacy than is possible with qRT-PCR or TBS microscopy alone, and critically, the genotypic analysis did not reveal associations between breakthrough infections that occurred in CIS43LS recipients and polymorphisms within or near the PfCSP epitope that CIS43LS targets.

We found that the aggregate 6-month CIS43LS efficacy estimates were similar in magnitude and precision when determined by the molFOI method and the standard HR method. However, when analyzed by month, the precision of efficacy estimates was higher with the molFOI method, and that precision increased with time relative to the HR method as trial participants experienced their first infections and were removed (censored) from the population at risk that contributed to the HR efficacy estimate. Notably, the lower bound of the 95% confidence interval for protective efficacy remained above 77% by molFOI as opposed to 3% by HR at month 6 in the 40 mg/kg CIS43LS arm, underscoring the ability of molFOI to assess efficacy with greater precision over the entire follow-up period, a feature that could have implications for future trial designs (e.g. smaller sample size) and for improving the accuracy of efficacy predictions beyond the follow-up period.

Importantly, by capturing total infection incidence with molFOI, and not censoring participants after their first infection, the size and risk profile of the study population included in the molFOI efficacy analysis was maintained throughout the follow-up period, a feature precluded by HR analysis that censors participants after their first infection, potentially introducing bias into efficacy estimates. For example, with HR analysis we found that censoring participants after their first infections resulted in more rapid depletion of participants who were infected at enrollment, a marker of higher infection risk. The depletion effect was relatively small in this trial and did not bias Eff_HR_ estimates. It will be important to assess the impact of depletion effects in future malaria intervention trials, particularly those involving children who have less acquired immunity to blood-stage malaria and are therefore more likely to experience patent parasitemia and symptomatic malaria earlier in a trial. Limited ability to control parasitemia (e.g., median 7,690 parasites/µl for first infections in 4-6 year-olds in the region (20)) may also strengthen the positive association between baseline infection risk and time to first detectable infection. This could accelerate the depletion of higher-risk individuals from less-protected study arms within a time-to-first infection analysis framework, possibly leading to biases in Eff_HR_ unobserved in the present study. As evidence of the potentially greater risk of encountering bias in Eff_HR_ in pediatric trials, a previous longitudinal study conducted at the same study site reports approximately two-fold higher molFOI in children and substantially greater inter-individual variability relative to that observed in this study (21).

Higher re-infection rates and greater heterogeneity in infection susceptibility may further reduce the extent to which time to first infection serves as a proxy for cumulative incidence over follow-up. These factors may have contributed to the observation of greater Eff_molFOI_ than Eff_HR_ (0.52 vs. 0.46) in a study of infants receiving RTS,S (22). Although the Eff_molFOI_ estimate was less precise, this may be because only the first TBS positive sample per child was genotyped in that study.

We also found that the efficacy estimate of CIS43LS against the mean number of infecting strains per exposure (Eff_FH_) was similar to the HR efficacy estimate, primarily because the mean number of strains at first infection was similar in the placebo and CIS43LS arms. Eff_FH_ was less precise than Eff_HR_ because Eff_FH_ accounts for uncertainty in estimating the mean strain count in the two arms. Previous analyses of two trials that evaluated genotyping endpoints in African infants receiving the RTS,S vaccine or a control vaccine estimated a lower strain count at first infection in RTS,S recipients (the mean ratio of strains at first infection in RTS,S versus control vaccine recipients was 0.88 (23) and 0.86 (22)). The accompanying statistical power analysis in the second study found that even with a mean ratio of 0.86, Eff_FH_ is only slightly more powerful than Eff_HR_ in some scenarios and is less powerful in others. In this study, the mean ratios near 1 may be because the treatments extend time to first infection, so treated participants become infected later in the trial when malaria transmission is higher. Thus, Eff_FH_ does not provide a clear advantage to Eff_HR_ although it requires more data.

The collective findings emerging from TBS microscopy (6), qRT-PCR (14), and molFOI-based analyses of this CIS43LS trial offer several lessons for the design of future malaria intervention trials. First, for interventions that aim to block sporozoite infection, qRT-PCR offers greatly enhanced sensitivity relative to TBS, and superior sensitivity to genotyping. Interpretive ambiguities related to the high sensitivity of qRT-PCR, for example distinguishing new infection from recrudescence following drug treatment of baseline parasitemia, may be clarified by genotyping (9). Second, the improved precision and potentially reduced bias in efficacy estimation offered by molFOI may warrant genotyping, particularly as new malaria interventions are deployed and clinical trials contend with measuring protection against the backdrop of an escalating standard of care. Third, genotyping only the first infection (the Follmann-Huang approach) is not advised, because the seasonal timing of first infections may differ systematically between study arms. Therefore, if a genotypic estimate of molFOI is planned, we recommend genotyping all infected samples collected during followup, ideally with infected samples first identified using qRT-PCR for cost-effectiveness. In summary, while molecular endpoints of infection such as qRT-PCR and genotyping carry additional costs beyond clinical disease or microscopy-based endpoints, they can uniquely inform whether candidate malaria interventions may support disease elimination by decreasing transmission risk, in addition to reducing clinical disease burden.

## MATERIALS AND METHODS

### Trial setting and temporal design

The double-blind, placebo-controlled, randomized phase 2 trial of CIS43LS (NCT04329104) involved healthy adults residing in the rural neighboring communities of Kalifabougou and Torodo, Mali, where *P. falciparum* transmission typically occurs from July through December (20). As described previously (6,14), 330 eligible adult participants were randomized and received a single intravenous (IV) infusion of normal saline placebo (n = 110) or CIS43LS at a dose of 10 mg/kg (n = 110) or 40 mg/kg (n = 110) between between May 6th and August 6th, 2021. All participants received a standard treatment course (AL) at enrollment, 7-15 days prior to study agent administration (‘day 0’). DBS and TBS sampling was scheduled for collection at enrollment and study days 0, 1, 3, 7, 14, 21, 28, 42, 56, 60, 84, 98, 112, 126, 140, 154, and 168 and during unscheduled illness visits when malaria was suspected. Follow-up visits were realized on exact scheduled dates in 30.2% of cases, with realized dates averaging 1.5 days before or after scheduled dates. In our analyses we therefore generally refer to the actual number of days elapsed since day 0 instead of the originally scheduled date. TBS microscopy was only conducted in real time when study participants exhibited signs or symptoms of malaria, and all participants who developed symptomatic malaria during the trial were provided standard treatment.

The trial followed the International Council for Harmonization Good Clinical Practice guidelines and applicable regulations in Mali. The participating communities granted permission to conduct the trial, and all participants provided written informed consent. The trial protocol and informed-consent forms were approved by the ethics committee at Faculté de Médecine et d’Odonto-Stomatologie and Faculté de Pharmacie at the University of Sciences, Techniques and Technologies of Bamako and by the national regulatory authorities of Mali.

Microscopy was performed for all TBS samples, while qRT-PCR testing was only performed on the subset of DBS needed to determine when the first qRT-PCR-detectable infection occurred. Because the qRT-PCR testing did not include all DBS samples, genotyping was triggered by TBS or qRT-PCR positivity at the timepoint of interest or before in the same person, and also included enrollment samples with positive or missing qRT-PCR.

Genotyping consisted of two separate AmpSeq assays: 1) an ‘Nterm’ PCR monoplex containing the CIS43LS epitope in the junctional region (JR) of PfCSP, and 2) a PCR multiplex called ‘SIMPLseq’ (24) which targets six highly polymorphic *P. falciparum* gene fragments. For both assays, we extracted total DNA from DBS using KingFisher Ready DNA Ultra 2.0 Prefilled Plates (ThermoFisher Scientific) and followed a standard nested PCR approach followed by paired-end sequencing on the Illumina MiSeq platform. We resolved microhaplotypes (alleles) from sequence output using DADA2 (v.1.14.0) (25) and the malaria amplicon processing pipeline available at https://github.com/broadinstitute/malaria-amplicon-pipeline.git. Details on sample inclusion, library preparation, and microhaplotype processing are provided in the Supplementary Materials.

To classify alleles as new or persistent at each detection timepoint per sample, we applied a Bayesian framework called DINEMITES (26). The model treats longitudinal allele observations as outcomes of latent infection states and estimates, via posterior inference, the probability that each observed allele reflects either persistence of an existing strain or acquisition of a new strain. For each participant timepoint, new allele presences inferred per locus are summarized into an expected number of newly acquired strains. DINEMITES also estimates the number of distinct infection events for each subject. An “infection event” is defined as transmission of malaria parasites from a mosquito to a human, potentially involving multiple strains, and is assigned to the sampling timepoint rather than the actual transmission time (expected to have occurred approximately 10 days earlier, based on the *P. falciparum* incubation period (27)). DINEMITES assignment of infection events accounts for the possibility that alleles introduced by a single infection become detectable over several visits due to imperfect detection and temporal variation in parasite density. At most one infection event is inferred per sampling timepoint, as multiple transmission events occurring within a sampling interval cannot be distinguished. DINEMITES included imputation for qRT-PCR positive samples with missing genotyping data; 50 augmented datasets were generated including expected new strain counts for each person-timepoint. See Supplementary Materials for further details about DINEMITES.

### Statistical analyses (molFOI modelling and efficacy estimation)

Efficacy estimates were calculated for five methods, described below, for each of the 50 augmented datasets and then combined using Rubin’s rules (28). The first two efficacy estimation methods are commonly applied and analyze only the time to first malaria infection, without incorporating data from later samples. The latter three methods incorporate information obtained from genotyping.

1. <u>Risk ratio (RR) efficacy</u>, denoted Eff_RR_, is estimated as one minus the risk ratio (1-RR), which is the CIS43LS-induced proportion reduction in probability of malaria infection during follow-up. In this analysis, malaria infection was defined as qRT-PCR positivity.
2. <u>Hazard ratio (HR) efficacy</u>, denoted Eff_HR_, against malaria infection is estimated as one minus the hazard ratio (1-HR), which is the CIS43LS-induced proportion reduction in hazard of malaria infection during follow-up. This measure incorporates information about the time to first infection but does not use information about later infections or number of strains at first infection. Eff_HR_ was calculated both for the hazard of qRT-PCR positivity, and for the hazard of TBS positivity during follow-up.
3. <u>Follmann-Huang (FH) efficacy</u>, denoted Eff_FH_, is defined as the CIS43LS-induced proportion reduction in number of infecting strains per exposure. In field trials, we do not observe exposure events of participants, so we cannot calculate mean strains per exposure. Follmann and Huang demonstrated that under randomization (which ensures similar exposure distributions in the study arms), we can estimate Eff_FH_ as 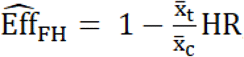 where 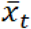 and 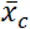 are mean numbers of strains for <u>observed infections</u> (rather than exposure events) in treated and controls, respectively, and HR is the hazard ratio for time to first observed infection obtained from a Cox model. The rationale for this is that a treatment that reduces the number of strains per exposure will convert some infections to non-infections (by converting exposure events with non-zero strains to exposure events with zero strains), which pushes back the time to first infection. This measure incorporates the time to first infection and number of strains present at the first infection. This is why, although the estimand is one minus the ratio of means, a valid estimator is one minus the HR times the ratio of truncated means, where the truncation excludes all exposures with zero infecting strains as these are unobservable.
4. <u>Molecular force of infection (molFOI) efficacy</u>, denoted Eff_molFOI_, is estimated as one minus the incidence of strains per unit time in treated vs. control participants. This measure incorporates information about the number of strains per participant acquired during follow-up but discards the timing information.
5. <u>Incidence rate ratio efficacy</u>, denoted Eff_IRR_, is estimated as one minus the incidence of new malaria infections determined by DINEMITES, where one “infection” may include multiple strains.

RR values were estimated by modified Poisson regression (29). In general, HR values and associated variance were estimated with Cox regression, except for the HR for time to first positive TBS, which was estimated with interval-censored survival analysis with a Weibull distribution to align with the primary analysis (6). FH estimates were calculated using the empirical mean number of strains at first infection and the HR estimated from Cox regression. molFOI was estimated by a Tweedie model with log-transformed total time-at-risk as an offset, using the glmmTMB() package (30,31) in R. Tweedie models are a class of models appropriate for random variables X that are discrete when X=0 and continuous when X>0 (32). Eff_IRR_ (where an infection may include multiple strains) was calculated analogously to molFOI.

To evaluate whether enrollment positivity is a proxy for exposure (23), Kaplan Meier curves for time to first infection were visualized by infection-status at enrollment (the visit prior to day 0). A similarly stratified analysis was performed for time from first infection to clinical malaria to assess whether participants with higher exposure have more natural immunity to symptoms. CIS43LS efficacy was calculated separately for participants who were positive vs. negative for infection at enrollment, and time-varying molFOI and time-varying efficacy (by both molFOI and HR methods) were visualized by enrollment infection-status. Additional details are in the Supplementary Materials.

Because censoring of participants after their first infections progressively depletes the sample size remaining under analysis, and may create non-random changes in the distribution of infection risk or other important participant traits between study arms, we used logistic regression with an interaction between treatment and a spline of time to visualize how the proportion of enrollment-positive participants remaining in the risk set varied over time by arm.

## Data Availability

The *P. falciparum* genotyping calls are available in Data file S3. All other data are available in the main text or the supplementary materials.

## List of Supplementary Materials

Supplementary Methods

Figs. S1 to S5

Tables S1 to S2

Data files S1 to S3

References *33*–*37*

## Acknowledgements

We thank the trial participants in Kalifabougou and Torodo, Mali, for their time and samples contributed to the clinical trial.

## Author contributions

AN, KK, AO, ACP, BT, RAS and PDC designed the clinical trial.

KK, AO, ACP, PDC, SD, and DD supervised and coordinated the clinical trial.

SL, MEP, and JS gathered and managed the clinical trial data.

SCM contributed the *P. falciparum* 18S rRNA qRT-PCR data.

PS, KAK, and JEA generated and analyzed the *P. falciparum* AmpSeq data, which DEN supervised.

PS, WN, GEP, VC, TMT, and AM analyzed and interpreted the data.

PS, VC, AM, GEP, PDC, and DEN conceptualized the paper.

PS, GEP, PDC, and DEN wrote the paper.

WN, VC, AM, SCM, and RAS reviewed and edited the paper.

All authors supported the review of the paper.

## Data and materials availability

*Please note that page numbering restarts below (Supplementary Materials section)*.

## Supplementary Materials

## Supplementary Methods

### Measurement schedules and methodology

#### Blood sample collection and *P. falciparum* detection by microscopy and qRT-PCR

The trial used finger prick blood (or venipuncture) samples to prepare Giemsa-stained TBS, as well as DBS on Protein Saver 903 DBS cards (Global Life Sciences Solutions). TBS were analyzed by two independent readers, and a third reader when discrepancies occurred. TBS were declared positive if two independent readers reported the presence of any number of *P falciparum* asexual parasites after counting 2500 leukocytes or examining 200 high-power fields. qRT-PCR targeting *P falciparum* 18S rRNA was performed on an Abbott m2000rtA module using 15 µl of extracted RNA. Absolute quantification was determined using an Armored RNA Calibrator encoding the full-length *P falciparum* 18S rRNA. While TBS was performed on all samples provided, qRT-PCR was performed on a subset sufficient to measure time-to-first-event. qRT-PCR was performed consecutively on DBS collected at all scheduled and unscheduled visits from enrollment through day 70 after study agent administration, except DBS collected on study day 1, which were not tested. For participants who did not have *P. falciparum* detected by qRT-PCR from days 28-70, qRT-PCR testing continued consecutively through all subsequent scheduled and unscheduled visits until either the last scheduled visit (s-day 168) or upon reaching a positive result of >100,000 parasites per milliliter of blood (p/ml) by qRT-PCR; 1.9% of all samples were not processed despite being designated for qRT-PCR (random error).

#### *falciparum* amplicon sequencing

We applied Illumina-based amplicon sequencing (AmpSeq) targeting *P. falciparum* to a pre-defined subset of enrollment and follow-up (≥day 0) DBS samples. This set included all DBS collected at enrollment and follow-up visits with a quantifiable (≥100 p/ml, i.e., ≥0.1 p/μl) *P. falciparum* qRT-PCR result or a *P. falciparum*-positive TBS. It also included DBS for which qRT-PCR or TBS had not been performed, including enrollment visits and all follow-up visits for which any previous follow-up visit from the same person had a quantifiable *P. falciparum* qRT-PCR result or a *P. falciparum*-positive TBS. Specifically, we screened qRT-PCR and TBS records for samples satisfying any of the criteria A-C:

A. follow-up visits with ≥0.1 p/μl by qRT-PCR and/or positive TBS
B. follow-up visits missing qRT-PCR data and with negative TBS but after any follow-up visit satisfying point A
C. enrollment visits with ≥0.1 p/μl by qRT-PCR, missing qRT-PCR, or positive TBS

There were 1083 records that met criteria A or B. Of these, 9 records did not have available DBS for sequencing (0106A-UV4, 0198K-UV1, 0236E-Day140, 0254B-UV1, 0261K-UV2, 0309S-UV3, 0329R-UV1, 0423C-Day21, 0423C-UV1), and 1 record occurred in duplicate (participant 0261K had two visits on the same day; of these, we sequenced only 0261K-Day56 and not 0261K-UV1). Thus, the final sequencing sample set for follow-up visits was 1073 from 186 participants (56.4% of total study population). There were 141 records that met criterion C. Of these, 7 records did not have available DBS for sequencing (0124W-Reenroll, 0270V-Enroll, 0255C-Enroll, 0254B-Enroll, 0184T-Reenroll, 0203R-Reenroll, 0394T-Enroll).

For both Nterm and SIMPLseq assays, we extracted total DNA from DBS via a KingFisher Flex instrument using KingFisher Ready DNA Ultra 2.0 Prefilled Plates (ThermoFisher Scientific). Both assays also followed a standard nested PCR approach (first round (PCR1) primers targeting the genomic regions of interest, and second round (PCR2) adding sample indexes and Illumina adapter components to the PCR1 products).

For Nterm assay PCR1, we used 3 µl DNA input, 5 µl 2x KAPA Master Mix (Roche), and 1 µl per 10 uM primer (p639_fw: 5′-TCGTCGGCAGCGTCAGATGTGTATAAGAGACAGGTCCATTACCTTGATTGTTTTTATTAG-3’ and p622_rv: 5′-GTCTCGTGGGCTCGGAGATGTGTATAAGAGACAGTGGGAAACAGGAAAATTGGT-3’), with an initial incubation step at 95 °C for 3 minutes (min) and touchdown PCR with denaturation at 98°C for 20 seconds (s), 20 s annealing between 61 °C to 56.5 °C (0.5 °C increments), and 72°C (15 s) extension.

Following touchdown, we used 20 standard amplification cycles at 98 °C (20 s), 56 °C (20 s), and 72 °C (15 s), and a final extension step at 72 °C (1 min). We diluted PCR1 products 1 to 9 in nuclease-free water prior to PCR2 (7 amplification cycles at 98 °C (20 s), 65 °C (30 s) and 72 °C (30 s) and a final extension step at 72 °C (1 min).

For SIMPLseq PCR1 (12 µl DNA input) and PCR2, we followed the default protocol available at https://www.protocols.io/view/simplseq-protocol-5qpvodd3zg4o/v2 (24). The assay targets short regions (188-320 base pairs (bp), excluding primer binding sites) within the genes encoding CSP (PF37_0304600), TRAP (PF3D7_1335900), SERA8 (PF3D7_0207300), SURFIN 4.2 (PF3D7_0424400), KELT (PF3D7_1475900) and WD-repeat containing protein (PF3D7_1410300).

For both SIMPLseq and Nterm assays, we purified PCR2 product pools using single-sided bead-based size selection (AMPure XP) before sequencing on the Illumina MiSeq platform (~384 samples per run) with 20% PhiX.

#### *P. falciparum* microhaplotypes and new vs. persistent allele calls

We resolved microhaplotypes (alleles) from sequence output using DADA2 (v.1.14.0), a denoising algorithm that tests whether nucleotide variation deviates from expected error distributions around abundant sequence types (25). We performed DADA2-denoising for SIMPLseq (overlapping paired-end reads) by running the malaria amplicon processing pipeline available at https://github.com/broadinstitute/malaria-amplicon-pipeline.git (default parameters). We subsequently masked polymorphisms that occurred in homopolymer tracts (≥ 5 bp) of the Pf3D7 reference sequence and filtered out microhaplotypes supported by < 5 read-pairs or by < 1% total read-pairs within a sample locus. For Nterm denoising (non-overlapping reads), we processed only the reverse read using standalone scripts. We rejected reads containing Nextera adapter sequence (cutadapt -b CTGTCTCTTATACACATCT --discard-trimmed) and then ran single-read DADA2 with a stricter expected error threshold (maxEE = 2) but without a subsequent homopolymer masking step. We additionally required Nterm reads to exhibit perfect PCR1 primer binding site matching to the Pf3D7 reference sequence, including also the first 5 bases adjacent to the primer (in contrast, the pipeline defaults used for SIMPLseq pre-processing allowed up to 2 mismatches (10% error tolerance) within the primer and did not filter based on adjacent bases; it also trims adapters, if present, instead of discarding such reads). We also filtered out Nterm microhaplotypes supported by <5 read-pairs or by <1% total read-pairs within a sample locus.

We used the DINEMITES (26) to classify alleles as new or persistent at each detection timepoint per sample. The framework explicitly accounts for polyclonal infections, allele sharing among strains, and stochastic allele detection, including allele imputation at qRT-PCR-positive visits for which genotyping was absent or incomplete. DINEMITES requires specifying two types of covariates: (1) “persistence covariates” that only affect the probability of observing continued prior infections and (2) “general covariates” that increase the probability of a new infection. Here, the persistence covariates used were whether an infection had been observed 30, 60, or 90 days prior, and whether treatment was given in the last 1-28 days or more than 28 days ago. The general covariates used were whether the visit fell in the rainy season and whether the visit was during the initial study phase (days 1-9 with more frequent visits), the mid study phase (days 10-35 with moderate frequency visits), or the late study phase. To determine the rainy season, precipitation data were fetched from the Open-Meteo ERA5 API (33) for the study location, and the rainy season for a given year was defined as the continuous window from the first day to the last day in that year where precipitation exceeded 1 mm. The DINEMITES process was run with 50 separate imputations (imputations for qRT-PCR-positive samples without sequencing data) to enable mean estimates and confidence intervals in molFOI-based analyses, as described in the next section.

For each participant and for each allele observed in the dataset, DINEMITES produces Bayesian estimates per timepoint (visit) for the probability that the allele is present (“p_present”) and for whether it is from a newly incident strain (“p_new”). For each of the 50 datasets containing these probabilities, we applied a counting procedure to consolidate multi-locus results (i.e., alleles detected at 7 different AmpSeq loci) into an estimate of the number of incident strains per participant-timepoint. Specifically, for each participant, timepoint, and genetic locus, we summed the product of p_new and p_present across alleles. Per participant-timepoint, this leads to seven “new_*time, locus*_” values, each indicating the newly incident strain count per AmpSeq locus. Taking the cumulative sum of new_*time, locus*_ over time and comparing sums across loci (an approach we call ‘sum-then-max’ (STM)) indicates which locus has shown the highest new strain signal up to any given timepoint. For each participant-timepoint, we retained only the new_*time, locus*_ value representing the locus with the highest cumulative new strain signal at the given timepoint. We used these pruned new_*time, locus*_ values (henceforth also termed ‘STM values’) as the single point estimates for the number of incident strains per participant-timepoint underlying molFOI analyses described in the next section.

We also used DINEMITES to identify distinct infection events for each subject. An “infection event” is defined as the transmission of malaria parasites from a mosquito to a human, possibly including multiple strains. We applied DINEMITES’ new_infection_bayes() calculation, which estimates whether novel allele detections at successive sampling visits are more likely to reflect a single infection event or multiple independent infection events. The estimation procedure accounts for the fact that alleles introduced by one infection may become detectable over several visits owing to imperfect detection and temporal variation in parasite densities. First, the p_new values of present alleles are summed within each visit, and the visits are partitioned over time into multi-visit chunks separated by local minima in the sums. Within each chunk whose largest per-visit sum reaches at least 1, the largest p_new value across the alleles and visits in that chunk (typically 1) is added to the infection event count. The maximum is used because a single infection is evidenced by how confidently at least one allele is new, without summing co-detected alleles from the same infection event. At each remaining visit in the chunk, the minimum p_new is added to the infection count, crediting additional infection events only to the extent that every allele at that visit appears new and thus avoiding re-counting alleles already attributed via the primary maximum or that were part of the same infection event but were missed in sequencing on the chunk’s maximum day. If no visit in a chunk has a p_new sum of at least 1, the chunk is treated as too weak for a clear infection event. For each visit in those chunks, the smallest allele p_new among alleles present is recorded, and the largest of those values across the chunk is added as a fractional residual.

We also used the p_new and p_present probabilities of the first DINEMITES dataset for allele persistence estimation and for escape-polymorphism analysis. For persistence estimation, using visits starting at day 8, we partitioned observations of recurrent alleles (i.e. alleles observed on ≥ 2 days within the same participant) into persistence intervals, with each observation at which p_new * p_present exceeded 0.9 initiating a new interval, i.e., persistence period. Analysis excluded participants with interrupted infections, i.e., participants who received AL treatment any time during follow-up. Possible differences in allele persistence between groups were assessed using Wilcoxon rank sum test. For escape-polymorphism analysis (CSP-JR amplicon), we again used the p_new * p_present > 0.9 and ≥ day 8 thresholds to identify newly acquired alleles. We then further classified newly acquired alleles based on whether or not they contained individual polymorphisms relative to 3D7. Analysis involved CSP-JR amplicon sequence starting 5 bp from the 3’ end of the left PCR1 primer (3D7 amino acid position 65) and continued to the end of the CSP epitope (position 115). Possible differences in polymorphism occurrence between groups was assessed statistically using Pearson’s Chi-squared test applied to counts of alleles with and without each polymorphism of interest.

### Statistical methods

#### The Follmann-Huang estimand and estimator

To define the Follmann-Huang estimand (17), the proportion reduction in mean strains per exposure, Let 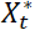 and 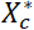 denote the number of strains from an exposure that develop into a blood-stage infection for treated participants and controls, respectively. Then Eff_FH_ is defined as:

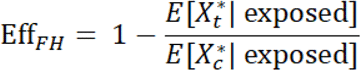

Naively, one would want to estimate this as one minus the ratio of empirical mean number of strains per exposure in each arm, but in field trials, we do not observe exposure events of participants. Follmann and Huang demonstrated that under randomization (which ensures similar exposure distributions in the two arms), we can estimate Eff_FH_ as 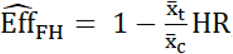 where 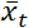 and 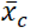 are mean numbers of strains for observed infections (rather than exposure events) in treated and controls, respectively, and HR is the hazard ratio for time to first observed infection obtained from a Cox model. The intuition for this is that a treatment that reduces the number of strains per exposure will convert some infections to non-infections (by converting exposure events with nonzero strains to exposure events with zero strains), which pushes back the time to first infection. For estimates of VE using the Follmann-Huang estimator, the standard error for *log(Δ)*was estimated as: 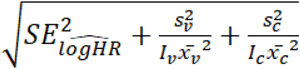, where 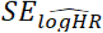 is the standard error of the log hazard ratio from Cox regression, 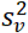 and 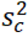 denote the variances of number of strains at first infection in treatment and control groups, respectively, and *I*_*v*_ and *I*_*c*_ denote numbers of treatment and control groups infected.

#### Analysis and modeling details

Analyses excluded two participants in the placebo group and one participant in the 40 mg/kg CIS43LS group who had no post-enrollment data. For all analyses, we started counting time at risk on day 8 after study-product administration to avoid counting pre-existing infections as new infections. Infection in FH and molFOI analyses was defined as the first instance where the expected number of incident strains exceeded 0.5.

Empirical molFOI estimates (calculated as the total number of strains per group, divided by the total time at risk per group) closely matched (+/-0.01) molFOI values estimated using a Tweedie distribution (log link) including an offset for log(time at risk) and treatment indicator variables. Variance estimates were from robust “sandwich” estimators.

Time-varying molFOI was modeled treating each person/timepoint as their own observation. We calculated time at risk (TAR) as the time between the sampling timepoint being modeled and the previous sampling timepoint for the same person. For example, a person with samples on Days 10, 28, and 78, would have TAR values 10, 18, and 50. We included a cubic spline for study day, as well as an interaction between a cubic spline of study day with treatment group in a Tweedie model. We used this fitted model to predict values for day 1-168 for each treatment group, and TAR of 1 year to calculate our point estimates. Variance estimates are from the predict() function.

We also calculated discretized monthly estimates of molFOI for each treatment group by summing the number of new strains in that month within that group and dividing by the sum of time at risk for all people with any data for that month/group. The time at risk by month was calculated for each person as their last follow-up day in that month minus the month start day (e.g. 1, 29, 57, etc.) plus 1. Follow-up month was calculated so that for each person, their first follow-up month is day 1-28 on study, second follow-up month is day 29-56, etc. Variance estimates were robust “sandwich” estimators and were estimated from a Tweedie model with a log link, with the total number of strains acquired over a month of follow-up per person as the response variable and the log-transformed total time-at-risk in that month as an offset. A separate Tweedie model was fit for each month independently. The calculation of VE_molFOI_ by follow-up month follows from the calculation of molFOI by study agent and follow-up month. VE_HR_ was calculated by follow-up month by splitting the data for each participant into 28-day intervals and including that stratification interval as an interaction with treatment in the Cox regression model to get a separate coefficient for each 28-day interval.

First infection for each participant for molFOI analyses was defined as the first instance where the expected number of incident strains exceeded 0.5 (on day 8 or later). All strains before the first infection were recoded to zero. For HR analyses, first infection was defined as the first positive qRT-PCR result (on day 8 or later) to align with the standard approach which does not incorporate genotyping information. Days were calculated to be relative to day 7 (for example, an infection on day 42 was recoded to 42-7 = 35).

To evaluate whether enrollment positivity is a proxy for exposure (23), monthly molFOI was calculated separately for participants who were infected or not at enrollment. Enrollment qRT-PCR positivity was defined using the visit prior to day 0. For subjects with multiple visits prior to antibody administration, the visit closest to antibody administration was used. Kaplan Meier curves for time-to-first infection were visualized by enrollment infection-status, and differences were assessed using a log-rank test, with infection defined as the first instance where the expected number of incident strains exceeded 0.5. A similarly stratified analysis was performed for time from first infection to clinical malaria to assess whether participants with higher exposure have more natural immunity to symptoms. The time of first clinical malaria was defined using the first date of antimalarial treatment, and anyone who had a date of antimalarial treatment was considered to have clinical malaria. For time from first infection to first clinical malaria, the infection start date was imputed to the midpoint between the first infection (expected number of incident strains > 0.5) and the prior negative. This analysis was restricted to those that had an infection, and then the start date (as defined above) became Day 1, and the time of clinical malaria (or end time, for those who did not have clinical malaria) was adjusted accordingly.

We also tested if CIS43LS efficacy differed by qRT-PCR-detected *P. falciparum* infection-status at enrollment by including an interaction between study arm assignment (CIS43LS or placebo) and enrollment infection-status in the model. Next, we extended this model to estimate time-varying molFOI and efficacy by including cubic splines for time and study arm by time interactions. Time was also treated discretely (as months) in analogous models estimating monthly Eff_molFOI_ and monthly Eff_HR_.

#### Multiple imputation

We averaged 50 imputed datasets and combined our estimates for all analyses using Rubin’s rules (28). Briefly, to obtain the pooled estimate of interest θ, we took the mean of the estimates across the 50 datasets.

We denote the estimate from each dataset as *θ*^*(i)*^for i=1, 2, …, M, where M is the number of datasets. For count data, this mean was rounded to the nearest integer.

The variance calculation takes both the within-imputation variance and the between-imputation variance into account. The within-imputation variance W was calculated as the mean of the variance from each imputed dataset. The between-imputation variance was defined as 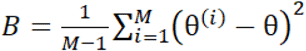. Then the overall variance estimate was 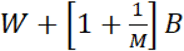.

The square root of the variance estimator was used as the standard error to calculate confidence intervals assuming normality, and p-values were calculated directly from the means and standard errors.

To calculate the p-value for the log-rank test using multiply imputed data using M=50 datasets (34,35), define the following:

- Let *R*_*M*_ denote the log rank test statistic for imputed data set M
- Let *E*_*M*_ denote the standard error of *R*_*M*_
- Let 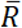 denote the average *R*_*M*_, so 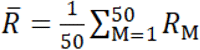
- Let *V*_*w*_ denote the within sample variance, 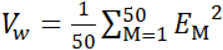
- Let *V*_*b*_ denote the between sample variance, 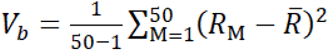
- Define r as 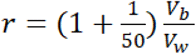
- Define t as t=50-1.

The log-rank test statistic is assumed to be normally distributed, and is calculated as:

(observed events in group 1 - expected events in group 1)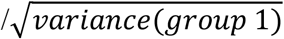. Under H_A_, this test statistic is normally distributed with variance 1 and mean equal to μ = log(HR) sqrt(D/4), where D is the total expected number of events in the two groups combined, and log(HR) is the log hazard ratio (10).

Under the null hypothesis that the two survival curves come from the same population, the below test statistic follows an F distribution with 1 and w degrees of freedom, where *w*=4+(*t*−4)(1+(1−2*t*^−1^)/*r*):

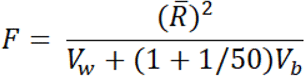

The p-value is the probability under the null hypothesis that the F statistic is greater than or equal to the observed value.

All analyses were performed in R version 4.5.2 (36), and p < 0.05 was considered significant.

## Supplementary Figures

**Fig. S1.**
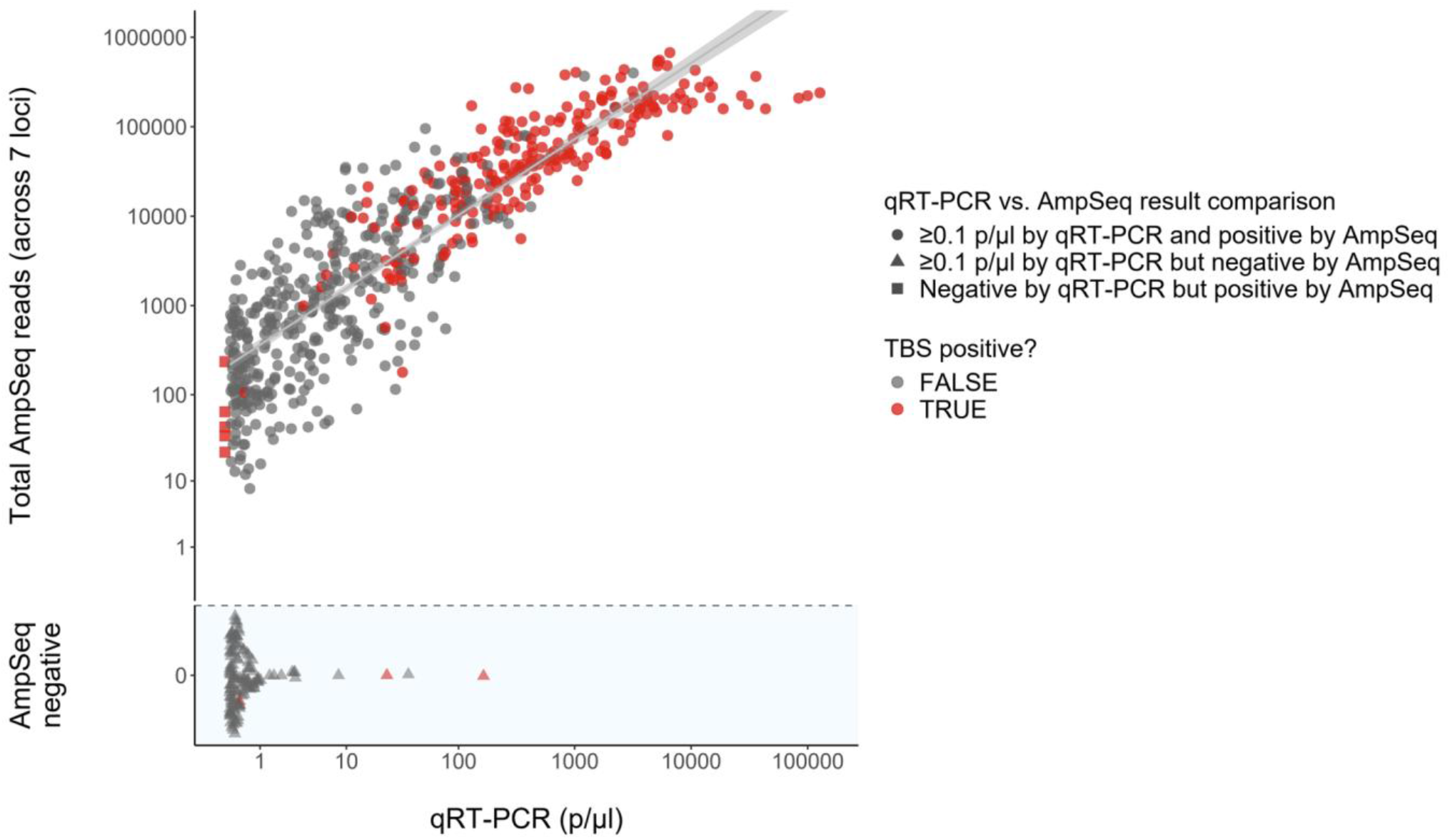
AmpSeq read signal for DBS with *P. falciparum* TBS positivity or qRT-PCR estimates at or above 0.1 *P. falciparum* parasites/µl. AmpSeq sensitivity per DBS sample is shown as total reads (y-axis, all 7 loci combined) with respect to parasitemia determined by qRT-PCR (continuous x-axis as opposed to binning used in Fig. 1B). AmpSeq-negative samples are shown separately at bottom with arbitrary vertical jitter. Point fill color indicates the result of TBS microscopy. Point shape categorizes samples by both AmpSeq and qRT-PCR result. Of 753 samples assayed by both AmpSeq and qRT-PCR, 603 (80.1%) had *P. falciparum* detected by both qRT-PCR and genotyping (any locus), whereas 13 samples were negative for *P. falciparum* by both methods. These 13 samples belonged to a set of 18 that underwent AmpSeq despite a negative qRT-PCR result because the TBS was positive. An additional 132 samples were negative by genotyping despite having ≥0.1 p/μl detected by qRT-PCR. Five qRT-PCR-negative samples were positive by genotyping, and each of these was positive by TBS. For samples with at least 10 p/µl, 99.2% (350 of 353) were successfully genotyped. For samples with 1 to 10 p/µl, 94.4% (151 of 160) samples were successfully genotyped, and for samples with 0.1 to 1 p/µl, the genotyping success rate was 45.9% (102 of 222).

**Fig. S2.**
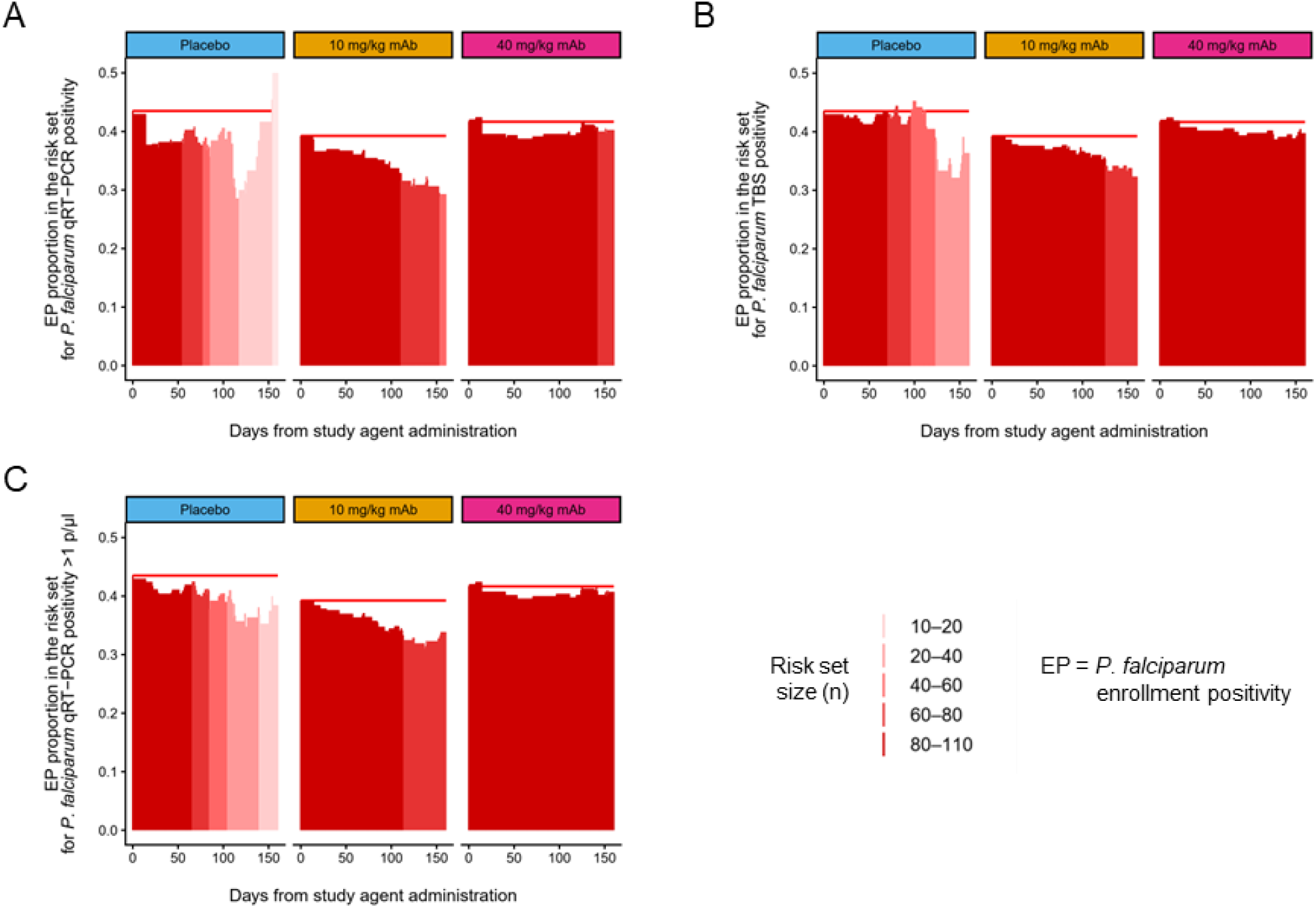
Change in the proportion of participants who had qRT-PCR-detected *P. falciparum* infection at enrollment over follow-up (survival analysis). (A) The proportion of *P. falciparum* enrollment-positive (EP) participants in the risk set for *P. falciparum* qRT-PCR positivity during follow-up per study arm. Red shading indicates sample size. (B) The proportion of EP participants in the risk set for *P. falciparum* TBS positivity during follow-up per study arm. Red shading indicates sample size. (C) The proportion of EP participants in the risk set for *P. falciparum* qRT-PCR positivity above 1 parasite/μl during follow-up per study arm (this endpoint is not used elsewhere in the work, only serving exploratory purposes here). Red shading indicates sample size.

**Fig. S3.**
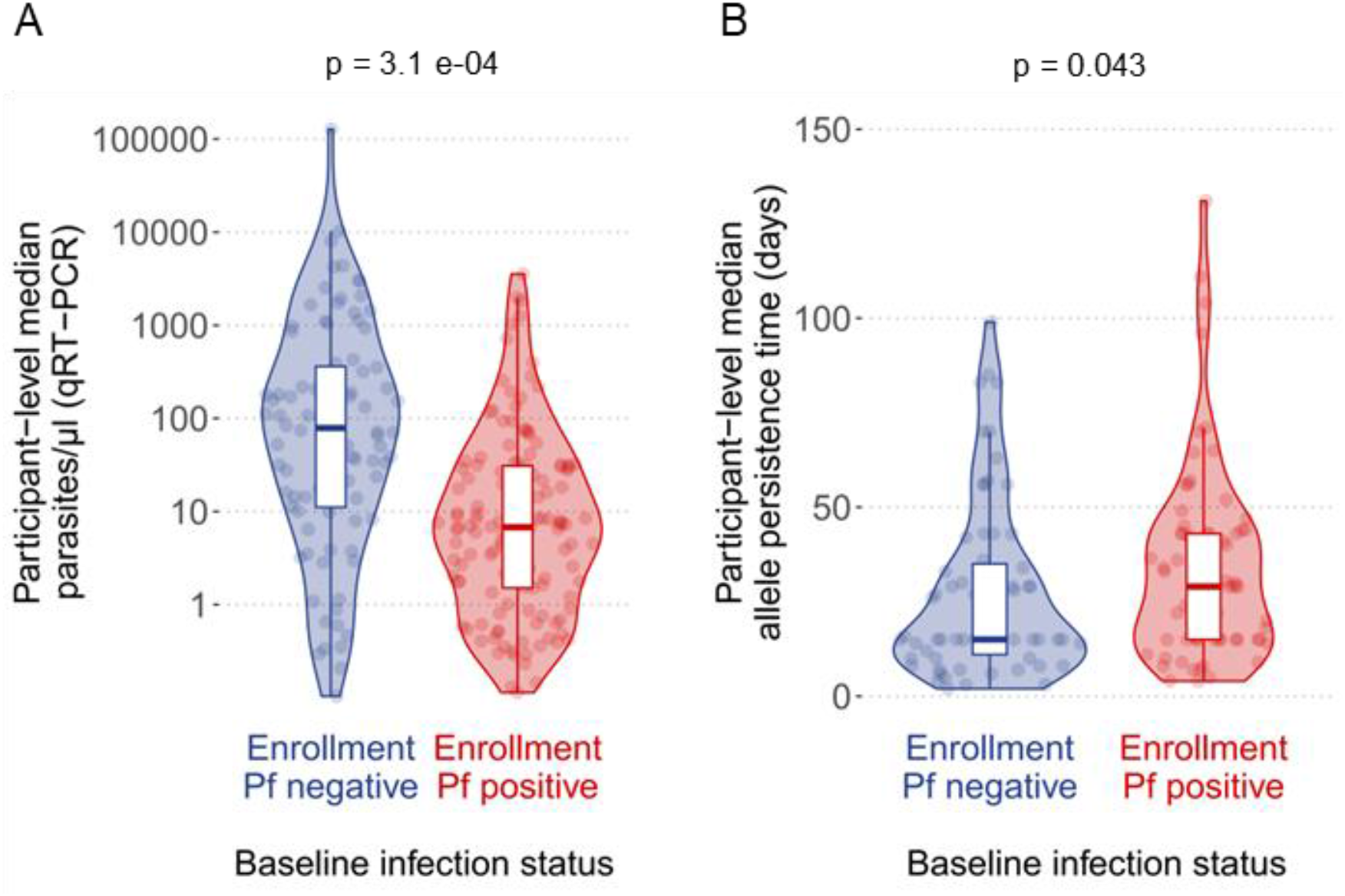
Relationship between baseline *P. falciparum* infection and infection traits (parasitemia, duration estimate) observed over follow-up. (A) Median *P. falciparum* parasitemia over follow-up per participant (points), grouped by whether or not *P. falciparum* infection was detected by qRT-PCR at enrollment. The figure pools observations across study arms, but a similar relationship occurs when restricting analysis to placebo (median = 15.4 parasites/μl for enrollment-positive participants vs. 135.0 parasites/μl for enrollment-negative participants, p = 0.045). (B) The length of allele persistence detected by genotyping over time, likewise grouped by whether or not *P. falciparum* infection was detected by qRT-PCR at enrollment. We assess only participants which never exhibited symptoms or signs of clinical malaria during follow-up and therefore never received antimalarial treatment during follow-up. Allele-persistence times thus approximate the duration of unmanipulated, asymptomatic blood-stage infection. The figure pools observations across study arms, but a similar relationship occurs when restricting analysis to placebo (median = 30.5 days for enrollment-positive participants vs. 16.0 days for enrollment-negative participants, p = 0.069).

**Fig. S4.**
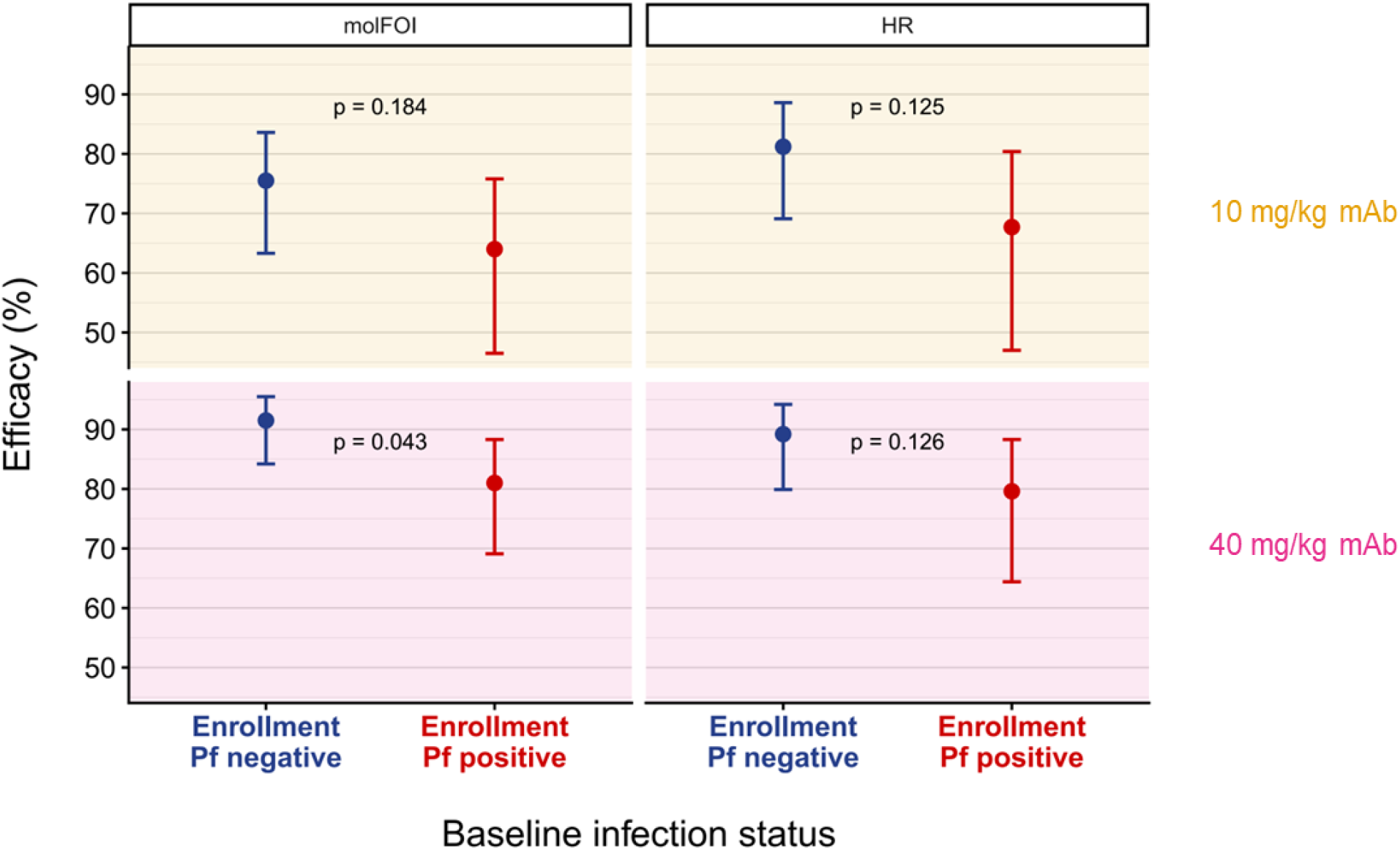
Relationship between baseline *P. falciparum* infection and composite CIS43LS efficacy by the molFOI and HR methods over the 6-month trial. Upper and lower panels represent efficacy results for 10 and 40 mg/kg CIS43LS, respectively. Left and right panels present the molFOI and HR (qRT-PCR positivity) methods, respectively. Vertical lines indicate 95% confidence intervals.

**Fig. S5.**
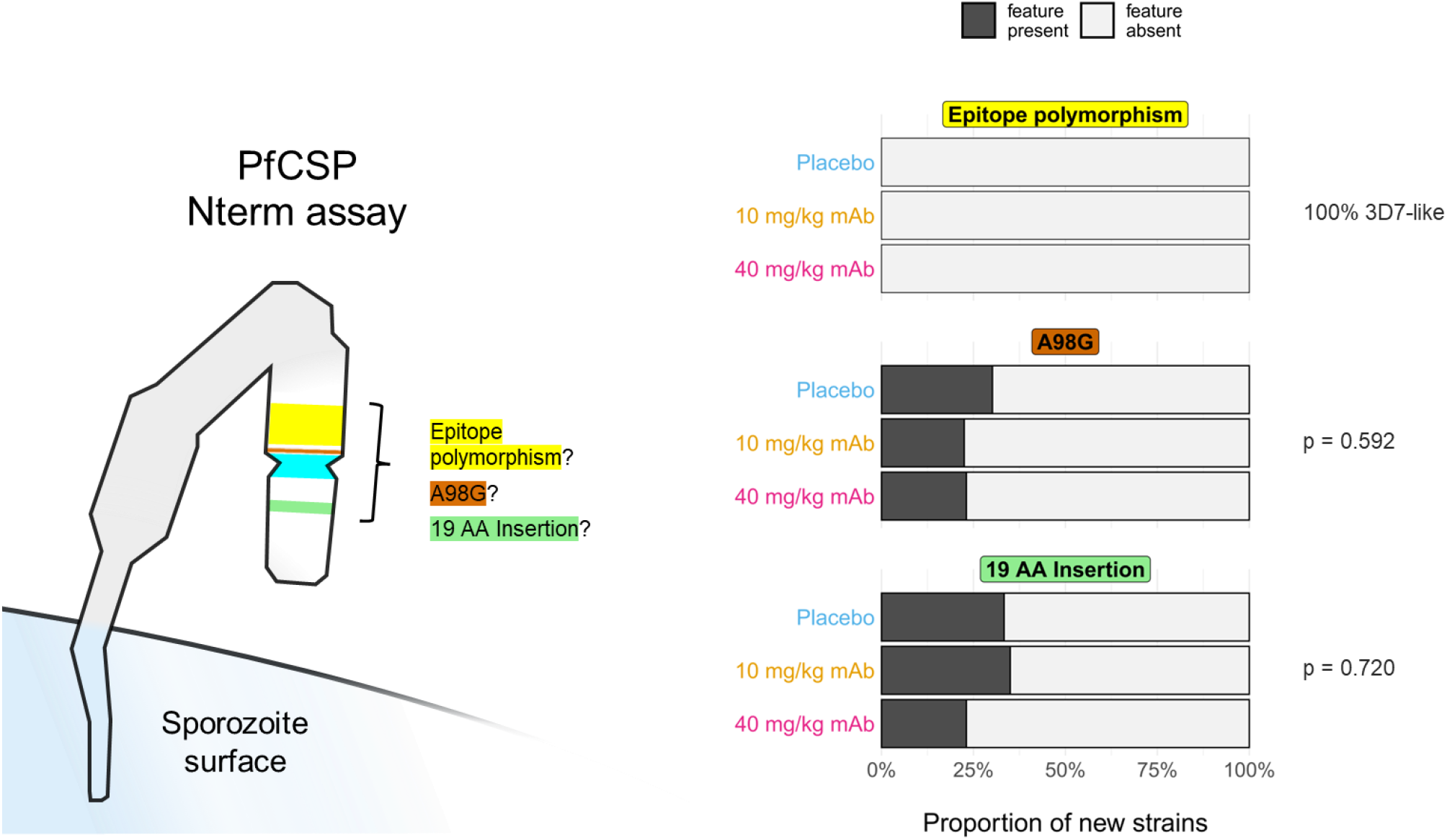
No evidence of escape-related polymorphism in PfCSP junctional region. As depicted at left, Nterm AmpSeq targets the PfCSP junctional region epitope targeted by CIS43LS (amino acid positions 101 to 115) but also encompasses up-stream sequence in which two polymorphisms occur: an Alanine to Glycine amino acid change at codon position 98 (A98G), and a 19 amino acid (AA) insertion between codon positions 80 and 81 (with respect to the Pf3D7 reference genome). The KLKQP motif, which undergoes essential proteolytic cleavage during hepatocyte invasion (37), is additionally shown in cyan. Stacked bars at right show – for newly acquired strains with AmpSeq signal at the Nterm locus (n 126 = for placebo, n = 40 for 10 mg/kg, n = 13 for 10 mg/kg) – the proportion which contained polymorphism in the epitope (none), A98G, or insertion at AA 80-81. A98G and insertion presence were not significantly different among arms (Chi-squared test).

## Supplementary Tables

**Table S1.**
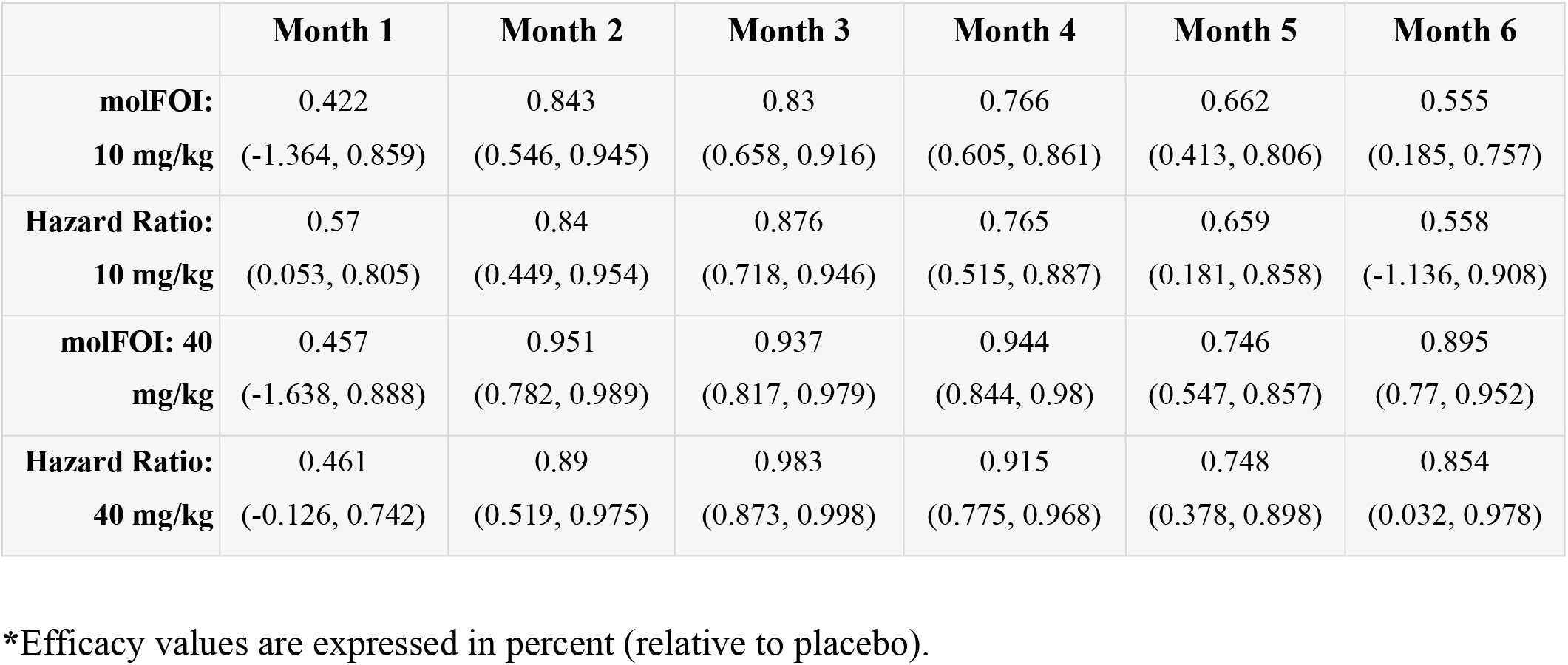
Monthly efficacy estimates by molFOI and Hazard Ratio.

|  | **Month 1** | **Month 2** | **Month 3** | **Month 4** | **Month 5** | **Month 6** |
|---|---|---|---|---|---|---|
| **molFOI:**<br>**10 mg/kg** | 0.422<br>(-1.364, 0.859) | 0.843<br>(0.546, 0.945) | 0.83<br>(0.658, 0.916) | 0.766<br>(0.605, 0.861) | 0.662<br>(0.413, 0.806) | 0.555<br>(0.185, 0.757) |
| **Hazard Ratio:**<br>**10 mg/kg** | 0.57<br>(0.053, 0.805) | 0.84<br>(0.449, 0.954) | 0.876<br>(0.718, 0.946) | 0.765<br>(0.515, 0.887) | 0.659<br>(0.181, 0.858) | 0.558<br>(-1.136, 0.908) |
| **molFOI: 40 mg/kg** | 0.457<br>(-1.638, 0.888) | 0.951<br>(0.782, 0.989) | 0.937<br>(0.817, 0.979) | 0.944<br>(0.844, 0.98) | 0.746<br>(0.547, 0.857) | 0.895<br>(0.77, 0.952) |
| **Hazard Ratio:**<br>**40 mg/kg** | 0.461<br>(-0.126, 0.742) | 0.89<br>(0.519, 0.975) | 0.983<br>(0.873, 0.998) | 0.915<br>(0.775, 0.968) | 0.748<br>(0.378, 0.898) | 0.854<br>(0.032, 0.978) |

\*Efficacy values are expressed in percent (relative to placebo).

**Table S2.** Aggregate efficacy estimates by molFOI and Hazard Ratio split by enrollment infection-status.

|  | **Enroll-negative** | **Enroll-positive** | **P-value** |
|---|---|---|---|
| **molFOI: 10 mg/kg** | 75.5 (63.3–83.6) | 64.0 (46.5–75.8) | 0.184 |
| **Hazard Ratio: 10 mg/kg** | 81.2 (69.1–88.6) | 67.7 (47.0–80.4) | 0.125 |
| **molFOI: 40 mg/kg** | 91.5 (84.2–95.5) | 81.0 (69.1–88.3) | 0.043 |
| **Hazard Ratio: 40 mg/kg** | 89.2 (79.9–94.2) | 79.6 (64.4–88.3) | 0.126 |

\*Efficacy values are expressed in percent (relative to placebo).

