## Supplementary material for "Genotypic infection endpoint analysis to understand efficacy and escape potential of the malaria monoclonal antibody CIS43LS": Data File S1: participant_0104X_with_imputation.pdf

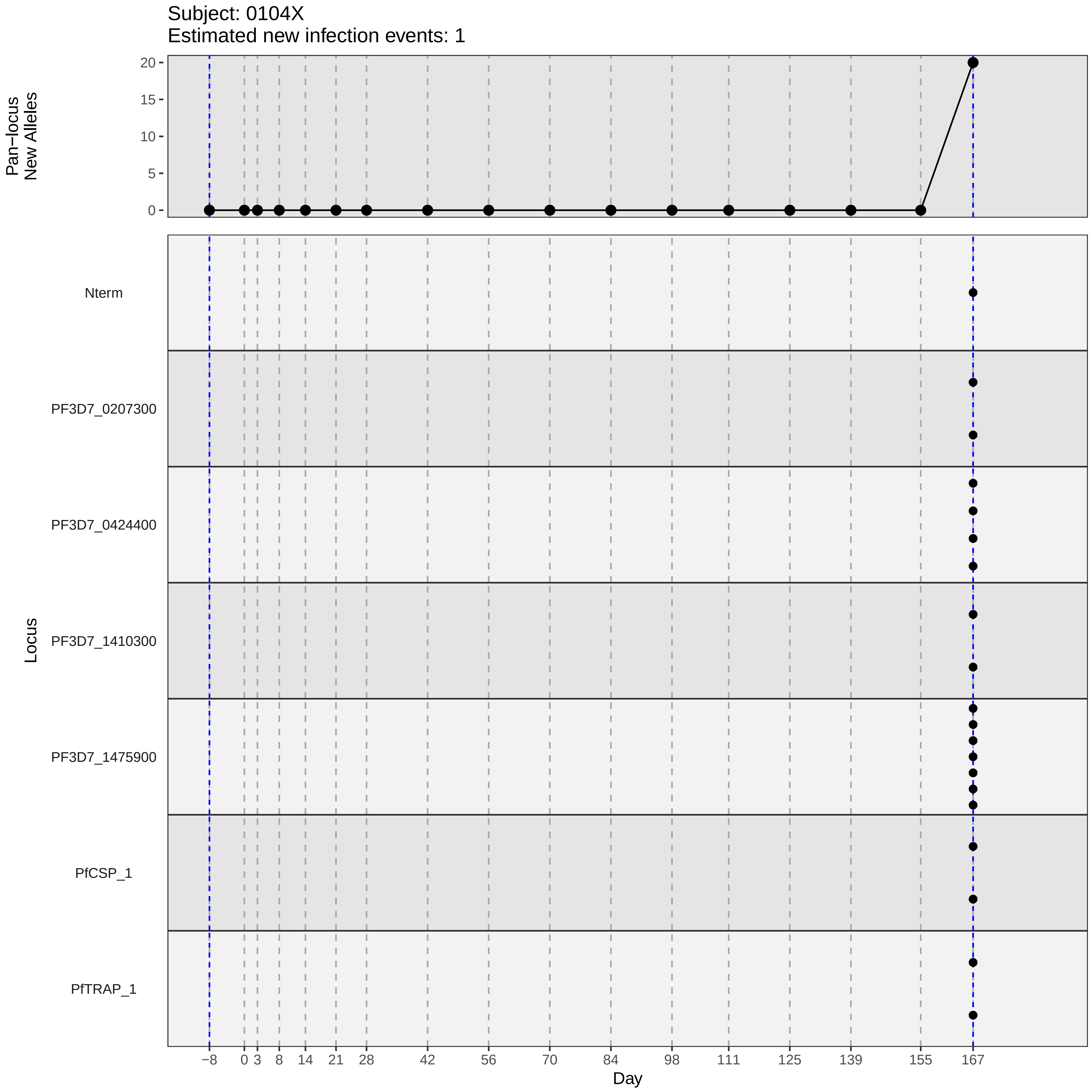
Subject: 0104X
Estimated new infection events: 1
Pan−locus
New Alleles
20
15
10
5
0
Locus
Nterm
PF3D7_0207300
PF3D7_0424400
PF3D7_1410300
PF3D7_1475900
PfCSP_1
PfTRAP_1
−8
0
3
8
14
21
28
42
56
70
84
98
111
125
139
155
167
Day
