## Supplementary material for "Genotypic infection endpoint analysis to understand efficacy and escape potential of the malaria monoclonal antibody CIS43LS": Data File S1: participant_0112G_with_imputation.pdf

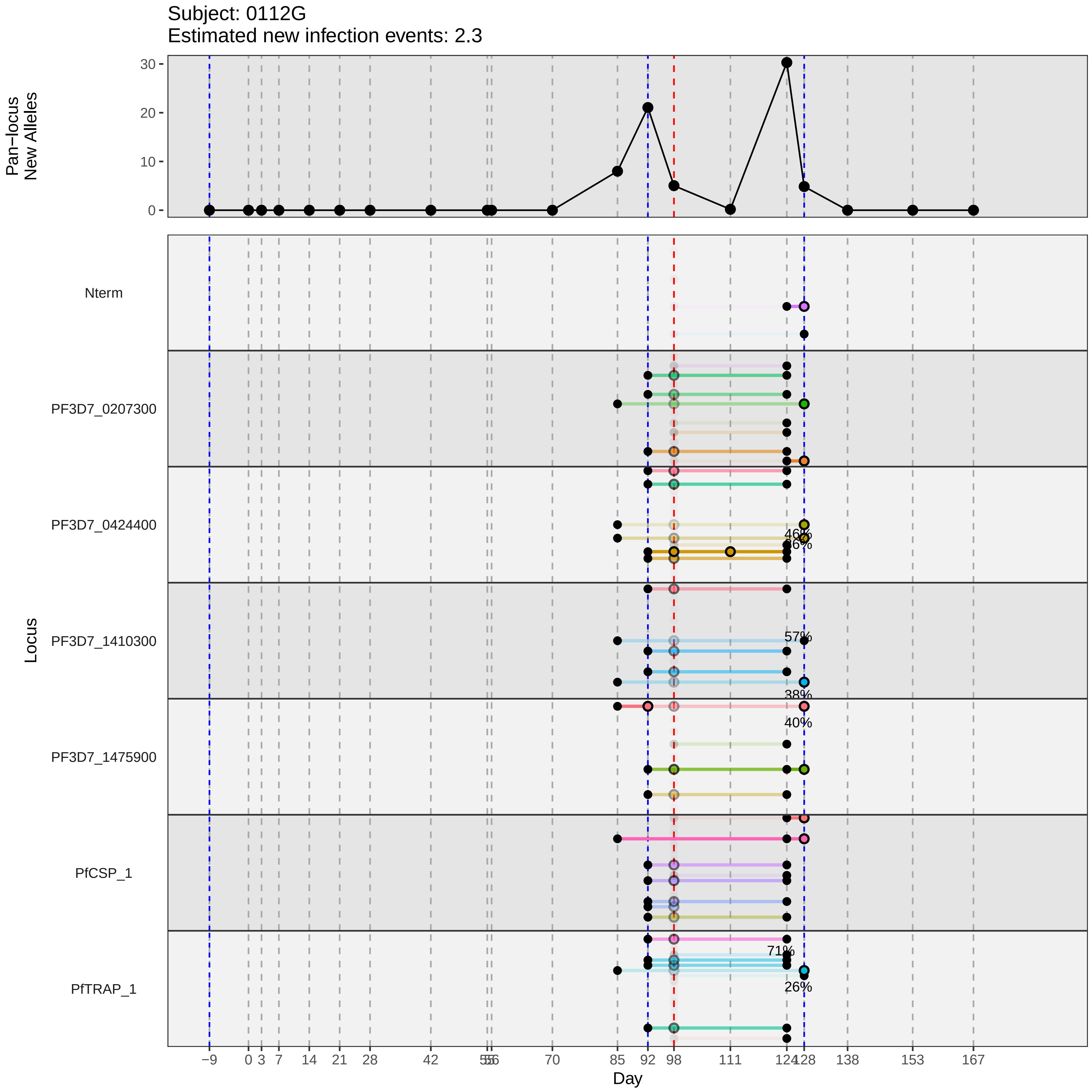
Subject: 0112G
Estimated new infection events: 2.3
Pan−locus
New Alleles
30
20
10
0
Locus
Nterm
PF3D7_0207300
PF3D7_0424400
PF3D7_1410300
PF3D7_1475900
PfCSP_1
PfTRAP_1
46%
26%
57%
38%
40%
71%
26%
−9
0
3
7
14
21
28
42
55
56
70
85
92
98
111
124
128
138
153
167
Day
