## Supplementary material for "Genotypic infection endpoint analysis to understand efficacy and escape potential of the malaria monoclonal antibody CIS43LS": Data File S1: participant_0121T_with_imputation.pdf

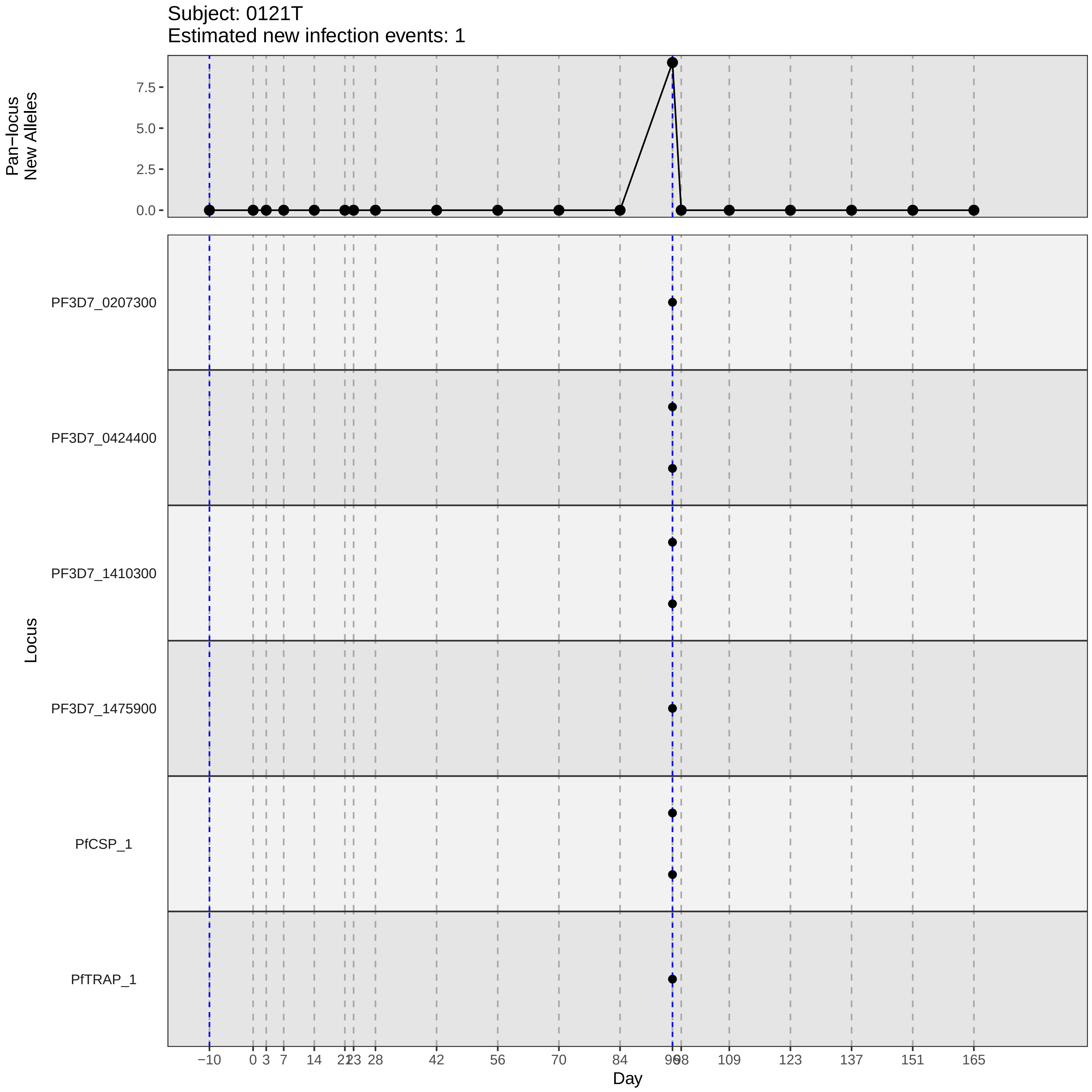
Subject: 0121T
Estimated new infection events: 1
Pan−locus
New Alleles
7.5
5.0
2.5
0.0
Locus
PF3D7_0207300
PF3D7_0424400
PF3D7_1410300
PF3D7_1475900
PfCSP_1
PfTRAP_1
−10
0
3
7
14
21
23
28
42
56
70
84
96
98
109
123
137
151
165
Day
