## Supplementary material for "Genotypic infection endpoint analysis to understand efficacy and escape potential of the malaria monoclonal antibody CIS43LS": Data File S1: participant_0126Y_with_imputation.pdf

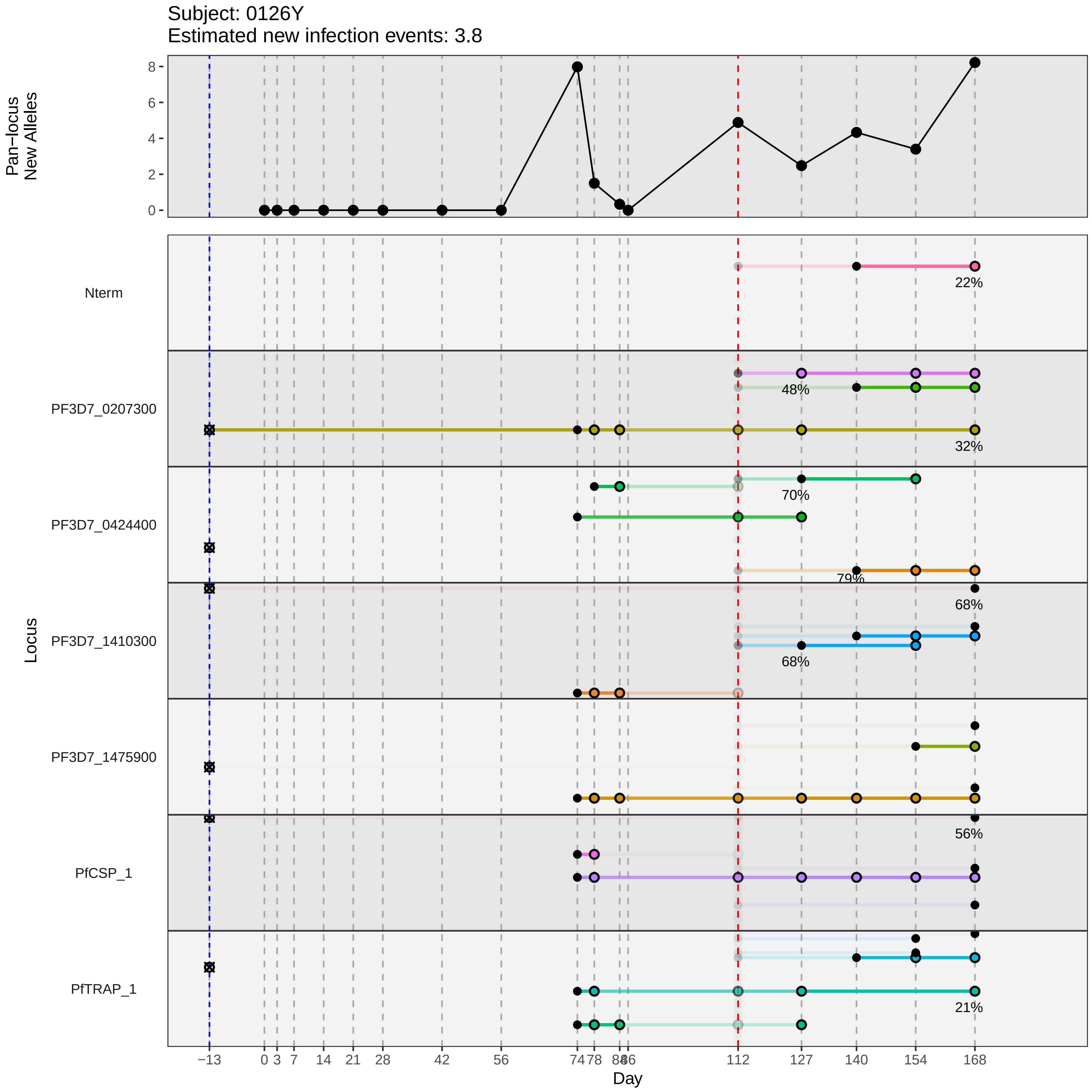

Subject: 0126Y
Estimated new infection events: 3.8
Pan−locus
New Alleles
0
2
4
6
8
Locus
Nterm
PF3D7_0207300
PF3D7_0424400
PF3D7_1410300
PF3D7_1475900
PfCSP_1
PfTRAP_1
22%
48%
32%
70%
79%
68%
68%
56%
21%
−13
0
3
7
14
21
28
42
56
74
78
112
127
140
154
168
Day
