## Supplementary material for "Genotypic infection endpoint analysis to understand efficacy and escape potential of the malaria monoclonal antibody CIS43LS": Data File S1: participant_0132F_with_imputation.pdf

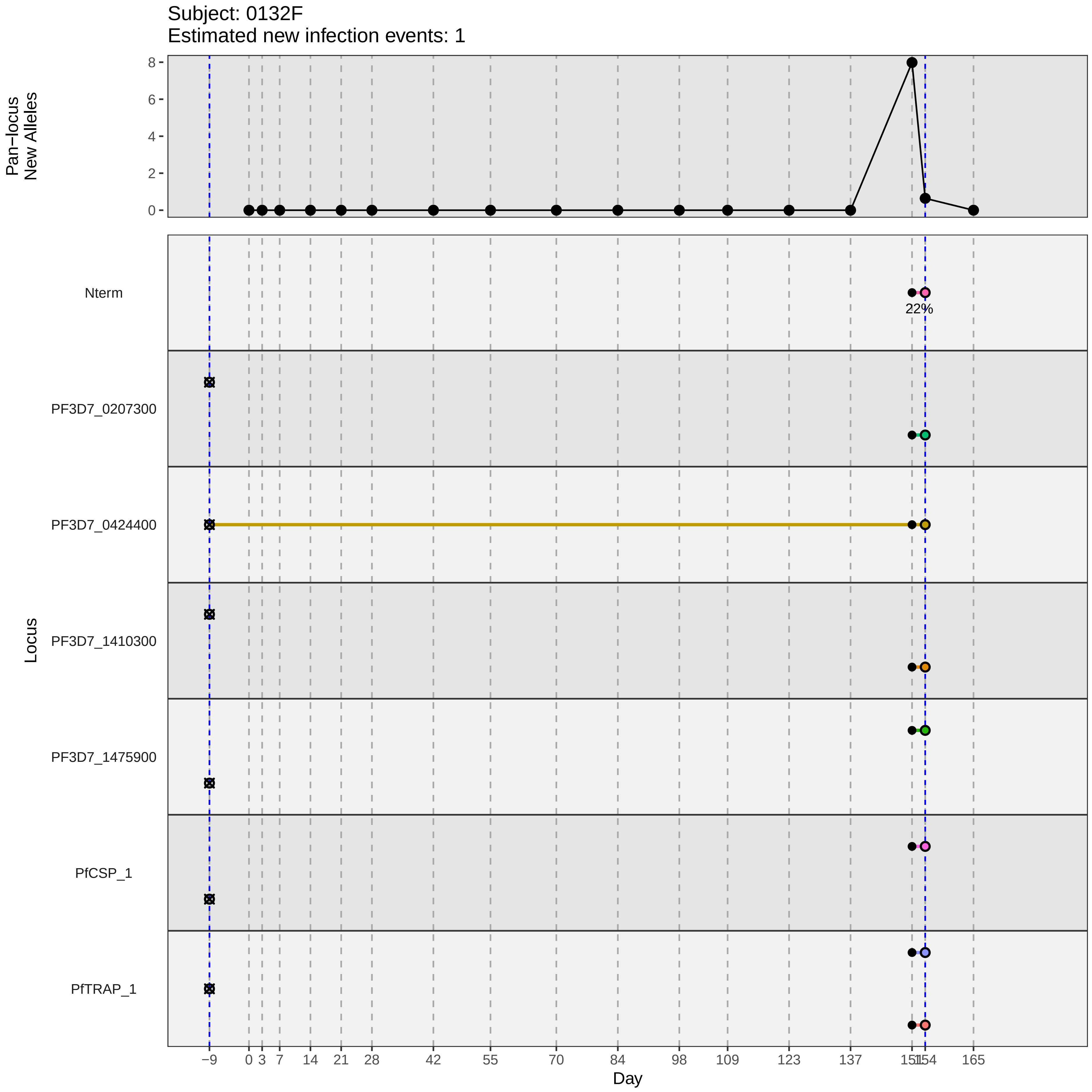
Subject: 0132F
Estimated new infection events: 1
Pan−locus New Alleles
Locus
Nterm
22%
PF3D7_0207300
PF3D7_0424400
PF3D7_1410300
PF3D7_1475900
PfCSP_1
PfTRAP_1
Day
−9
0
3
7
14
21
28
42
55
70
84
98
109
123
137
151
154
165
