## Supplementary material for "Genotypic infection endpoint analysis to understand efficacy and escape potential of the malaria monoclonal antibody CIS43LS": Data File S1: participant_0143U_with_imputation.pdf

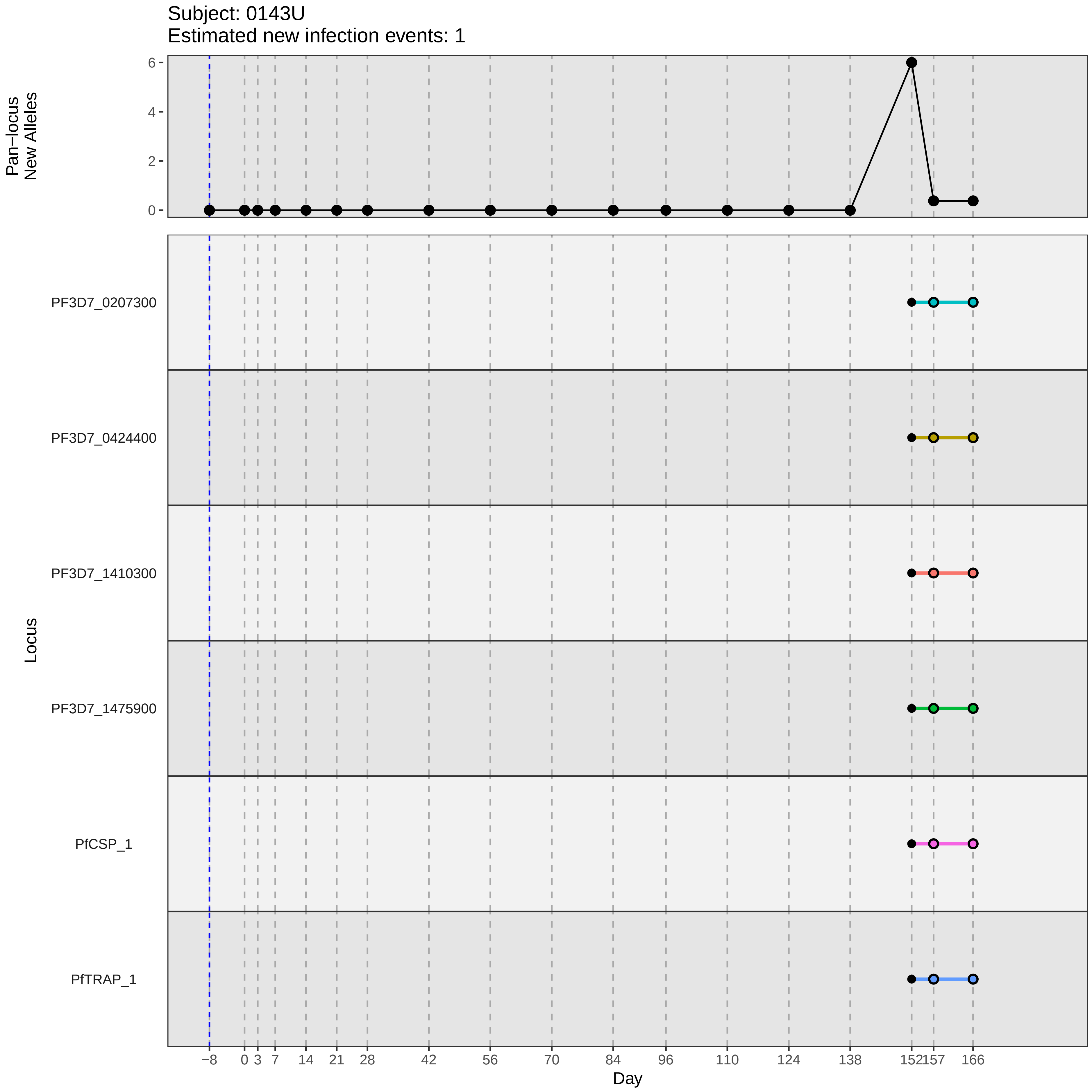
Subject: 0143U
Estimated new infection events: 1
Pan−locus
New Alleles
6
4
2
0
Locus
PF3D7_0207300
PF3D7_0424400
PF3D7_1410300
PF3D7_1475900
PfCSP_1
PfTRAP_1
−8
0
3
7
14
21
28
42
56
70
84
96
110
124
138
152
157
166
Day
