## Supplementary material for "Genotypic infection endpoint analysis to understand efficacy and escape potential of the malaria monoclonal antibody CIS43LS": Data File S1: participant_0145W_with_imputation.pdf

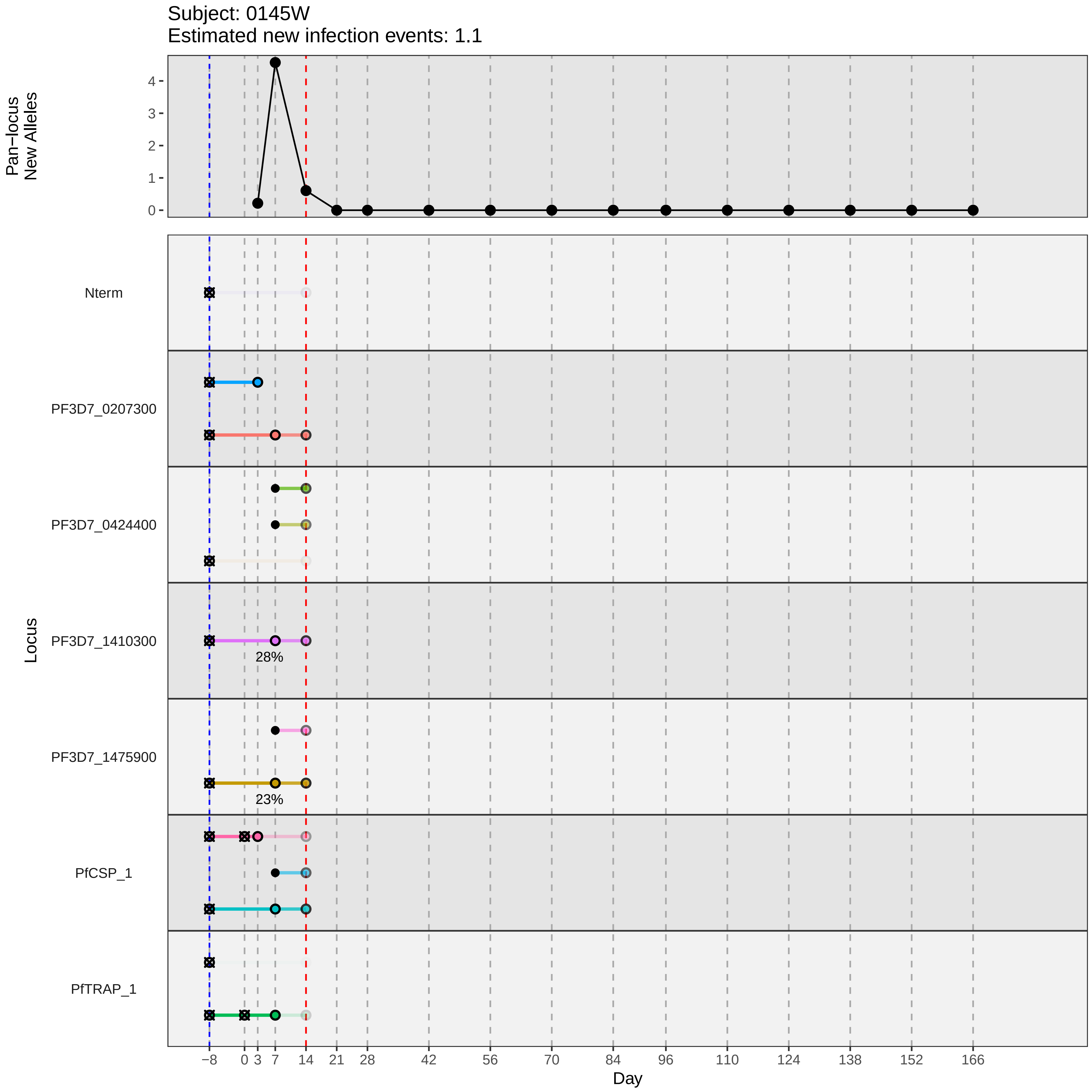

Subject: 0145W
Estimated new infection events: 1.1
Pan−locus
New Alleles
4
3
2
1
0
Locus
Nterm
PF3D7_0207300
PF3D7_0424400
PF3D7_1410300
28%
PF3D7_1475900
23%
PfCSP_1
PfTRAP_1
−8
0
3
7
14
21
28
42
56
70
84
96
110
124
138
152
166
Day
