## Supplementary material for "Genotypic infection endpoint analysis to understand efficacy and escape potential of the malaria monoclonal antibody CIS43LS": Data File S1: participant_0146X_with_imputation.pdf

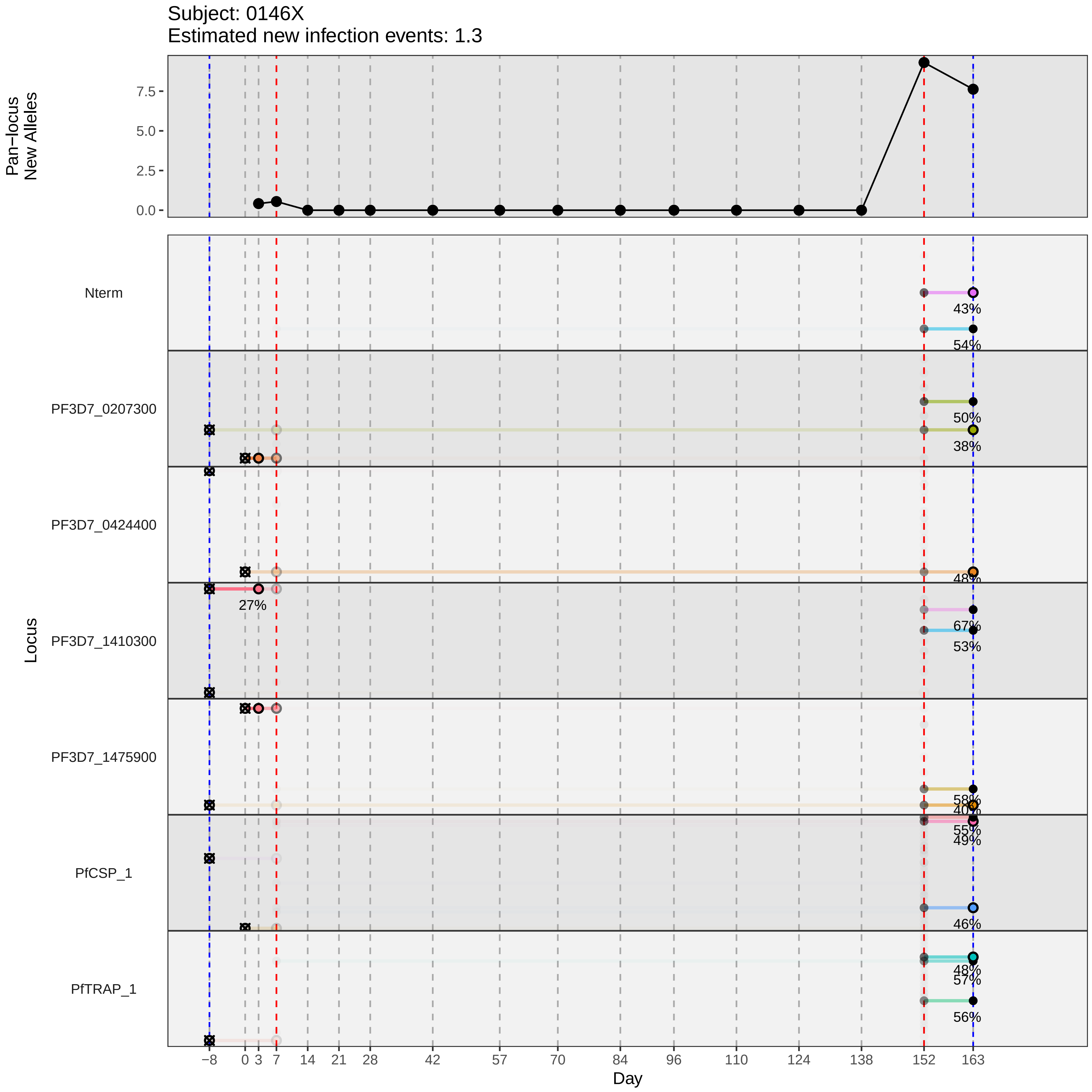

Subject: 0146X
Estimated new infection events: 1.3
Pan−locus New Alleles
7.5
5.0
2.5
0.0
Locus
Nterm
PF3D7_0207300
PF3D7_0424400
PF3D7_1410300
PF3D7_1475900
PfCSP_1
PfTRAP_1
43%
54%
50%
38%
48%
27%
67%
53%
58%
40%
55%
49%
46%
48%
57%
56%
−8
0
3
7
14
21
28
42
57
70
84
96
110
124
138
152
163
Day
