## Supplementary material for "Genotypic infection endpoint analysis to understand efficacy and escape potential of the malaria monoclonal antibody CIS43LS": Data File S1: participant_0152E_with_imputation.pdf

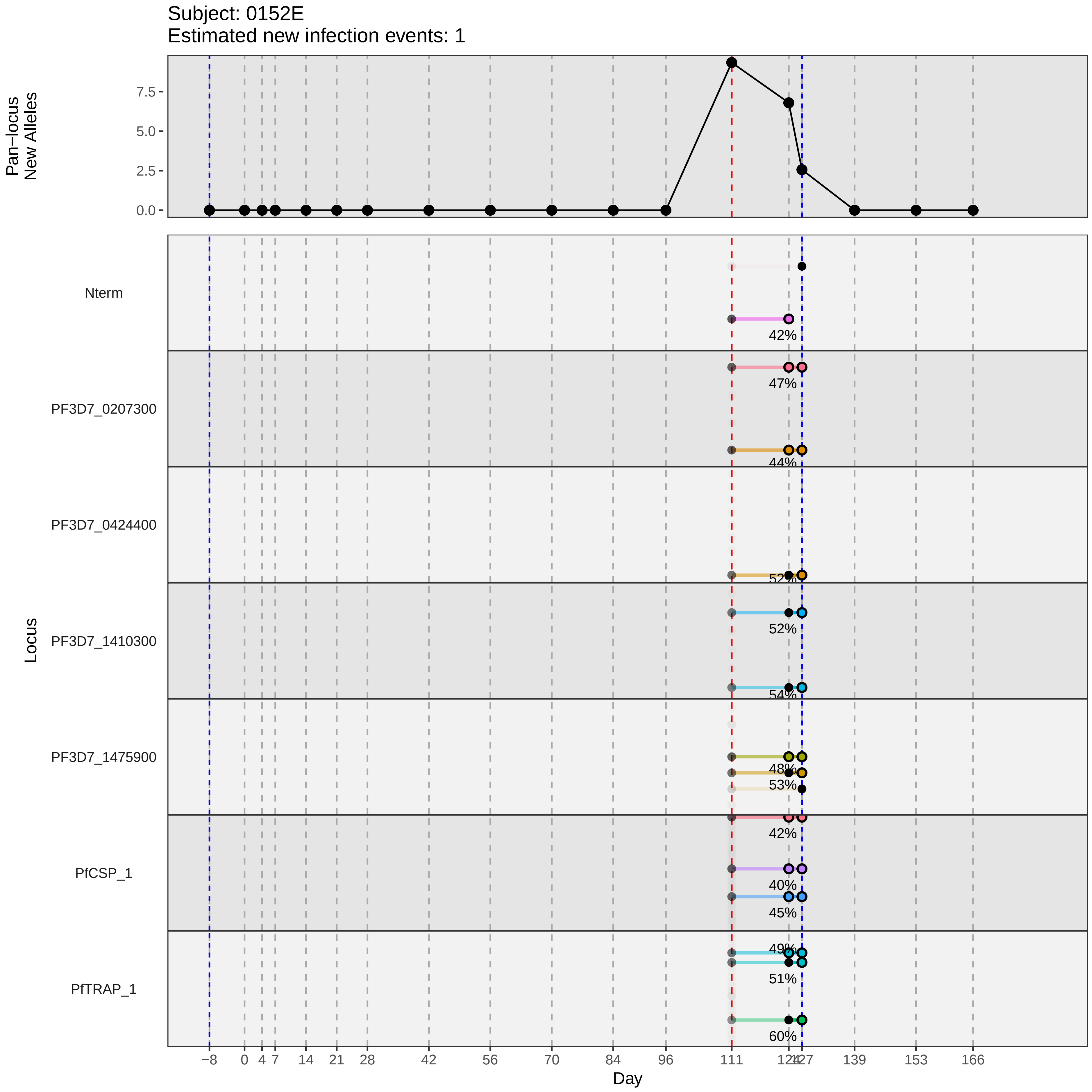
Subject: 0152E
Estimated new infection events: 1
Pan−locus New Alleles
7.5
5.0
2.5
0.0
Locus
Nterm
PF3D7_0207300
PF3D7_0424400
PF3D7_1410300
PF3D7_1475900
PfCSP_1
PfTRAP_1
42%
47%
44%
52%
52%
54%
48%
53%
42%
40%
45%
49%
51%
60%
−8
0
4
7
14
21
28
42
56
70
84
96
111
124
127
139
153
166
Day
