## Supplementary material for "Genotypic infection endpoint analysis to understand efficacy and escape potential of the malaria monoclonal antibody CIS43LS": Data File S1: participant_0159P_with_imputation.pdf

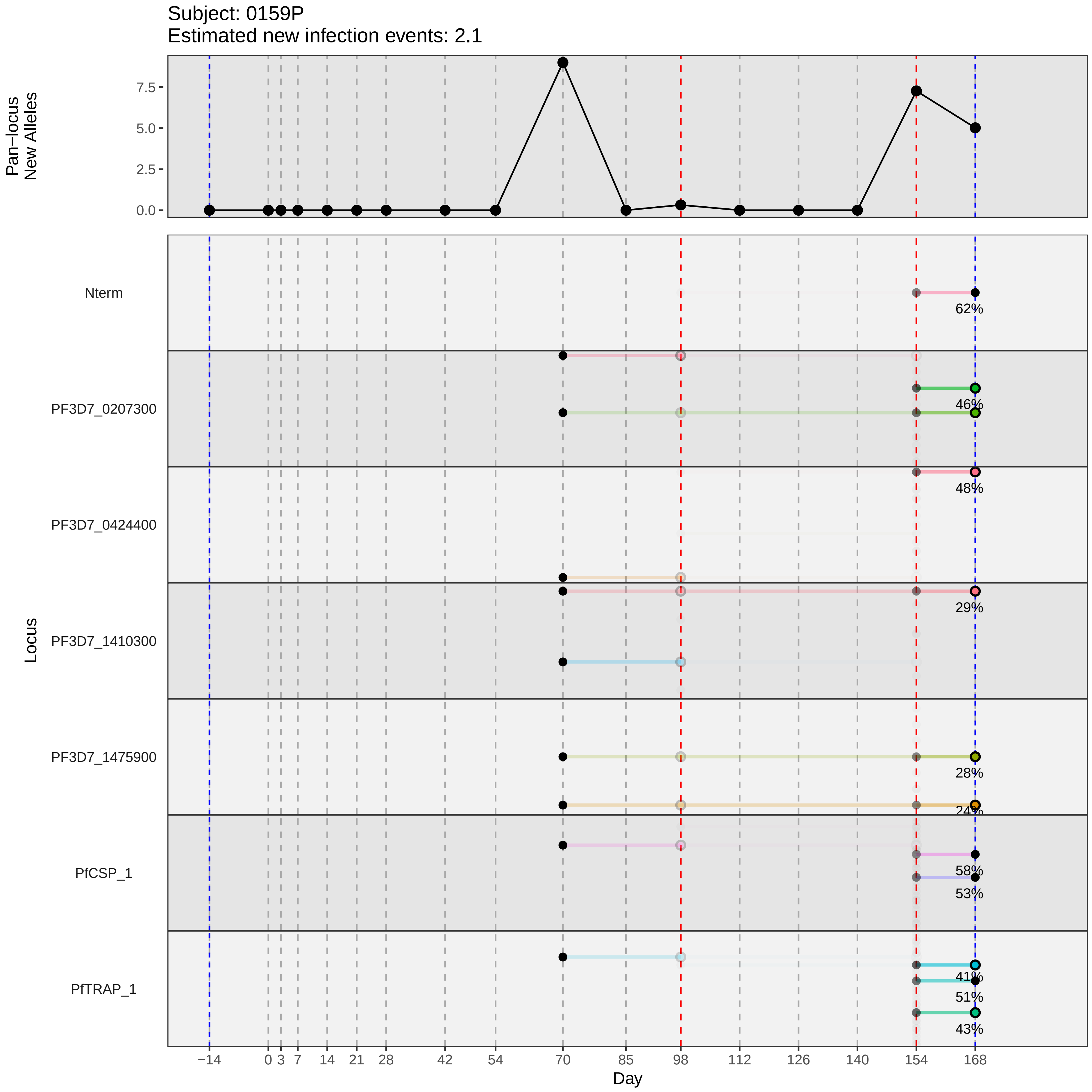

Subject: 0159P
Estimated new infection events: 2.1
Pan−locus New Alleles
0.0
2.5
5.0
7.5
Locus
Nterm
PF3D7_0207300
PF3D7_0424400
PF3D7_1410300
PF3D7_1475900
PfCSP_1
PfTRAP_1
62%
46%
48%
29%
28%
24%
58%
53%
41%
51%
43%
−14
0
3
7
14
21
28
42
54
70
85
98
112
126
140
154
168
Day
