## Supplementary material for "Genotypic infection endpoint analysis to understand efficacy and escape potential of the malaria monoclonal antibody CIS43LS": Data File S1: participant_0162S_with_imputation.pdf

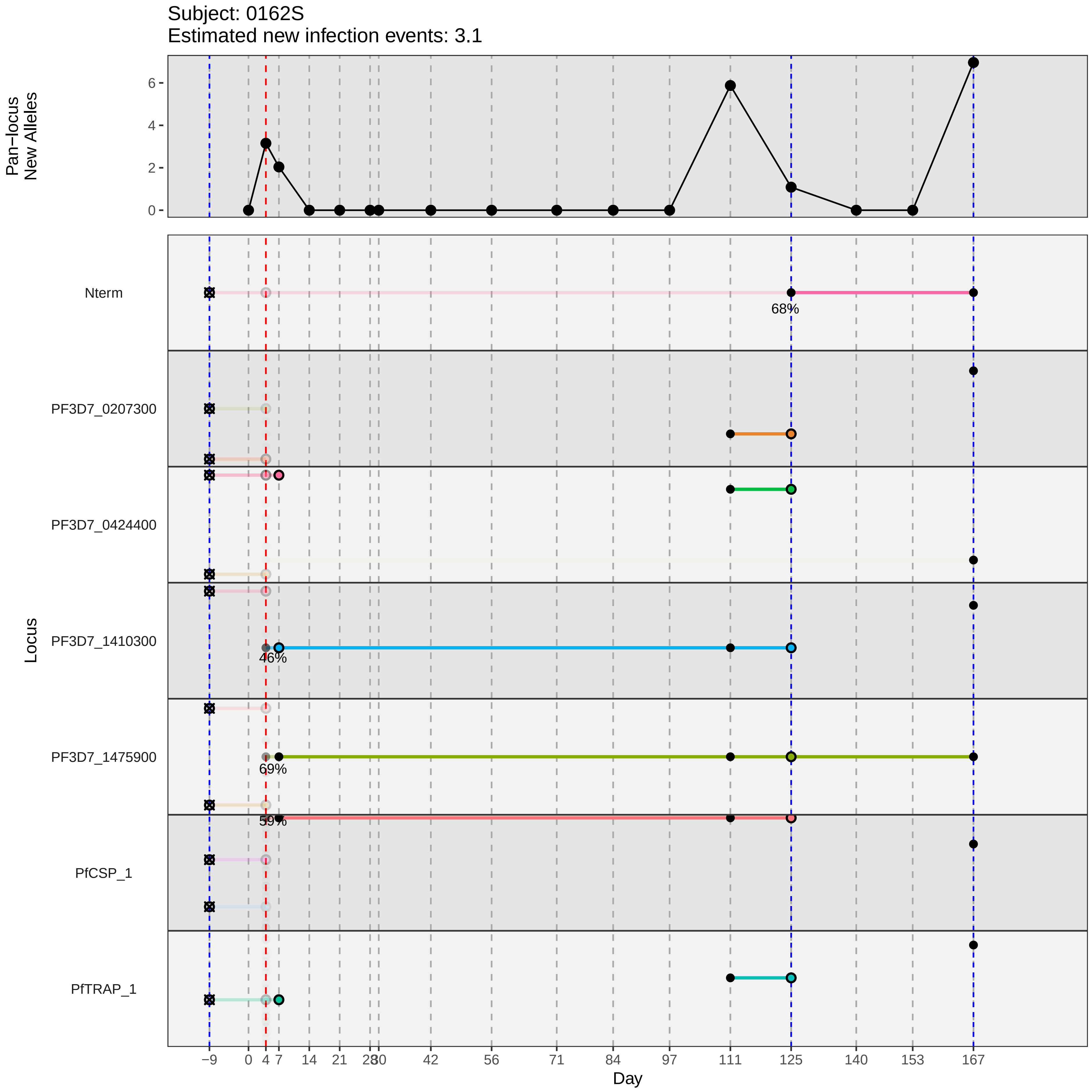
Subject: 0162S
Estimated new infection events: 3.1
Pan−locus New Alleles
Locus
Nterm
PF3D7_0207300
PF3D7_0424400
PF3D7_1410300
PF3D7_1475900
PfCSP_1
PfTRAP_1
68%
46%
69%
59%
Day
−9
0
4
7
14
21
28
30
42
56
71
84
97
111
125
140
153
167
0
2
4
6
