## Supplementary material for "Genotypic infection endpoint analysis to understand efficacy and escape potential of the malaria monoclonal antibody CIS43LS": Data File S1: participant_0167X_with_imputation.pdf

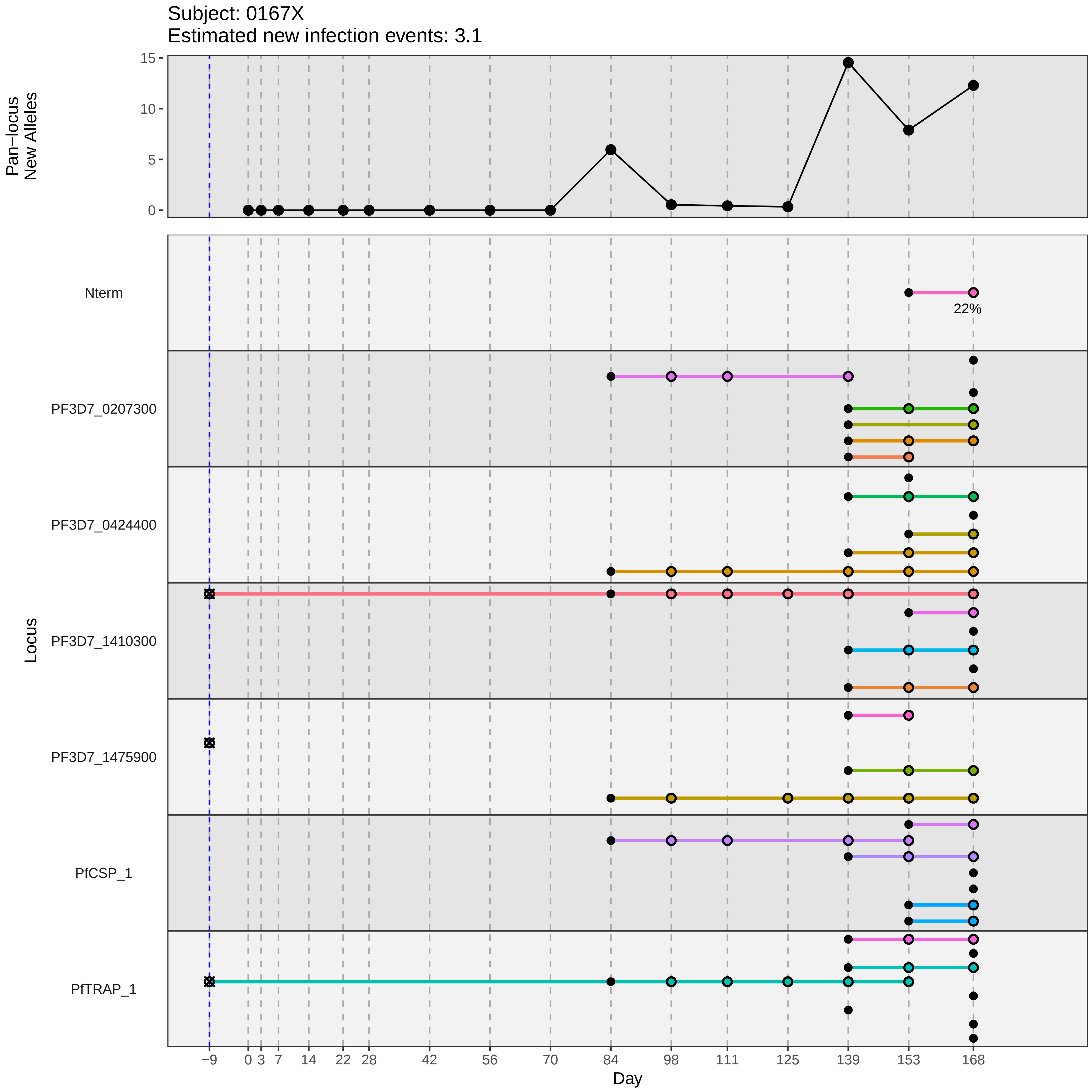

Subject: 0167X
Estimated new infection events: 3.1
Pan−locus New Alleles
15
10
5
0
Locus
Nterm
22%
PF3D7_0207300
PF3D7_0424400
PF3D7_1410300
PF3D7_1475900
PfCSP_1
PfTRAP_1
−9
0
3
7
14
22
28
42
56
70
84
98
111
125
139
153
168
Day
