## Supplementary material for "Genotypic infection endpoint analysis to understand efficacy and escape potential of the malaria monoclonal antibody CIS43LS": Data File S1: participant_0168Y_with_imputation.pdf

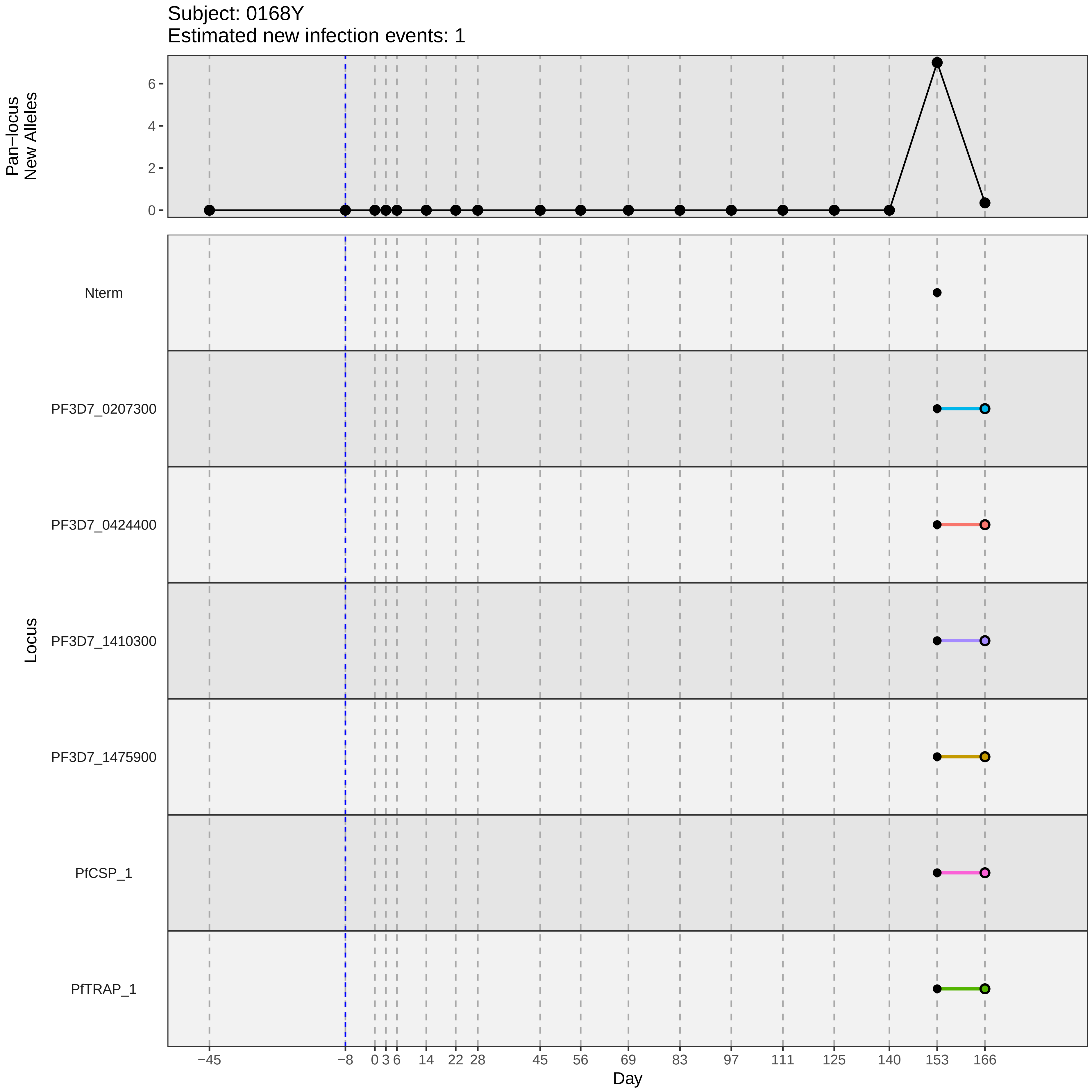

Subject: 0168Y
Estimated new infection events: 1
Pan−locus
New Alleles
6
4
2
0
Locus
Nterm
PF3D7_0207300
PF3D7_0424400
PF3D7_1410300
PF3D7_1475900
PfCSP_1
PfTRAP_1
−45
−8
0
3
6
14
22
28
45
56
69
83
97
111
125
140
153
166
Day
