## Supplementary material for "Genotypic infection endpoint analysis to understand efficacy and escape potential of the malaria monoclonal antibody CIS43LS": Data File S1: participant_0175G_with_imputation.pdf

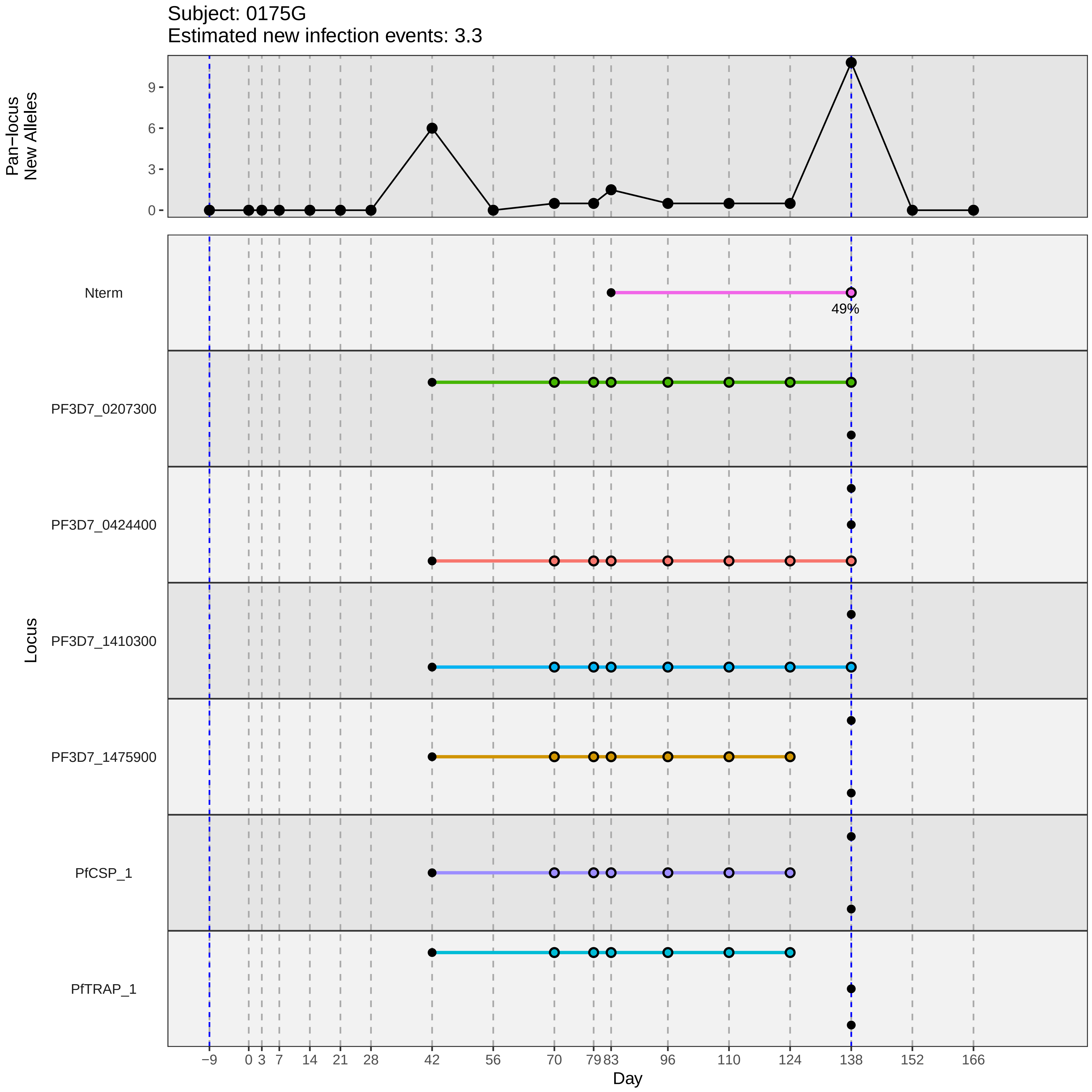

Subject: 0175G
Estimated new infection events: 3.3
Pan−locus
New Alleles
0
3
6
9
Locus
Nterm
49%
PF3D7_0207300
PF3D7_0424400
PF3D7_1410300
PF3D7_1475900
PfCSP_1
PfTRAP_1
−9
0
3
7
14
21
28
42
56
70
79
83
96
110
124
138
152
166
Day
