## Supplementary material for "Genotypic infection endpoint analysis to understand efficacy and escape potential of the malaria monoclonal antibody CIS43LS": Data File S1: participant_0176H_with_imputation.pdf

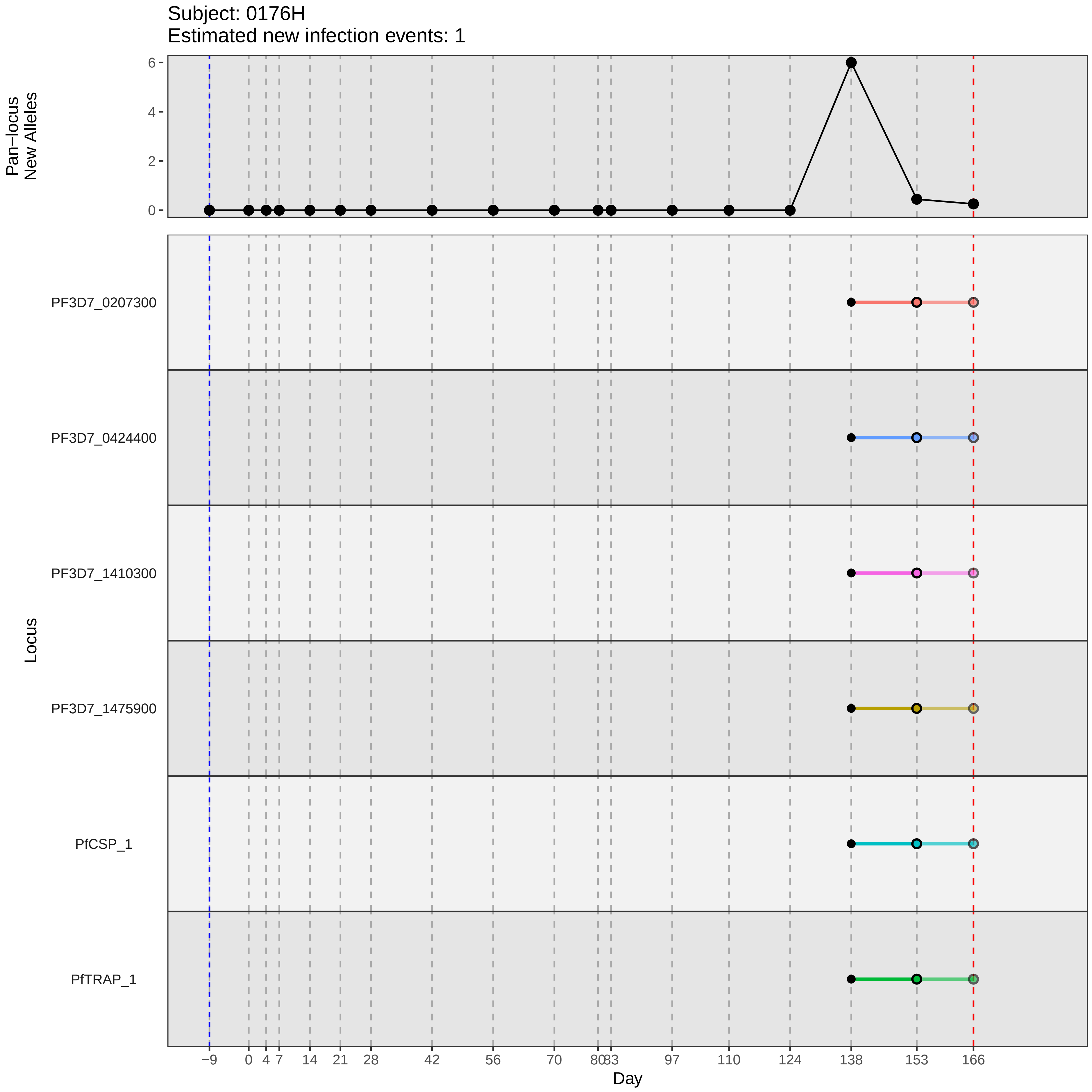
Subject: 0176H
Estimated new infection events: 1
Pan−locus New Alleles
0
2
4
6
Locus
PF3D7_0207300
PF3D7_0424400
PF3D7_1410300
PF3D7_1475900
PfCSP_1
PfTRAP_1
−9
0
4
7
14
21
28
42
56
70
80
83
97
110
124
138
153
166
Day
