## Supplementary material for "Genotypic infection endpoint analysis to understand efficacy and escape potential of the malaria monoclonal antibody CIS43LS": Data File S1: participant_0180P_with_imputation.pdf

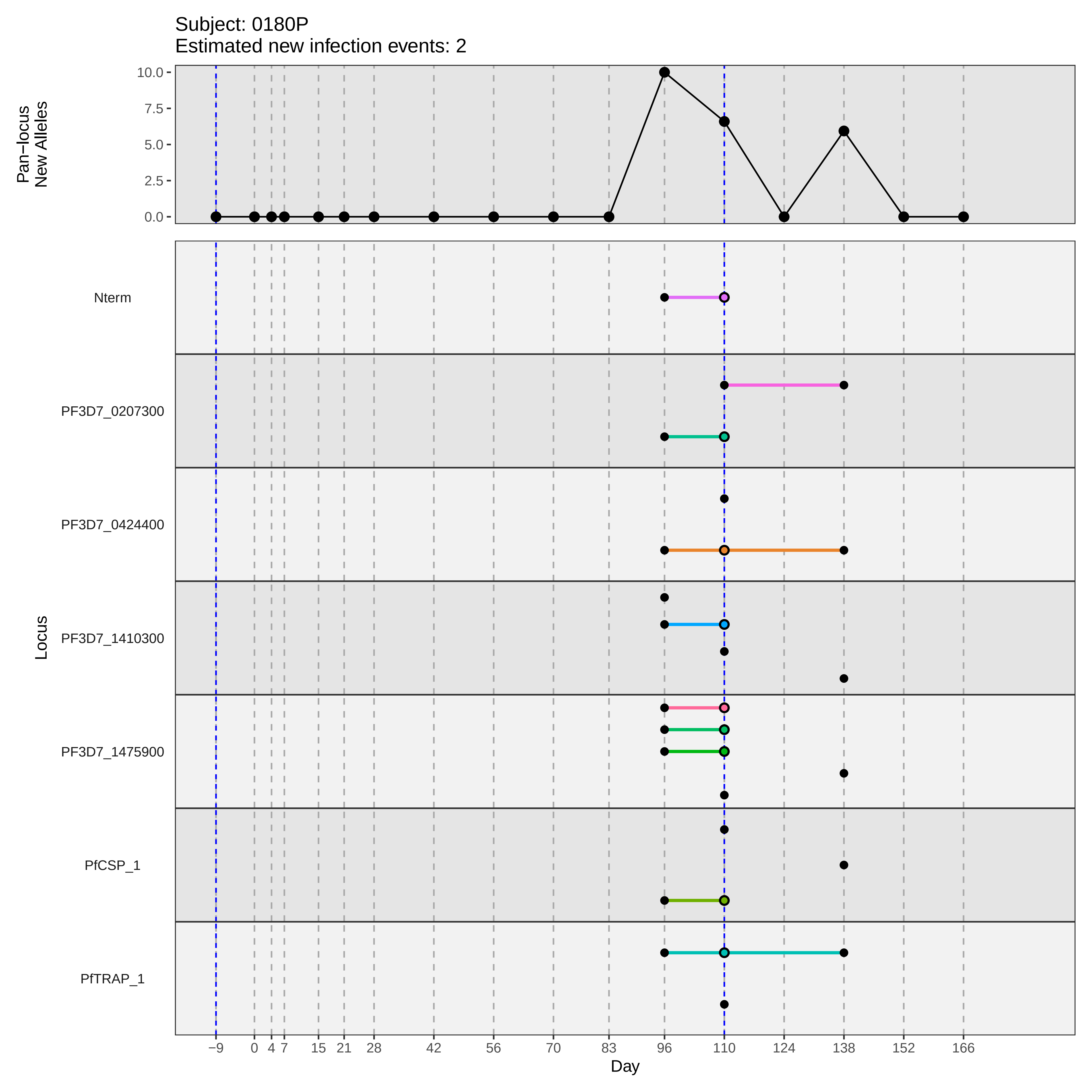

Subject: 0180P
Estimated new infection events: 2
Pan−locus New Alleles
10.0
7.5
5.0
2.5
0.0
Locus
Nterm
PF3D7_0207300
PF3D7_0424400
PF3D7_1410300
PF3D7_1475900
PfCSP_1
PfTRAP_1
−9 0 4 7 15 21 28 42 56 70 83 96 110 124 138 152 166
Day
