## Supplementary material for "Genotypic infection endpoint analysis to understand efficacy and escape potential of the malaria monoclonal antibody CIS43LS": Data File S1: participant_0184T_with_imputation.pdf

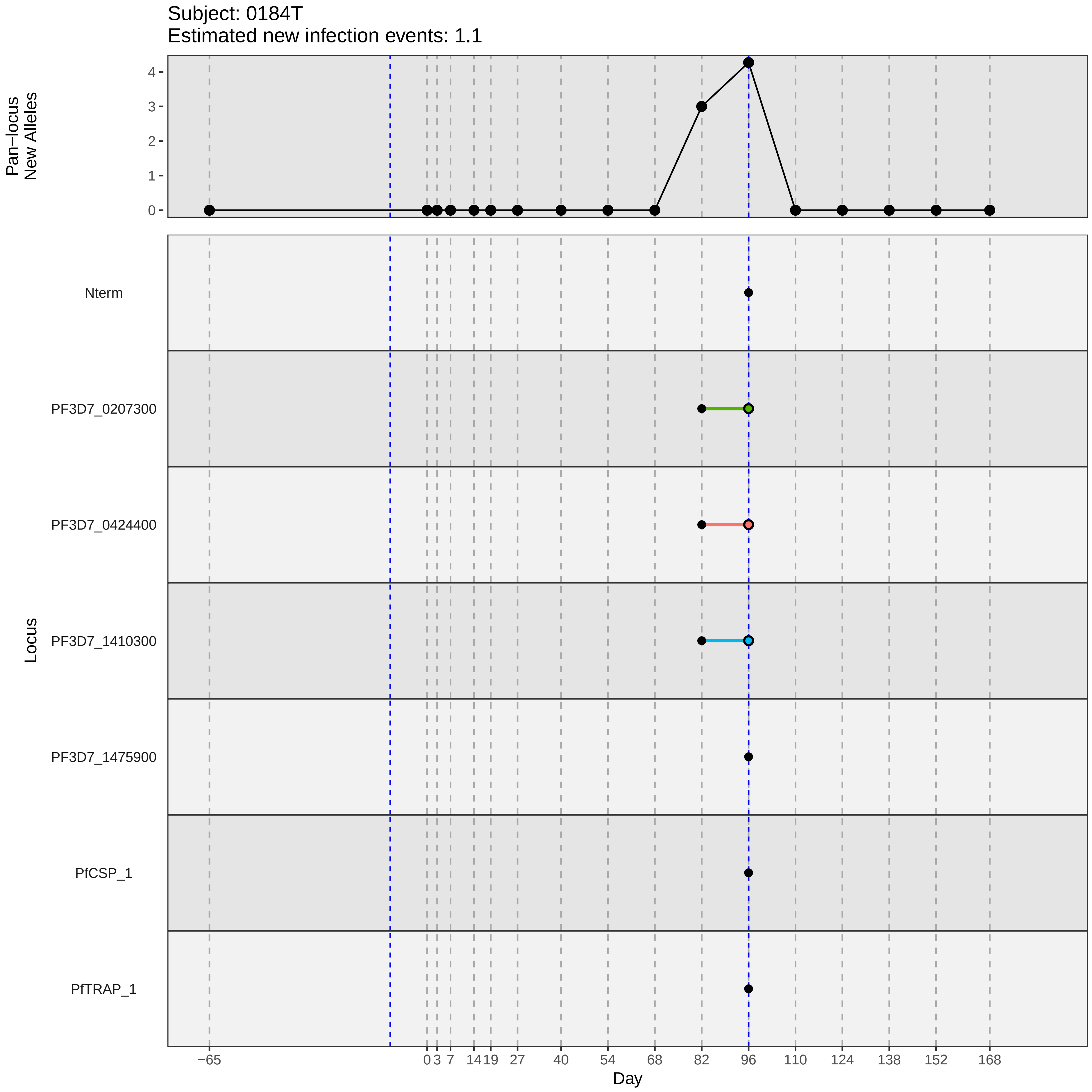

Subject: 0184T
Estimated new infection events: 1.1
Pan−locus New Alleles
Locus
Nterm
PF3D7_0207300
PF3D7_0424400
PF3D7_1410300
PF3D7_1475900
PfCSP_1
PfTRAP_1
Day
−65
0
3
7
14
19
27
40
54
68
82
96
110
124
138
152
168
0
1
2
3
4
