## Supplementary material for "Genotypic infection endpoint analysis to understand efficacy and escape potential of the malaria monoclonal antibody CIS43LS": Data File S1: participant_0185U_with_imputation.pdf

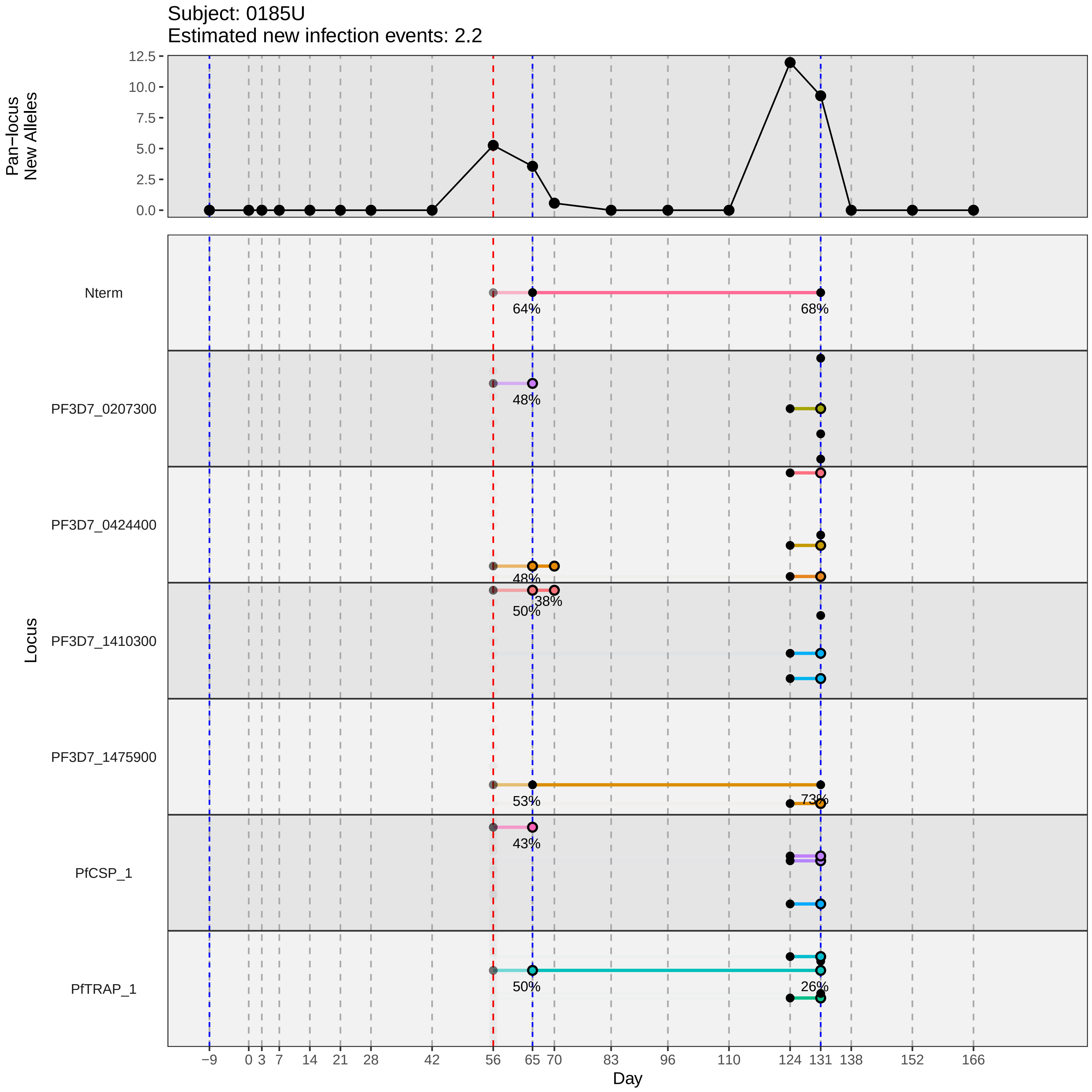
Subject: 0185U
Estimated new infection events: 2.2
Pan−locus New Alleles
12.5
10.0
7.5
5.0
2.5
0.0
Locus
Nterm
PF3D7_0207300
PF3D7_0424400
PF3D7_1410300
PF3D7_1475900
PfCSP_1
PfTRAP_1
64%
68%
48%
48%
38%
50%
53%
73%
43%
50%
26%
−9
0
3
7
14
21
28
42
56
65
70
83
96
110
124
131
138
152
166
Day
