## Supplementary material for "Genotypic infection endpoint analysis to understand efficacy and escape potential of the malaria monoclonal antibody CIS43LS": Data File S1: participant_0191B_with_imputation.pdf

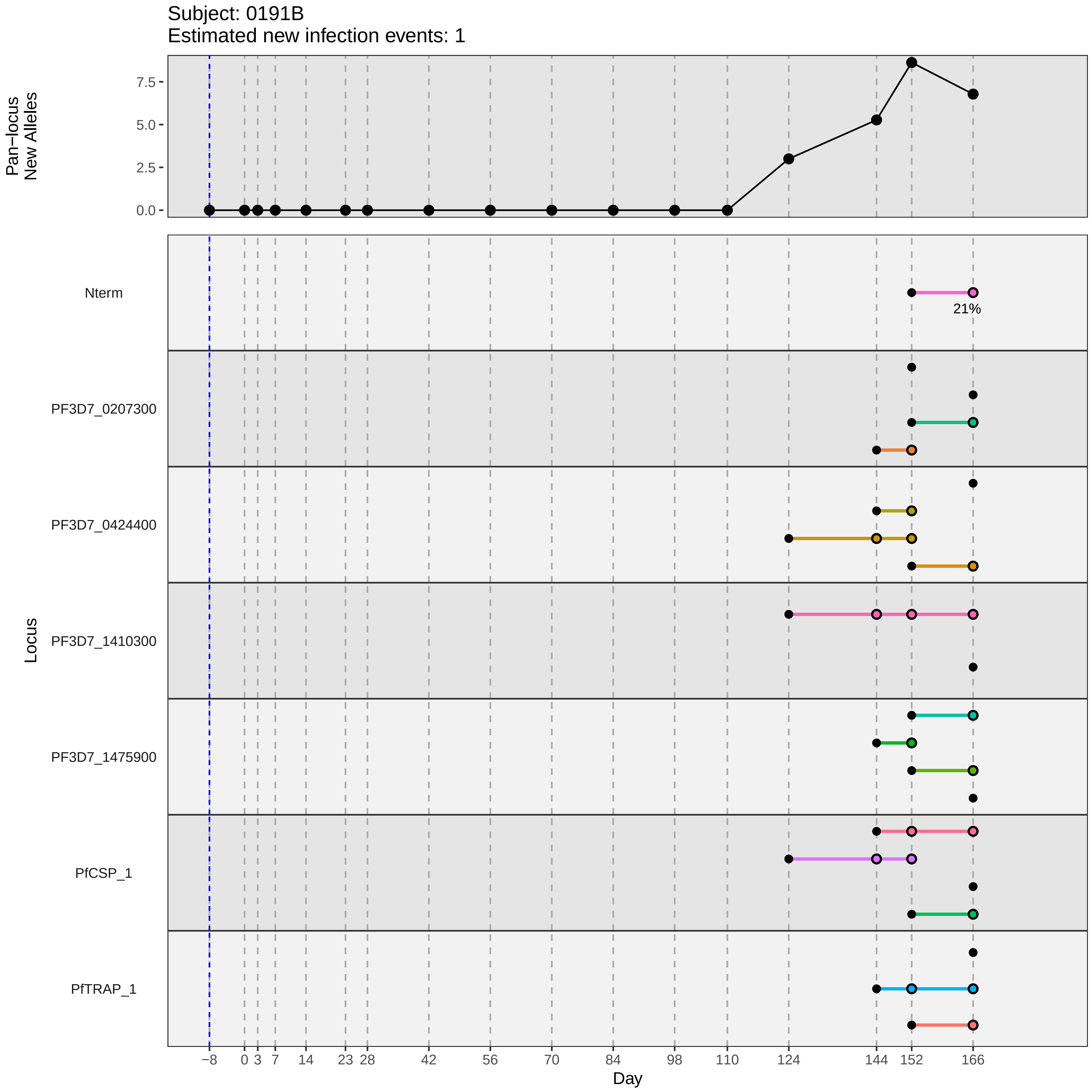

Subject: 0191B
Estimated new infection events: 1
Pan−locus New Alleles
7.5
5.0
2.5
0.0
Locus
Nterm
21%
PF3D7_0207300
PF3D7_0424400
PF3D7_1410300
PF3D7_1475900
PfCSP_1
PfTRAP_1
−8 0 3 7 14 23 28 42 56 70 84 98 110 124 144 152 166
Day
