## Supplementary material for "Genotypic infection endpoint analysis to understand efficacy and escape potential of the malaria monoclonal antibody CIS43LS": Data File S1: participant_0193D_with_imputation.pdf

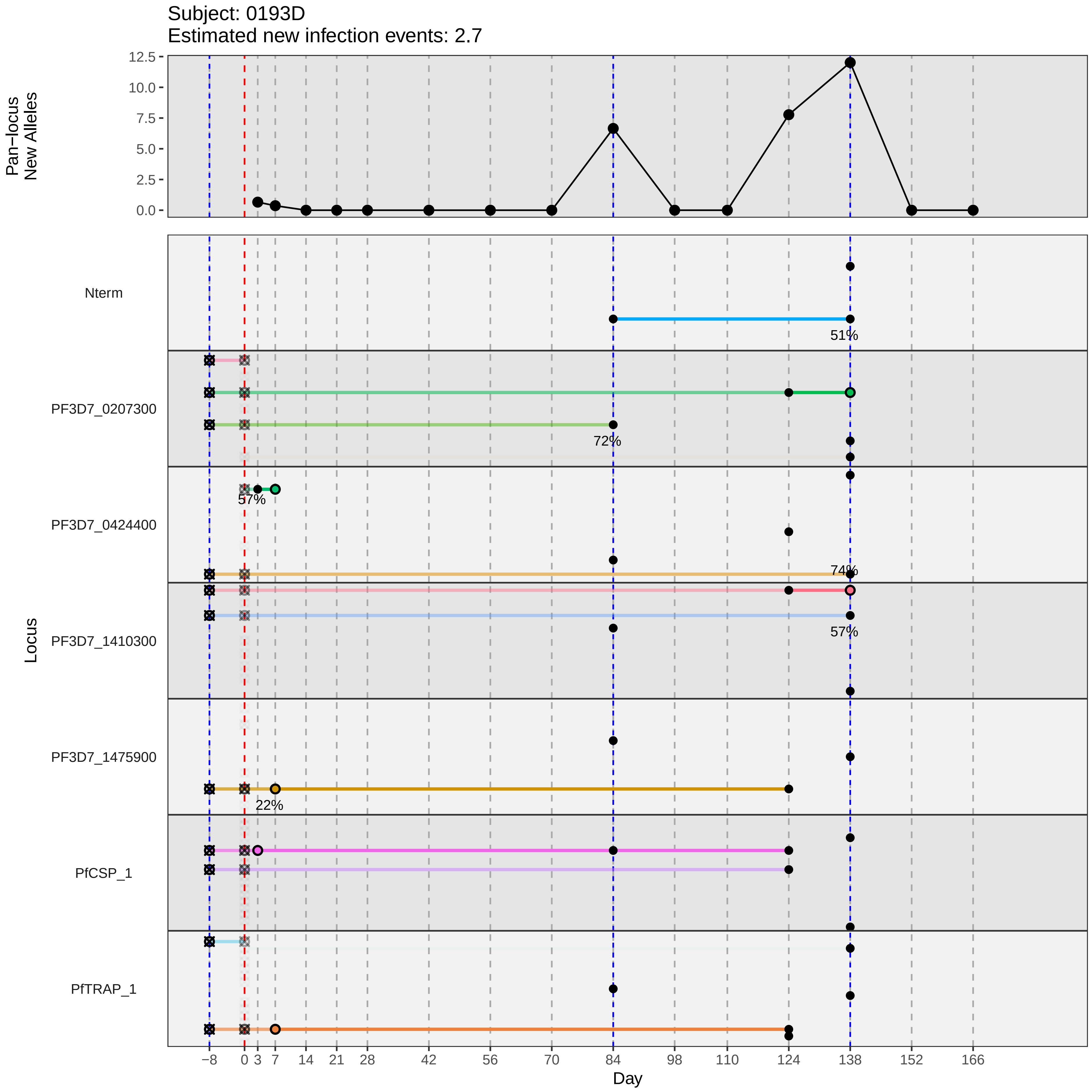

Subject: 0193D
Estimated new infection events: 2.7
Pan−locus
New Alleles
12.5
10.0
7.5
5.0
2.5
0.0
Locus
Nterm
PF3D7_0207300
PF3D7_0424400
PF3D7_1410300
PF3D7_1475900
PfCSP_1
PfTRAP_1
51%
72%
57%
74%
57%
22%
−8
0
3
7
14
21
28
42
56
70
84
98
110
124
138
152
166
Day
