## Supplementary material for "Genotypic infection endpoint analysis to understand efficacy and escape potential of the malaria monoclonal antibody CIS43LS": Data File S1: participant_0194E_with_imputation.pdf

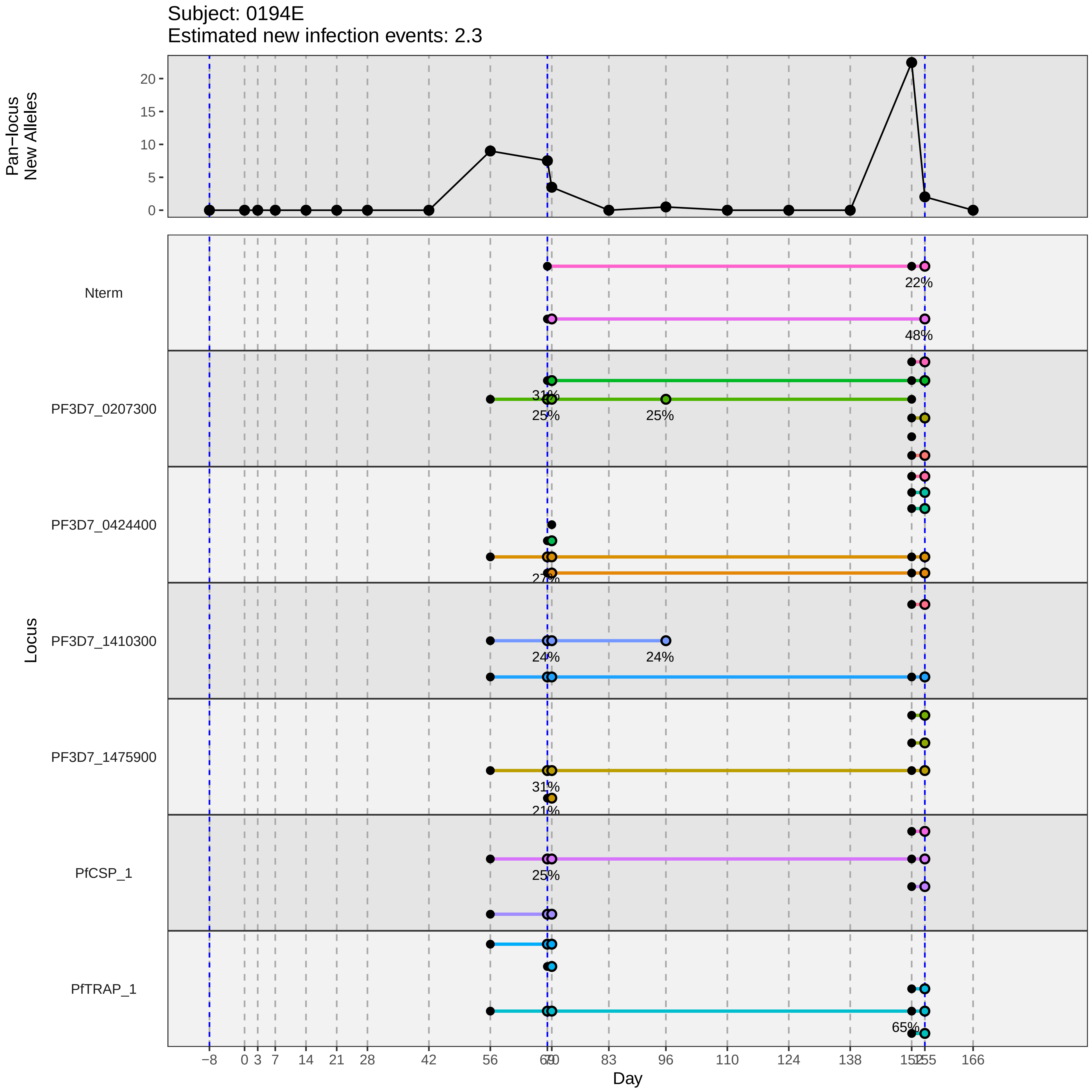

Subject: 0194E
Estimated new infection events: 2.3
Pan−locus New Alleles
0
5
10
15
20
Locus
Nterm
PF3D7_0207300
PF3D7_0424400
PF3D7_1410300
PF3D7_1475900
PfCSP_1
PfTRAP_1
22%
48%
31%
25%
25%
27%
24%
24%
31%
21%
25%
65%
Day
−8
0
3
7
14
21
28
42
56
83
96
110
124
138
166
