## Supplementary material for "Genotypic infection endpoint analysis to understand efficacy and escape potential of the malaria monoclonal antibody CIS43LS": Data File S1: participant_0198K_with_imputation.pdf

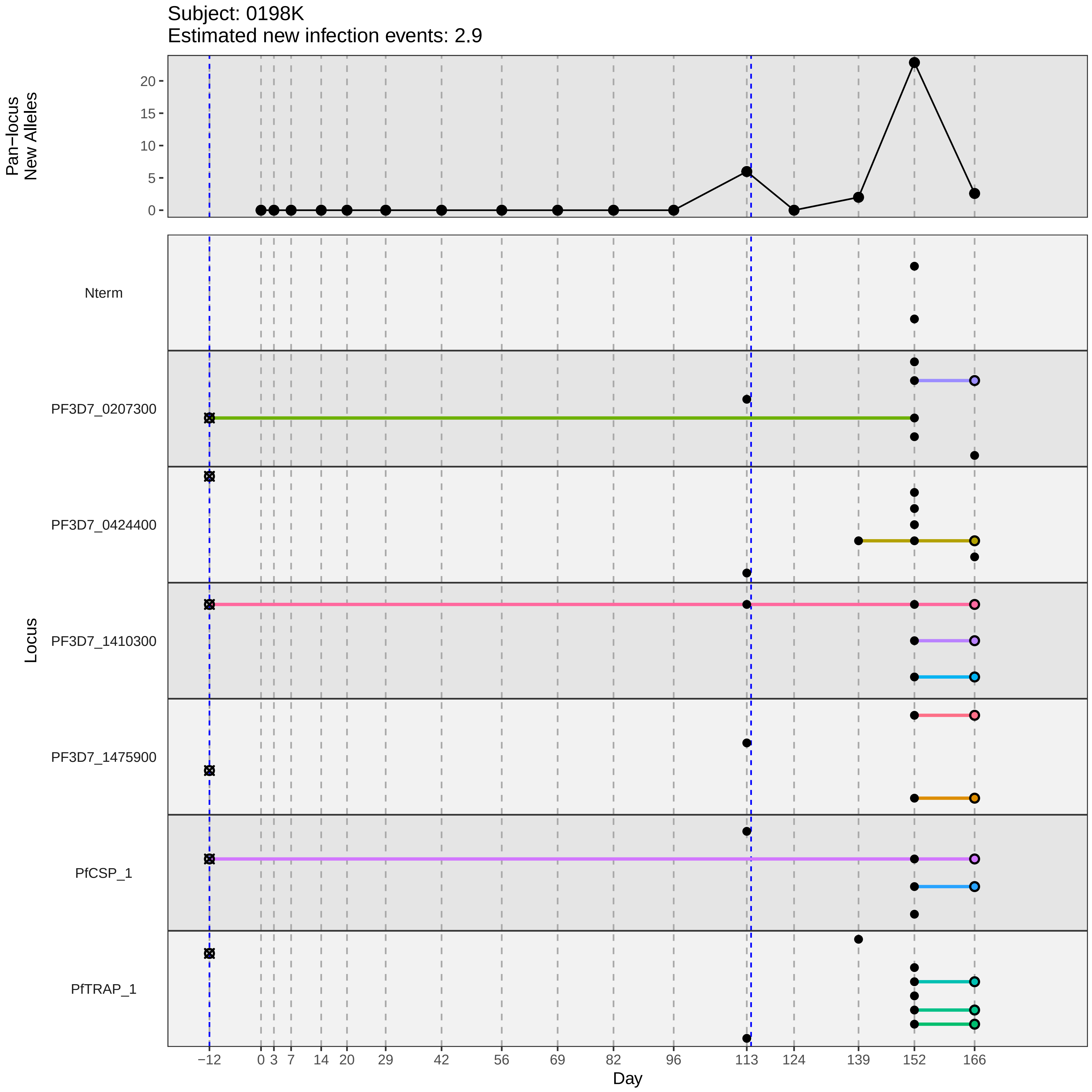

Subject: 0198K
Estimated new infection events: 2.9
Pan–locus
New Alleles
0
5
10
15
20
Locus
Nterm
PF3D7_0207300
PF3D7_0424400
PF3D7_1410300
PF3D7_1475900
PfCSP_1
PfTRAP_1
−12
0
3
7
14
20
29
42
56
69
82
96
113
124
139
152
166
Day
