## Supplementary material for "Genotypic infection endpoint analysis to understand efficacy and escape potential of the malaria monoclonal antibody CIS43LS": Data File S1: participant_0200N_with_imputation.pdf

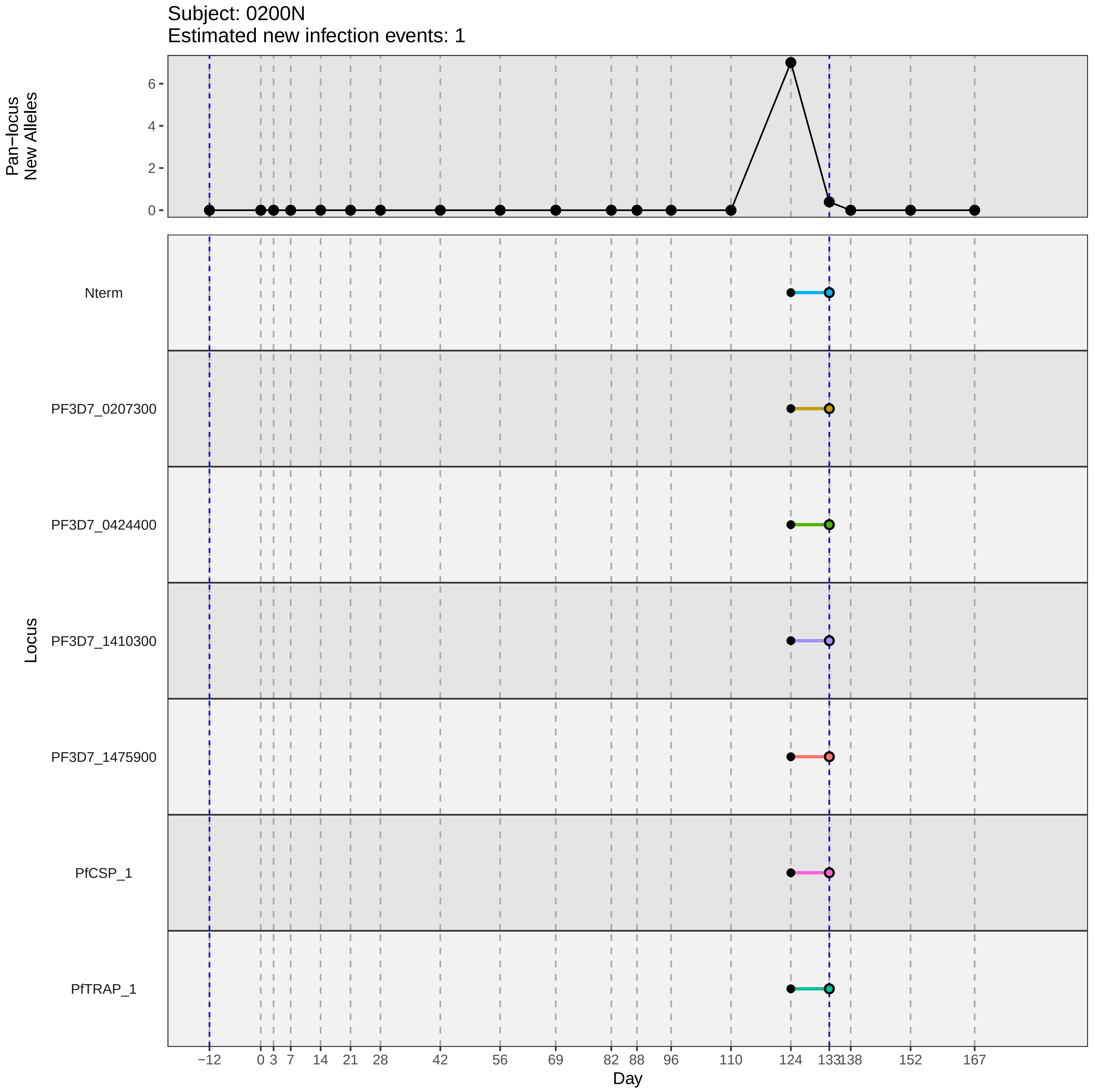
Subject: 0200N
Estimated new infection events: 1
Pan−locus New Alleles
0
2
4
6
Locus
Nterm
PF3D7_0207300
PF3D7_0424400
PF3D7_1410300
PF3D7_1475900
PfCSP_1
PfTRAP_1
−12
0
3
7
14
21
28
42
56
69
82
88
96
110
124
133
138
152
167
Day
