## Supplementary material for "Genotypic infection endpoint analysis to understand efficacy and escape potential of the malaria monoclonal antibody CIS43LS": Data File S1: participant_0201P_with_imputation.pdf

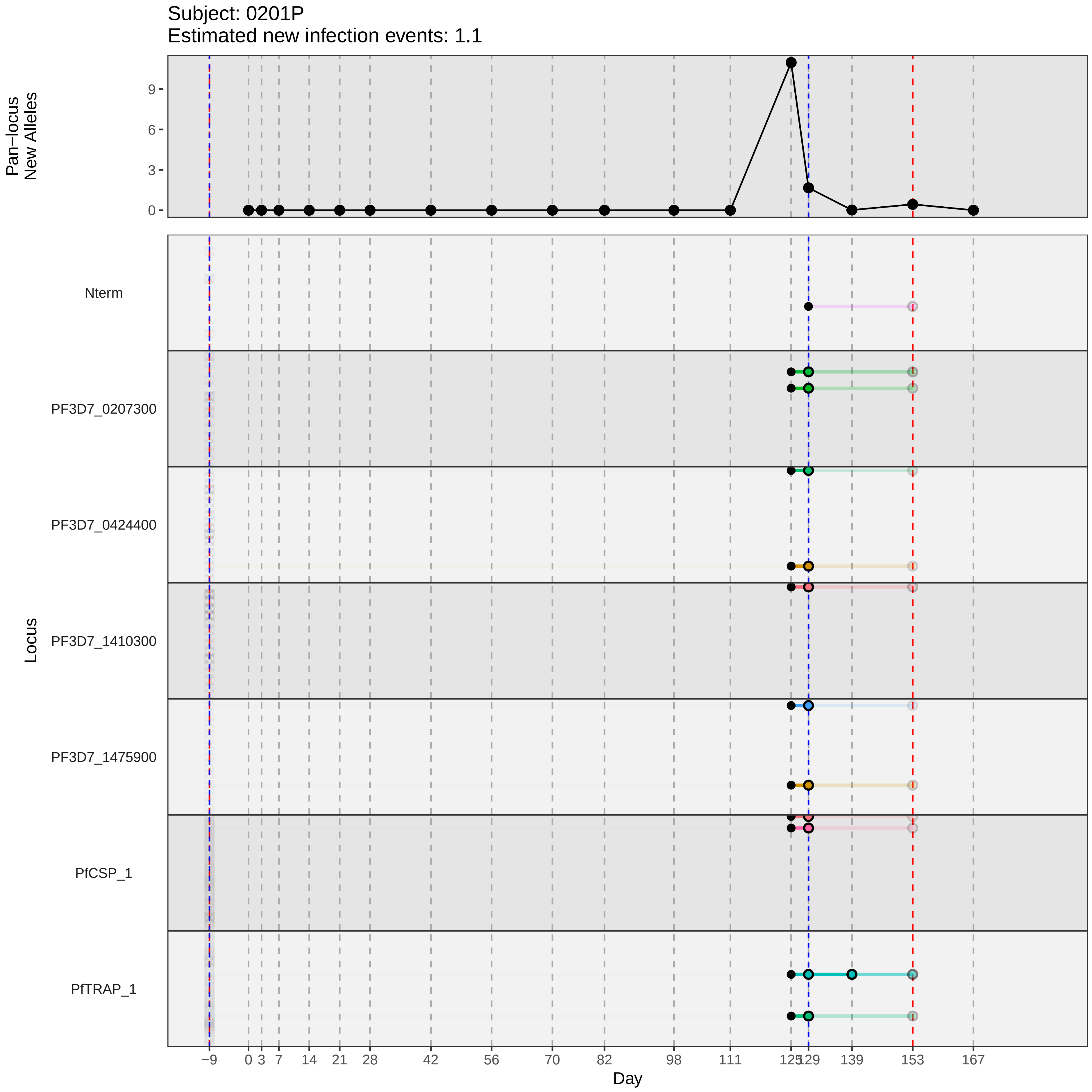

Subject: 0201P
Estimated new infection events: 1.1
Pan−locus
New Alleles
0
3
6
9
Locus
Nterm
PF3D7_0207300
PF3D7_0424400
PF3D7_1410300
PF3D7_1475900
PfCSP_1
PfTRAP_1
−9
0
3
7
14
21
28
42
56
70
82
98
111
125
129
139
153
167
Day
