## Supplementary material for "Genotypic infection endpoint analysis to understand efficacy and escape potential of the malaria monoclonal antibody CIS43LS": Data File S1: participant_0202Q_with_imputation.pdf

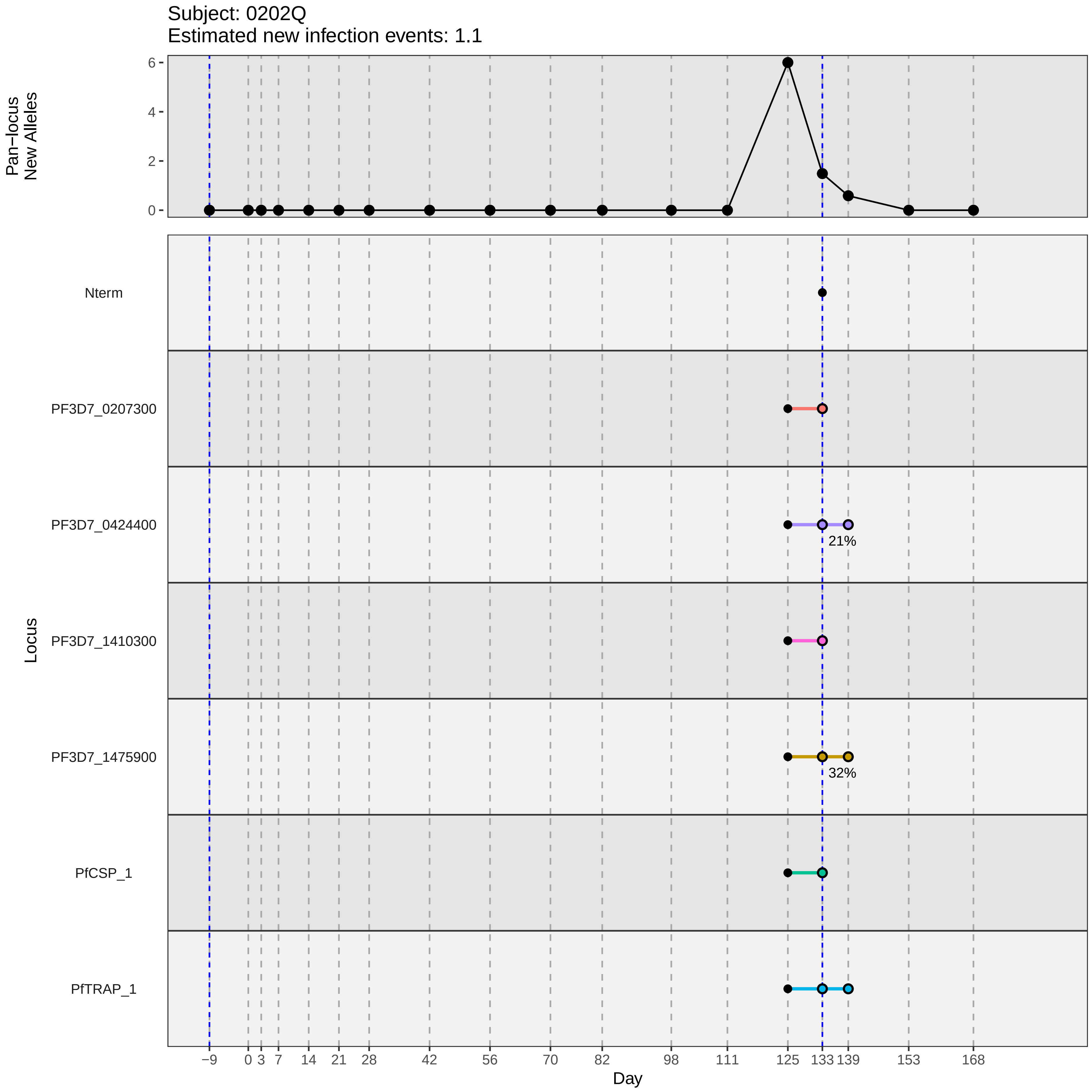
Subject: 0202Q
Estimated new infection events: 1.1
Pan−locus
New Alleles
6
4
2
0
Locus
Nterm
PF3D7_0207300
PF3D7_0424400
21%
PF3D7_1410300
PF3D7_1475900
32%
PfCSP_1
PfTRAP_1
−9
0
3
7
14
21
28
42
56
70
82
98
111
125
133
139
153
168
Day
