## Supplementary material for "Genotypic infection endpoint analysis to understand efficacy and escape potential of the malaria monoclonal antibody CIS43LS": Data File S1: participant_0206U_with_imputation.pdf

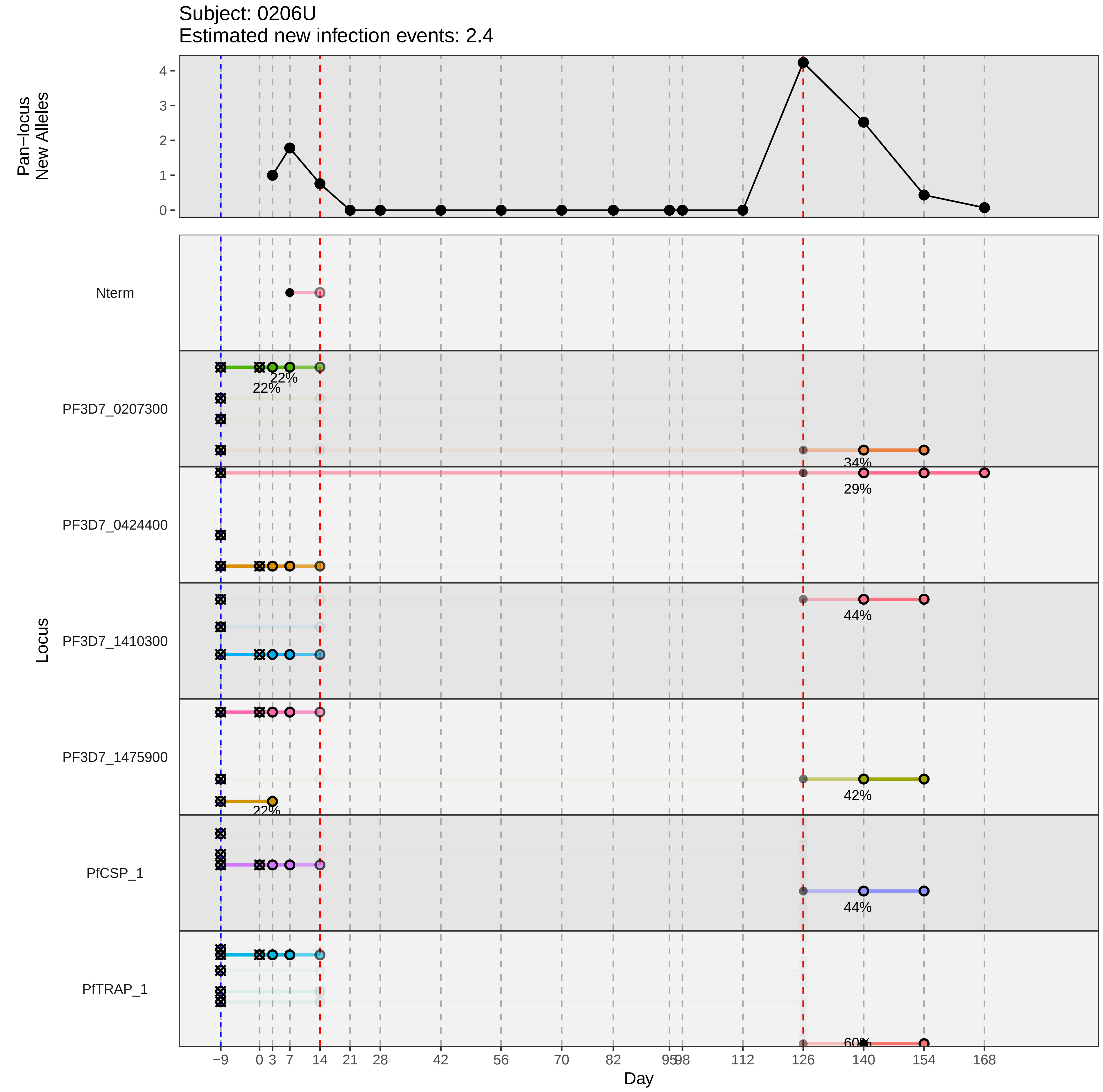

Subject: 0206U
Estimated new infection events: 2.4
Pan−locus
New Alleles
0
1
2
3
4
Nterm
PF3D7_0207300
22%
22%
34%
PF3D7_0424400
29%
PF3D7_1410300
44%
PF3D7_1475900
42%
22%
PfCSP_1
44%
PfTRAP_1
60%
Locus
−9
0
3
7
14
21
28
42
56
70
82
95
98
112
126
140
154
168
Day
