## Supplementary material for "Genotypic infection endpoint analysis to understand efficacy and escape potential of the malaria monoclonal antibody CIS43LS": Data File S1: participant_0211A_with_imputation.pdf

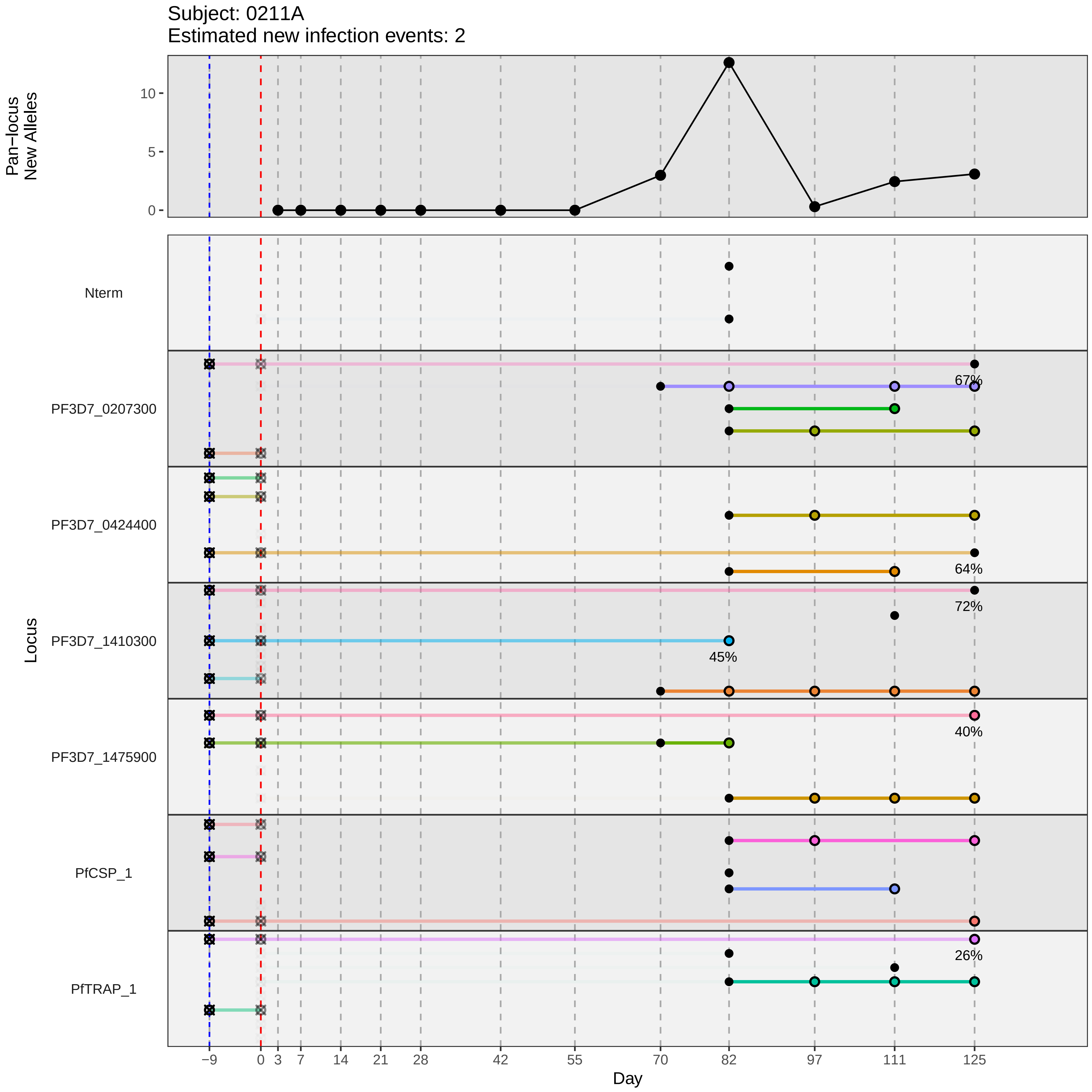
Subject: 0211A
Estimated new infection events: 2
Pan−locus New Alleles
10
5
0
Locus
Nterm
PF3D7_0207300
PF3D7_0424400
PF3D7_1410300
PF3D7_1475900
PfCSP_1
PfTRAP_1
67%
64%
72%
45%
40%
26%
−9
0
3
7
14
21
28
42
55
70
82
97
111
125
Day
