## Supplementary material for "Genotypic infection endpoint analysis to understand efficacy and escape potential of the malaria monoclonal antibody CIS43LS": Data File S1: participant_0213C_with_imputation.pdf

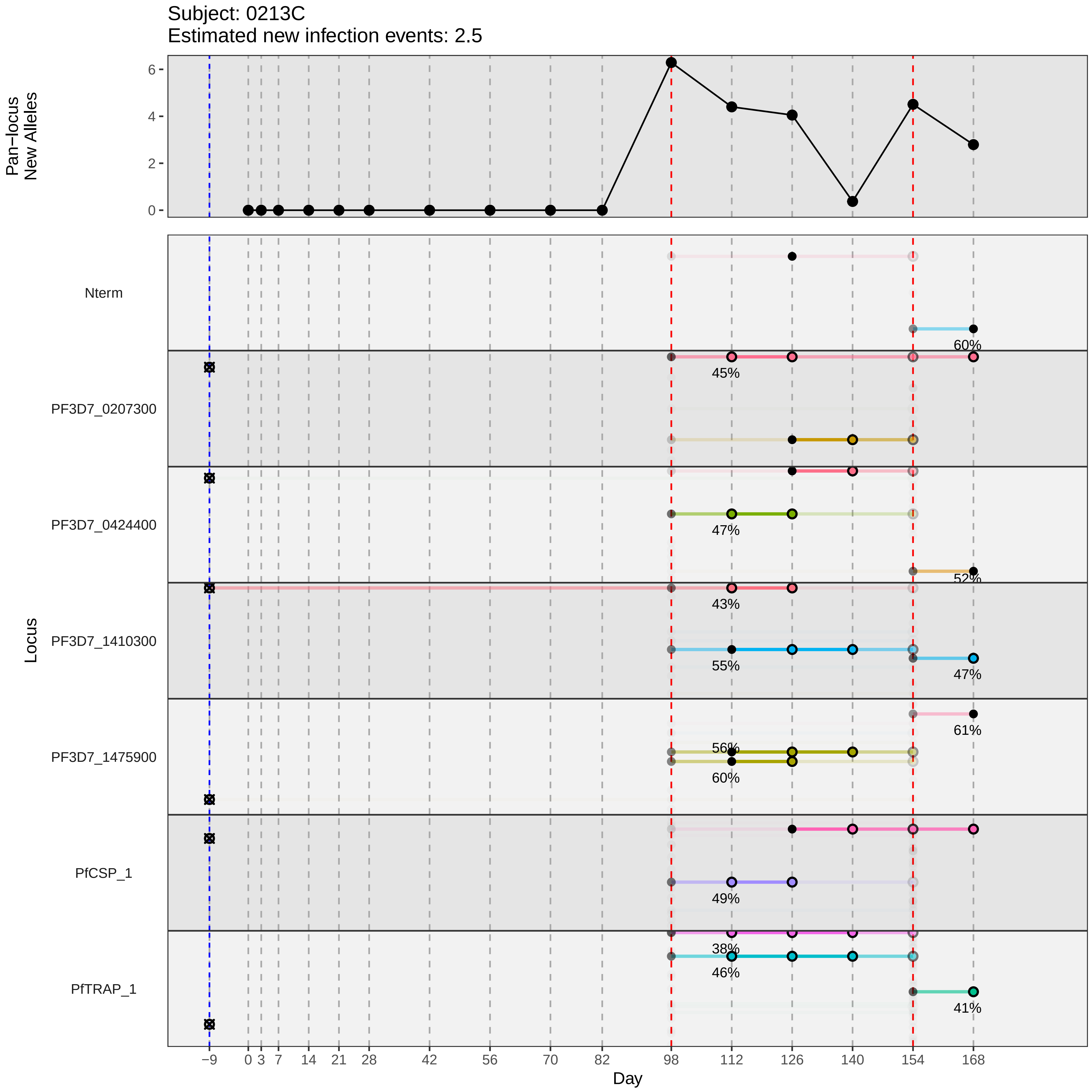

Subject: 0213C
Estimated new infection events: 2.5
Pan−locus New Alleles
0
2
4
6
Locus
Nterm
PF3D7_0207300
PF3D7_0424400
PF3D7_1410300
PF3D7_1475900
PfCSP_1
PfTRAP_1
60%
45%
47%
52%
43%
55%
47%
61%
56%
60%
49%
38%
46%
41%
−9
0
3
7
14
21
28
42
56
70
82
98
112
126
140
154
168
Day
