## Supplementary material for "Genotypic infection endpoint analysis to understand efficacy and escape potential of the malaria monoclonal antibody CIS43LS": Data File S1: participant_0222P_with_imputation.pdf

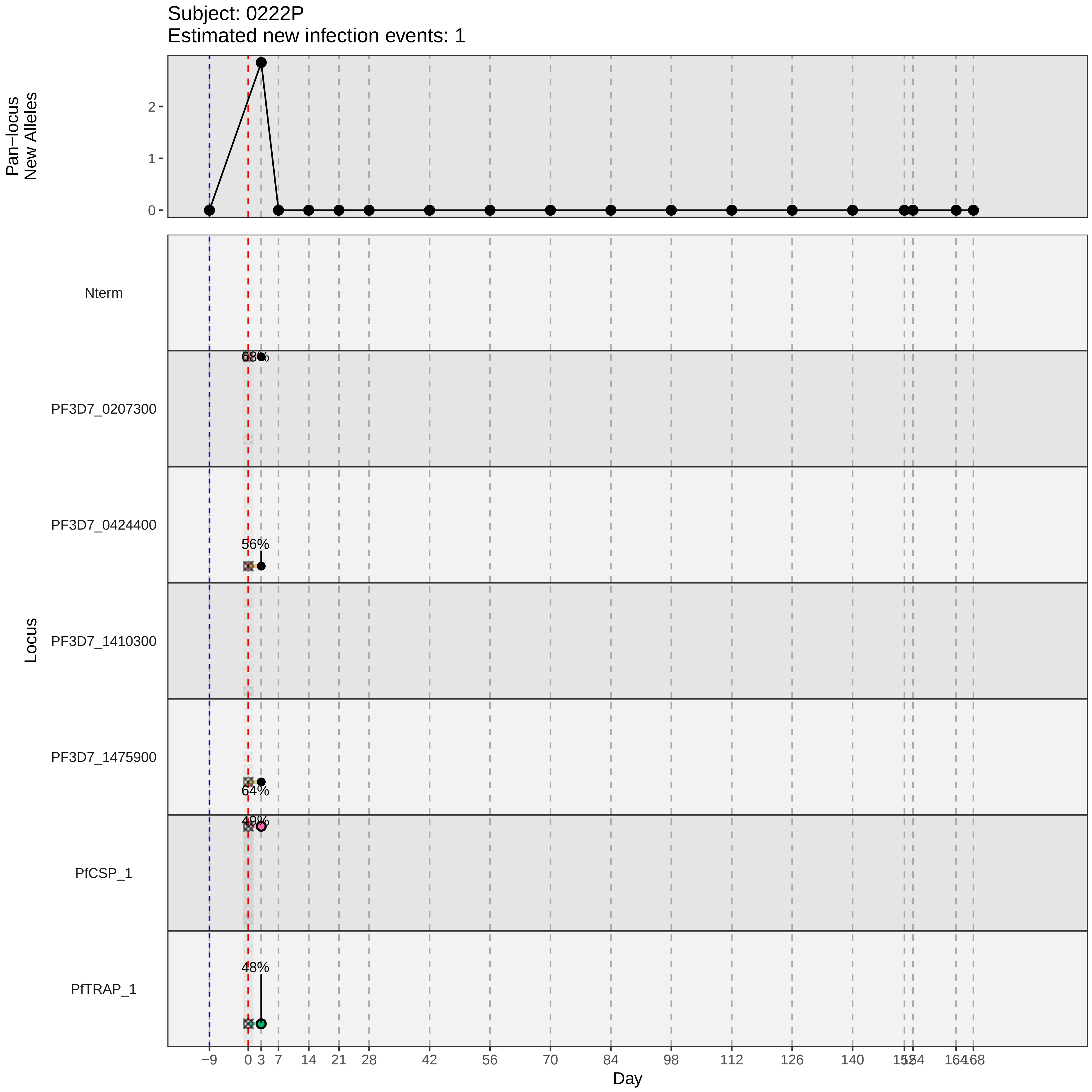

Subject: 0222P
Estimated new infection events: 1
Pan−locus
New Alleles
2
1
0
Nterm
PF3D7_0207300
68%
PF3D7_0424400
56%
PF3D7_1410300
PF3D7_1475900
64%
49%
PfCSP_1
48%
PfTRAP_1
Locus
−9
0
3
7
14
21
28
42
56
70
84
98
112
126
140
168
Day
