## Supplementary material for "Genotypic infection endpoint analysis to understand efficacy and escape potential of the malaria monoclonal antibody CIS43LS": Data File S1: participant_0223Q_with_imputation.pdf

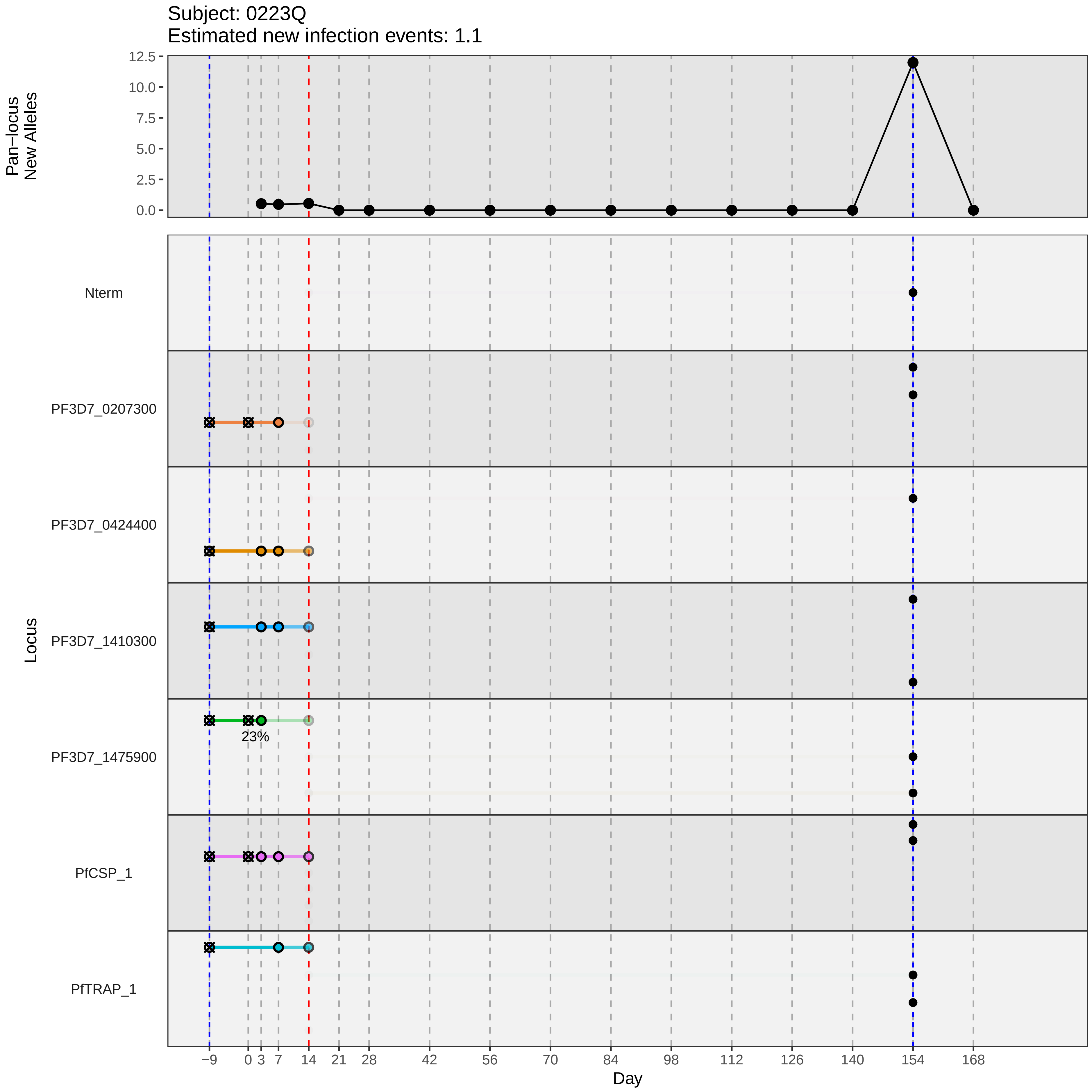

Subject: 0223Q
Estimated new infection events: 1.1
Pan−locus New Alleles
12.5
10.0
7.5
5.0
2.5
0.0
Locus
Nterm
PF3D7_0207300
PF3D7_0424400
PF3D7_1410300
PF3D7_1475900
23%
PfCSP_1
PfTRAP_1
−9
0
3
7
14
21
28
42
56
70
84
98
112
126
140
154
168
Day
