## Supplementary material for "Genotypic infection endpoint analysis to understand efficacy and escape potential of the malaria monoclonal antibody CIS43LS": Data File S1: participant_0225S_with_imputation.pdf

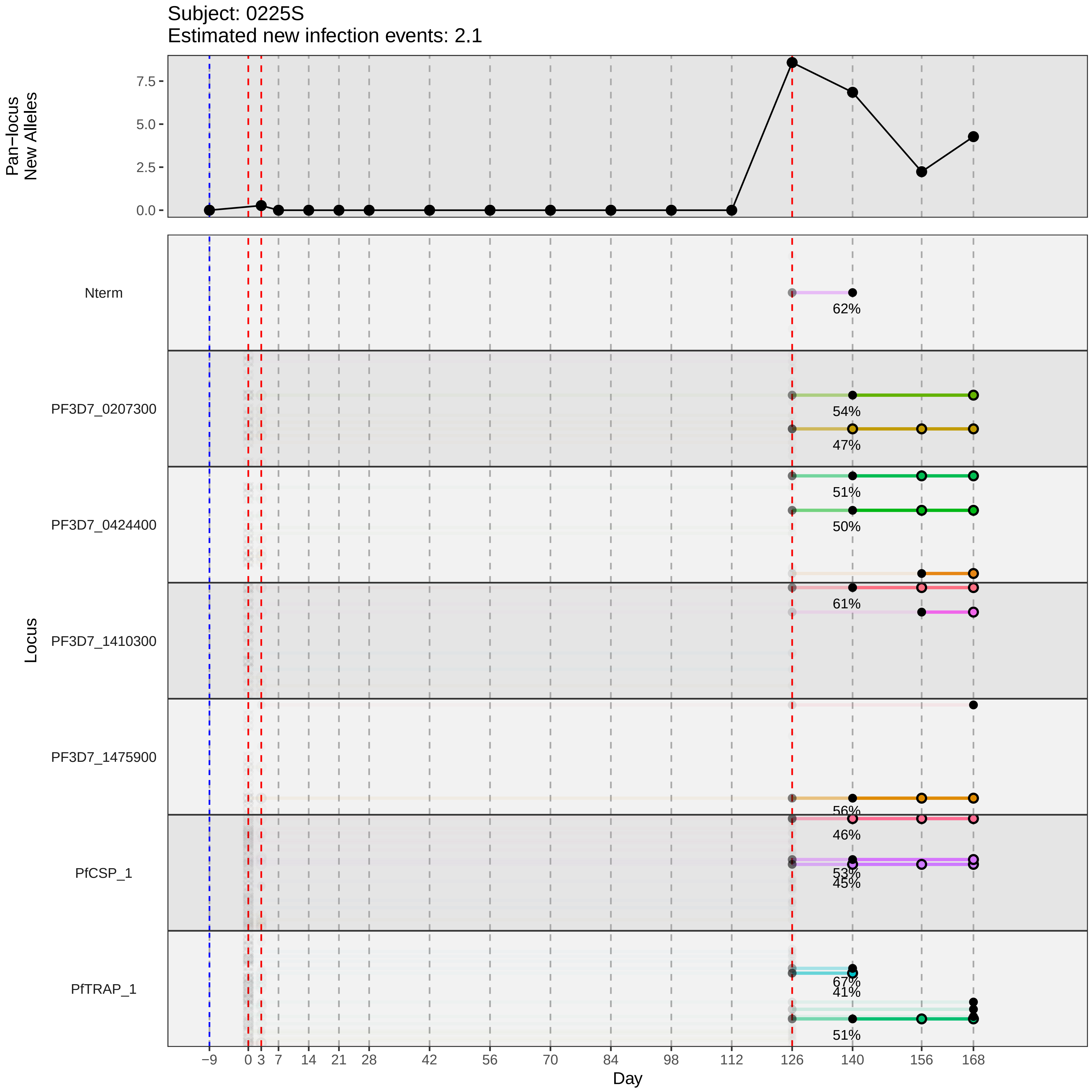
Subject: 0225S
Estimated new infection events: 2.1
Pan−locus New Alleles
7.5
5.0
2.5
0.0
Locus
Nterm
PF3D7_0207300
PF3D7_0424400
PF3D7_1410300
PF3D7_1475900
PfCSP_1
PfTRAP_1
62%
54%
47%
51%
50%
61%
56%
46%
53%
45%
67%
41%
51%
−9
0
3
7
14
21
28
42
56
70
84
98
112
126
140
156
168
Day
