## Supplementary material for "Genotypic infection endpoint analysis to understand efficacy and escape potential of the malaria monoclonal antibody CIS43LS": Data File S1: participant_0231Y_with_imputation.pdf

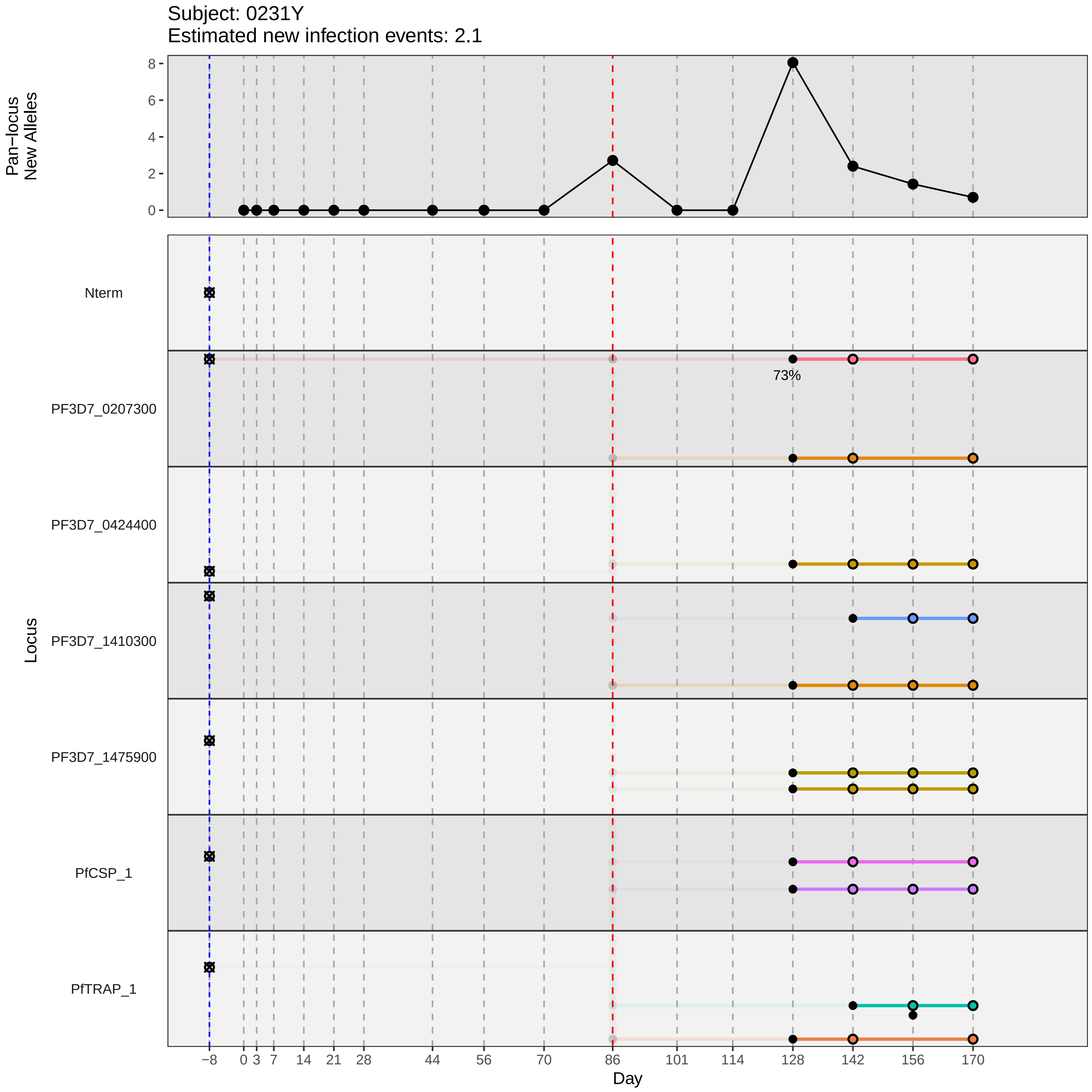
Subject: 0231Y
Estimated new infection events: 2.1
Pan−locus
New Alleles
8
6
4
2
0
Nterm
PF3D7_0207300
73%
PF3D7_0424400
PF3D7_1410300
PF3D7_1475900
PfCSP_1
PfTRAP_1
Locus
−8
0
3
7
14
21
28
44
56
70
86
101
114
128
142
156
170
Day
