## Supplementary material for "Genotypic infection endpoint analysis to understand efficacy and escape potential of the malaria monoclonal antibody CIS43LS": Data File S1: participant_0245R_with_imputation.pdf

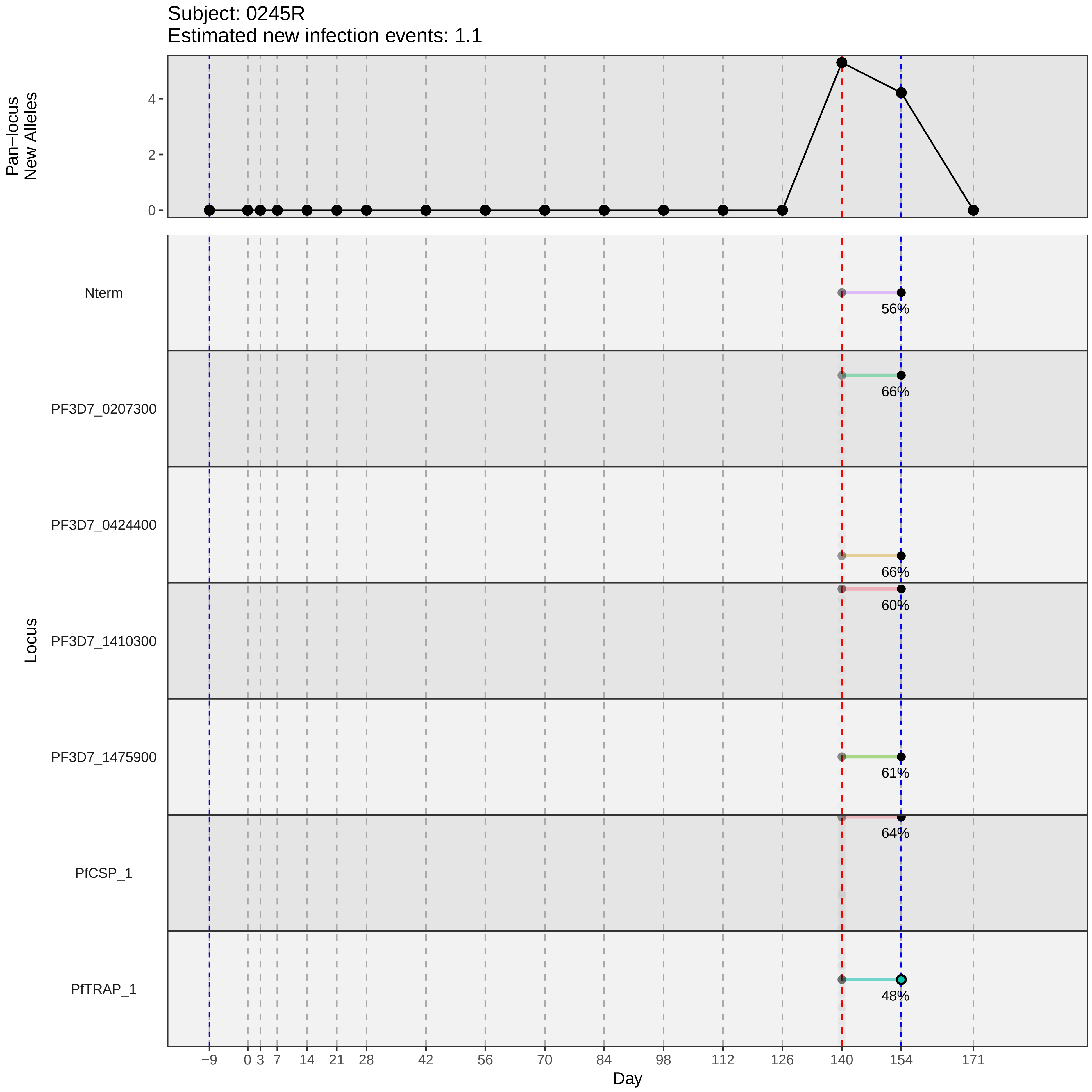

Subject: 0245R
Estimated new infection events: 1.1
Pan−locus New Alleles
4
2
0
Nterm
56%
PF3D7_0207300
66%
PF3D7_0424400
66%
60%
PF3D7_1410300
PF3D7_1475900
61%
64%
PfCSP_1
PfTRAP_1
48%
Locus
−9
0
3
7
14
21
28
42
56
70
84
98
112
126
140
154
171
Day
