## Supplementary material for "Genotypic infection endpoint analysis to understand efficacy and escape potential of the malaria monoclonal antibody CIS43LS": Data File S1: participant_0253A_with_imputation.pdf

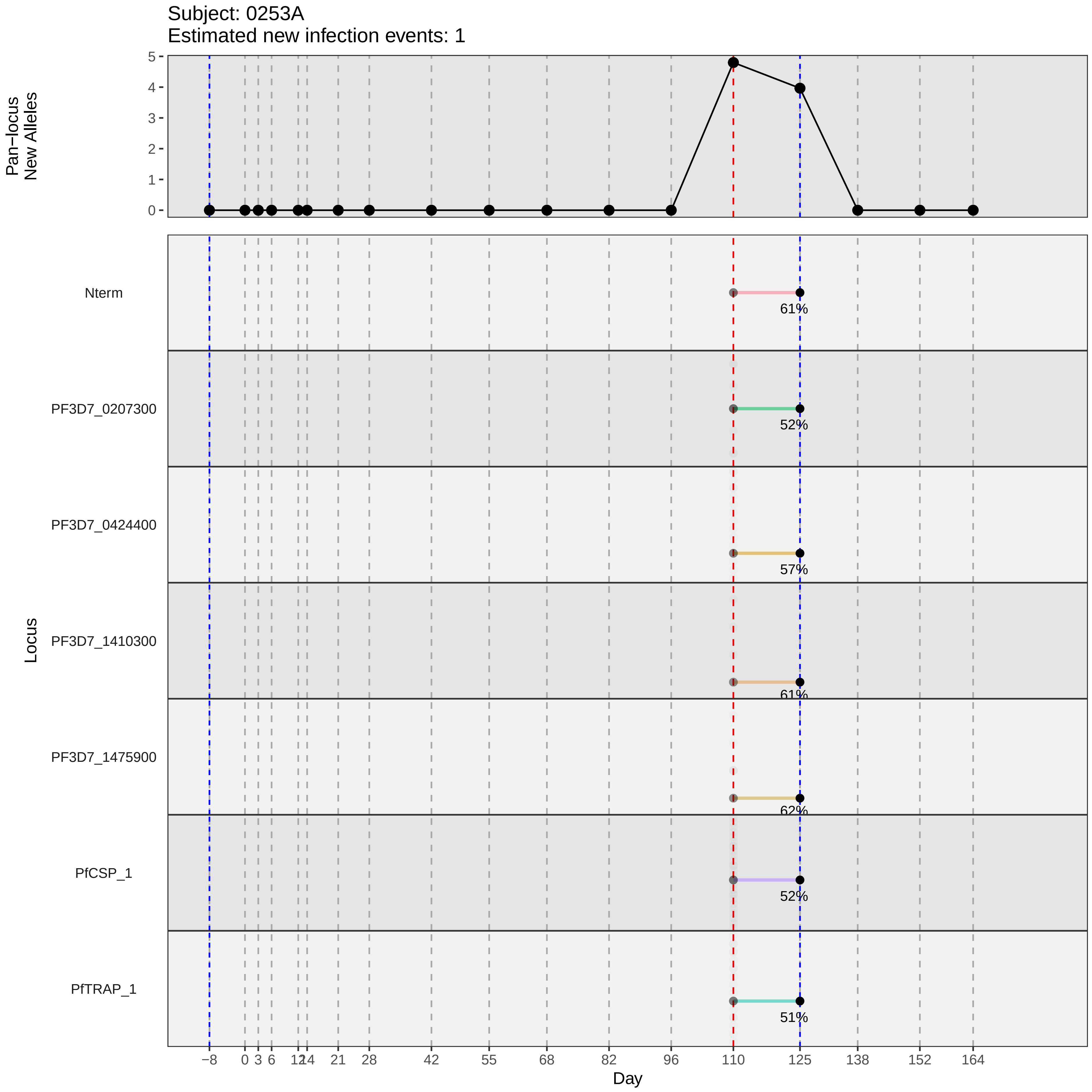

Subject: 0253A
Estimated new infection events: 1
Pan−locus New Alleles
5
4
3
2
1
0
Locus
Nterm
61%
PF3D7_0207300
52%
PF3D7_0424400
57%
PF3D7_1410300
61%
PF3D7_1475900
62%
PfCSP_1
52%
PfTRAP_1
51%
−8
0
3
6
12
14
21
28
42
55
68
82
96
110
125
138
152
164
Day
