## Supplementary material for "Genotypic infection endpoint analysis to understand efficacy and escape potential of the malaria monoclonal antibody CIS43LS": Data File S1: participant_0255C_with_imputation.pdf

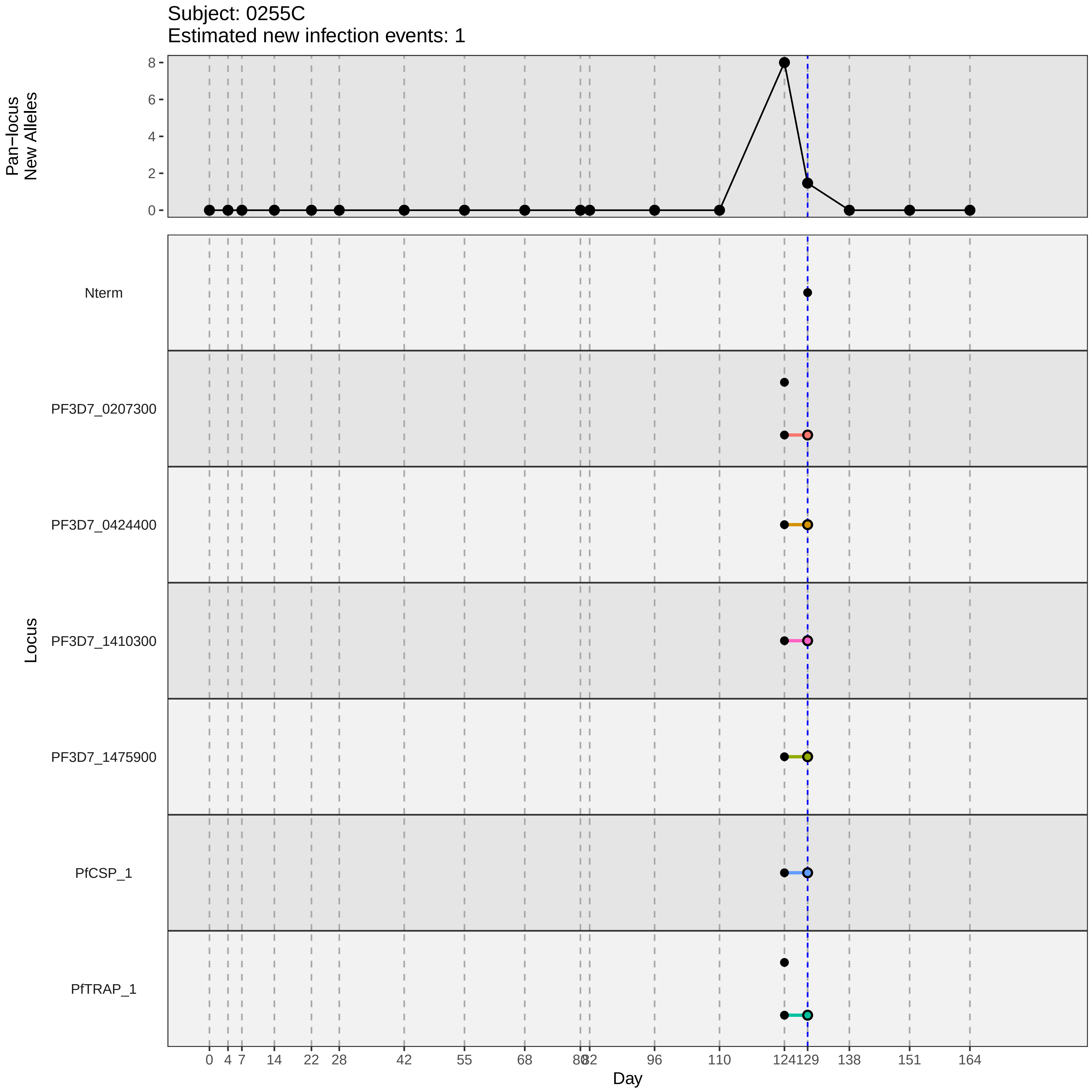

Subject: 0255C
Estimated new infection events: 1
Pan−locus New Alleles
8
6
4
2
0
Locus
Nterm
PF3D7_0207300
PF3D7_0424400
PF3D7_1410300
PF3D7_1475900
PfCSP_1
PfTRAP_1
0 4 7 14 22 28 42 55 68 80 82 96 110 124 129 138 151 164
Day
