## Supplementary material for "Genotypic infection endpoint analysis to understand efficacy and escape potential of the malaria monoclonal antibody CIS43LS": Data File S1: participant_0266R_with_imputation.pdf

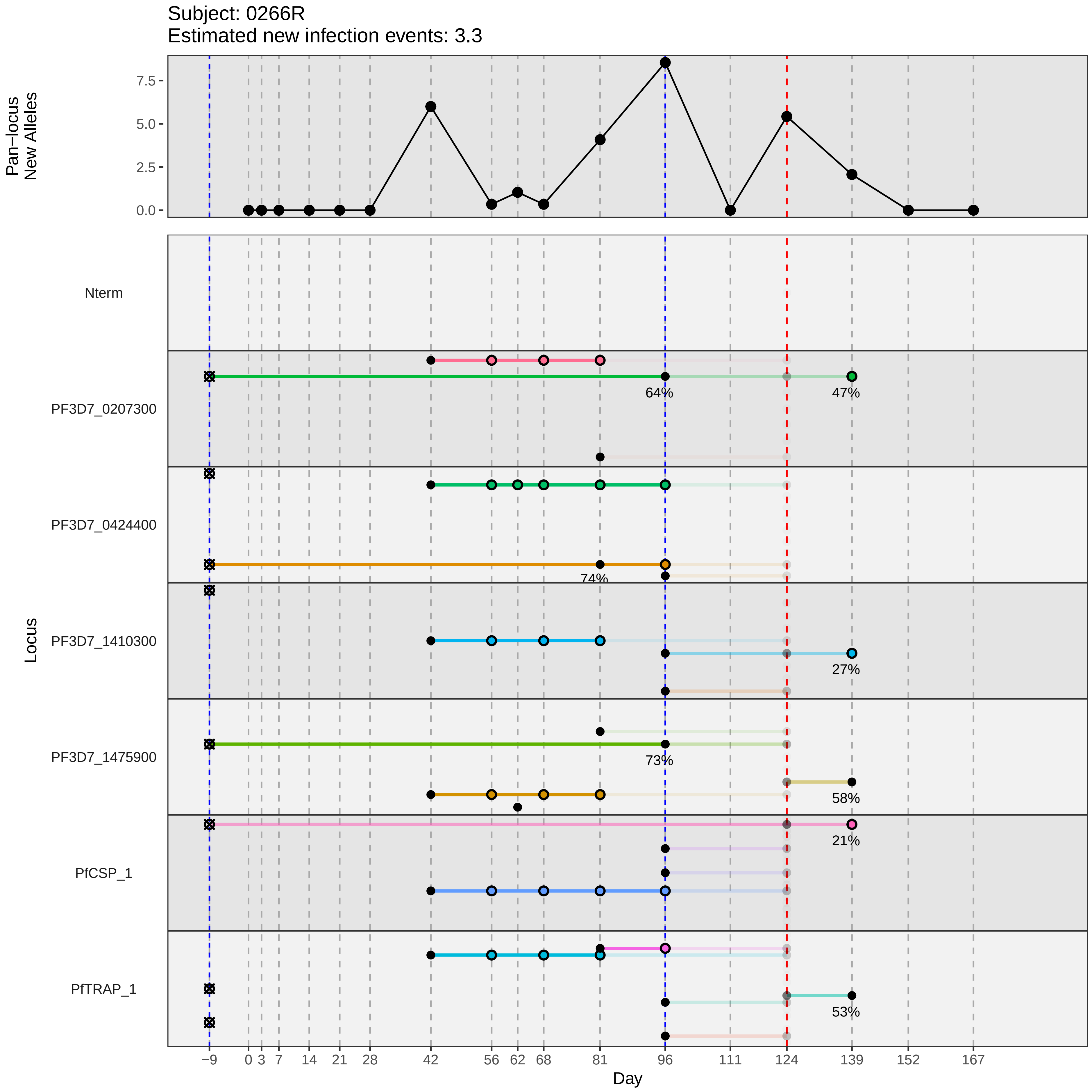

Subject: 0266R
Estimated new infection events: 3.3
Pan−locus New Alleles
7.5
5.0
2.5
0.0
Locus
Nterm
PF3D7_0207300
64%
47%
PF3D7_0424400
74%
PF3D7_1410300
27%
PF3D7_1475900
73%
58%
PfCSP_1
21%
PfTRAP_1
53%
−9 0 3 7 14 21 28 42 56 62 68 81 96 111 124 139 152 167
Day
