## Supplementary material for "Genotypic infection endpoint analysis to understand efficacy and escape potential of the malaria monoclonal antibody CIS43LS": Data File S1: participant_0267S_with_imputation.pdf

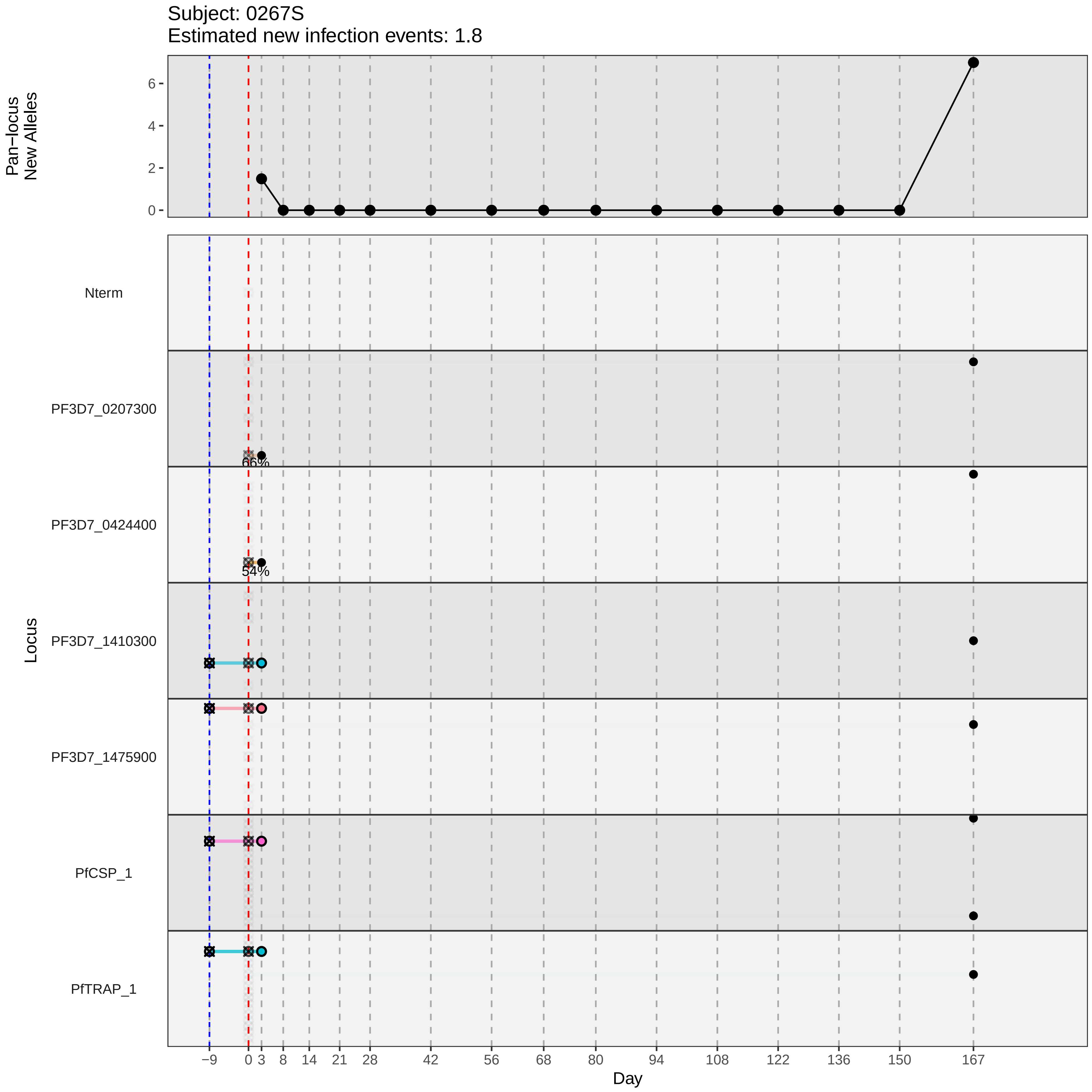

Subject: 0267S
Estimated new infection events: 1.8
Pan−locus
New Alleles
6
4
2
0
Locus
Nterm
PF3D7_0207300
66%
PF3D7_0424400
54%
PF3D7_1410300
PF3D7_1475900
PfCSP_1
PfTRAP_1
−9
0
3
8
14
21
28
42
56
68
80
94
108
122
136
150
167
Day
