## Supplementary material for "Genotypic infection endpoint analysis to understand efficacy and escape potential of the malaria monoclonal antibody CIS43LS": Data File S1: participant_0272X_with_imputation.pdf

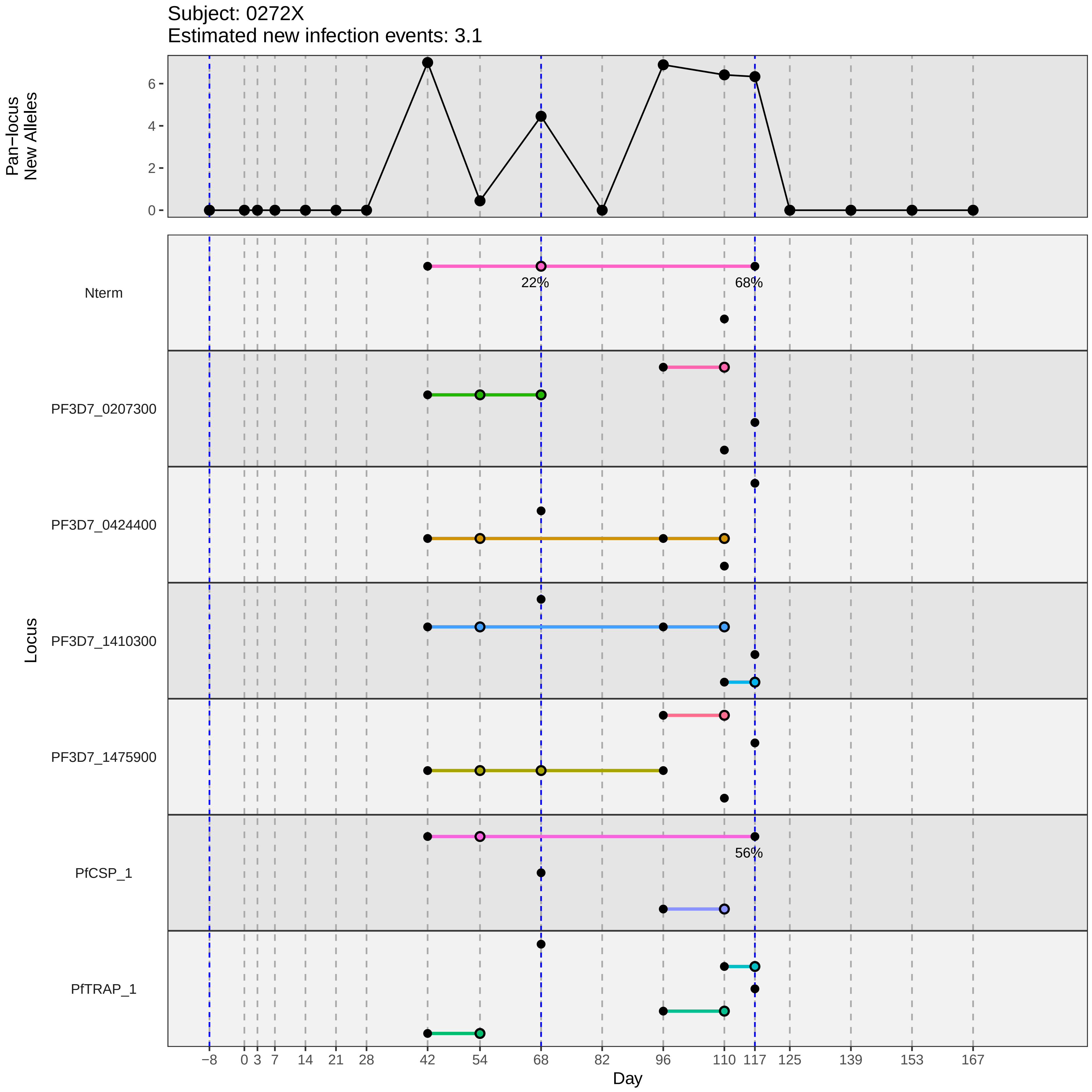

Subject: 0272X
Estimated new infection events: 3.1
Pan−locus
New Alleles
0
2
4
6
Locus
Nterm
PF3D7_0207300
PF3D7_0424400
PF3D7_1410300
PF3D7_1475900
PfCSP_1
PfTRAP_1
22%
68%
56%
−8
0
3
7
14
21
28
42
54
68
82
96
110
117
125
139
153
167
Day
