## Supplementary material for "Genotypic infection endpoint analysis to understand efficacy and escape potential of the malaria monoclonal antibody CIS43LS": Data File S1: participant_0276C_with_imputation.pdf

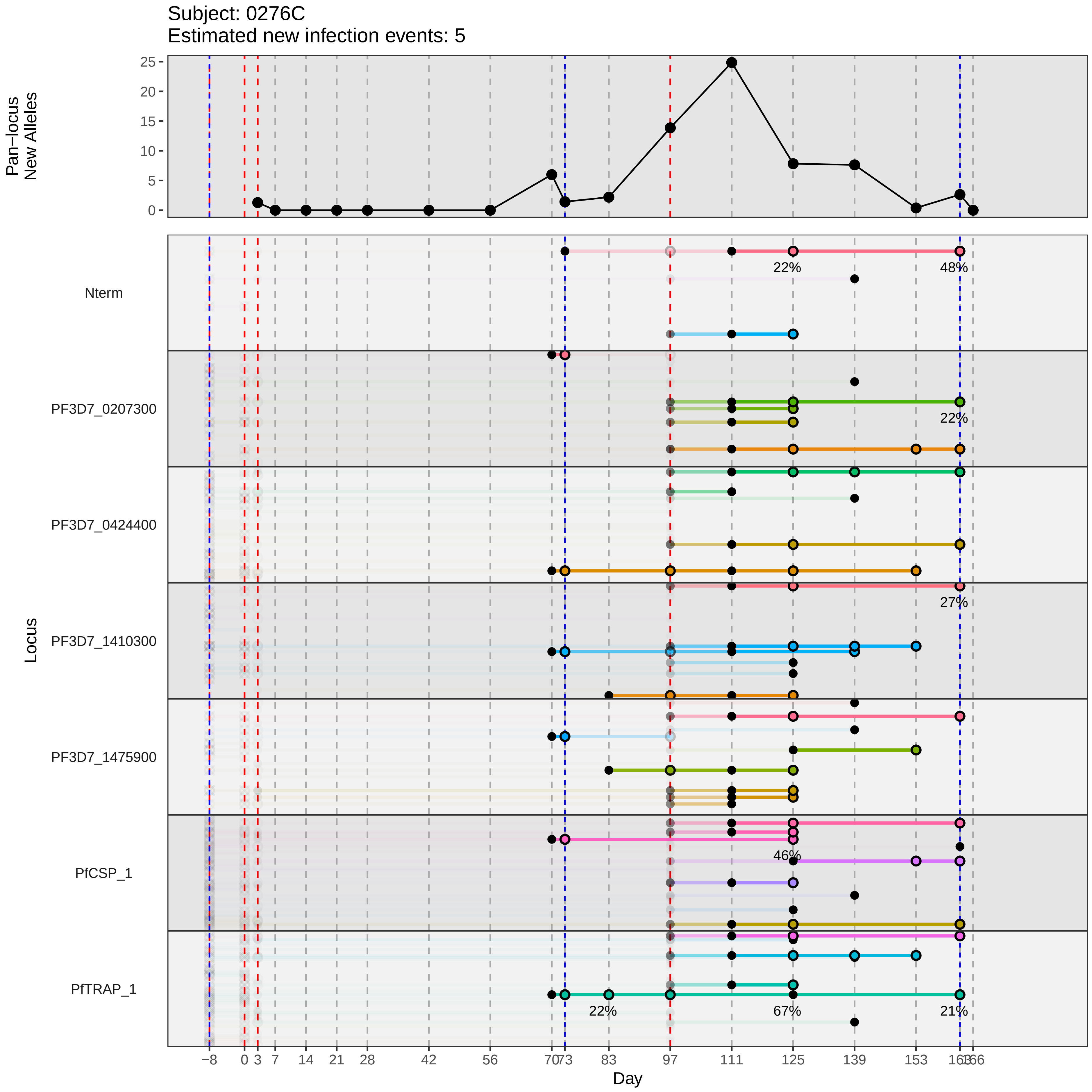

Subject: 0276C
Estimated new infection events: 5
Pan−locus New Alleles
25
20
15
10
5
0
Nterm
PF3D7_0207300
PF3D7_0424400
PF3D7_1410300
PF3D7_1475900
PfCSP_1
PfTRAP_1
Locus
22%
48%
22%
27%
46%
22%
67%
21%
−8
0
3
7
14
21
28
42
56
70
73
83
97
111
125
139
153
163
166
Day
