## Supplementary material for "Genotypic infection endpoint analysis to understand efficacy and escape potential of the malaria monoclonal antibody CIS43LS": Data File S1: participant_0279F_with_imputation.pdf

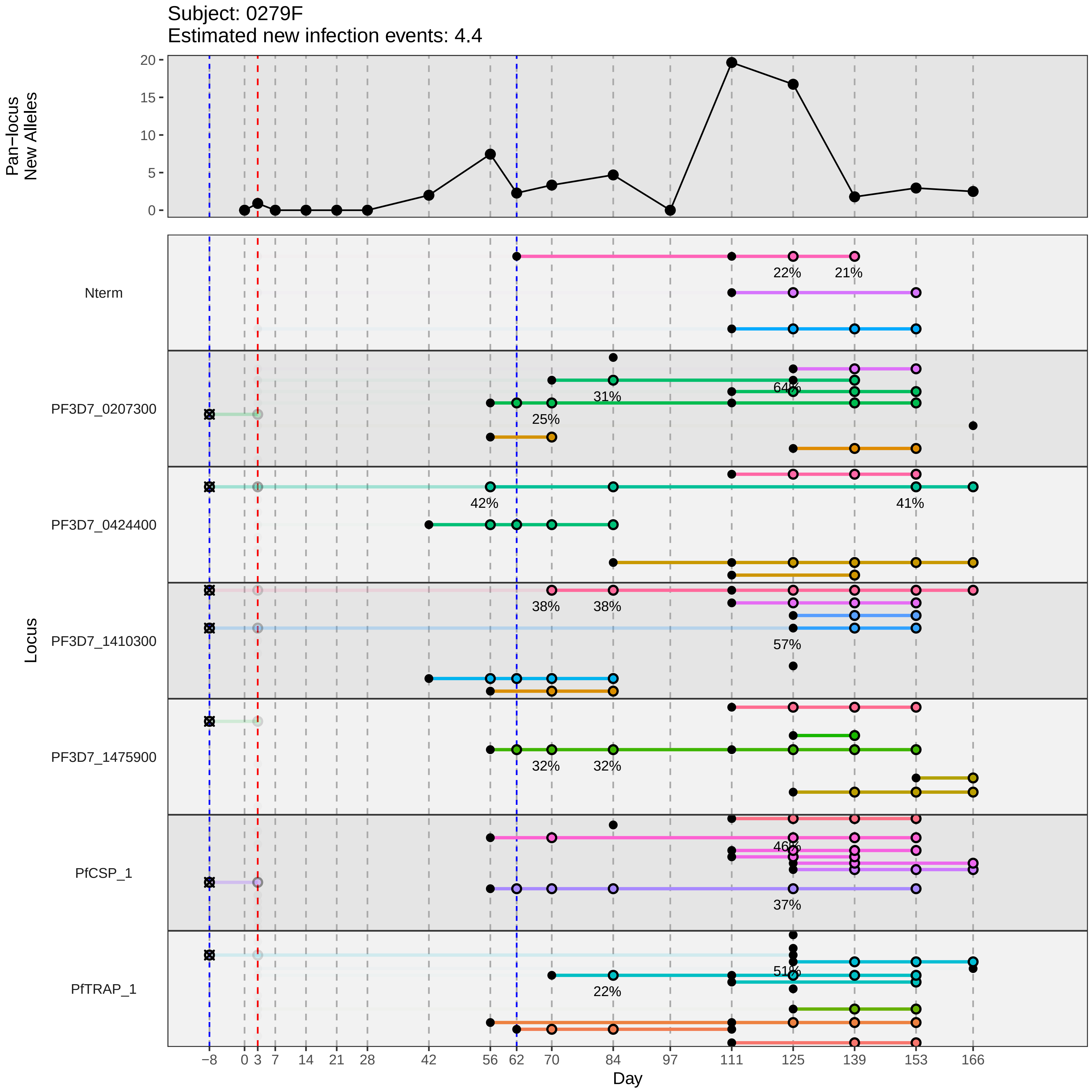

Subject: 0279F
Estimated new infection events: 4.4
Pan−locus New Alleles
20
15
10
5
0
Locus
Nterm
PF3D7_0207300
PF3D7_0424400
PF3D7_1410300
PF3D7_1475900
PfCSP_1
PfTRAP_1
22%
21%
31%
64%
25%
42%
41%
38%
38%
57%
32%
32%
46%
37%
51%
22%
−8
0
3
7
14
21
28
42
56
62
70
84
97
111
125
139
153
166
Day
