## Supplementary material for "Genotypic infection endpoint analysis to understand efficacy and escape potential of the malaria monoclonal antibody CIS43LS": Data File S1: participant_0284N_with_imputation.pdf

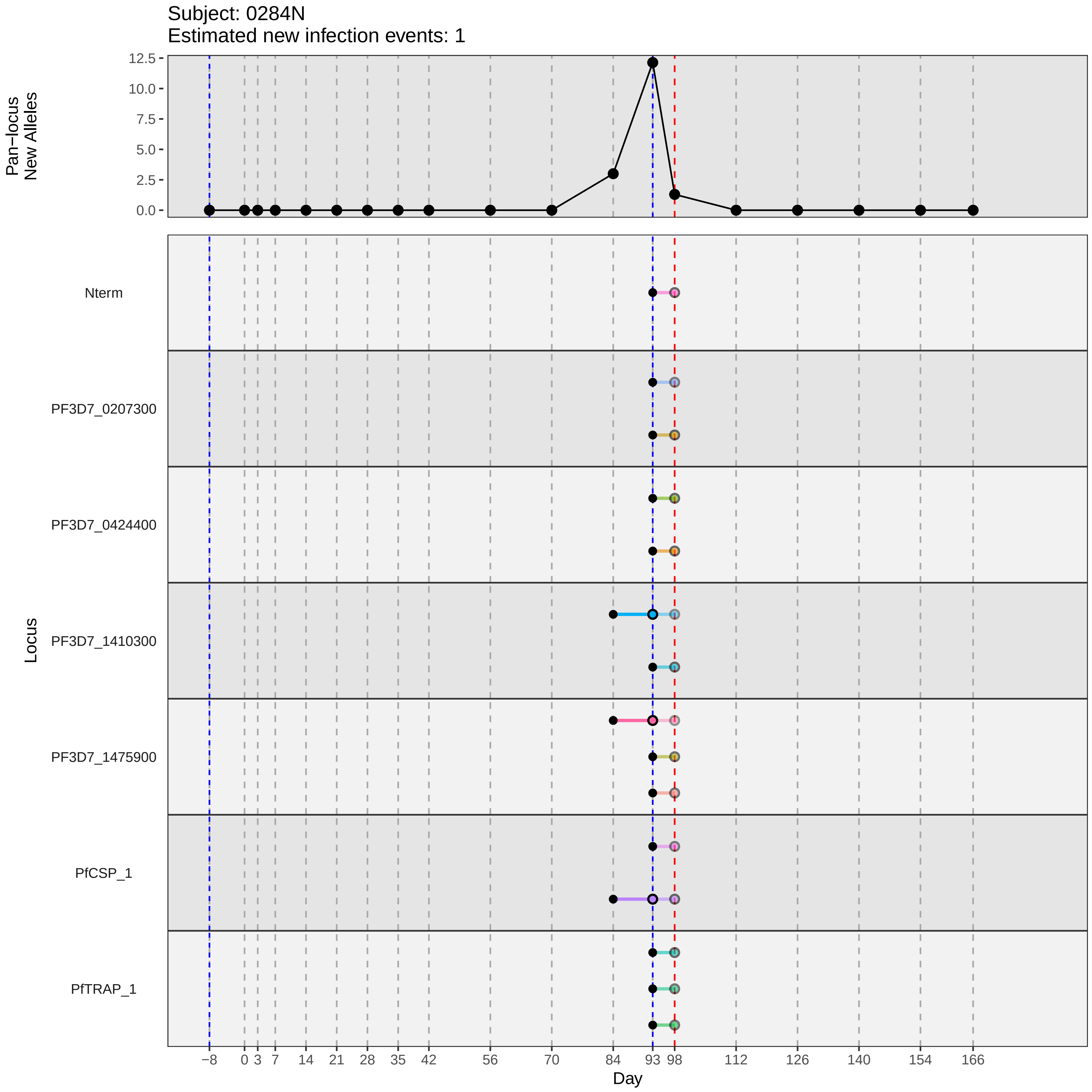

Subject: 0284N
Estimated new infection events: 1
Pan−locus New Alleles
12.5
10.0
7.5
5.0
2.5
0.0
Locus
Nterm
PF3D7_0207300
PF3D7_0424400
PF3D7_1410300
PF3D7_1475900
PfCSP_1
PfTRAP_1
−8 0 3 7 14 21 28 35 42 56 70 84 93 98 112 126 140 154 166
Day
