## Supplementary material for "Genotypic infection endpoint analysis to understand efficacy and escape potential of the malaria monoclonal antibody CIS43LS": Data File S1: participant_0286Q_with_imputation.pdf

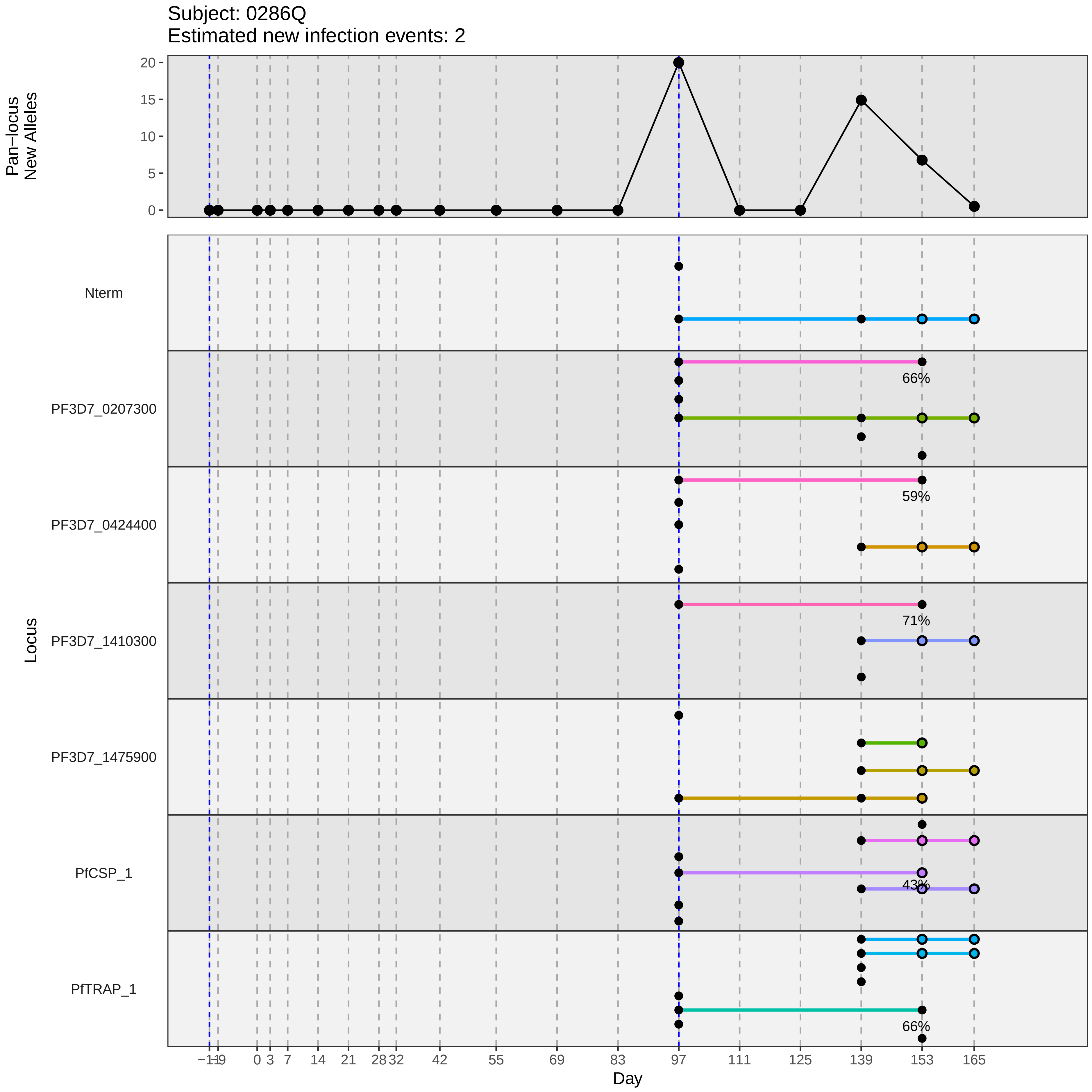

Subject: 0286Q
Estimated new infection events: 2
Pan−locus New Alleles
20
15
10
5
0
Locus
Nterm
PF3D7_0207300
66%
PF3D7_0424400
59%
PF3D7_1410300
71%
PF3D7_1475900
PfCSP_1
43%
PfTRAP_1
66%
−11
−9
0
3
7
14
21
28
32
42
55
69
83
97
111
125
139
153
165
Day
