## Supplementary material for "Genotypic infection endpoint analysis to understand efficacy and escape potential of the malaria monoclonal antibody CIS43LS": Data File S1: participant_0289T_with_imputation.pdf

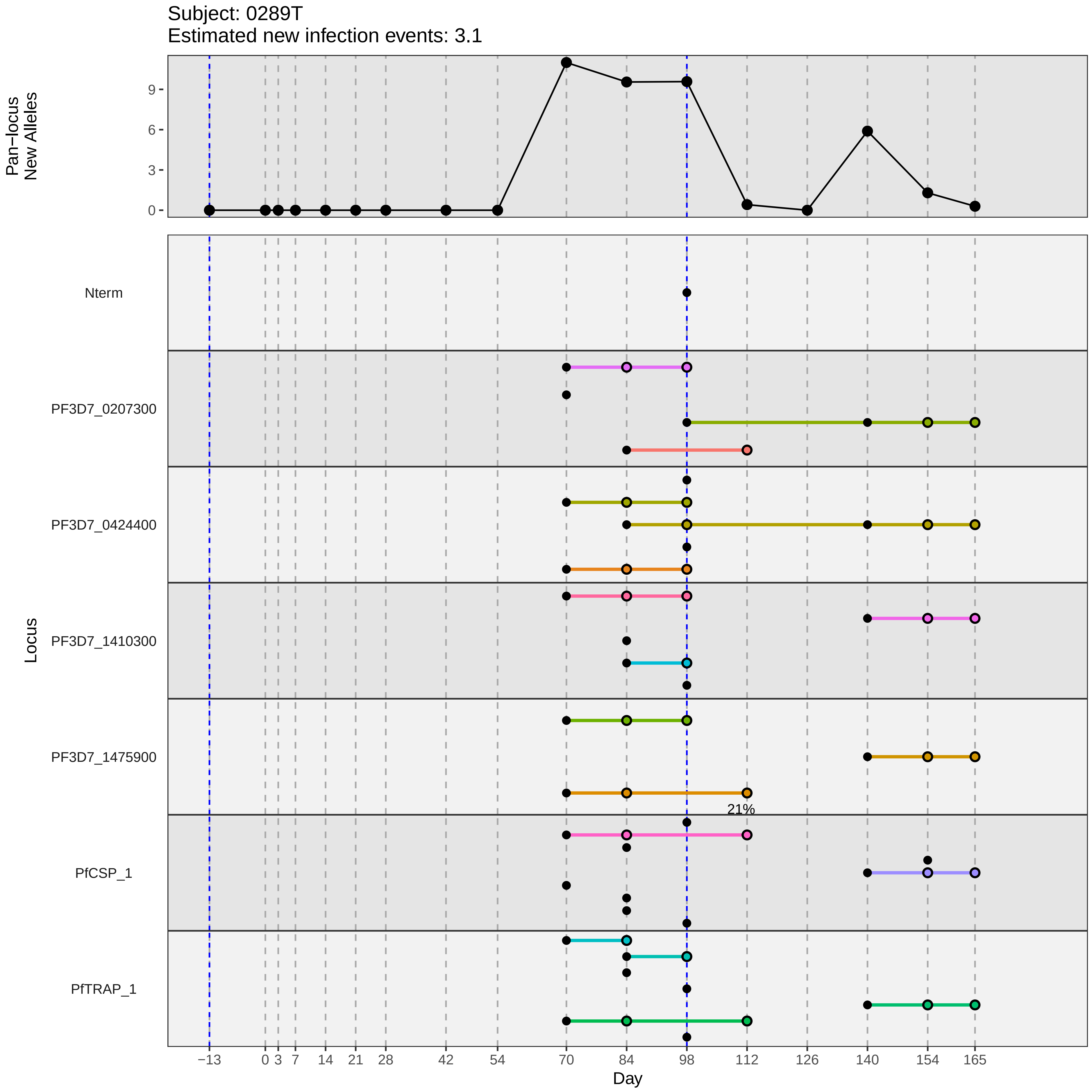
Subject: 0289T
Estimated new infection events: 3.1
Pan−locus New Alleles
9
6
3
0
Locus
Nterm
PF3D7_0207300
PF3D7_0424400
PF3D7_1410300
PF3D7_1475900
PfCSP_1
PfTRAP_1
21%
−13
0
3
7
14
21
28
42
54
70
84
98
112
126
140
154
165
Day
