## Supplementary material for "Genotypic infection endpoint analysis to understand efficacy and escape potential of the malaria monoclonal antibody CIS43LS": Data File S1: participant_0294Y_with_imputation.pdf

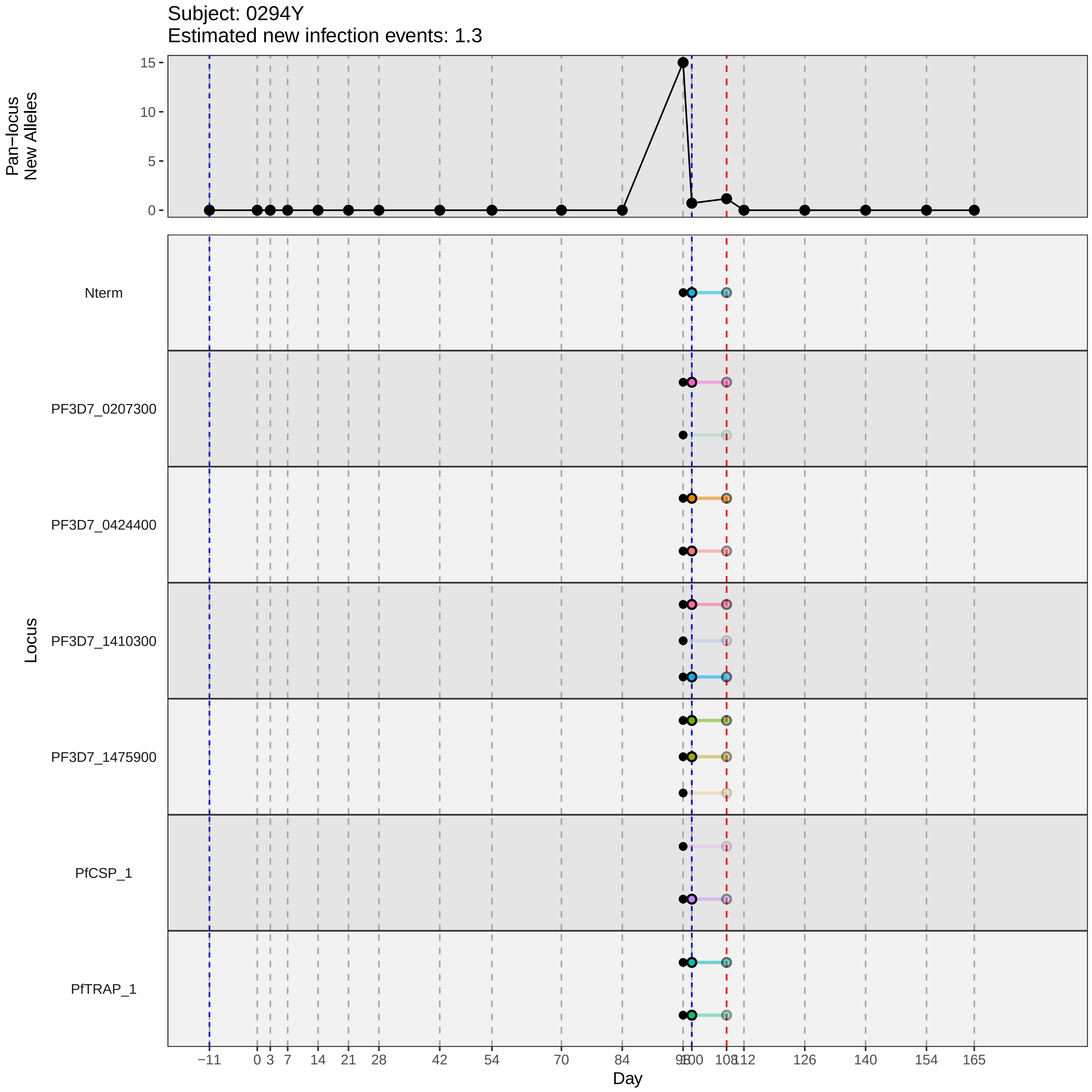

Subject: 0294Y
Estimated new infection events: 1.3
Pan−locus
New Alleles
15
10
5
0
Locus
Nterm
PF3D7_0207300
PF3D7_0424400
PF3D7_1410300
PF3D7_1475900
PfCSP_1
PfTRAP_1
−11
0
3
7
14
21
28
42
54
70
84
98
100
108
112
126
140
154
165
Day
