## Supplementary material for "Genotypic infection endpoint analysis to understand efficacy and escape potential of the malaria monoclonal antibody CIS43LS": Data File S1: participant_0301G_with_imputation.pdf

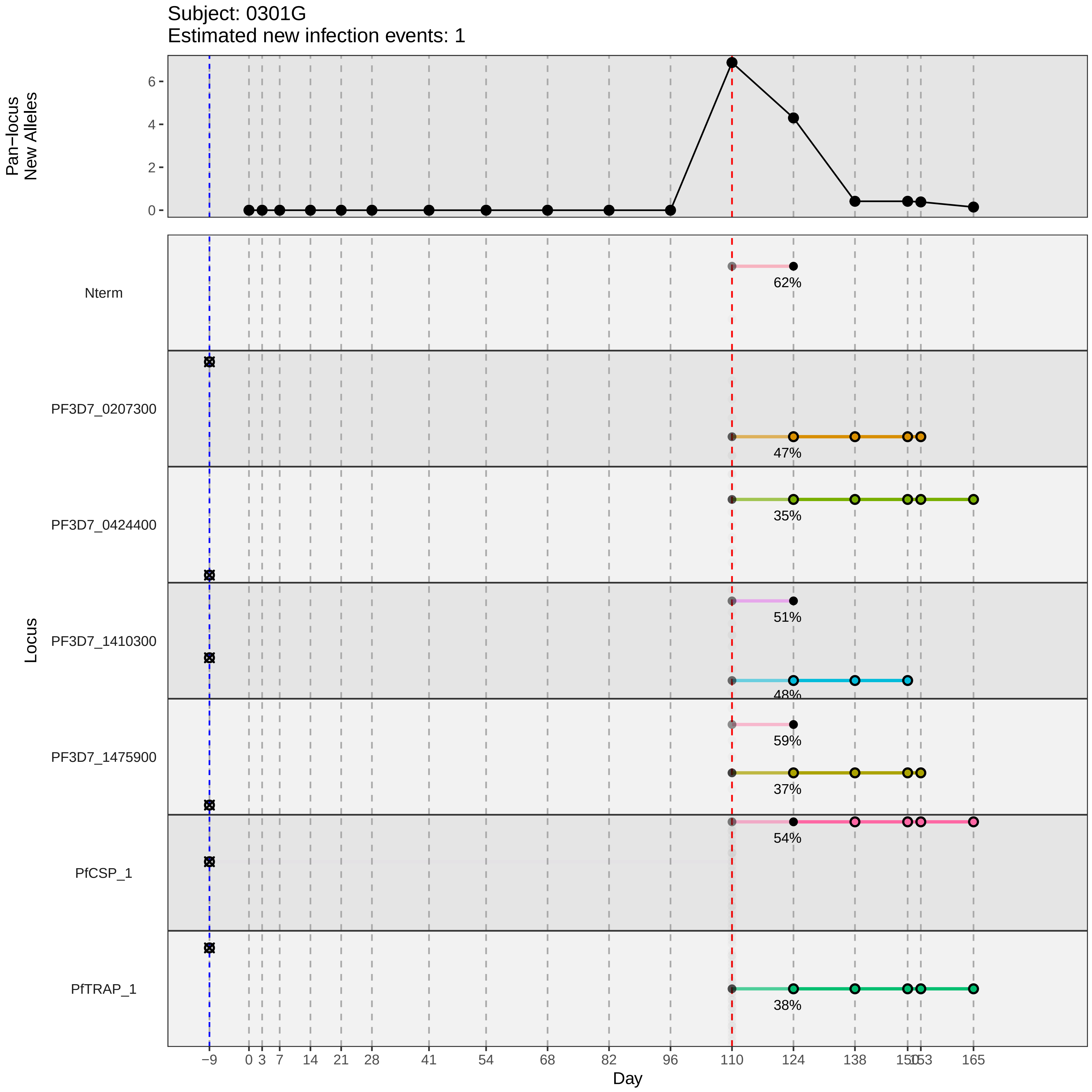

Subject: 0301G
Estimated new infection events: 1
Pan−locus New Alleles
0
2
4
6
Locus
Nterm
PF3D7_0207300
PF3D7_0424400
PF3D7_1410300
PF3D7_1475900
PfCSP_1
PfTRAP_1
62%
47%
35%
51%
48%
59%
37%
54%
38%
−9
0
3
7
14
21
28
41
54
68
82
96
110
124
138
150
153
165
Day
