## Supplementary material for "Genotypic infection endpoint analysis to understand efficacy and escape potential of the malaria monoclonal antibody CIS43LS": Data File S1: participant_0302H_with_imputation.pdf

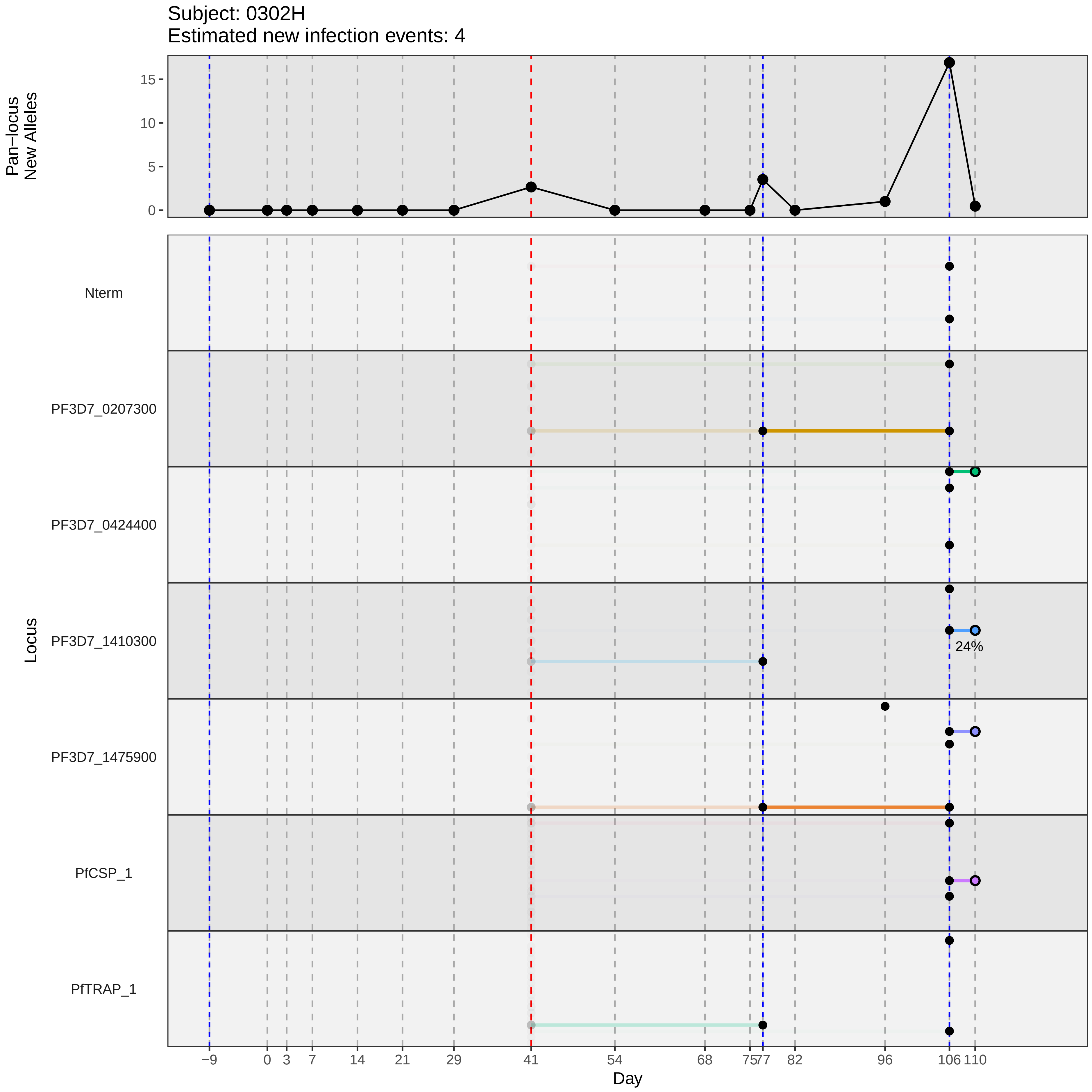

Subject: 0302H
Estimated new infection events: 4
Pan−locus New Alleles
0
5
10
15
Locus
Nterm
PF3D7_0207300
PF3D7_0424400
PF3D7_1410300
PF3D7_1475900
PfCSP_1
PfTRAP_1
24%
−9
0
3
7
14
21
29
41
54
68
75
77
82
96
106
110
Day
