## Supplementary material for "Genotypic infection endpoint analysis to understand efficacy and escape potential of the malaria monoclonal antibody CIS43LS": Data File S1: participant_0303K_with_imputation.pdf

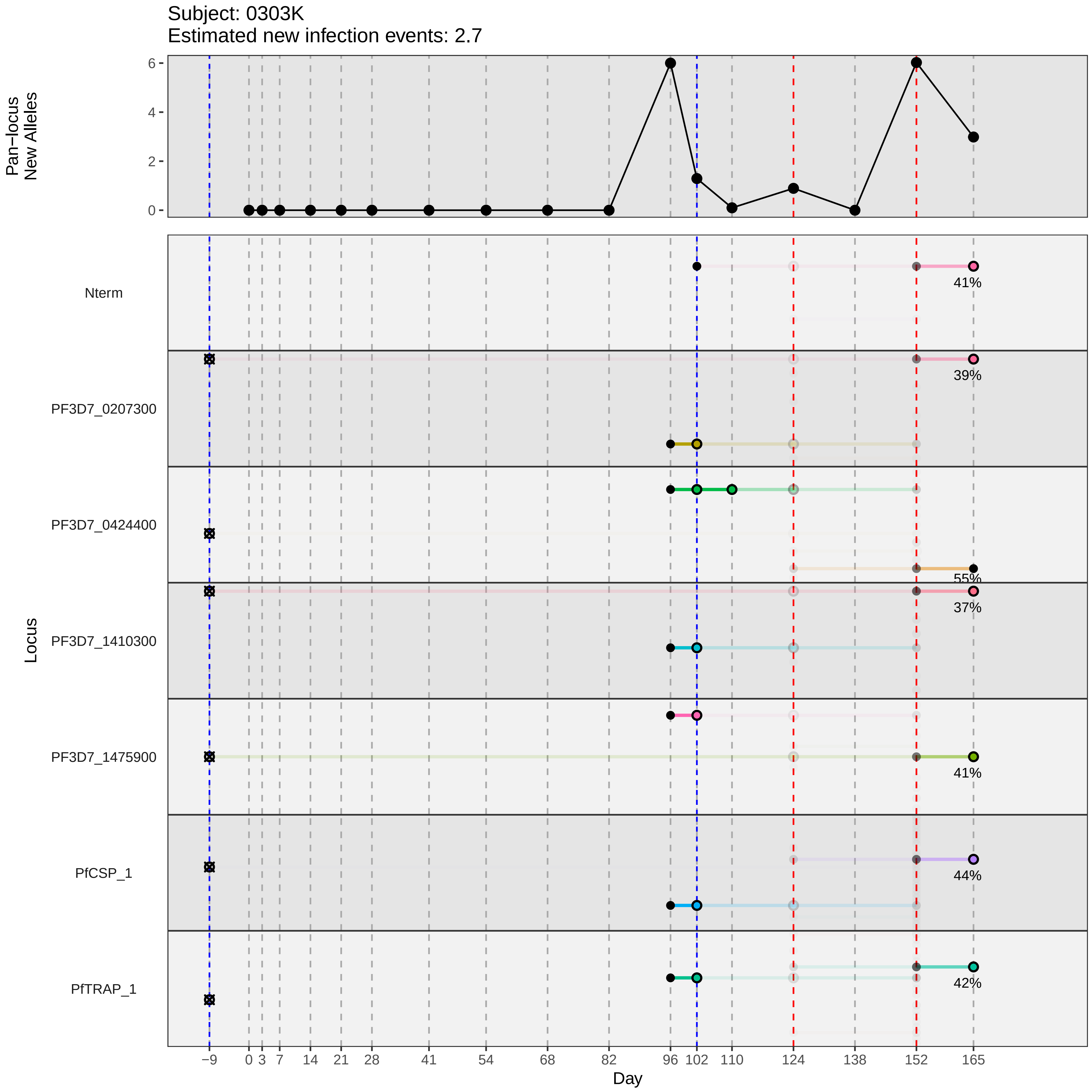

Subject: 0303K
Estimated new infection events: 2.7
Pan−locus
New Alleles
6
4
2
0
Nterm
41%
PF3D7_0207300
39%
PF3D7_0424400
55%
PF3D7_1410300
37%
PF3D7_1475900
41%
PfCSP_1
44%
PfTRAP_1
42%
Locus
−9
0
3
7
14
21
28
41
54
68
82
96
102
110
124
138
152
165
Day
