## Supplementary material for "Genotypic infection endpoint analysis to understand efficacy and escape potential of the malaria monoclonal antibody CIS43LS": Data File S1: participant_0306P_with_imputation.pdf

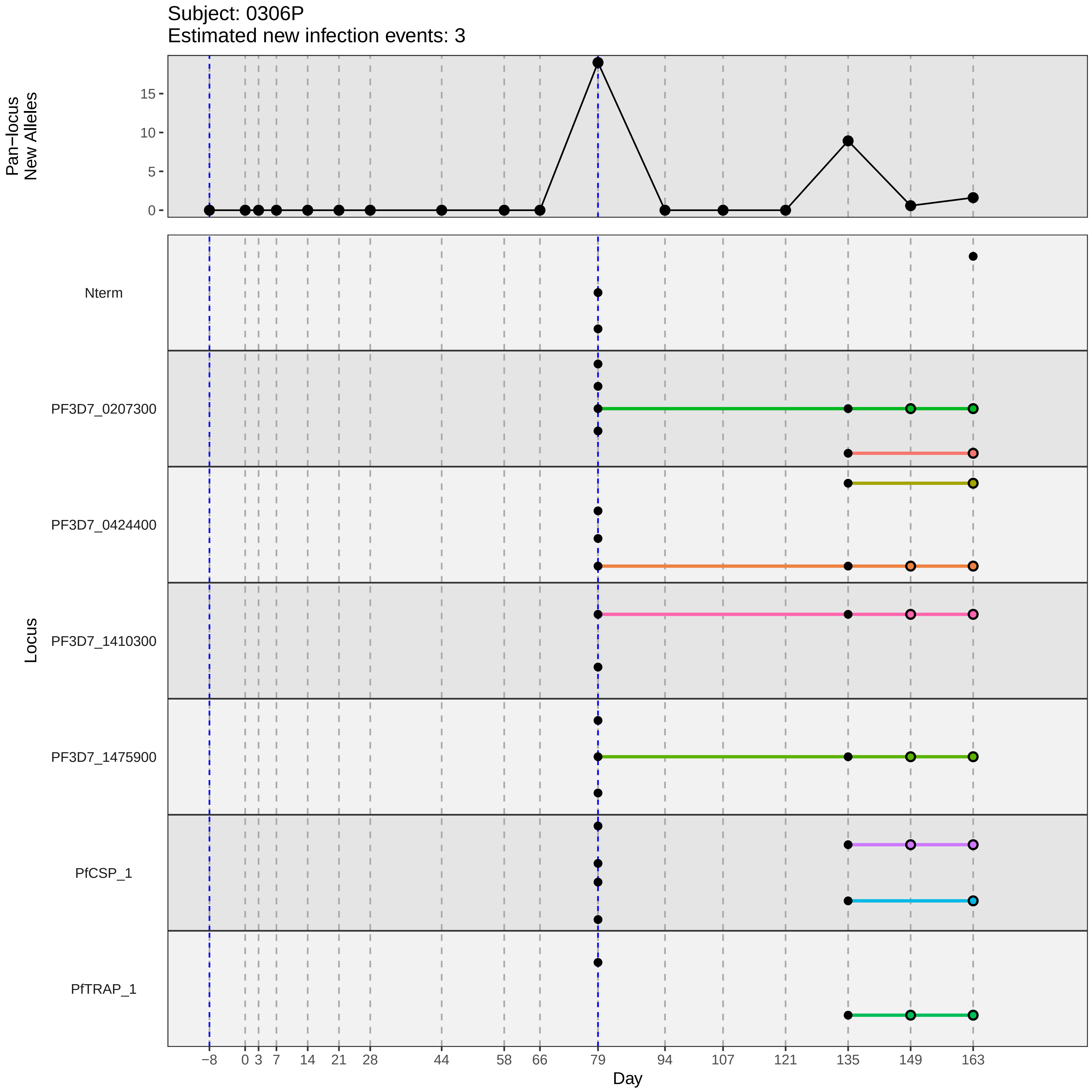

Subject: 0306P
Estimated new infection events: 3
Pan−locus New Alleles
0
5
10
15
Locus
Nterm
PF3D7_0207300
PF3D7_0424400
PF3D7_1410300
PF3D7_1475900
PfCSP_1
PfTRAP_1
−8
0
3
7
14
21
28
44
58
66
79
94
107
121
135
149
163
Day
