## Supplementary material for "Genotypic infection endpoint analysis to understand efficacy and escape potential of the malaria monoclonal antibody CIS43LS": Data File S1: participant_0309S_with_imputation.pdf

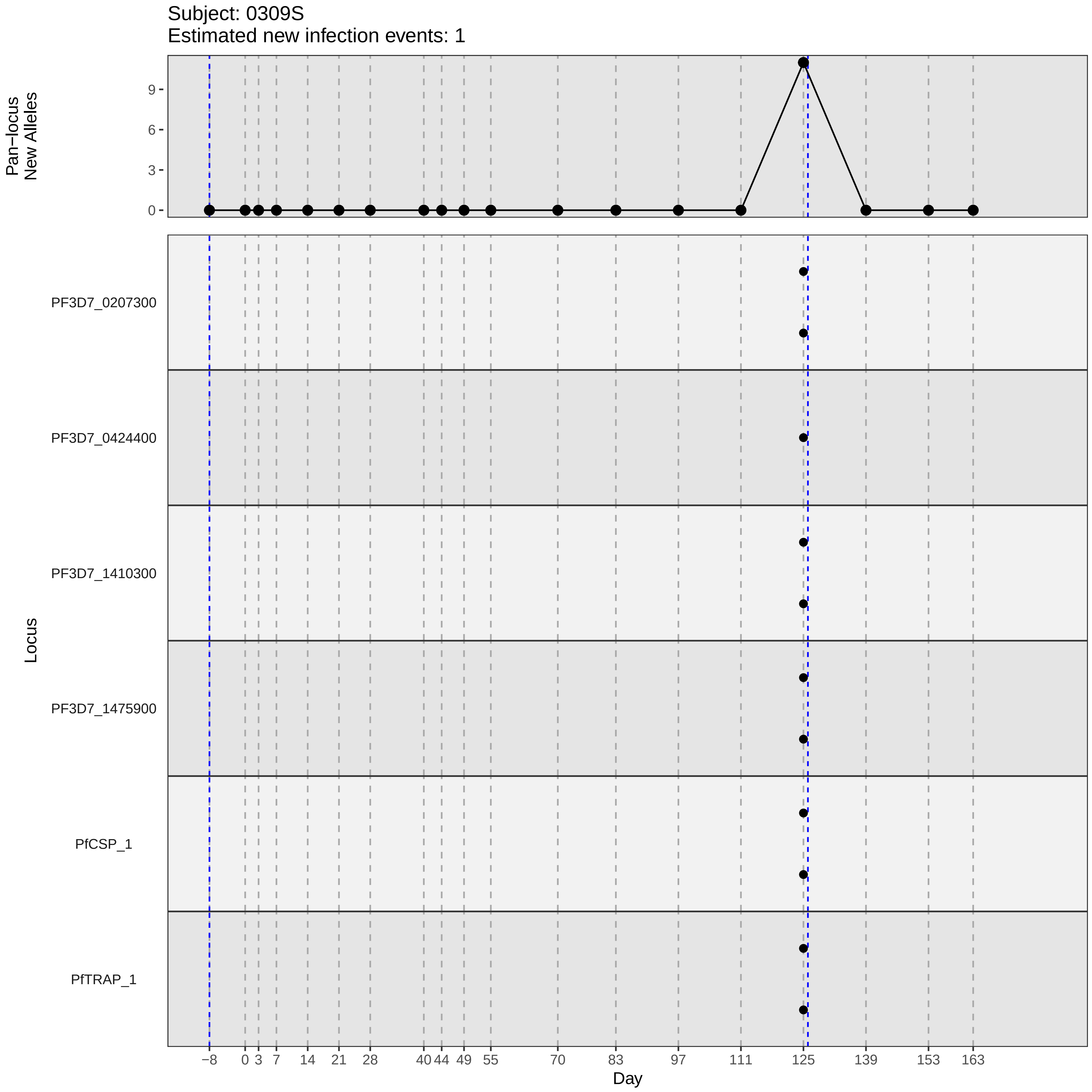

Subject: 0309S
Estimated new infection events: 1
Pan−locus
New Alleles
0
3
6
9
Locus
PF3D7_0207300
PF3D7_0424400
PF3D7_1410300
PF3D7_1475900
PfCSP_1
PfTRAP_1
−8
0
3
7
14
21
28
40
44
49
55
70
83
97
111
125
139
153
163
Day
