## Supplementary material for "Genotypic infection endpoint analysis to understand efficacy and escape potential of the malaria monoclonal antibody CIS43LS": Data File S1: participant_0313W_with_imputation.pdf

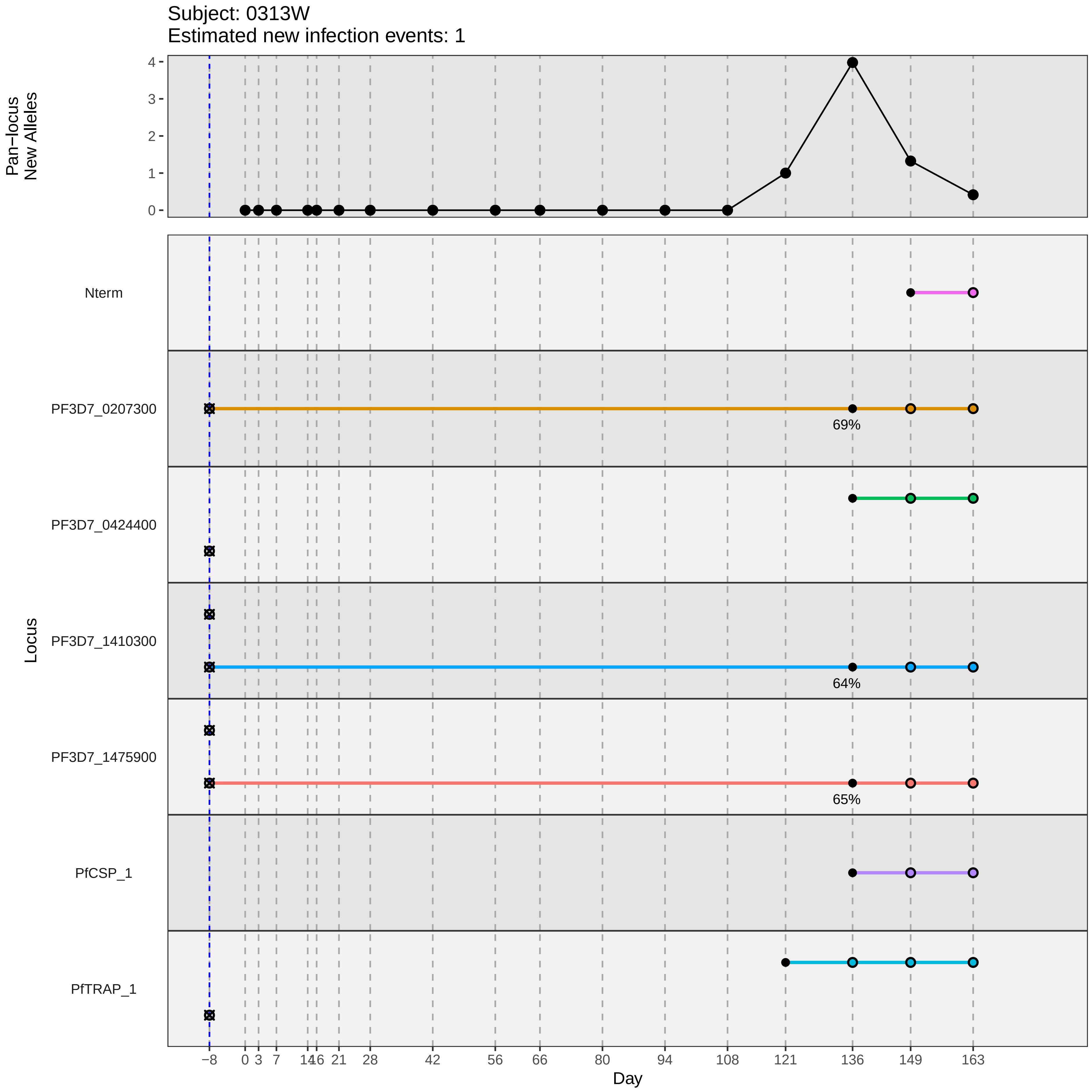

Subject: 0313W
Estimated new infection events: 1
Pan−locus
New Alleles
0
1
2
3
4
Locus
Nterm
PF3D7_0207300
PF3D7_0424400
PF3D7_1410300
PF3D7_1475900
PfCSP_1
PfTRAP_1
69%
64%
65%
−8
0
3
7
14
16
21
28
42
56
66
80
94
108
121
136
149
163
Day
