## Supplementary material for "Genotypic infection endpoint analysis to understand efficacy and escape potential of the malaria monoclonal antibody CIS43LS": Data File S1: participant_0319D_with_imputation.pdf

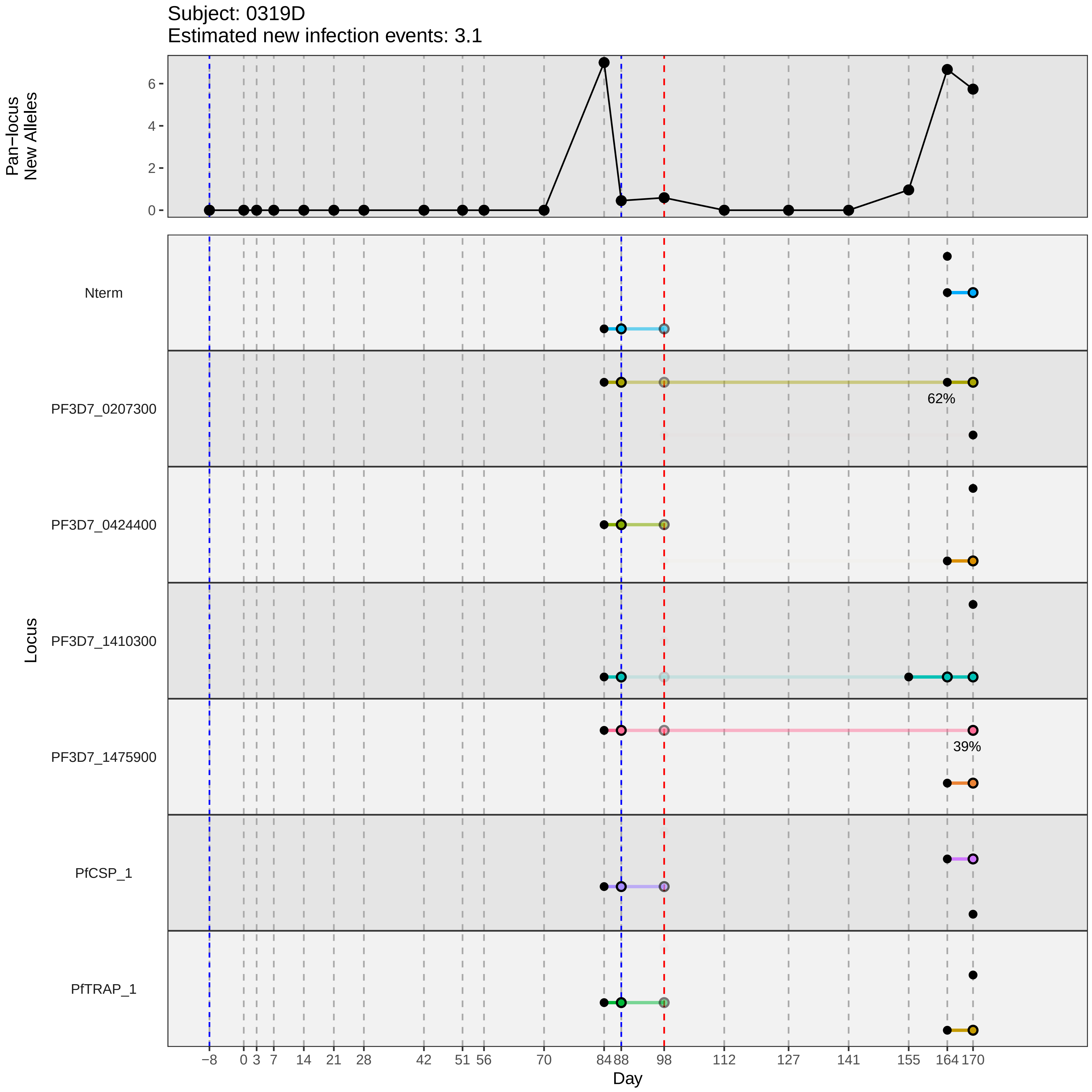

Subject: 0319D
Estimated new infection events: 3.1
Pan−locus New Alleles
0
2
4
6
Locus
Nterm
PF3D7_0207300
62%
PF3D7_0424400
PF3D7_1410300
PF3D7_1475900
39%
PfCSP_1
PfTRAP_1
−8
0
3
7
14
21
28
42
51
56
70
84
88
98
112
127
141
155
164
170
Day
