## Supplementary material for "Genotypic infection endpoint analysis to understand efficacy and escape potential of the malaria monoclonal antibody CIS43LS": Data File S1: participant_0321F_with_imputation.pdf

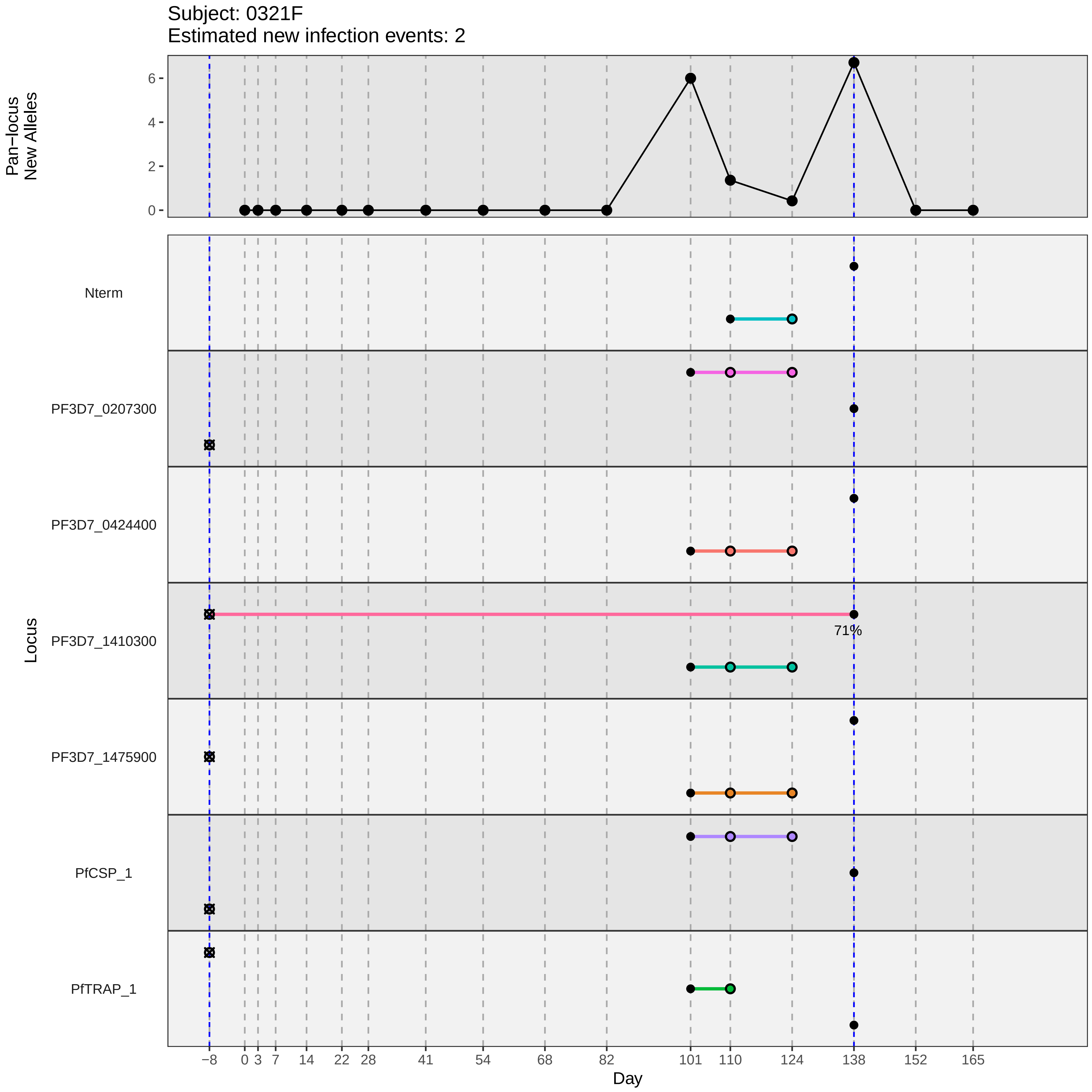

Subject: 0321F
Estimated new infection events: 2
Pan−locus
New Alleles
0
2
4
6
Locus
Nterm
PF3D7_0207300
PF3D7_0424400
PF3D7_1410300
71%
PF3D7_1475900
PfCSP_1
PfTRAP_1
−8
0
3
7
14
22
28
41
54
68
82
101
110
124
138
152
165
Day
