## Supplementary material for "Genotypic infection endpoint analysis to understand efficacy and escape potential of the malaria monoclonal antibody CIS43LS": Data File S1: participant_0325M_with_imputation.pdf

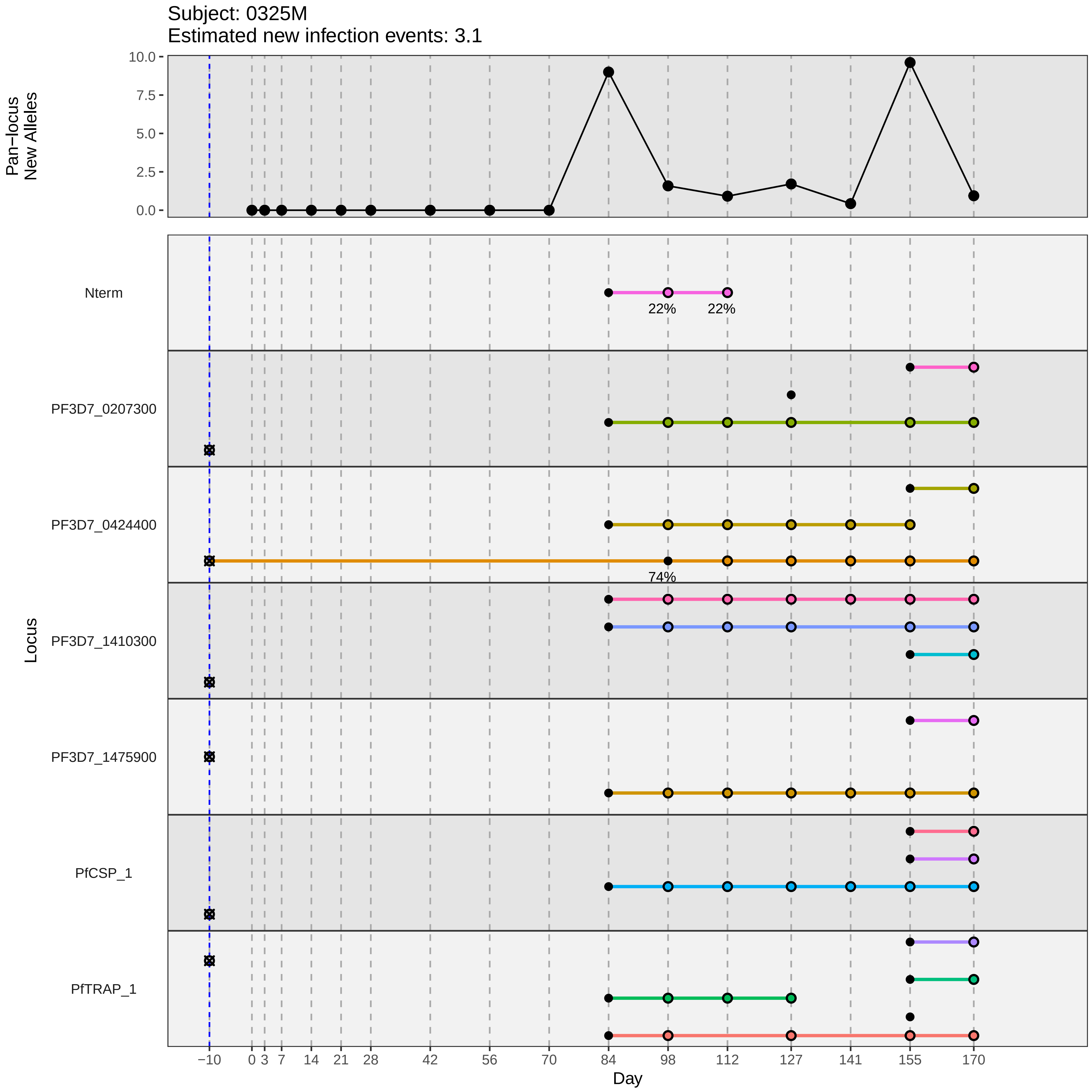
Subject: 0325M
Estimated new infection events: 3.1
Pan−locus
New Alleles
10.0
7.5
5.0
2.5
0.0
Nterm
22%
22%
PF3D7_0207300
PF3D7_0424400
74%
PF3D7_1410300
PF3D7_1475900
PfCSP_1
PfTRAP_1
Locus
−10
0
3
7
14
21
28
42
56
70
84
98
112
127
141
155
170
Day
