## Supplementary material for "Genotypic infection endpoint analysis to understand efficacy and escape potential of the malaria monoclonal antibody CIS43LS": Data File S1: participant_0333V_with_imputation.pdf

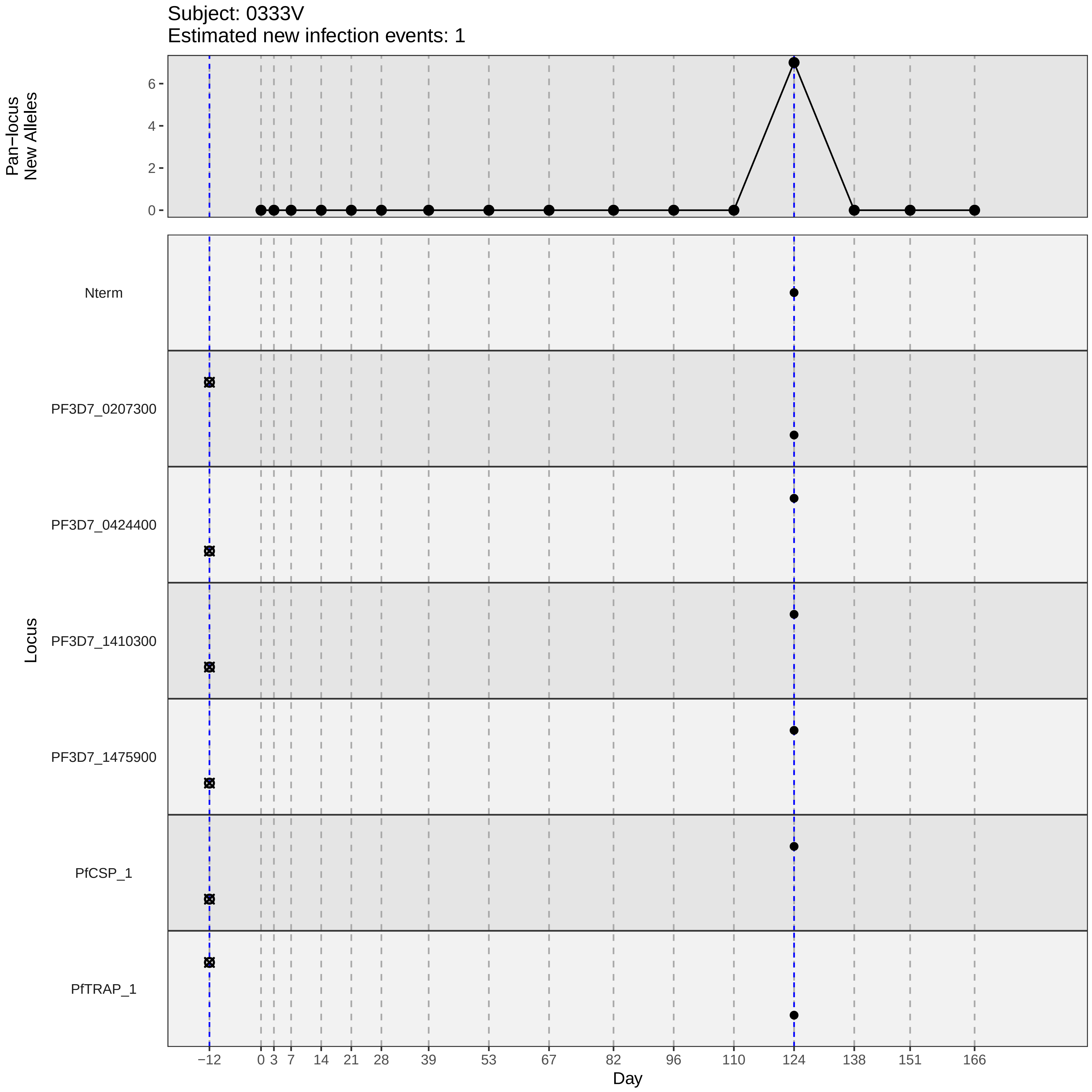

Subject: 0333V
Estimated new infection events: 1
Pan−locus
New Alleles
0
2
4
6
Locus
Nterm
PF3D7_0207300
PF3D7_0424400
PF3D7_1410300
PF3D7_1475900
PfCSP_1
PfTRAP_1
−12
0
3
7
14
21
28
39
53
67
82
96
110
124
138
151
166
Day
