## Supplementary material for "Genotypic infection endpoint analysis to understand efficacy and escape potential of the malaria monoclonal antibody CIS43LS": Data File S1: participant_0334W_with_imputation.pdf

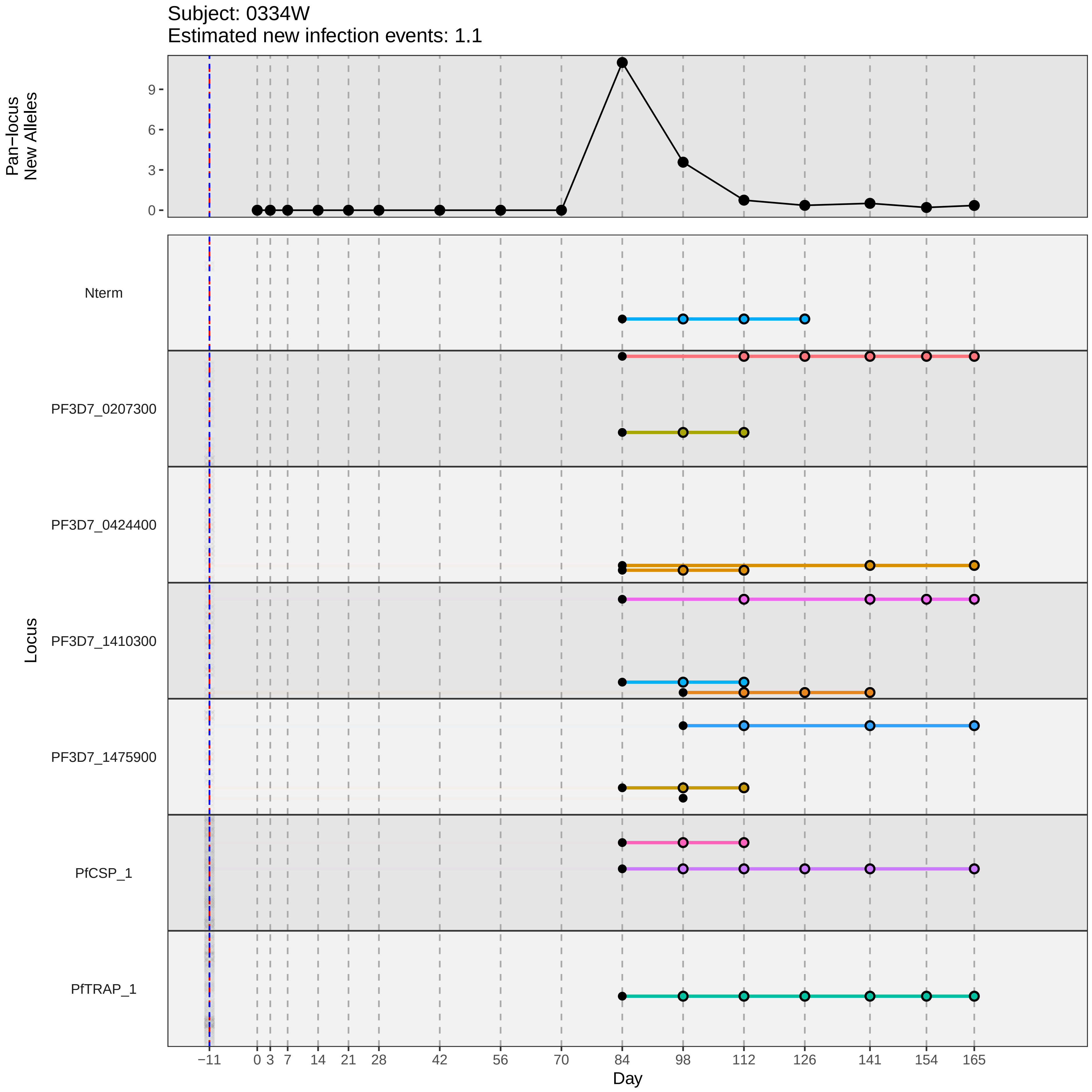

Subject: 0334W
Estimated new infection events: 1.1
Pan−locus
New Alleles
0
3
6
9
Locus
Nterm
PF3D7_0207300
PF3D7_0424400
PF3D7_1410300
PF3D7_1475900
PfCSP_1
PfTRAP_1
−11
0
3
7
14
21
28
42
56
70
84
98
112
126
141
154
165
Day
