## Supplementary material for "Genotypic infection endpoint analysis to understand efficacy and escape potential of the malaria monoclonal antibody CIS43LS": Data File S1: participant_0336Y_with_imputation.pdf

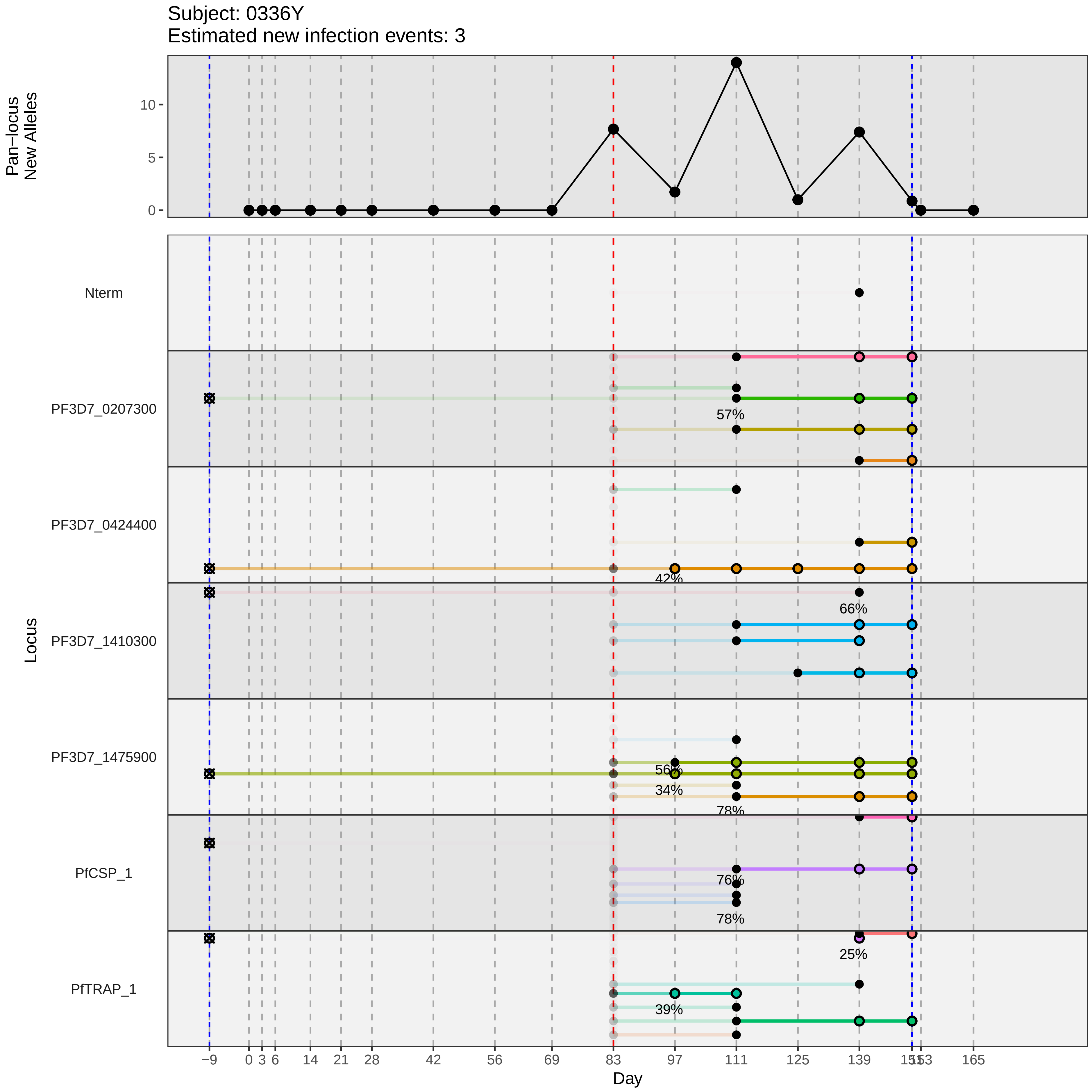

Subject: 0336Y
Estimated new infection events: 3
Pan−locus New Alleles
0
5
10
Locus
Nterm
PF3D7_0207300
PF3D7_0424400
PF3D7_1410300
PF3D7_1475900
PfCSP_1
PfTRAP_1
57%
42%
66%
56%
34%
78%
76%
78%
25%
39%
−9
0
3
6
14
21
28
42
56
69
83
97
111
125
139
153
165
Day
