## Supplementary material for "Genotypic infection endpoint analysis to understand efficacy and escape potential of the malaria monoclonal antibody CIS43LS": Data File S1: participant_0337A_with_imputation.pdf

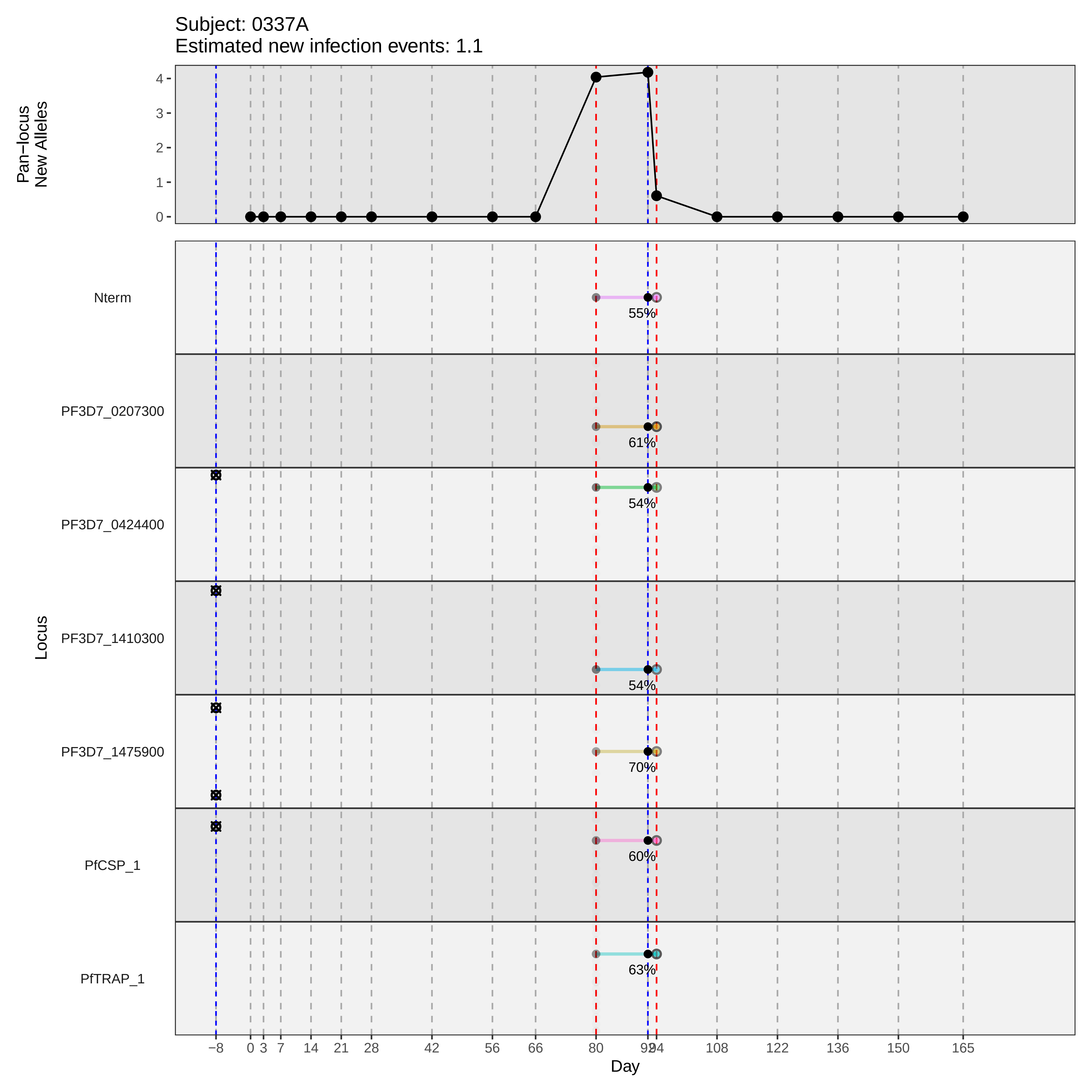

Subject: 0337A
Estimated new infection events: 1.1
Pan−locus New Alleles
0
1
2
3
4
Locus
Nterm
PF3D7_0207300
PF3D7_0424400
PF3D7_1410300
PF3D7_1475900
PfCSP_1
PfTRAP_1
55%
61%
54%
54%
70%
60%
63%
−8
0
3
7
14
21
28
42
56
66
80
92
94
108
122
136
150
165
Day
