## Supplementary material for "Genotypic infection endpoint analysis to understand efficacy and escape potential of the malaria monoclonal antibody CIS43LS": Data File S1: participant_0341E_with_imputation.pdf

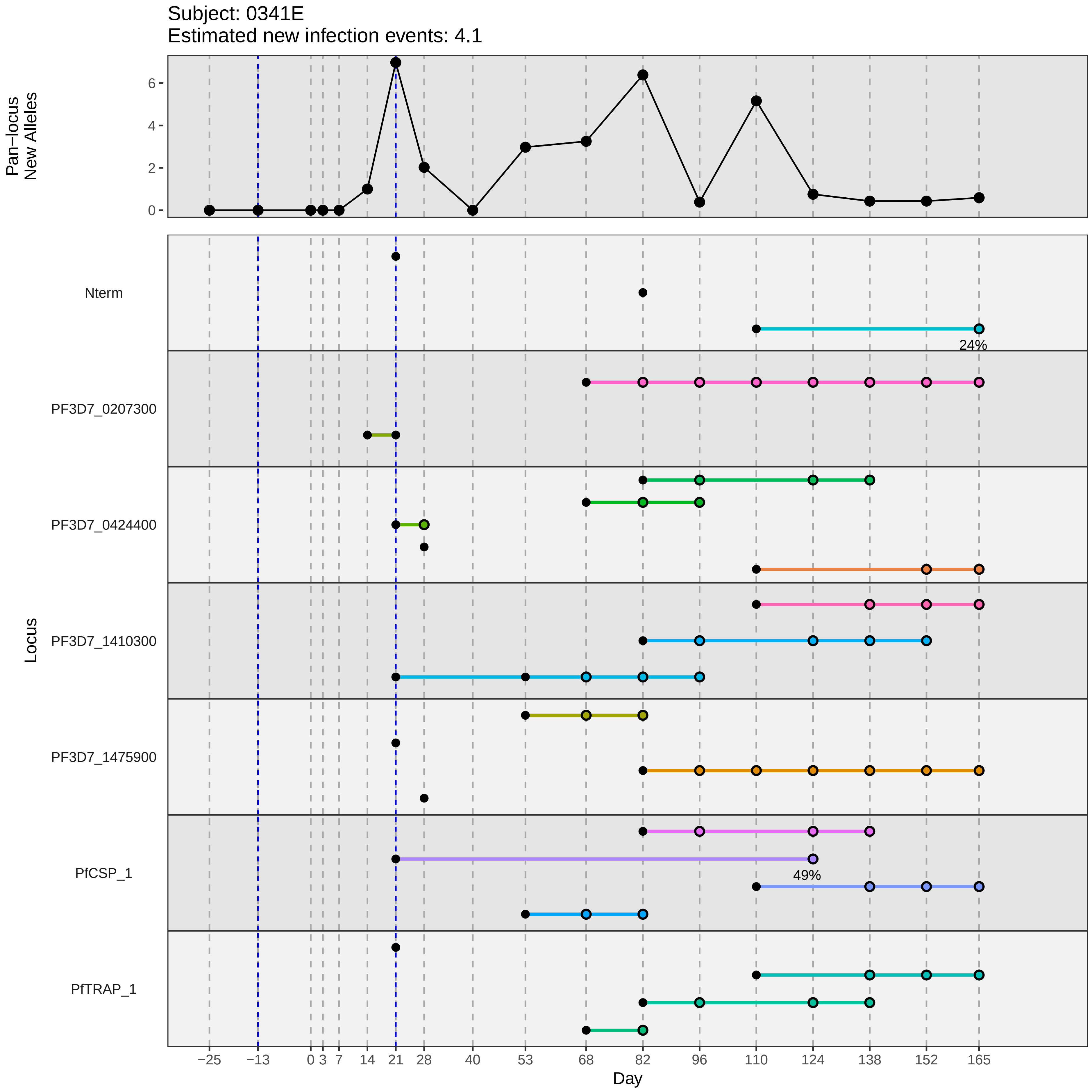

Subject: 0341E
Estimated new infection events: 4.1
Pan−locus New Alleles
Locus
Nterm
PF3D7_0207300
PF3D7_0424400
PF3D7_1410300
PF3D7_1475900
PfCSP_1
PfTRAP_1
24%
49%
Day
−25
−13
0
3
7
14
21
28
40
53
68
82
96
110
124
138
152
165
0
2
4
6
