## Supplementary material for "Genotypic infection endpoint analysis to understand efficacy and escape potential of the malaria monoclonal antibody CIS43LS": Data File S1: participant_0342F_with_imputation.pdf

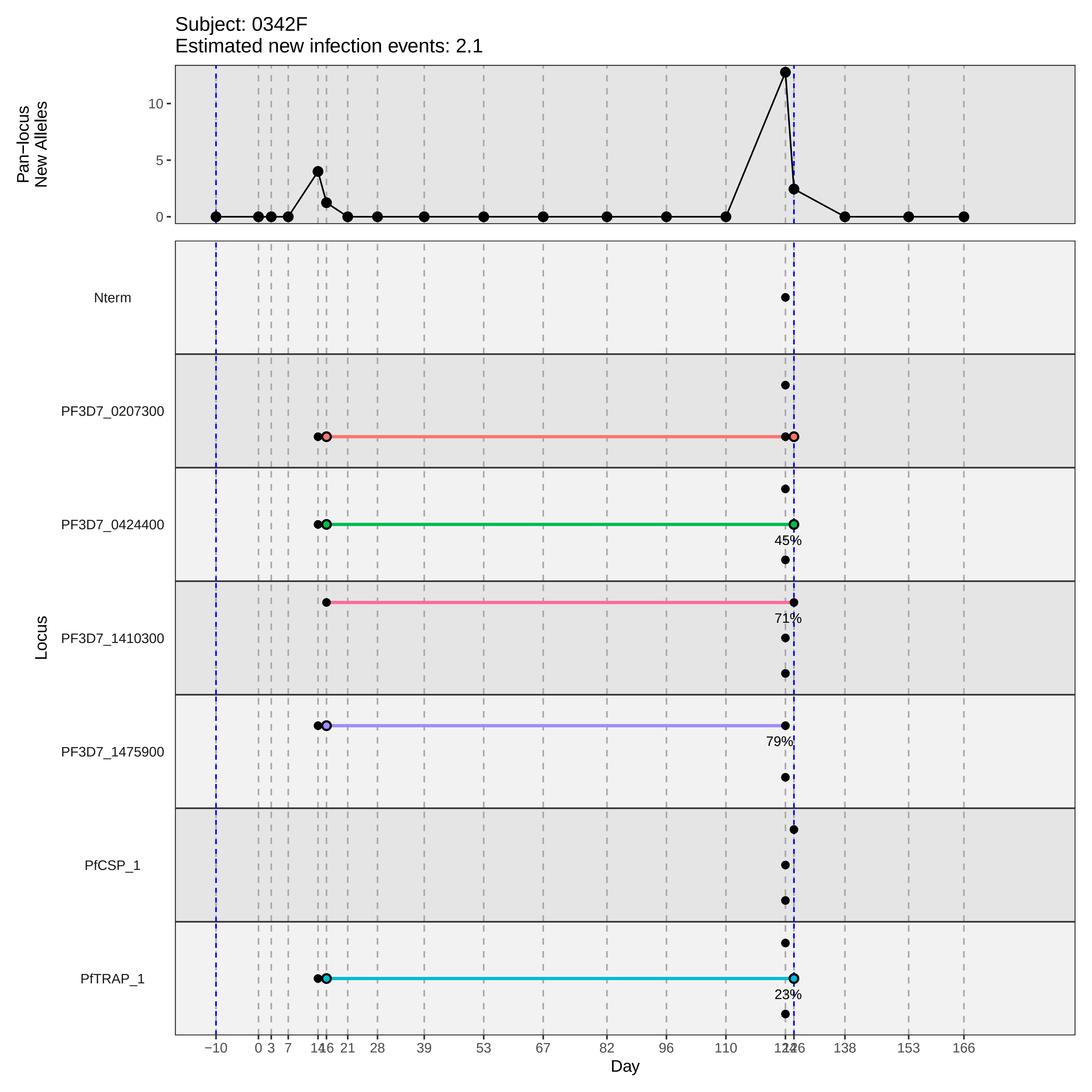

Subject: 0342F
Estimated new infection events: 2.1
Pan−locus
New Alleles
10
5
0
Nterm
PF3D7_0207300
PF3D7_0424400
45%
PF3D7_1410300
71%
PF3D7_1475900
79%
PfCSP_1
PfTRAP_1
23%
Locus
−10
0
3
7
14
16
21
28
39
53
67
82
96
110
124
126
138
153
166
Day
