## Supplementary material for "Genotypic infection endpoint analysis to understand efficacy and escape potential of the malaria monoclonal antibody CIS43LS": Data File S1: participant_0345K_with_imputation.pdf

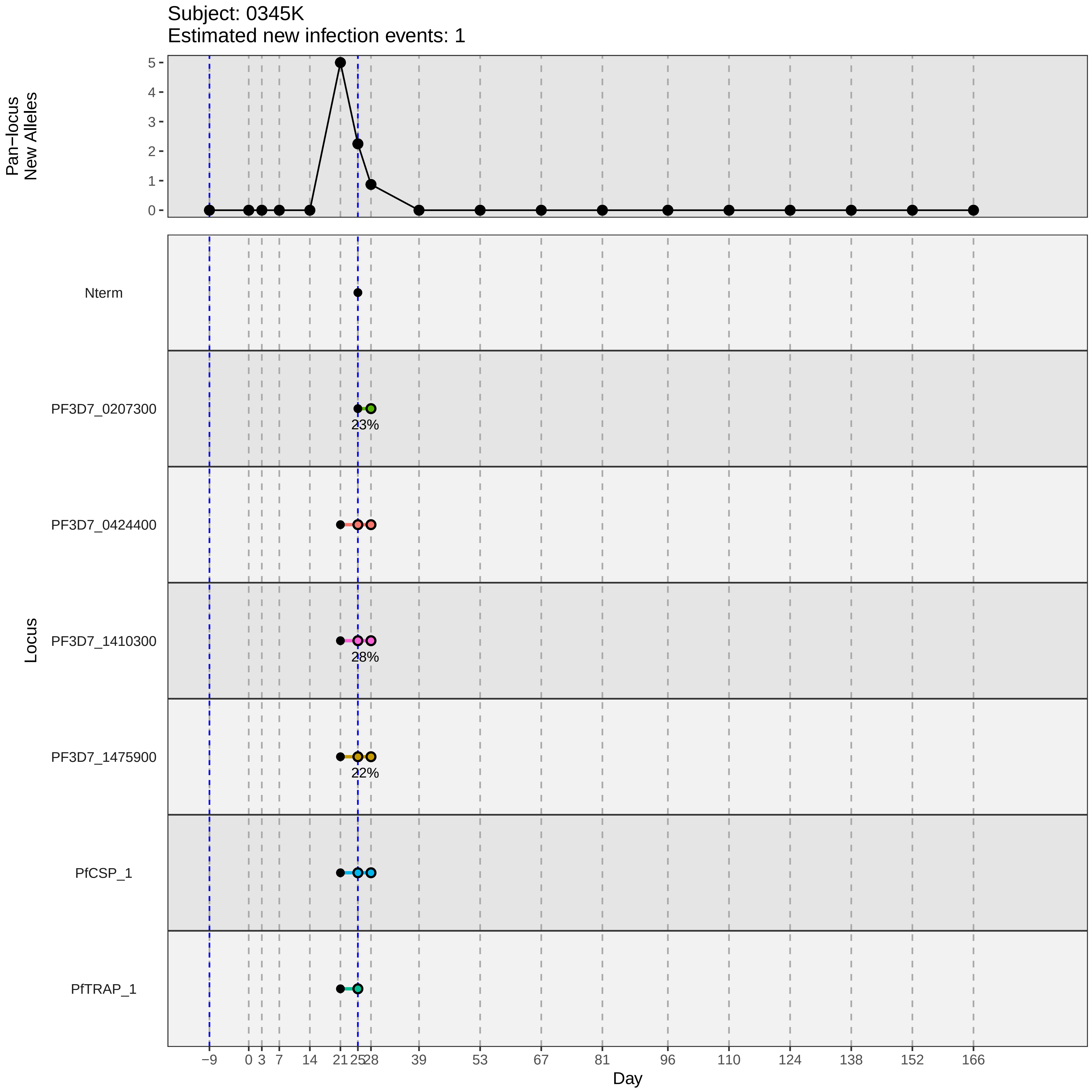
Subject: 0345K
Estimated new infection events: 1
Pan−locus New Alleles
5
4
3
2
1
0
Locus
Nterm
PF3D7_0207300
23%
PF3D7_0424400
PF3D7_1410300
28%
PF3D7_1475900
22%
PfCSP_1
PfTRAP_1
−9
0
3
7
14
21
25
28
39
53
67
81
96
110
124
138
152
166
Day
