## Supplementary material for "Genotypic infection endpoint analysis to understand efficacy and escape potential of the malaria monoclonal antibody CIS43LS": Data File S1: participant_0348P_with_imputation.pdf

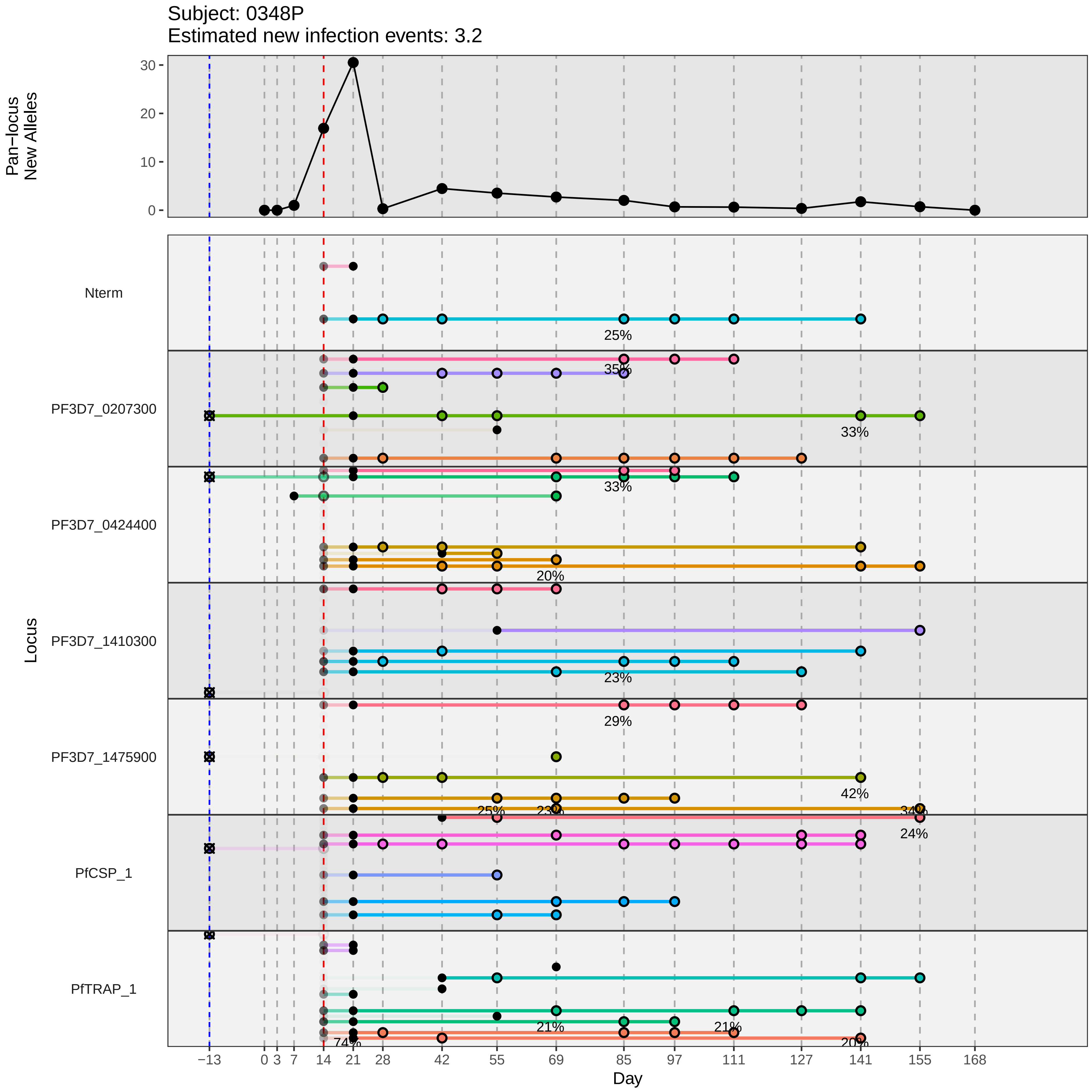
Subject: 0348P
Estimated new infection events: 3.2
Pan−locus New Alleles
30
20
10
0
Locus
Nterm
PF3D7_0207300
PF3D7_0424400
PF3D7_1410300
PF3D7_1475900
PfCSP_1
PfTRAP_1
25%
35%
33%
33%
20%
23%
29%
42%
25%
23%
34%
24%
21%
21%
74%
20%
−13
0
3
7
14
21
28
42
55
69
85
97
111
127
141
155
168
Day
