## Supplementary material for "Genotypic infection endpoint analysis to understand efficacy and escape potential of the malaria monoclonal antibody CIS43LS": Data File S1: participant_0352T_with_imputation.pdf

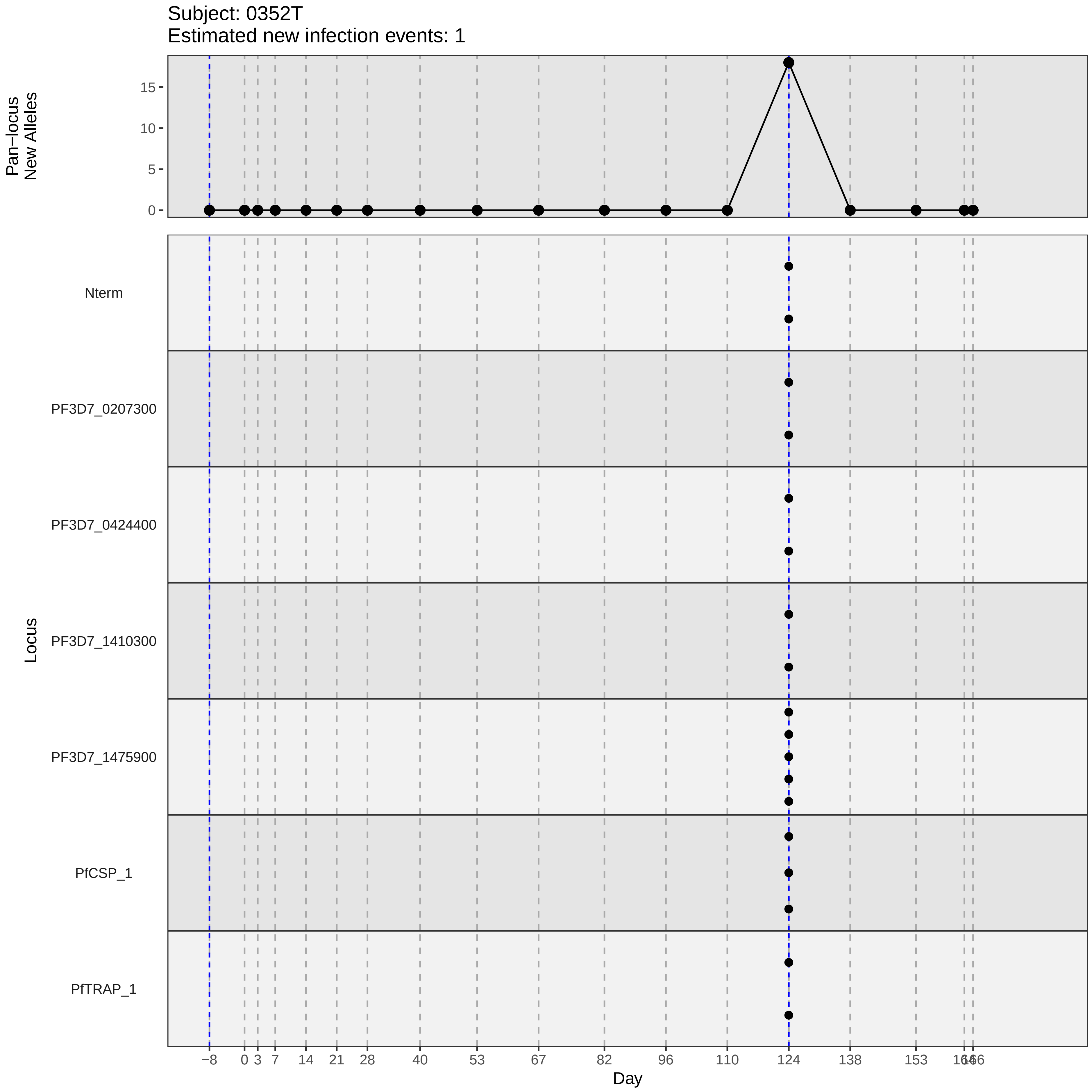

Subject: 0352T
Estimated new infection events: 1
Pan−locus New Alleles
0
5
10
15
Locus
Nterm
PF3D7_0207300
PF3D7_0424400
PF3D7_1410300
PF3D7_1475900
PfCSP_1
PfTRAP_1
−8
0
3
7
14
21
28
40
53
67
82
96
110
124
138
153
Day
