## Supplementary material for "Genotypic infection endpoint analysis to understand efficacy and escape potential of the malaria monoclonal antibody CIS43LS": Data File S1: participant_0358A_with_imputation.pdf

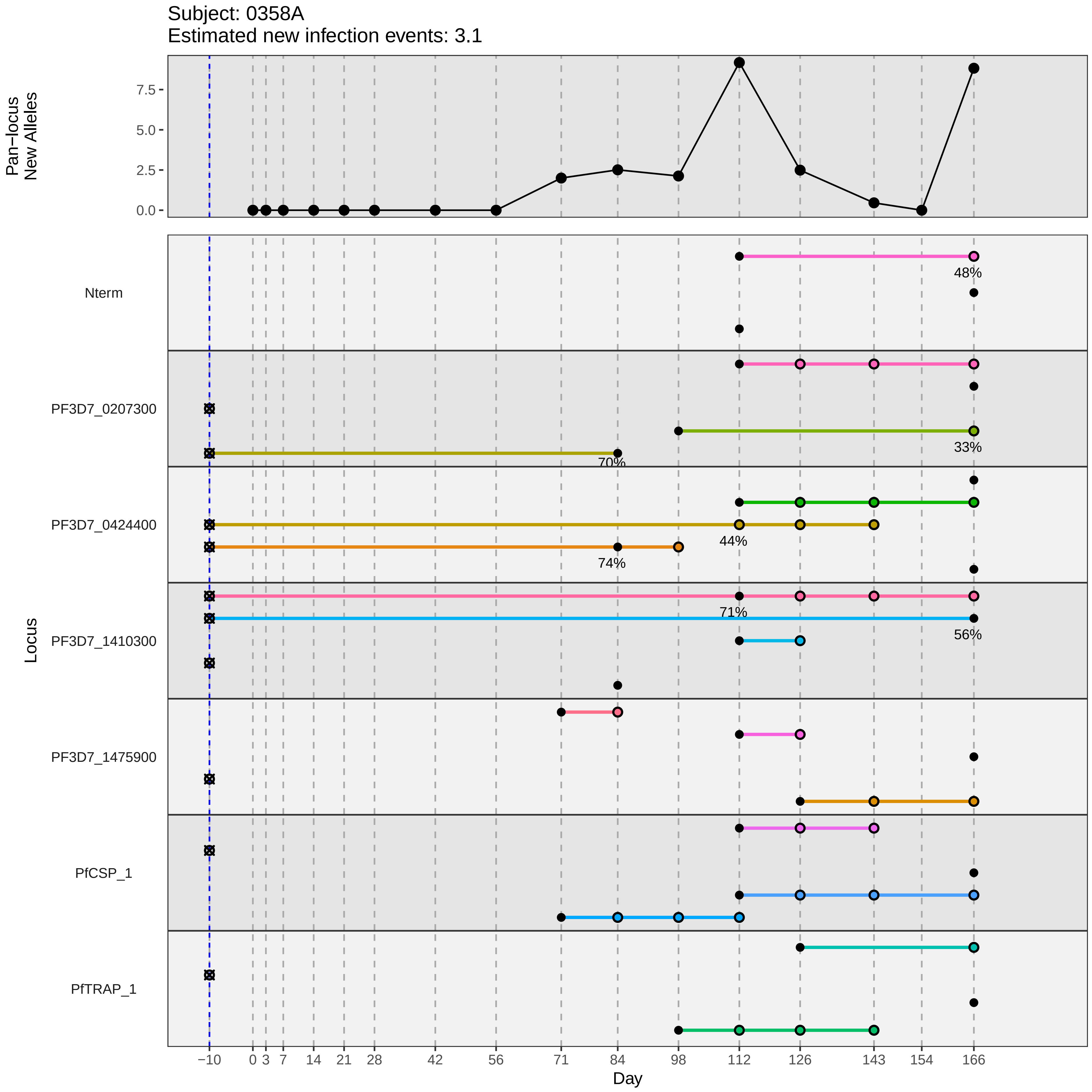

Subject: 0358A
Estimated new infection events: 3.1
Pan−locus
New Alleles
7.5
5.0
2.5
0.0
Locus
Nterm
PF3D7_0207300
PF3D7_0424400
PF3D7_1410300
PF3D7_1475900
PfCSP_1
PfTRAP_1
48%
33%
70%
44%
74%
71%
56%
−10
0
3
7
14
21
28
42
56
71
84
98
112
126
143
154
166
Day
