## Supplementary material for "Genotypic infection endpoint analysis to understand efficacy and escape potential of the malaria monoclonal antibody CIS43LS": Data File S1: participant_0361D_with_imputation.pdf

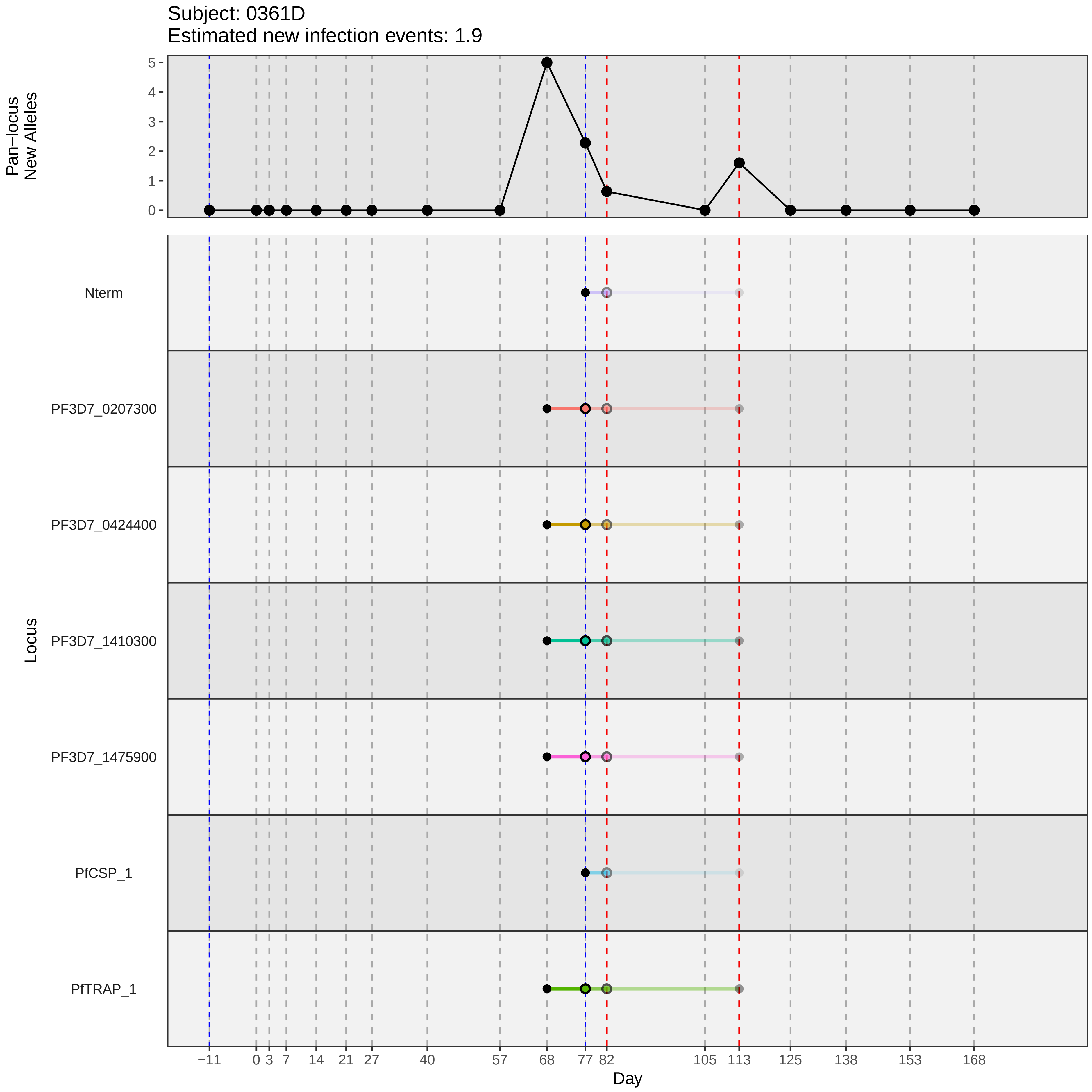
Subject: 0361D
Estimated new infection events: 1.9
Pan−locus
New Alleles
5
4
3
2
1
0
Locus
Nterm
PF3D7_0207300
PF3D7_0424400
PF3D7_1410300
PF3D7_1475900
PfCSP_1
PfTRAP_1
−11
0
3
7
14
21
27
40
57
68
77
82
105
113
125
138
153
168
Day
