## Supplementary material for "Genotypic infection endpoint analysis to understand efficacy and escape potential of the malaria monoclonal antibody CIS43LS": Data File S1: participant_0366K_with_imputation.pdf

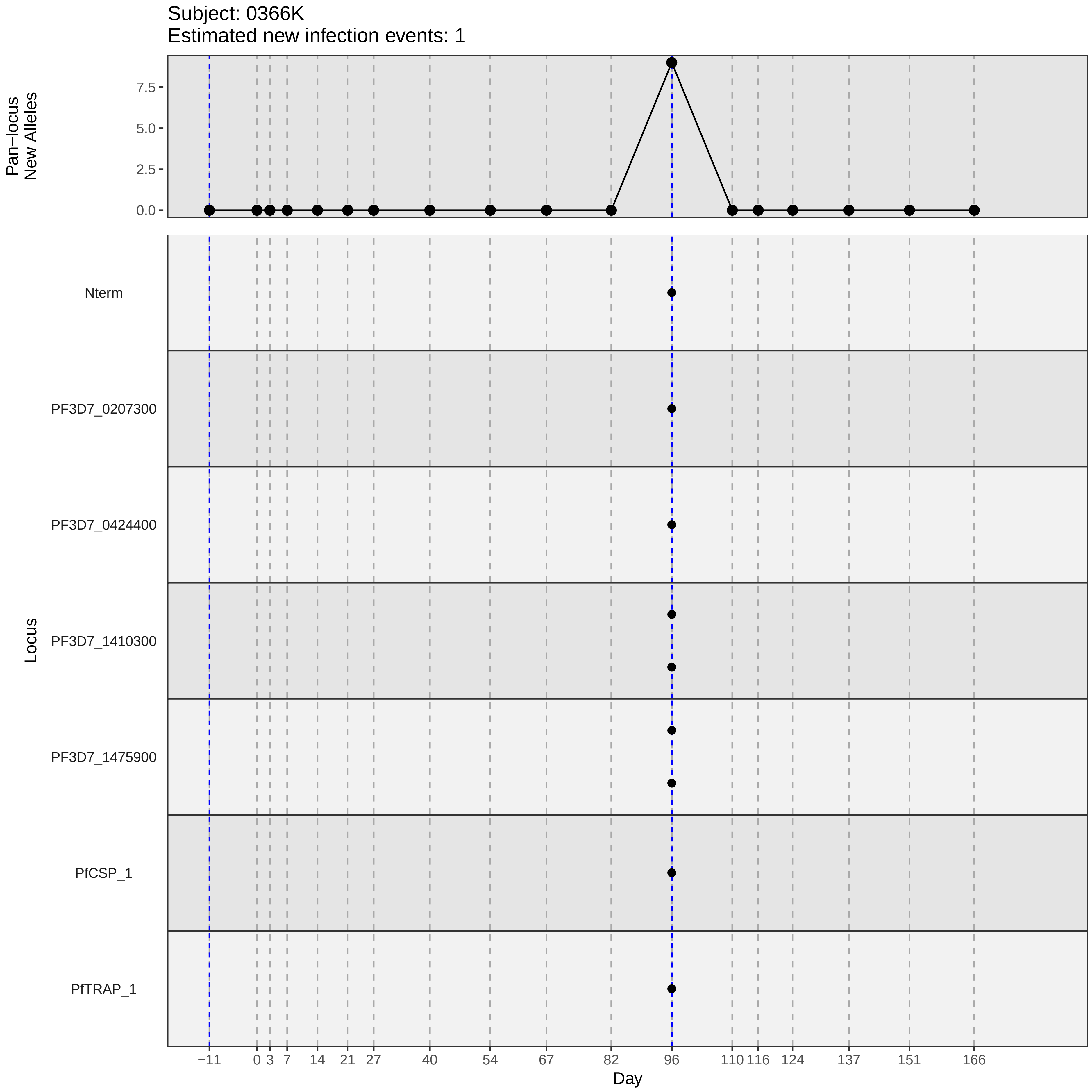
Subject: 0366K
Estimated new infection events: 1
Pan−locus
New Alleles
7.5
5.0
2.5
0.0
Nterm
PF3D7_0207300
PF3D7_0424400
PF3D7_1410300
PF3D7_1475900
PfCSP_1
PfTRAP_1
Locus
−11
0
3
7
14
21
27
40
54
67
82
96
110
116
124
137
151
166
Day
