## Supplementary material for "Genotypic infection endpoint analysis to understand efficacy and escape potential of the malaria monoclonal antibody CIS43LS": Data File S1: participant_0367M_with_imputation.pdf

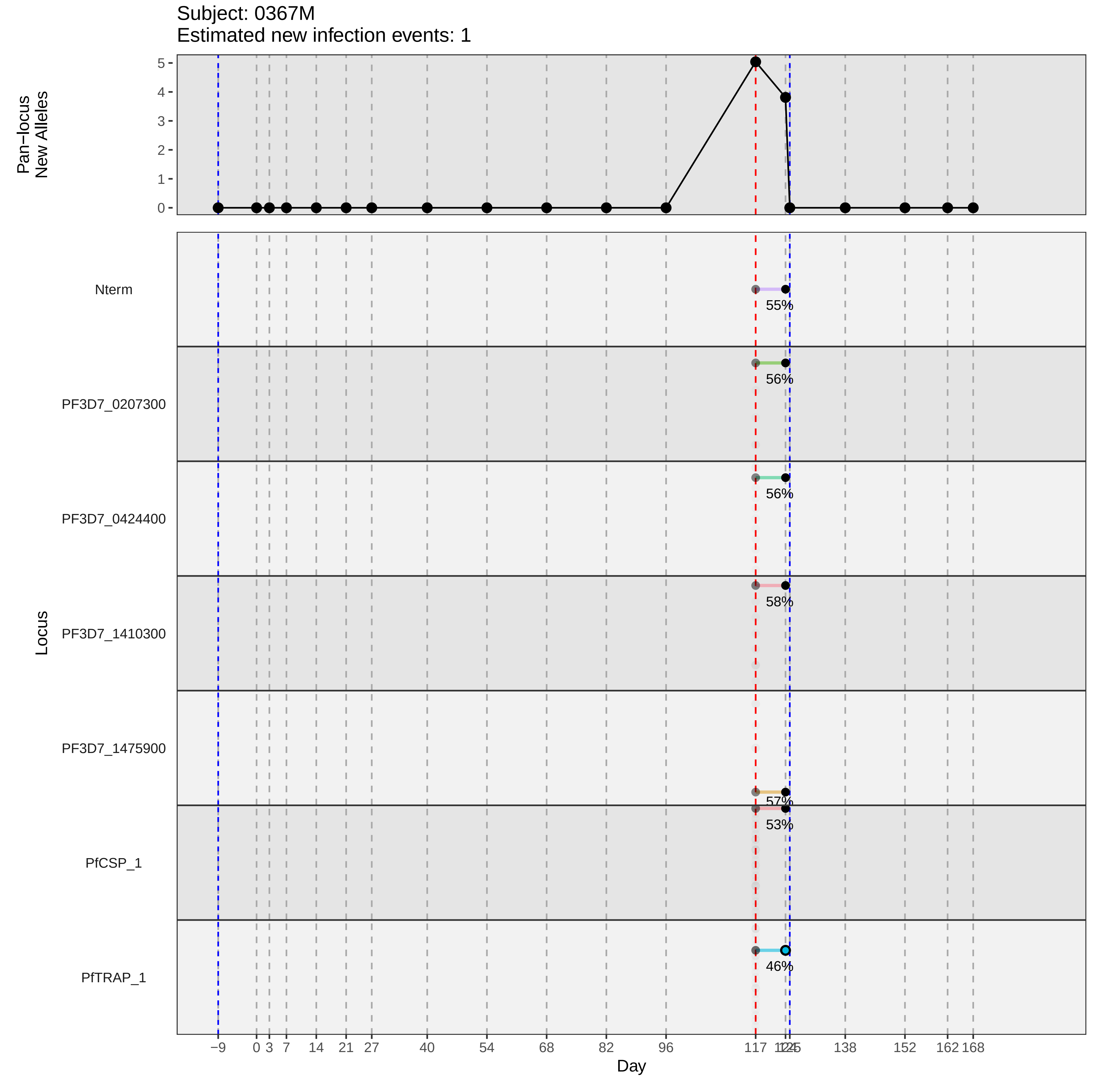
Subject: 0367M
Estimated new infection events: 1
Pan−locus
New Alleles
5
4
3
2
1
0
Locus
Nterm
PF3D7_0207300
PF3D7_0424400
PF3D7_1410300
PF3D7_1475900
PfCSP_1
PfTRAP_1
55%
56%
56%
58%
57%
53%
46%
−9
0
3
7
14
21
27
40
54
68
82
96
117
138
152
162
168
Day
