## Supplementary material for "Genotypic infection endpoint analysis to understand efficacy and escape potential of the malaria monoclonal antibody CIS43LS": Data File S1: participant_0384F_with_imputation.pdf

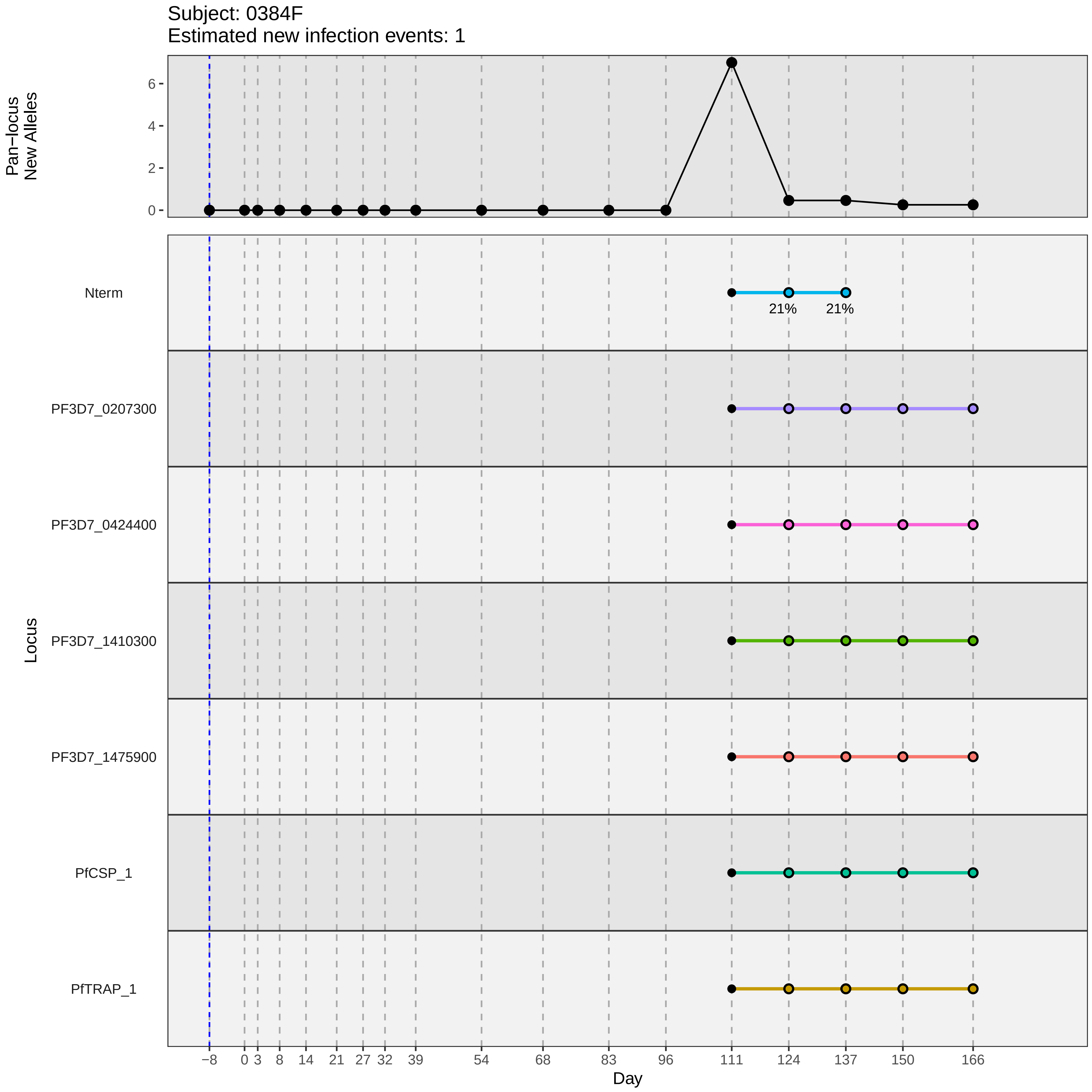
Subject: 0384F
Estimated new infection events: 1
Pan−locus New Alleles
0
2
4
6
Nterm
21%
21%
PF3D7_0207300
PF3D7_0424400
PF3D7_1410300
PF3D7_1475900
PfCSP_1
PfTRAP_1
Locus
−8
0
3
8
14
21
27
32
39
54
68
83
96
111
124
137
150
166
Day
