## Supplementary material for "Genotypic infection endpoint analysis to understand efficacy and escape potential of the malaria monoclonal antibody CIS43LS": Data File S1: participant_0386H_with_imputation.pdf

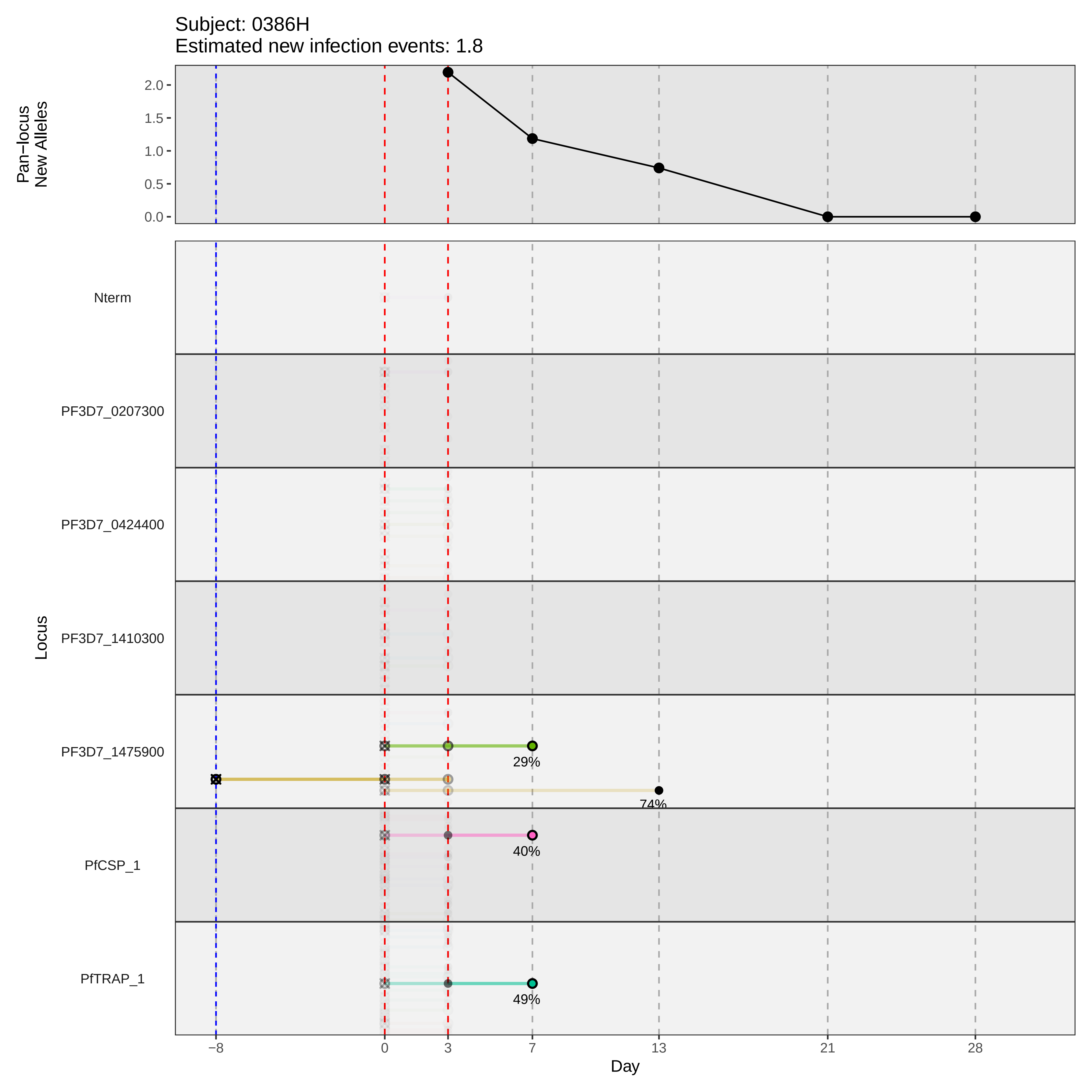

Subject: 0386H
Estimated new infection events: 1.8
Pan−locus New Alleles
2.0
1.5
1.0
0.5
0.0
Locus
Nterm
PF3D7_0207300
PF3D7_0424400
PF3D7_1410300
PF3D7_1475900
PfCSP_1
PfTRAP_1
29%
74%
40%
49%
−8
0
3
7
13
21
28
Day
