## Supplementary material for "Genotypic infection endpoint analysis to understand efficacy and escape potential of the malaria monoclonal antibody CIS43LS": Data File S1: participant_0387K_with_imputation.pdf

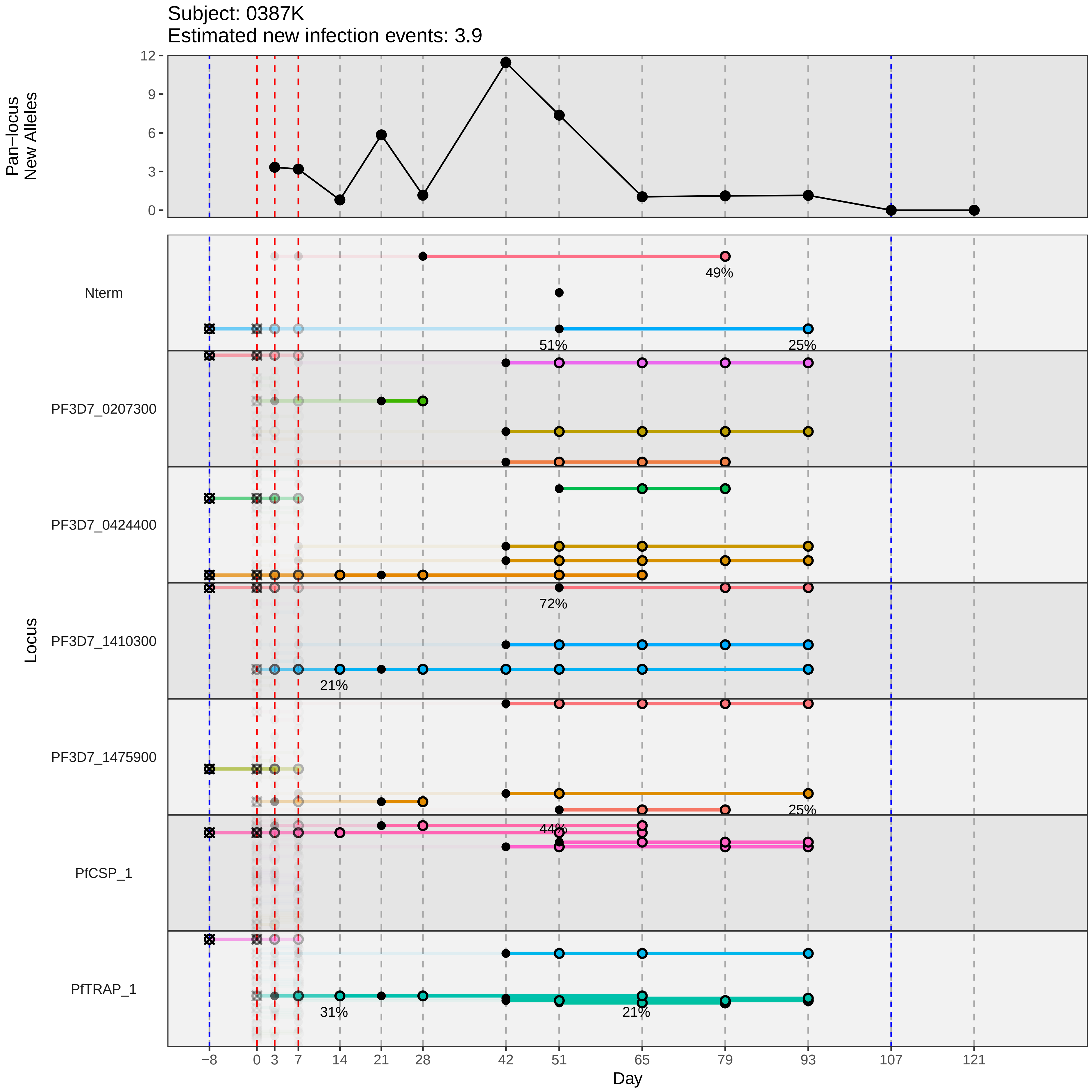

Subject: 0387K
Estimated new infection events: 3.9
Pan−locus New Alleles
12
9
6
3
0
Locus
Nterm
PF3D7_0207300
PF3D7_0424400
PF3D7_1410300
PF3D7_1475900
PfCSP_1
PfTRAP_1
49%
51%
25%
72%
21%
25%
44%
31%
21%
−8
0
3
7
14
21
28
42
51
65
79
93
107
121
Day
