## Supplementary material for "Genotypic infection endpoint analysis to understand efficacy and escape potential of the malaria monoclonal antibody CIS43LS": Data File S1: participant_0390P_with_imputation.pdf

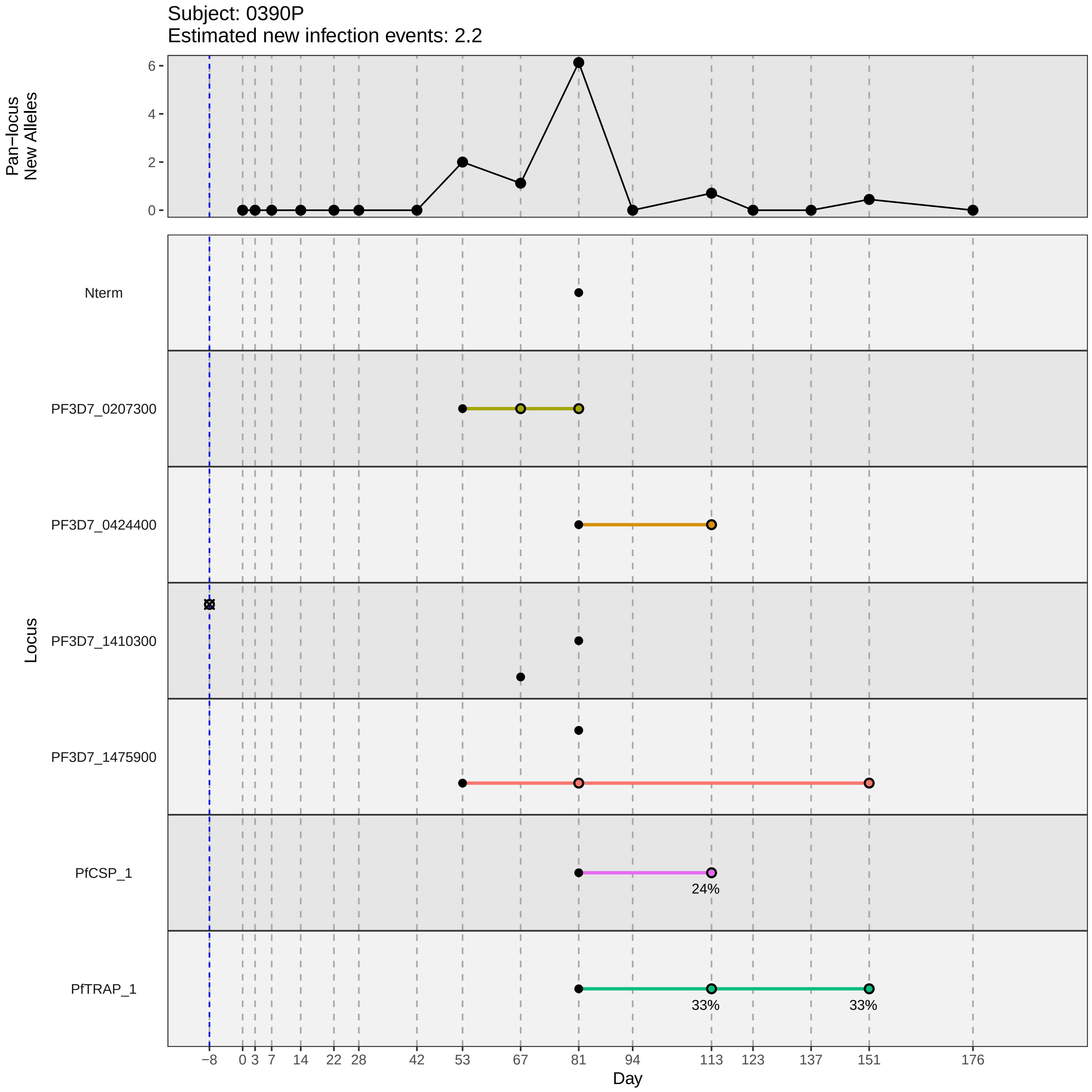
Subject: 0390P
Estimated new infection events: 2.2
Pan−locus
New Alleles
6
4
2
0
Locus
Nterm
PF3D7_0207300
PF3D7_0424400
PF3D7_1410300
PF3D7_1475900
PfCSP_1
PfTRAP_1
24%
33%
33%
−8
0
3
7
14
22
28
42
53
67
81
94
113
123
137
151
176
Day
