## Supplementary material for "Genotypic infection endpoint analysis to understand efficacy and escape potential of the malaria monoclonal antibody CIS43LS": Data File S1: participant_0397W_with_imputation.pdf

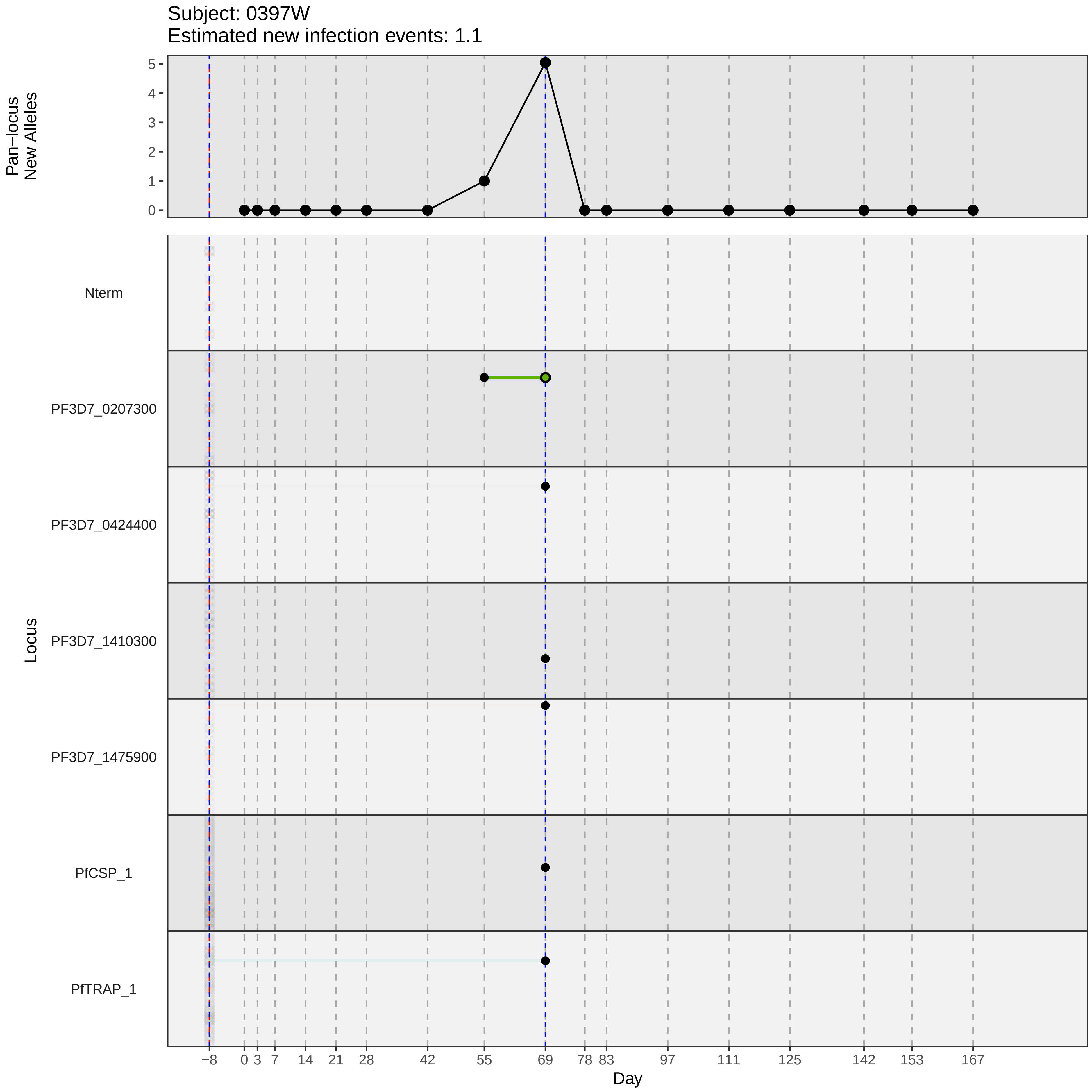

Subject: 0397W
Estimated new infection events: 1.1
Pan−locus New Alleles
5
4
3
2
1
0
Locus
Nterm
PF3D7_0207300
PF3D7_0424400
PF3D7_1410300
PF3D7_1475900
PfCSP_1
PfTRAP_1
−8
0
3
7
14
21
28
42
55
69
78
83
97
111
125
142
153
167
Day
