## Supplementary material for "Genotypic infection endpoint analysis to understand efficacy and escape potential of the malaria monoclonal antibody CIS43LS": Data File S1: participant_0398X_with_imputation.pdf

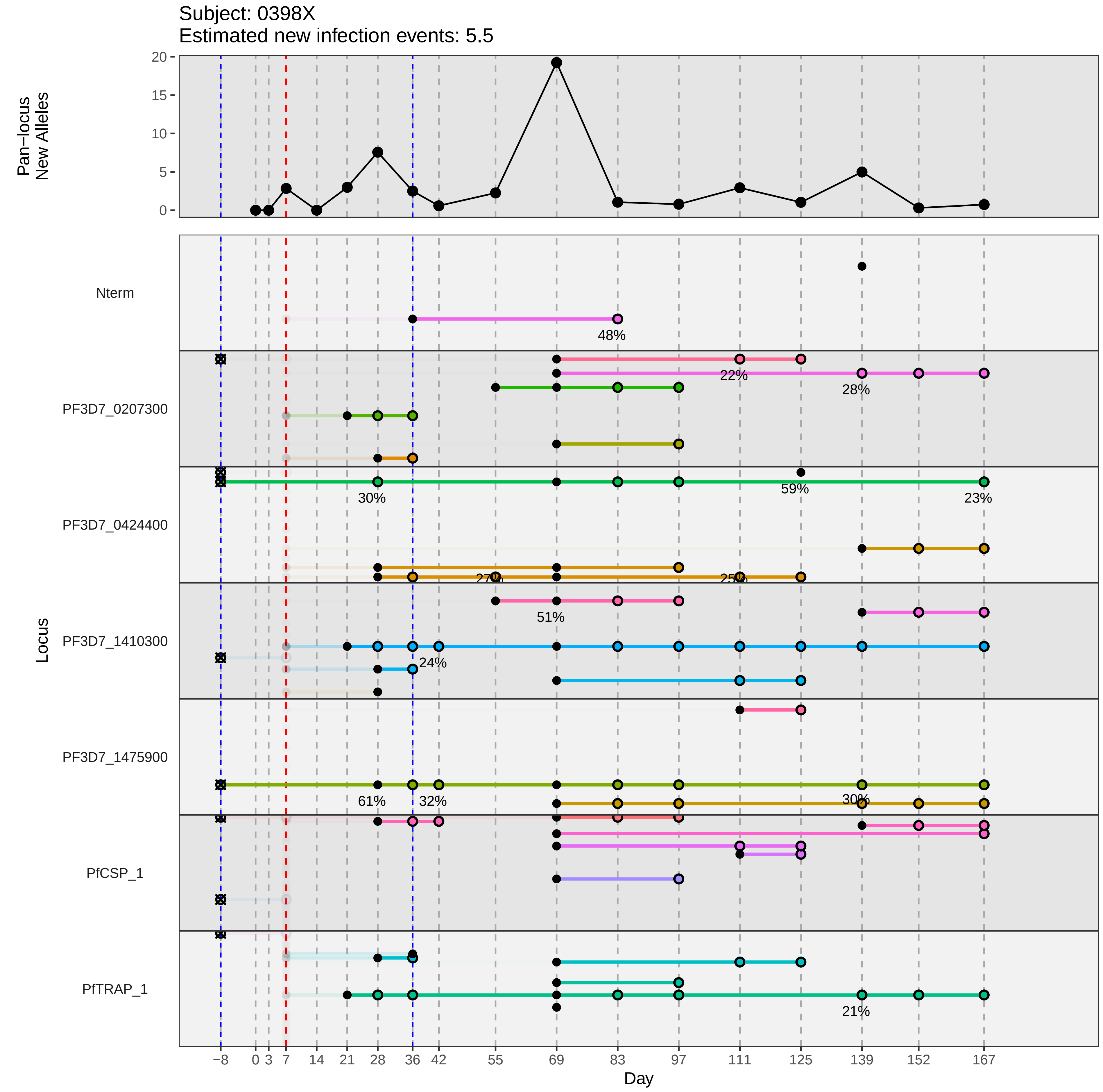

Subject: 0398X
Estimated new infection events: 5.5
Pan−locus New Alleles
20
15
10
5
0
Nterm
48%
PF3D7_0207300
22%
28%
PF3D7_0424400
30%
59%
23%
27%
25%
PF3D7_1410300
51%
24%
PF3D7_1475900
61%
32%
30%
PfCSP_1
PfTRAP_1
21%
Locus
−8
0
3
7
14
21
28
36
42
55
69
83
97
111
125
139
152
167
Day
