## Supplementary material for "Genotypic infection endpoint analysis to understand efficacy and escape potential of the malaria monoclonal antibody CIS43LS": Data File S1: participant_0406G_with_imputation.pdf

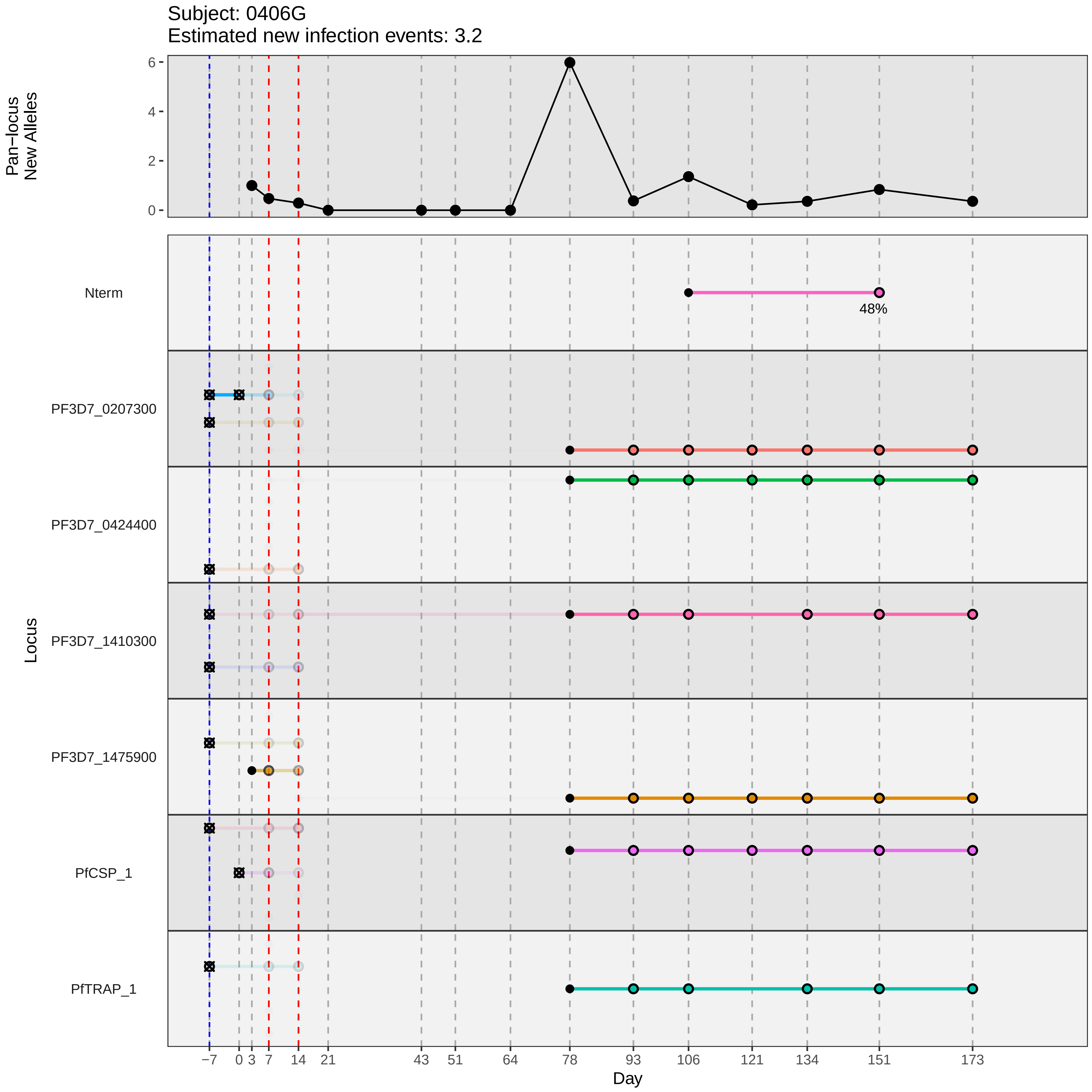

Subject: 0406G
Estimated new infection events: 3.2
Pan−locus New Alleles
0
2
4
6
Locus
Nterm
48%
PF3D7_0207300
PF3D7_0424400
PF3D7_1410300
PF3D7_1475900
PfCSP_1
PfTRAP_1
−7
0
3
7
14
21
43
51
64
78
93
106
121
134
151
173
Day
