## Supplementary material for "Genotypic infection endpoint analysis to understand efficacy and escape potential of the malaria monoclonal antibody CIS43LS": Data File S1: participant_0409M_with_imputation.pdf

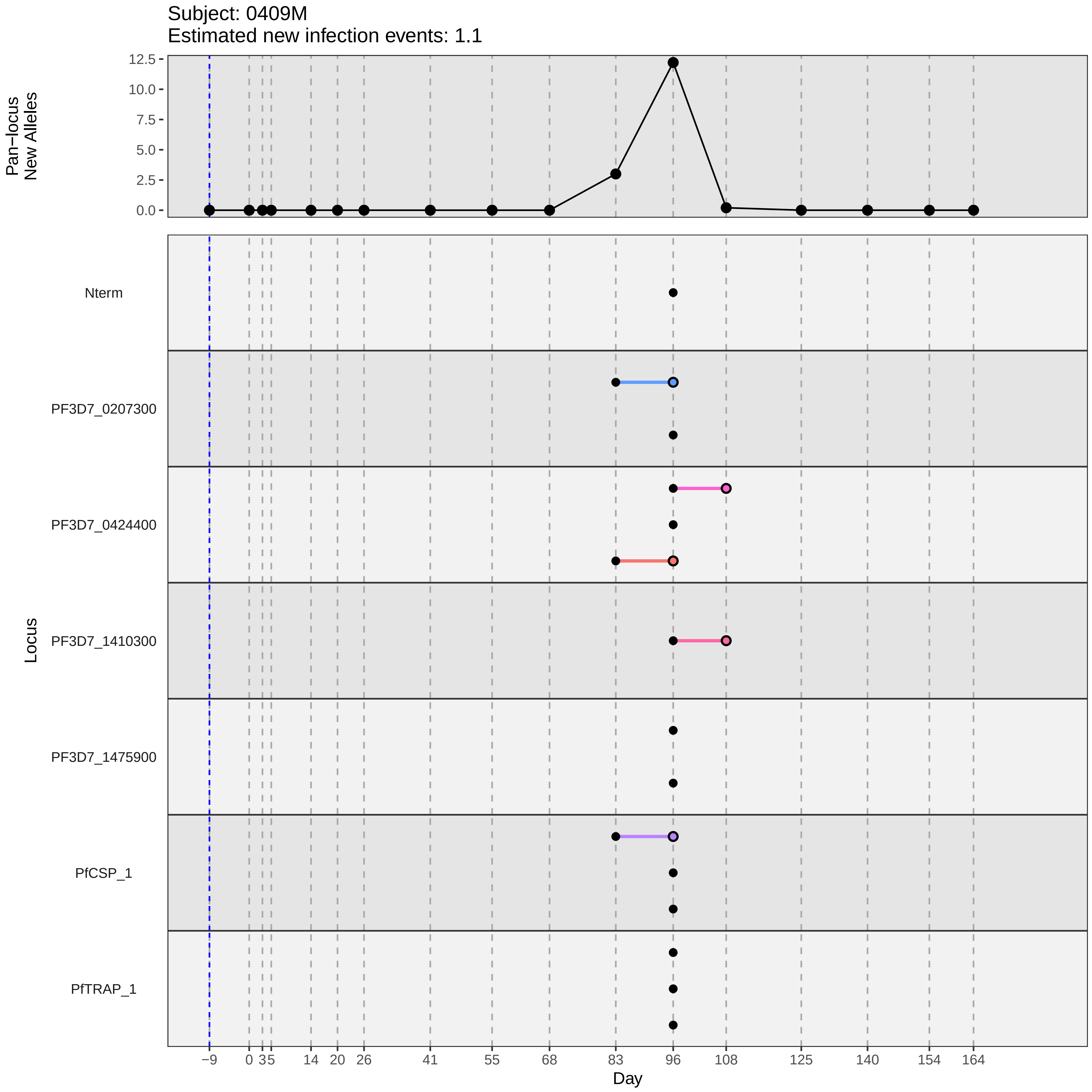
Subject: 0409M
Estimated new infection events: 1.1
Pan−locus
New Alleles
12.5
10.0
7.5
5.0
2.5
0.0
Locus
Nterm
PF3D7_0207300
PF3D7_0424400
PF3D7_1410300
PF3D7_1475900
PfCSP_1
PfTRAP_1
−9
0
3
5
14
20
26
41
55
68
83
96
108
125
140
154
164
Day
