## Supplementary material for "Genotypic infection endpoint analysis to understand efficacy and escape potential of the malaria monoclonal antibody CIS43LS": Data File S1: participant_0412Q_with_imputation.pdf

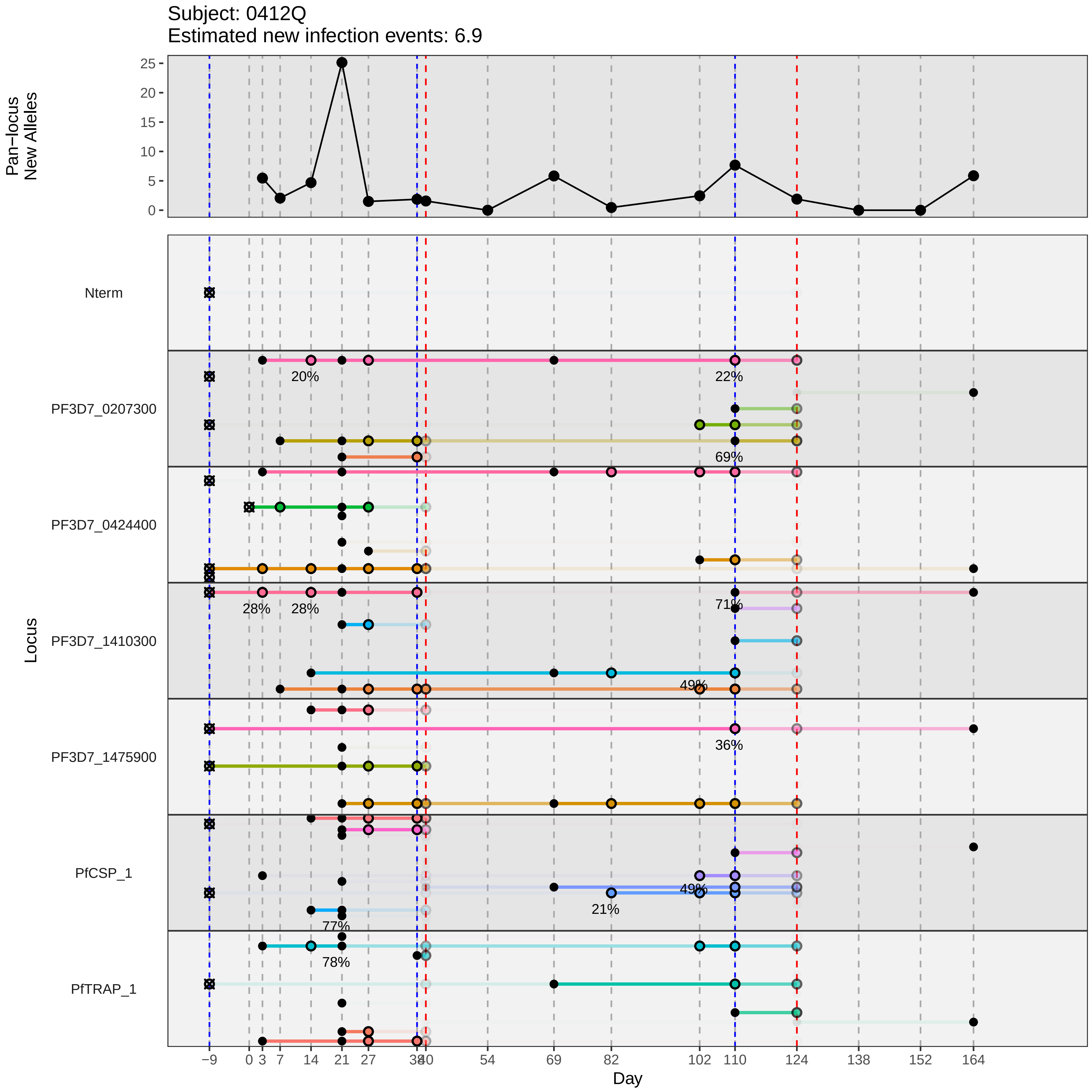

Subject: 0412Q
Estimated new infection events: 6.9
Pan−locus New Alleles
25
20
15
10
5
0
Locus
Nterm
PF3D7_0207300
PF3D7_0424400
PF3D7_1410300
PF3D7_1475900
PfCSP_1
PfTRAP_1
20%
22%
69%
28%
28%
71%
49%
36%
49%
21%
77%
78%
−9
0
3
7
14
21
27
38
40
54
69
82
102
110
124
138
152
164
Day
