## Supplementary material for "Genotypic infection endpoint analysis to understand efficacy and escape potential of the malaria monoclonal antibody CIS43LS": Data File S1: participant_0418W_with_imputation.pdf

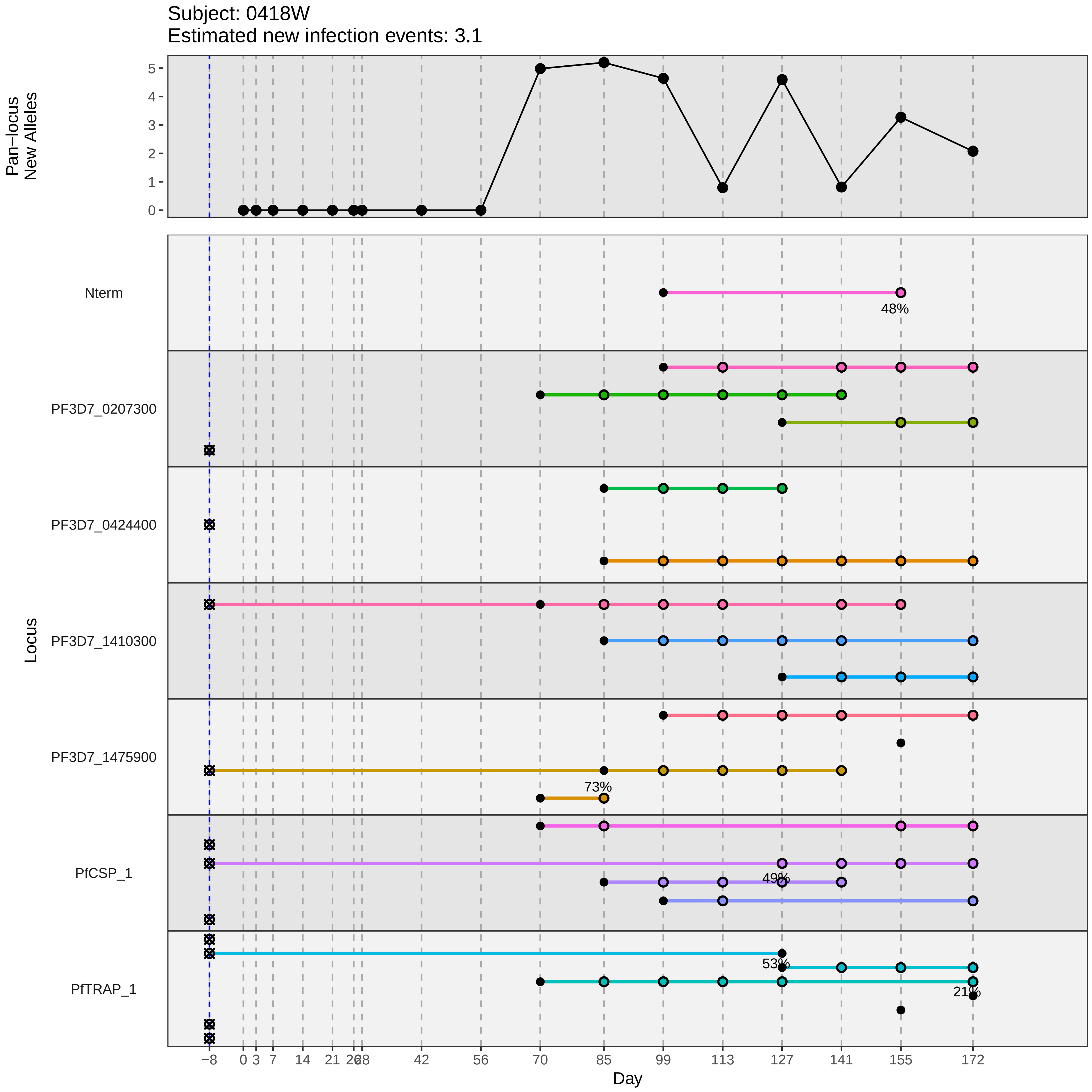

Subject: 0418W
Estimated new infection events: 3.1
Pan−locus New Alleles
Locus
Nterm
PF3D7_0207300
PF3D7_0424400
PF3D7_1410300
PF3D7_1475900
PfCSP_1
PfTRAP_1
48%
73%
49%
53%
21%
Day
−8
0
3
7
14
21
26
28
42
56
70
85
99
113
127
141
155
172
