## Supplementary material for "Genotypic infection endpoint analysis to understand efficacy and escape potential of the malaria monoclonal antibody CIS43LS": Data File S1: participant_0419X_with_imputation.pdf

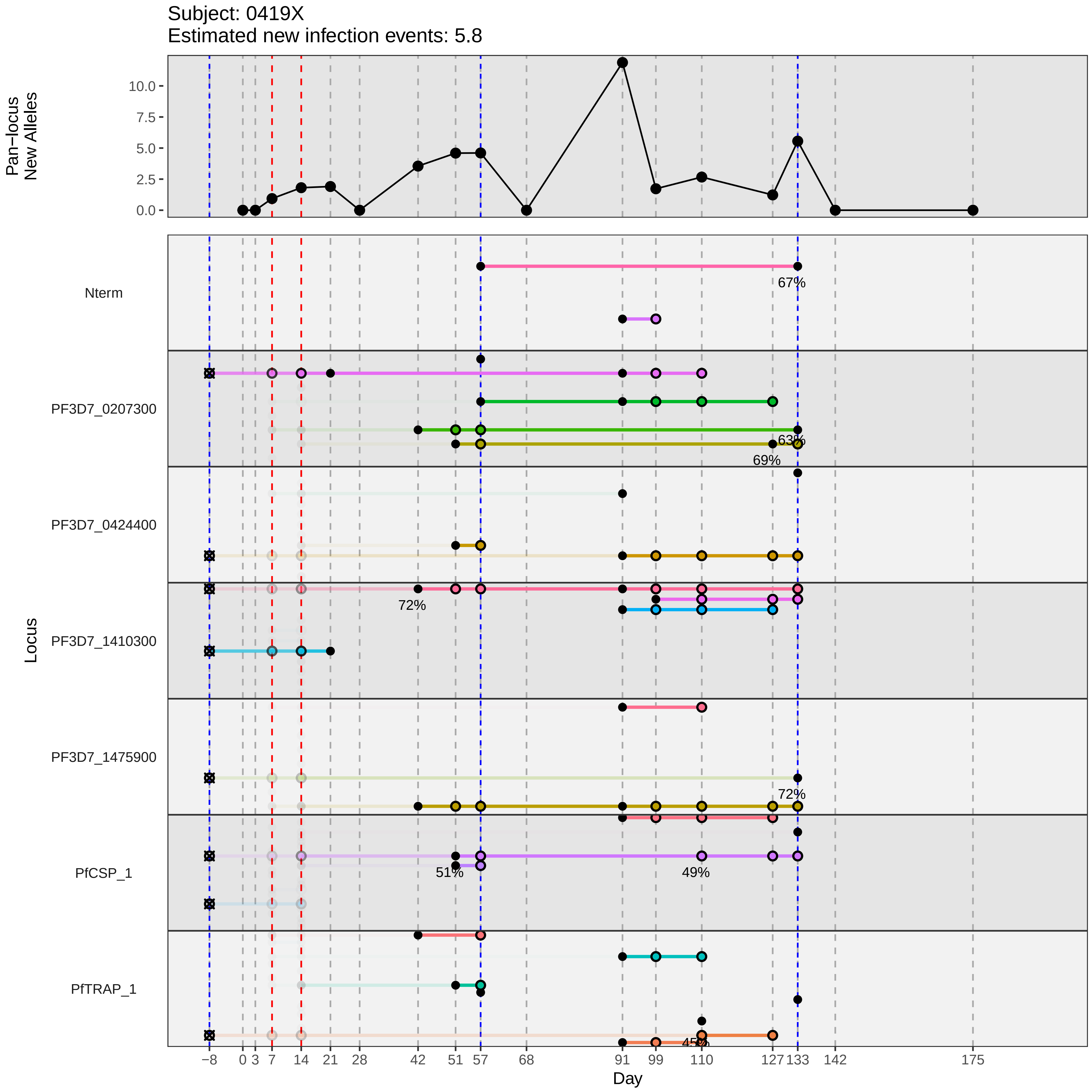
Subject: 0419X
Estimated new infection events: 5.8
Pan−locus
New Alleles
10.0
7.5
5.0
2.5
0.0
Locus
Nterm
PF3D7_0207300
PF3D7_0424400
PF3D7_1410300
PF3D7_1475900
PfCSP_1
PfTRAP_1
67%
63%
69%
72%
72%
51%
49%
45%
−8
0
3
7
14
21
28
42
51
57
68
91
99
110
127
133
142
175
Day
