## Supplementary material for "Genotypic infection endpoint analysis to understand efficacy and escape potential of the malaria monoclonal antibody CIS43LS": Data File S1: participant_0420Y_with_imputation.pdf

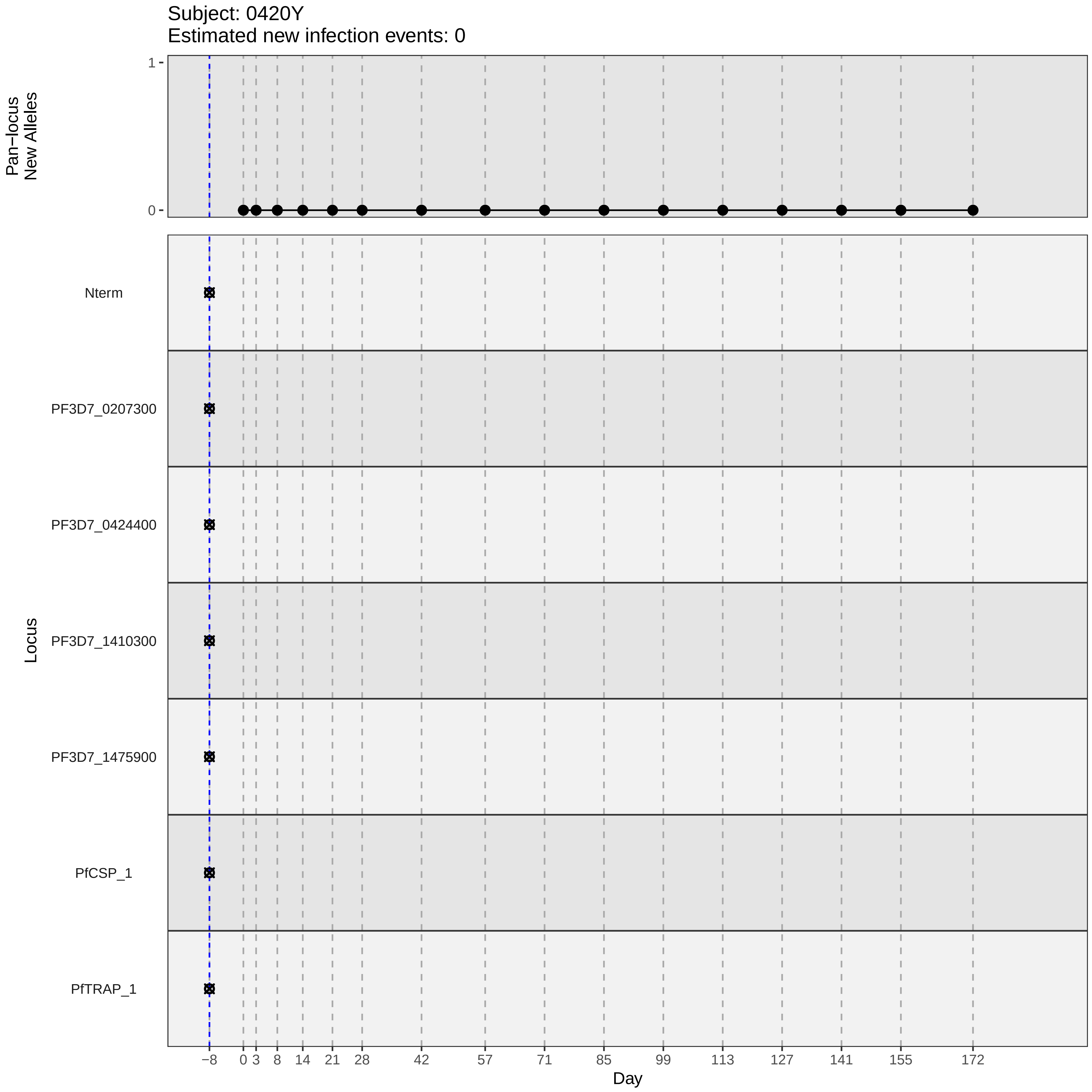

Subject: 0420Y
Estimated new infection events: 0
Pan−locus
New Alleles
1
0
Locus
Nterm
PF3D7_0207300
PF3D7_0424400
PF3D7_1410300
PF3D7_1475900
PfCSP_1
PfTRAP_1
−8
0
3
8
14
21
28
42
57
71
85
99
113
127
141
155
172
Day
