## Supplementary material for "Genotypic infection endpoint analysis to understand efficacy and escape potential of the malaria monoclonal antibody CIS43LS": Data File S1: participant_0422B_with_imputation.pdf

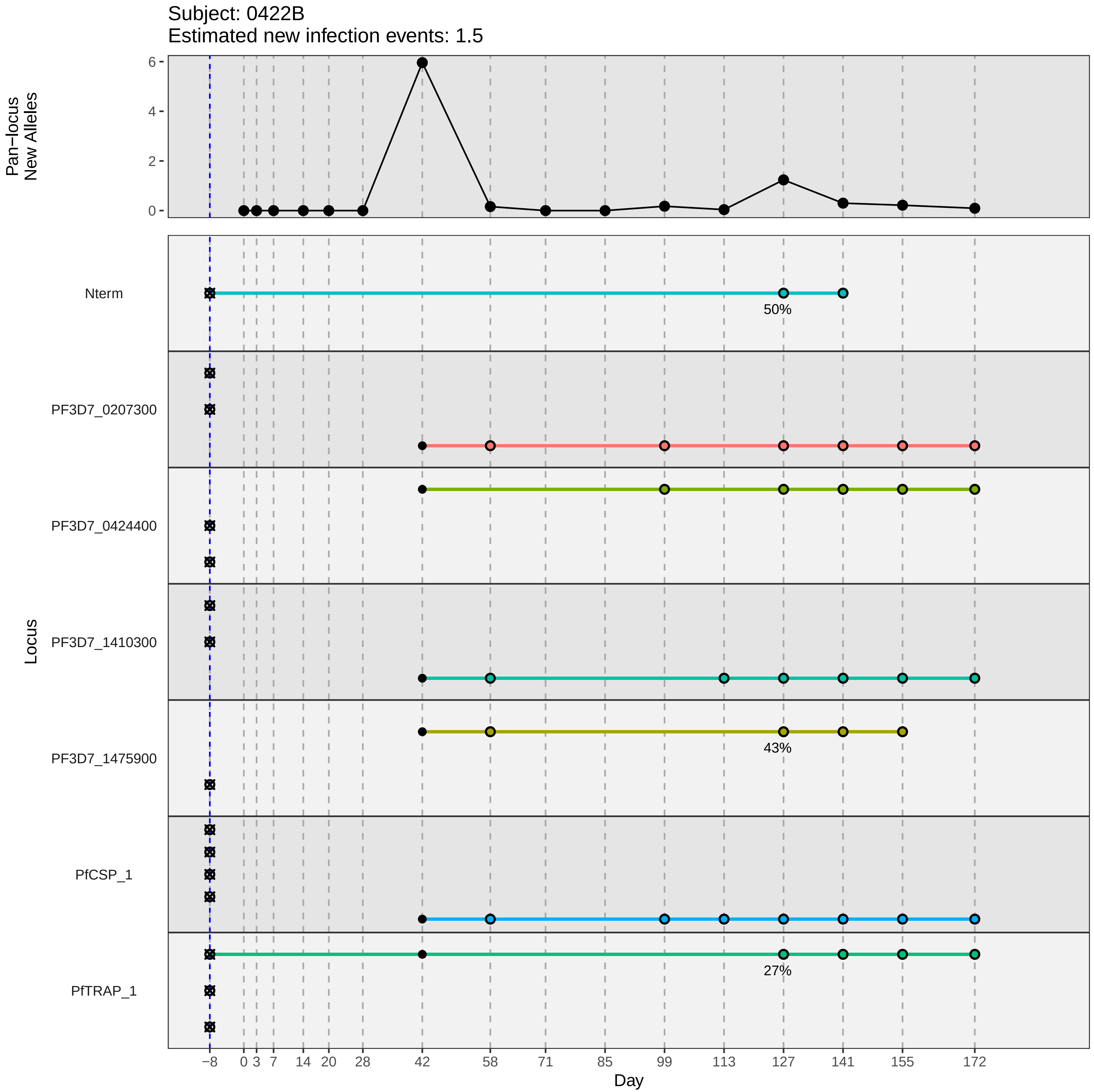

Subject: 0422B
Estimated new infection events: 1.5
Pan−locus
New Alleles
6
4
2
0
Locus
Nterm
PF3D7_0207300
PF3D7_0424400
PF3D7_1410300
PF3D7_1475900
PfCSP_1
PfTRAP_1
50%
43%
27%
−8
0
3
7
14
20
28
42
58
71
85
99
113
127
141
155
172
Day
