## Supplementary material for "Genotypic infection endpoint analysis to understand efficacy and escape potential of the malaria monoclonal antibody CIS43LS": Data File S1: participant_0429K_with_imputation.pdf

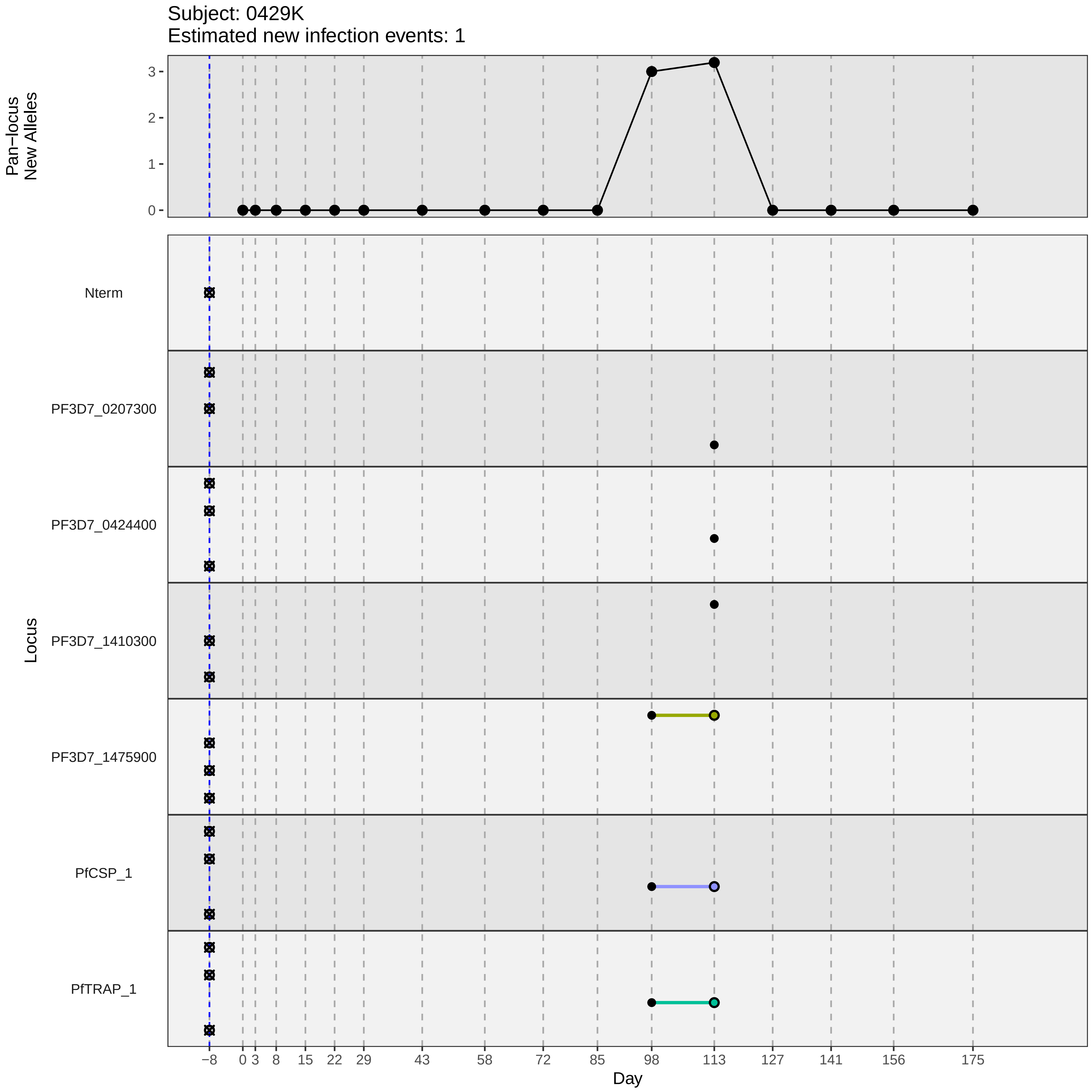

Subject: 0429K
Estimated new infection events: 1
Pan−locus
New Alleles
3
2
1
0
Locus
Nterm
PF3D7_0207300
PF3D7_0424400
PF3D7_1410300
PF3D7_1475900
PfCSP_1
PfTRAP_1
−8
0
3
8
15
22
29
43
58
72
85
98
113
127
141
156
175
Day
