## Supplementary figures and images for "Genotypic infection endpoint analysis to understand efficacy and escape potential of the malaria monoclonal antibody CIS43LS"

### participant_0111F_with_imputation.pdf

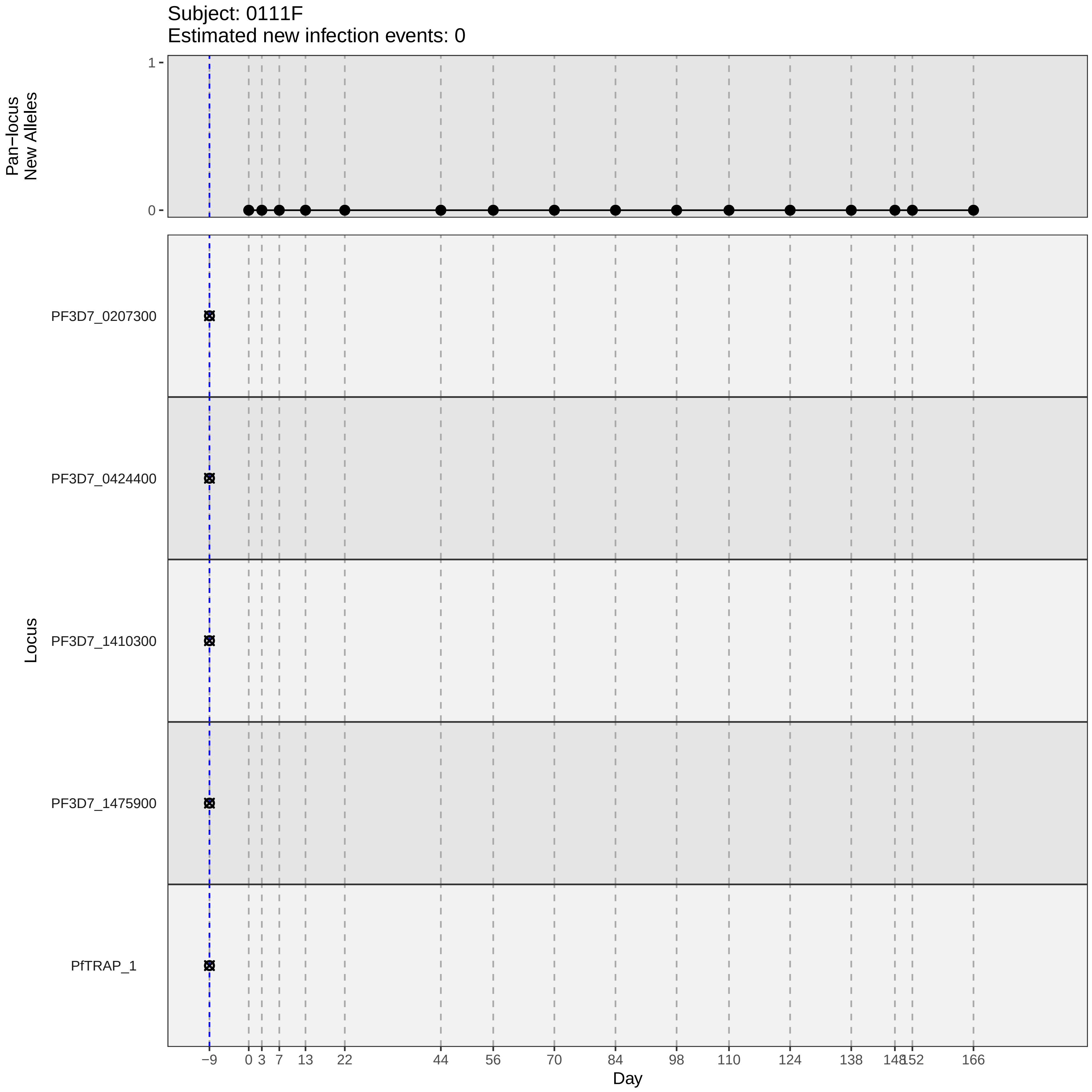
Subject: 0111F
Estimated new infection events: 0
Pan−locus New Alleles
1
0
Locus
PF3D7_0207300
PF3D7_0424400
PF3D7_1410300
PF3D7_1475900
PfTRAP_1
−9
0
3
7
13
22
44
56
70
84
98
110
124
138
148
152
166
Day

### participant_0111F_without_imputation.pdf

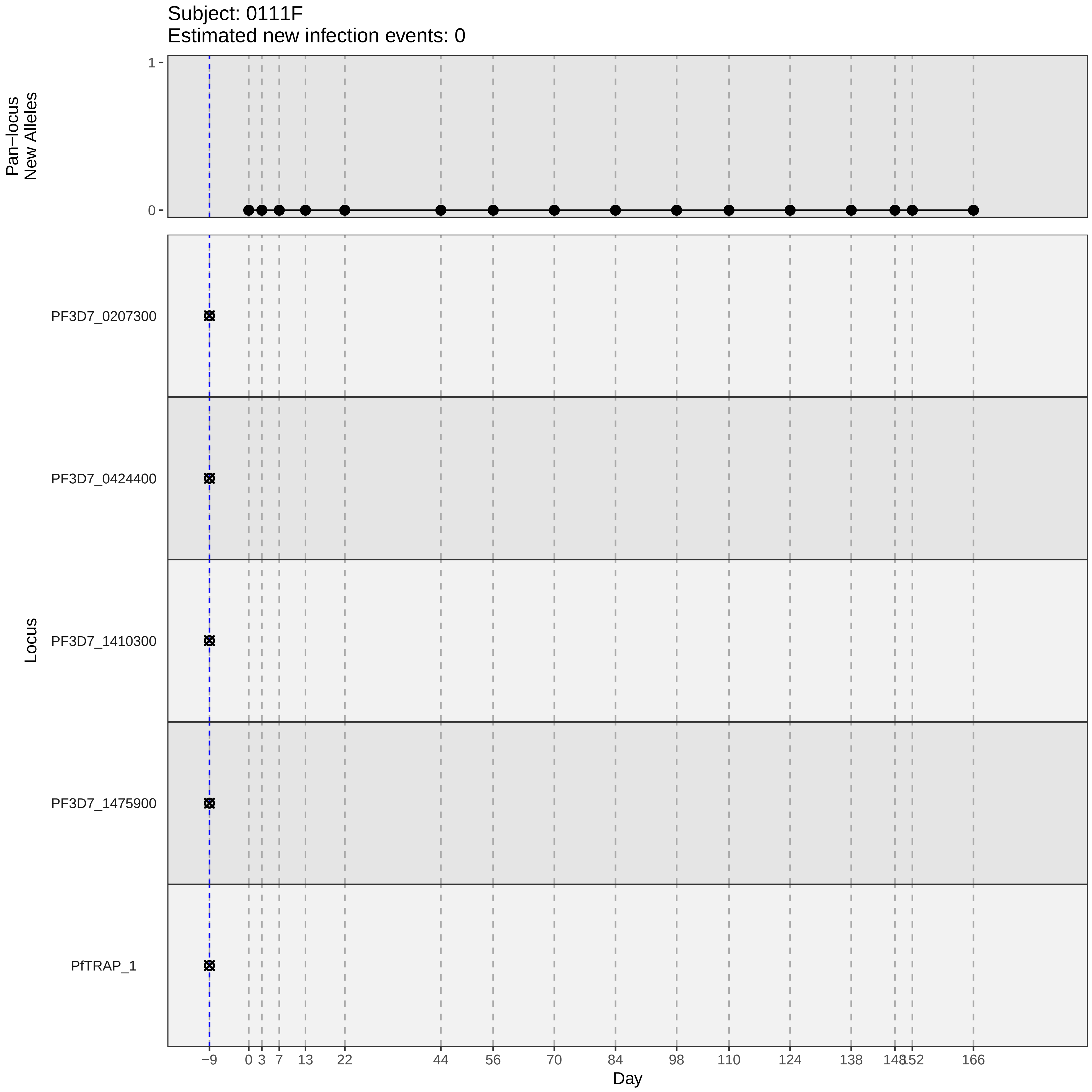

Subject: 0111F
Estimated new infection events: 0
Pan−locus
New Alleles
1
0
Locus
PF3D7_0207300
PF3D7_0424400
PF3D7_1410300
PF3D7_1475900
PfTRAP_1
−9
0
3
7
13
22
44
56
70
84
98
110
124
138
148
152
166
Day

### participant_0133G_without_imputation.pdf

Subject: 0133G
Estimated new infection events: 0

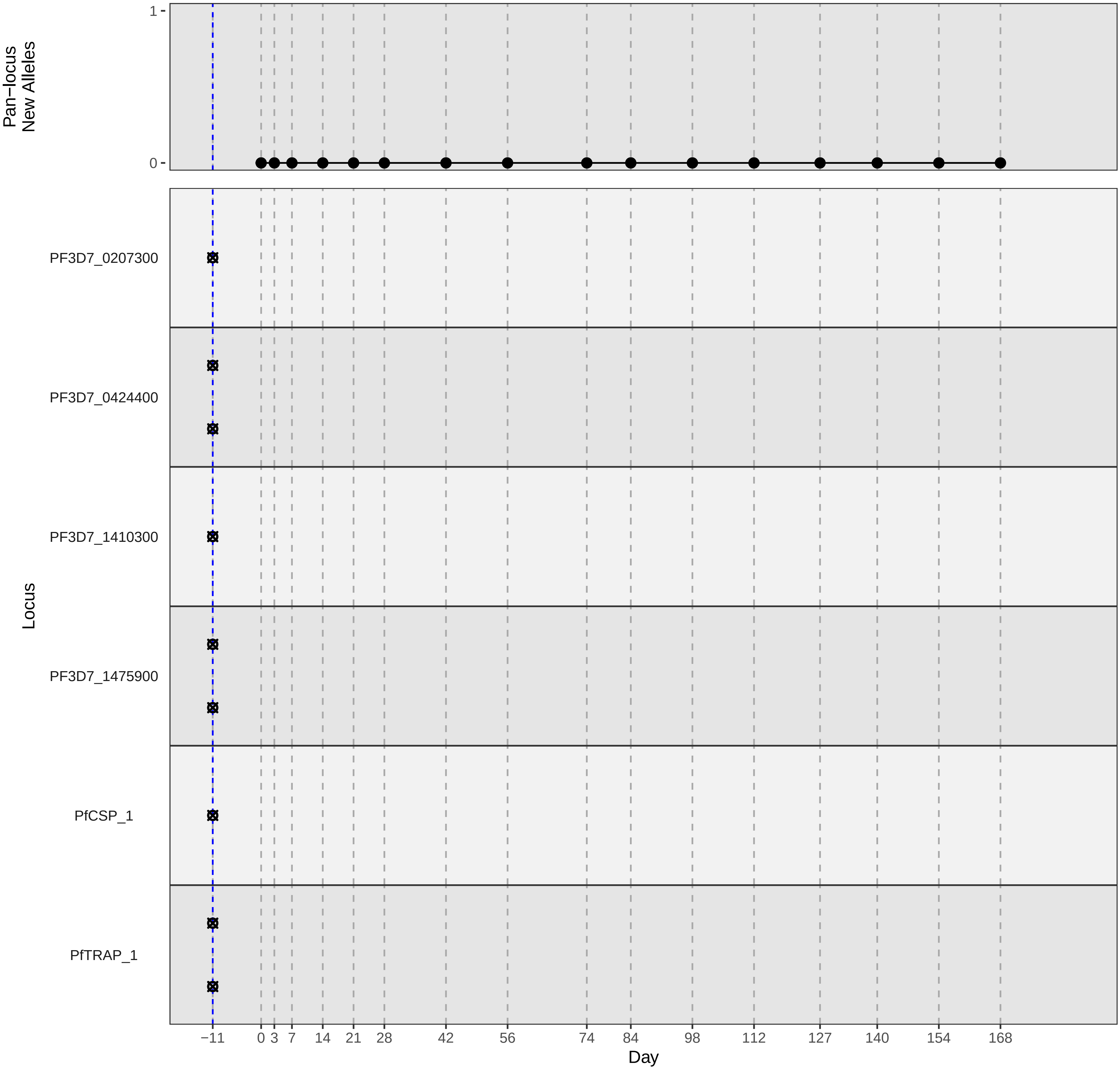

### participant_0164U_with_imputation.pdf

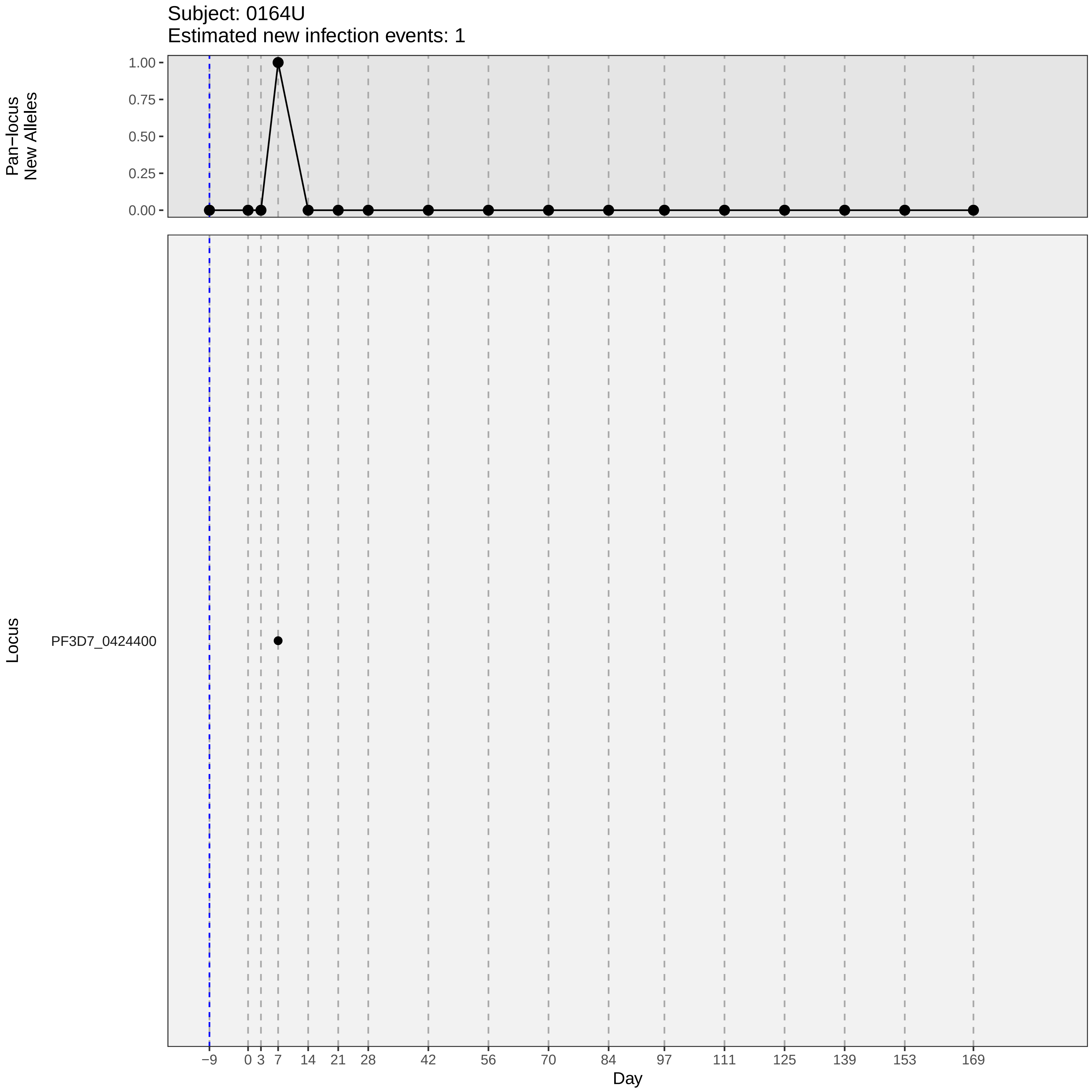
Subject: 0164U
Estimated new infection events: 1
Pan−locus New Alleles
1.00
0.75
0.50
0.25
0.00
Locus
PF3D7_0424400
−9
0
3
7
14
21
28
42
56
70
84
97
111
125
139
153
169
Day

### participant_0164U_without_imputation.pdf

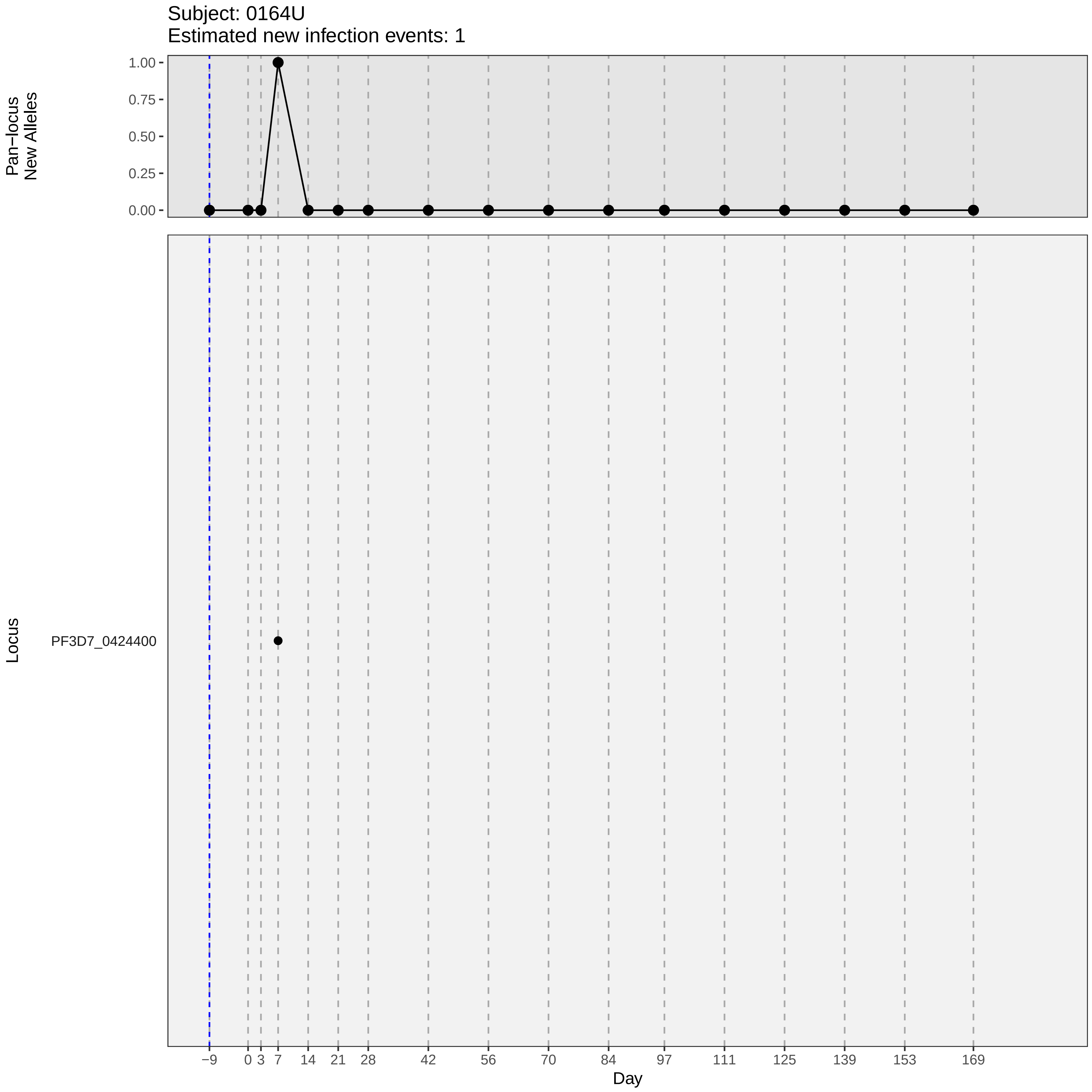

Subject: 0164U
Estimated new infection events: 1
Pan−locus New Alleles
1.00
0.75
0.50
0.25
0.00
Locus
PF3D7_0424400
−9
0
3
7
14
21
28
42
56
70
84
97
111
125
139
153
169
Day

### participant_0168Y_without_imputation.pdf

Subject: 0168Y
Estimated new infection events: 1

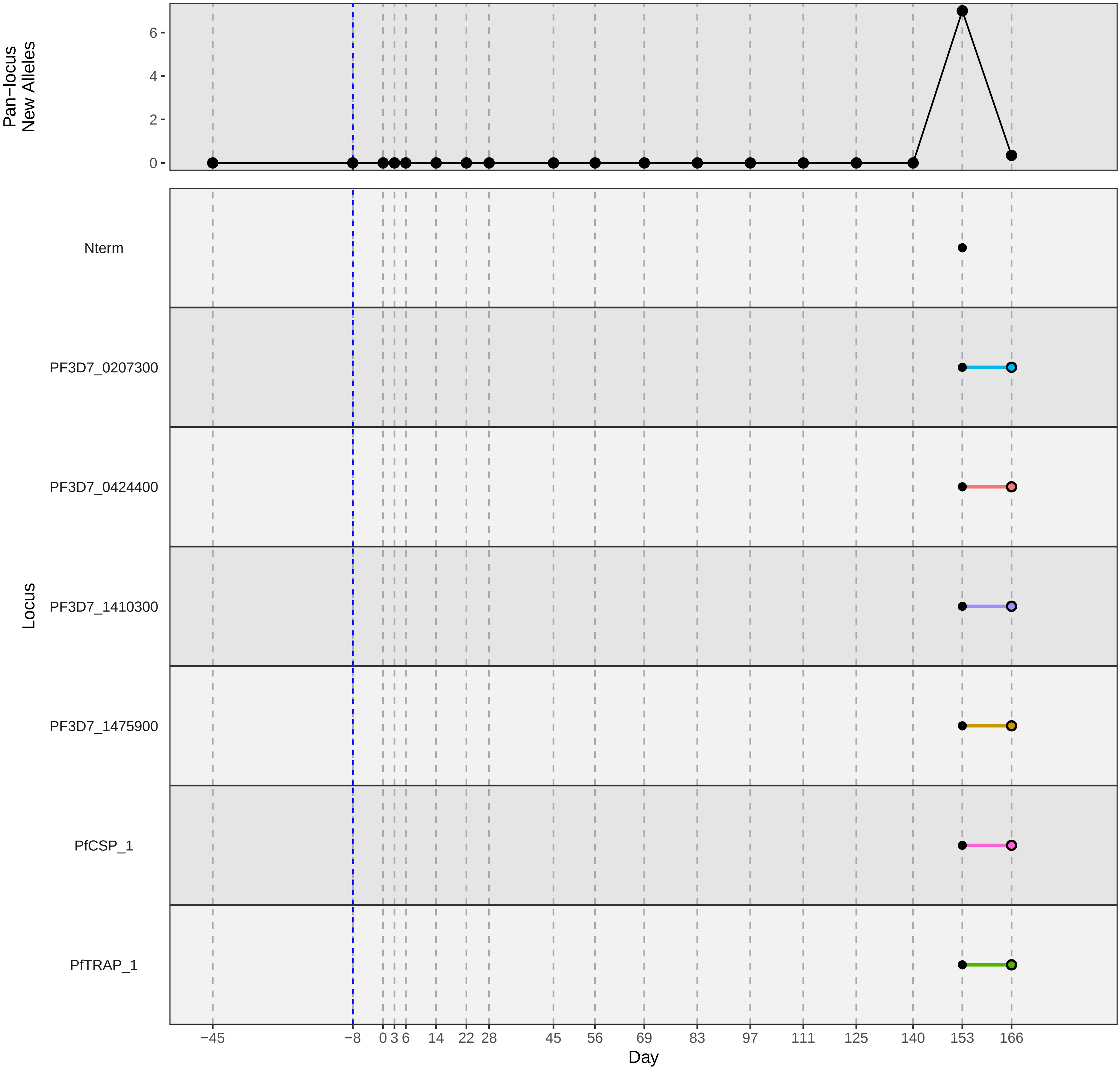

### participant_0181Q_with_imputation.pdf

Subject: 0181Q
Estimated new infection events: 1

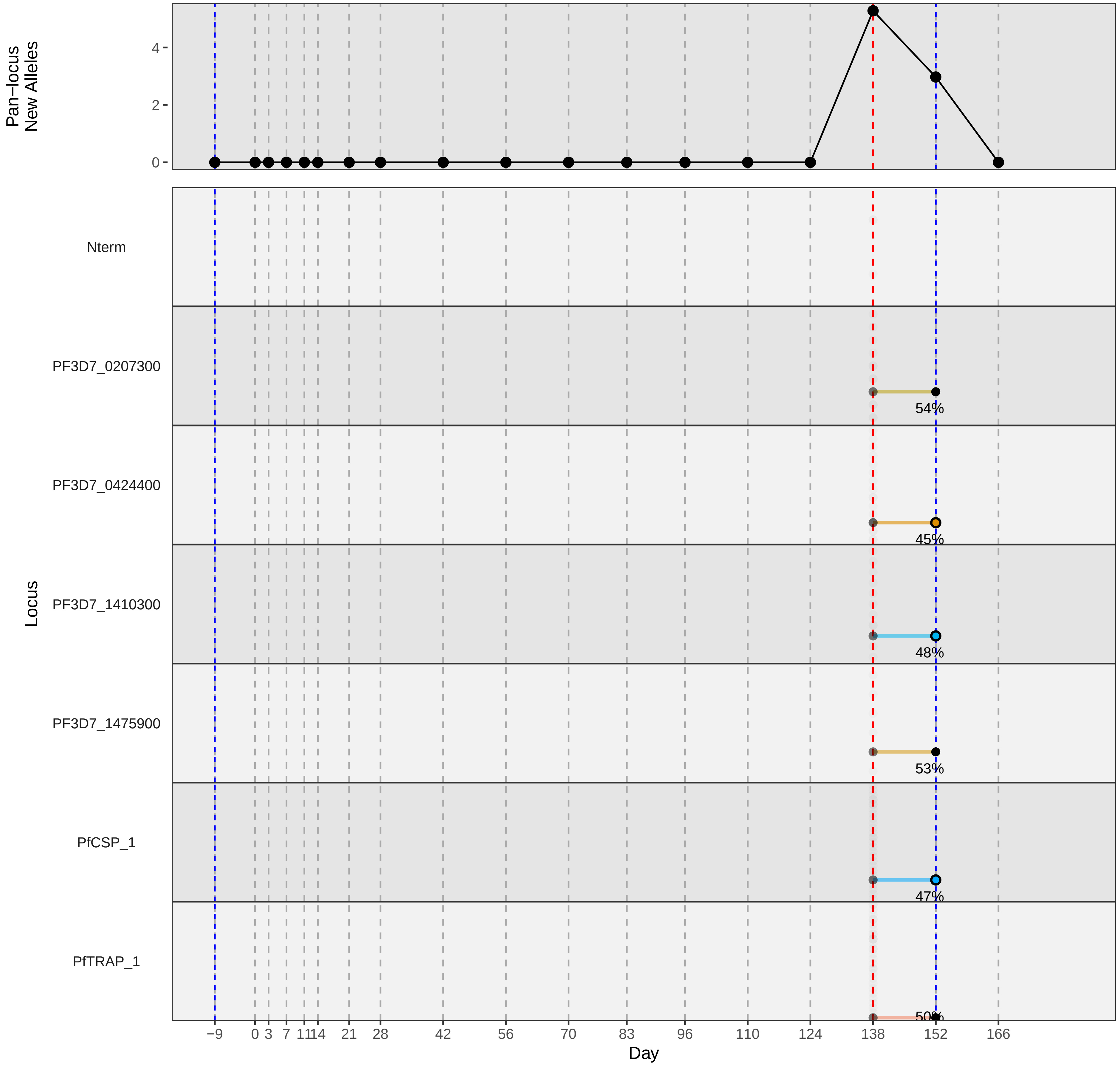

### participant_0186V_with_imputation.pdf

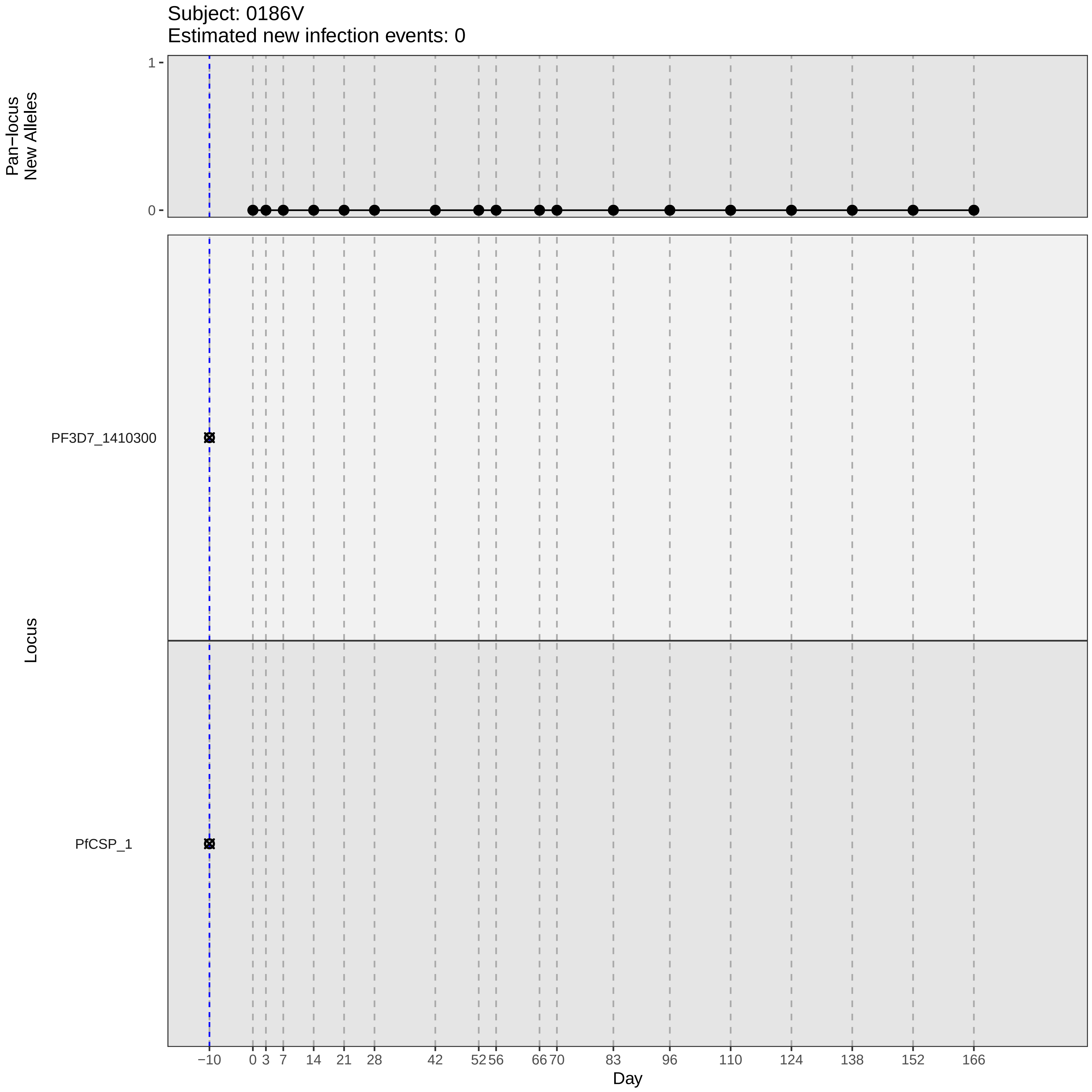
Subject: 0186V
Estimated new infection events: 0
Pan−locus New Alleles
1
0
Locus
PF3D7_1410300
PfCSP_1
−10
0
3
7
14
21
28
42
52
56
66
70
83
96
110
124
138
152
166
Day

### participant_0186V_without_imputation.pdf

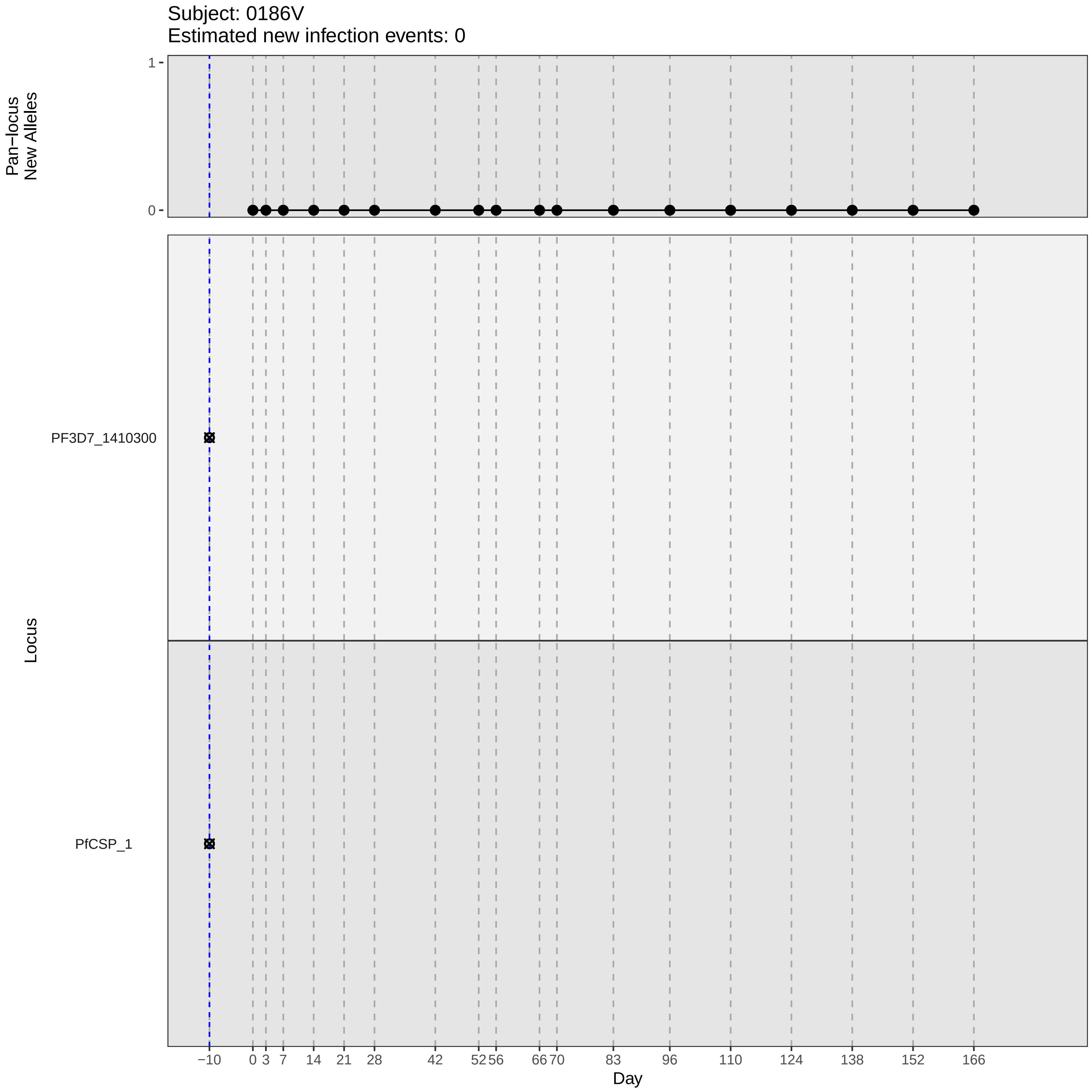

Subject: 0186V
Estimated new infection events: 0
Pan−locus
New Alleles
1
0
Locus
PF3D7_1410300
PfCSP_1
−10
0
3
7
14
21
28
42
52
56
66
70
83
96
110
124
138
152
166
Day

### participant_0187W_with_imputation.pdf

Subject: 0187W
Estimated new infection events: 1

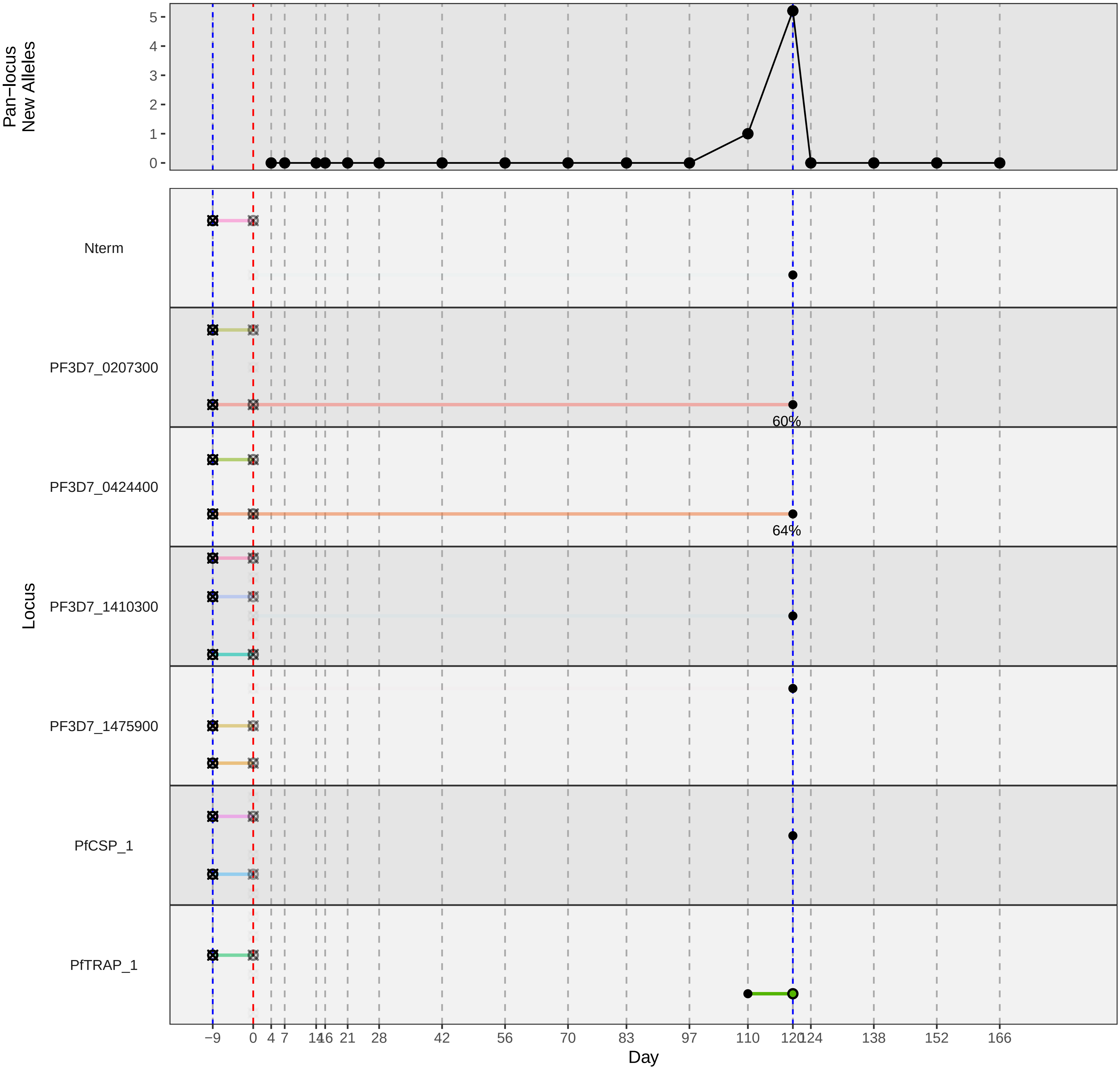

### participant_0224R_with_imputation.pdf

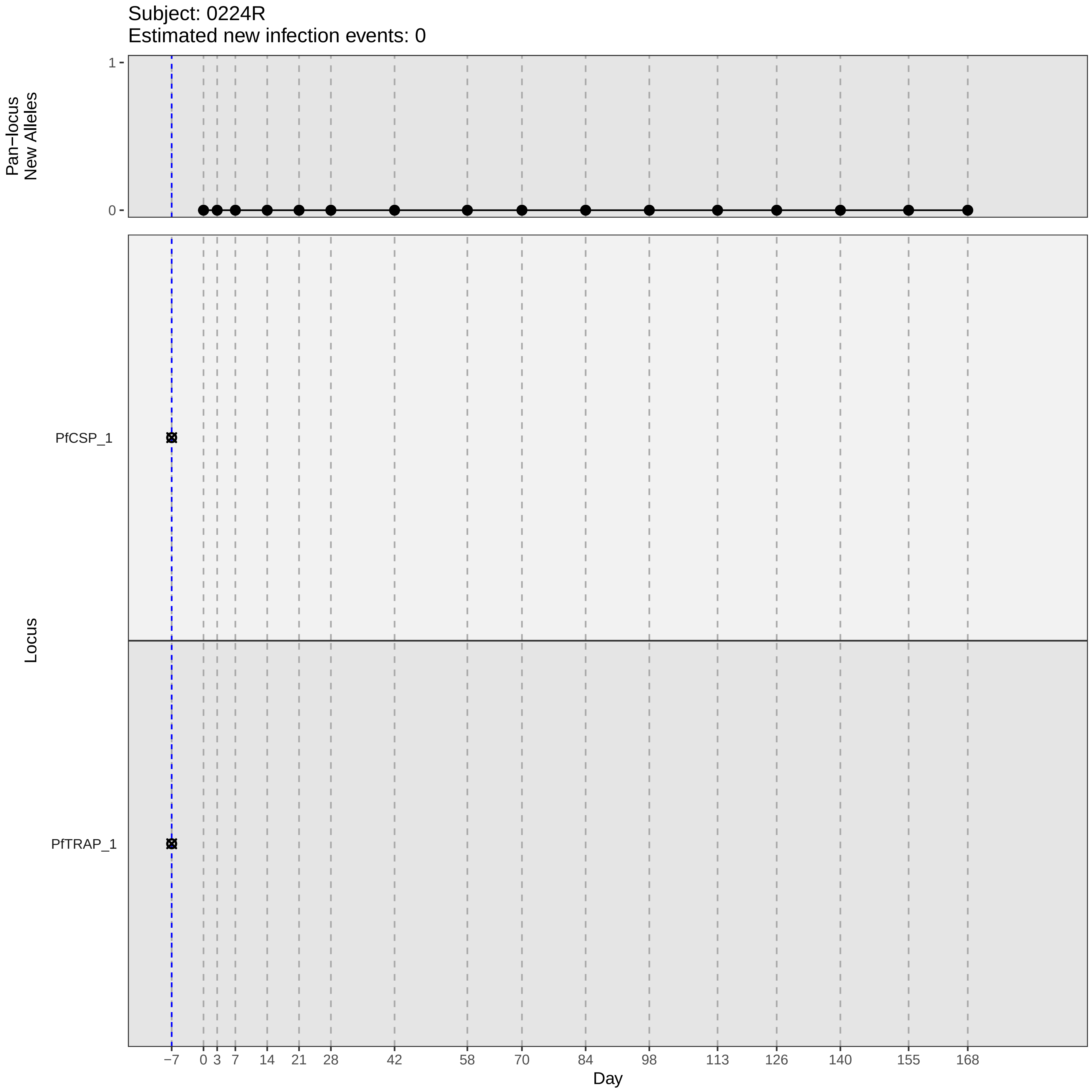
Subject: 0224R
Estimated new infection events: 0
Pan−locus New Alleles
1
0
Locus
PfCSP_1
PfTRAP_1
−7
0
3
7
14
21
28
42
58
70
84
98
113
126
140
155
168
Day

### participant_0224R_without_imputation.pdf

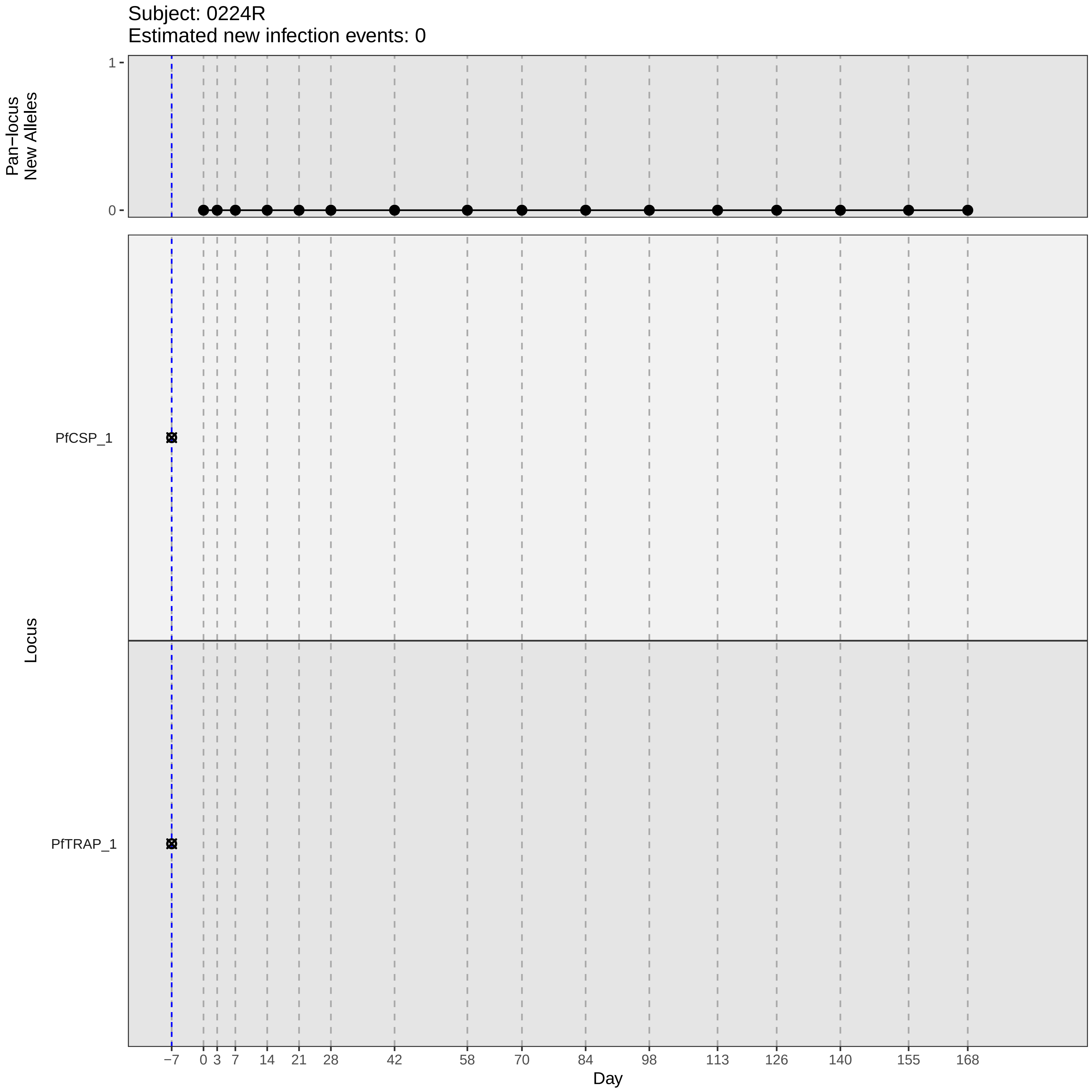
Subject: 0224R
Estimated new infection events: 0
Pan−locus
New Alleles
1
0
Locus
PfCSP_1
PfTRAP_1
−7
0
3
7
14
21
28
42
58
70
84
98
113
126
140
155
168
Day

### participant_0227U_with_imputation.pdf

Subject: 0227U
Estimated new infection events: 1.1

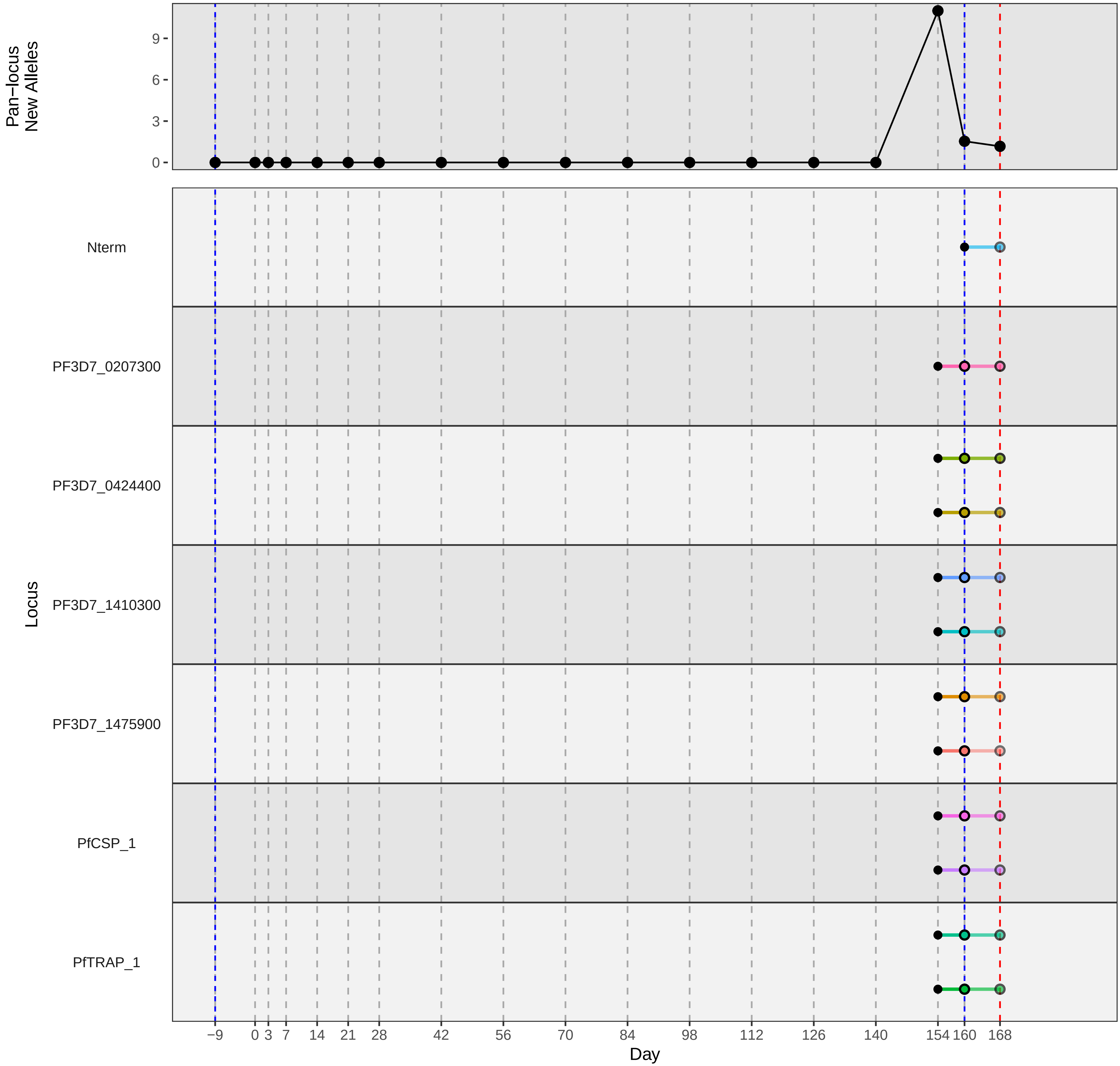

### participant_0232A_with_imputation.pdf

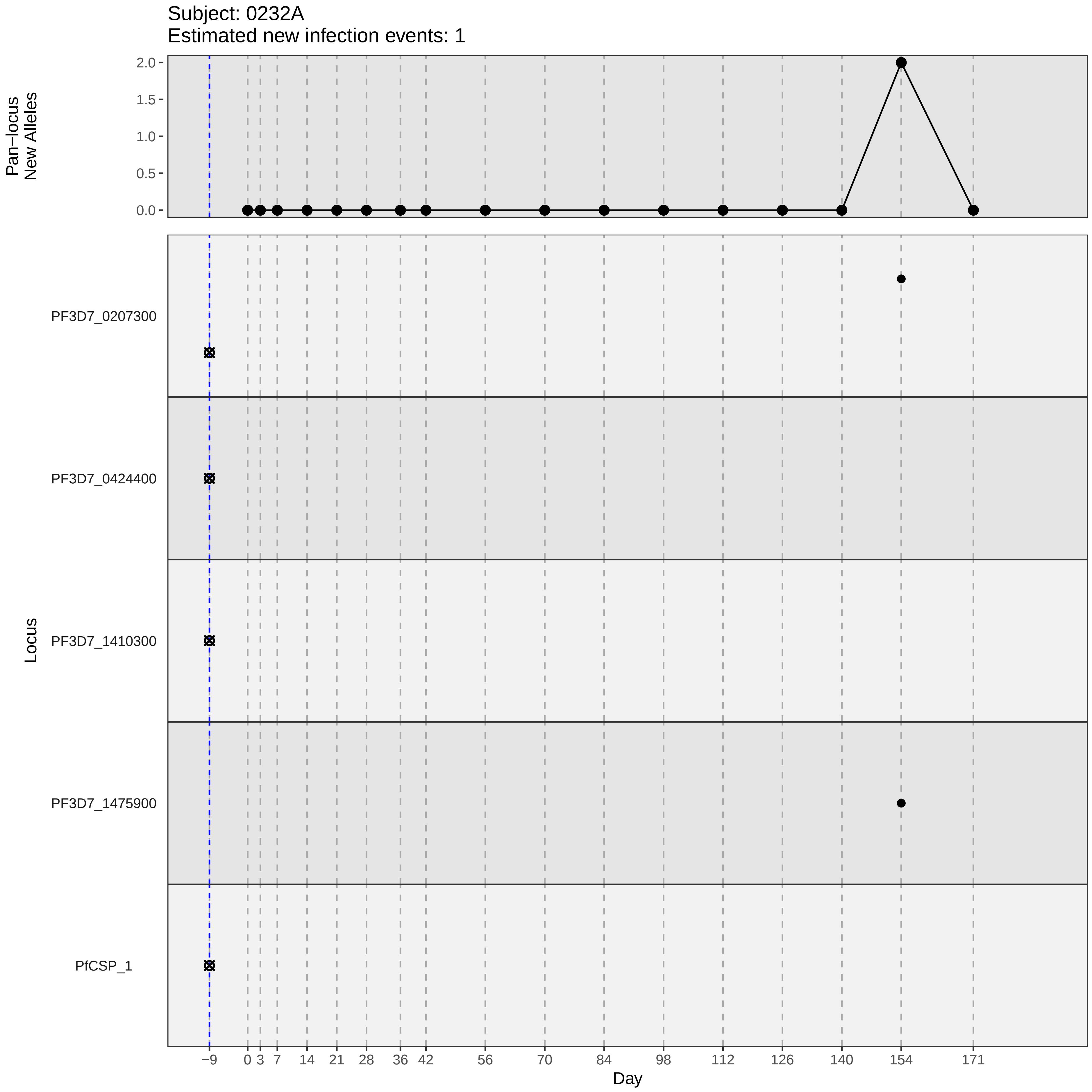

Subject: 0232A
Estimated new infection events: 1
Pan−locus
New Alleles
2.0
1.5
1.0
0.5
0.0
Locus
PF3D7_0207300
PF3D7_0424400
PF3D7_1410300
PF3D7_1475900
PfCSP_1
−9
0
3
7
14
21
28
36
42
56
70
84
98
112
126
140
154
171
Day

### participant_0232A_without_imputation.pdf

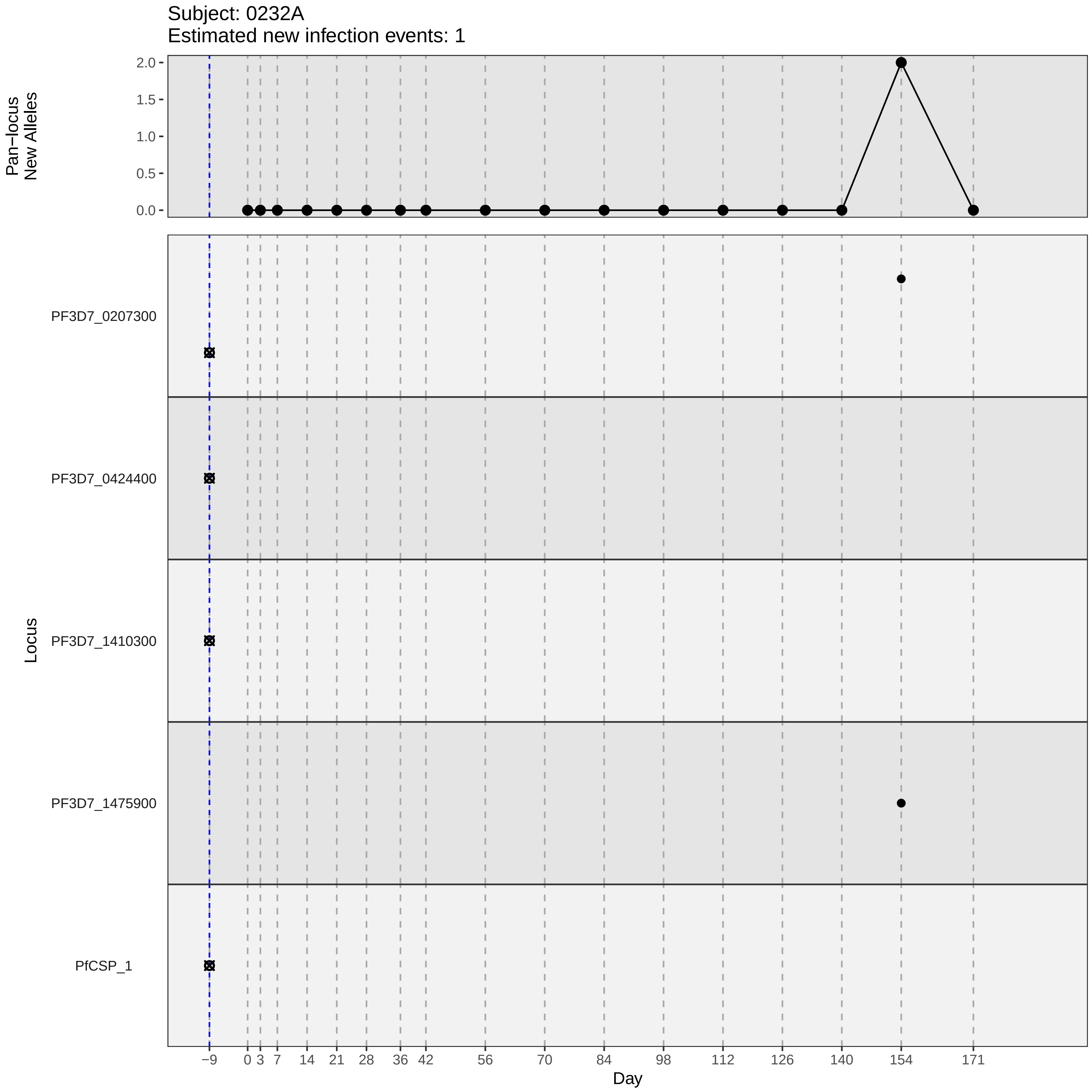

Subject: 0232A
Estimated new infection events: 1
Pan−locus New Alleles
2.0
1.5
1.0
0.5
0.0
Locus
PF3D7_0207300
PF3D7_0424400
PF3D7_1410300
PF3D7_1475900
PfCSP_1
−9
0
3
7
14
21
28
36
42
56
70
84
98
112
126
140
154
171
Day

### participant_0235D_with_imputation.pdf

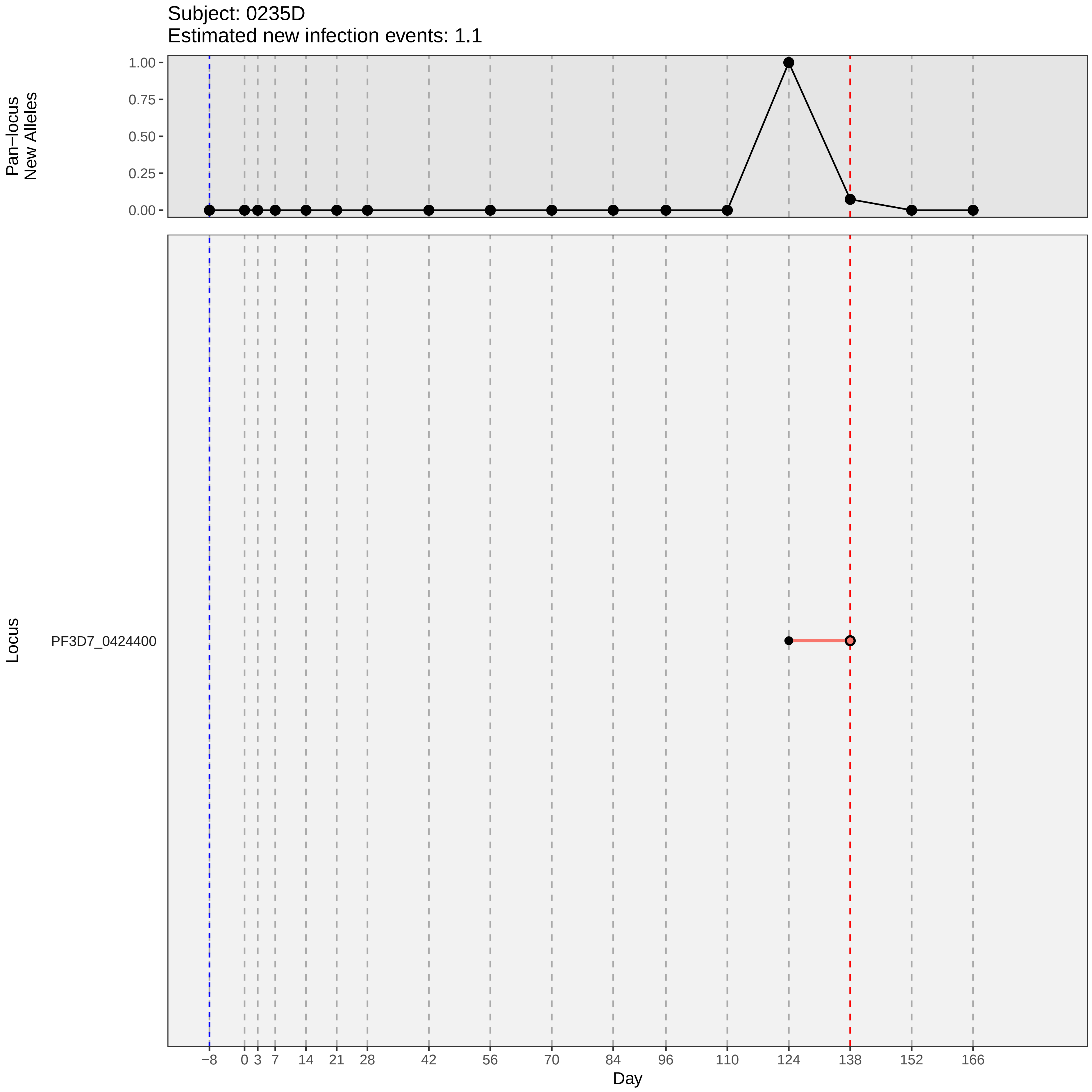

Subject: 0235D
Estimated new infection events: 1.1
Pan−locus New Alleles
1.00
0.75
0.50
0.25
0.00
Locus
PF3D7_0424400
−8
0
3
7
14
21
28
42
56
70
84
96
110
124
138
152
166
Day

### participant_0235D_without_imputation.pdf

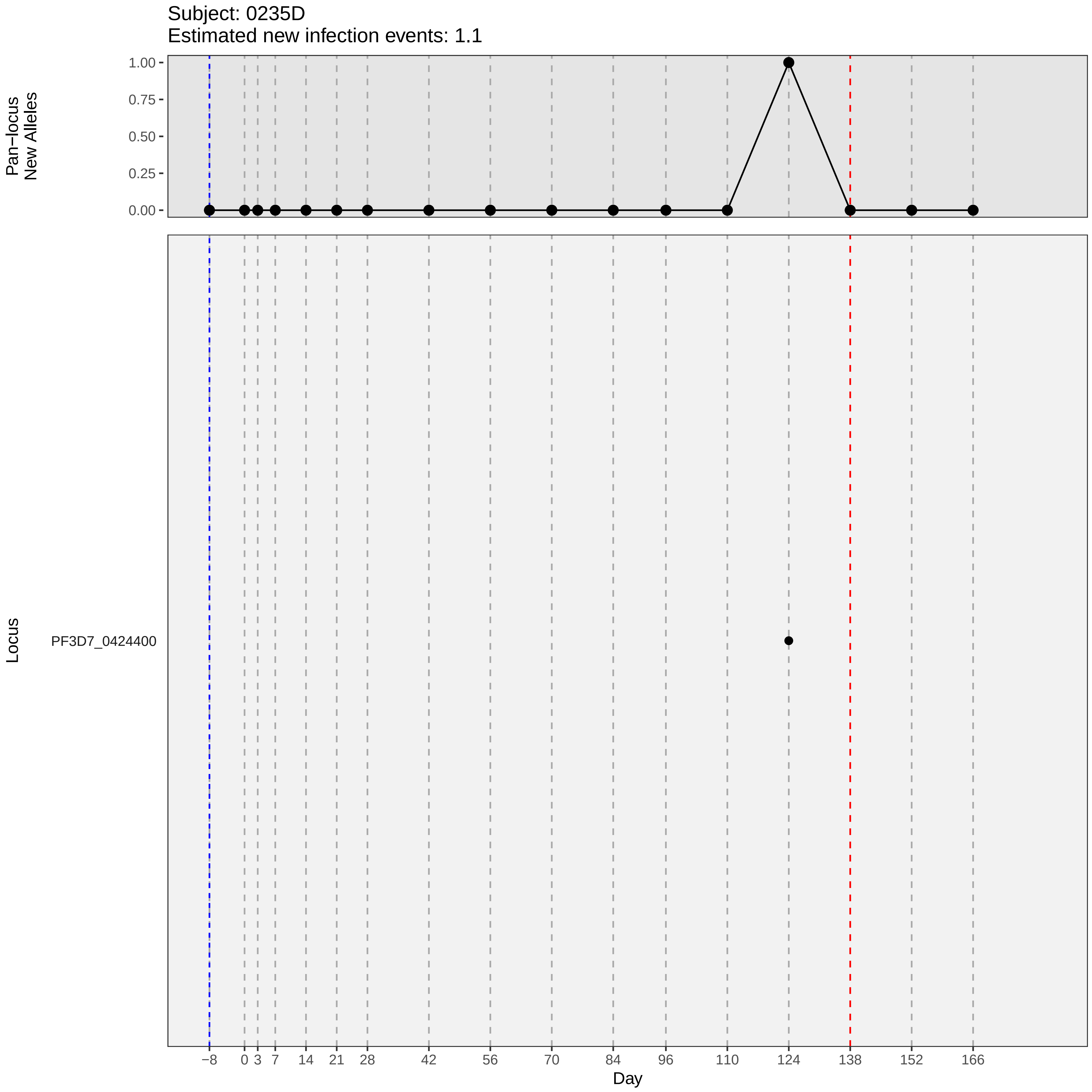
Subject: 0235D
Estimated new infection events: 1.1
Pan−locus
New Alleles
1.00
0.75
0.50
0.25
0.00
Locus
PF3D7_0424400
−8
0
3
7
14
21
28
42
56
70
84
96
110
124
138
152
166
Day

### participant_0237F_with_imputation.pdf

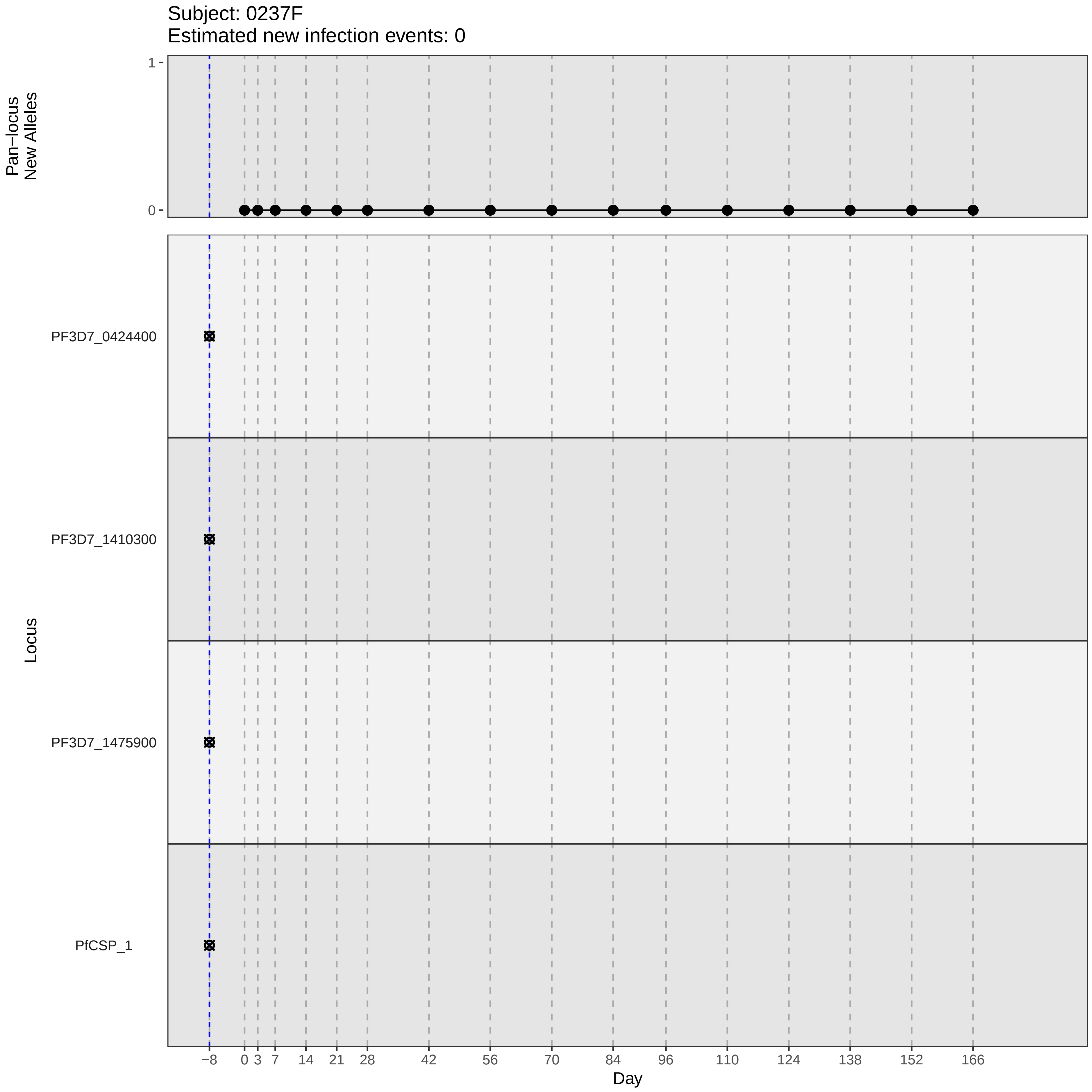
Subject: 0237F
Estimated new infection events: 0
Pan−locus
New Alleles
1
0
Locus
PF3D7_0424400
PF3D7_1410300
PF3D7_1475900
PfCSP_1
−8
0
3
7
14
21
28
42
56
70
84
96
110
124
138
152
166
Day

### participant_0237F_without_imputation.pdf

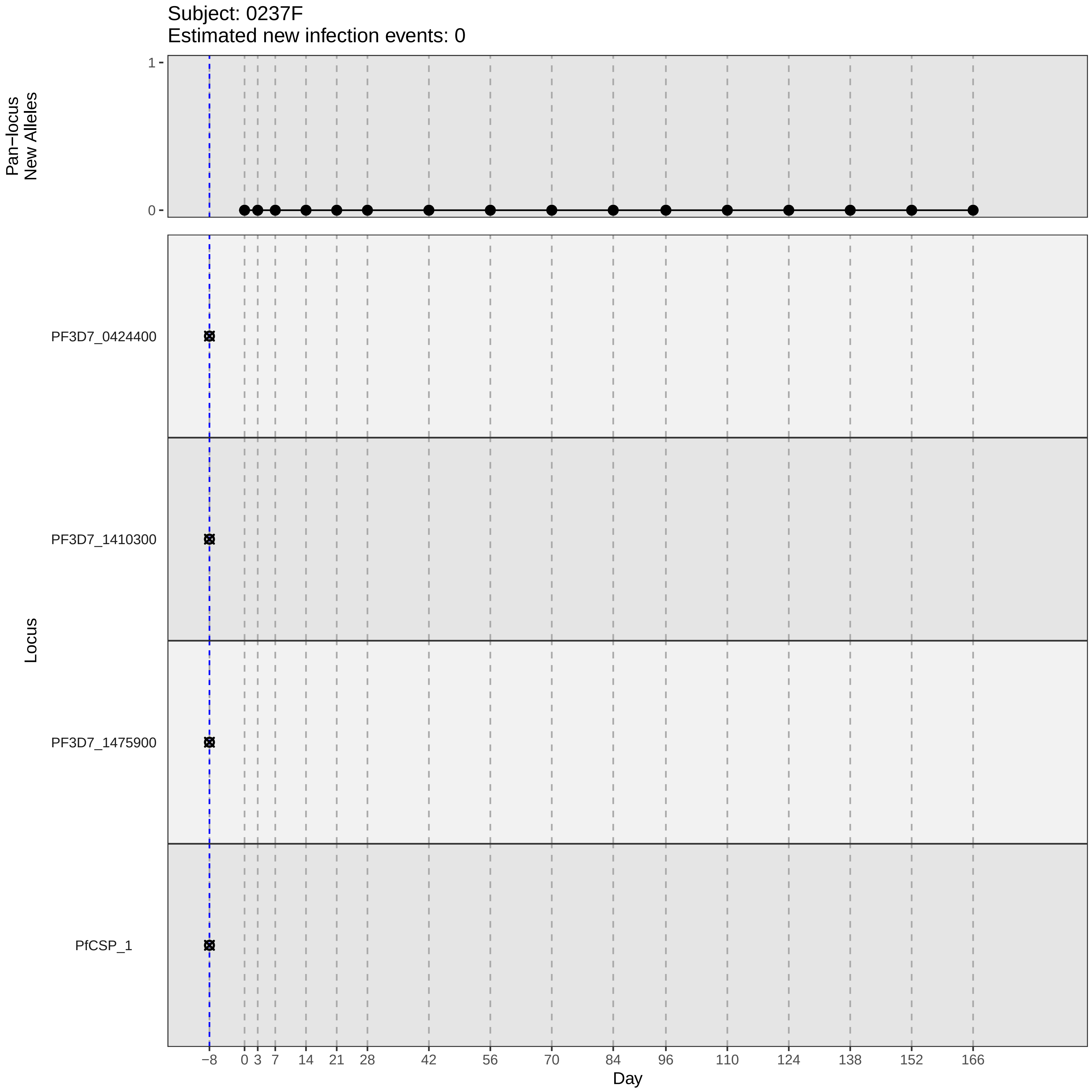

Subject: 0237F
Estimated new infection events: 0
Pan−locus New Alleles
1
0
Locus
PF3D7_0424400
PF3D7_1410300
PF3D7_1475900
PfCSP_1
−8
0
3
7
14
21
28
42
56
70
84
96
110
124
138
152
166
Day

### participant_0242N_with_imputation.pdf

Subject: 0242N
Estimated new infection events: 3.1

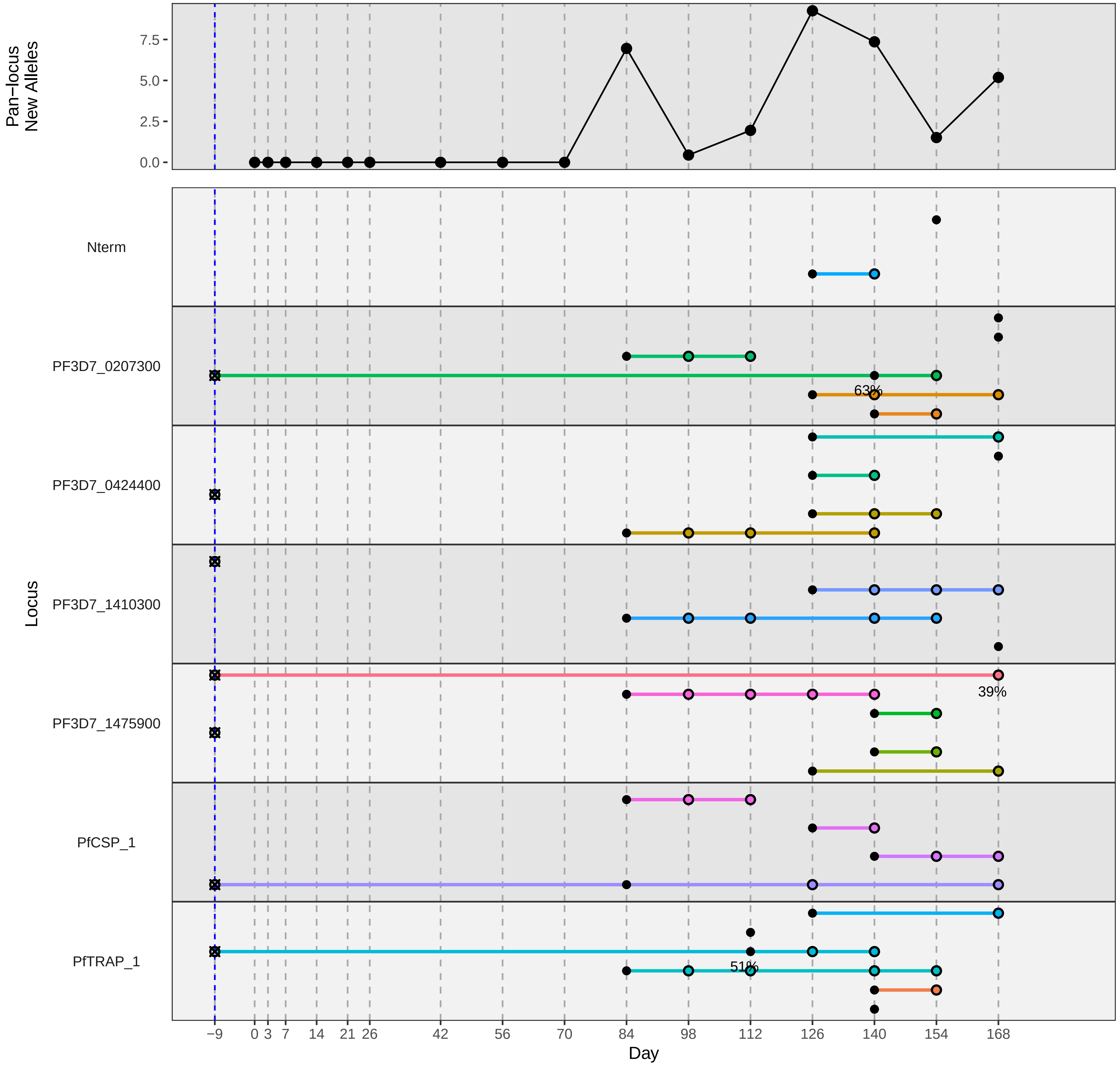

### participant_0247T_with_imputation.pdf

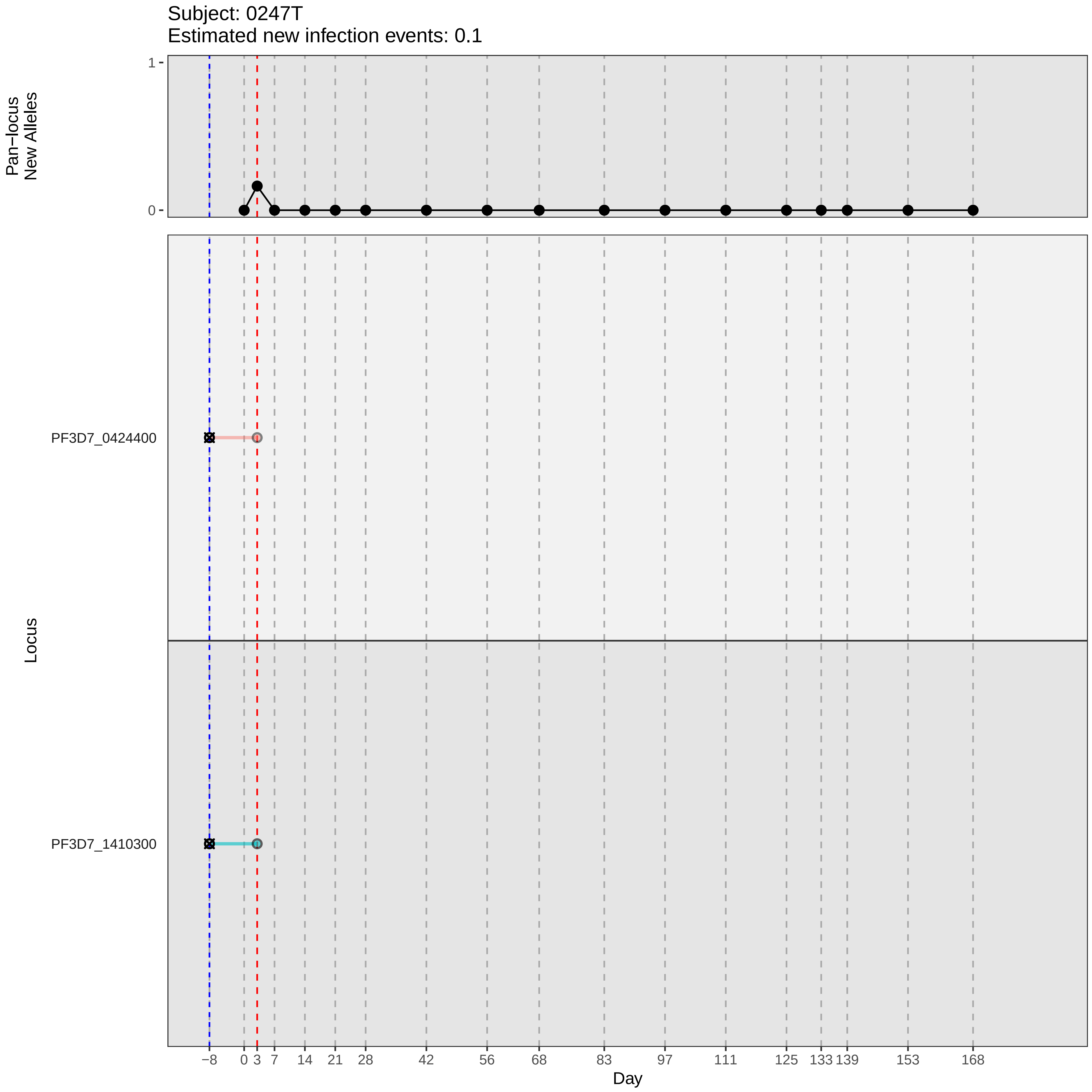

Subject: 0247T
Estimated new infection events: 0.1
Pan−locus New Alleles
1
0
Locus
PF3D7_0424400
PF3D7_1410300
−8
0
3
7
14
21
28
42
56
68
83
97
111
125
133
139
153
168
Day

### participant_0247T_without_imputation.pdf

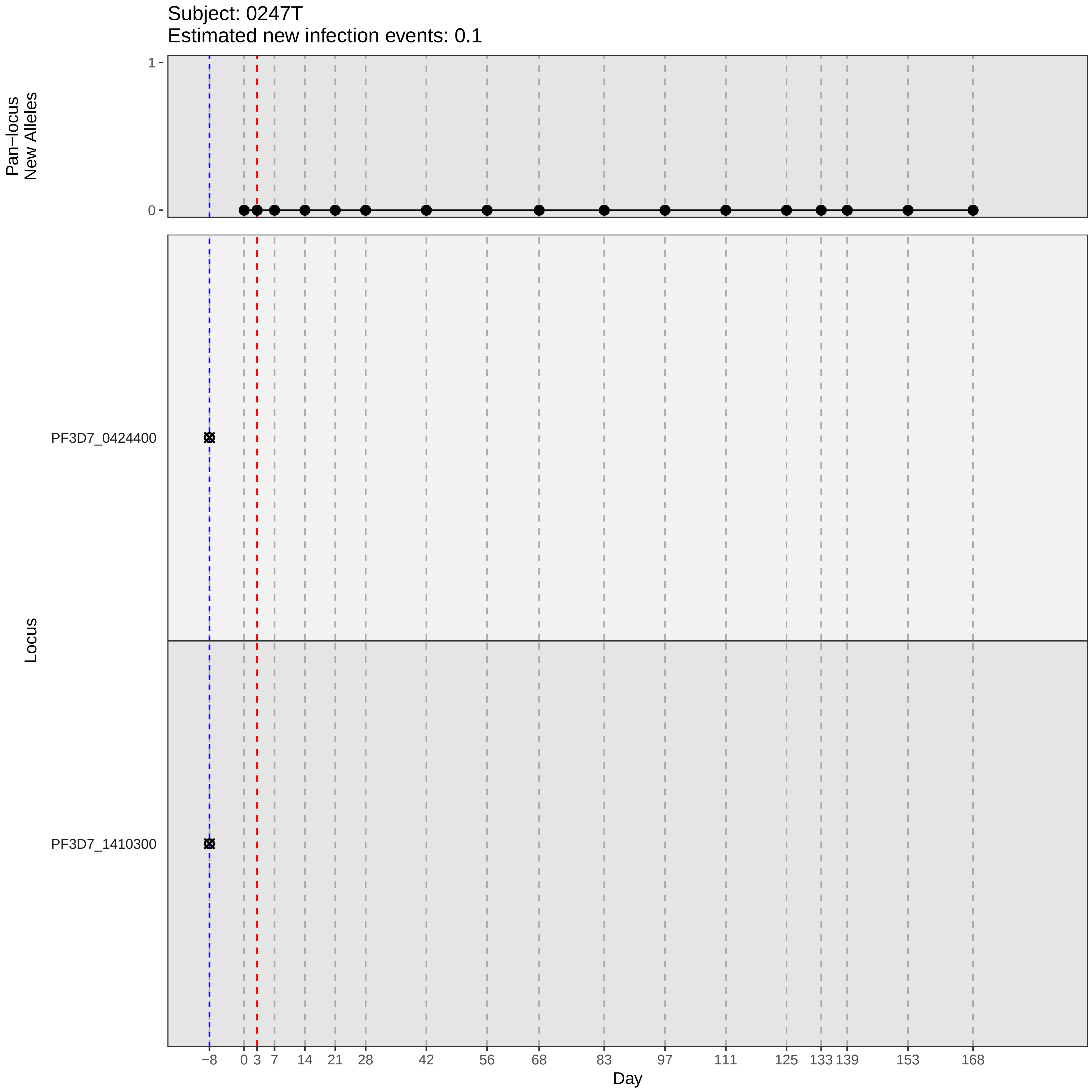

Subject: 0247T
Estimated new infection events: 0.1
Pan−locus
New Alleles
1
0
Locus
PF3D7_0424400
PF3D7_1410300
−8
0
3
7
14
21
28
42
56
68
83
97
111
125
133
139
153
168
Day

### participant_0252Y_with_imputation.pdf

Subject: 0252Y
Estimated new infection events: 1

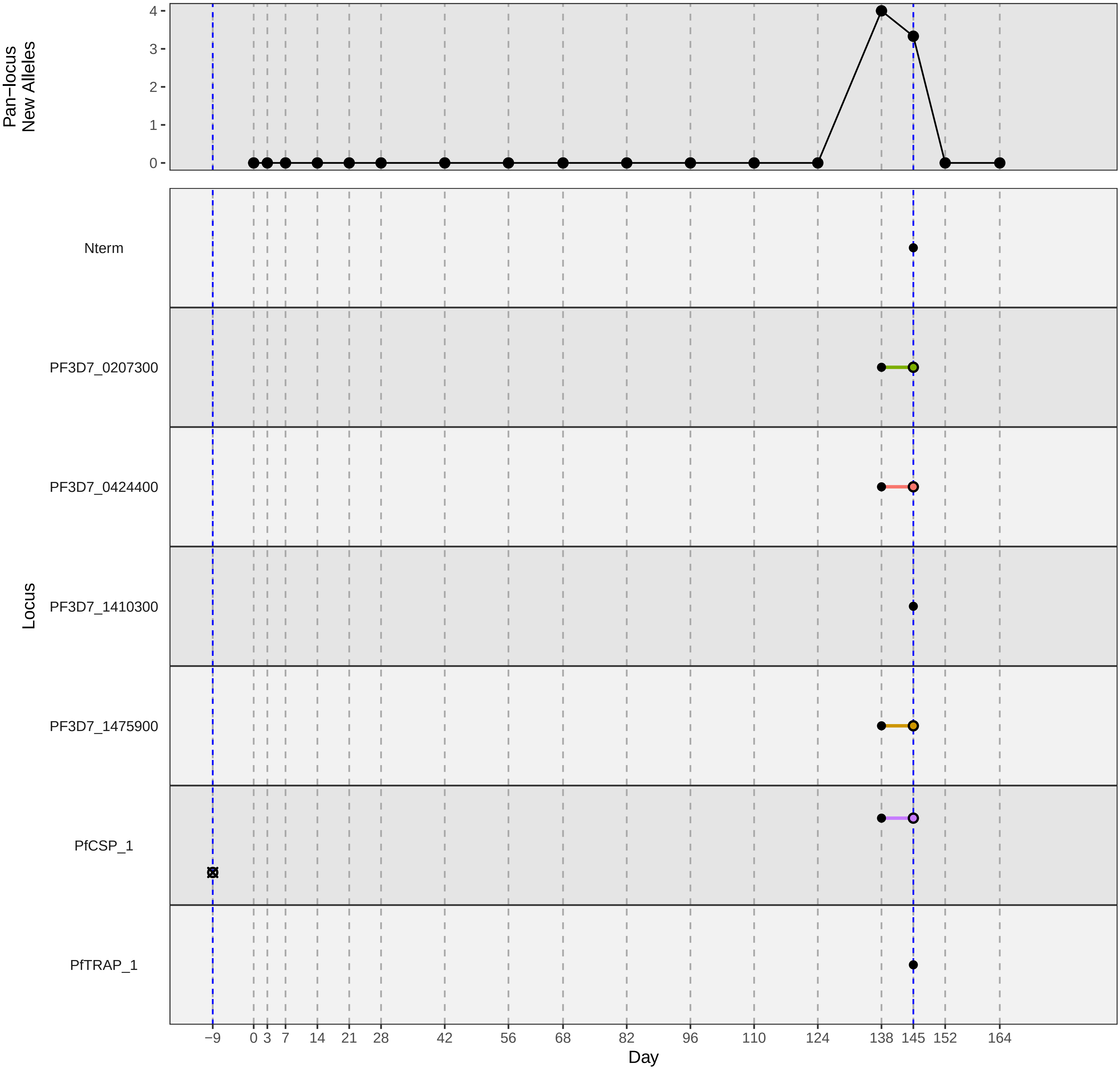

### participant_0258F_with_imputation.pdf

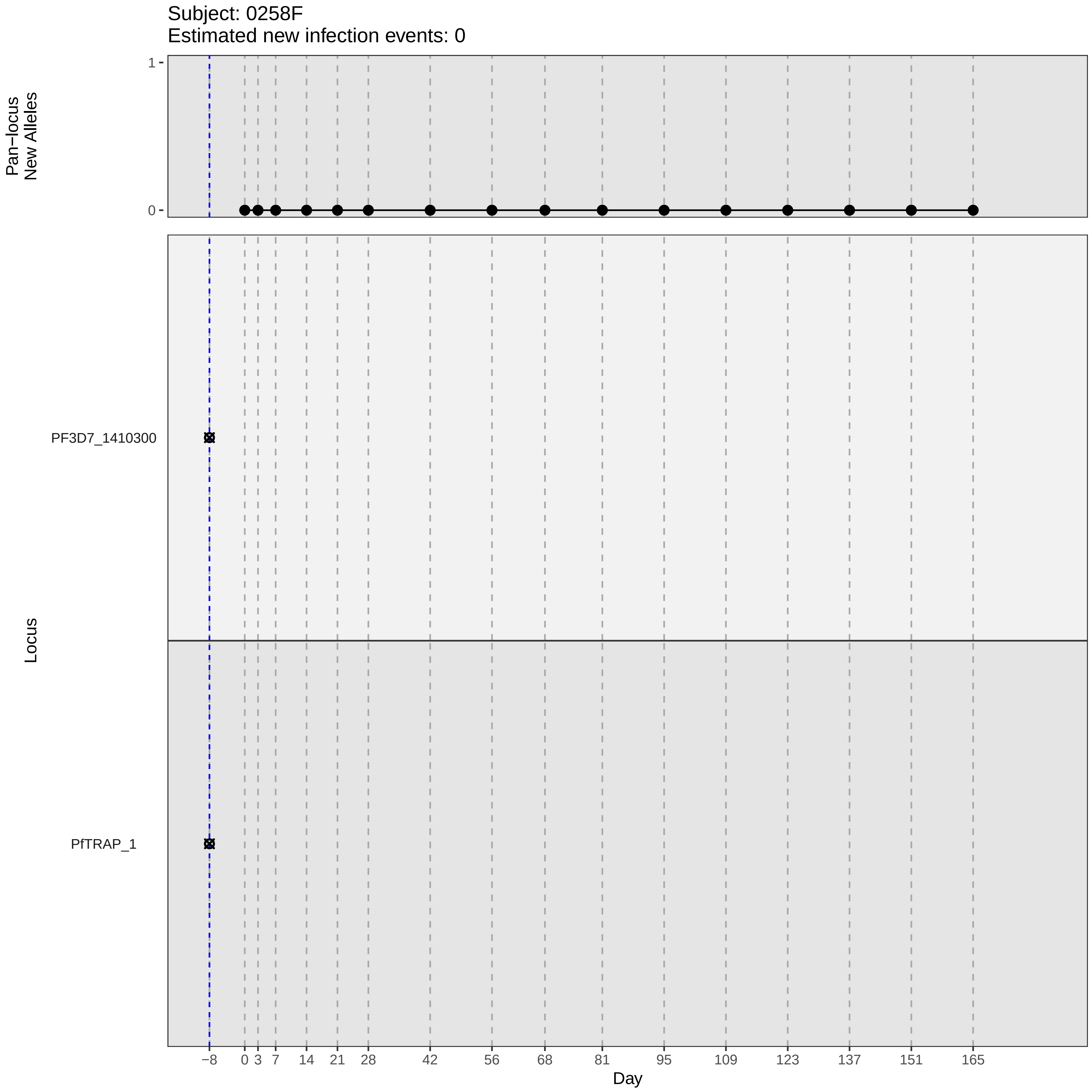

Subject: 0258F
Estimated new infection events: 0
Pan−locus
New Alleles
1
0
Locus
PF3D7_1410300
PfTRAP_1
−8
0
3
7
14
21
28
42
56
68
81
95
109
123
137
151
165
Day

### participant_0258F_without_imputation.pdf

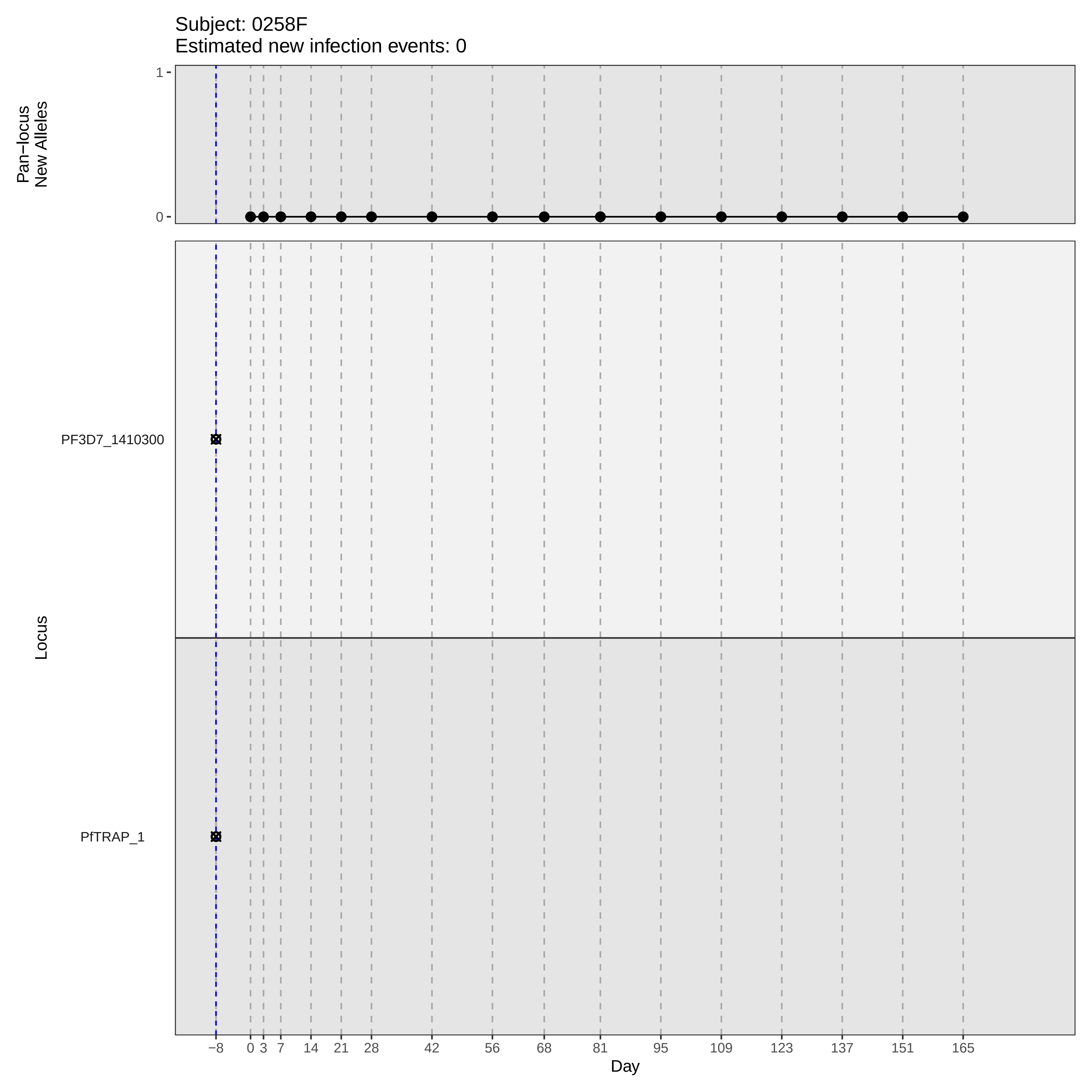
Subject: 0258F
Estimated new infection events: 0
Pan−locus
New Alleles
1
0
Locus
PF3D7_1410300
PfTRAP_1
−8
0
3
7
14
21
28
42
56
68
81
95
109
123
137
151
165
Day

### participant_0260H_without_imputation.pdf

Subject: 0260H
Estimated new infection events: 0

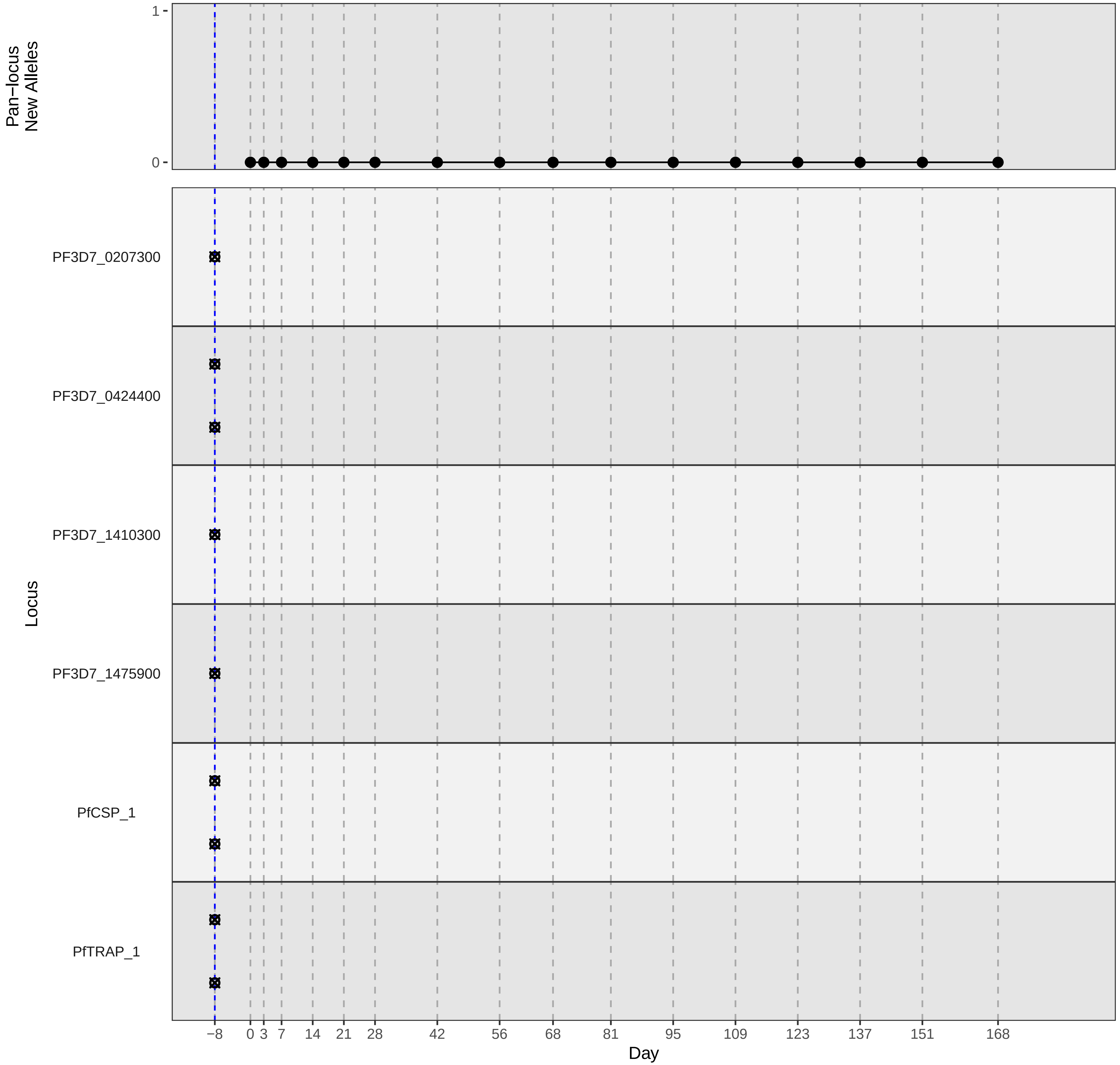

### participant_0267S_without_imputation.pdf

Subject: 0267S
Estimated new infection events: 1.8

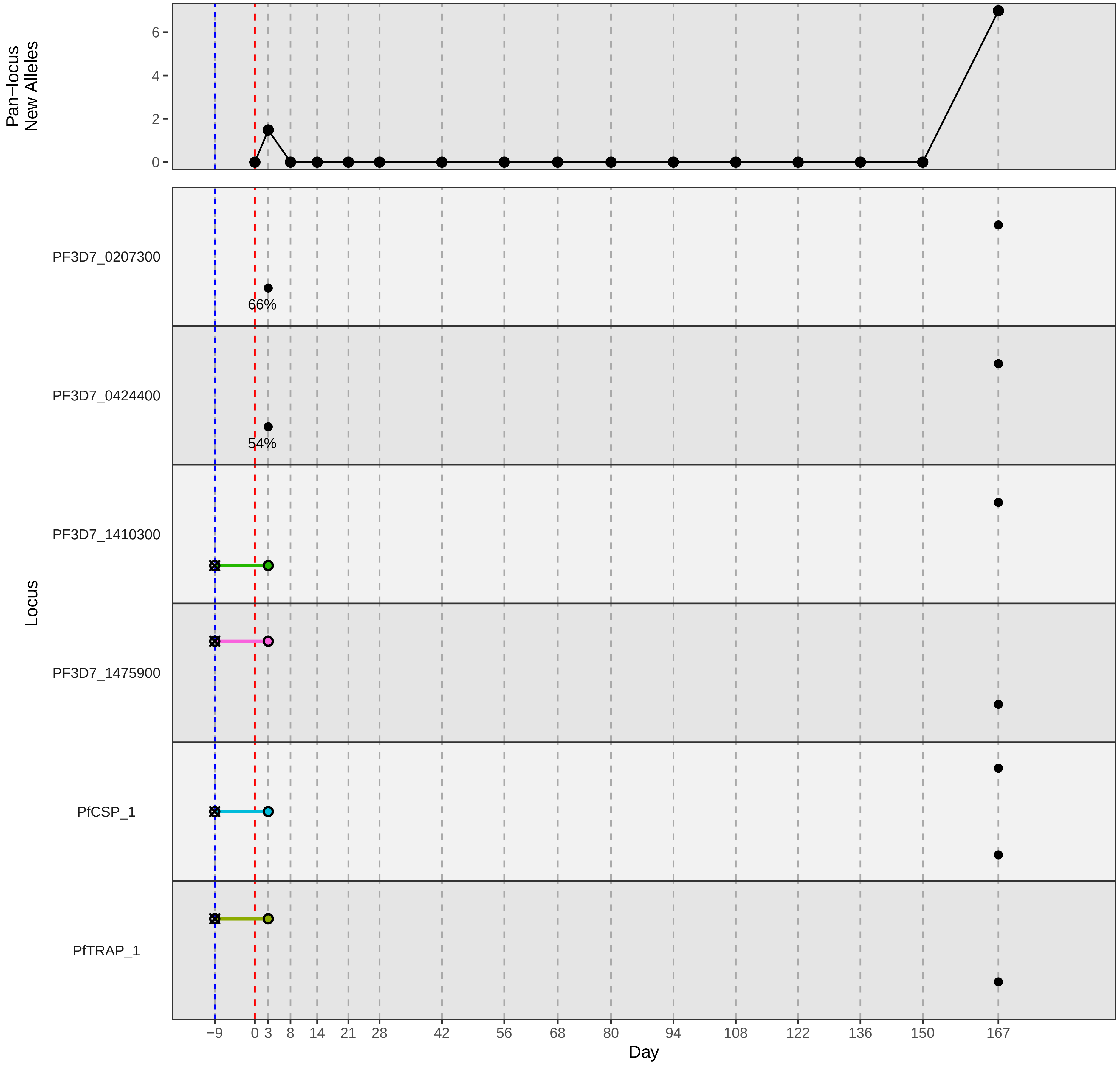

### participant_0270V_with_imputation.pdf

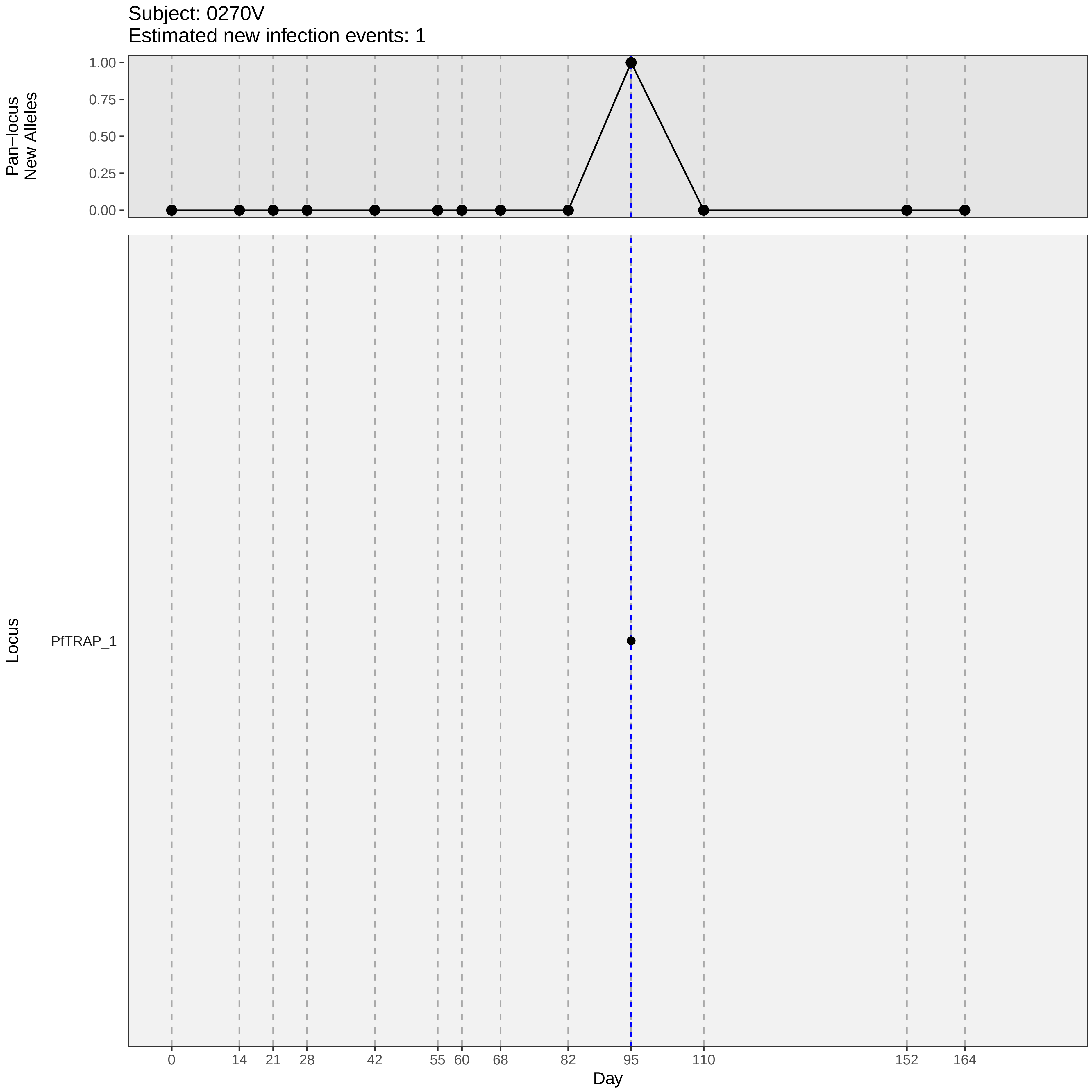

Subject: 0270V
Estimated new infection events: 1
Pan−locus
New Alleles
1.00
0.75
0.50
0.25
0.00
Locus
PfTRAP_1
0
14
21
28
42
55
60
68
82
95
110
152
164
Day

### participant_0270V_without_imputation.pdf

Subject: 0270V
Estimated new infection events: 1

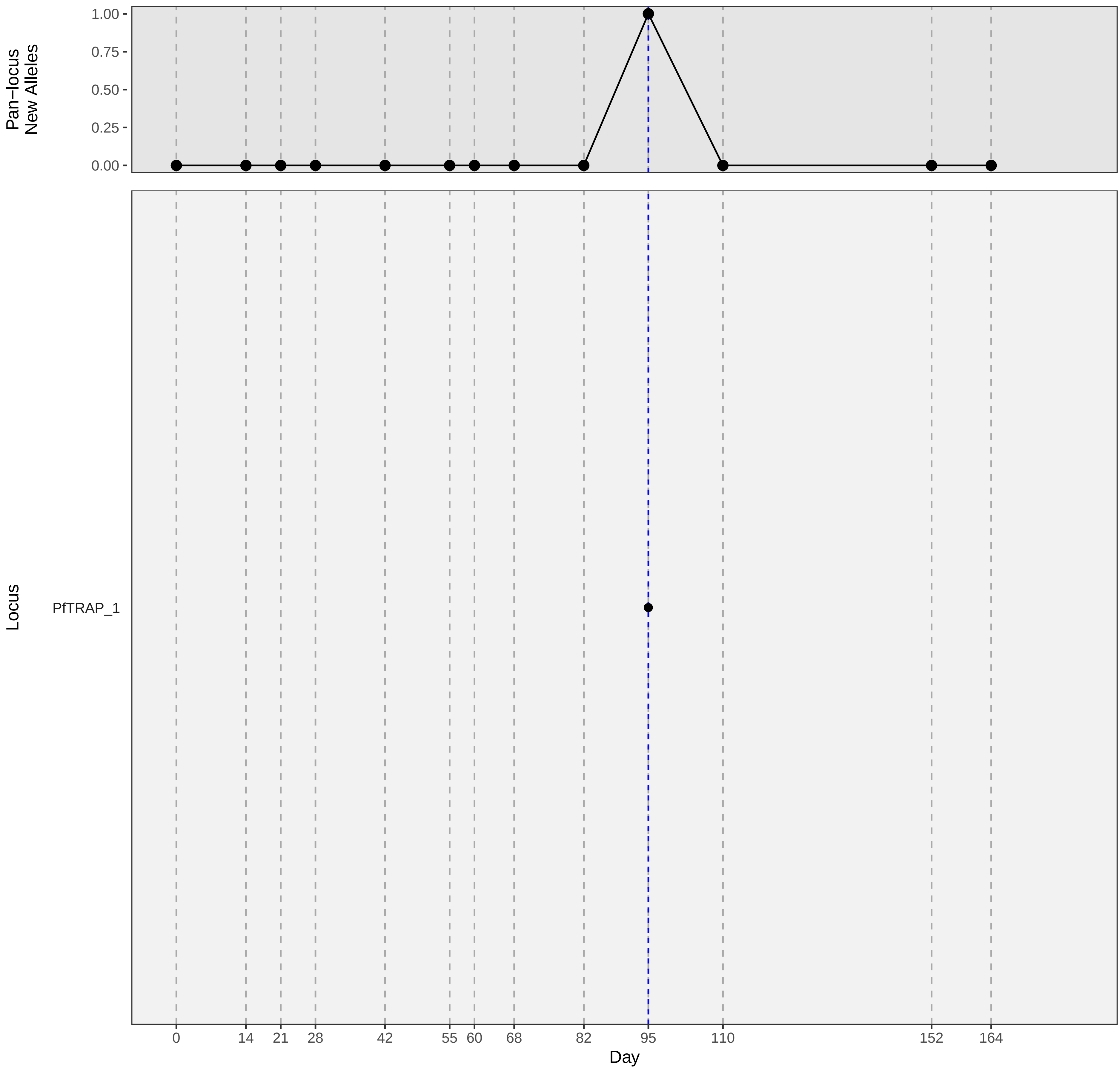

### participant_0272X_without_imputation.pdf

Subject: 0272X
Estimated new infection events: 3.1

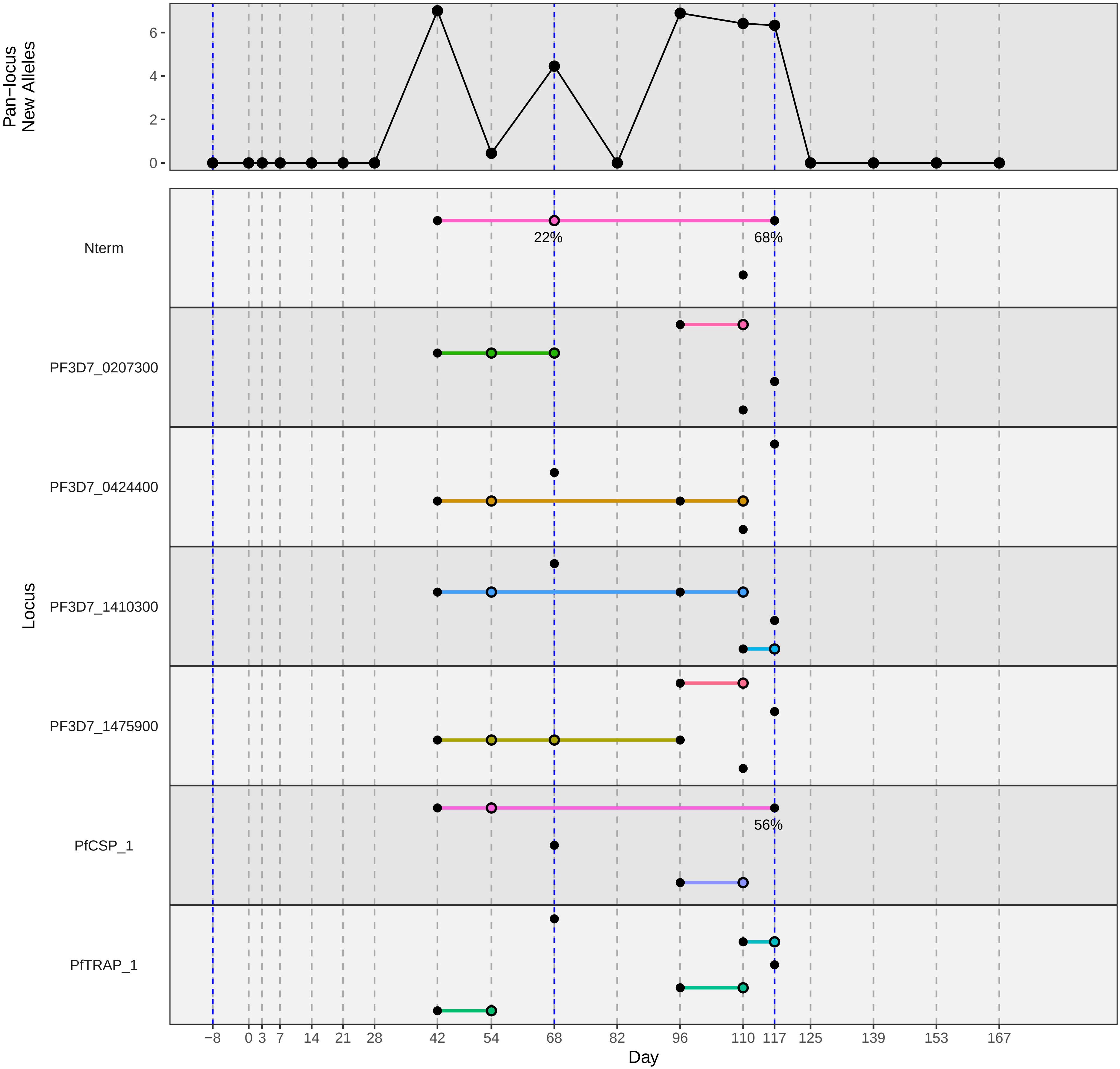

### participant_0278E_with_imputation.pdf

Subject: 0278E
Estimated new infection events: 2.1

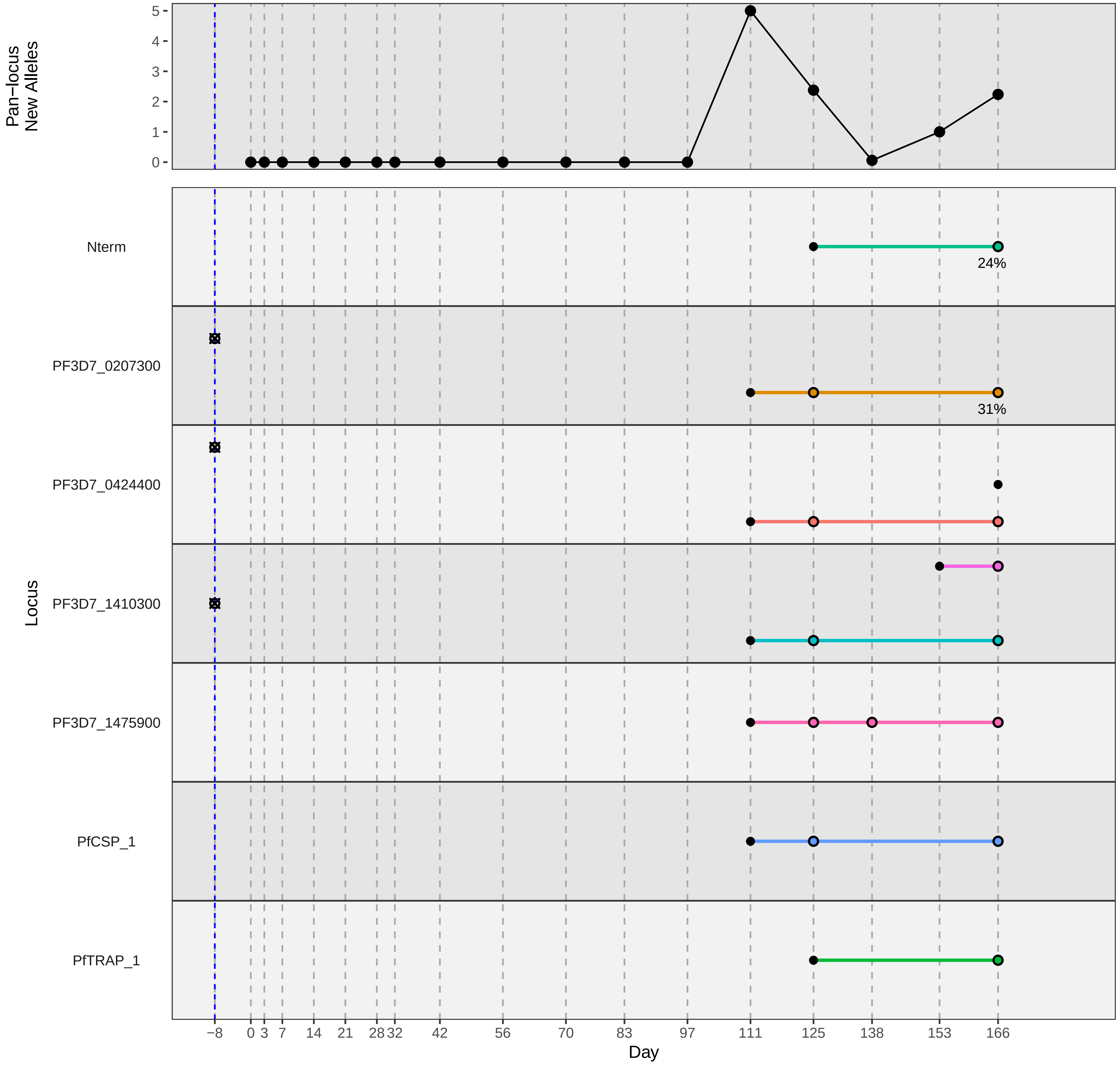

### participant_0285P_with_imputation.pdf

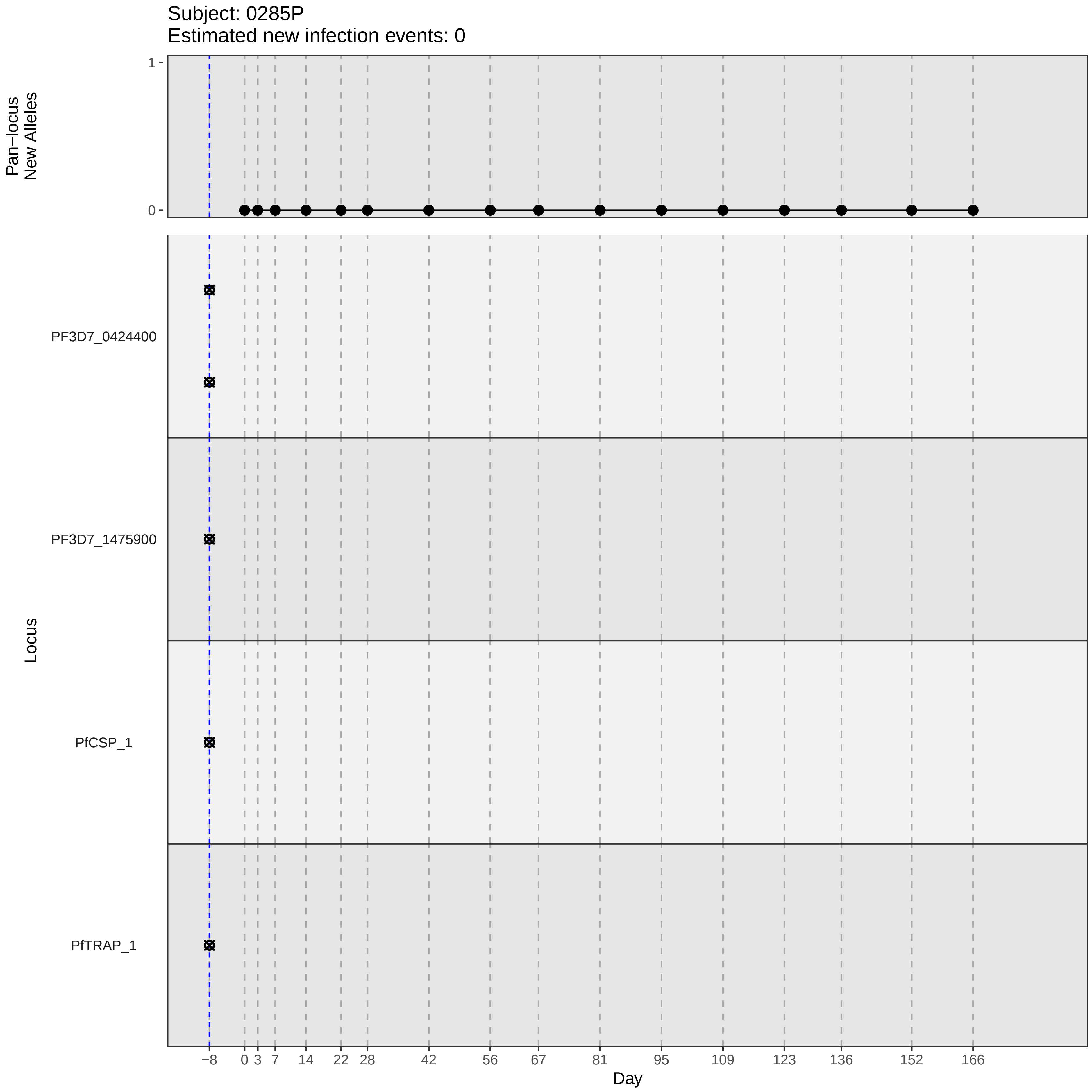
Subject: 0285P
Estimated new infection events: 0
Pan−locus
New Alleles
1
0
Locus
PF3D7_0424400
PF3D7_1475900
PfCSP_1
PfTRAP_1
−8
0
3
7
14
22
28
42
56
67
81
95
109
123
136
152
166
Day

### participant_0285P_without_imputation.pdf

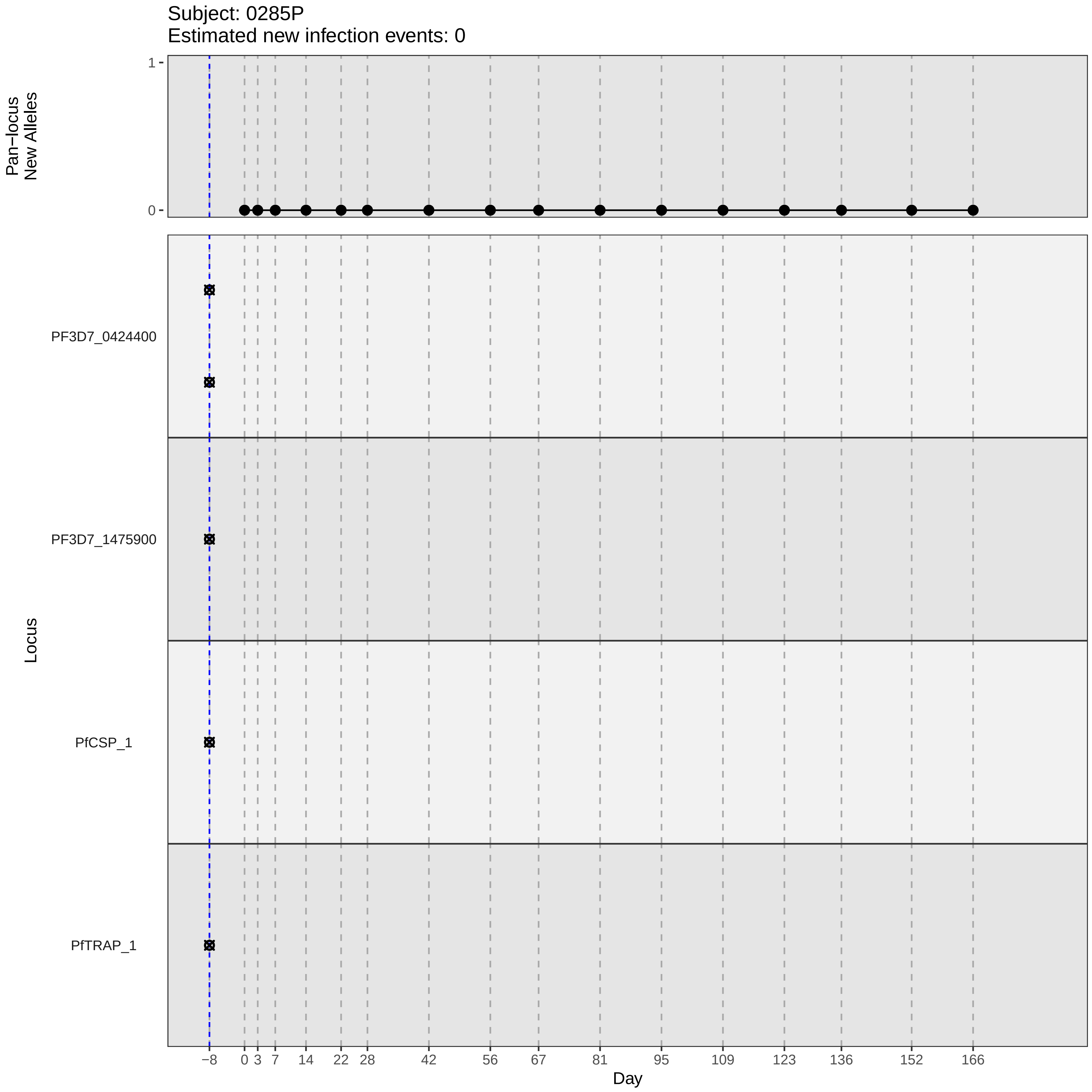
Subject: 0285P
Estimated new infection events: 0
Pan−locus New Alleles
1
0
Locus
PF3D7_0424400
PF3D7_1475900
PfCSP_1
PfTRAP_1
−8
0
3
7
14
22
28
42
56
67
81
95
109
123
136
152
166
Day

### participant_0291V_with_imputation.pdf

Subject: 0291V
Estimated new infection events: 5.7

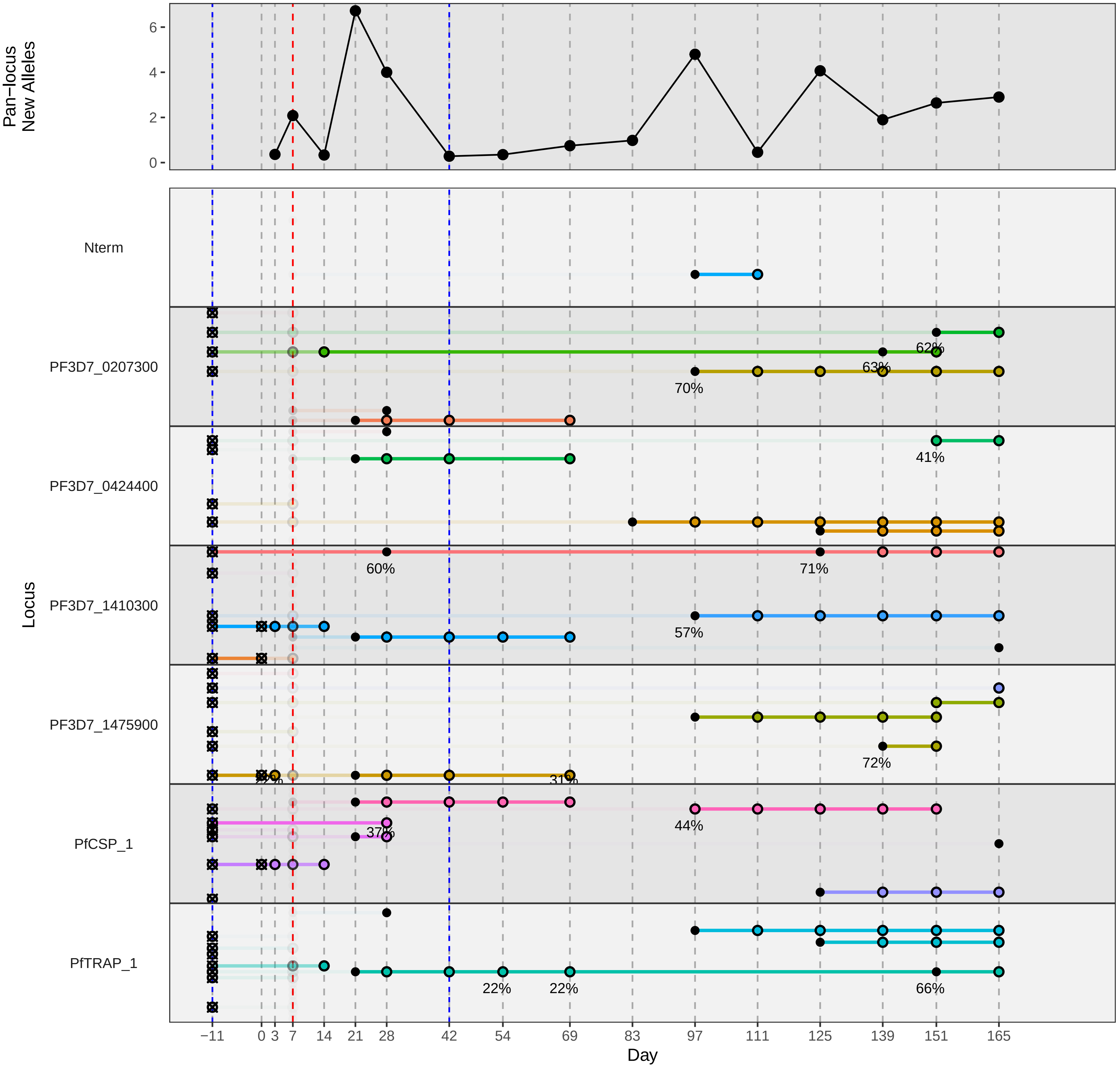

### participant_0305N_with_imputation.pdf

Subject: 0305N
Estimated new infection events: 4.7

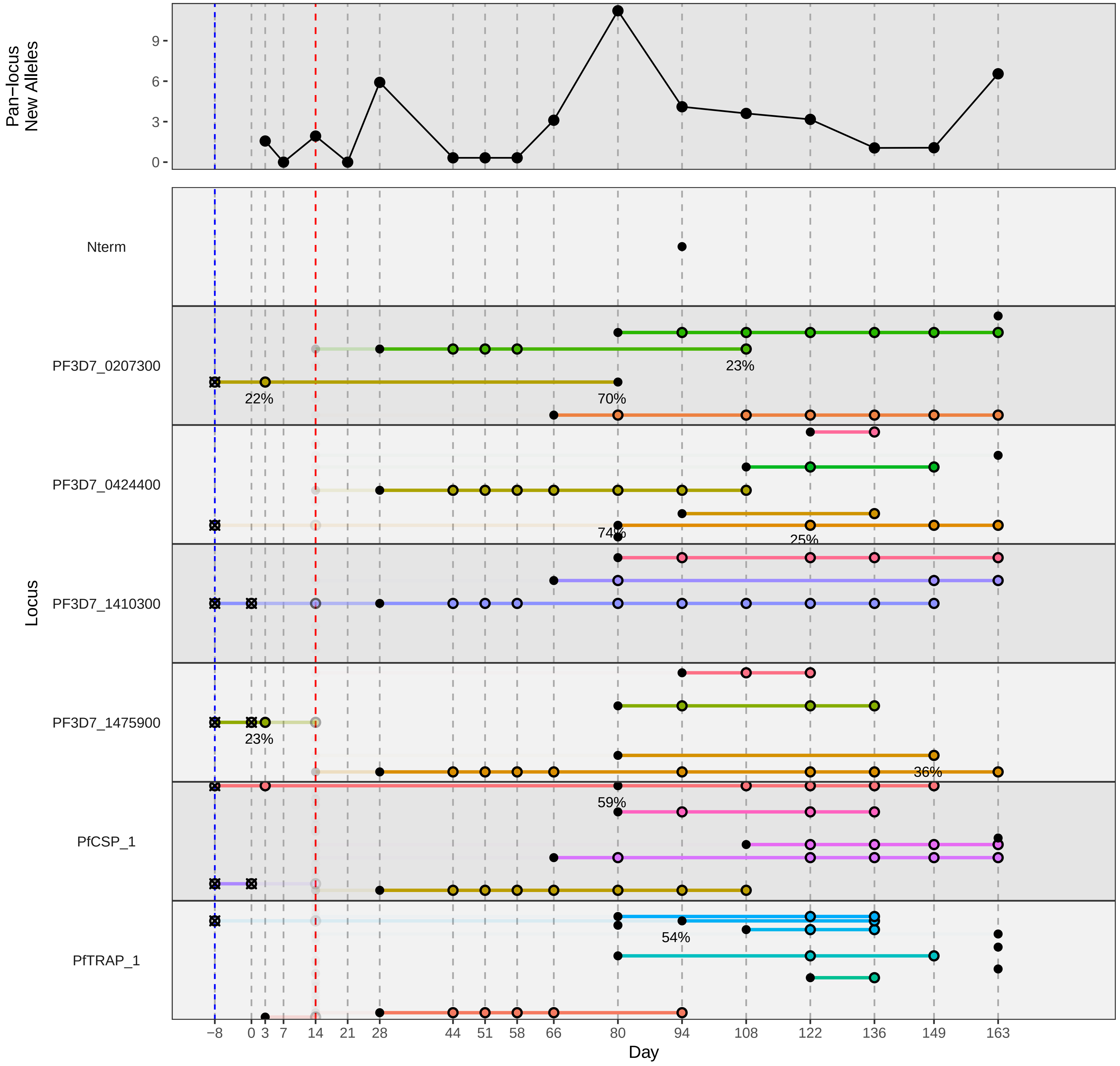

### participant_0307Q_with_imputation.pdf

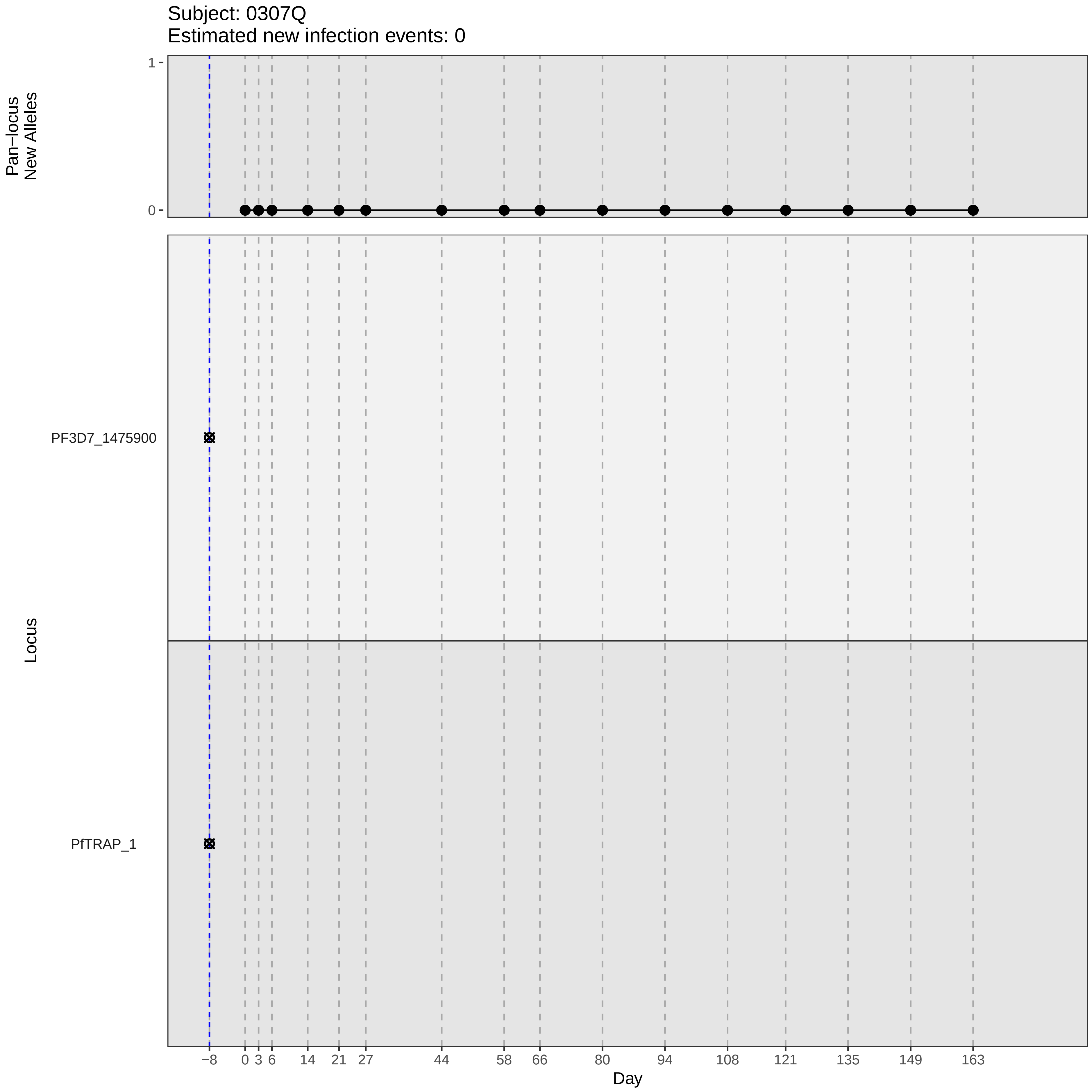
Subject: 0307Q
Estimated new infection events: 0
Pan−locus
New Alleles
1
0
Locus
PF3D7_1475900
PfTRAP_1
−8
0
3
6
14
21
27
44
58
66
80
94
108
121
135
149
163
Day

### participant_0307Q_without_imputation.pdf

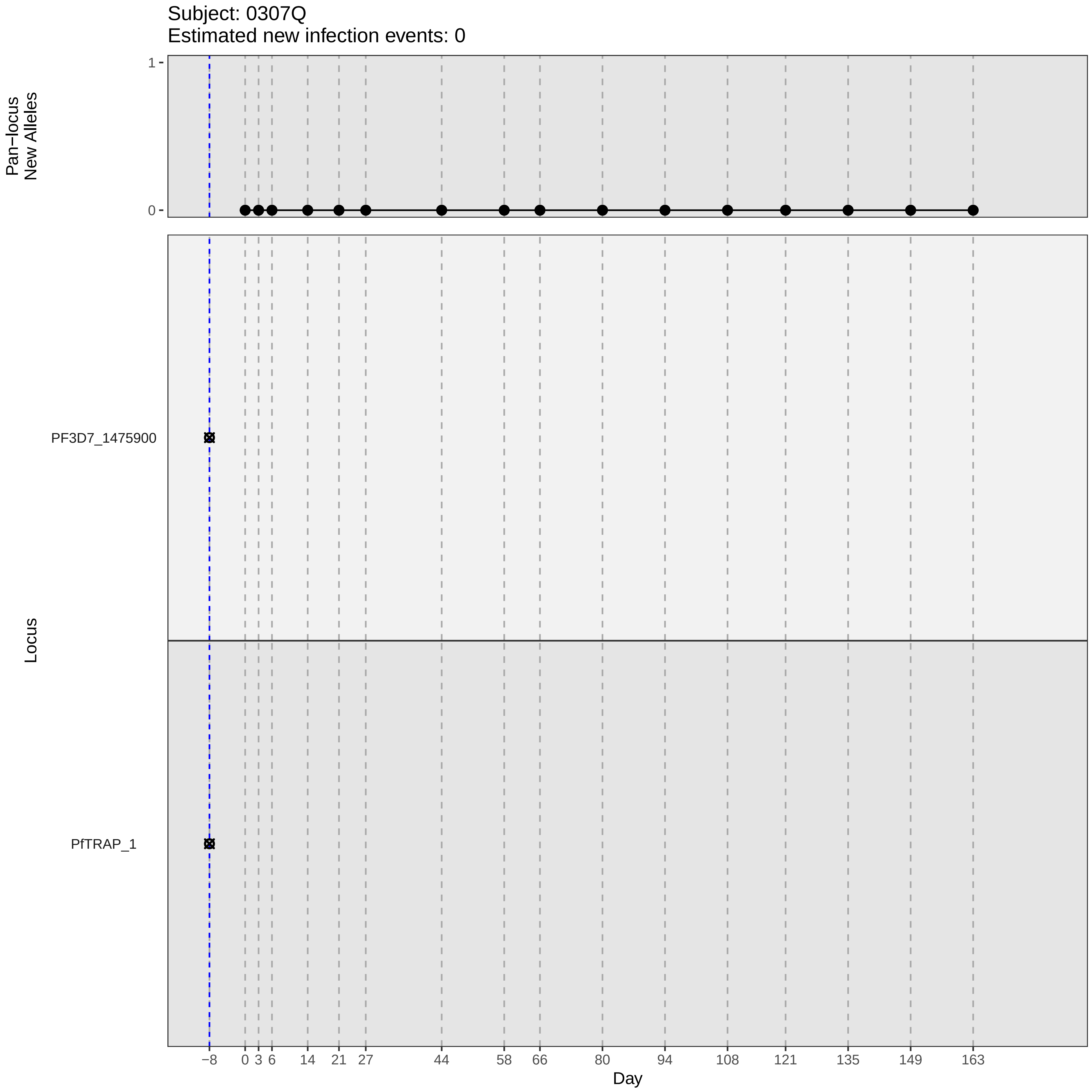

Subject: 0307Q
Estimated new infection events: 0
Pan−locus
New Alleles
1
0
Locus
PF3D7_1475900
PfTRAP_1
−8
0
3
6
14
21
27
44
58
66
80
94
108
121
135
149
163
Day

### participant_0311U_with_imputation.pdf

Subject: 0311U
Estimated new infection events: 3.1

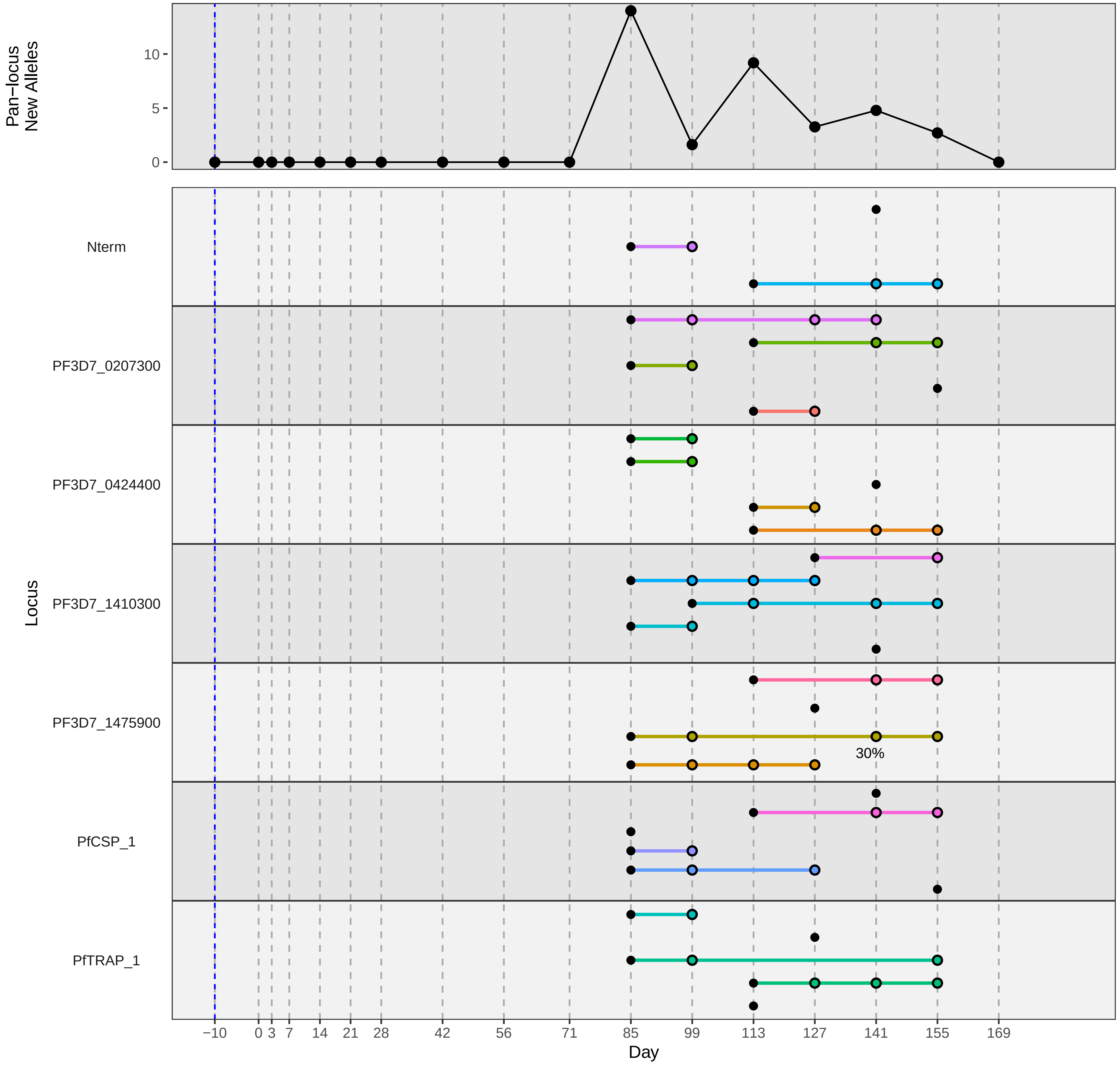

### participant_0314X_without_imputation.pdf

Subject: 0314X
Estimated new infection events: 1

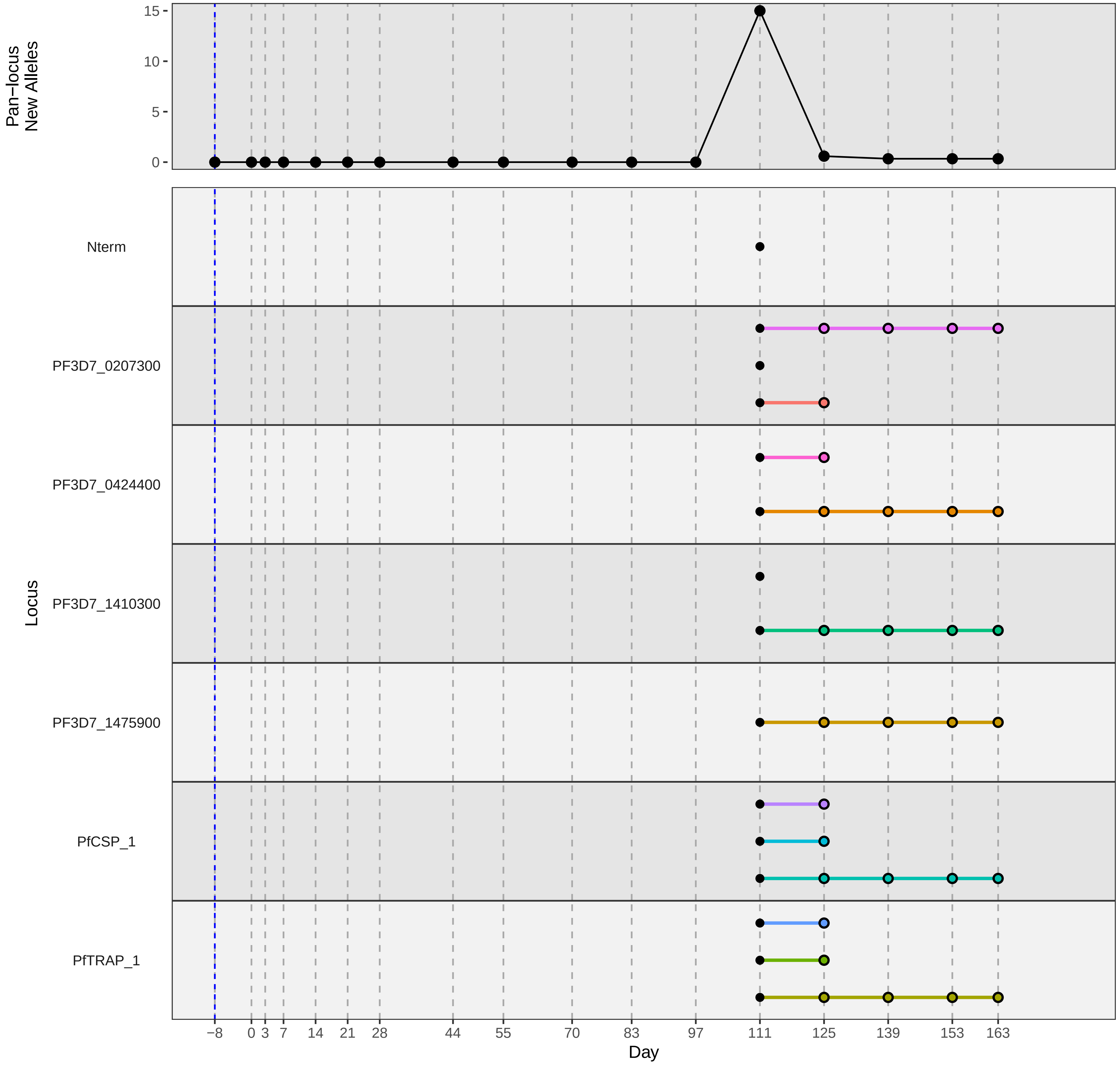

### participant_0339C_with_imputation.pdf

Subject: 0339C
Estimated new infection events: 3.1

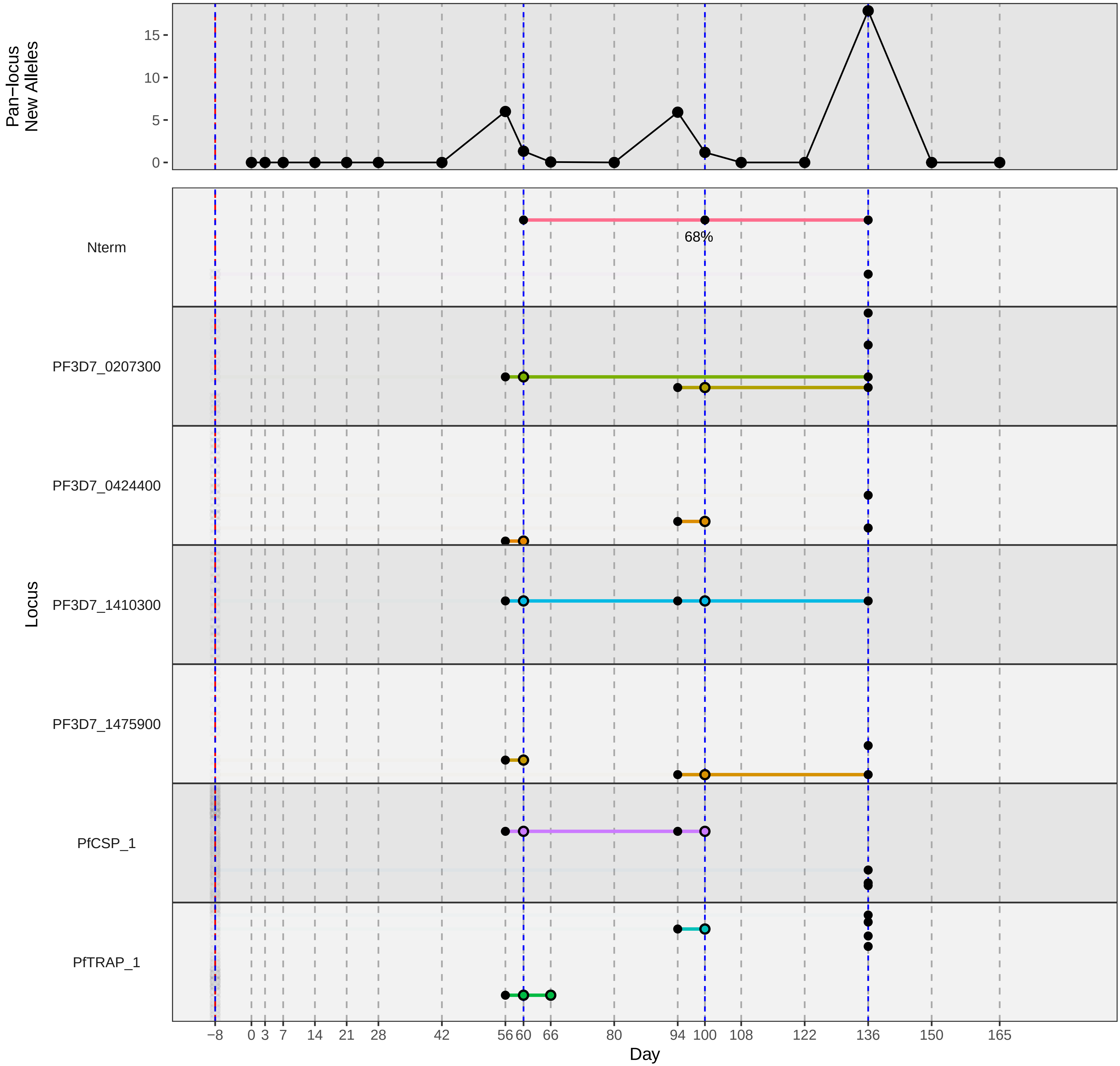

### participant_0341E_without_imputation.pdf

Subject: 0341E
Estimated new infection events: 4.1

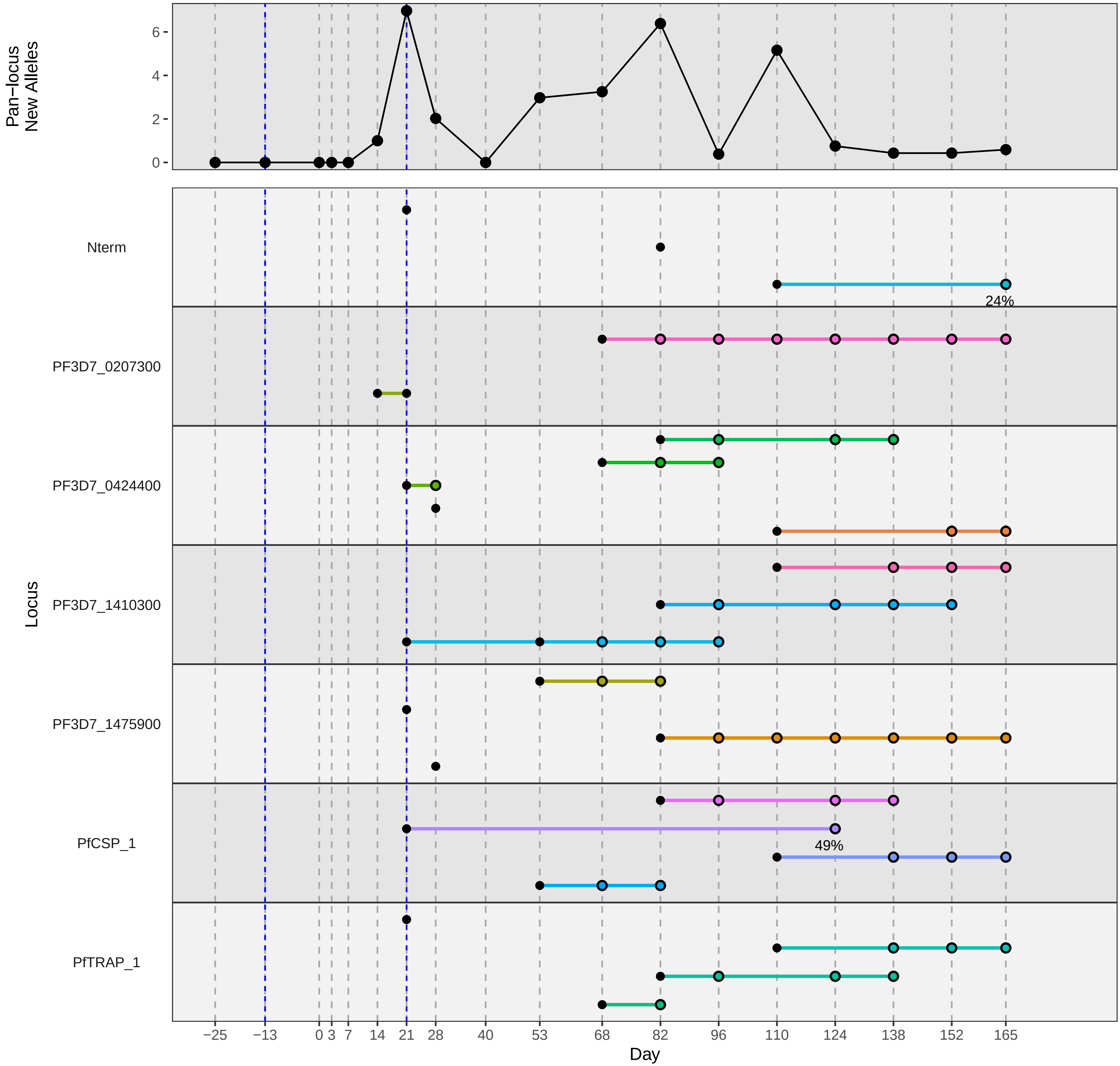

### participant_0353U_with_imputation.pdf

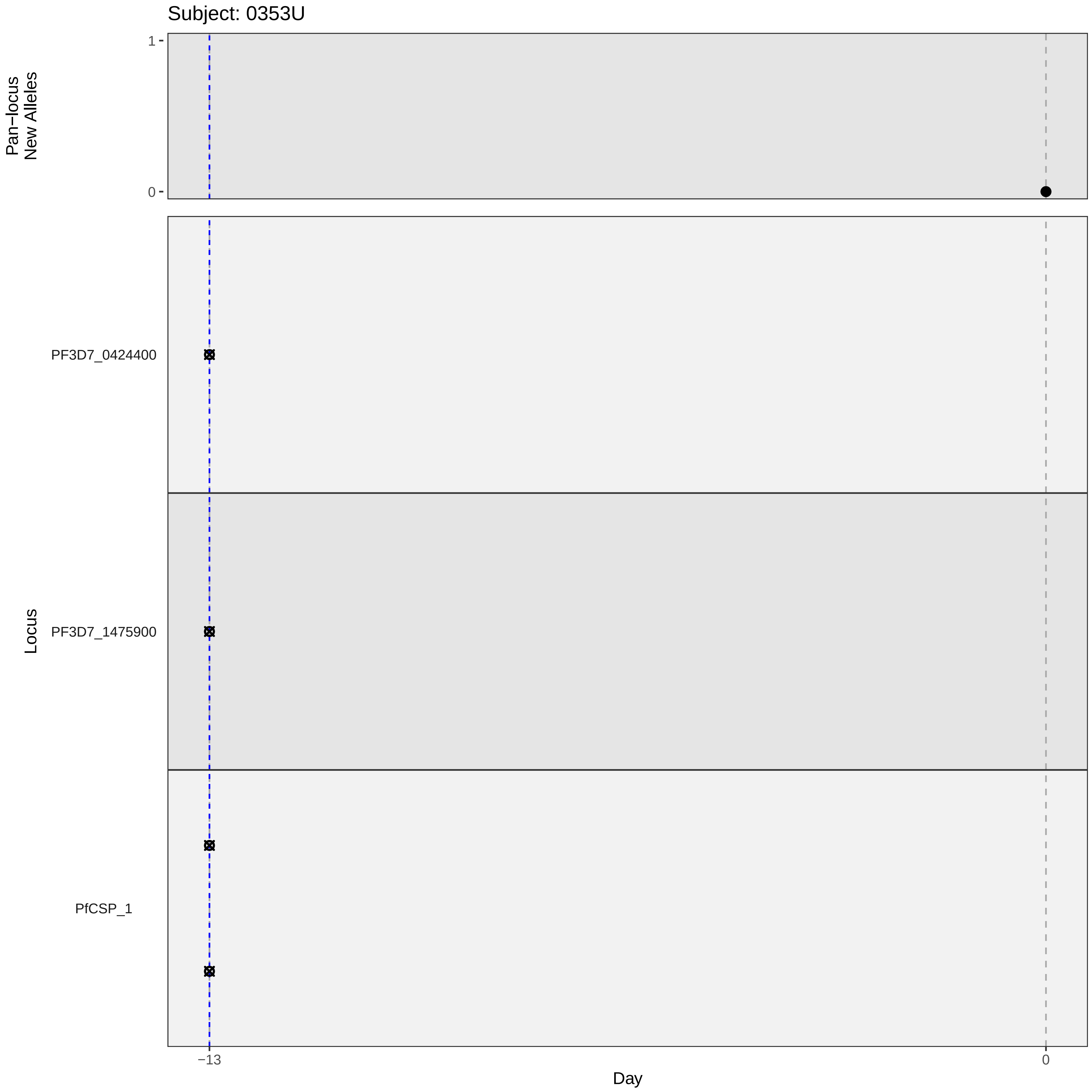

Subject: 0353U
Pan−locus
New Alleles
1
0
Locus
PF3D7_0424400
PF3D7_1475900
PfCSP_1
−13
0
Day

### participant_0353U_without_imputation.pdf

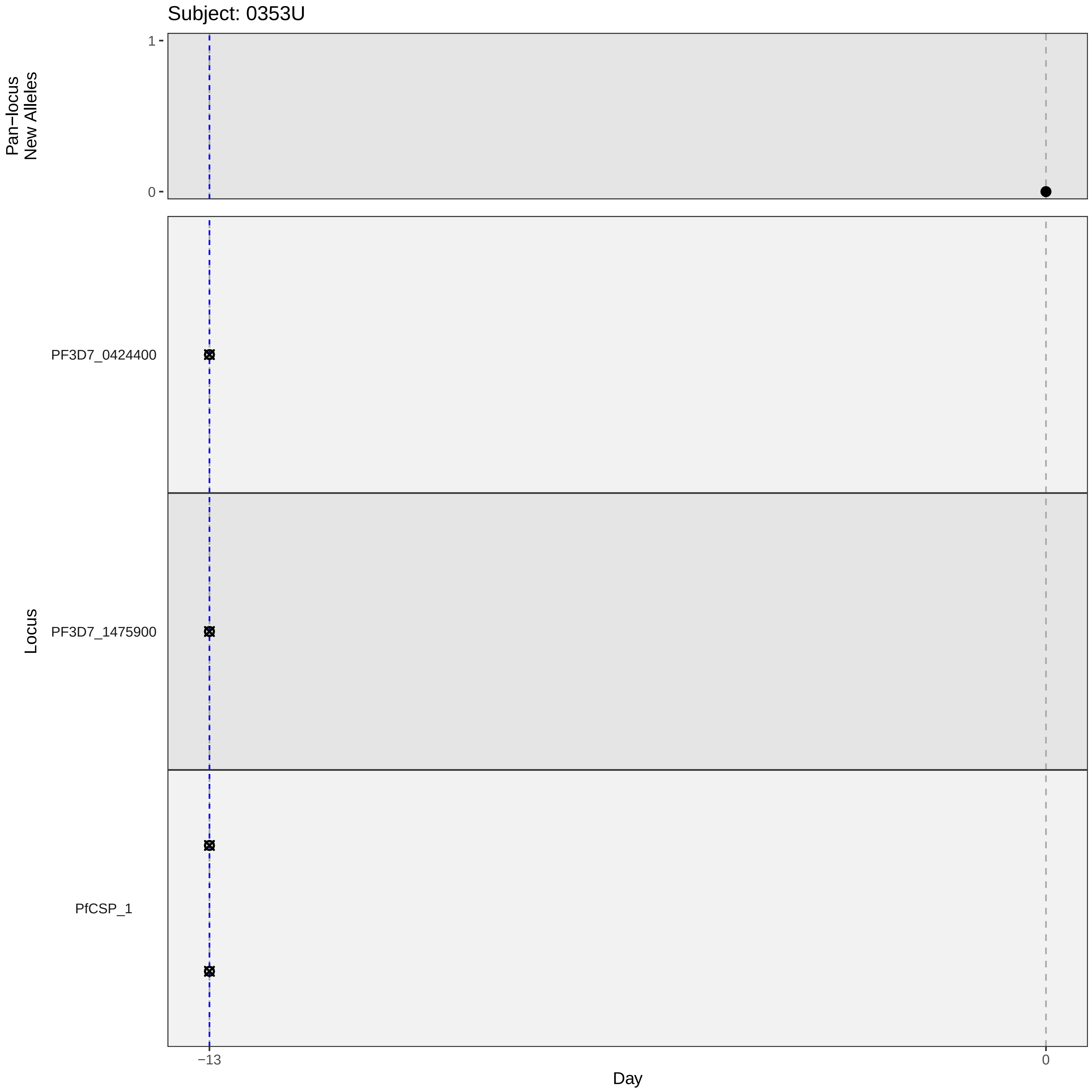

Subject: 0353U
Pan−locus
New Alleles
1
0
Locus
PF3D7_0424400
PF3D7_1475900
PfCSP_1
−13
0
Day

### participant_0374U_with_imputation.pdf

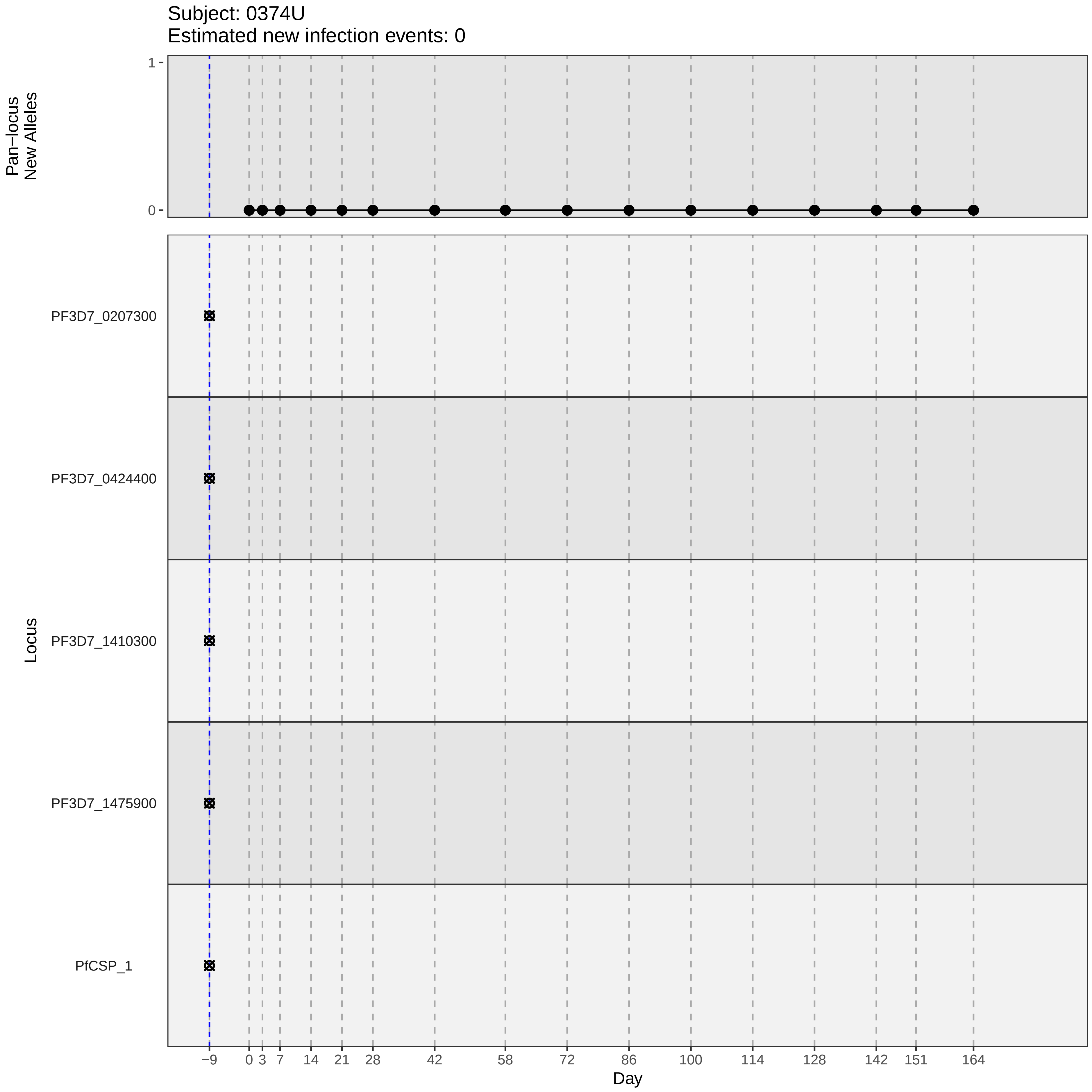

Subject: 0374U
Estimated new infection events: 0
Pan−locus New Alleles
1
0
Locus
PF3D7_0207300
PF3D7_0424400
PF3D7_1410300
PF3D7_1475900
PfCSP_1
−9
0
3
7
14
21
28
42
58
72
86
100
114
128
142
151
164
Day

### participant_0374U_without_imputation.pdf

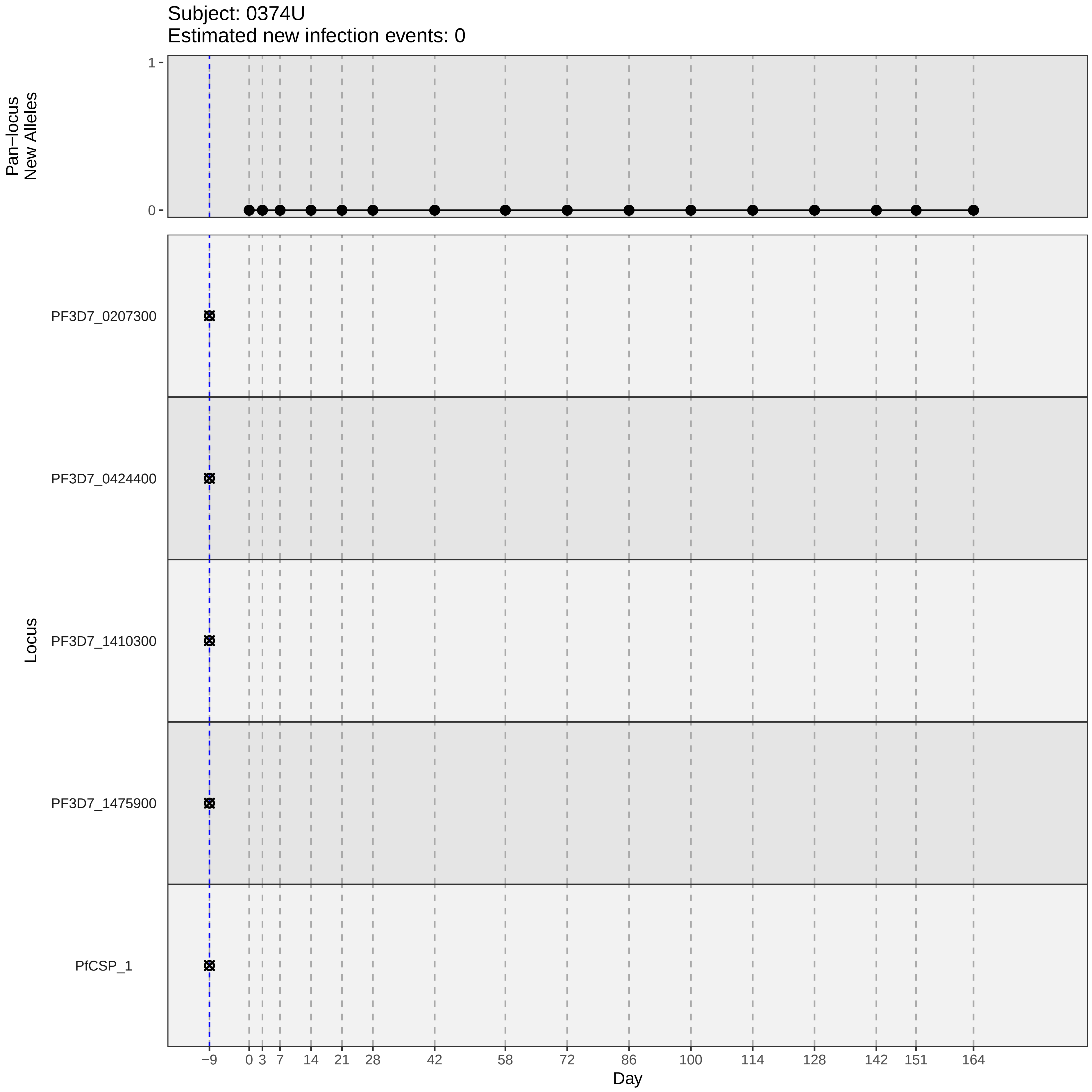

Subject: 0374U
Estimated new infection events: 0
Pan−locus
New Alleles
1
0
Locus
PF3D7_0207300
PF3D7_0424400
PF3D7_1410300
PF3D7_1475900
PfCSP_1
−9
0
3
7
14
21
28
42
58
72
86
100
114
128
142
151
164
Day

### participant_0386H_without_imputation.pdf

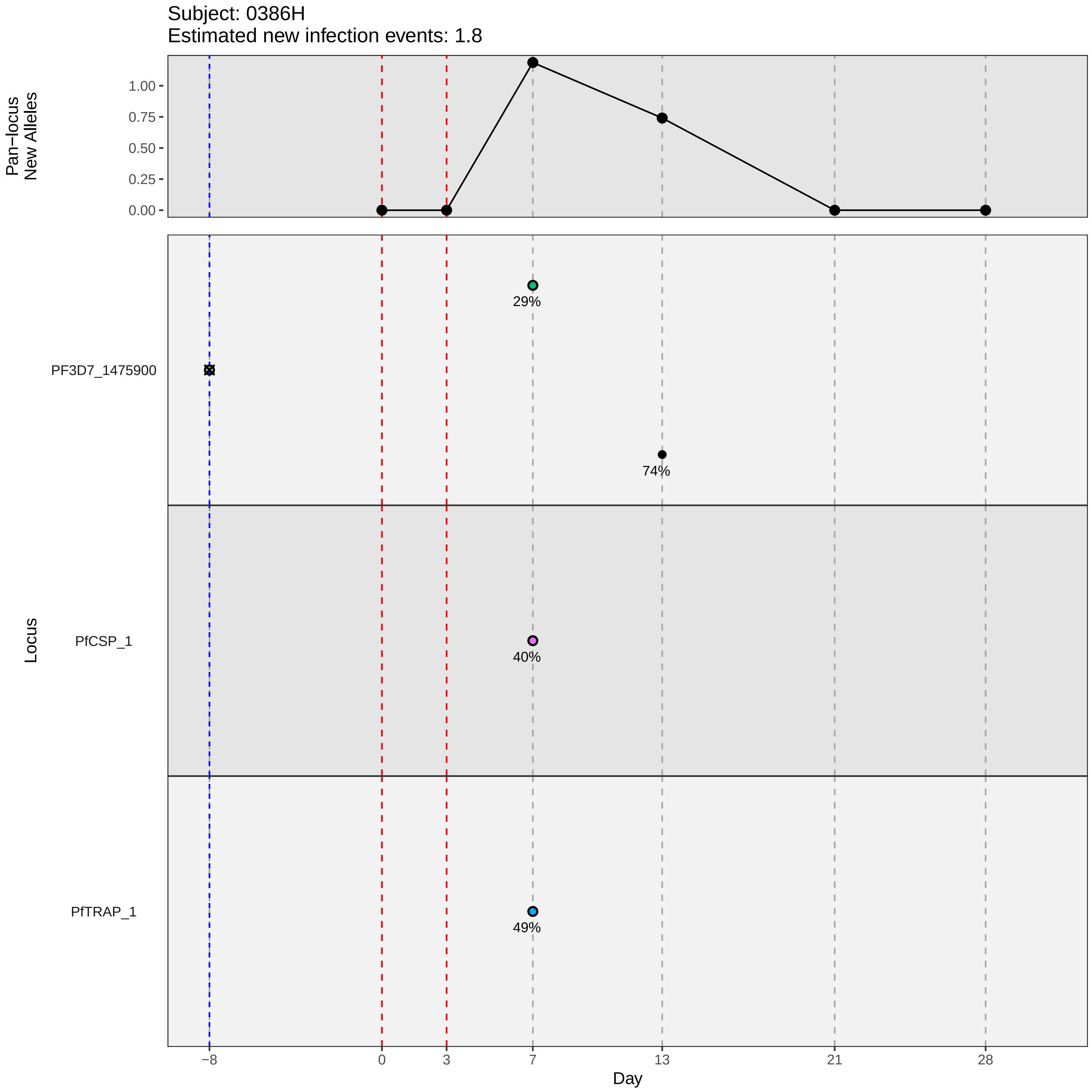

Subject: 0386H
Estimated new infection events: 1.8
Pan−locus New Alleles
1.00
0.75
0.50
0.25
0.00
Locus
PF3D7_1475900
PfCSP_1
PfTRAP_1
29%
74%
40%
49%
−8
0
3
7
13
21
28
Day

### participant_0388M_with_imputation.pdf

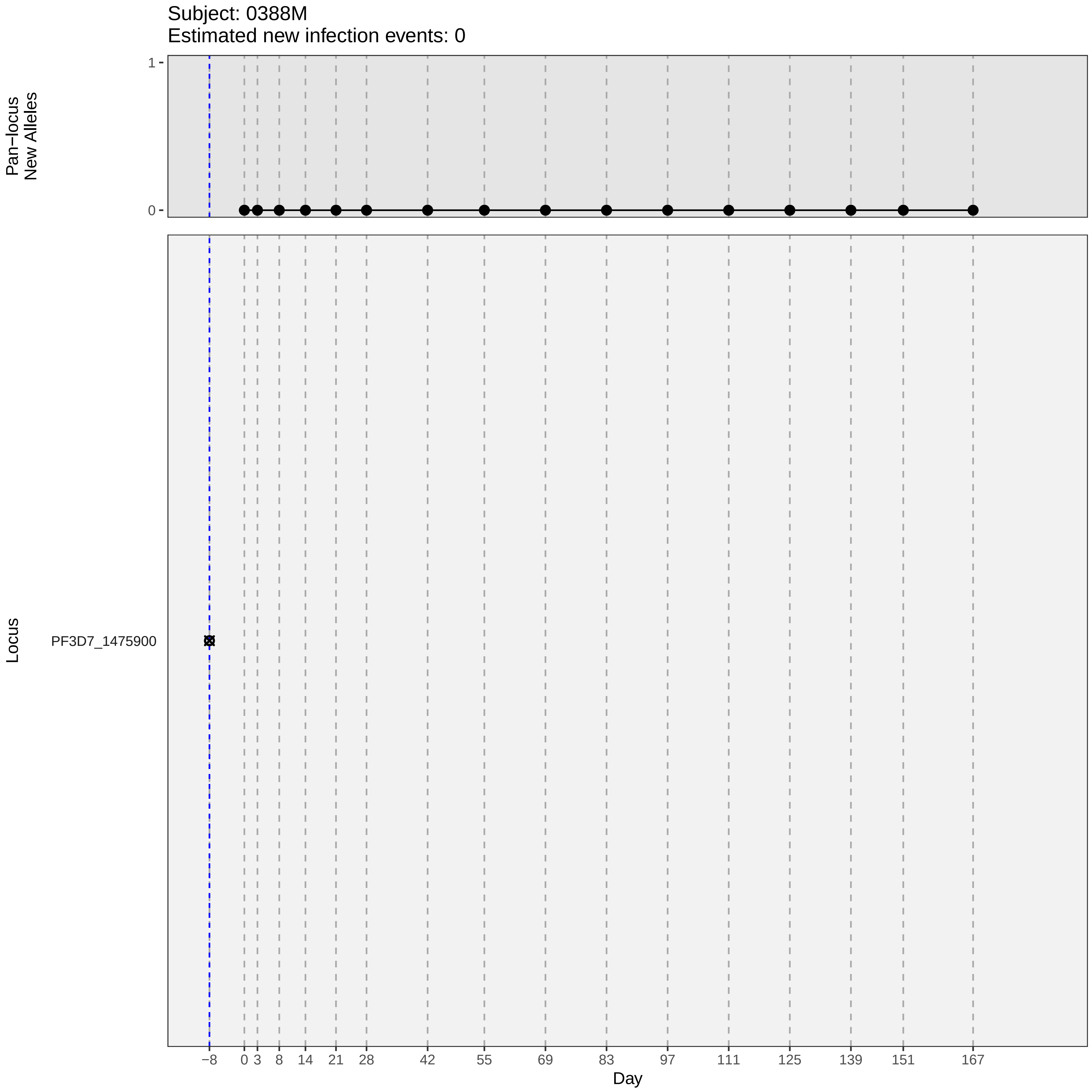
Subject: 0388M
Estimated new infection events: 0
Pan−locus
New Alleles
1
0
Locus
PF3D7_1475900
−8
0
3
8
14
21
28
42
55
69
83
97
111
125
139
151
167
Day

### participant_0388M_without_imputation.pdf

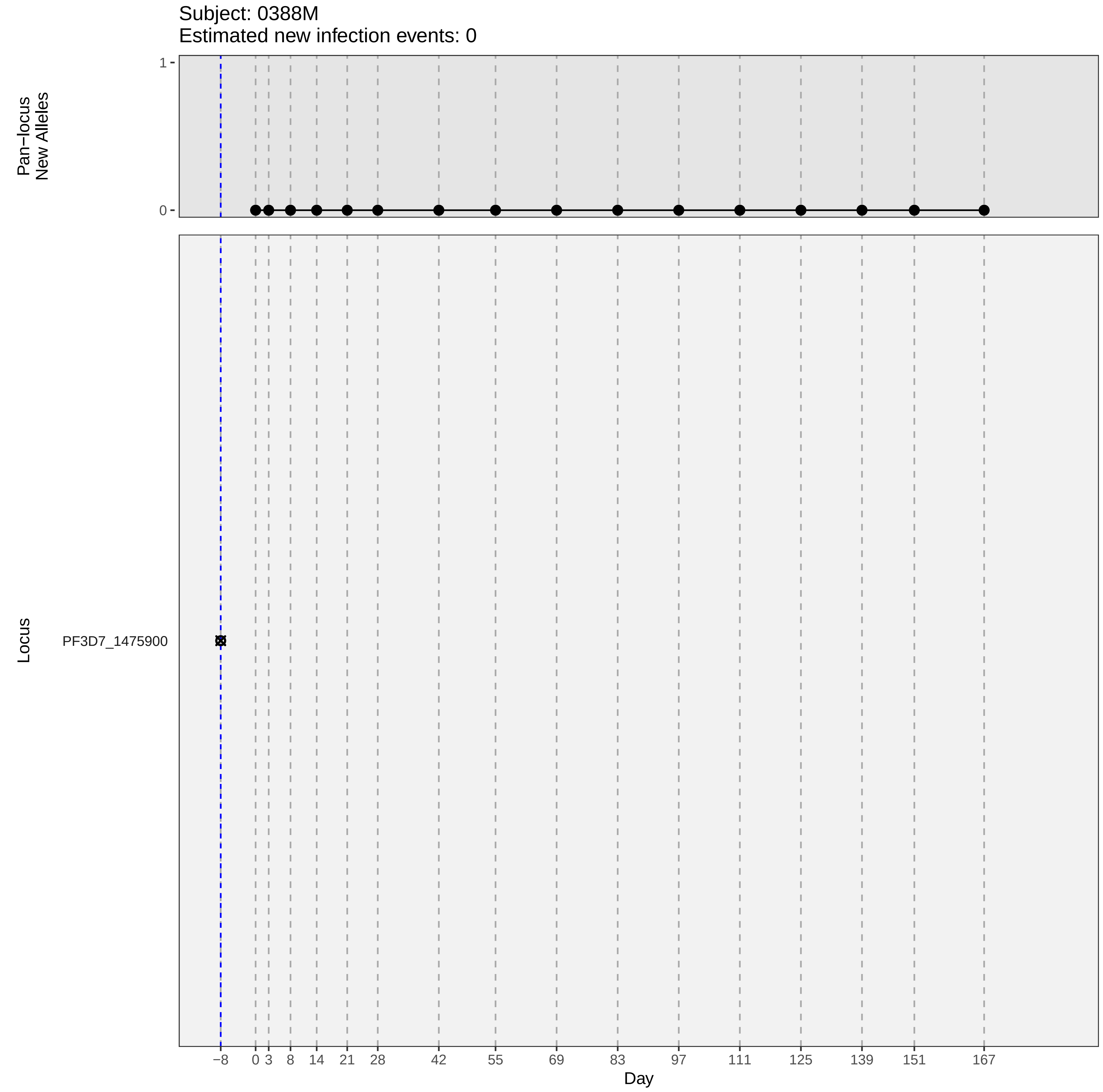

Subject: 0388M
Estimated new infection events: 0
Pan−locus
New Alleles
1
0
Locus
PF3D7_1475900
−8
0
3
8
14
21
28
42
55
69
83
97
111
125
139
151
167
Day

### participant_0405F_with_imputation.pdf

Subject: 0405F
Estimated new infection events: 0

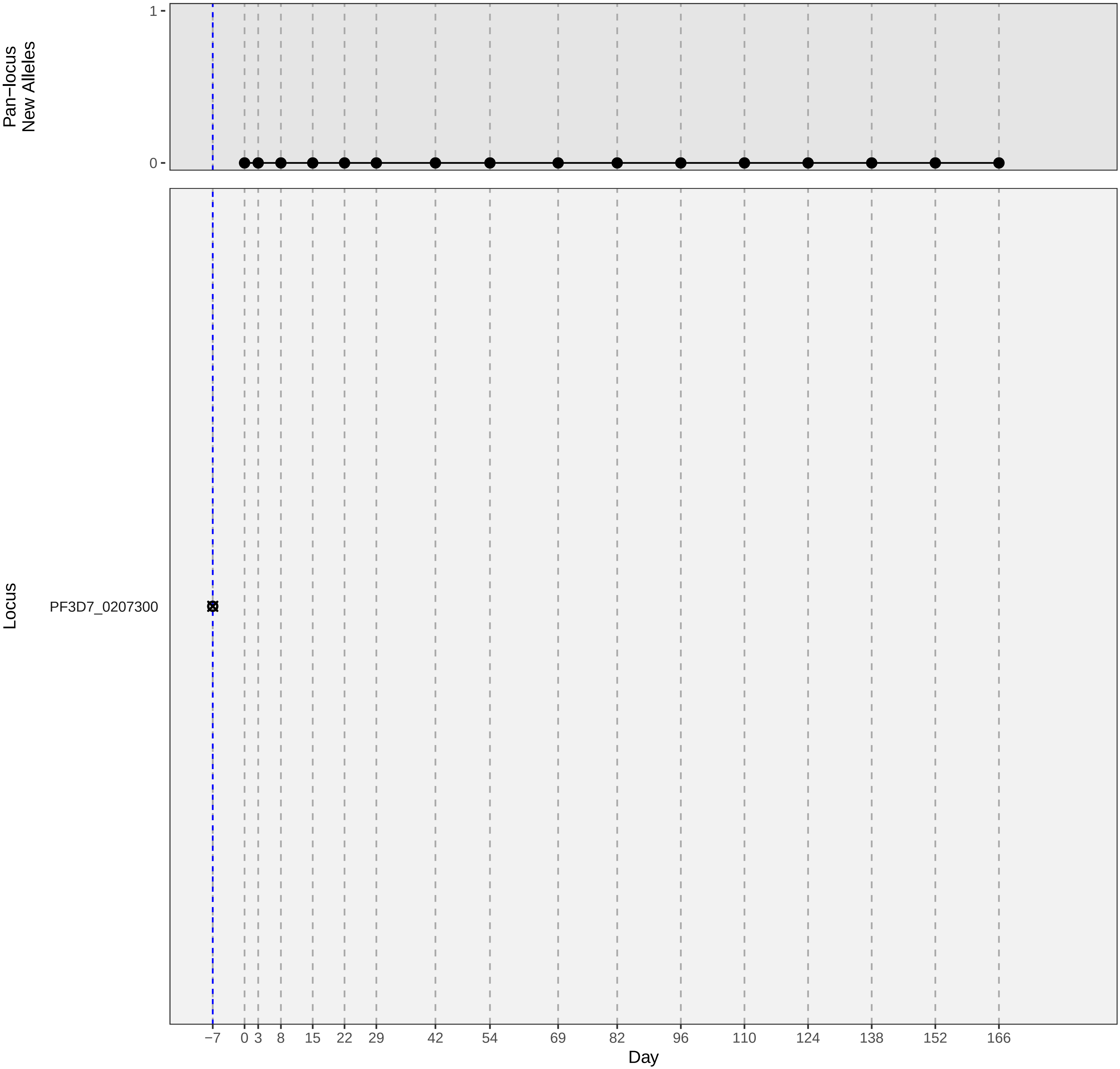

### participant_0405F_without_imputation.pdf

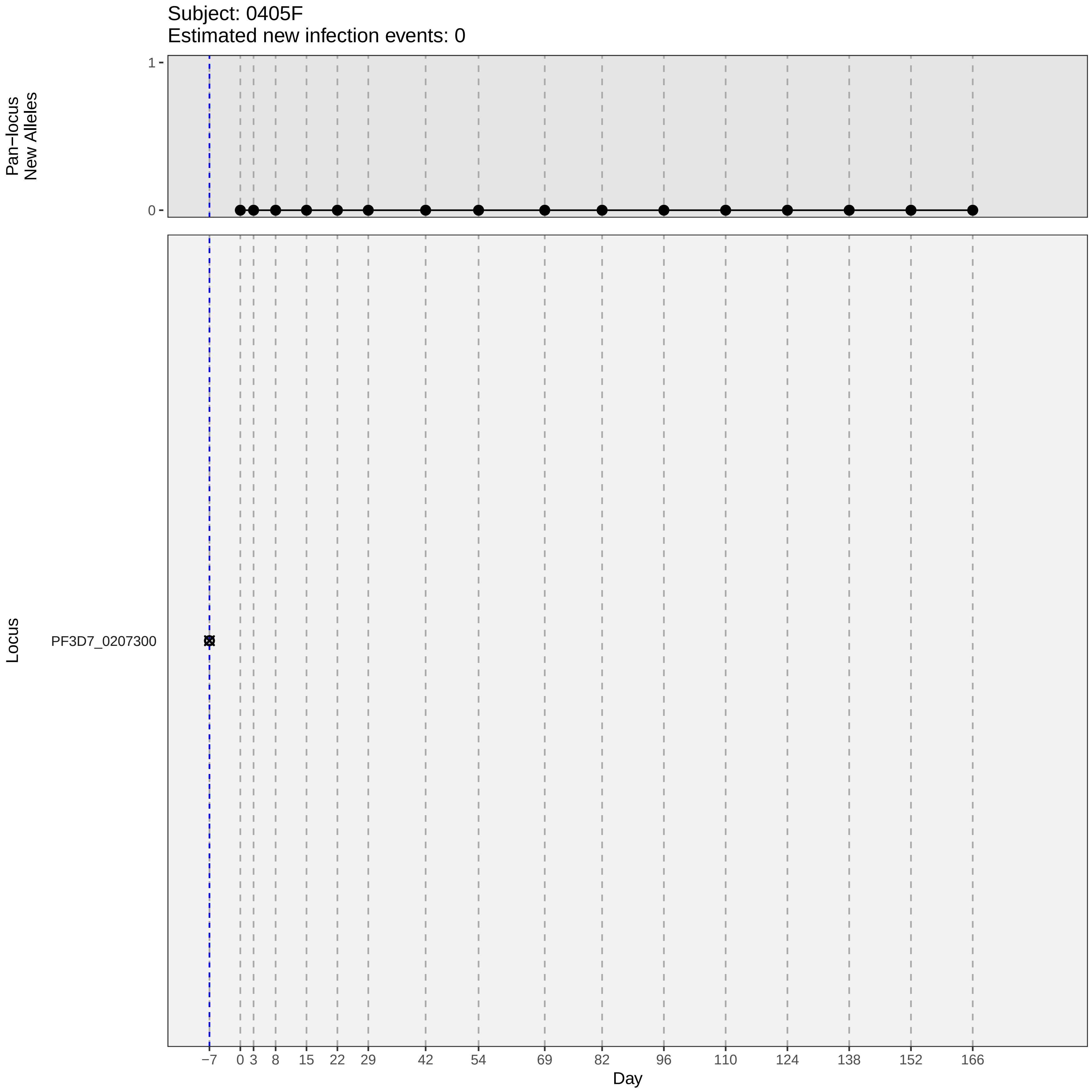
Subject: 0405F
Estimated new infection events: 0
Pan−locus
New Alleles
1
0
Locus
PF3D7_0207300
−7
0
3
8
15
22
29
42
54
69
82
96
110
124
138
152
166
Day

### participant_0423C_with_imputation.pdf

Subject: 0423C
Estimated new infection events: 4

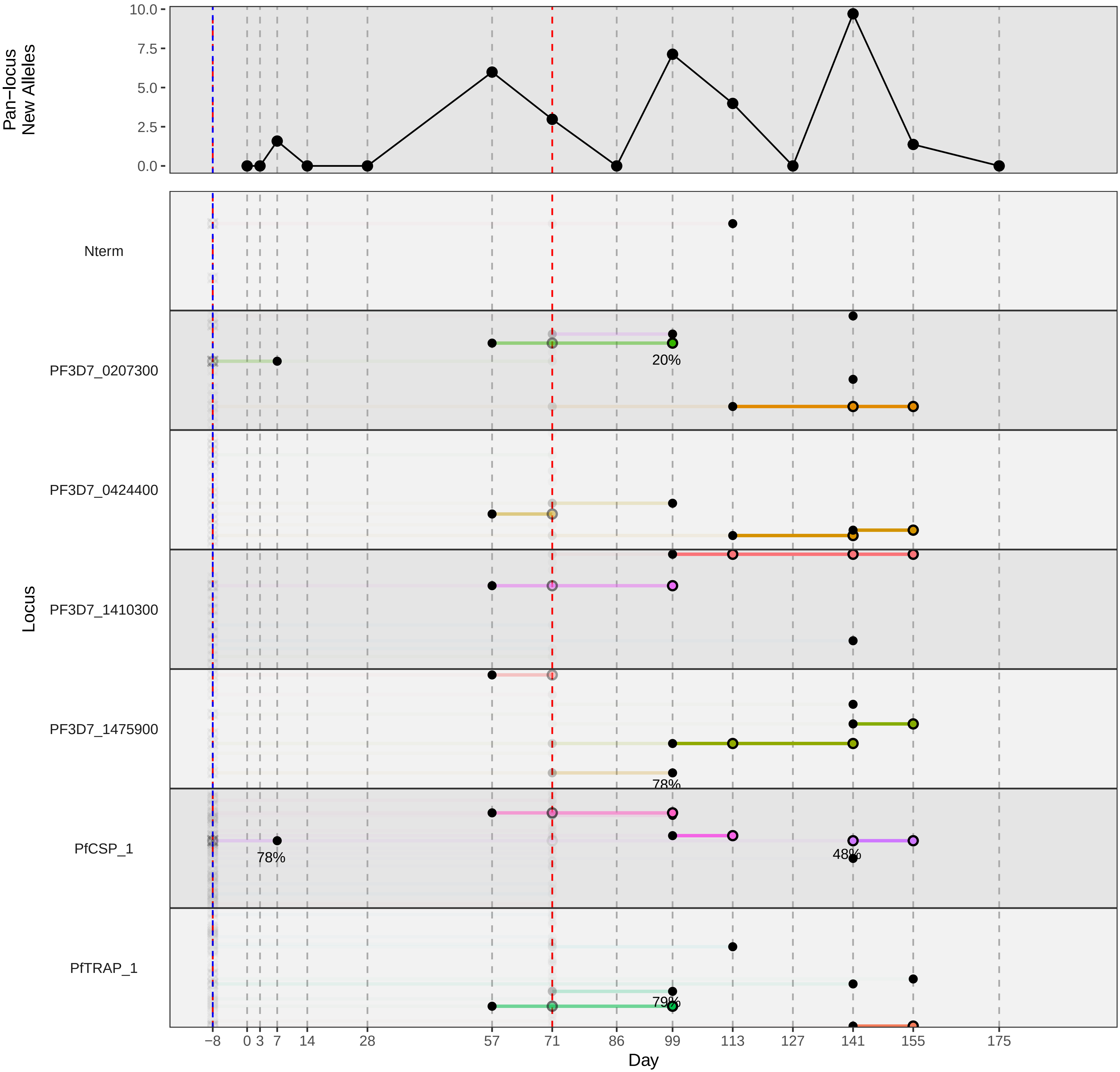

### participant_0423C_without_imputation.pdf

Subject: 0423C
Estimated new infection events: 4

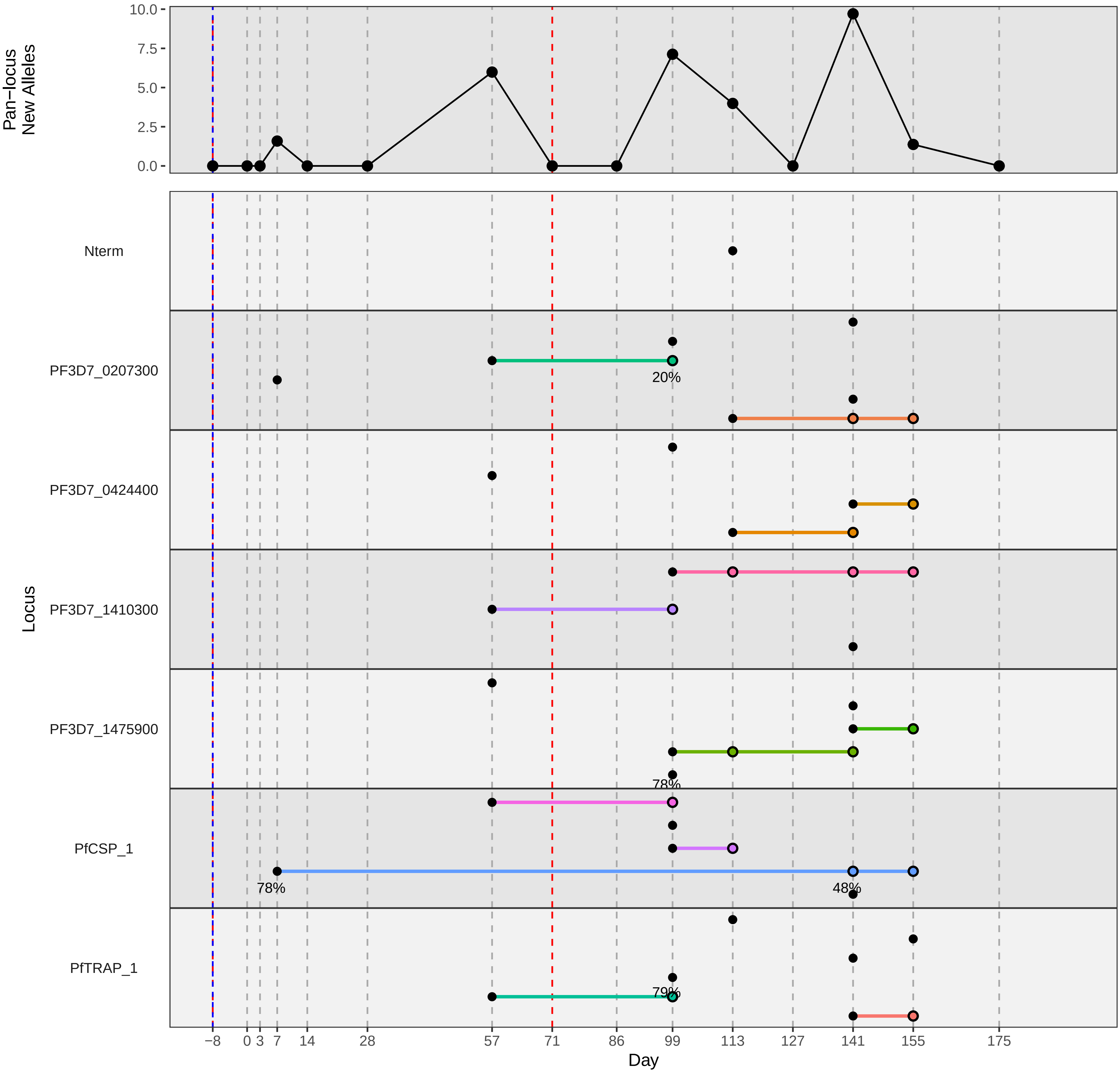
