## Supplementary material for "Genotypic infection endpoint analysis to understand efficacy and escape potential of the malaria monoclonal antibody CIS43LS": Data File S2: participant_0115M_without_imputation.pdf

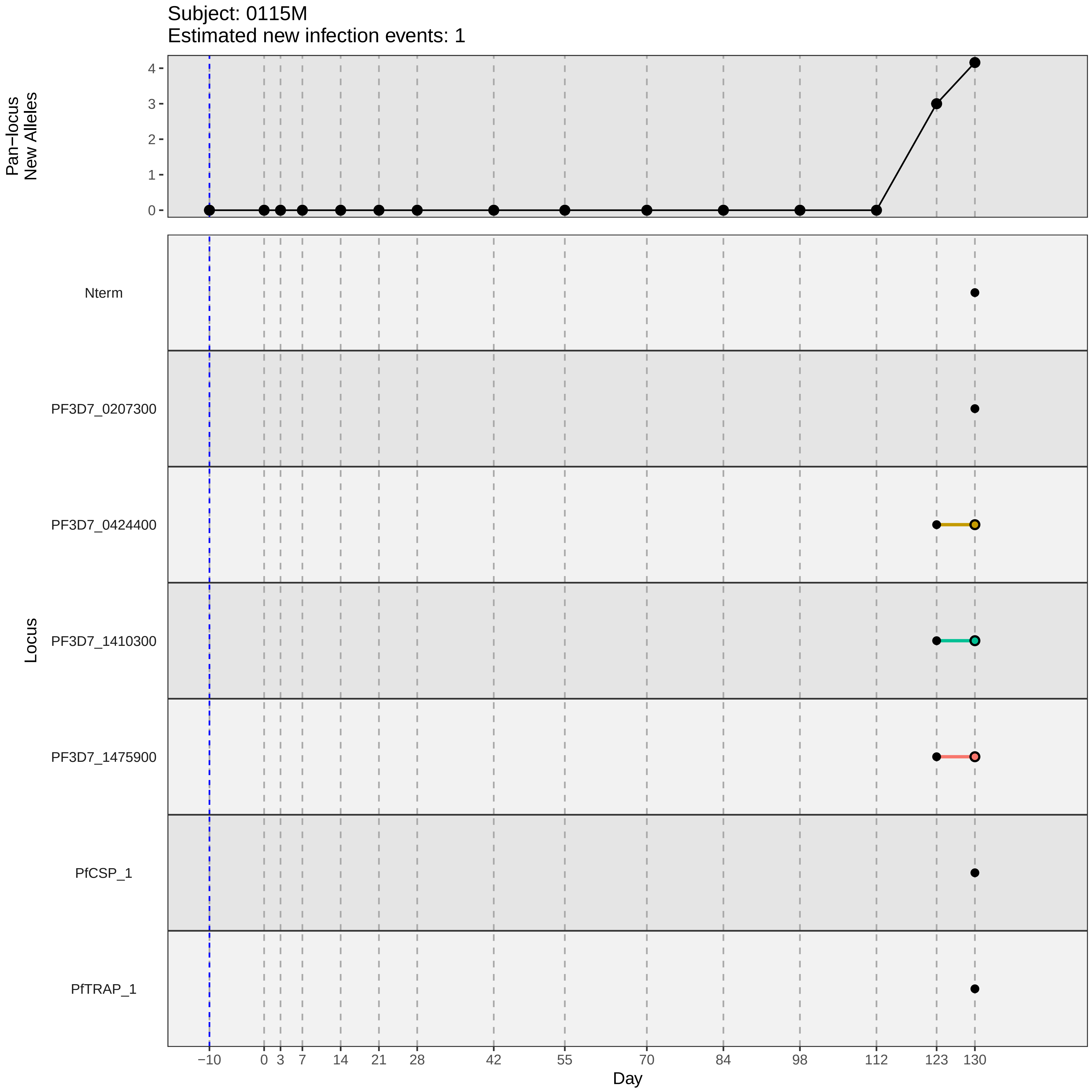

Subject: 0115M
Estimated new infection events: 1
Pan−locus
New Alleles
4
3
2
1
0
Nterm
PF3D7_0207300
PF3D7_0424400
PF3D7_1410300
PF3D7_1475900
PfCSP_1
PfTRAP_1
Locus
−10
0
3
7
14
21
28
42
55
70
84
98
112
123
130
Day
