## Supplementary material for "Genotypic infection endpoint analysis to understand efficacy and escape potential of the malaria monoclonal antibody CIS43LS": Data File S2: participant_0119R_without_imputation.pdf

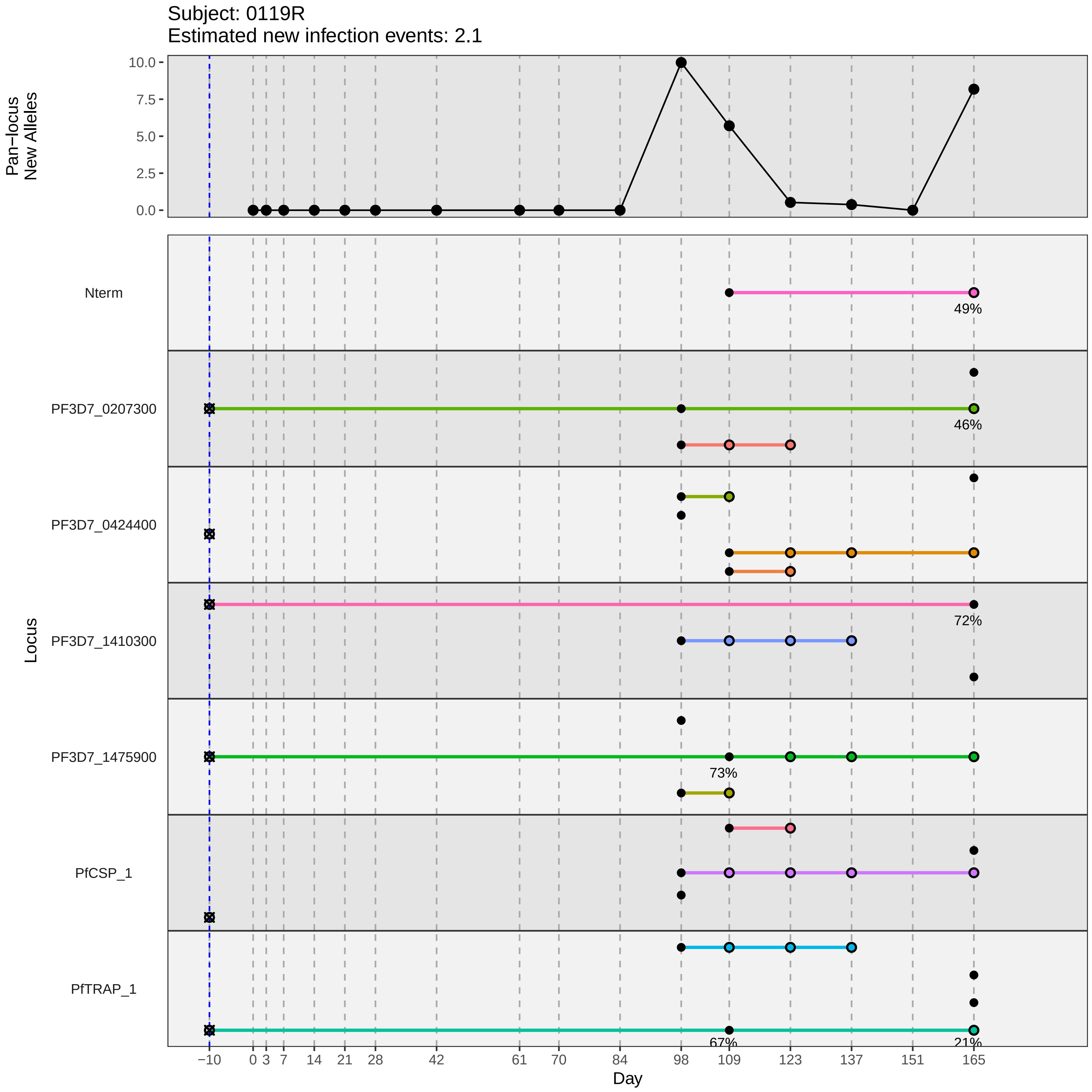

Subject: 0119R
Estimated new infection events: 2.1
Pan−locus New Alleles
10.0
7.5
5.0
2.5
0.0
Locus
Nterm
PF3D7_0207300
PF3D7_0424400
PF3D7_1410300
PF3D7_1475900
PfCSP_1
PfTRAP_1
49%
46%
72%
73%
67%
21%
−10
0
3
7
14
21
28
42
61
70
84
98
109
123
137
151
165
Day
