## Supplementary material for "Genotypic infection endpoint analysis to understand efficacy and escape potential of the malaria monoclonal antibody CIS43LS": Data File S2: participant_0125X_without_imputation.pdf

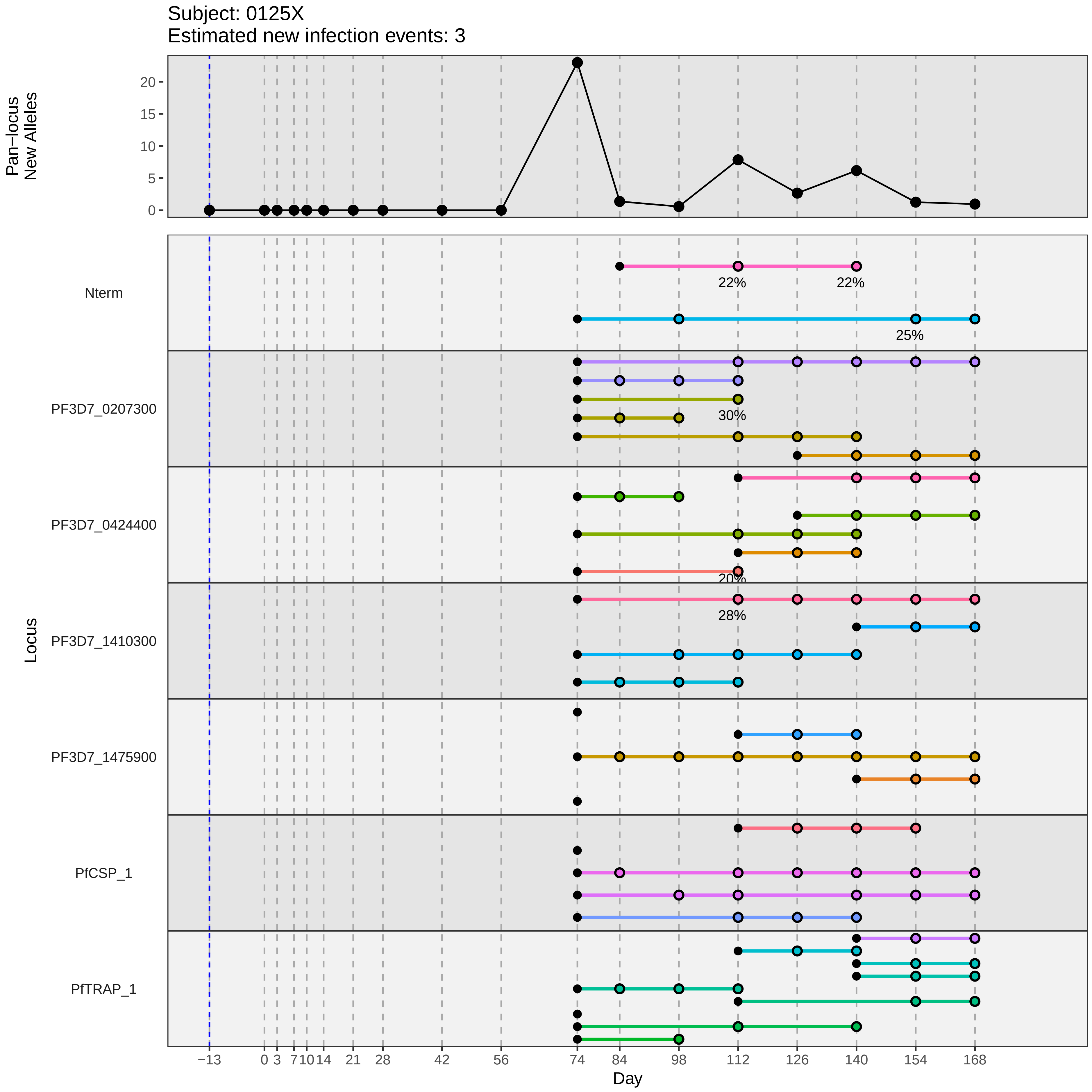
Subject: 0125X
Estimated new infection events: 3
Pan−locus New Alleles
0
5
10
15
20
Locus
Nterm
PF3D7_0207300
PF3D7_0424400
PF3D7_1410300
PF3D7_1475900
PfCSP_1
PfTRAP_1
22%
22%
25%
30%
20%
28%
−13
0
3
7
10
14
21
28
42
56
74
84
98
112
126
140
154
168
Day
