## Supplementary material for "Genotypic infection endpoint analysis to understand efficacy and escape potential of the malaria monoclonal antibody CIS43LS": Data File S2: participant_0130D_without_imputation.pdf

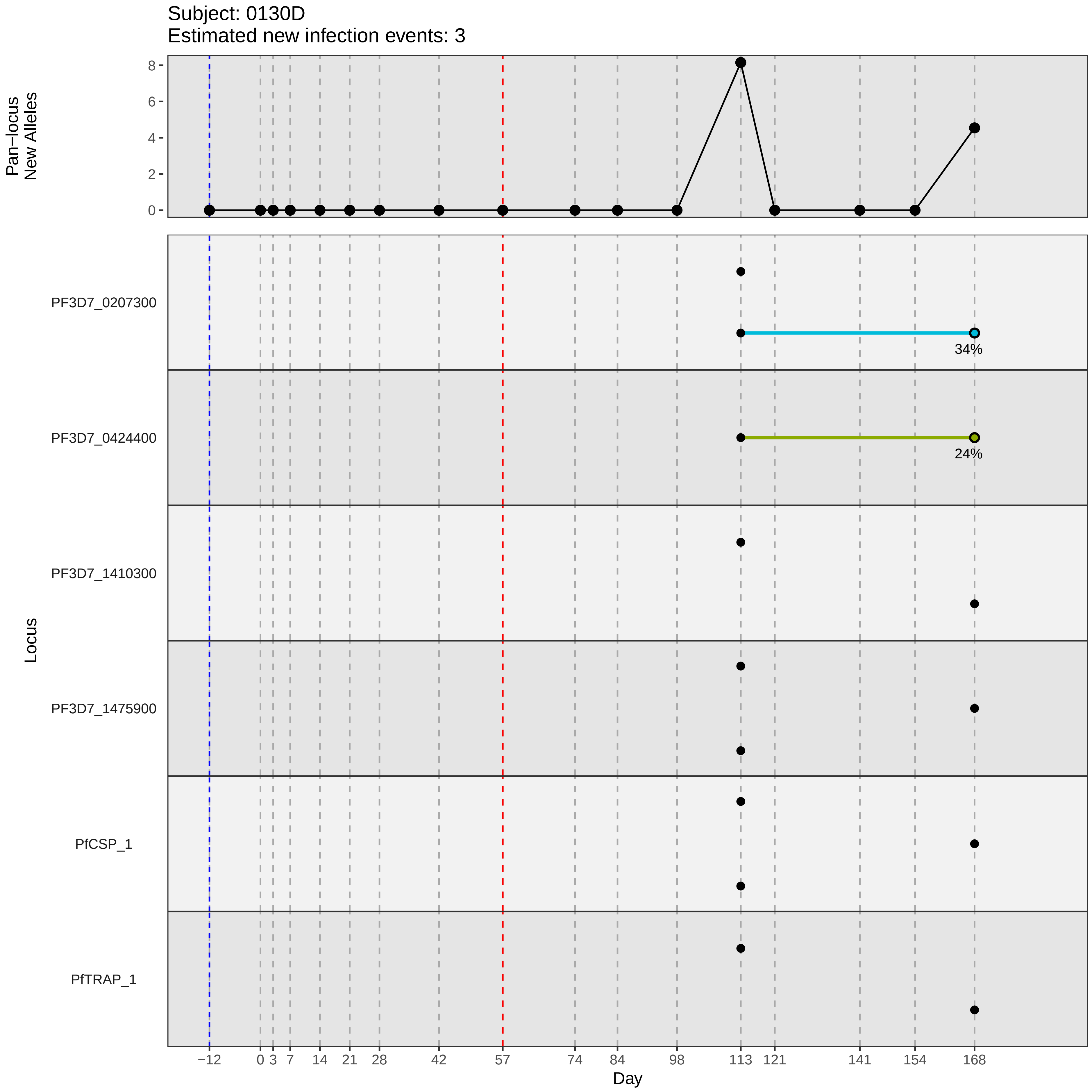

Subject: 0130D
Estimated new infection events: 3
Pan−locus New Alleles
8
6
4
2
0
Locus
PF3D7_0207300
34%
PF3D7_0424400
24%
PF3D7_1410300
PF3D7_1475900
PfCSP_1
PfTRAP_1
−12
0
3
7
14
21
28
42
57
74
84
98
113
121
141
154
168
Day
