## Supplementary material for "Genotypic infection endpoint analysis to understand efficacy and escape potential of the malaria monoclonal antibody CIS43LS": Data File S2: participant_0131E_without_imputation.pdf

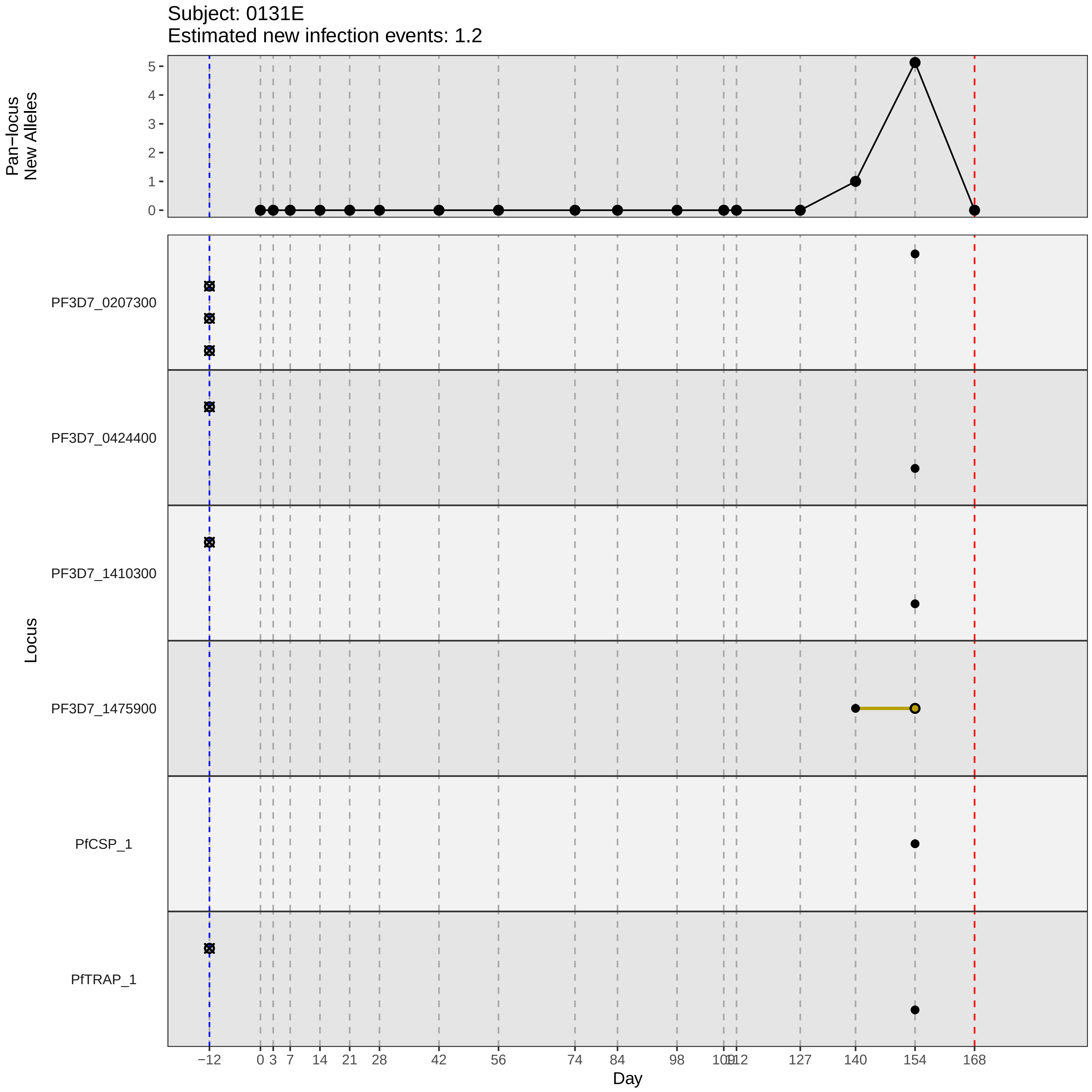
Subject: 0131E
Estimated new infection events: 1.2
Pan−locus New Alleles
Locus
PF3D7_0207300
PF3D7_0424400
PF3D7_1410300
PF3D7_1475900
PfCSP_1
PfTRAP_1
Day
−12
0
3
7
14
21
28
42
56
74
84
98
127
140
154
168
