## Supplementary material for "Genotypic infection endpoint analysis to understand efficacy and escape potential of the malaria monoclonal antibody CIS43LS": Data File S2: participant_0134H_without_imputation.pdf

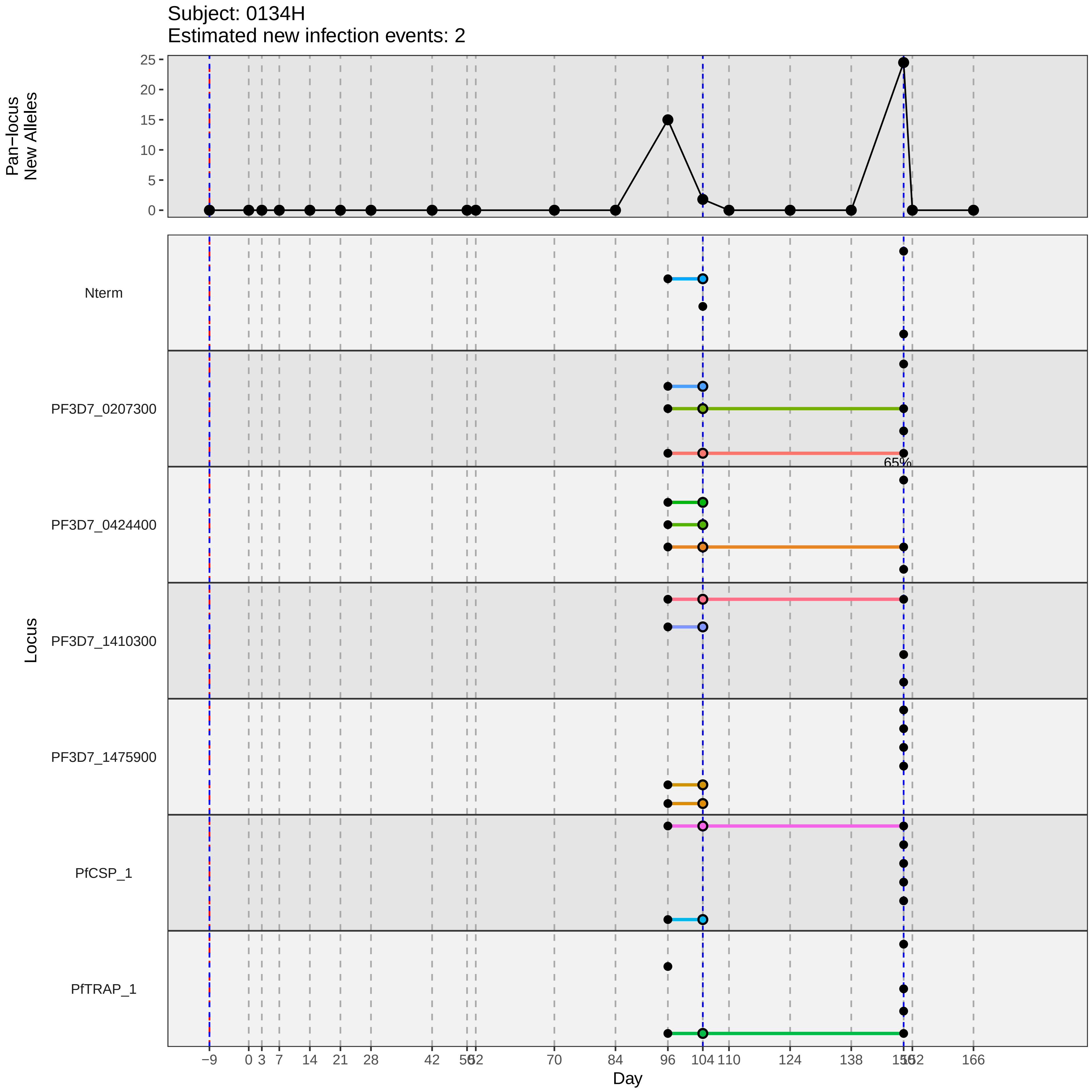

Subject: 0134H
Estimated new infection events: 2
Pan−locus New Alleles
25
20
15
10
5
0
Locus
Nterm
PF3D7_0207300
65%
PF3D7_0424400
PF3D7_1410300
PF3D7_1475900
PfCSP_1
PfTRAP_1
−9
0
3
7
14
21
28
42
56
62
70
84
96
104
110
124
138
156
162
166
Day
