## Supplementary material for "Genotypic infection endpoint analysis to understand efficacy and escape potential of the malaria monoclonal antibody CIS43LS": Data File S2: participant_0142T_without_imputation.pdf

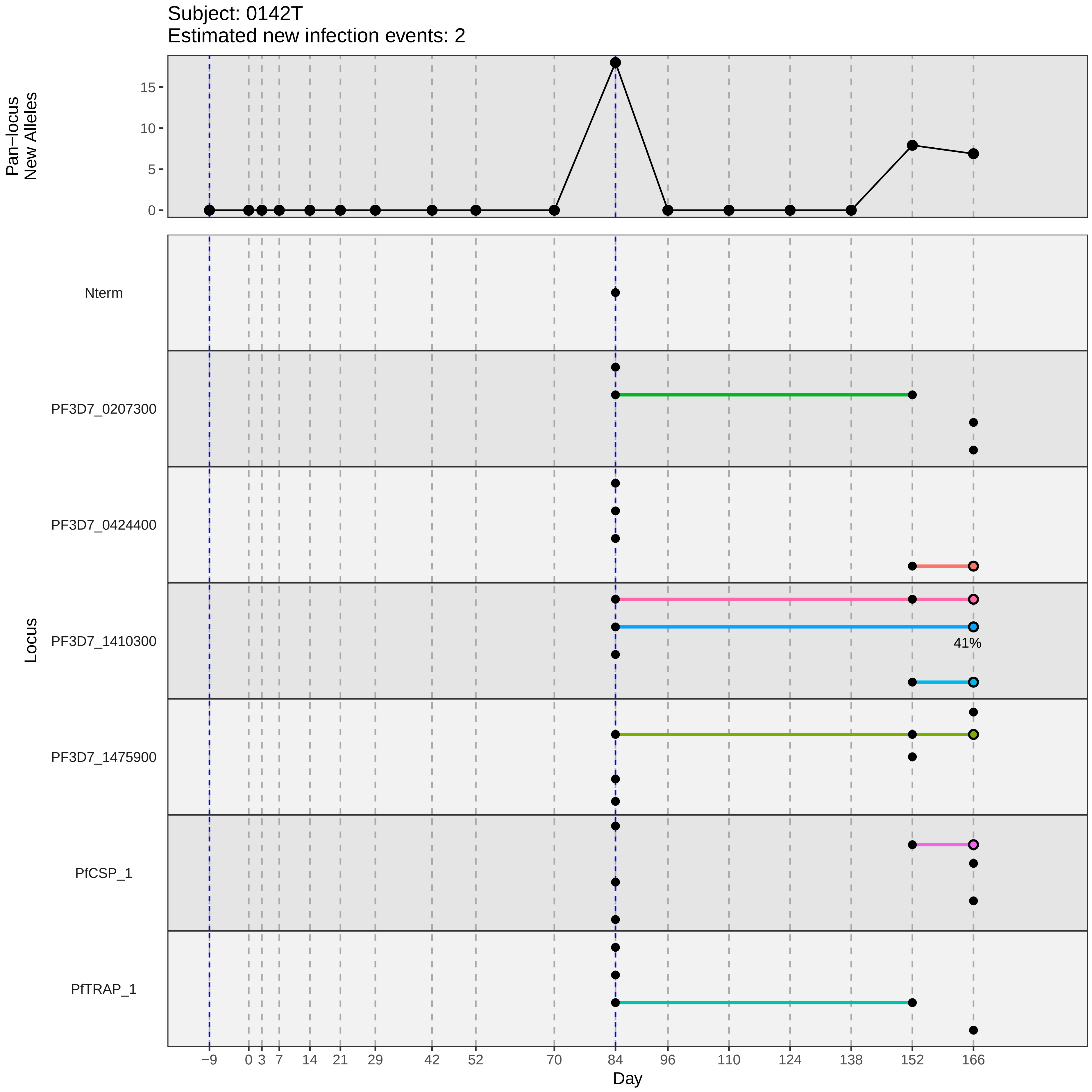
Subject: 0142T
Estimated new infection events: 2
Pan−locus
New Alleles
0
5
10
15
Locus
Nterm
PF3D7_0207300
PF3D7_0424400
PF3D7_1410300
41%
PF3D7_1475900
PfCSP_1
PfTRAP_1
−9
0
3
7
14
21
29
42
52
70
84
96
110
124
138
152
166
Day
