## Supplementary material for "Genotypic infection endpoint analysis to understand efficacy and escape potential of the malaria monoclonal antibody CIS43LS": Data File S2: participant_0145W_without_imputation.pdf

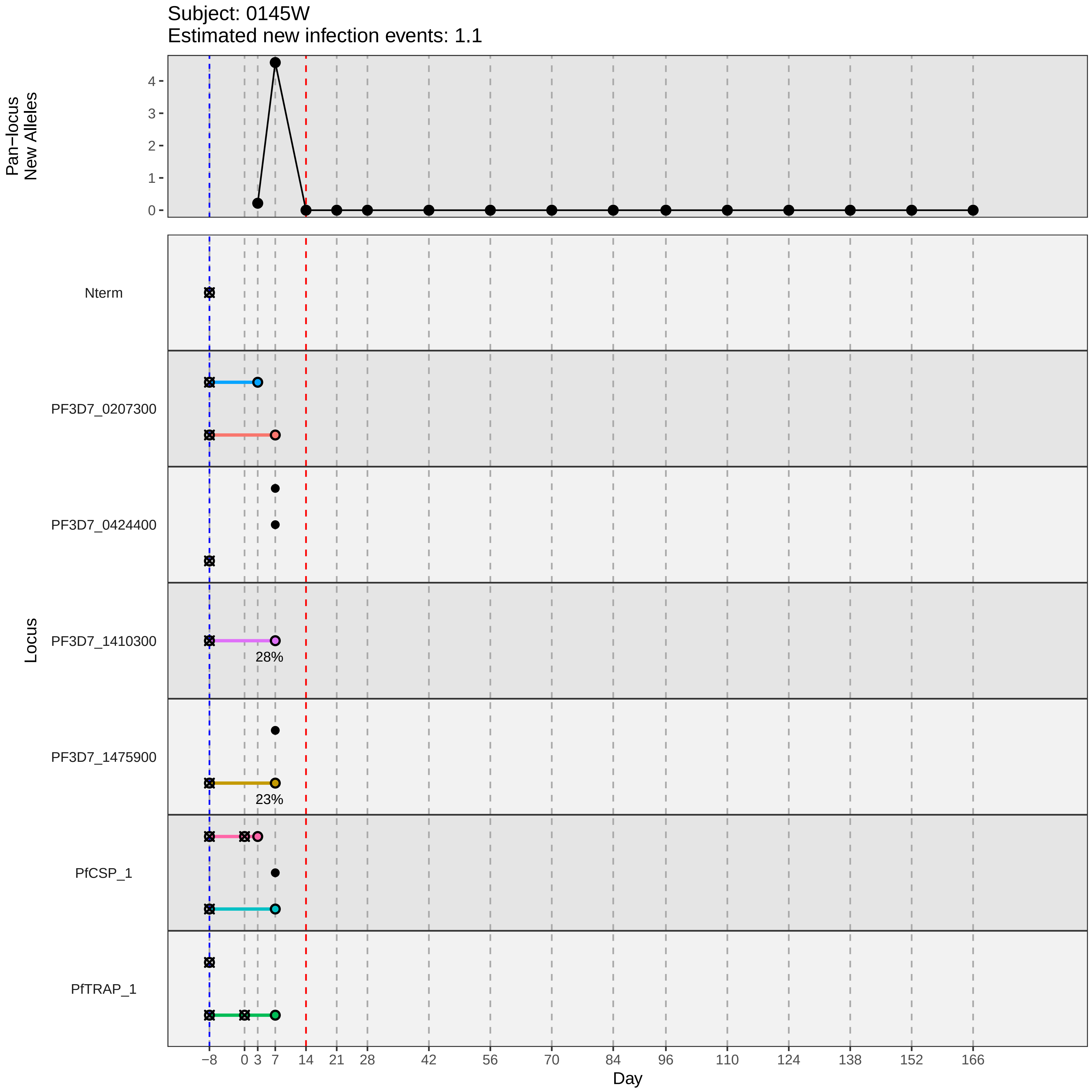

Subject: 0145W
Estimated new infection events: 1.1
Pan−locus New Alleles
0
1
2
3
4
Locus
Nterm
PF3D7_0207300
PF3D7_0424400
PF3D7_1410300
28%
PF3D7_1475900
23%
PfCSP_1
PfTRAP_1
−8 0 3 7 14 21 28 42 56 70 84 96 110 124 138 152 166
Day
