## Supplementary material for "Genotypic infection endpoint analysis to understand efficacy and escape potential of the malaria monoclonal antibody CIS43LS": Data File S2: participant_0148A_without_imputation.pdf

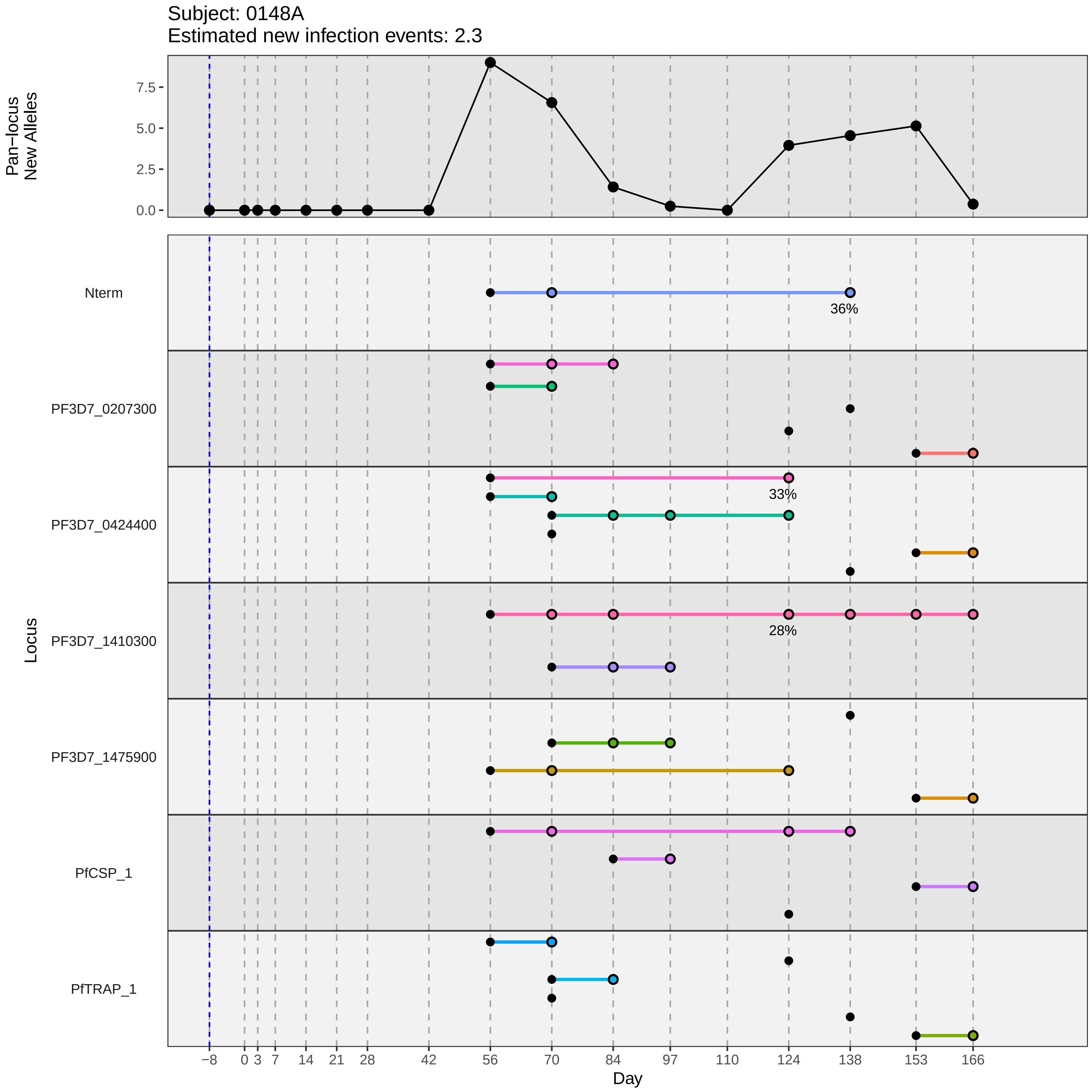

Subject: 0148A
Estimated new infection events: 2.3
Pan−locus New Alleles
0.0
2.5
5.0
7.5
Locus
Nterm
PF3D7_0207300
PF3D7_0424400
PF3D7_1410300
PF3D7_1475900
PfCSP_1
PfTRAP_1
36%
33%
28%
−8
0
3
7
14
21
28
42
56
70
84
97
110
124
138
153
166
Day
