## Supplementary material for "Genotypic infection endpoint analysis to understand efficacy and escape potential of the malaria monoclonal antibody CIS43LS": Data File S2: participant_0150C_without_imputation.pdf

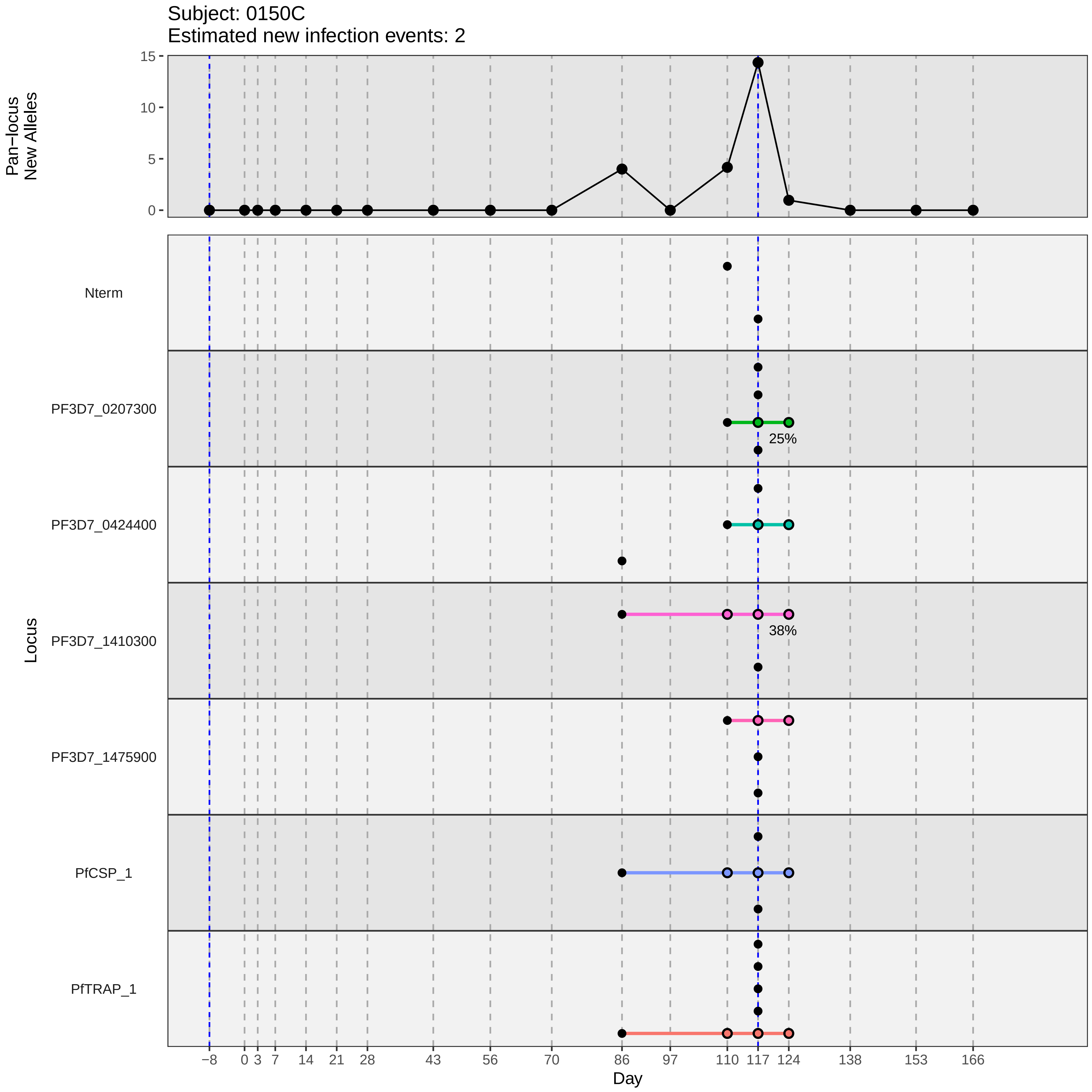
Subject: 0150C
Estimated new infection events: 2
Pan−locus
New Alleles
15
10
5
0
Locus
Nterm
PF3D7_0207300
25%
PF3D7_0424400
PF3D7_1410300
38%
PF3D7_1475900
PfCSP_1
PfTRAP_1
−8
0
3
7
14
21
28
43
56
70
86
97
110
117
124
138
153
166
Day
