## Supplementary material for "Genotypic infection endpoint analysis to understand efficacy and escape potential of the malaria monoclonal antibody CIS43LS": Data File S2: participant_0154G_without_imputation.pdf

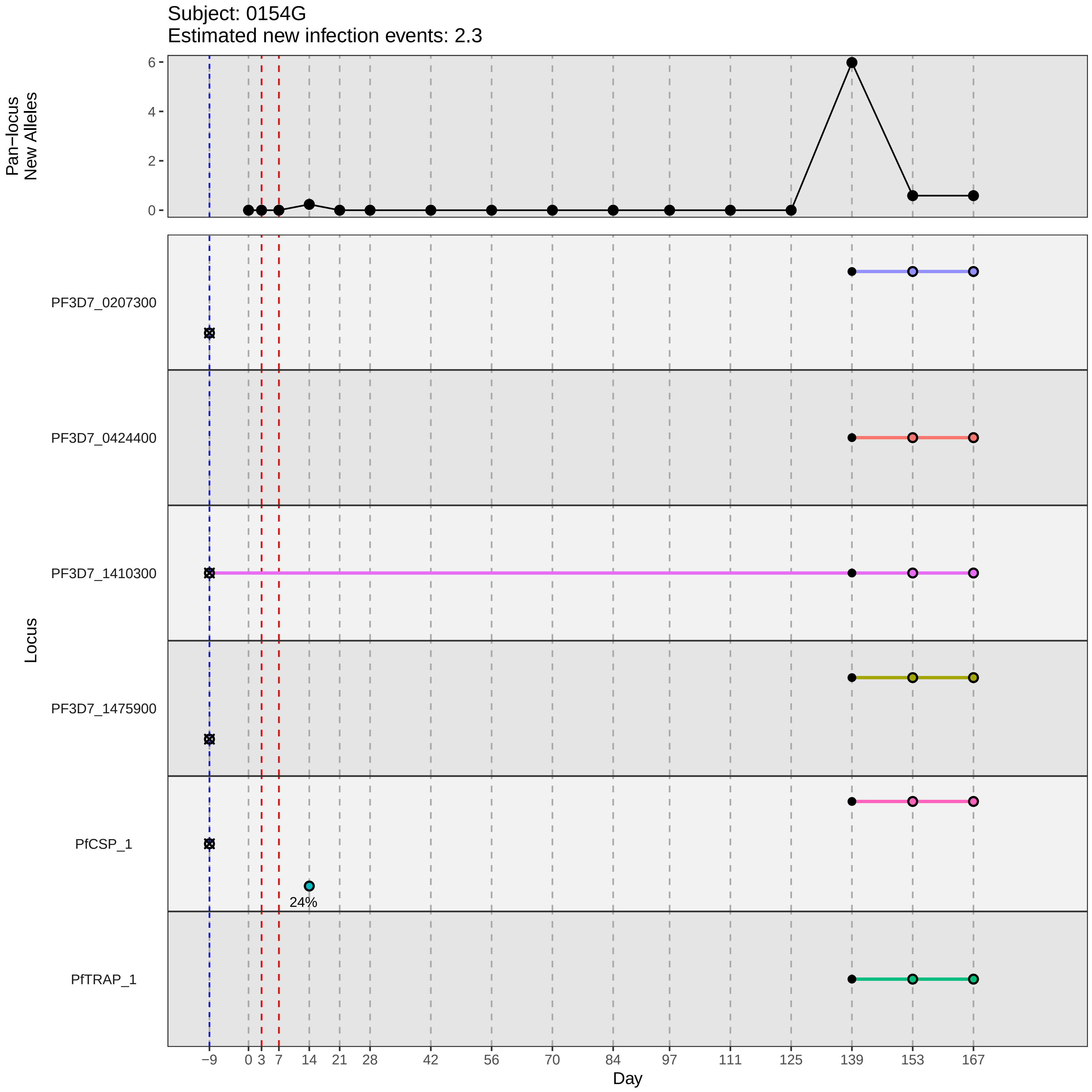
Subject: 0154G
Estimated new infection events: 2.3
Pan−locus
New Alleles
6
4
2
0
Locus
PF3D7_0207300
PF3D7_0424400
PF3D7_1410300
PF3D7_1475900
PfCSP_1
24%
PfTRAP_1
−9
0
3
7
14
21
28
42
56
70
84
97
111
125
139
153
167
Day
