## Supplementary material for "Genotypic infection endpoint analysis to understand efficacy and escape potential of the malaria monoclonal antibody CIS43LS": Data File S2: participant_0157M_without_imputation.pdf

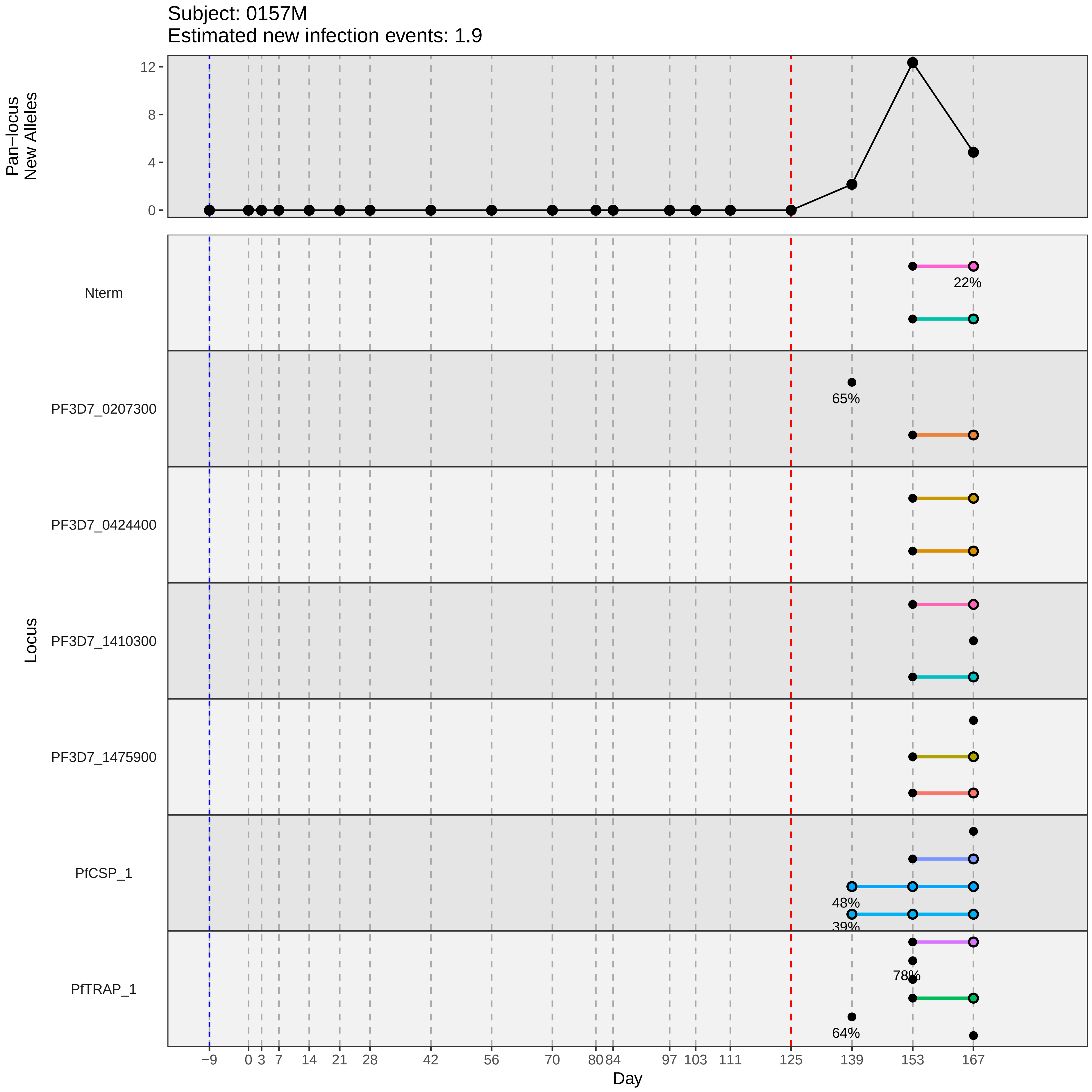

Subject: 0157M
Estimated new infection events: 1.9
Pan−locus New Alleles
0
4
8
12
Locus
Nterm
PF3D7_0207300
PF3D7_0424400
PF3D7_1410300
PF3D7_1475900
PfCSP_1
PfTRAP_1
22%
65%
48%
39%
78%
64%
−9
0
3
7
14
21
28
42
56
70
80
84
97
103
111
125
139
153
167
Day
