## Supplementary material for "Genotypic infection endpoint analysis to understand efficacy and escape potential of the malaria monoclonal antibody CIS43LS": Data File S2: participant_0162S_without_imputation.pdf

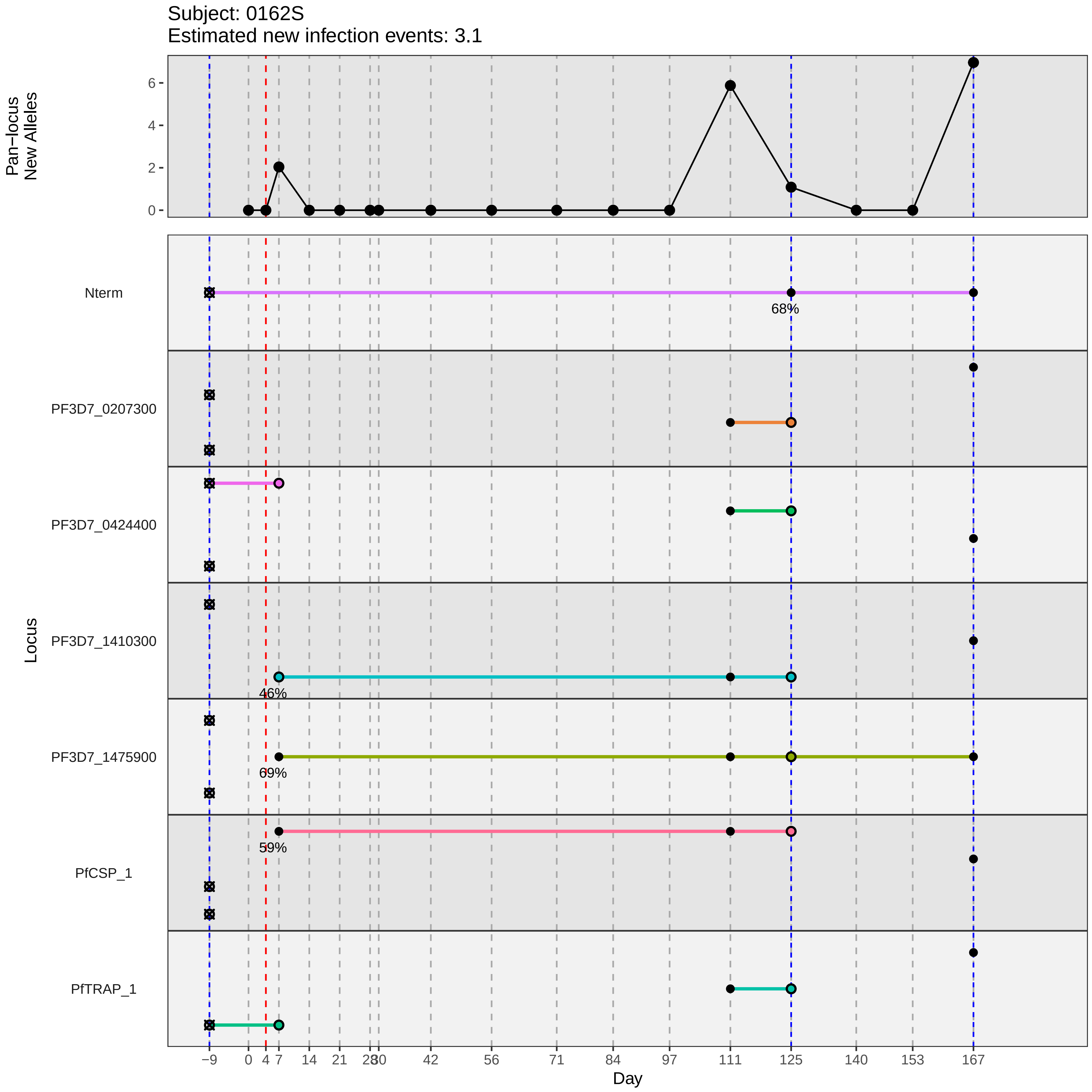

Subject: 0162S
Estimated new infection events: 3.1
Pan−locus New Alleles
0
2
4
6
Locus
Nterm
68%
PF3D7_0207300
PF3D7_0424400
PF3D7_1410300
46%
PF3D7_1475900
69%
PfCSP_1
59%
PfTRAP_1
−9
0
4
7
14
21
28
30
42
56
71
84
97
111
125
140
153
167
Day
