## Supplementary material for "Genotypic infection endpoint analysis to understand efficacy and escape potential of the malaria monoclonal antibody CIS43LS": Data File S2: participant_0174F_without_imputation.pdf

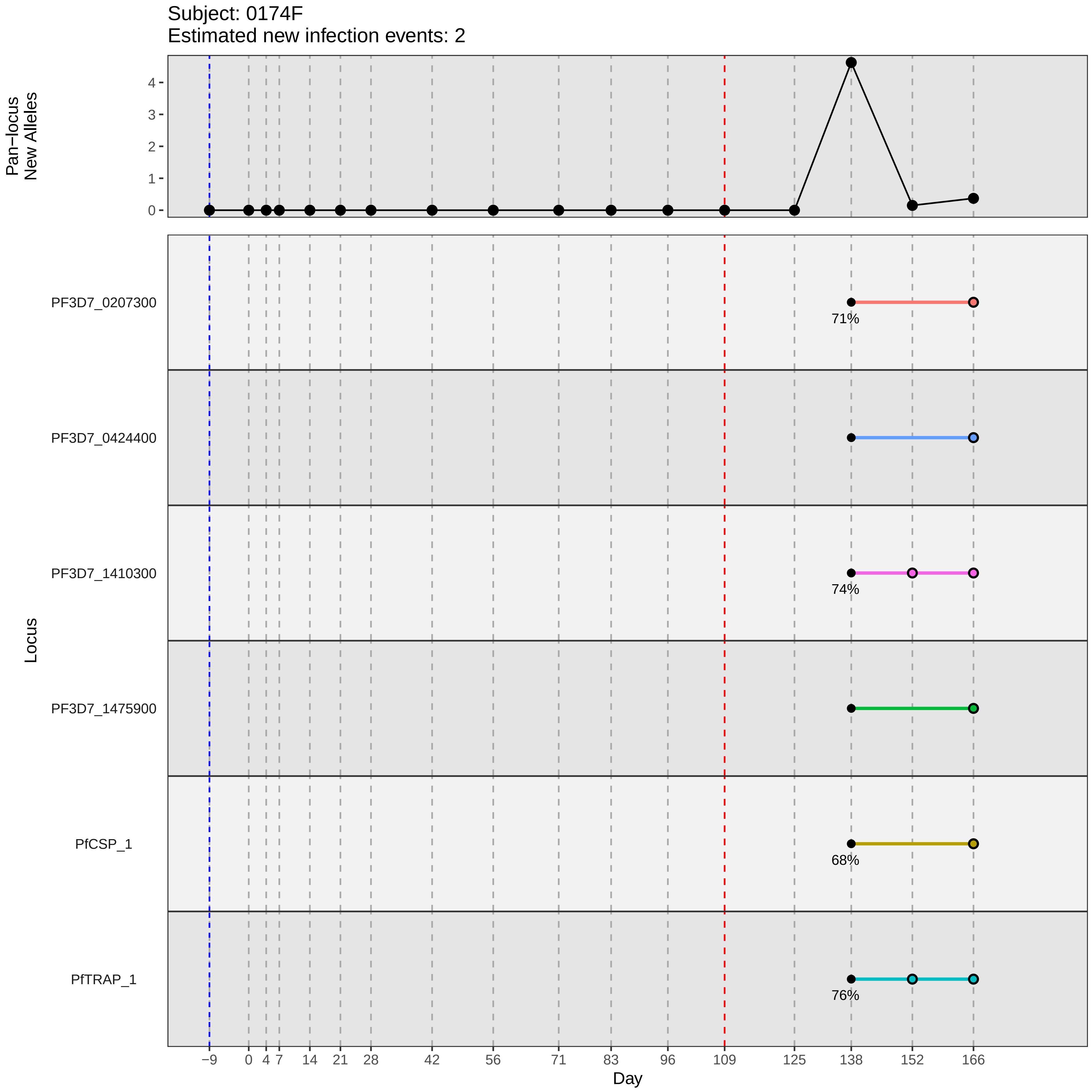

Subject: 0174F
Estimated new infection events: 2
Pan−locus
New Alleles
0
1
2
3
4
Locus
PF3D7_0207300
PF3D7_0424400
PF3D7_1410300
PF3D7_1475900
PfCSP_1
PfTRAP_1
71%
74%
68%
76%
−9
0
4
7
14
21
28
42
56
71
83
96
109
125
138
152
166
Day
