## Supplementary material for "Genotypic infection endpoint analysis to understand efficacy and escape potential of the malaria monoclonal antibody CIS43LS": Data File S2: participant_0181Q_without_imputation.pdf

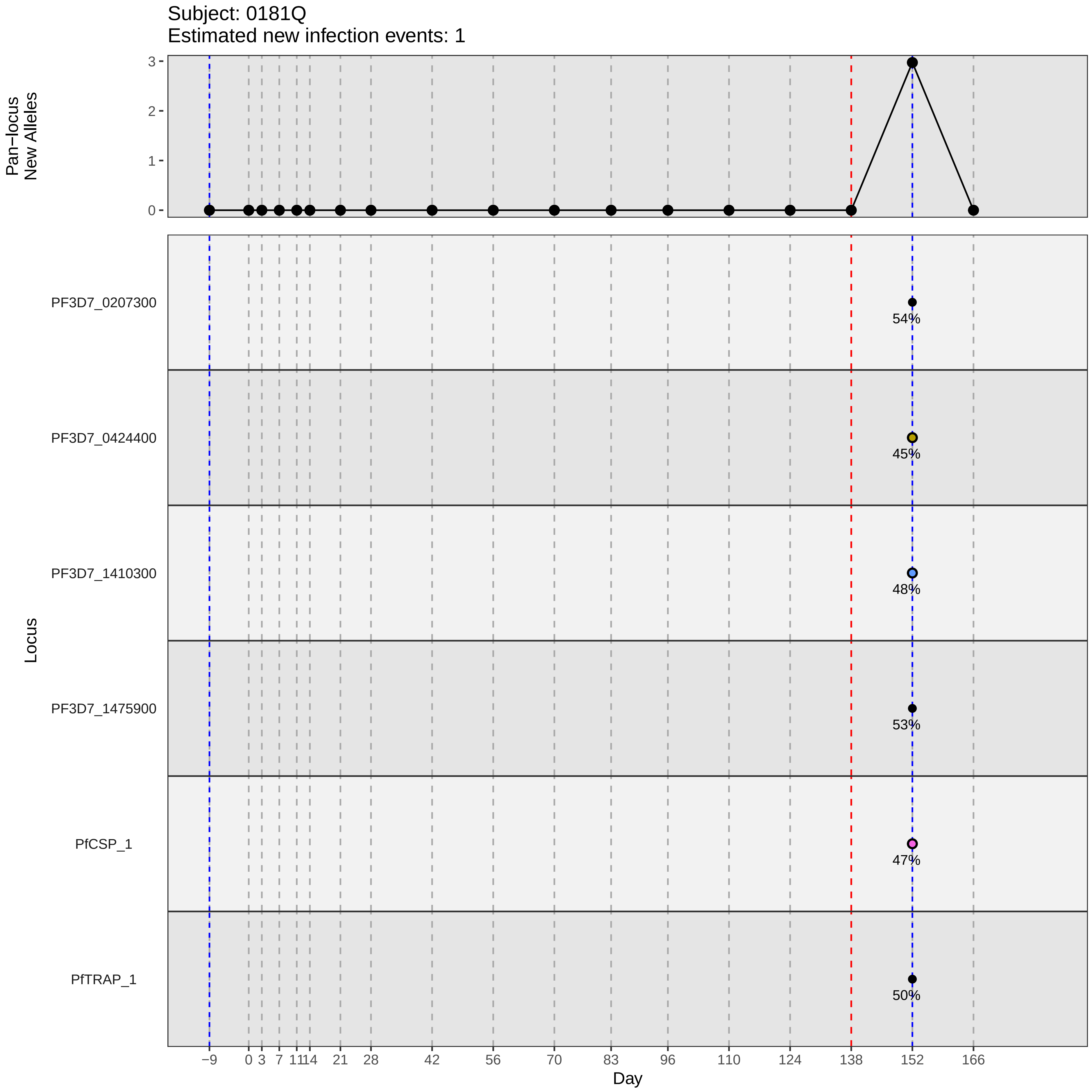

Subject: 0181Q
Estimated new infection events: 1
Pan−locus New Alleles
3
2
1
0
Locus
PF3D7_0207300
54%
PF3D7_0424400
45%
PF3D7_1410300
48%
PF3D7_1475900
53%
PfCSP_1
47%
PfTRAP_1
50%
−9 0 3 7 11 14 21 28 42 56 70 83 96 110 124 138 152 166
Day
