## Supplementary material for "Genotypic infection endpoint analysis to understand efficacy and escape potential of the malaria monoclonal antibody CIS43LS": Data File S2: participant_0187W_without_imputation.pdf

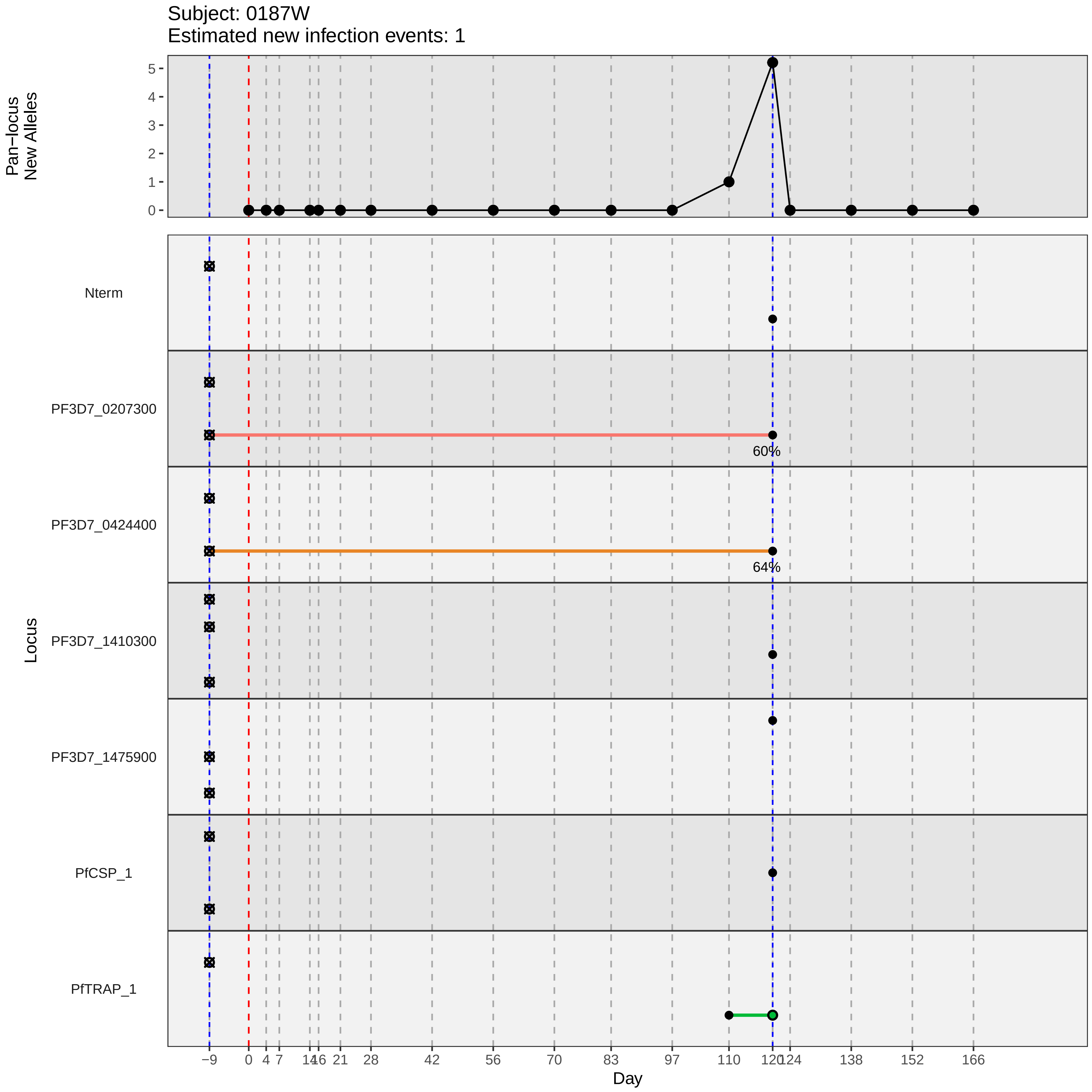
Subject: 0187W
Estimated new infection events: 1
Pan−locus
New Alleles
5
4
3
2
1
0
Locus
Nterm
PF3D7_0207300
60%
PF3D7_0424400
64%
PF3D7_1410300
PF3D7_1475900
PfCSP_1
PfTRAP_1
−9
0
4
7
14
16
21
28
42
56
70
83
97
110
120
124
138
152
166
Day
