## Supplementary material for "Genotypic infection endpoint analysis to understand efficacy and escape potential of the malaria monoclonal antibody CIS43LS": Data File S2: participant_0191B_without_imputation.pdf

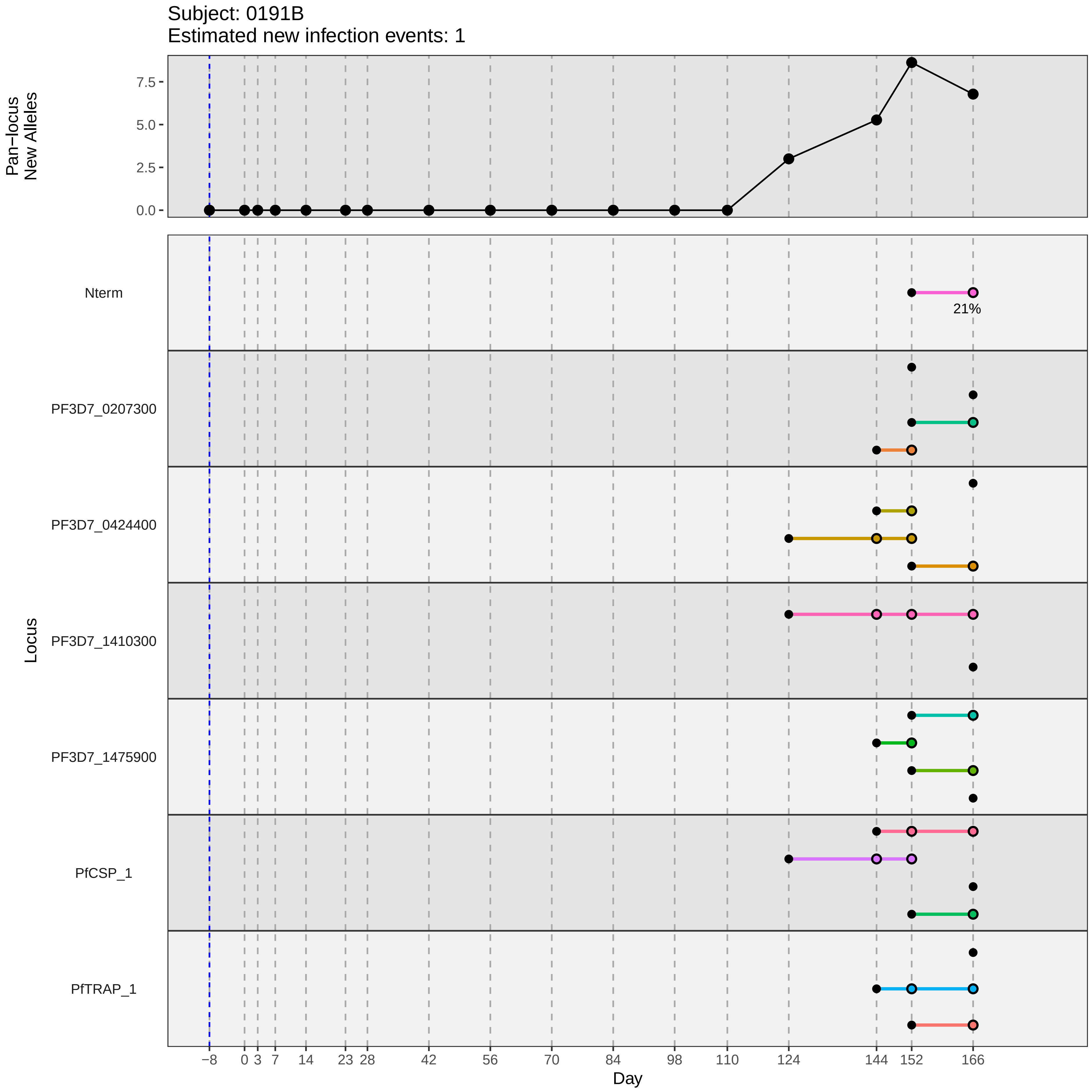
Subject: 0191B
Estimated new infection events: 1
Pan−locus
New Alleles
7.5
5.0
2.5
0.0
Locus
Nterm
21%
PF3D7_0207300
PF3D7_0424400
PF3D7_1410300
PF3D7_1475900
PfCSP_1
PfTRAP_1
−8
0
3
7
14
23
28
42
56
70
84
98
110
124
144
152
166
Day
