## Supplementary material for "Genotypic infection endpoint analysis to understand efficacy and escape potential of the malaria monoclonal antibody CIS43LS": Data File S2: participant_0199M_without_imputation.pdf

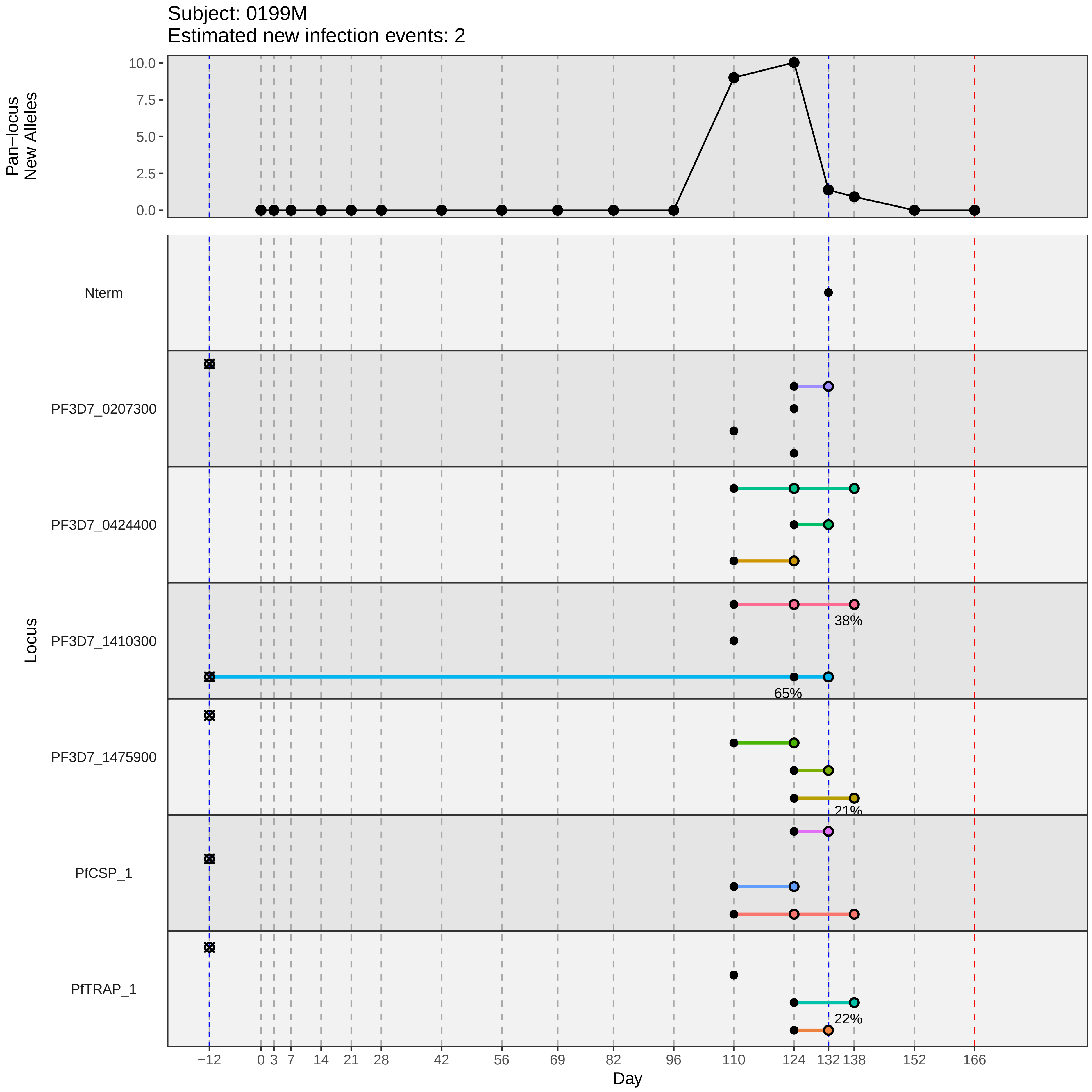
Subject: 0199M
Estimated new infection events: 2
Pan−locus New Alleles
10.0
7.5
5.0
2.5
0.0
Locus
Nterm
PF3D7_0207300
PF3D7_0424400
PF3D7_1410300
PF3D7_1475900
PfCSP_1
PfTRAP_1
38%
65%
21%
22%
−12
0
3
7
14
21
28
42
56
69
82
96
110
124
132
138
152
166
Day
