## Supplementary material for "Genotypic infection endpoint analysis to understand efficacy and escape potential of the malaria monoclonal antibody CIS43LS": Data File S2: participant_0204S_without_imputation.pdf

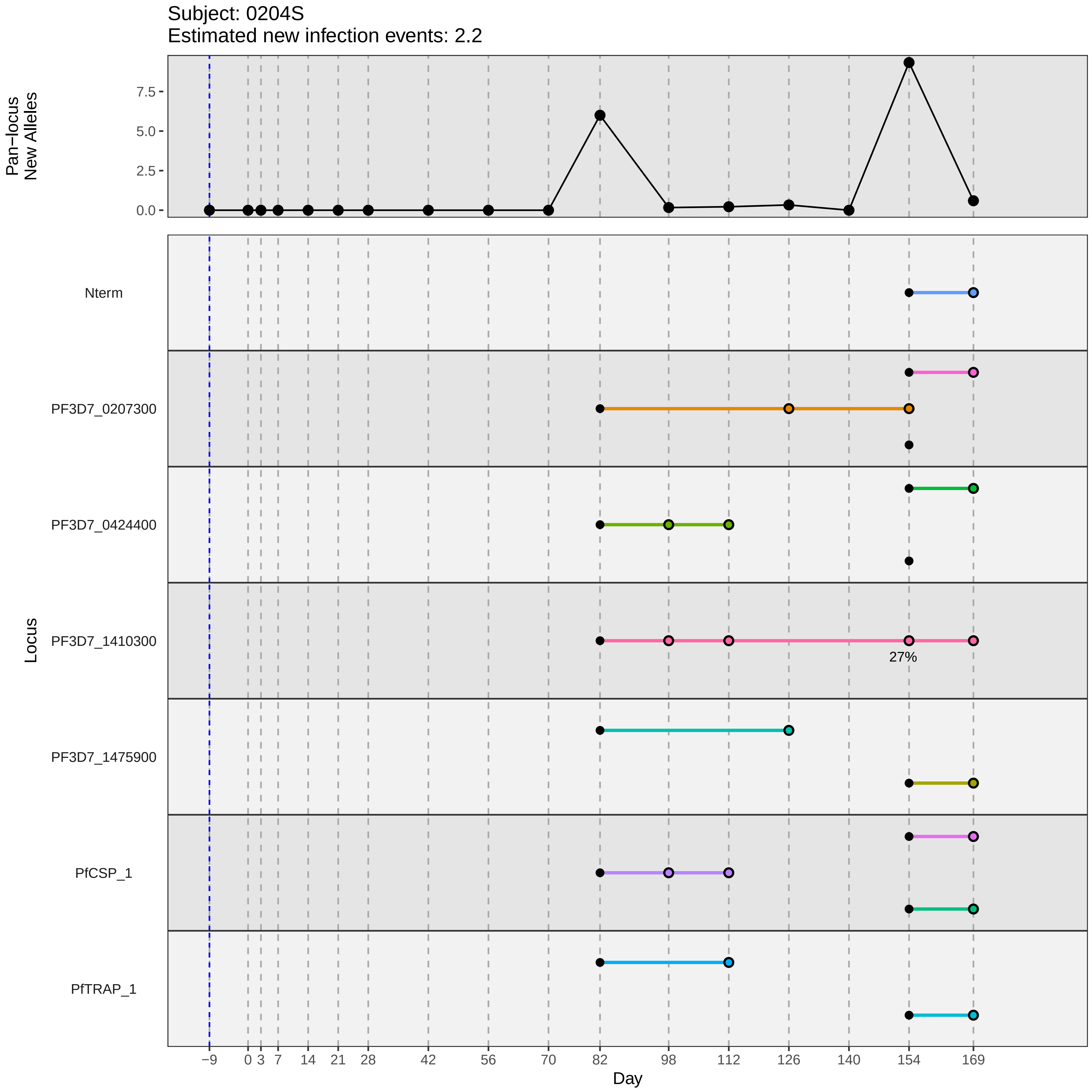

Subject: 0204S
Estimated new infection events: 2.2
Pan−locus New Alleles
7.5
5.0
2.5
0.0
Locus
Nterm
PF3D7_0207300
PF3D7_0424400
PF3D7_1410300
27%
PF3D7_1475900
PfCSP_1
PfTRAP_1
−9 0 3 7 14 21 28 42 56 70 82 98 112 126 140 154 169
Day
