## Supplementary material for "Genotypic infection endpoint analysis to understand efficacy and escape potential of the malaria monoclonal antibody CIS43LS": Data File S2: participant_0209X_without_imputation.pdf

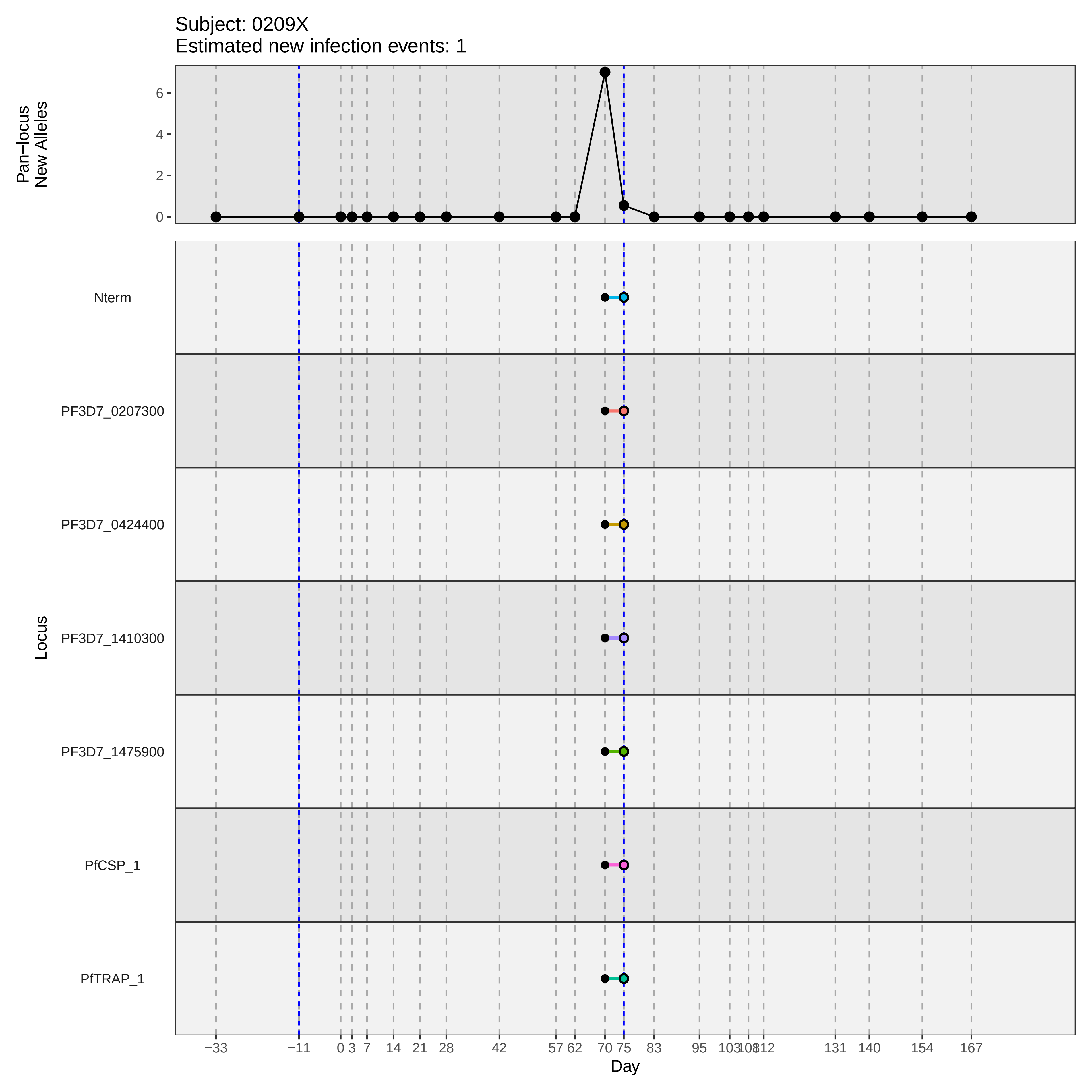
Subject: 0209X
Estimated new infection events: 1
Pan−locus
New Alleles
Locus
Nterm
PF3D7_0207300
PF3D7_0424400
PF3D7_1410300
PF3D7_1475900
PfCSP_1
PfTRAP_1
Day
−33
−11
0
3
7
14
21
28
42
57
62
70
75
83
95
103
108
112
131
140
154
167
0
2
4
6
