## Supplementary material for "Genotypic infection endpoint analysis to understand efficacy and escape potential of the malaria monoclonal antibody CIS43LS": Data File S2: participant_0210Y_without_imputation.pdf

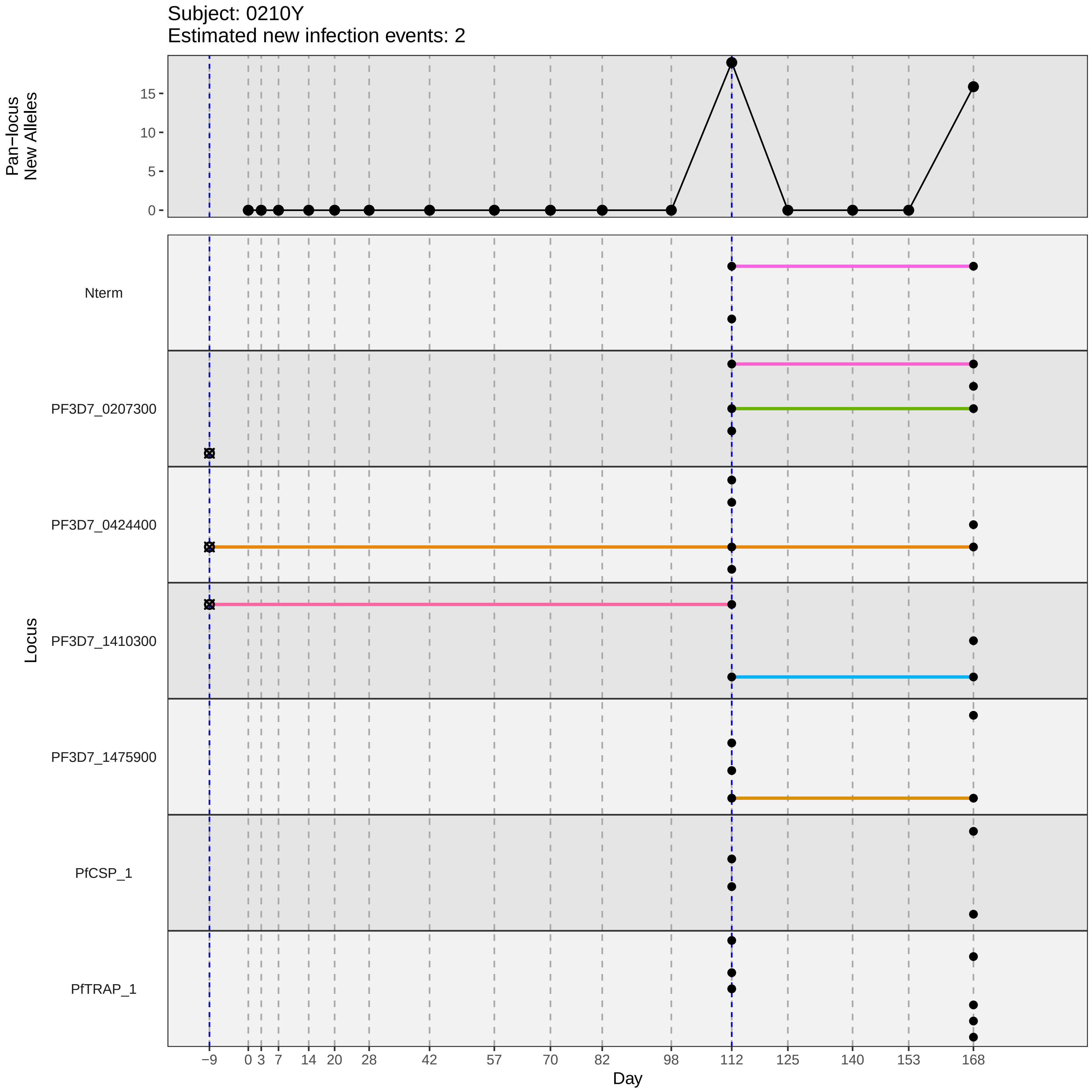
Subject: 0210Y
Estimated new infection events: 2
Pan−locus New Alleles
0
5
10
15
Locus
Nterm
PF3D7_0207300
PF3D7_0424400
PF3D7_1410300
PF3D7_1475900
PfCSP_1
PfTRAP_1
−9
0
3
7
14
20
28
42
57
70
82
98
112
125
140
153
168
Day
