## Supplementary material for "Genotypic infection endpoint analysis to understand efficacy and escape potential of the malaria monoclonal antibody CIS43LS": Data File S2: participant_0211A_without_imputation.pdf

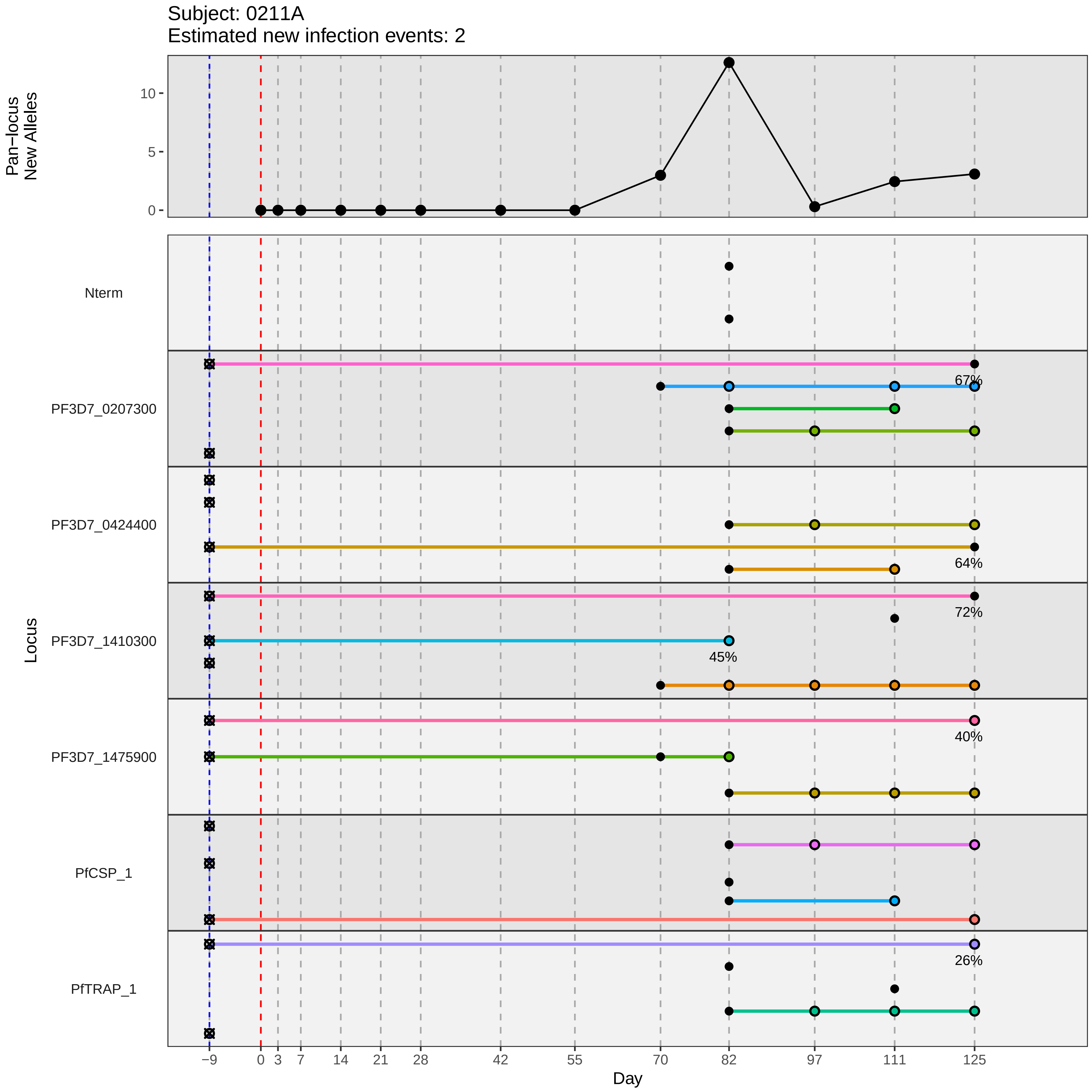

Subject: 0211A
Estimated new infection events: 2
Pan−locus New Alleles
10
5
0
Nterm
PF3D7_0207300
67%
PF3D7_0424400
64%
PF3D7_1410300
72%
45%
PF3D7_1475900
40%
PfCSP_1
PfTRAP_1
26%
Locus
−9
0
3
7
14
21
28
42
55
70
82
97
111
125
Day
