## Supplementary material for "Genotypic infection endpoint analysis to understand efficacy and escape potential of the malaria monoclonal antibody CIS43LS": Data File S2: participant_0217G_without_imputation.pdf

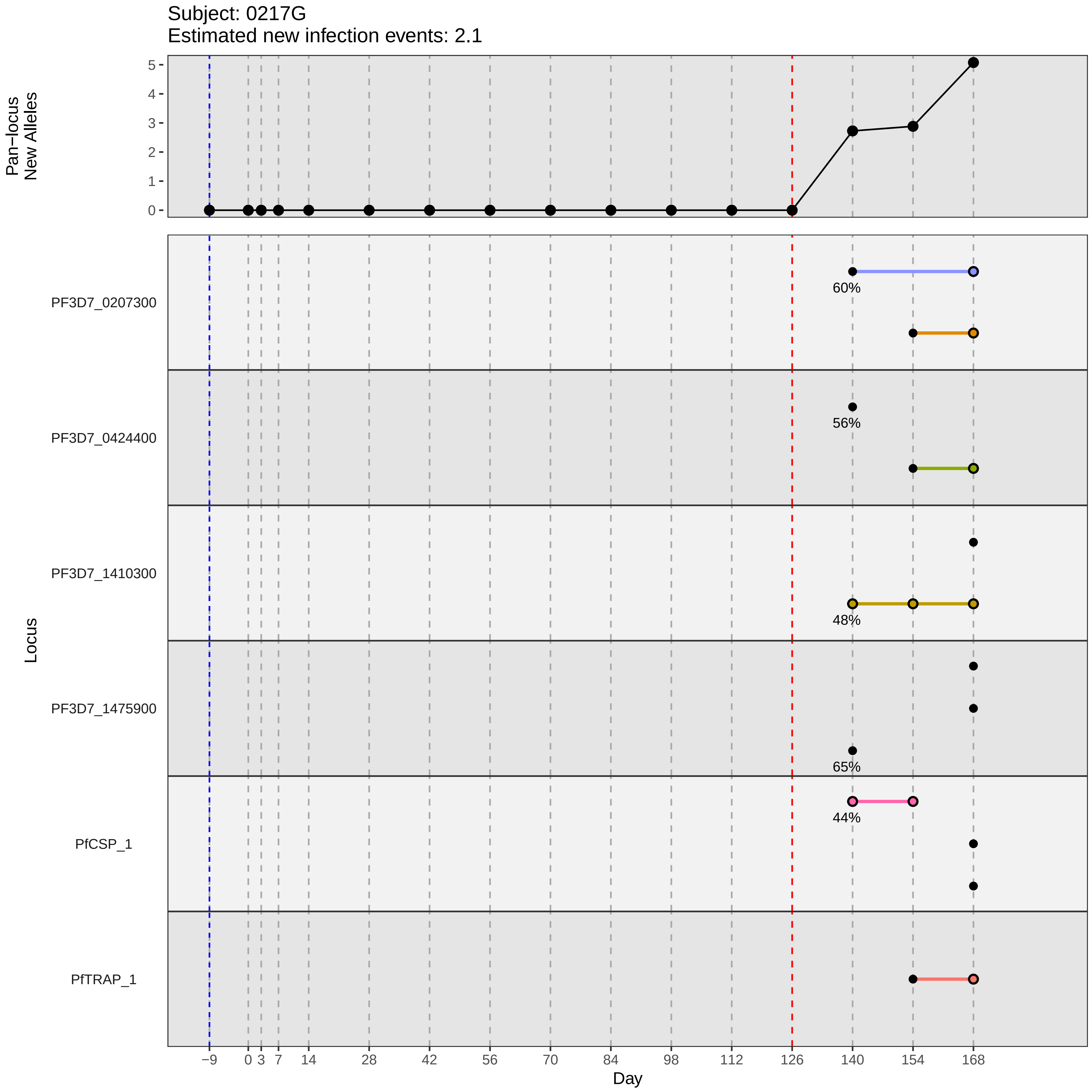

Subject: 0217G
Estimated new infection events: 2.1
Pan−locus New Alleles
0
1
2
3
4
5
Locus
PF3D7_0207300
PF3D7_0424400
PF3D7_1410300
PF3D7_1475900
PfCSP_1
PfTRAP_1
60%
56%
48%
65%
44%
−9
0
3
7
14
28
42
56
70
84
98
112
126
140
154
168
Day
