## Supplementary material for "Genotypic infection endpoint analysis to understand efficacy and escape potential of the malaria monoclonal antibody CIS43LS": Data File S2: participant_0218H_without_imputation.pdf

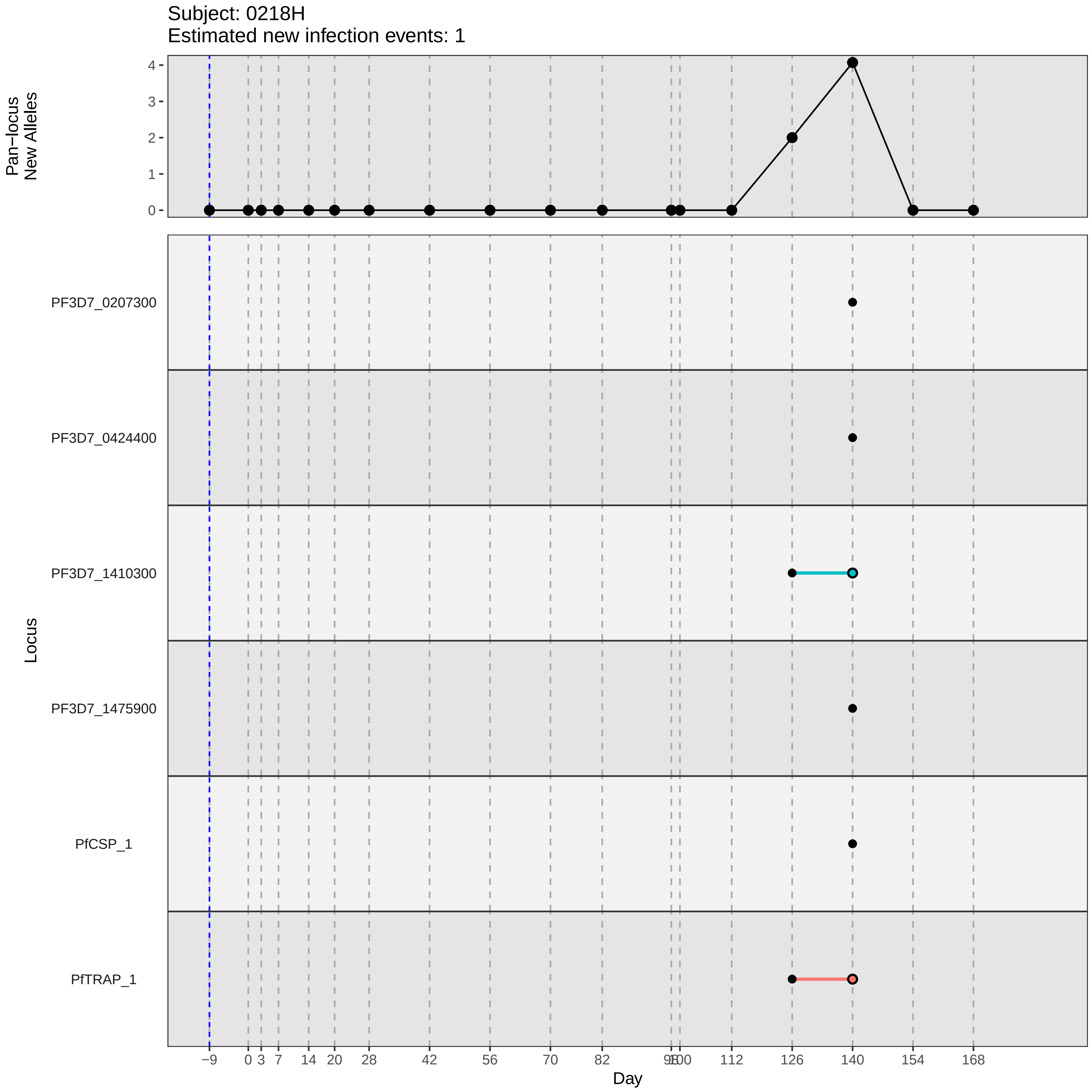
Subject: 0218H
Estimated new infection events: 1
Pan−locus
New Alleles
4
3
2
1
0
Locus
PF3D7_0207300
PF3D7_0424400
PF3D7_1410300
PF3D7_1475900
PfCSP_1
PfTRAP_1
−9
0
3
7
14
20
28
42
56
70
82
112
126
140
154
168
Day
