## Supplementary material for "Genotypic infection endpoint analysis to understand efficacy and escape potential of the malaria monoclonal antibody CIS43LS": Data File S2: participant_0219K_without_imputation.pdf

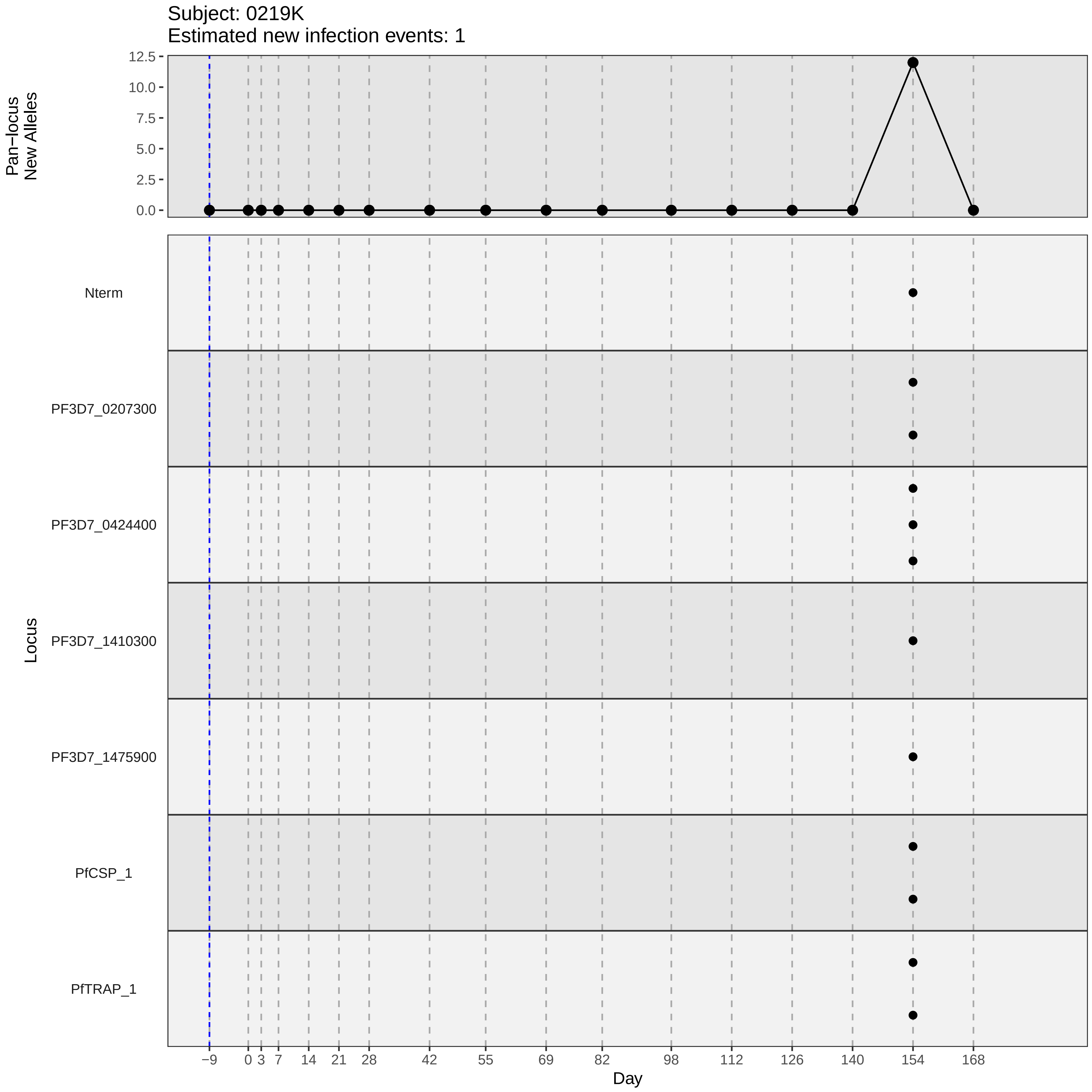

Subject: 0219K
Estimated new infection events: 1
Pan−locus
New Alleles
12.5
10.0
7.5
5.0
2.5
0.0
Locus
Nterm
PF3D7_0207300
PF3D7_0424400
PF3D7_1410300
PF3D7_1475900
PfCSP_1
PfTRAP_1
−9
0
3
7
14
21
28
42
55
69
82
98
112
126
140
154
168
Day
