## Supplementary material for "Genotypic infection endpoint analysis to understand efficacy and escape potential of the malaria monoclonal antibody CIS43LS": Data File S2: participant_0236E_without_imputation.pdf

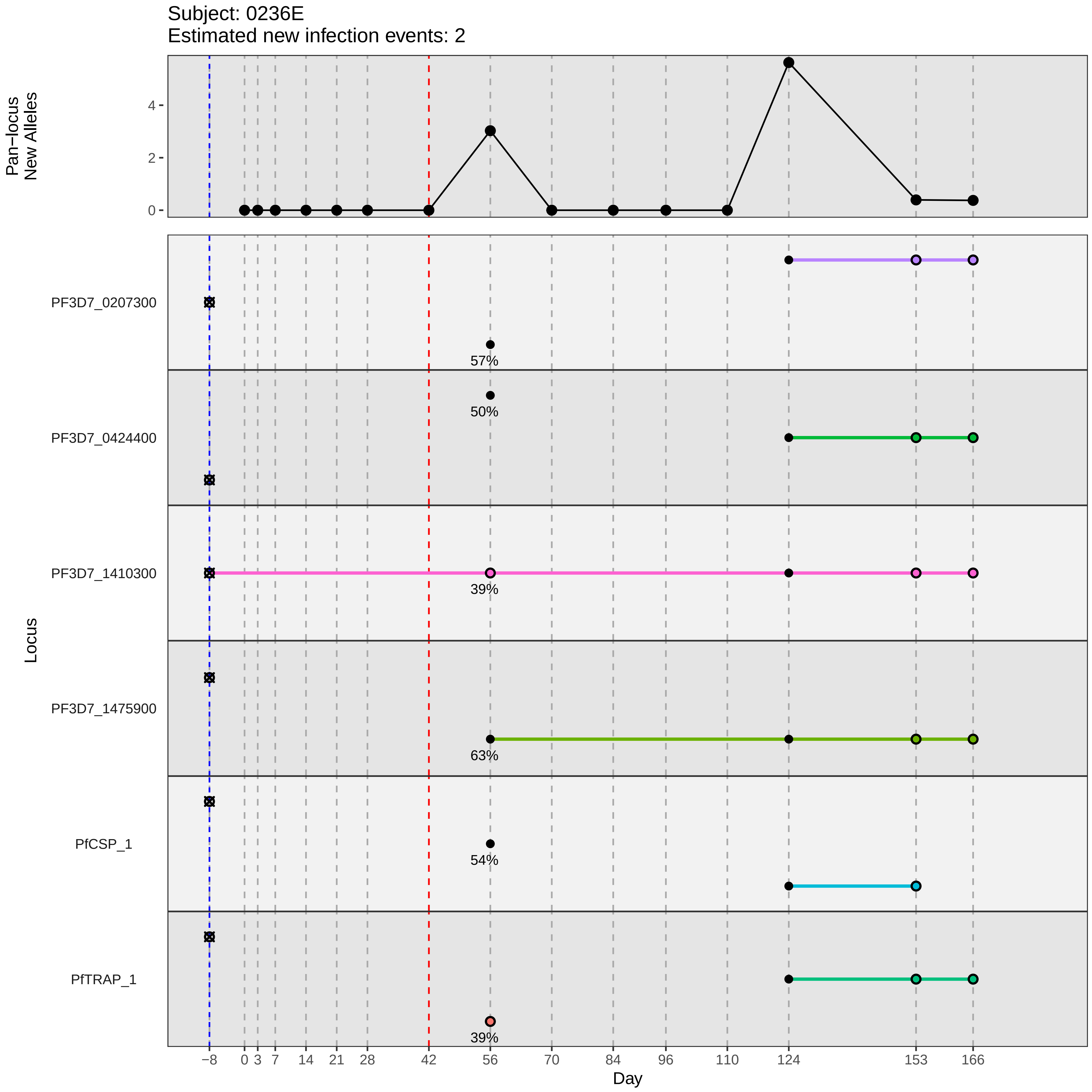
Subject: 0236E
Estimated new infection events: 2
Pan−locus New Alleles
0
2
4
Locus
PF3D7_0207300
PF3D7_0424400
PF3D7_1410300
PF3D7_1475900
PfCSP_1
PfTRAP_1
57%
50%
39%
63%
54%
39%
−8
0
3
7
14
21
28
42
56
70
84
96
110
124
153
166
Day
