## Supplementary material for "Genotypic infection endpoint analysis to understand efficacy and escape potential of the malaria monoclonal antibody CIS43LS": Data File S2: participant_0240K_without_imputation.pdf

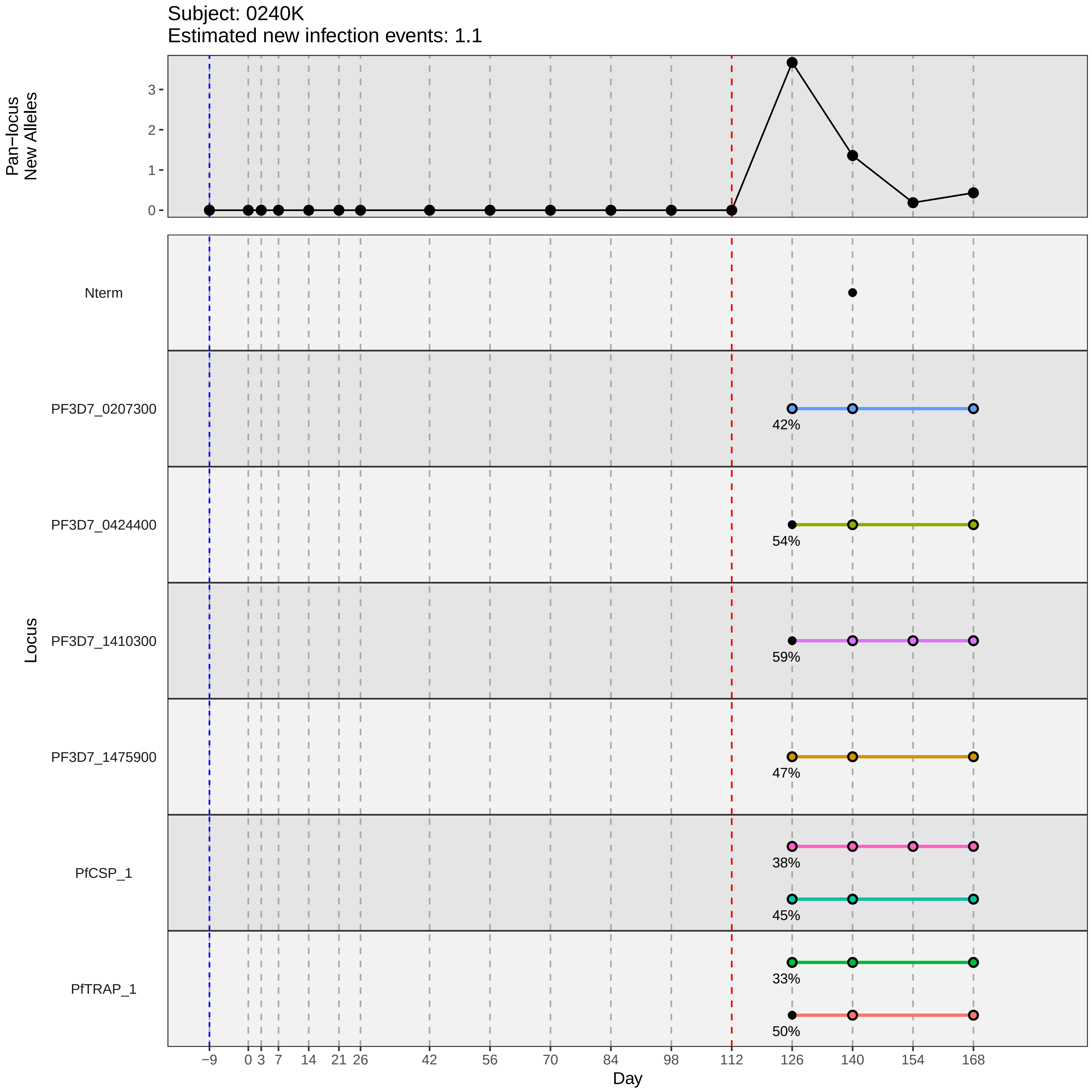

Subject: 0240K
Estimated new infection events: 1.1
Pan−locus New Alleles
0
1
2
3
Locus
Nterm
PF3D7_0207300
PF3D7_0424400
PF3D7_1410300
PF3D7_1475900
PfCSP_1
PfTRAP_1
42%
54%
59%
47%
38%
45%
33%
50%
−9
0
3
7
14
21
26
42
56
70
84
98
112
126
140
154
168
Day
