## Supplementary material for "Genotypic infection endpoint analysis to understand efficacy and escape potential of the malaria monoclonal antibody CIS43LS": Data File S2: participant_0241M_without_imputation.pdf

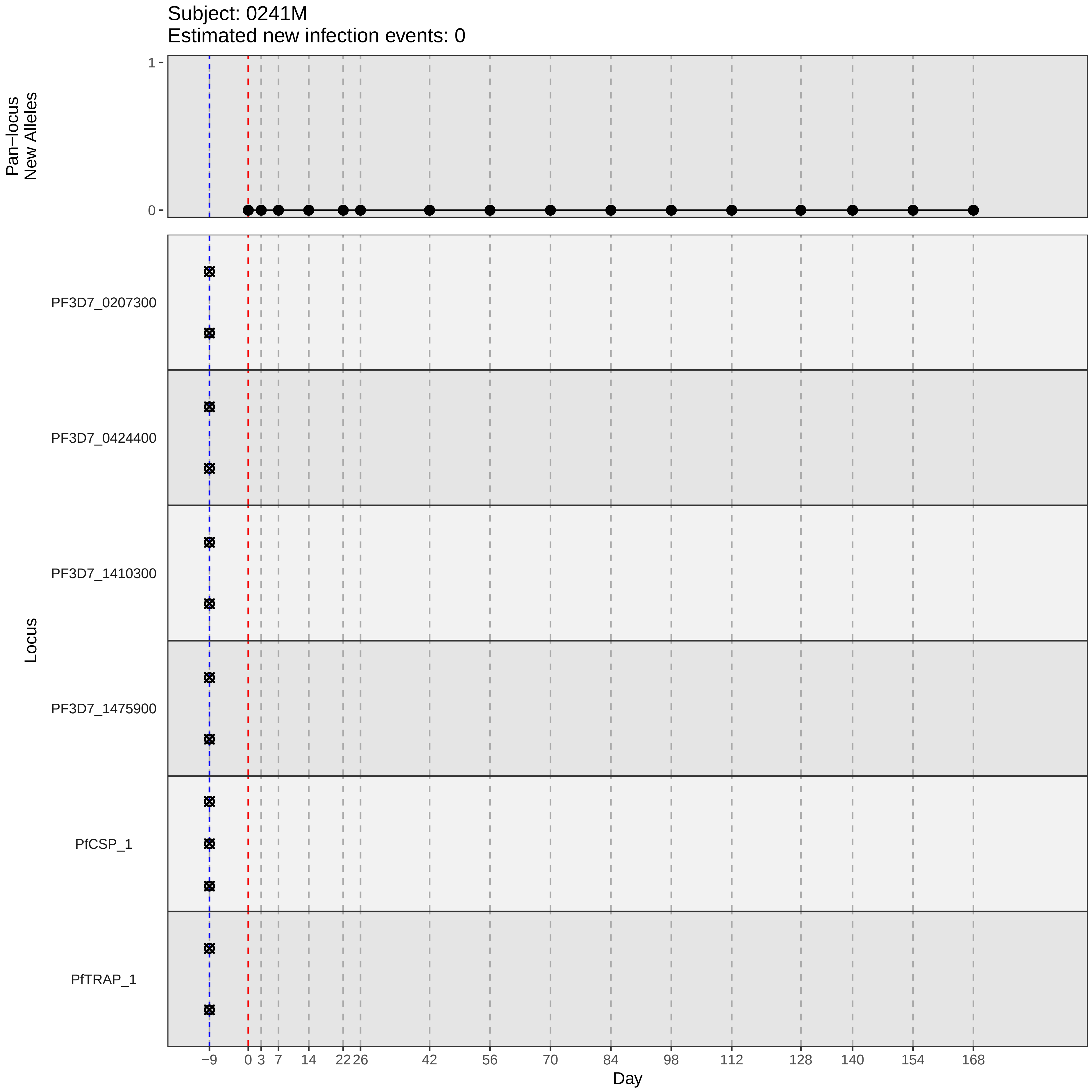

Subject: 0241M
Estimated new infection events: 0
Pan−locus New Alleles
1
0
Locus
PF3D7_0207300
PF3D7_0424400
PF3D7_1410300
PF3D7_1475900
PfCSP_1
PfTRAP_1
−9
0
3
7
14
22
26
42
56
70
84
98
112
128
140
154
168
Day
