## Supplementary material for "Genotypic infection endpoint analysis to understand efficacy and escape potential of the malaria monoclonal antibody CIS43LS": Data File S2: participant_0243P_without_imputation.pdf

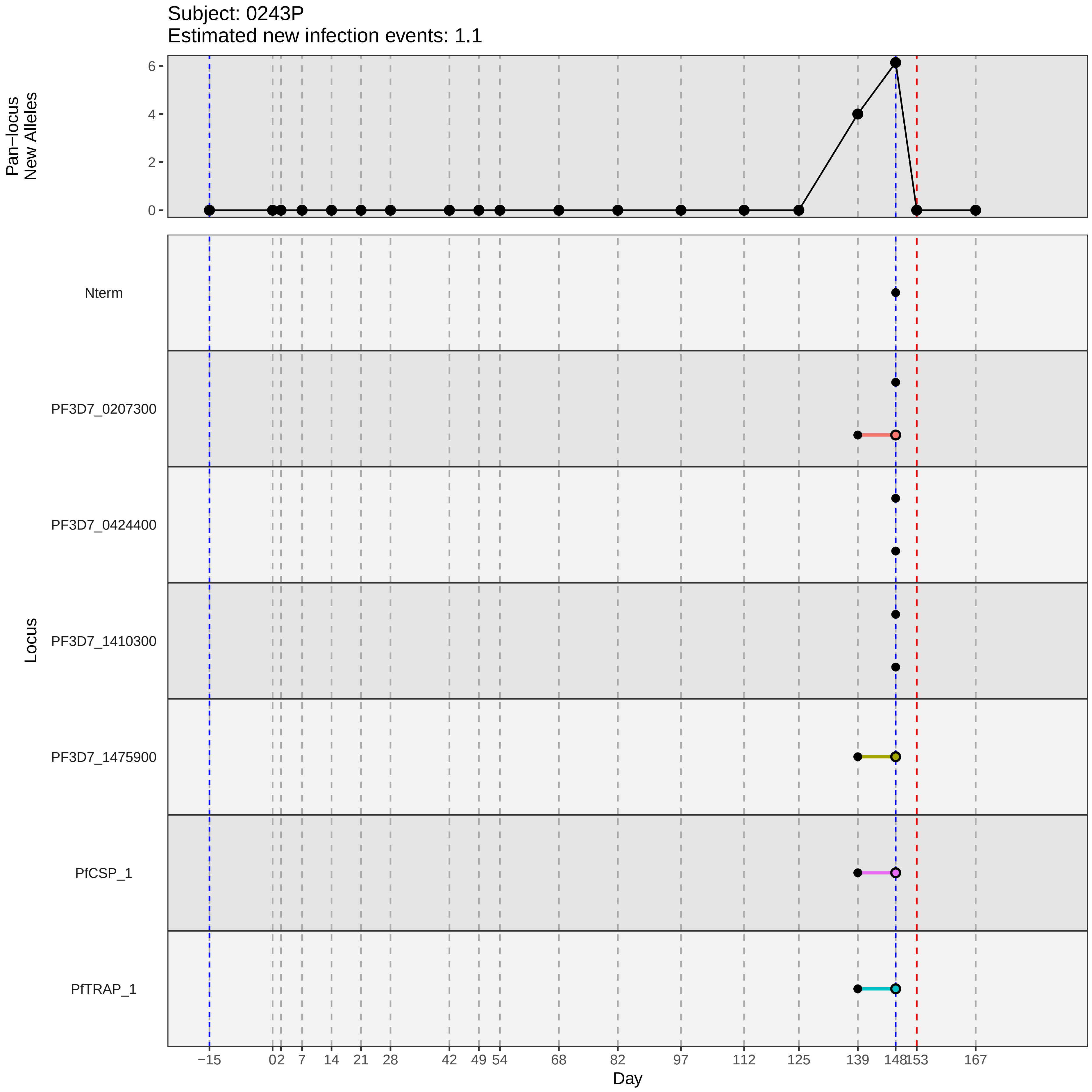

Subject: 0243P
Estimated new infection events: 1.1
Pan−locus New Alleles
0
2
4
6
Locus
Nterm
PF3D7_0207300
PF3D7_0424400
PF3D7_1410300
PF3D7_1475900
PfCSP_1
PfTRAP_1
−15
0
2
7
14
21
28
42
49
54
68
82
97
112
125
139
148
153
167
Day
