## Supplementary material for "Genotypic infection endpoint analysis to understand efficacy and escape potential of the malaria monoclonal antibody CIS43LS": Data File S2: participant_0244Q_without_imputation.pdf

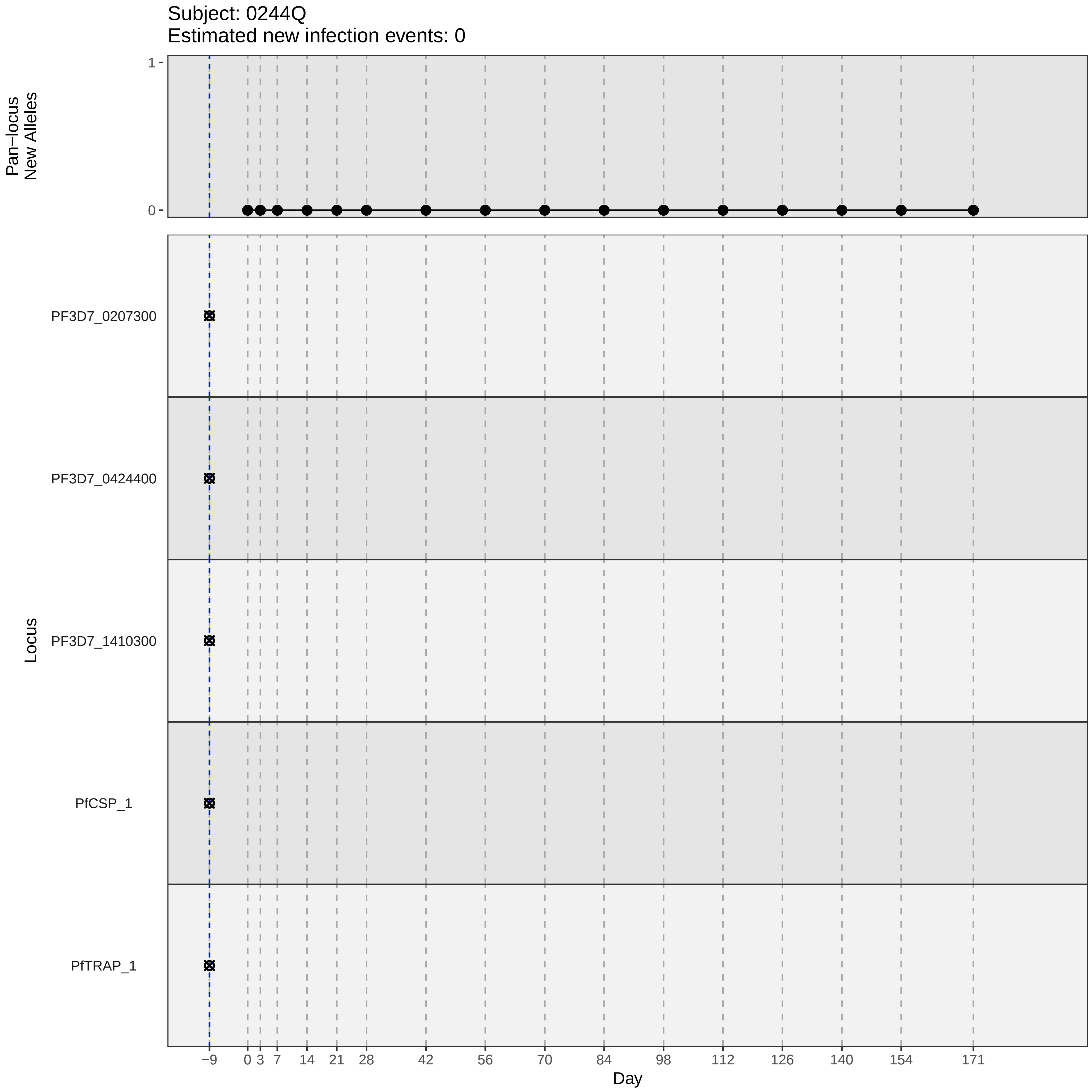

Subject: 0244Q
Estimated new infection events: 0
Pan−locus
New Alleles
1
0
Locus
PF3D7_0207300
PF3D7_0424400
PF3D7_1410300
PfCSP_1
PfTRAP_1
−9
0
3
7
14
21
28
42
56
70
84
98
112
126
140
154
171
Day
