## Supplementary material for "Genotypic infection endpoint analysis to understand efficacy and escape potential of the malaria monoclonal antibody CIS43LS": Data File S2: participant_0245R_without_imputation.pdf

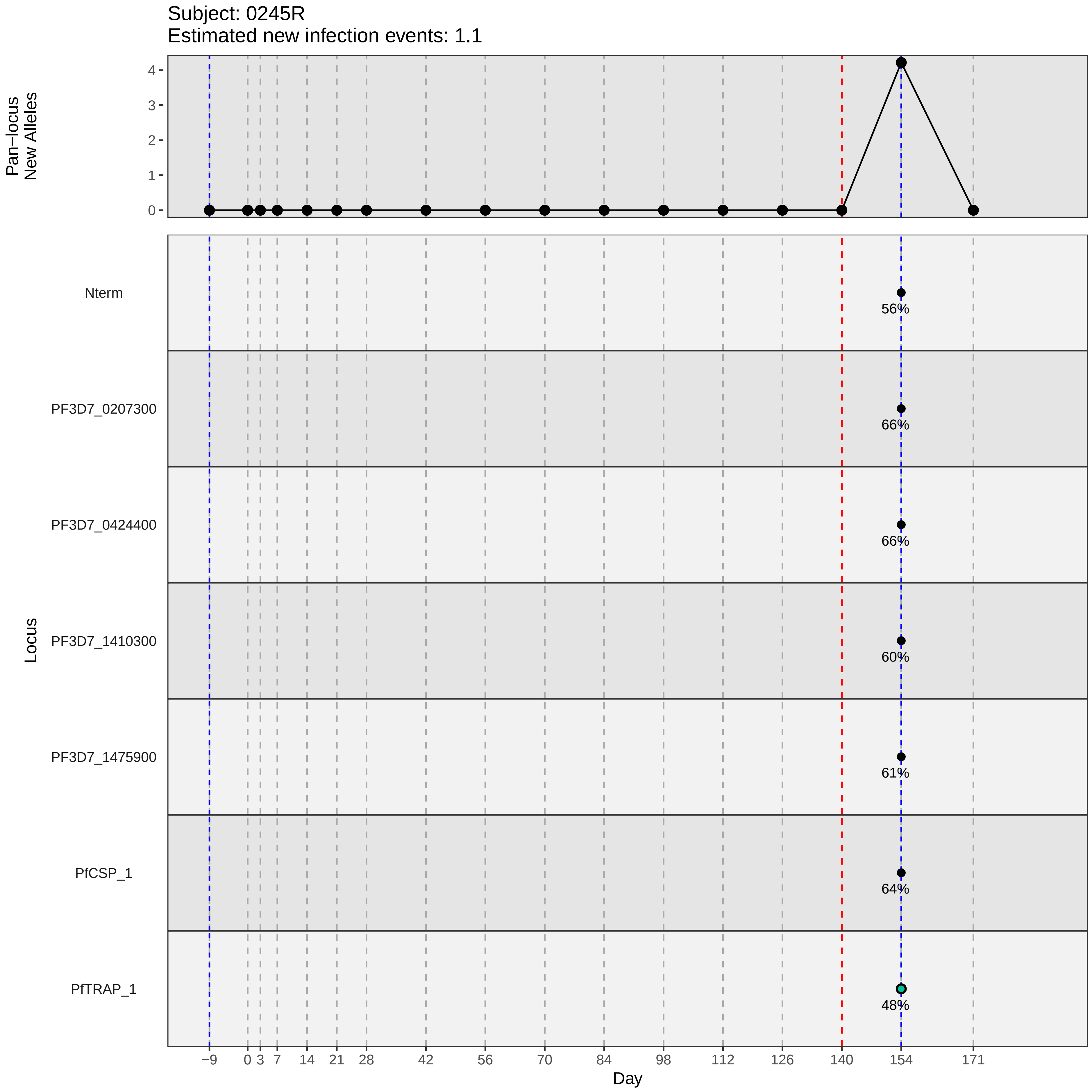
Subject: 0245R
Estimated new infection events: 1.1
Pan−locus
New Alleles
0
1
2
3
4
Locus
Nterm
PF3D7_0207300
PF3D7_0424400
PF3D7_1410300
PF3D7_1475900
PfCSP_1
PfTRAP_1
56%
66%
66%
60%
61%
64%
48%
−9
0
3
7
14
21
28
42
56
70
84
98
112
126
140
154
171
Day
