## Supplementary material for "Genotypic infection endpoint analysis to understand efficacy and escape potential of the malaria monoclonal antibody CIS43LS": Data File S2: participant_0252Y_without_imputation.pdf

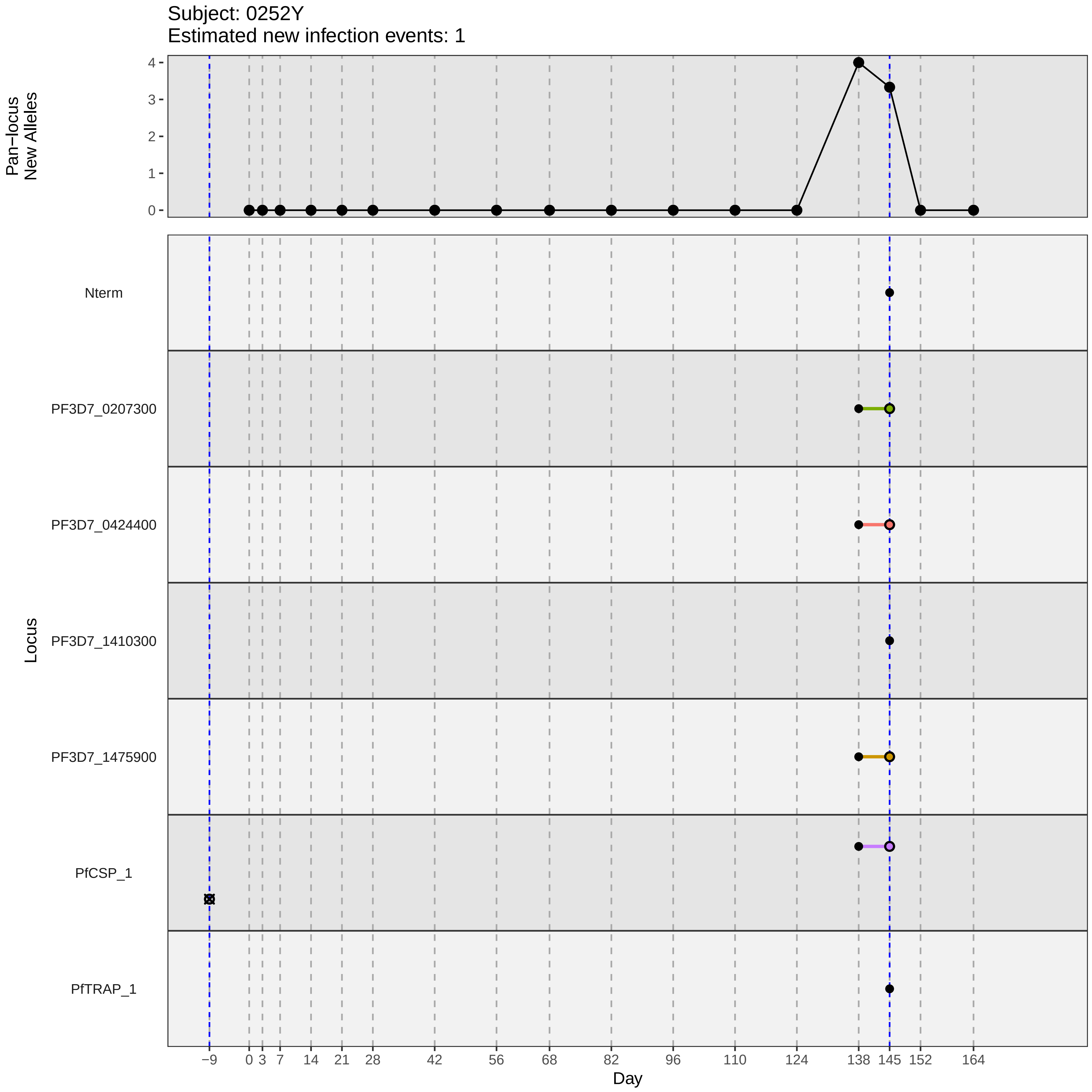
Subject: 0252Y
Estimated new infection events: 1
Pan−locus New Alleles
Locus
Nterm
PF3D7_0207300
PF3D7_0424400
PF3D7_1410300
PF3D7_1475900
PfCSP_1
PfTRAP_1
Day
−9 0 3 7 14 21 28 42 56 68 82 96 110 124 138 145 152 164
0 1 2 3 4
