## Supplementary material for "Genotypic infection endpoint analysis to understand efficacy and escape potential of the malaria monoclonal antibody CIS43LS": Data File S2: participant_0254B_without_imputation.pdf

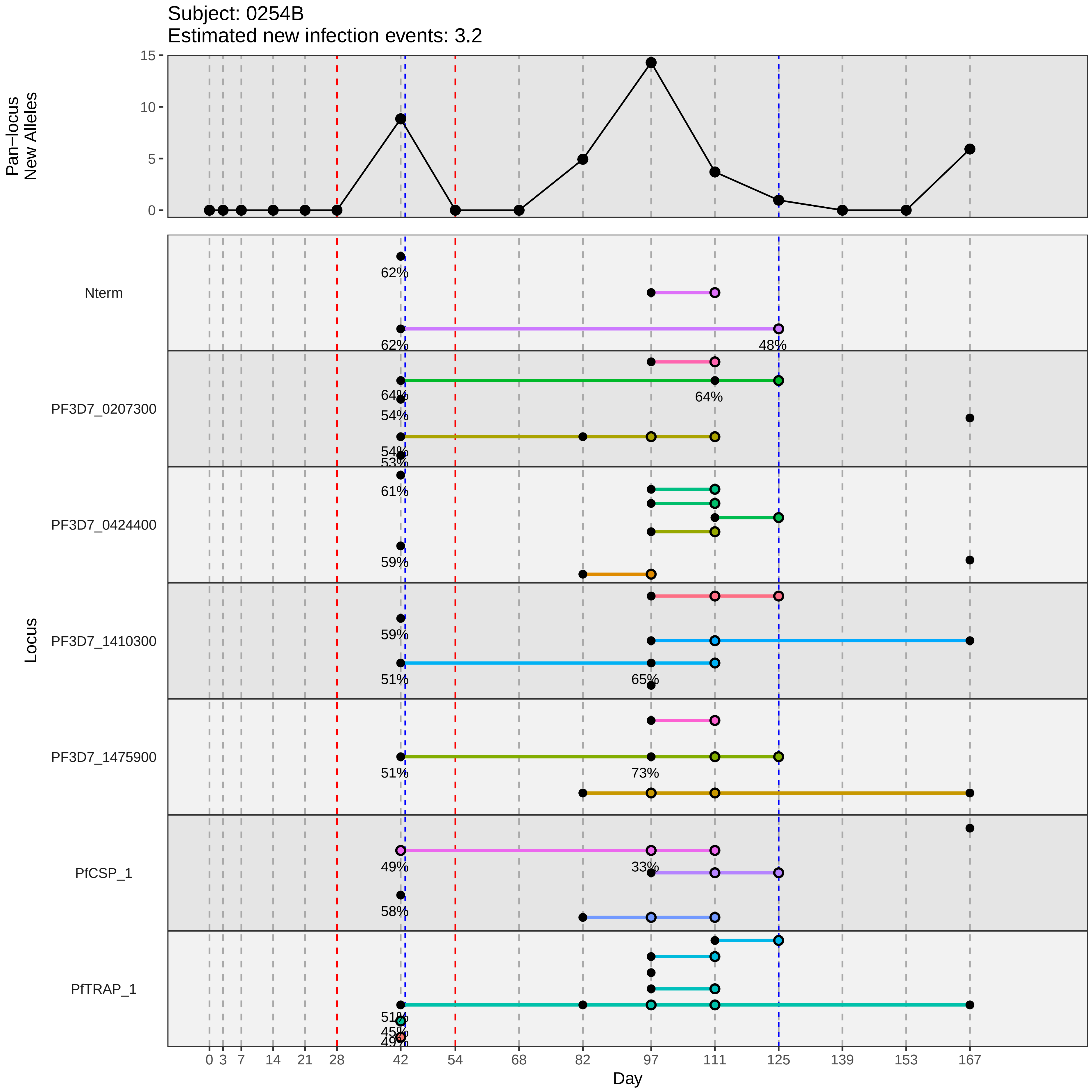

Subject: 0254B
Estimated new infection events: 3.2
Pan−locus
New Alleles
15
10
5
0
Locus
Nterm
PF3D7_0207300
PF3D7_0424400
PF3D7_1410300
PF3D7_1475900
PfCSP_1
PfTRAP_1
62%
62%
48%
64%
64%
54%
54%
53%
61%
59%
59%
51%
65%
51%
73%
49%
33%
58%
51%
45%
49%
0
3
7
14
21
28
42
54
68
82
97
111
125
139
153
167
Day
