## Supplementary material for "Genotypic infection endpoint analysis to understand efficacy and escape potential of the malaria monoclonal antibody CIS43LS": Data File S2: participant_0255C_without_imputation.pdf

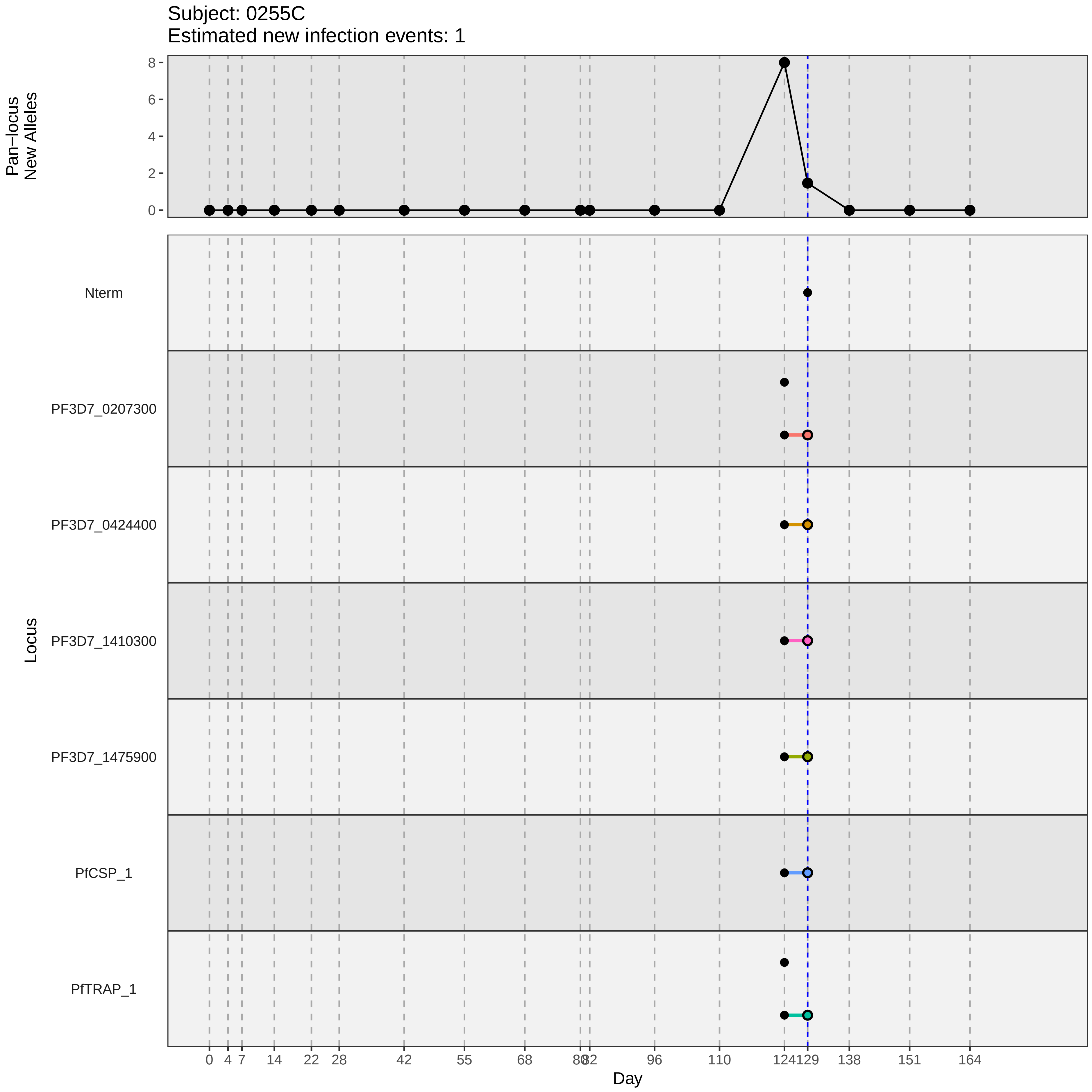
Subject: 0255C
Estimated new infection events: 1
Pan−locus
New Alleles
8
6
4
2
0
Locus
Nterm
PF3D7_0207300
PF3D7_0424400
PF3D7_1410300
PF3D7_1475900
PfCSP_1
PfTRAP_1
0
4
7
14
22
28
42
55
68
80
82
96
110
124
129
138
151
164
Day
