## Supplementary material for "Genotypic infection endpoint analysis to understand efficacy and escape potential of the malaria monoclonal antibody CIS43LS": Data File S2: participant_0259G_without_imputation.pdf

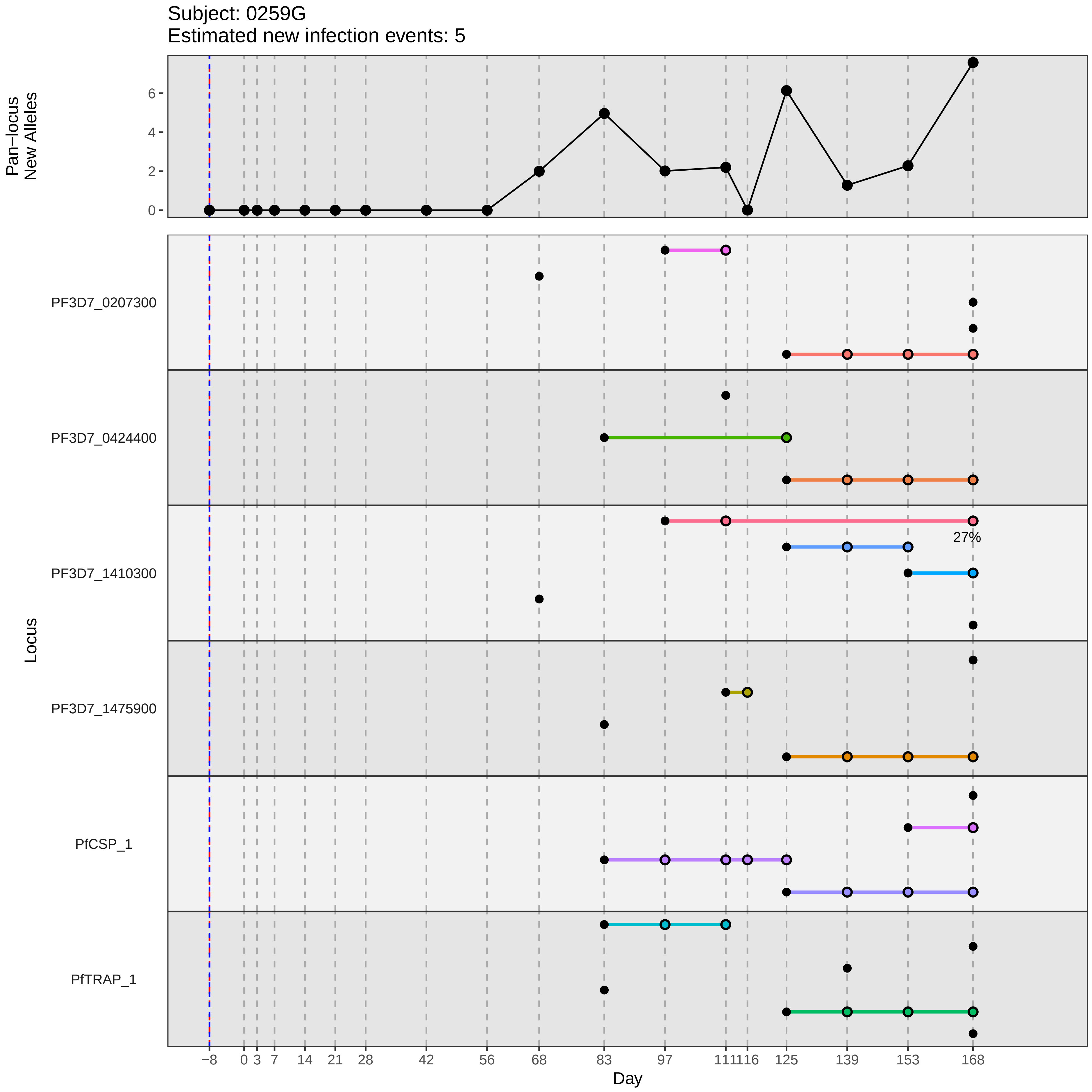
Subject: 0259G
Estimated new infection events: 5
Pan−locus New Alleles
0
2
4
6
Locus
PF3D7_0207300
PF3D7_0424400
PF3D7_1410300
PF3D7_1475900
PfCSP_1
PfTRAP_1
27%
−8
0
3
7
14
21
28
42
56
68
83
97
111116
125
139
153
168
Day
