## Supplementary material for "Genotypic infection endpoint analysis to understand efficacy and escape potential of the malaria monoclonal antibody CIS43LS": Data File S2: participant_0261K_without_imputation.pdf

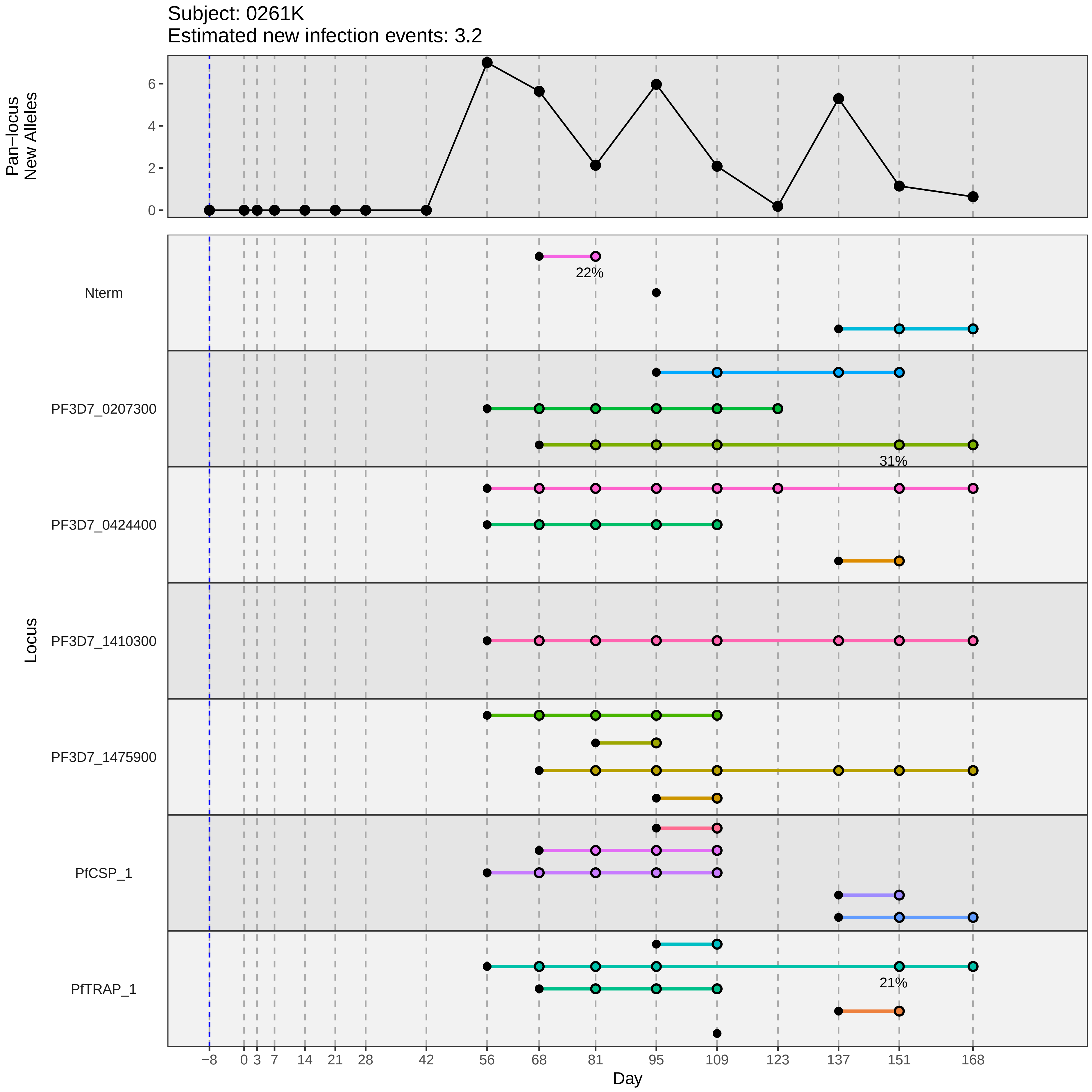

Subject: 0261K
Estimated new infection events: 3.2
Pan−locus New Alleles
0
2
4
6
Locus
Nterm
22%
PF3D7_0207300
31%
PF3D7_0424400
PF3D7_1410300
PF3D7_1475900
PfCSP_1
PfTRAP_1
21%
−8
0
3
7
14
21
28
42
56
68
81
95
109
123
137
151
168
Day
