## Supplementary material for "Genotypic infection endpoint analysis to understand efficacy and escape potential of the malaria monoclonal antibody CIS43LS": Data File S2: participant_0262M_without_imputation.pdf

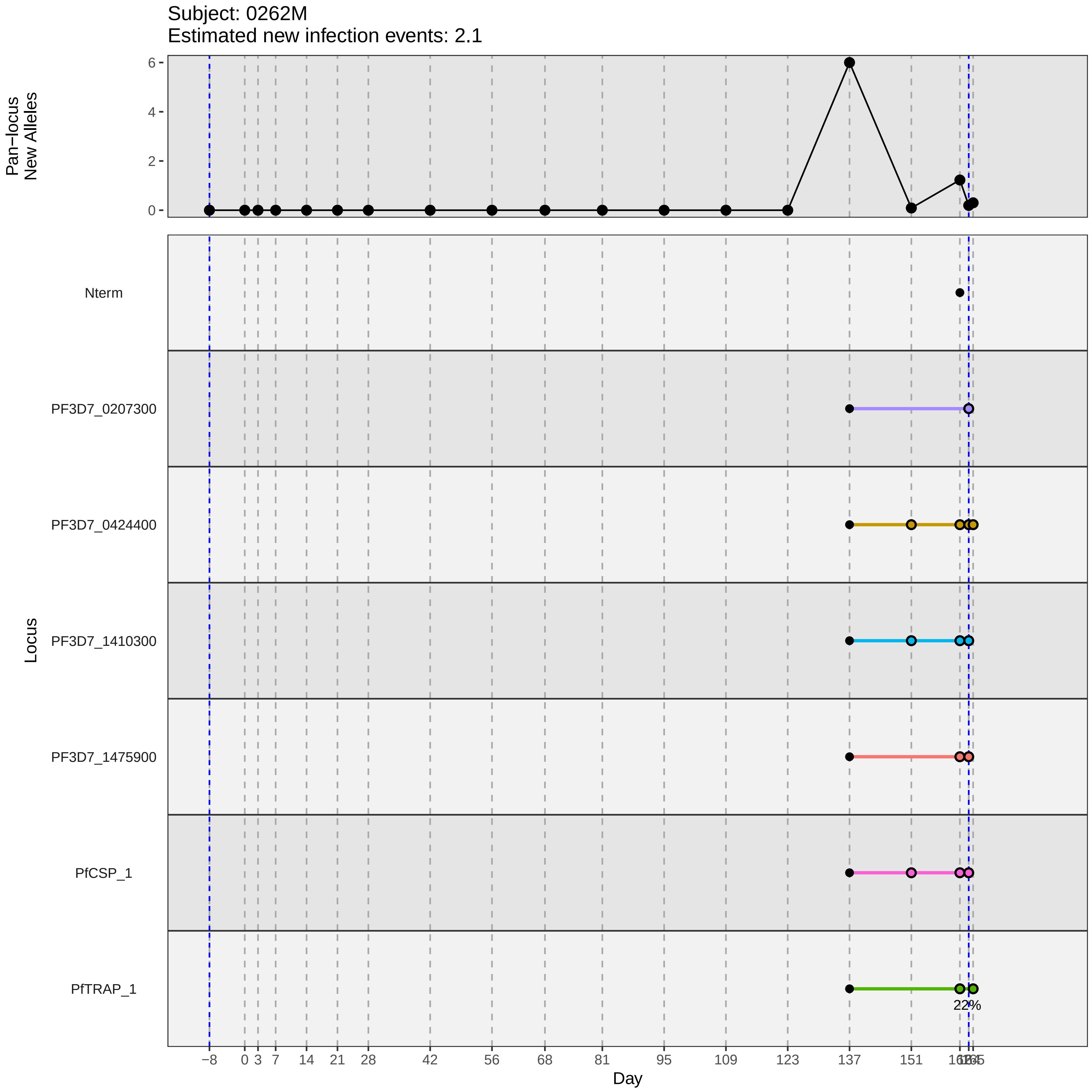
Subject: 0262M
Estimated new infection events: 2.1
Pan−locus
New Alleles
6
4
2
0
Locus
Nterm
PF3D7_0207300
PF3D7_0424400
PF3D7_1410300
PF3D7_1475900
PfCSP_1
PfTRAP_1
22%
−8
0
3
7
14
21
28
42
56
68
81
95
109
123
137
151
Day
