## Supplementary material for "Genotypic infection endpoint analysis to understand efficacy and escape potential of the malaria monoclonal antibody CIS43LS": Data File S2: participant_0266R_without_imputation.pdf

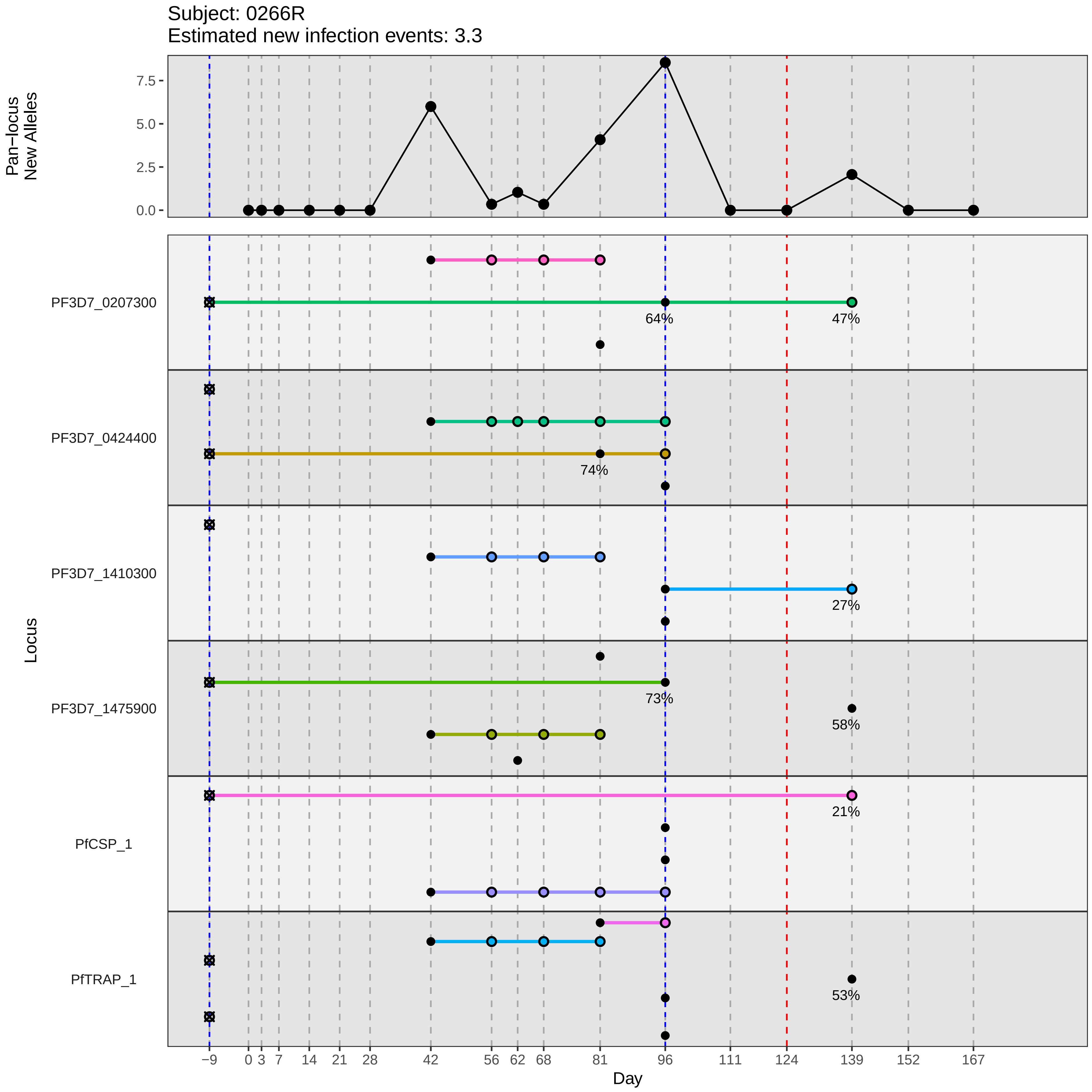

Subject: 0266R
Estimated new infection events: 3.3
Pan−locus
New Alleles
7.5
5.0
2.5
0.0
Locus
PF3D7_0207300
PF3D7_0424400
PF3D7_1410300
PF3D7_1475900
PfCSP_1
PfTRAP_1
64%
47%
74%
27%
73%
58%
21%
53%
−9
0
3
7
14
21
28
42
56
62
68
81
96
111
124
139
152
167
Day
