## Supplementary material for "Genotypic infection endpoint analysis to understand efficacy and escape potential of the malaria monoclonal antibody CIS43LS": Data File S2: participant_0271W_without_imputation.pdf

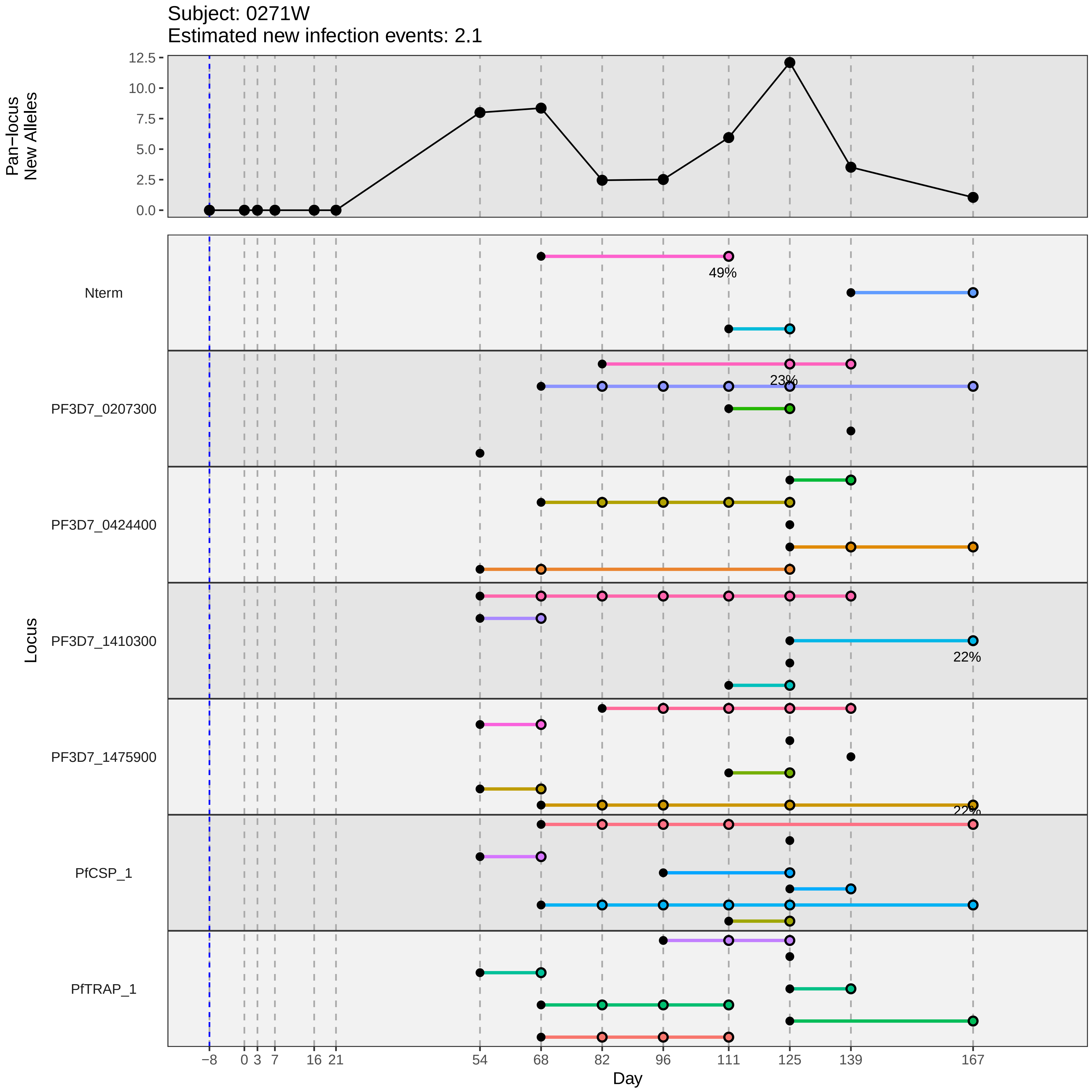
Subject: 0271W
Estimated new infection events: 2.1
Pan−locus New Alleles
12.5
10.0
7.5
5.0
2.5
0.0
Locus
Nterm
PF3D7_0207300
PF3D7_0424400
PF3D7_1410300
PF3D7_1475900
PfCSP_1
PfTRAP_1
49%
23%
22%
22%
−8
0
3
7
16
21
54
68
82
96
111
125
139
167
Day
