## Supplementary material for "Genotypic infection endpoint analysis to understand efficacy and escape potential of the malaria monoclonal antibody CIS43LS": Data File S2: participant_0274A_without_imputation.pdf

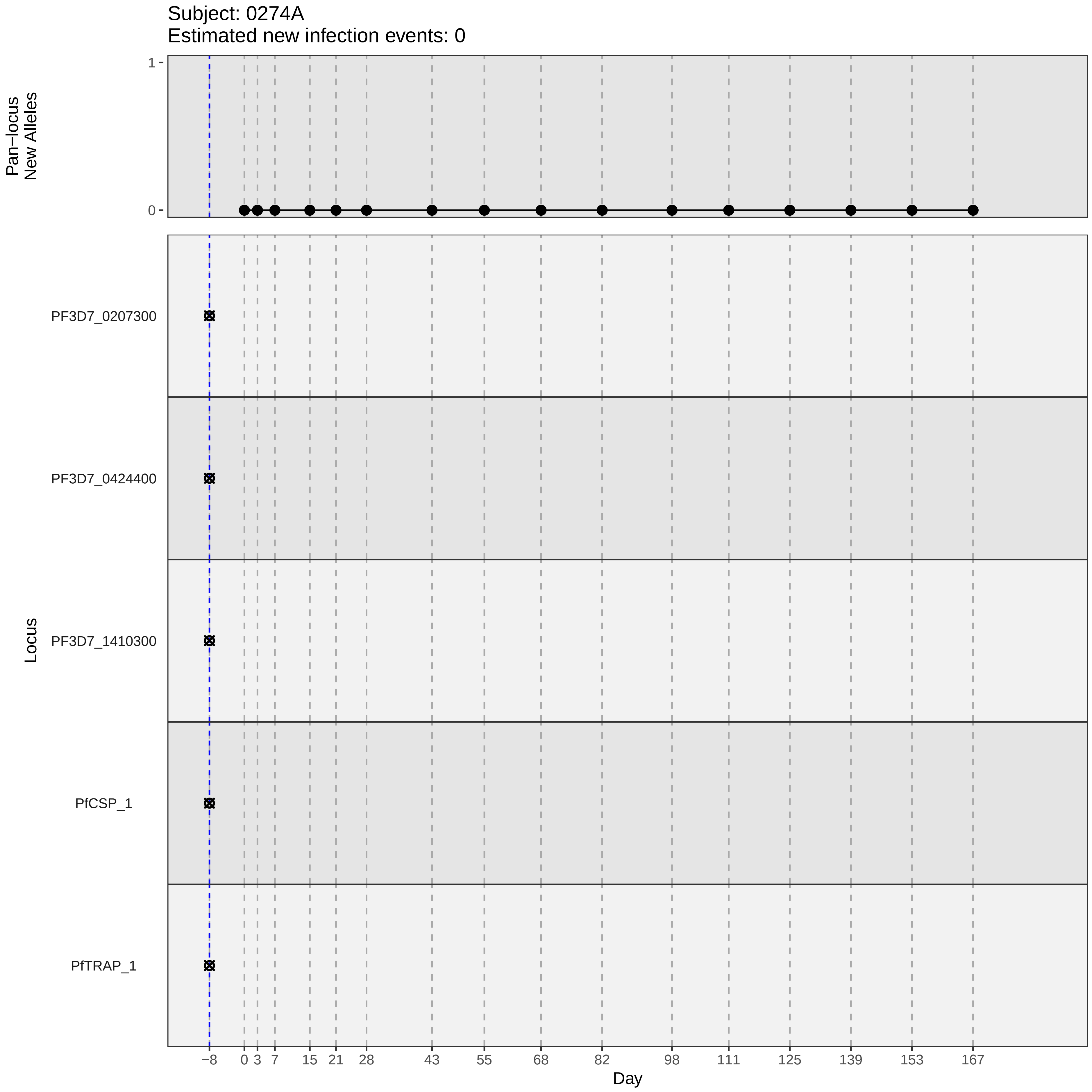
Subject: 0274A
Estimated new infection events: 0
Pan−locus New Alleles
1
0
Locus
PF3D7_0207300
PF3D7_0424400
PF3D7_1410300
PfCSP_1
PfTRAP_1
−8
0
3
7
15
21
28
43
55
68
82
98
111
125
139
153
167
Day
