## Supplementary material for "Genotypic infection endpoint analysis to understand efficacy and escape potential of the malaria monoclonal antibody CIS43LS": Data File S2: participant_0277D_without_imputation.pdf

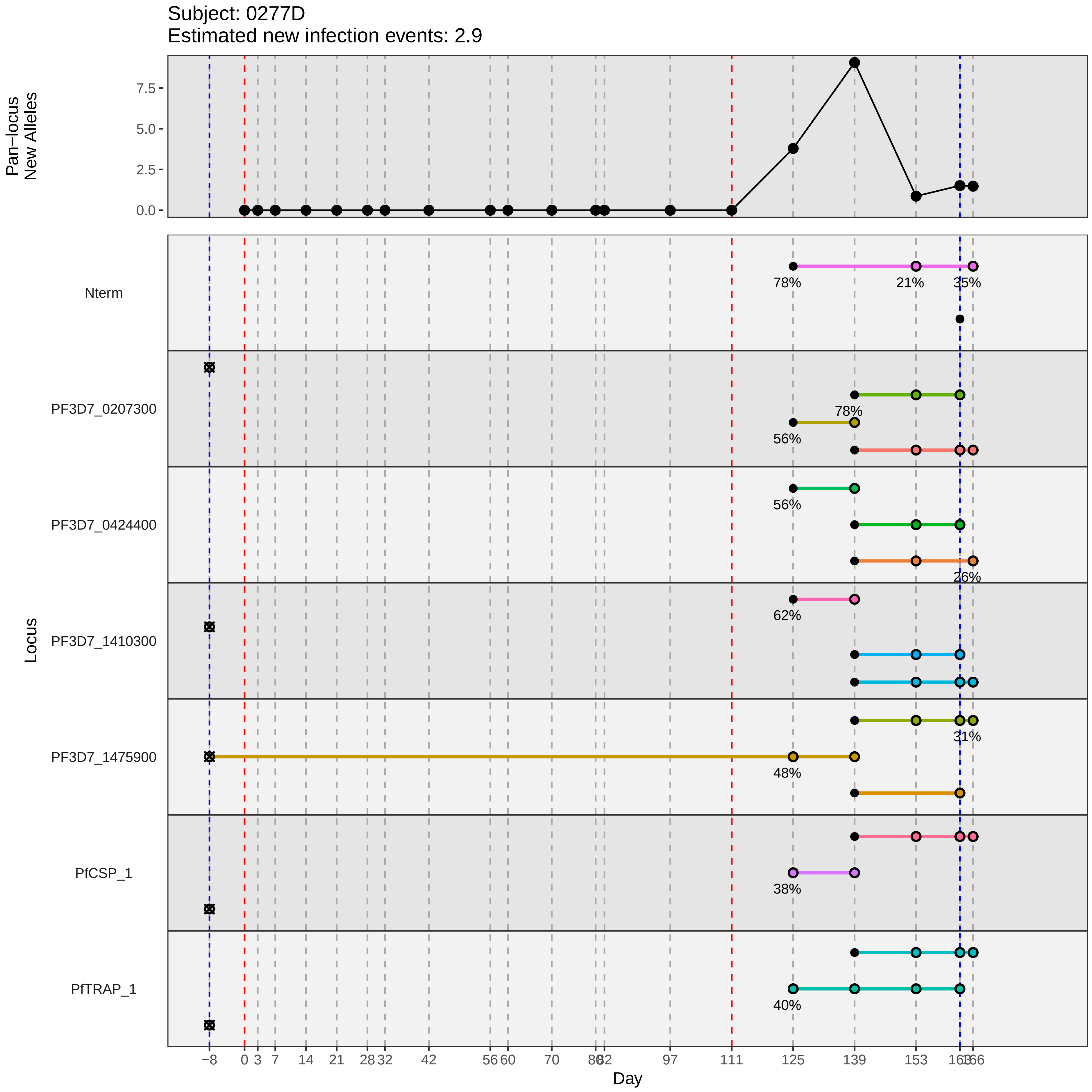

Subject: 0277D
Estimated new infection events: 2.9
Pan−locus New Alleles
7.5
5.0
2.5
0.0
Locus
Nterm
PF3D7_0207300
PF3D7_0424400
PF3D7_1410300
PF3D7_1475900
PfCSP_1
PfTRAP_1
78%
21%
35%
78%
56%
56%
26%
62%
31%
48%
38%
40%
−8
0
3
7
14
21
28
32
42
56
60
70
97
111
125
139
153
Day
