## Supplementary material for "Genotypic infection endpoint analysis to understand efficacy and escape potential of the malaria monoclonal antibody CIS43LS": Data File S2: participant_0278E_without_imputation.pdf

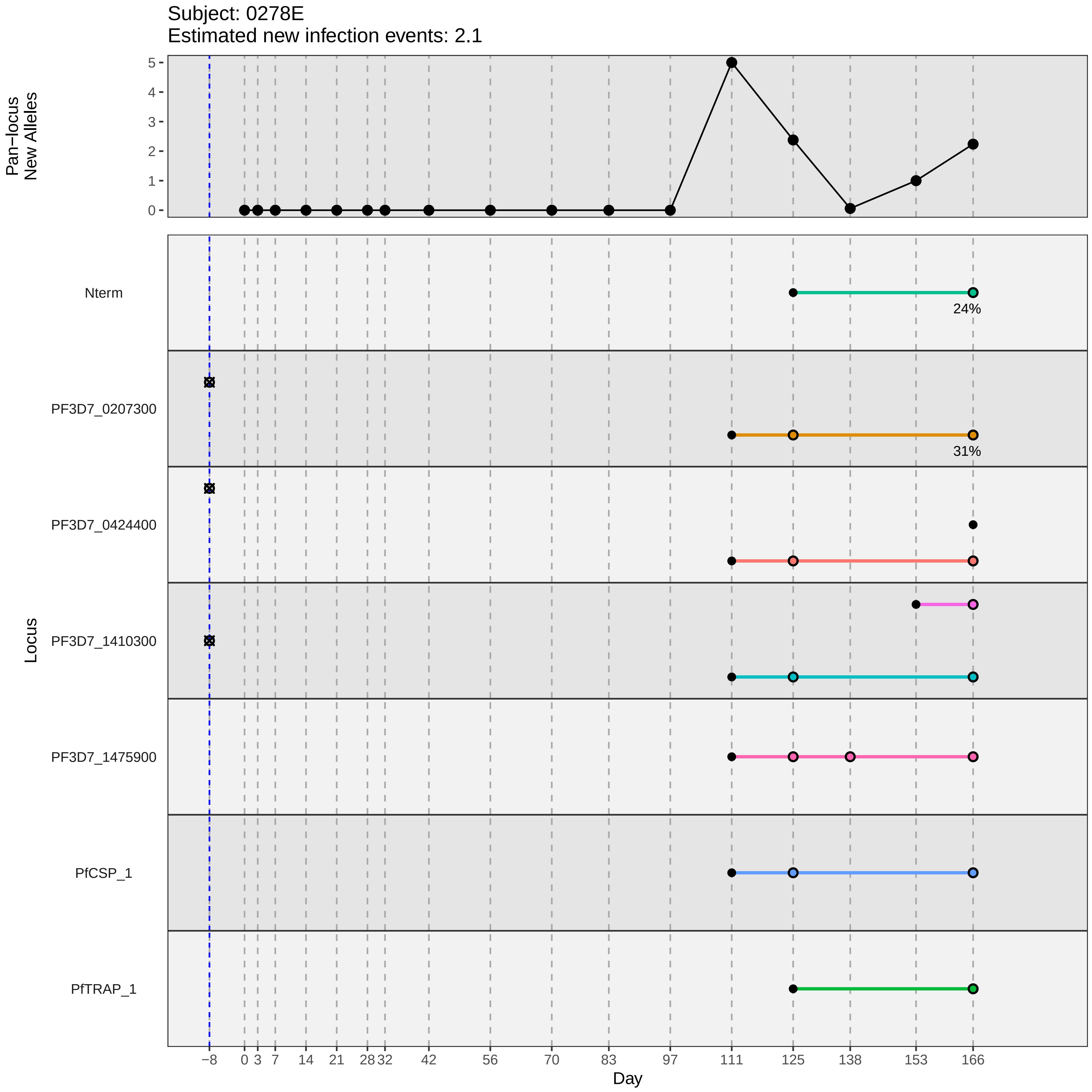

Subject: 0278E
Estimated new infection events: 2.1
Pan−locus
New Alleles
5
4
3
2
1
0
Locus
Nterm
24%
PF3D7_0207300
31%
PF3D7_0424400
PF3D7_1410300
PF3D7_1475900
PfCSP_1
PfTRAP_1
−8
0
3
7
14
21
28
32
42
56
70
83
97
111
125
138
153
166
Day
