## Supplementary material for "Genotypic infection endpoint analysis to understand efficacy and escape potential of the malaria monoclonal antibody CIS43LS": Data File S2: participant_0286Q_without_imputation.pdf

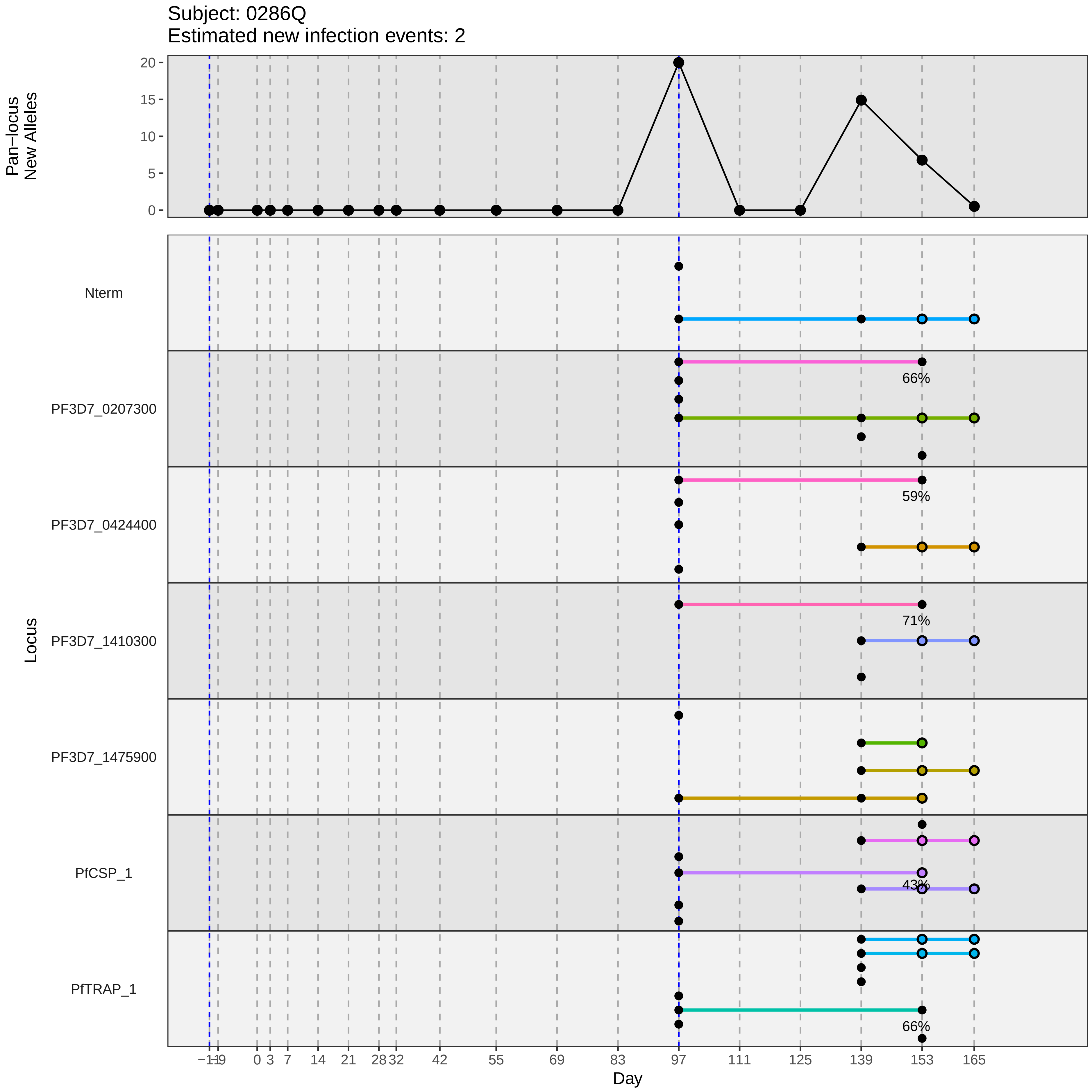

Subject: 0286Q
Estimated new infection events: 2
Pan−locus
New Alleles
20
15
10
5
0
Locus
Nterm
PF3D7_0207300
PF3D7_0424400
PF3D7_1410300
PF3D7_1475900
PfCSP_1
PfTRAP_1
66%
59%
71%
43%
66%
−1
−19
0
3
7
14
21
28
32
42
55
69
83
97
111
125
139
153
165
Day
