## Supplementary material for "Genotypic infection endpoint analysis to understand efficacy and escape potential of the malaria monoclonal antibody CIS43LS": Data File S2: participant_0288S_without_imputation.pdf

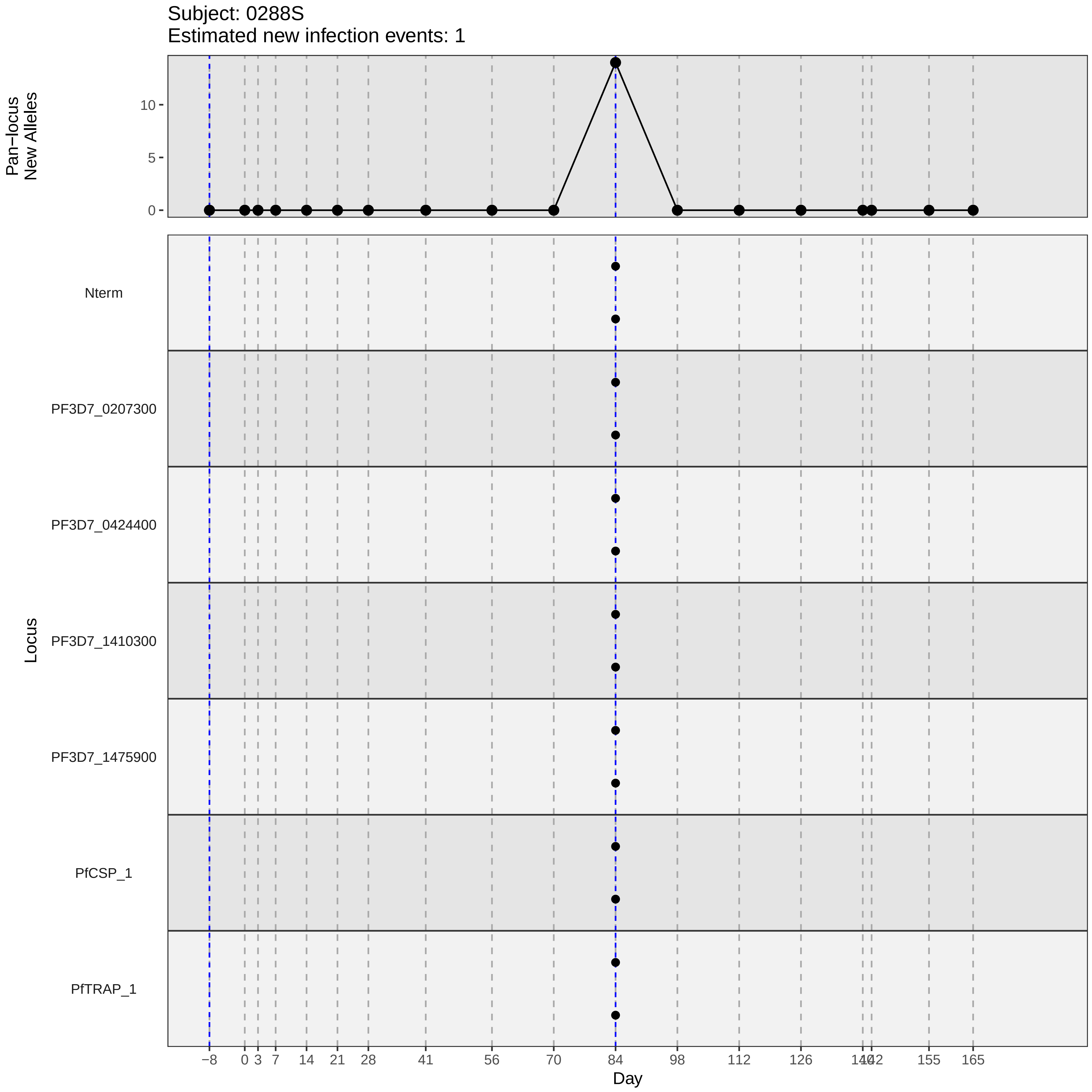

Subject: 0288S
Estimated new infection events: 1
Pan−locus
New Alleles
10
5
0
Locus
Nterm
PF3D7_0207300
PF3D7_0424400
PF3D7_1410300
PF3D7_1475900
PfCSP_1
PfTRAP_1
−8
0
3
7
14
21
28
41
56
70
84
98
112
126
155
165
Day
