## Supplementary material for "Genotypic infection endpoint analysis to understand efficacy and escape potential of the malaria monoclonal antibody CIS43LS": Data File S2: participant_0291V_without_imputation.pdf

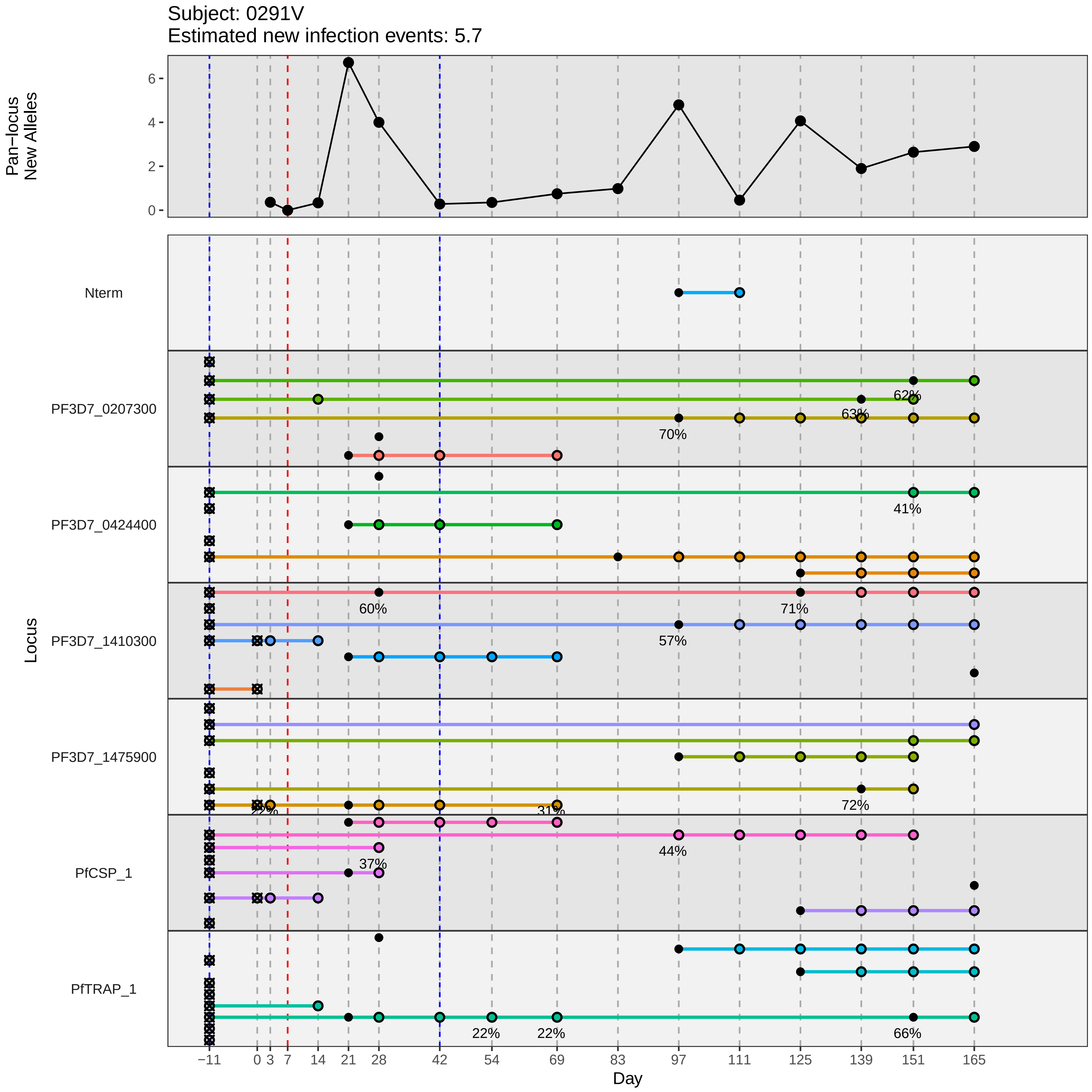

Subject: 0291V
Estimated new infection events: 5.7
Pan−locus New Alleles
0
2
4
6
Locus
Nterm
PF3D7_0207300
PF3D7_0424400
PF3D7_1410300
PF3D7_1475900
PfCSP_1
PfTRAP_1
62%
63%
70%
41%
60%
71%
57%
72%
31%
44%
37%
22%
22%
66%
−11
0
3
7
14
21
28
42
54
69
83
97
111
125
139
151
165
Day
