## Supplementary material for "Genotypic infection endpoint analysis to understand efficacy and escape potential of the malaria monoclonal antibody CIS43LS": Data File S2: participant_0297C_without_imputation.pdf

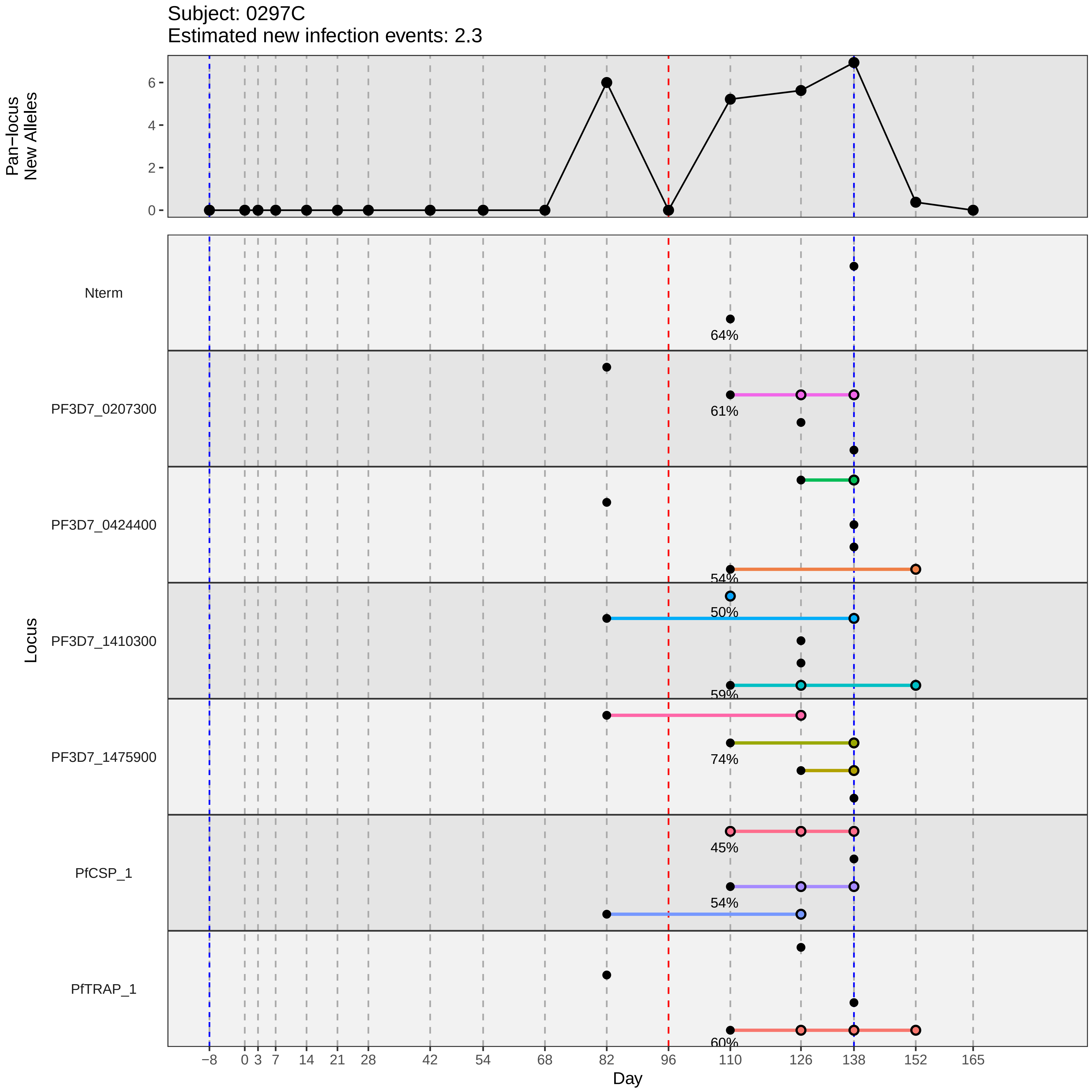

Subject: 0297C
Estimated new infection events: 2.3
Pan−locus
New Alleles
0
2
4
6
Locus
Nterm
PF3D7_0207300
PF3D7_0424400
PF3D7_1410300
PF3D7_1475900
PfCSP_1
PfTRAP_1
64%
61%
54%
50%
59%
74%
45%
54%
60%
−8
0
3
7
14
21
28
42
54
68
82
96
110
126
138
152
165
Day
