## Supplementary material for "Genotypic infection endpoint analysis to understand efficacy and escape potential of the malaria monoclonal antibody CIS43LS": Data File S2: participant_0305N_without_imputation.pdf

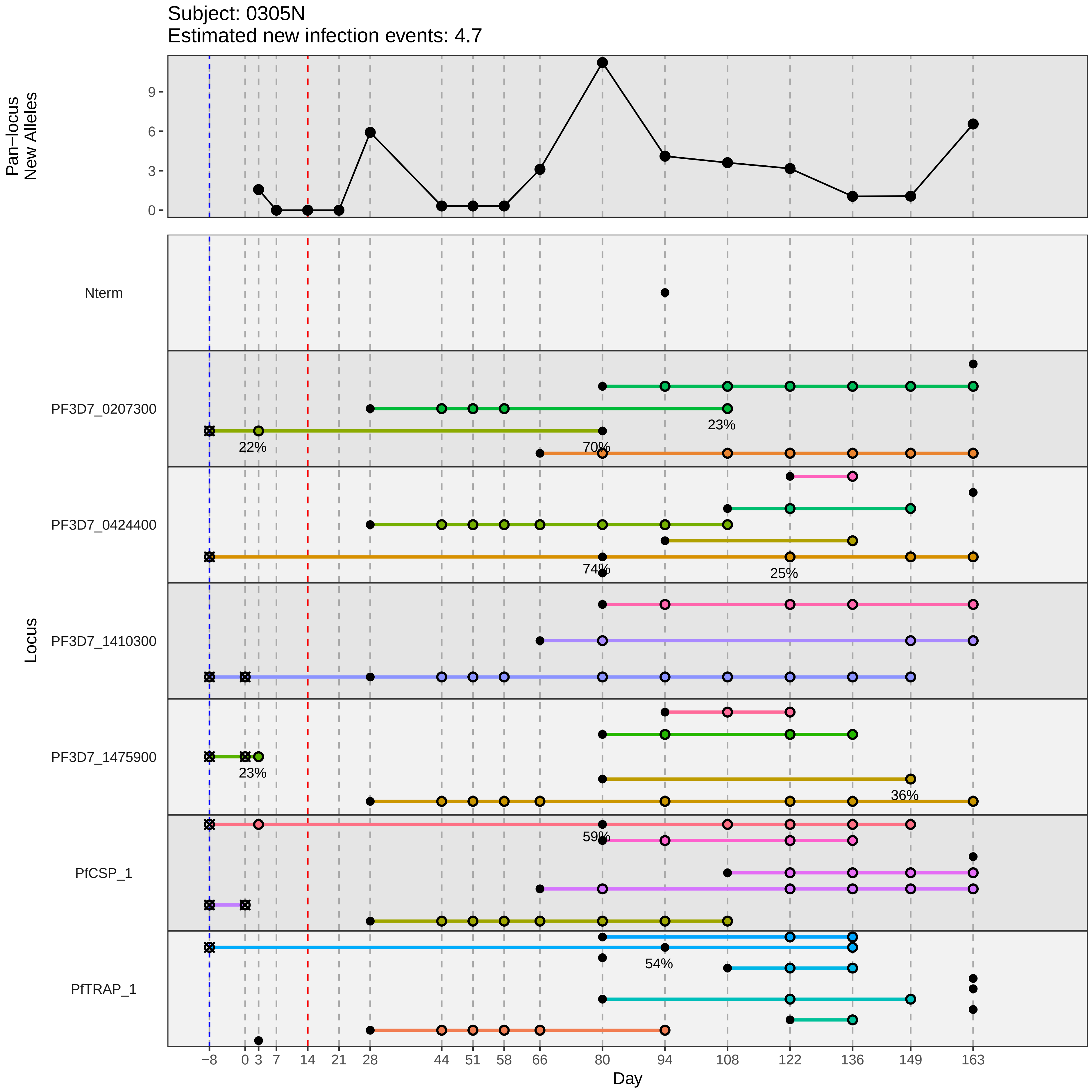

Subject: 0305N
Estimated new infection events: 4.7
Pan−locus
New Alleles
0
3
6
9
Locus
Nterm
PF3D7_0207300
PF3D7_0424400
PF3D7_1410300
PF3D7_1475900
PfCSP_1
PfTRAP_1
22%
70%
23%
74%
25%
23%
36%
59%
54%
−8
0
3
7
14
21
28
44
51
58
66
80
94
108
122
136
149
163
Day
