## Supplementary material for "Genotypic infection endpoint analysis to understand efficacy and escape potential of the malaria monoclonal antibody CIS43LS": Data File S2: participant_0311U_without_imputation.pdf

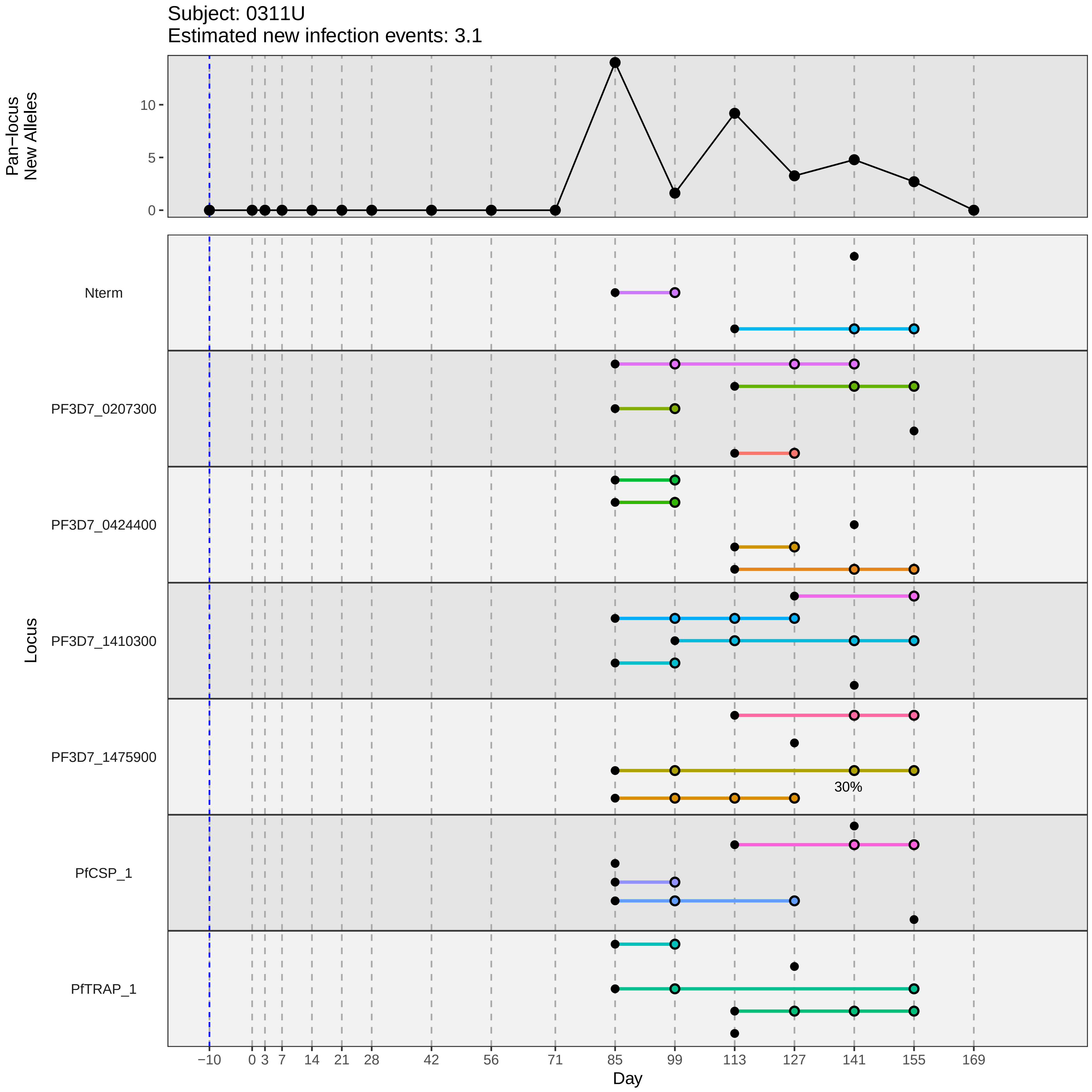
Subject: 0311U
Estimated new infection events: 3.1
Pan−locus New Alleles
Locus
Nterm
PF3D7_0207300
PF3D7_0424400
PF3D7_1410300
PF3D7_1475900
PfCSP_1
PfTRAP_1
30%
Day
−10 0 3 7 14 21 28 42 56 71 85 99 113 127 141 155 169
0 5 10
