## Supplementary material for "Genotypic infection endpoint analysis to understand efficacy and escape potential of the malaria monoclonal antibody CIS43LS": Data File S2: participant_0326N_without_imputation.pdf

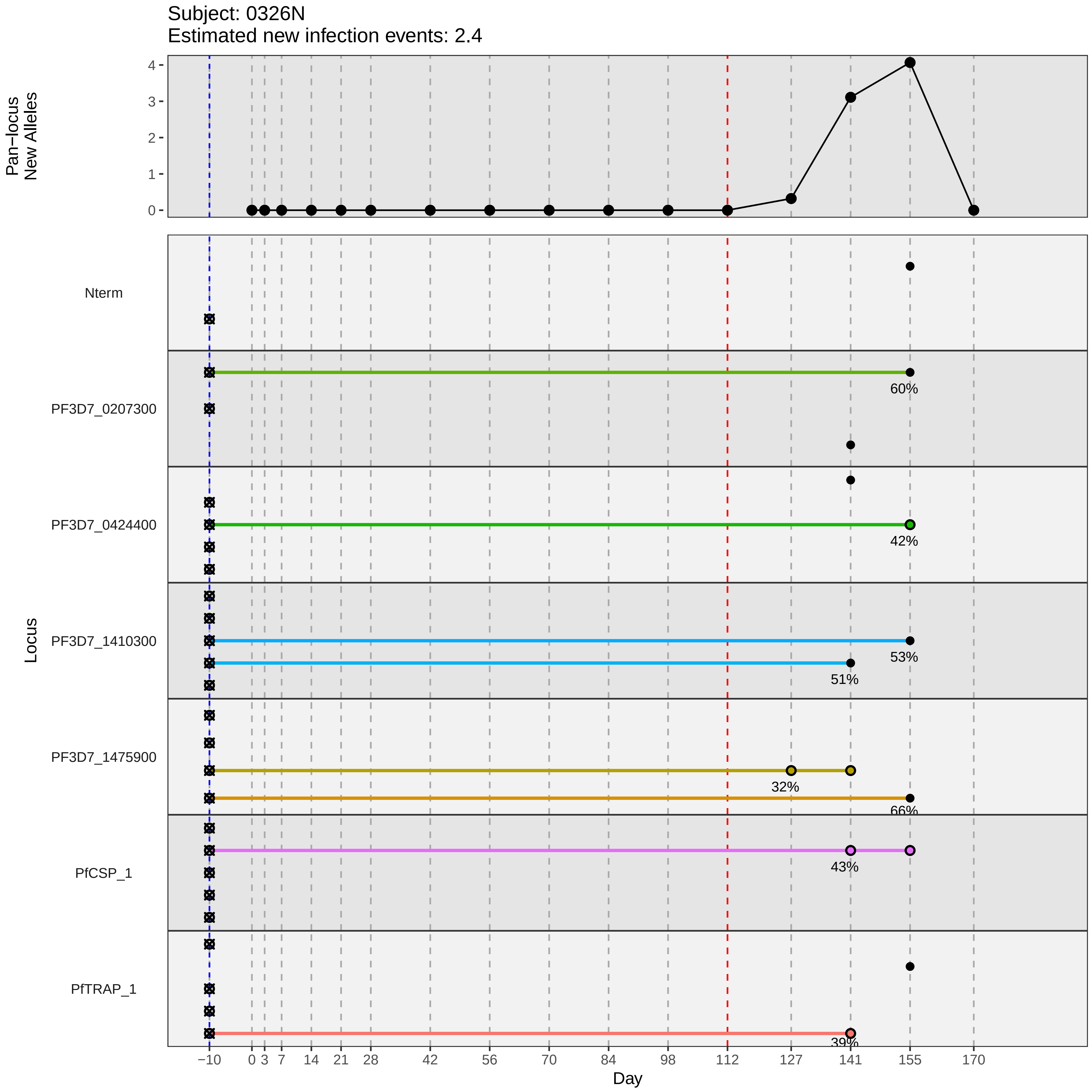
Subject: 0326N
Estimated new infection events: 2.4
Pan−locus
New Alleles
0
1
2
3
4
Locus
Nterm
PF3D7_0207300
PF3D7_0424400
PF3D7_1410300
PF3D7_1475900
PfCSP_1
PfTRAP_1
60%
42%
53%
51%
32%
66%
43%
39%
−10
0
3
7
14
21
28
42
56
70
84
98
112
127
141
155
170
Day
