## Supplementary material for "Genotypic infection endpoint analysis to understand efficacy and escape potential of the malaria monoclonal antibody CIS43LS": Data File S2: participant_0336Y_without_imputation.pdf

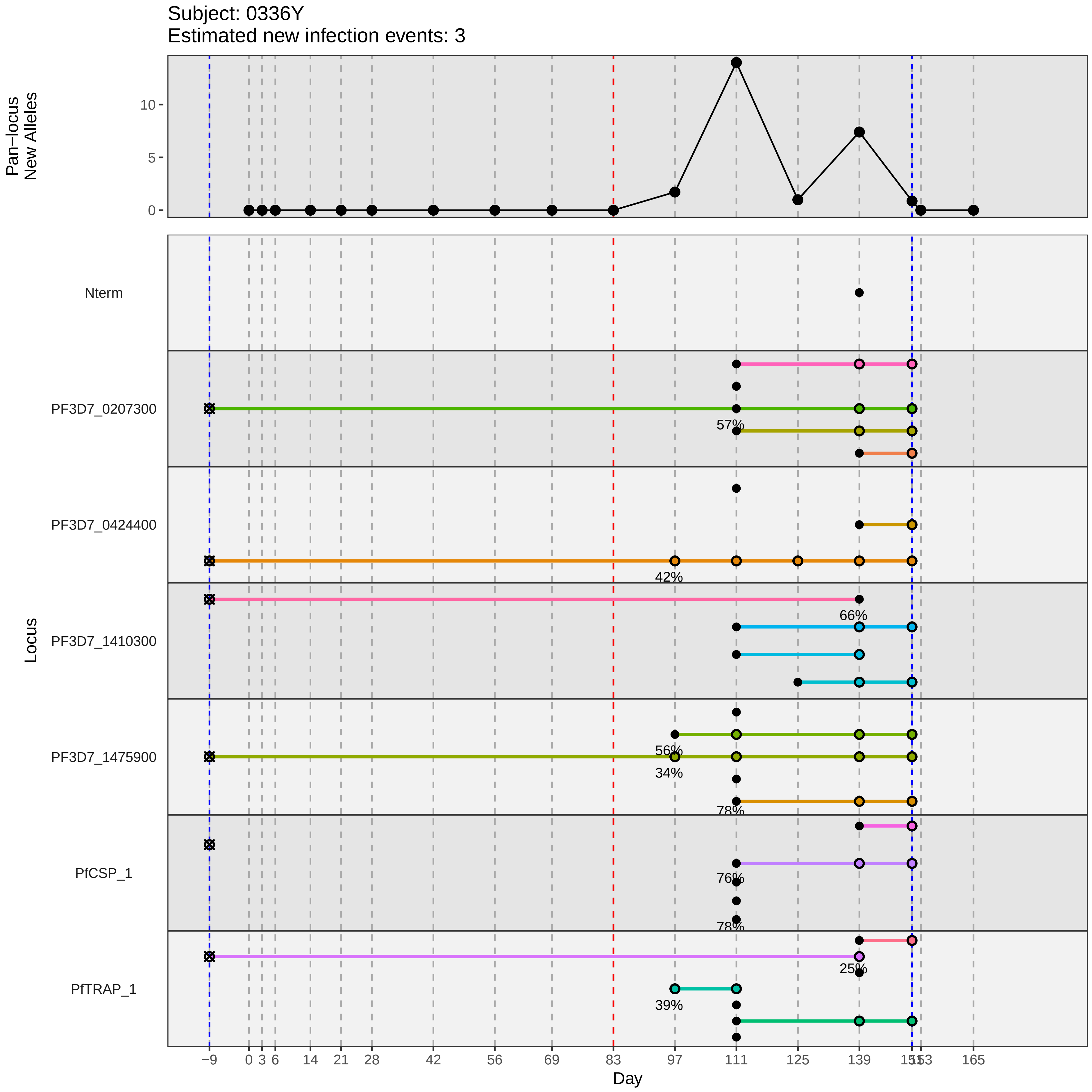
Subject: 0336Y
Estimated new infection events: 3
Pan−locus New Alleles
10
5
0
Locus
Nterm
PF3D7_0207300
57%
PF3D7_0424400
42%
PF3D7_1410300
66%
PF3D7_1475900
56%
34%
78%
PfCSP_1
76%
78%
PfTRAP_1
25%
39%
−9
0
3
6
14
21
28
42
56
69
83
97
111
125
139
165
Day
