## Supplementary material for "Genotypic infection endpoint analysis to understand efficacy and escape potential of the malaria monoclonal antibody CIS43LS": Data File S2: participant_0339C_without_imputation.pdf

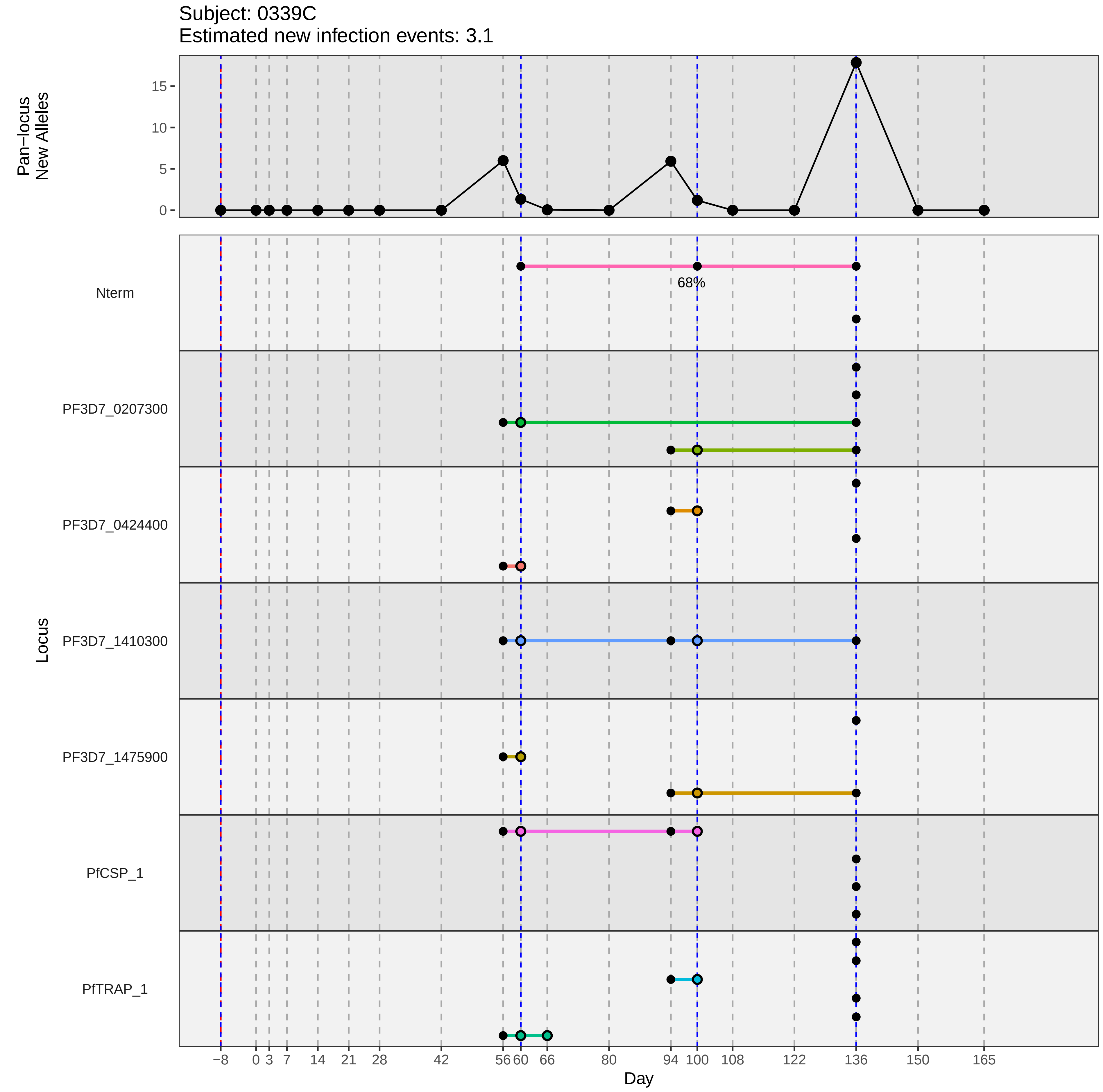
Subject: 0339C
Estimated new infection events: 3.1
Pan−locus
New Alleles
15
10
5
0
Nterm
68%
PF3D7_0207300
PF3D7_0424400
PF3D7_1410300
PF3D7_1475900
PfCSP_1
PfTRAP_1
Locus
−8
0
3
7
14
21
28
42
56
60
66
80
94
100
108
122
136
150
165
Day
