## Supplementary material for "Genotypic infection endpoint analysis to understand efficacy and escape potential of the malaria monoclonal antibody CIS43LS": Data File S2: participant_0342F_without_imputation.pdf

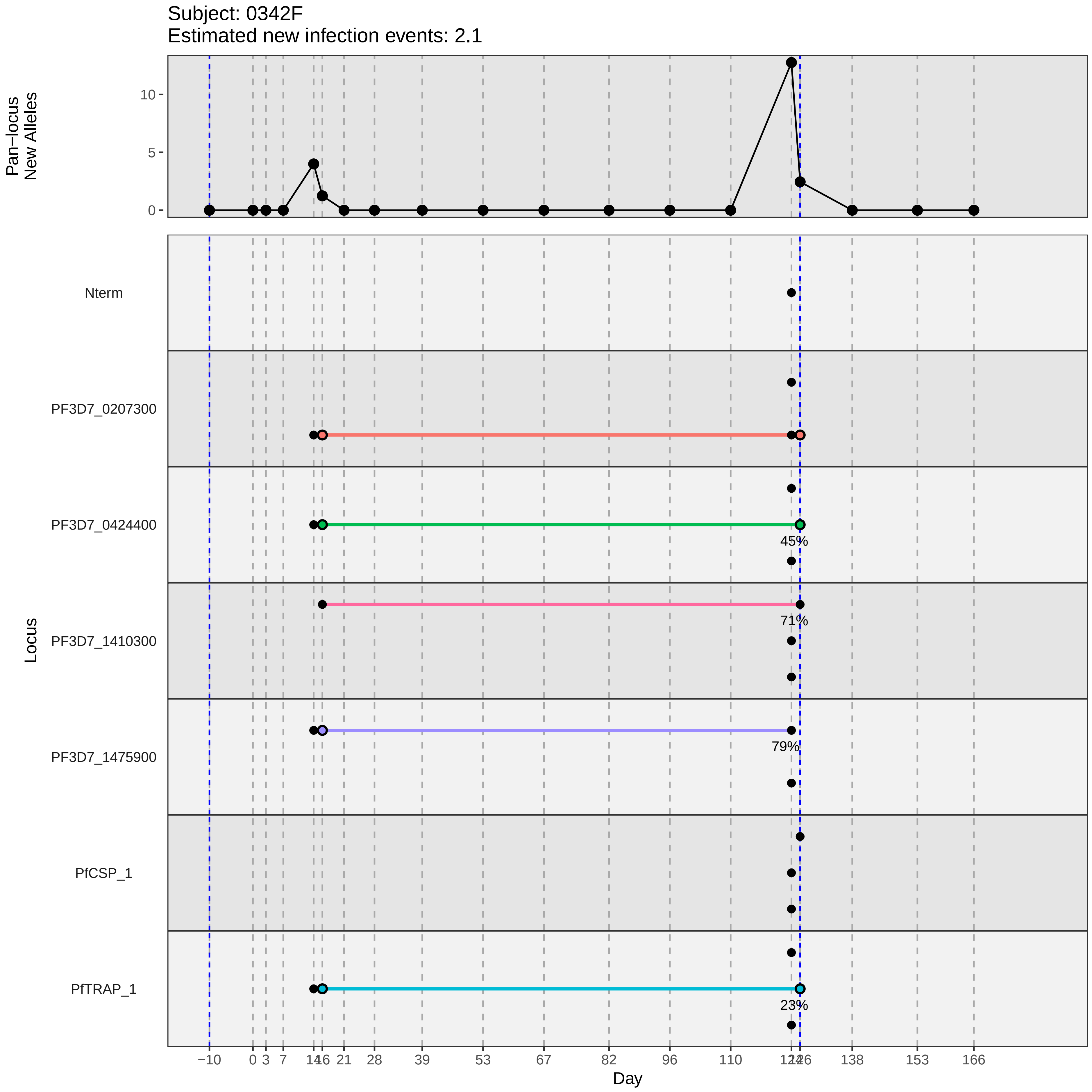

Subject: 0342F
Estimated new infection events: 2.1
Pan−locus New Alleles
10
5
0
Locus
Nterm
PF3D7_0207300
PF3D7_0424400
PF3D7_1410300
PF3D7_1475900
PfCSP_1
PfTRAP_1
45%
71%
79%
23%
−10
0
3
7
14
16
21
28
39
53
67
82
96
110
138
153
166
Day
