## Supplementary material for "Genotypic infection endpoint analysis to understand efficacy and escape potential of the malaria monoclonal antibody CIS43LS": Data File S2: participant_0343G_without_imputation.pdf

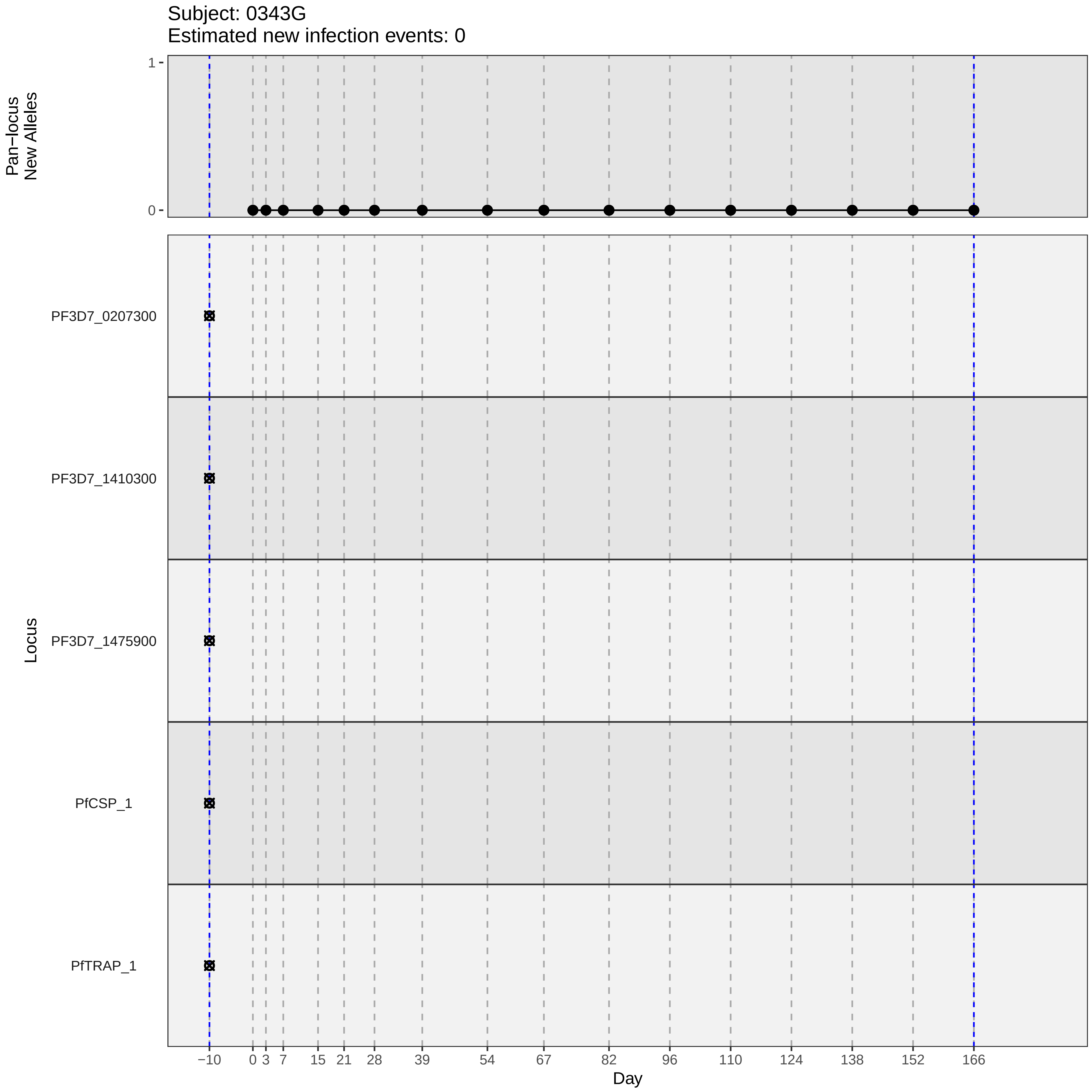
Subject: 0343G
Estimated new infection events: 0
Pan−locus New Alleles
1
0
Locus
PF3D7_0207300
PF3D7_1410300
PF3D7_1475900
PfCSP_1
PfTRAP_1
−10
0
3
7
15
21
28
39
54
67
82
96
110
124
138
152
166
Day
