## Supplementary material for "Genotypic infection endpoint analysis to understand efficacy and escape potential of the malaria monoclonal antibody CIS43LS": Data File S2: participant_0354V_without_imputation.pdf

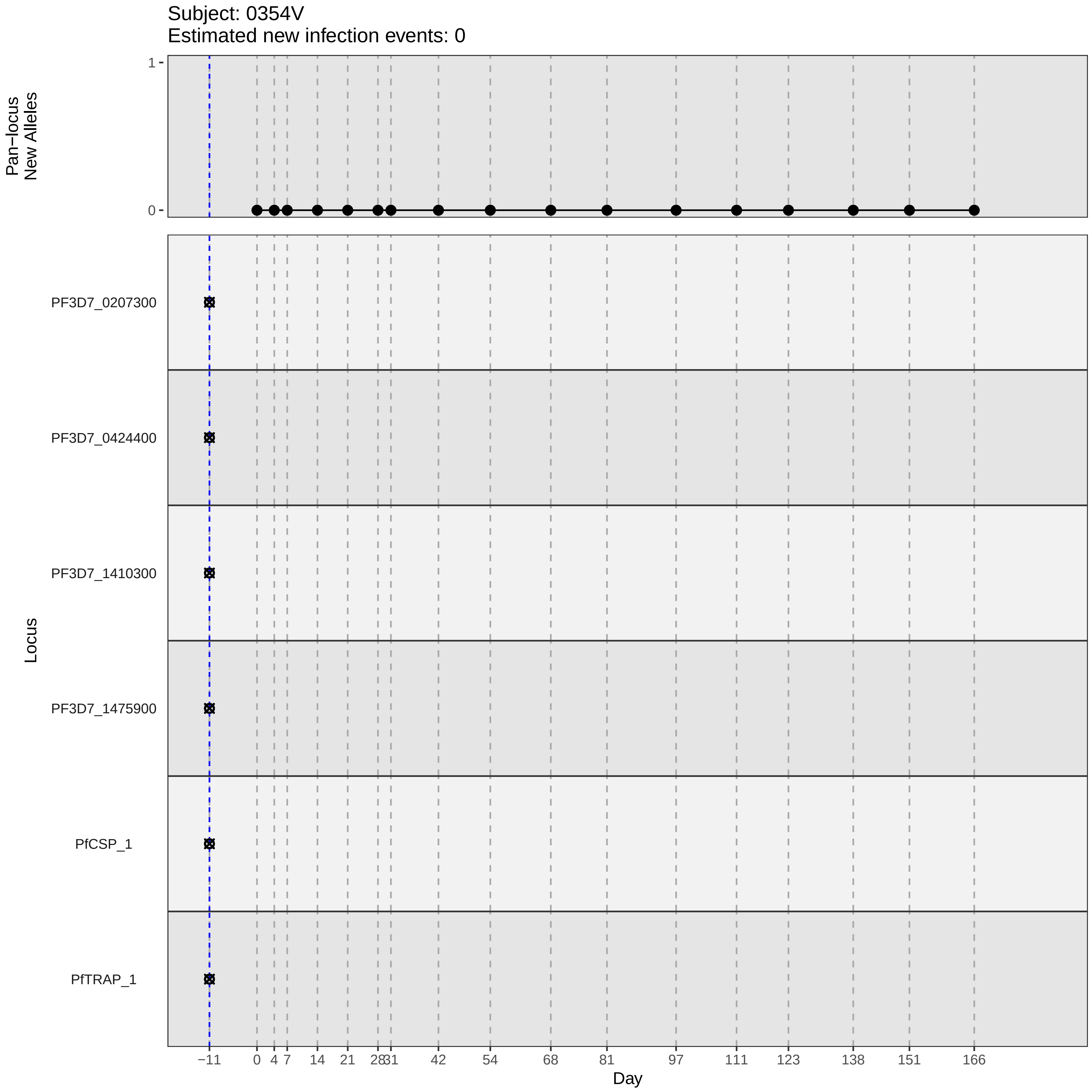
Subject: 0354V
Estimated new infection events: 0
Pan–locus New Alleles
1
0
Locus
PF3D7_0207300
PF3D7_0424400
PF3D7_1410300
PF3D7_1475900
PfCSP_1
PfTRAP_1
–11
0
4
7
14
21
28
31
42
54
68
81
97
111
123
138
151
166
Day
