## Supplementary material for "Genotypic infection endpoint analysis to understand efficacy and escape potential of the malaria monoclonal antibody CIS43LS": Data File S2: participant_0355W_without_imputation.pdf

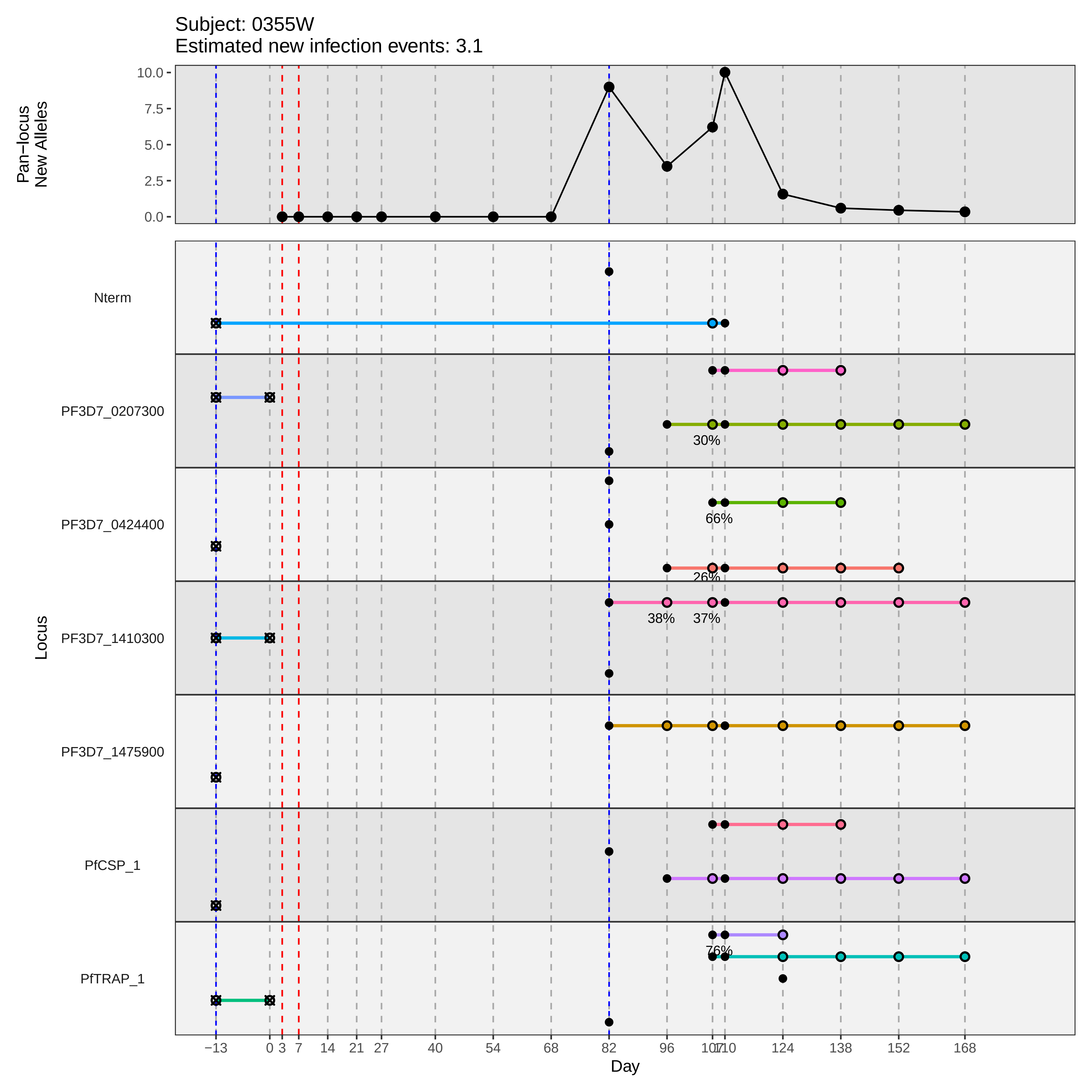

Subject: 0355W
Estimated new infection events: 3.1
Pan−locus New Alleles
10.0
7.5
5.0
2.5
0.0
Locus
Nterm
PF3D7_0207300
PF3D7_0424400
PF3D7_1410300
PF3D7_1475900
PfCSP_1
PfTRAP_1
30%
66%
26%
38%
37%
76%
−13
0
3
7
14
21
27
40
54
68
82
96
107
110
124
138
152
168
Day
