## Supplementary material for "Genotypic infection endpoint analysis to understand efficacy and escape potential of the malaria monoclonal antibody CIS43LS": Data File S2: participant_0358A_without_imputation.pdf

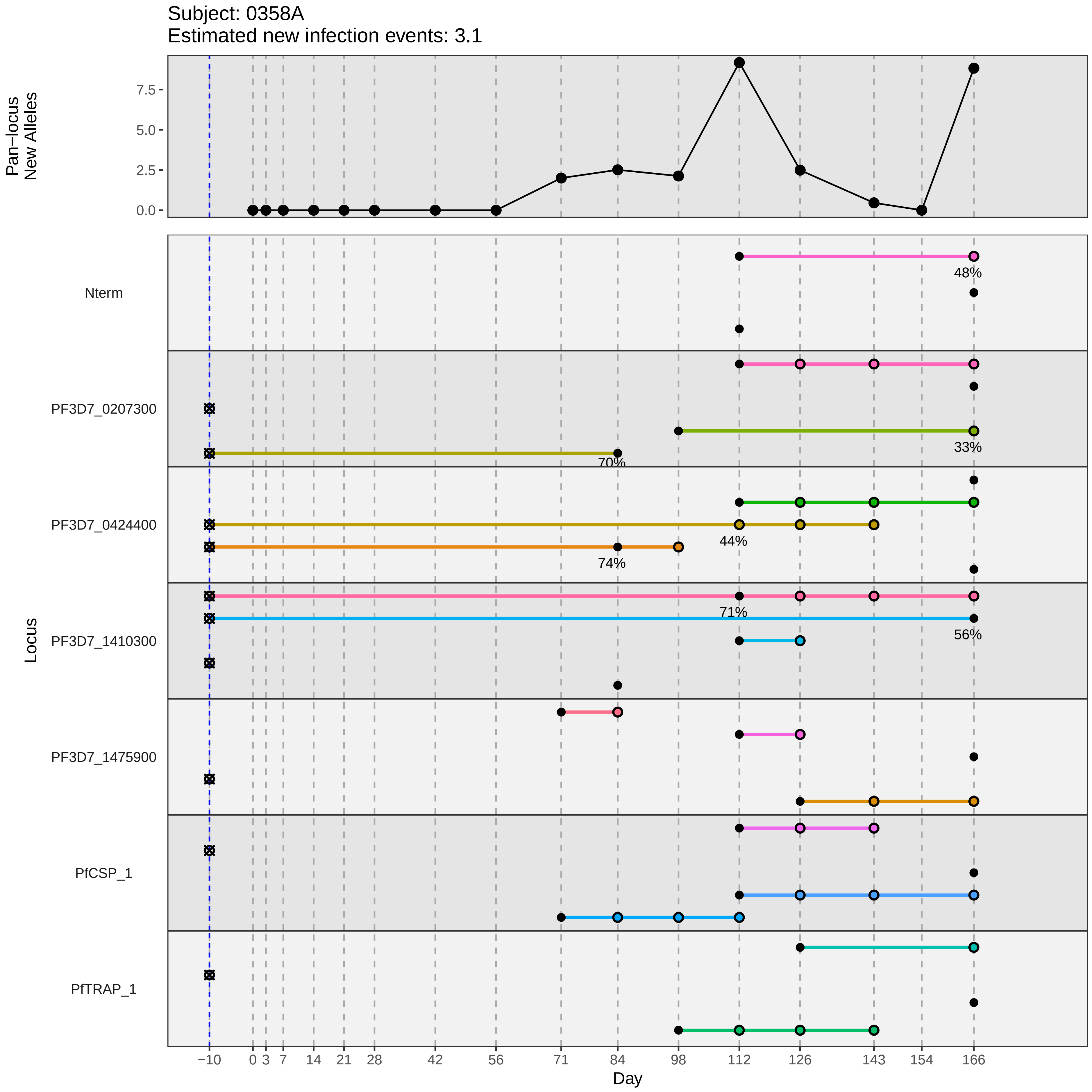

Subject: 0358A
Estimated new infection events: 3.1
Pan−locus
New Alleles
7.5
5.0
2.5
0.0
Nterm
48%
PF3D7_0207300
33%
70%
PF3D7_0424400
44%
74%
PF3D7_1410300
71%
56%
PF3D7_1475900
PfCSP_1
PfTRAP_1
Locus
−10
0
3
7
14
21
28
42
56
71
84
98
112
126
143
154
166
Day
