## Supplementary material for "Genotypic infection endpoint analysis to understand efficacy and escape potential of the malaria monoclonal antibody CIS43LS": Data File S2: participant_0367M_without_imputation.pdf

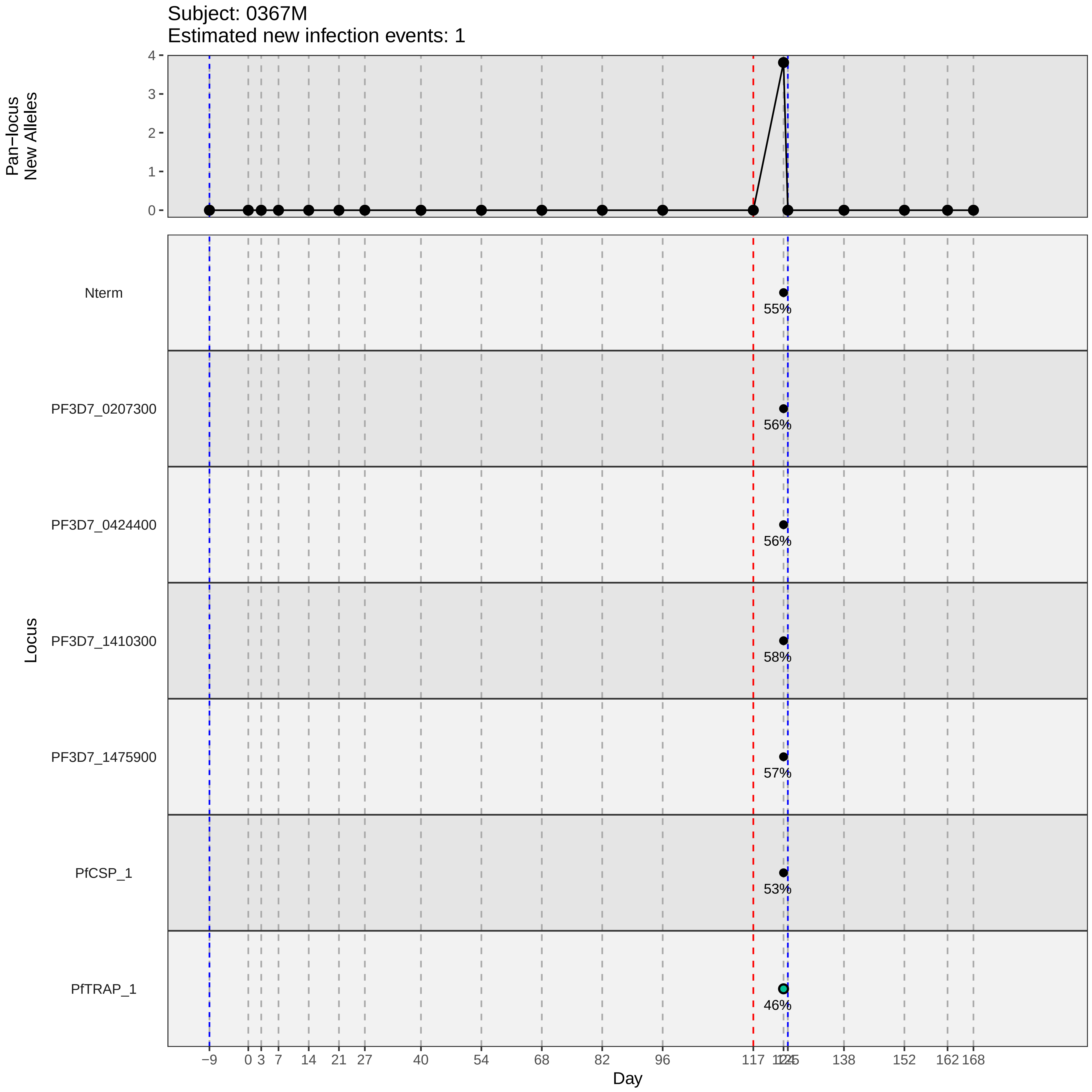

Subject: 0367M
Estimated new infection events: 1
Pan−locus New Alleles
4
3
2
1
0
Locus
Nterm
55%
PF3D7_0207300
56%
PF3D7_0424400
56%
PF3D7_1410300
58%
PF3D7_1475900
57%
PfCSP_1
53%
PfTRAP_1
46%
−9
0
3
7
14
21
27
40
54
68
82
96
117
138
152
162
168
Day
