## Supplementary material for "Genotypic infection endpoint analysis to understand efficacy and escape potential of the malaria monoclonal antibody CIS43LS": Data File S2: participant_0371R_without_imputation.pdf

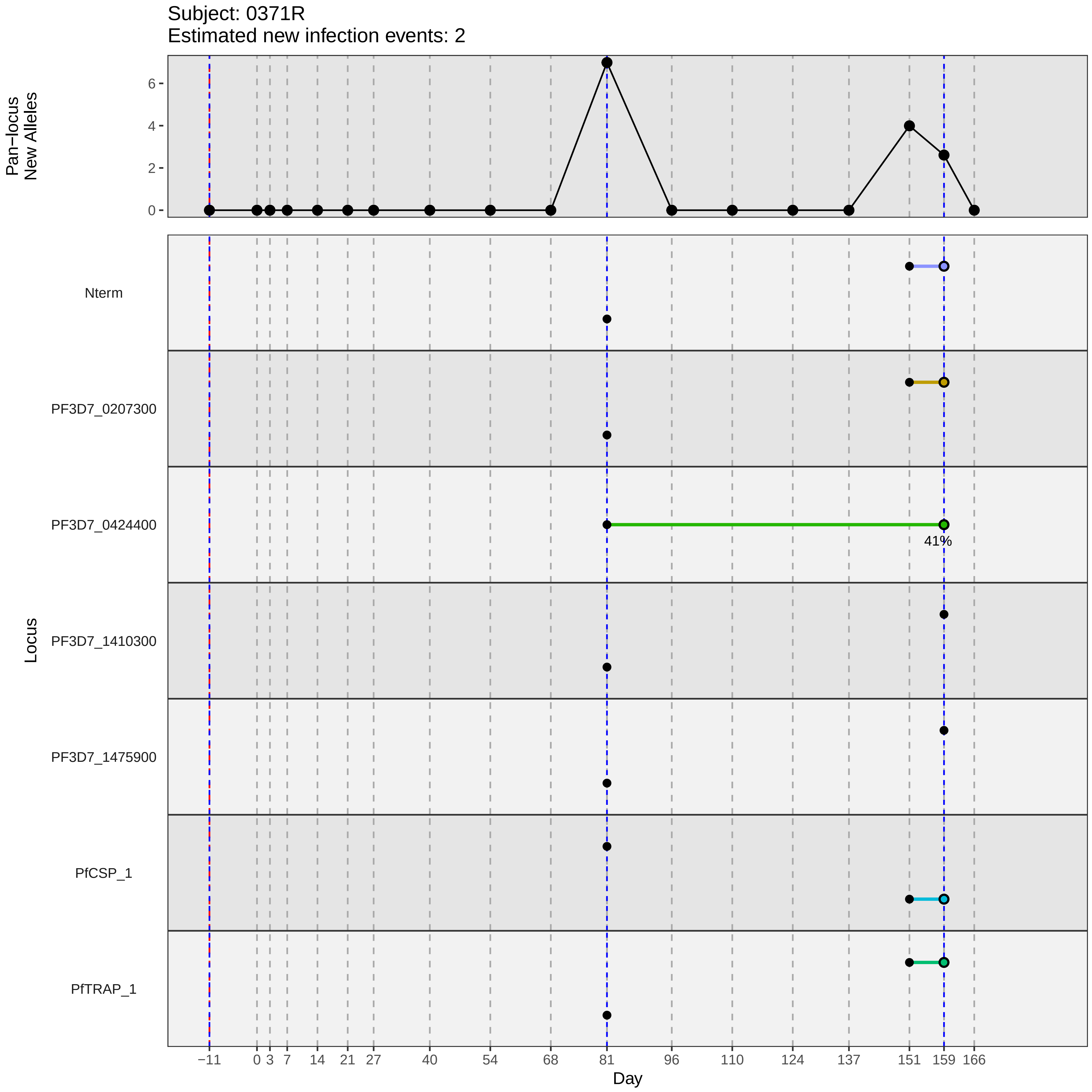

Subject: 0371R
Estimated new infection events: 2
Pan−locus
New Alleles
6
4
2
0
Locus
Nterm
PF3D7_0207300
PF3D7_0424400
41%
PF3D7_1410300
PF3D7_1475900
PfCSP_1
PfTRAP_1
−11
0
3
7
14
21
27
40
54
68
81
96
110
124
137
151
159
166
Day
