## Supplementary material for "Genotypic infection endpoint analysis to understand efficacy and escape potential of the malaria monoclonal antibody CIS43LS": Data File S2: participant_0372S_without_imputation.pdf

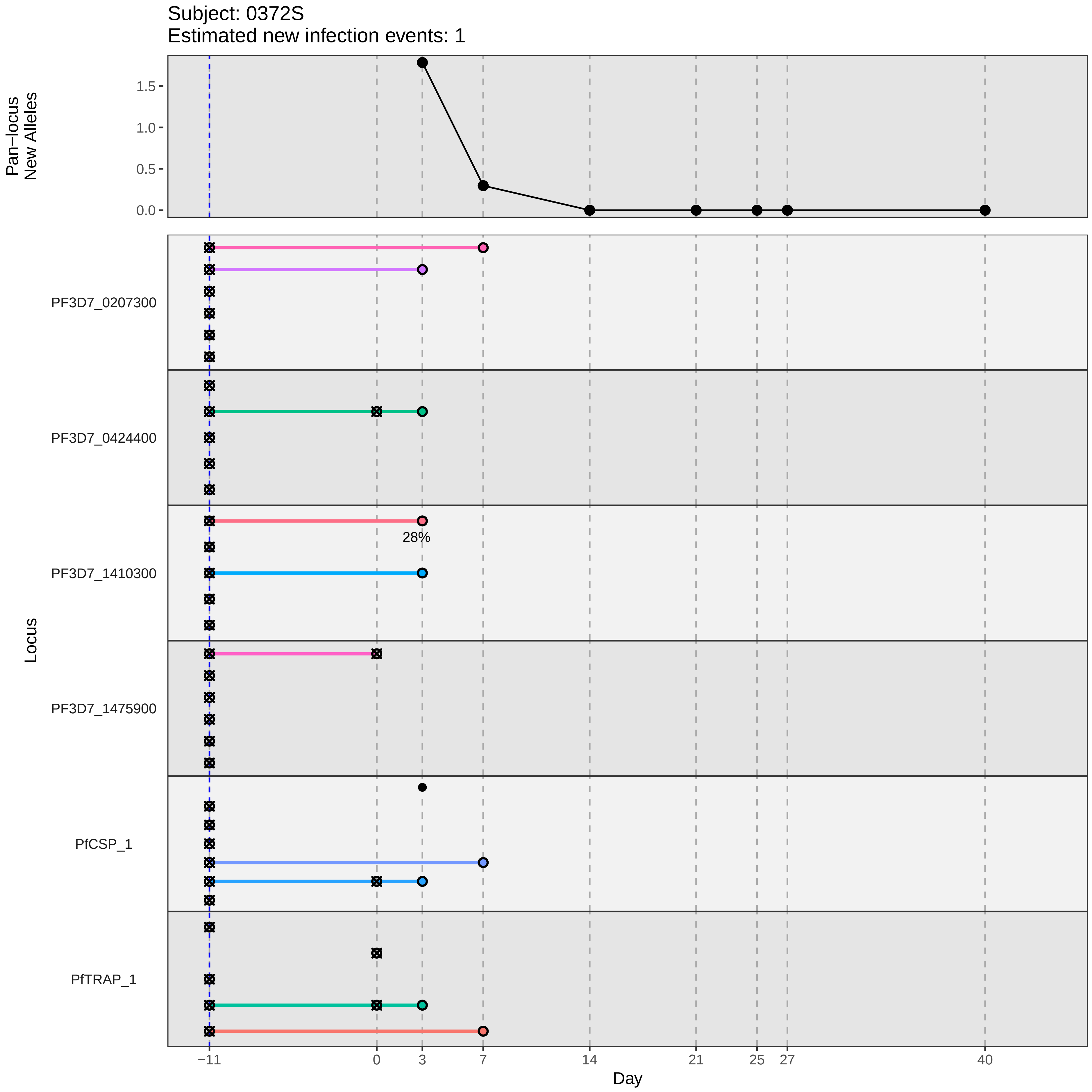

Subject: 0372S
Estimated new infection events: 1
Pan−locus New Alleles
1.5
1.0
0.5
0.0
Locus
PF3D7_0207300
PF3D7_0424400
PF3D7_1410300
28%
PF3D7_1475900
PfCSP_1
PfTRAP_1
−11
0
3
7
14
21
25
27
40
Day
