## Supplementary material for "Genotypic infection endpoint analysis to understand efficacy and escape potential of the malaria monoclonal antibody CIS43LS": Data File S2: participant_0375V_without_imputation.pdf

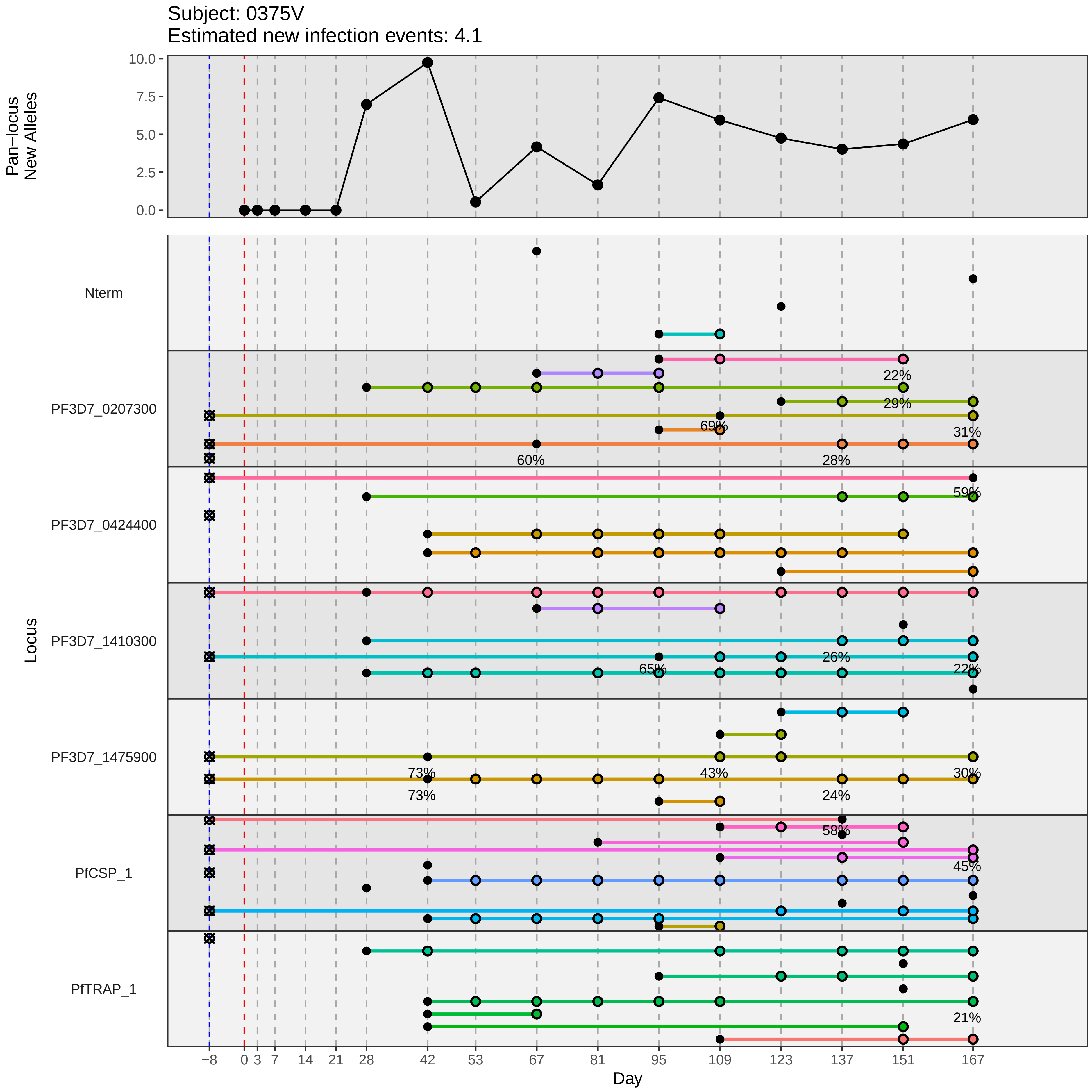

Subject: 0375V
Estimated new infection events: 4.1
Pan−locus New Alleles
10.0
7.5
5.0
2.5
0.0
Locus
Nterm
PF3D7_0207300
PF3D7_0424400
PF3D7_1410300
PF3D7_1475900
PfCSP_1
PfTRAP_1
22%
29%
69%
31%
60%
28%
59%
26%
65%
22%
73%
43%
30%
73%
24%
58%
45%
21%
−8
0
3
7
14
21
28
42
53
67
81
95
109
123
137
151
167
Day
