## Supplementary material for "Genotypic infection endpoint analysis to understand efficacy and escape potential of the malaria monoclonal antibody CIS43LS": Data File S2: participant_0377X_without_imputation.pdf

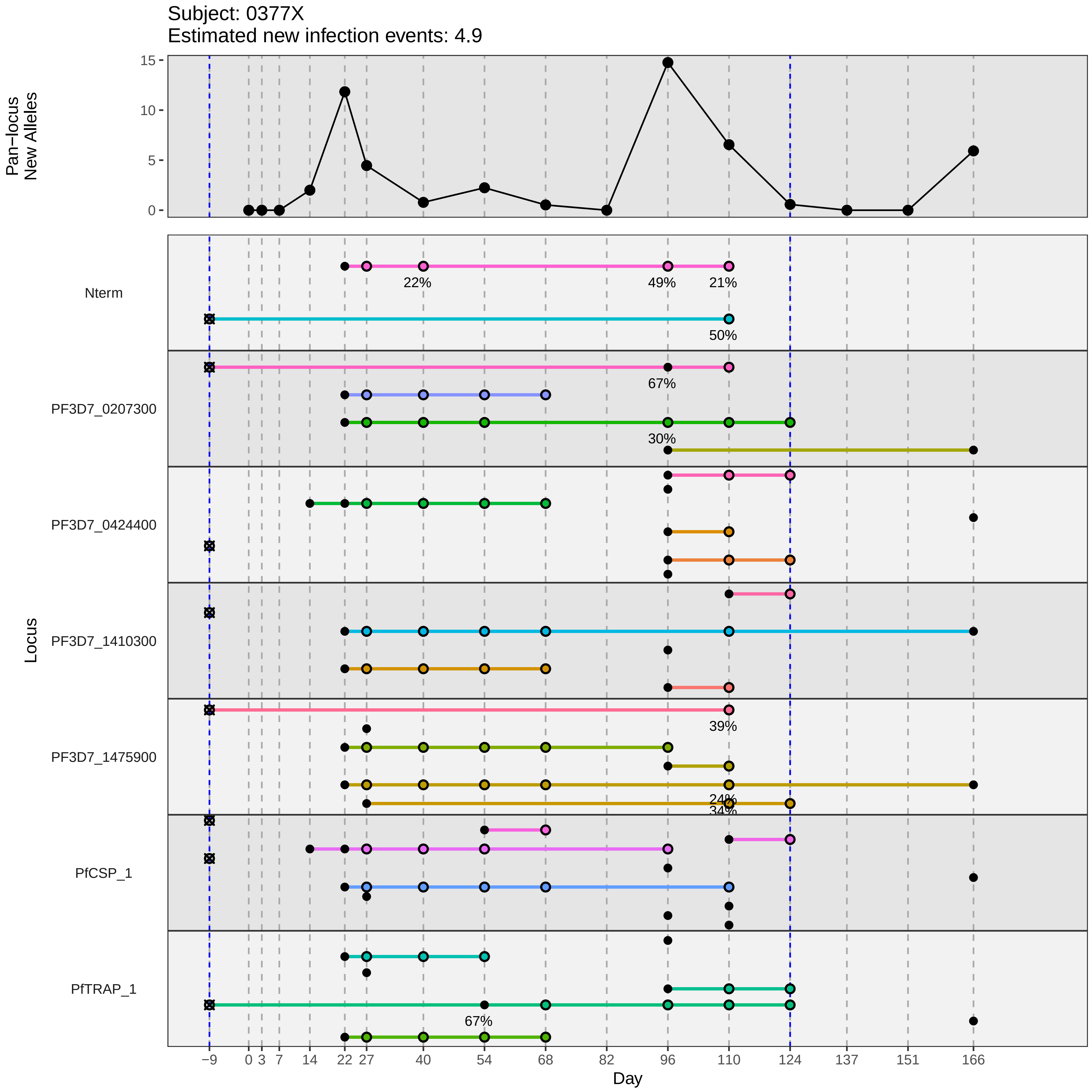

Subject: 0377X
Estimated new infection events: 4.9
Pan−locus New Alleles
Locus
Nterm
PF3D7_0207300
PF3D7_0424400
PF3D7_1410300
PF3D7_1475900
PfCSP_1
PfTRAP_1
22%
49%
21%
50%
67%
30%
39%
24%
34%
67%
Day
−9
0
3
7
14
22
27
40
54
68
82
96
110
124
137
151
166
0
5
10
15
