## Supplementary material for "Genotypic infection endpoint analysis to understand efficacy and escape potential of the malaria monoclonal antibody CIS43LS": Data File S2: participant_0383E_without_imputation.pdf

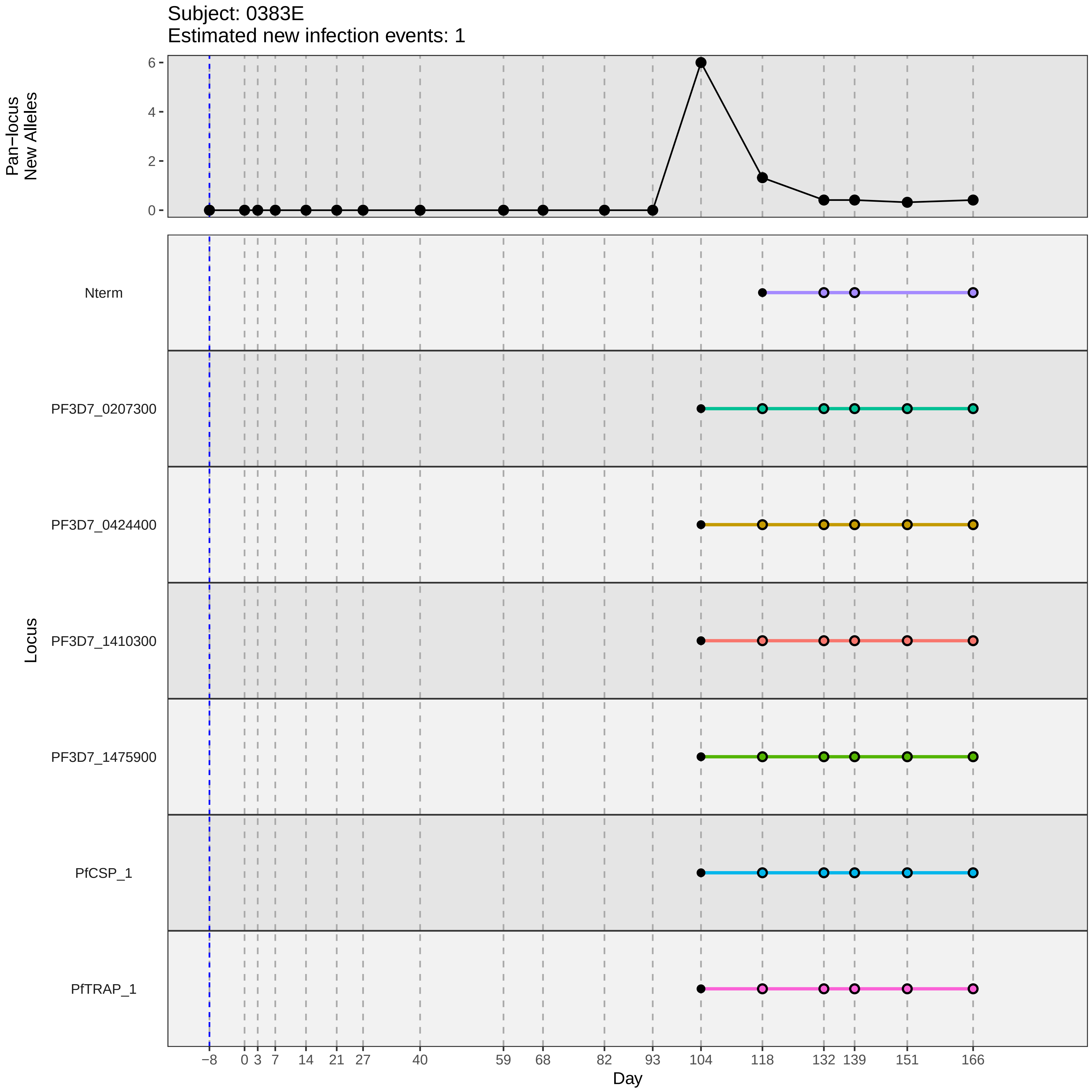

Subject: 0383E
Estimated new infection events: 1
Pan−locus
New Alleles
0
2
4
6
Nterm
PF3D7_0207300
PF3D7_0424400
PF3D7_1410300
PF3D7_1475900
PfCSP_1
PfTRAP_1
Locus
−8
0
3
7
14
21
27
40
59
68
82
93
104
118
132
139
151
166
Day
