## Supplementary material for "Genotypic infection endpoint analysis to understand efficacy and escape potential of the malaria monoclonal antibody CIS43LS": Data File S2: participant_0384F_without_imputation.pdf

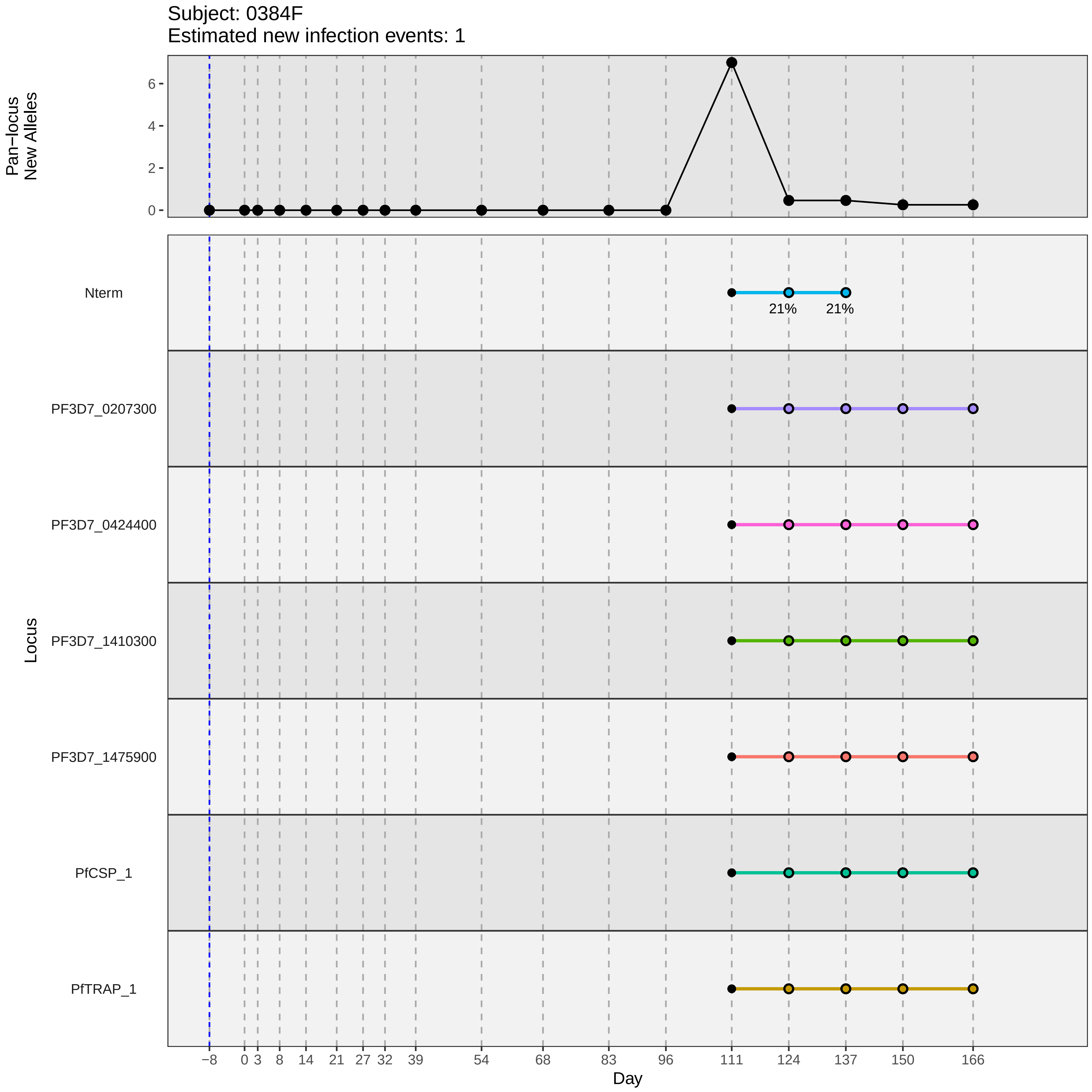

Subject: 0384F
Estimated new infection events: 1
Pan−locus
New Alleles
6
4
2
0
Locus
Nterm
21%
21%
PF3D7_0207300
PF3D7_0424400
PF3D7_1410300
PF3D7_1475900
PfCSP_1
PfTRAP_1
−8
0
3
8
14
21
27
32
39
54
68
83
96
111
124
137
150
166
Day
