## Supplementary material for "Genotypic infection endpoint analysis to understand efficacy and escape potential of the malaria monoclonal antibody CIS43LS": Data File S2: participant_0391Q_without_imputation.pdf

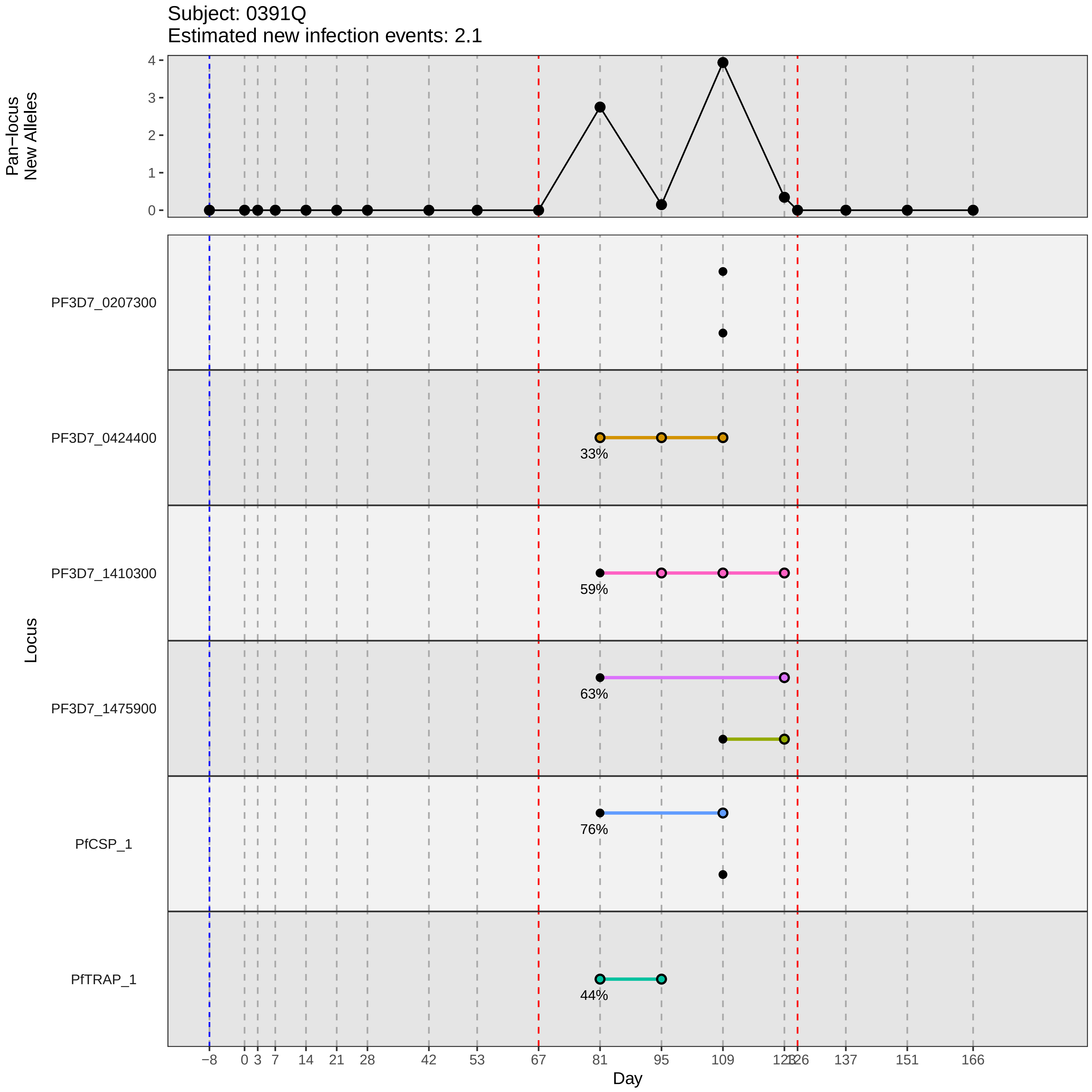

Subject: 0391Q
Estimated new infection events: 2.1
Pan−locus
New Alleles
Locus
PF3D7_0207300
PF3D7_0424400
PF3D7_1410300
PF3D7_1475900
PfCSP_1
PfTRAP_1
33%
59%
63%
76%
44%
Day
−8
0
3
7
14
21
28
42
53
67
81
95
109
123
126
137
151
166
