## Supplementary material for "Genotypic infection endpoint analysis to understand efficacy and escape potential of the malaria monoclonal antibody CIS43LS": Data File S2: participant_0392R_without_imputation.pdf

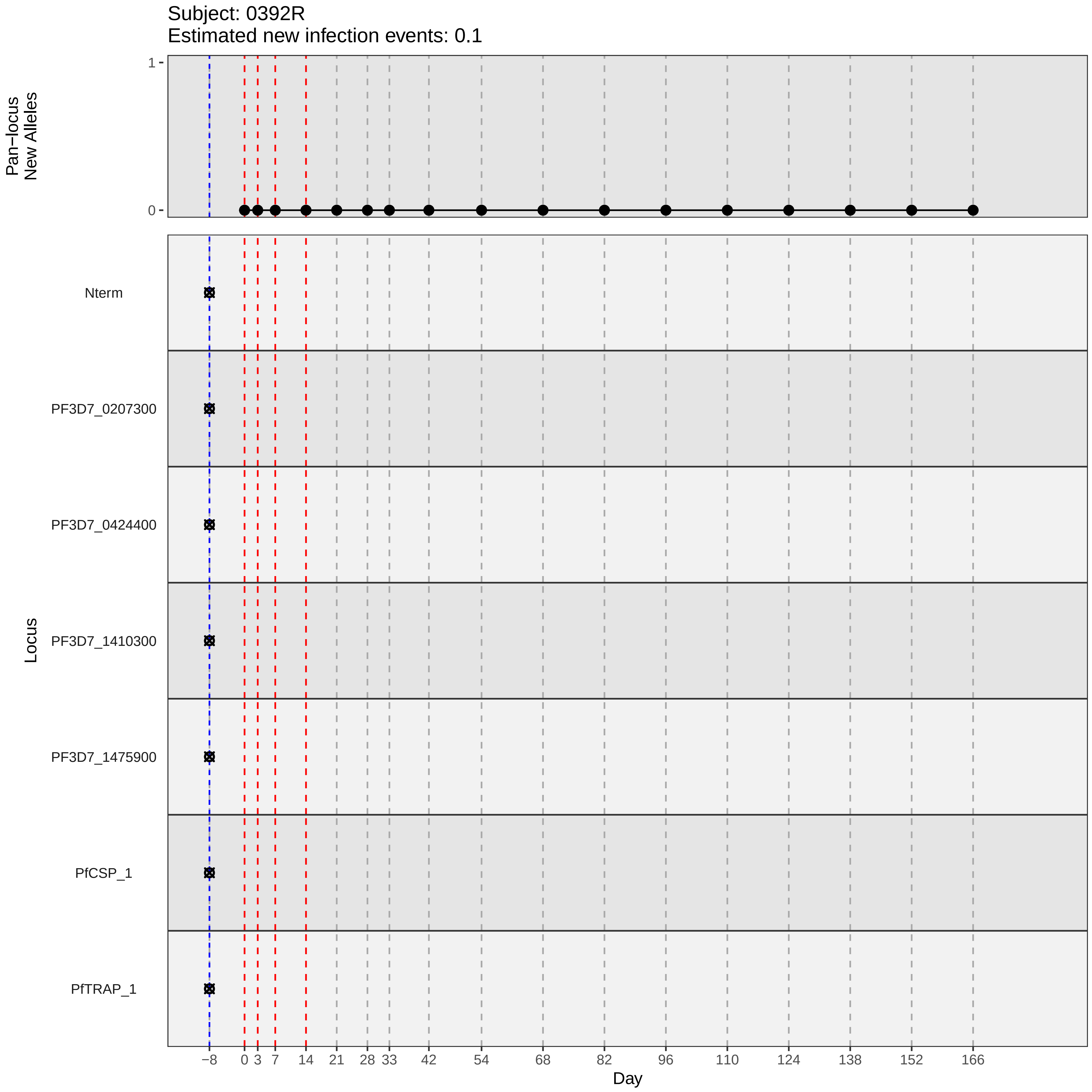

Subject: 0392R
Estimated new infection events: 0.1
Pan−locus
New Alleles
1
0
Locus
Nterm
PF3D7_0207300
PF3D7_0424400
PF3D7_1410300
PF3D7_1475900
PfCSP_1
PfTRAP_1
−8
0
3
7
14
21
28
33
42
54
68
82
96
110
124
138
152
166
Day
