## Supplementary material for "Genotypic infection endpoint analysis to understand efficacy and escape potential of the malaria monoclonal antibody CIS43LS": Data File S2: participant_0398X_without_imputation.pdf

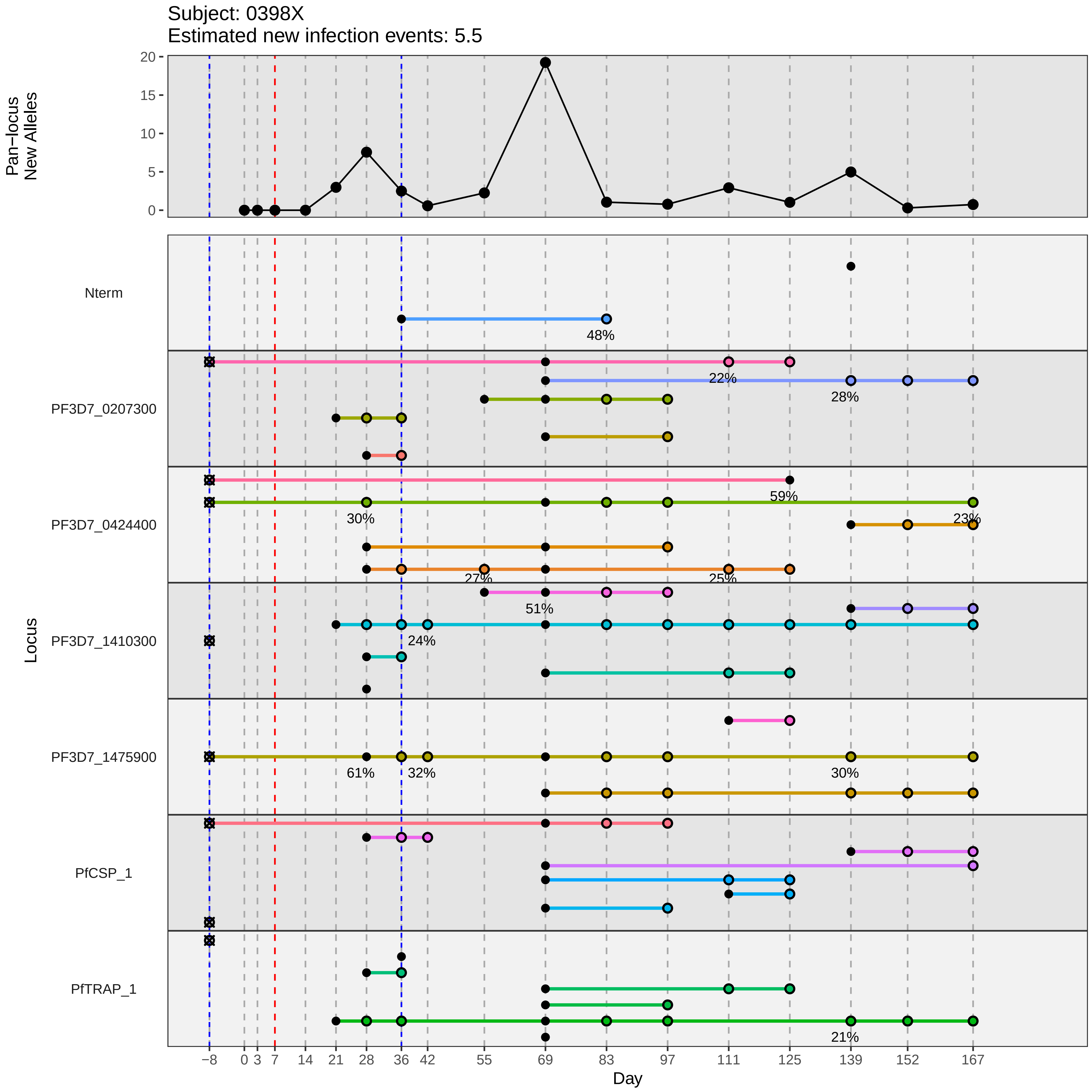
Subject: 0398X
Estimated new infection events: 5.5
Pan−locus New Alleles
0
5
10
15
20
Locus
Nterm
PF3D7_0207300
PF3D7_0424400
PF3D7_1410300
PF3D7_1475900
PfCSP_1
PfTRAP_1
48%
22%
28%
59%
30%
23%
27%
25%
51%
24%
61%
32%
30%
21%
−8
0
3
7
14
21
28
36
42
55
69
83
97
111
125
139
152
167
Day
