## Supplementary material for "Genotypic infection endpoint analysis to understand efficacy and escape potential of the malaria monoclonal antibody CIS43LS": Data File S2: participant_0402C_without_imputation.pdf

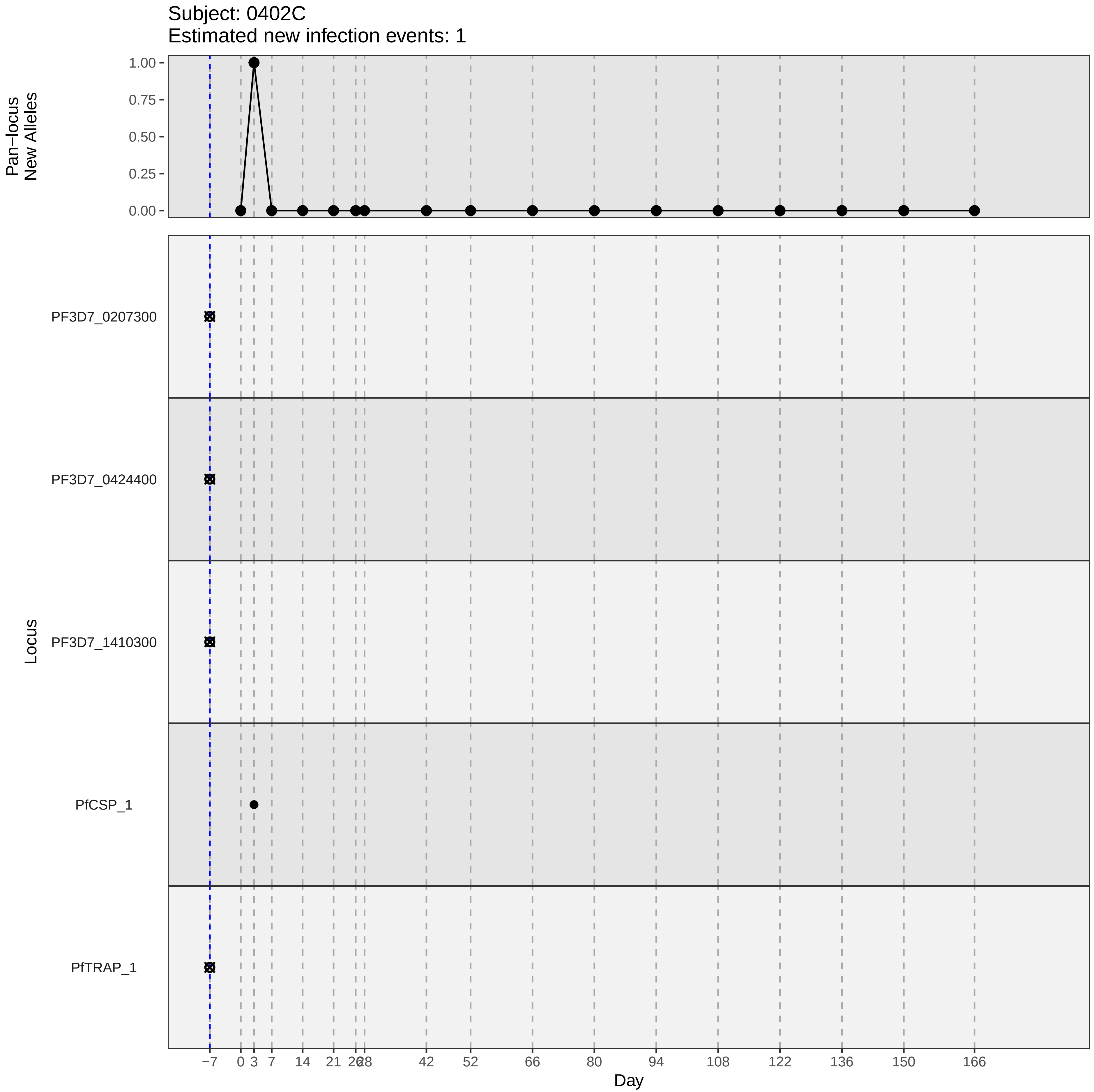

Subject: 0402C
Estimated new infection events: 1
Pan−locus New Alleles
1.00
0.75
0.50
0.25
0.00
Locus
PF3D7_0207300
PF3D7_0424400
PF3D7_1410300
PfCSP_1
PfTRAP_1
−7
0
3
7
14
21
26
28
42
52
66
80
94
108
122
136
150
166
Day
