## Supplementary material for "Genotypic infection endpoint analysis to understand efficacy and escape potential of the malaria monoclonal antibody CIS43LS": Data File S2: participant_0407H_without_imputation.pdf

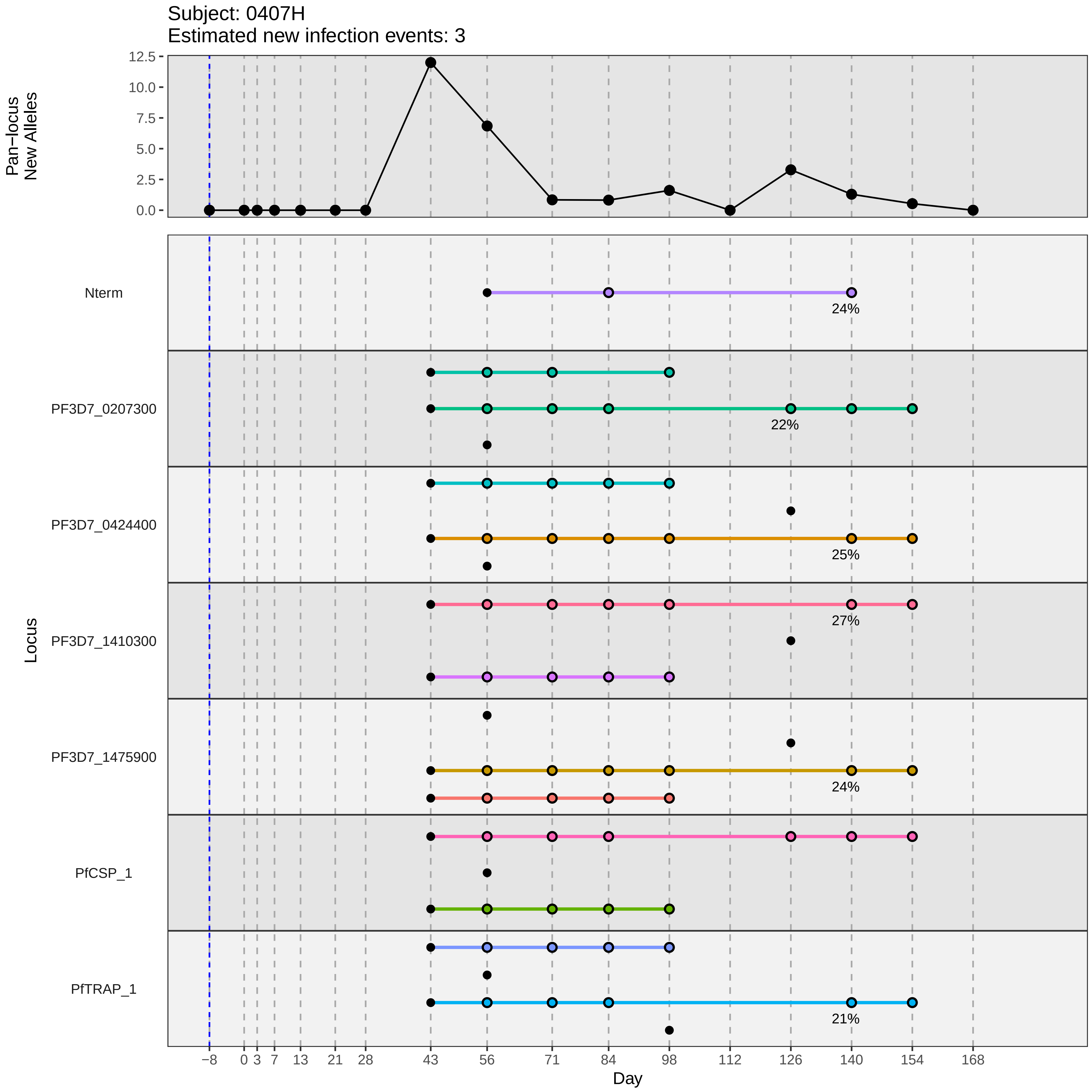
Subject: 0407H
Estimated new infection events: 3
Pan−locus
New Alleles
12.5
10.0
7.5
5.0
2.5
0.0
Locus
Nterm
24%
PF3D7_0207300
22%
PF3D7_0424400
25%
PF3D7_1410300
27%
PF3D7_1475900
24%
PfCSP_1
PfTRAP_1
21%
−8
0
3
7
13
21
28
43
56
71
84
98
112
126
140
154
168
Day
