## Supplementary material for "Genotypic infection endpoint analysis to understand efficacy and escape potential of the malaria monoclonal antibody CIS43LS": Data File S2: participant_0413R_without_imputation.pdf

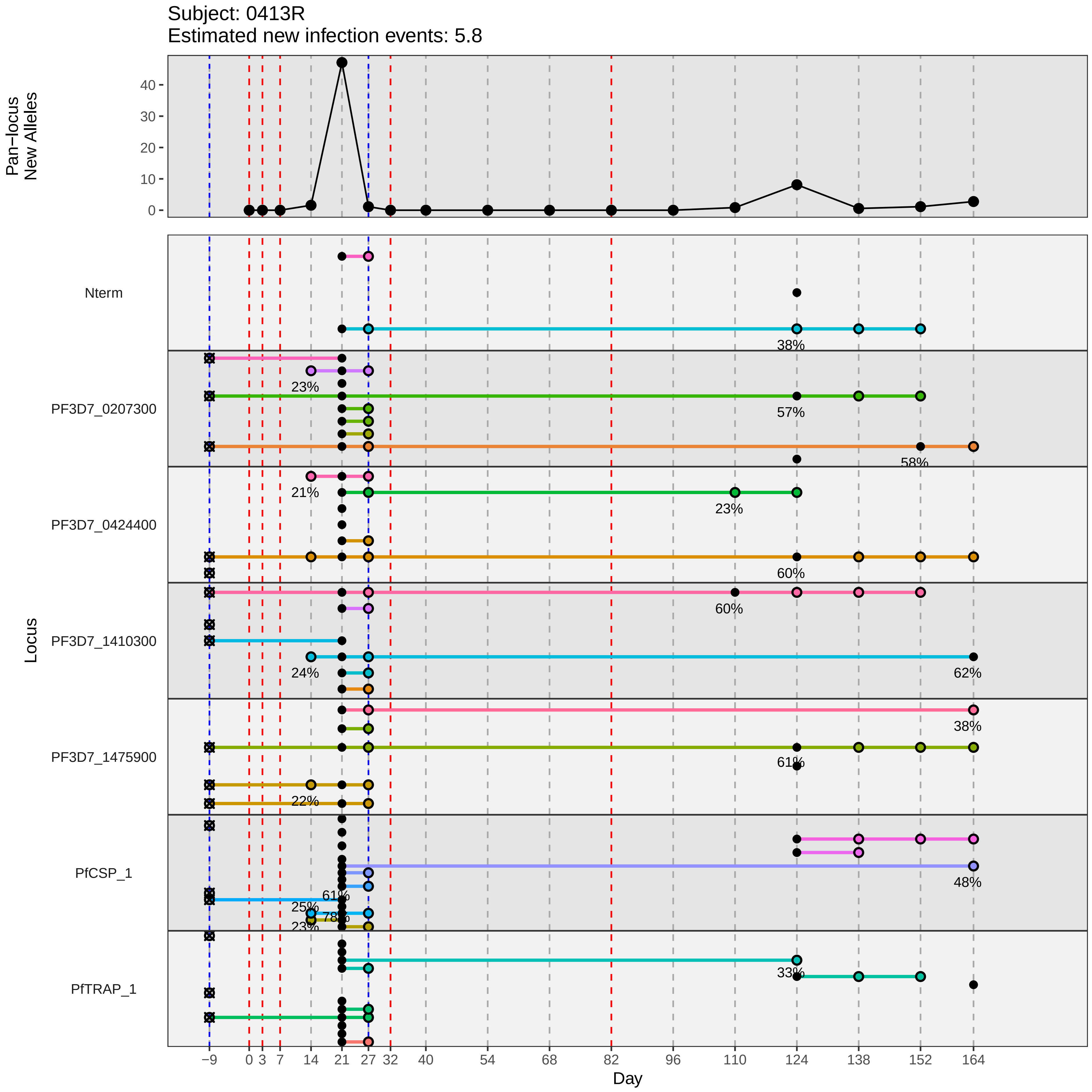
Subject: 0413R
Estimated new infection events: 5.8
Pan−locus
New Alleles
0
10
20
30
40
Locus
Nterm
PF3D7_0207300
PF3D7_0424400
PF3D7_1410300
PF3D7_1475900
PfCSP_1
PfTRAP_1
38%
23%
57%
58%
21%
23%
60%
60%
24%
62%
38%
61%
22%
48%
61%
25%
78%
23%
33%
−9
0
3
7
14
21
27
32
40
54
68
82
96
110
124
138
152
164
Day
