## Supplementary material for "Genotypic infection endpoint analysis to understand efficacy and escape potential of the malaria monoclonal antibody CIS43LS": Data File S2: participant_0414S_without_imputation.pdf

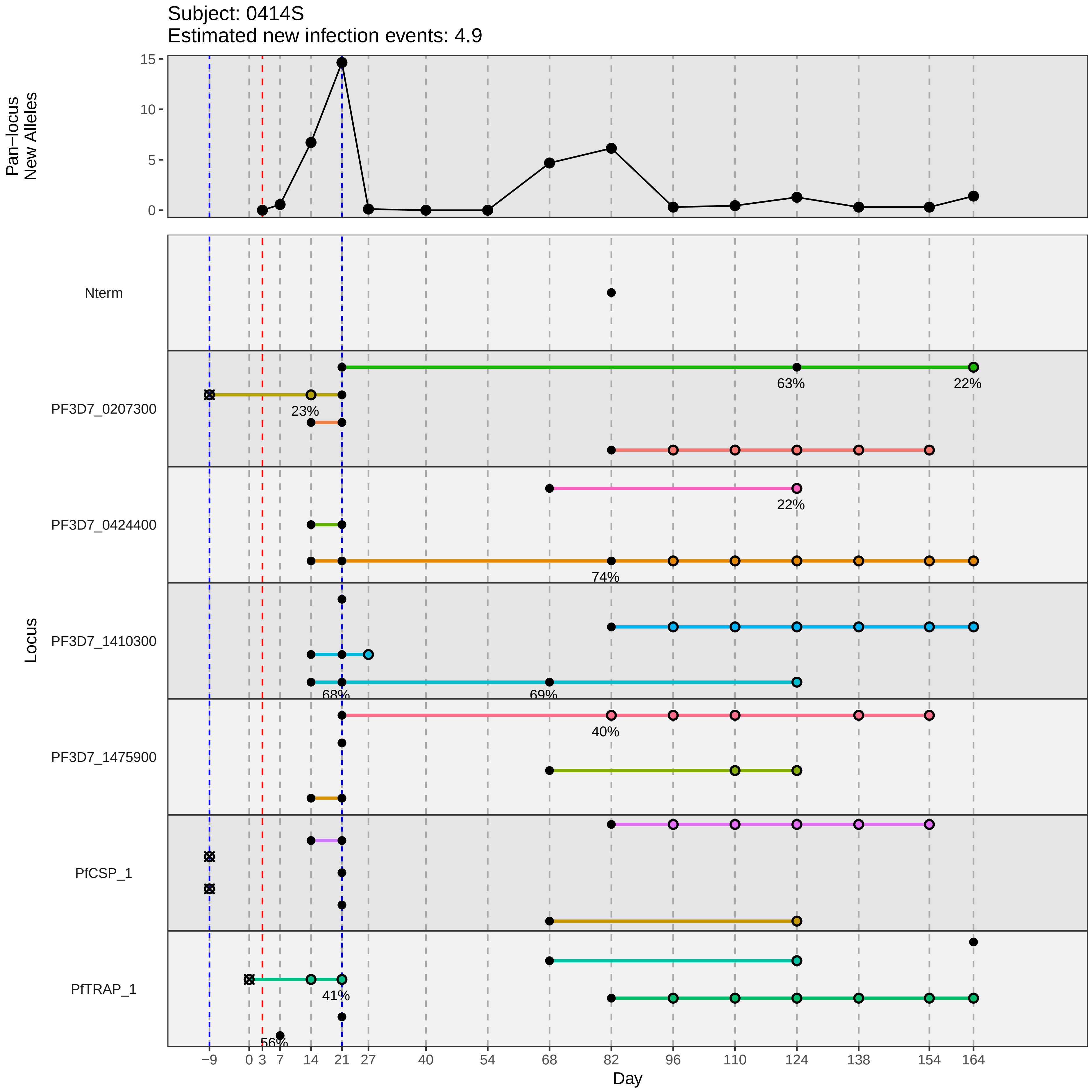
Subject: 0414S
Estimated new infection events: 4.9
Pan−locus New Alleles
15
10
5
0
Locus
Nterm
PF3D7_0207300
PF3D7_0424400
PF3D7_1410300
PF3D7_1475900
PfCSP_1
PfTRAP_1
63%
22%
23%
22%
74%
68%
69%
40%
41%
56%
−9
0
3
7
14
21
27
40
54
68
82
96
110
124
138
154
164
Day
