## Supplementary material for "Genotypic infection endpoint analysis to understand efficacy and escape potential of the malaria monoclonal antibody CIS43LS": Data File S2: participant_0416U_without_imputation.pdf

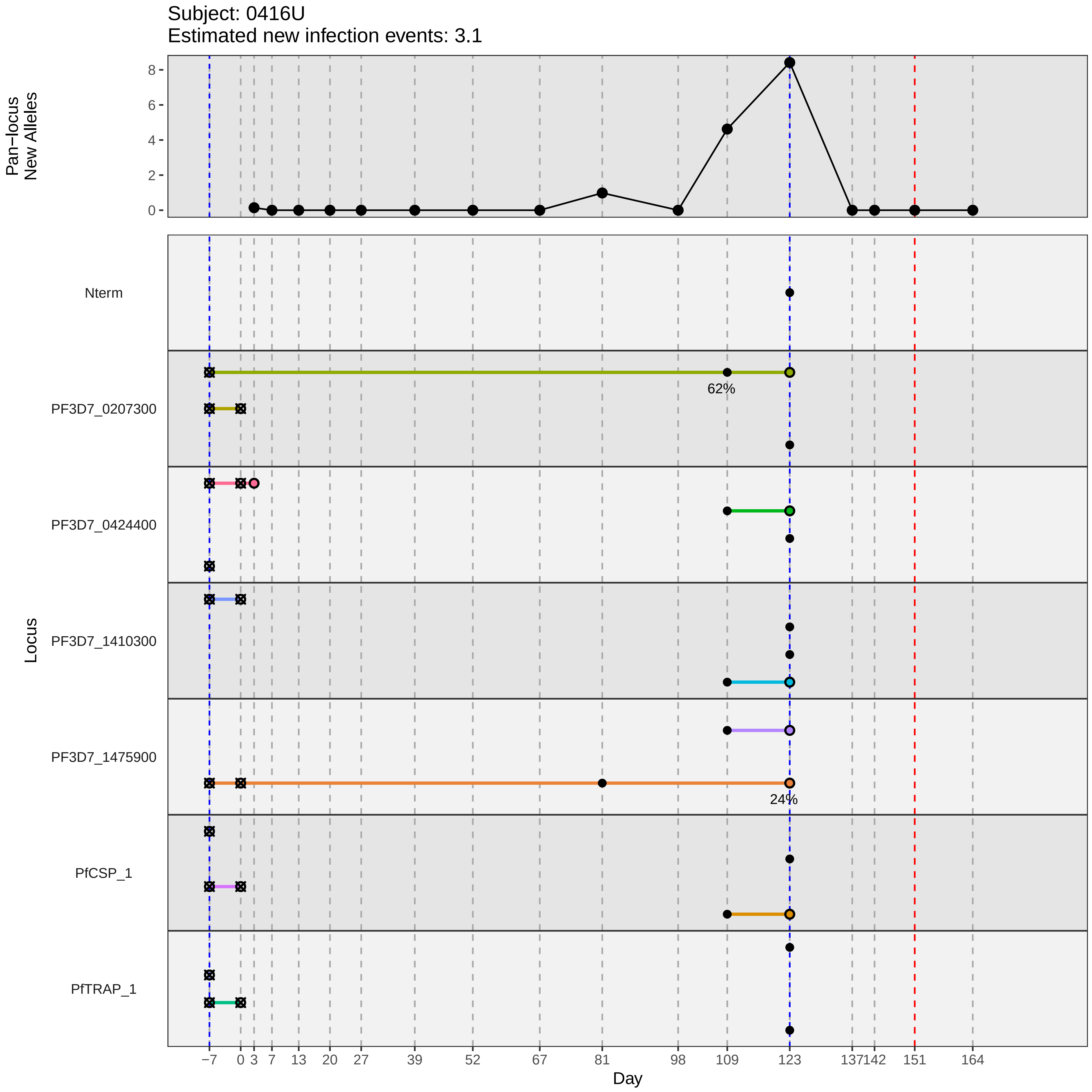
Subject: 0416U
Estimated new infection events: 3.1
Pan–locus New Alleles
8
6
4
2
0
Locus
Nterm
PF3D7_0207300
62%
PF3D7_0424400
PF3D7_1410300
PF3D7_1475900
24%
PfCSP_1
PfTRAP_1
−7
0
3
7
13
20
27
39
52
67
81
98
109
123
137
142
151
164
Day
