## Supplementary material for "Genotypic infection endpoint analysis to understand efficacy and escape potential of the malaria monoclonal antibody CIS43LS": Data File S2: participant_0418W_without_imputation.pdf

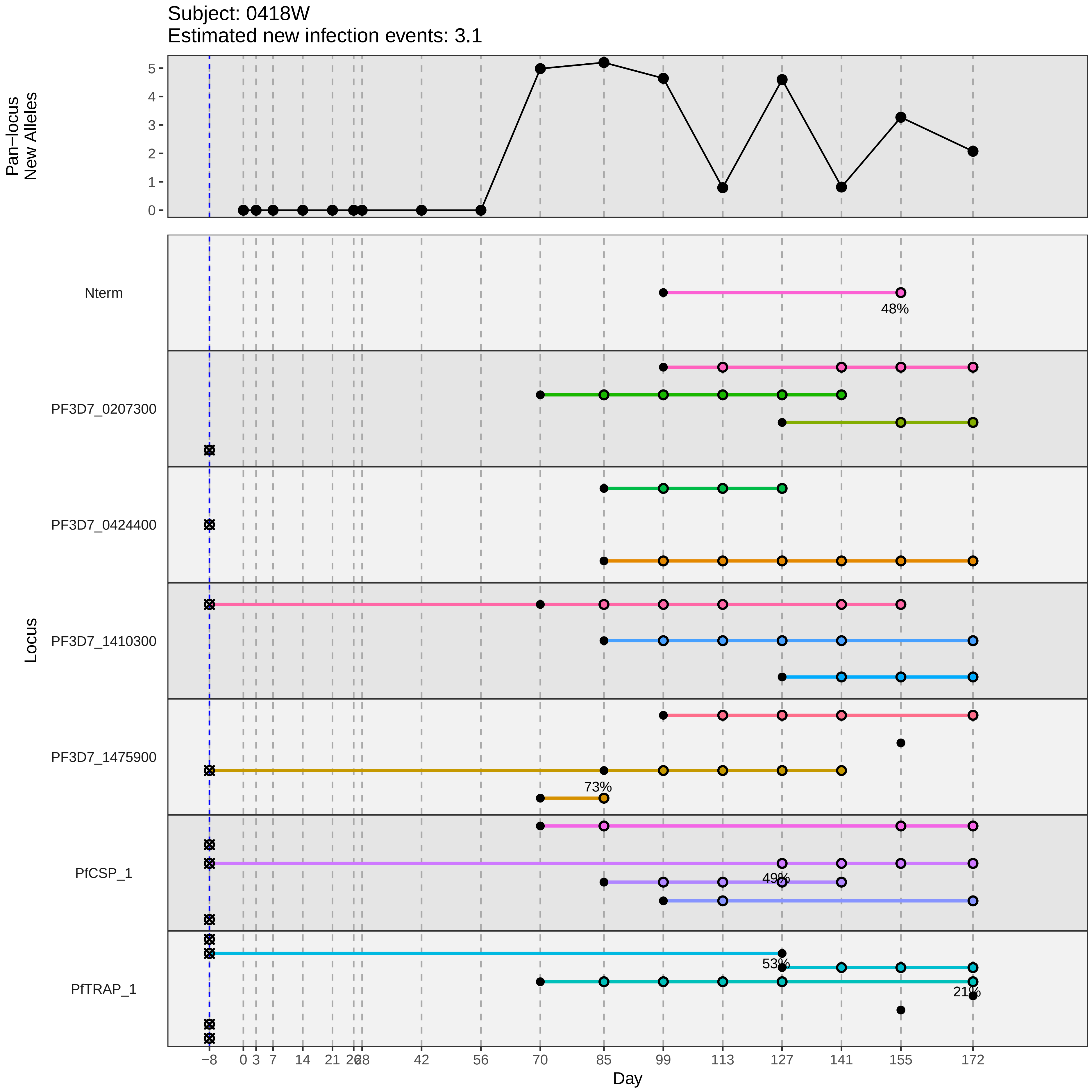

Subject: 0418W
Estimated new infection events: 3.1
Pan−locus New Alleles
0
1
2
3
4
5
Locus
Nterm
48%
PF3D7_0207300
PF3D7_0424400
PF3D7_1410300
PF3D7_1475900
73%
PfCSP_1
49%
PfTRAP_1
53%
21%
−8
0
3
7
14
21
28
42
56
70
85
99
113
127
141
155
172
Day
