## Supplementary material for "Genotypic infection endpoint analysis to understand efficacy and escape potential of the malaria monoclonal antibody CIS43LS": Data File S2: participant_0427G_without_imputation.pdf

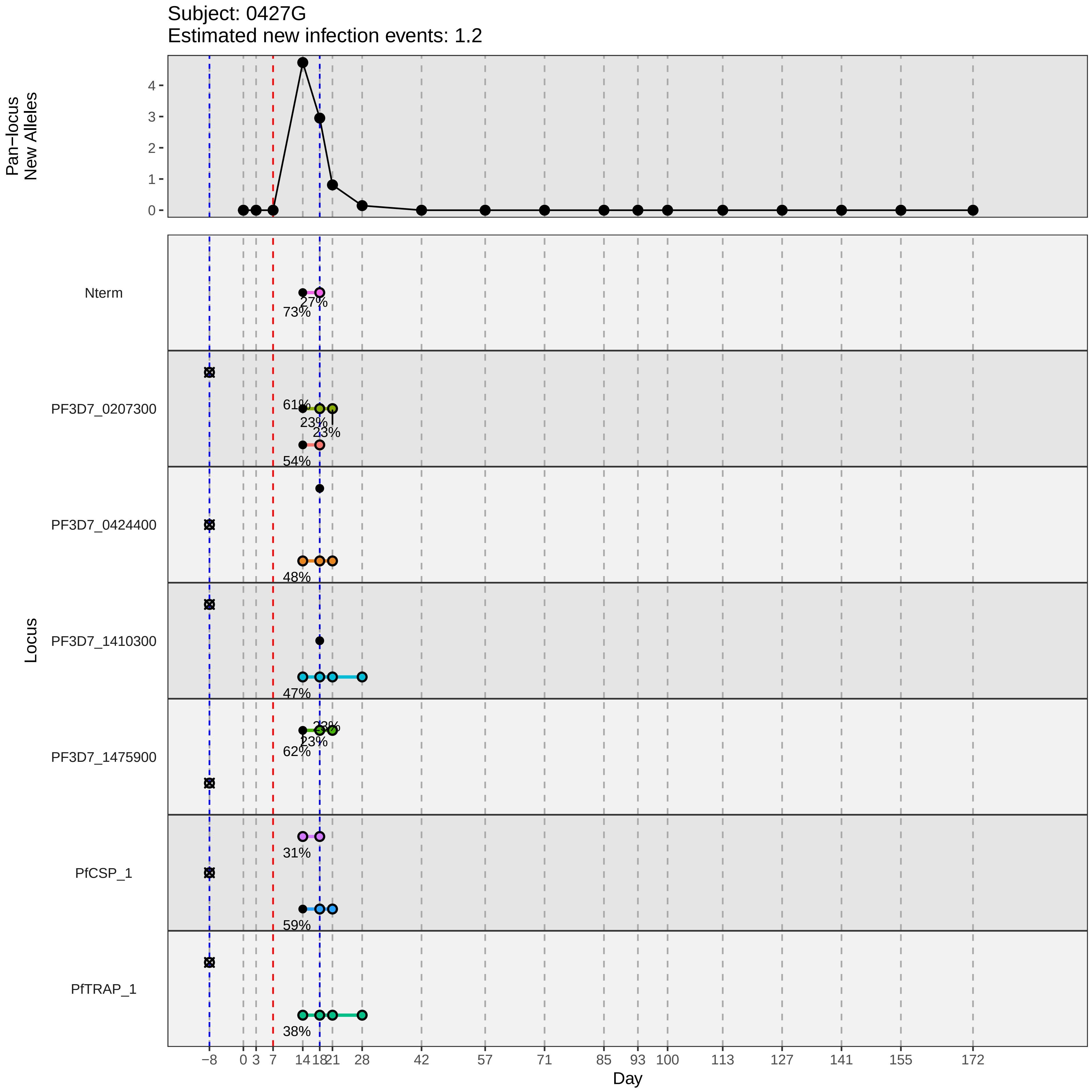
Subject: 0427G
Estimated new infection events: 1.2
Pan−locus New Alleles
4
3
2
1
0
Locus
Nterm
PF3D7_0207300
PF3D7_0424400
PF3D7_1410300
PF3D7_1475900
PfCSP_1
PfTRAP_1
27%
73%
61%
23%
23%
54%
48%
47%
23%
23%
62%
31%
59%
38%
−8
0
3
7
14
18
21
28
42
57
71
85
93
100
113
127
141
155
172
Day
