## Supplementary material for "Genotypic infection endpoint analysis to understand efficacy and escape potential of the malaria monoclonal antibody CIS43LS": Data File S2: participant_0430M_without_imputation.pdf

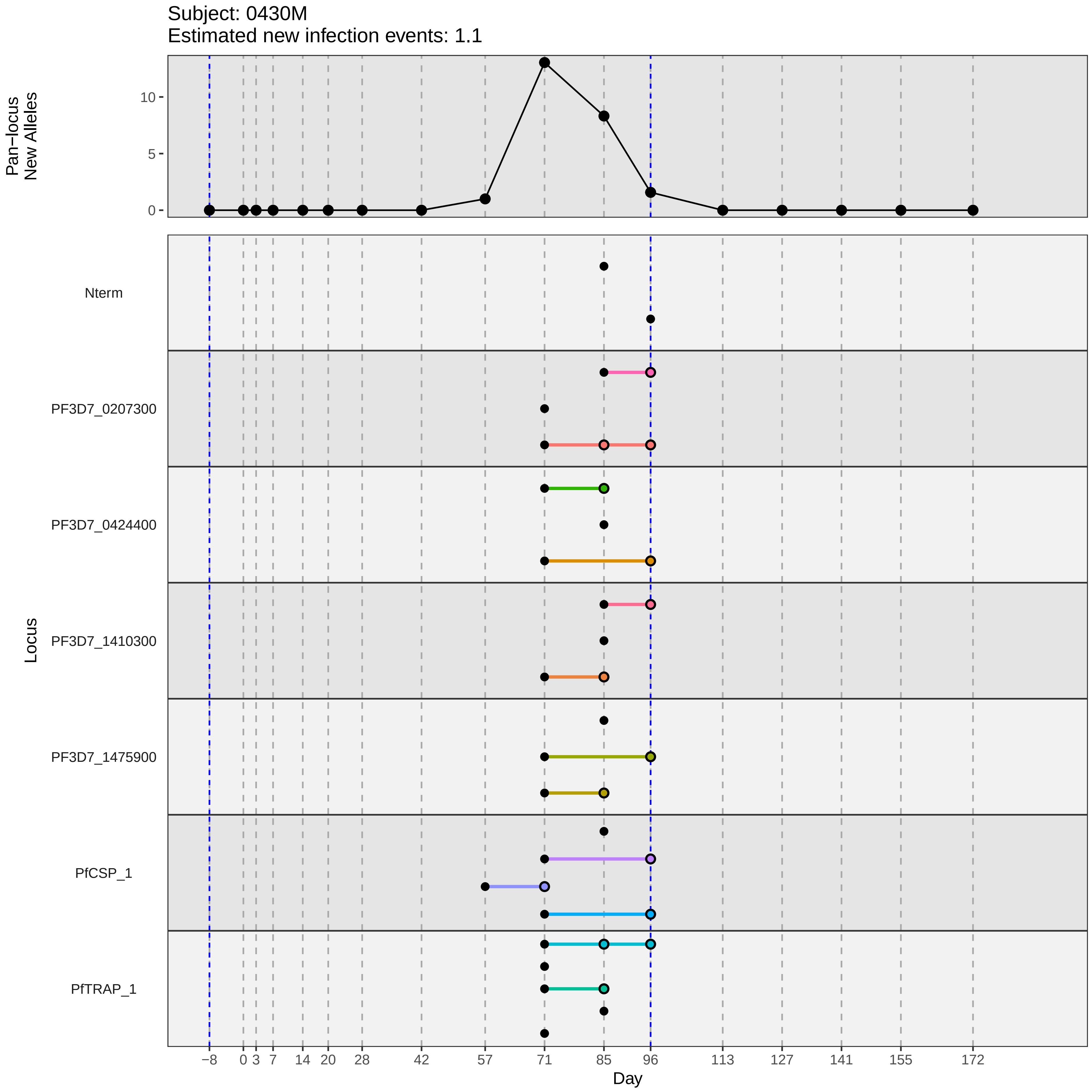

Subject: 0430M
Estimated new infection events: 1.1
Pan−locus New Alleles
0
5
10
Locus
Nterm
PF3D7_0207300
PF3D7_0424400
PF3D7_1410300
PF3D7_1475900
PfCSP_1
PfTRAP_1
−8
0
3
7
14
20
28
42
57
71
85
96
113
127
141
155
172
Day
